# Maternal and Fetal Complications Among Pregnant Women with Congenital Heart Disease

**DOI:** 10.1101/2025.02.17.25322188

**Authors:** Shayli Patel, Cheryl Raskind-Hood, Travis Wilson, Fred H. Rodriguez, Alex Haffner, Vijaya Kancherla, Lindsey Ivey, Wendy M. Book

## Abstract

**Background:** Survival to reproductive age is now common among women with congenital heart disease (CHD). Pregnancies to women with CHD may be at higher risk for maternal and fetal complications. This study examines maternal and fetal outcomes among pregnant women with CHD covered by Georgia Medicaid.

**Methods:** This case-control study identified 3,461 pregnant women with CHD, 11-50 years, covered by Georgia Medicaid from 2008-2019, who had 5,904 pregnancies. Maternal and fetal outcomes were determined by ICD-9-CM and ICD-10-CM codes at the encounter level. The association of select covariables, including race/ethnicity, social deprivation, CHD native anatomy, rurality, and presence of pre-existing maternal comorbidities on maternal and fetal outcomes were analyzed using chi-square. Multivariate logistic regression models adjusting for covariates were conducted.

**Results:** Women with severe CHD had more fetal complications and fetal deaths/stillbirths than women with non-severe CHD (53.9% vs. 50.2% p=0.01 and 4.4% vs. 3.0%, p=0.007, respectively), but no difference in maternal complications were revealed (60.7% vs. 59.9%, p=ns). In multivariate analysis, pregnancies to women with severe CHD were 25% more likely to have fetal complications than pregnancies to women with non-severe CHD (aOR 1.25, 95CI 1.07-1.47). The most common fetal complications by maternal CHD anatomy were fetal distress, fetal growth restriction, and preterm delivery (27.2% vs. 23.1%, 27.6% vs. 19.4%, 23.1% vs. 19.6% for severe vs. non-severe, respectively, p=<0.05).

**Conclusions:** Pregnancies to women with CHD were associated with a high rate of maternal and fetal complications. Fetal complications, fetal death and stillbirth were highest among women with severe CHD.

## BACKGROUND

Survival to reproductive age is now common for women born with congenital heart disease (CHD).^1,2^ Today, more than 500,000 women over the age of 18 are living with CHD in the U.S., and the number of pregnancies among this growing population continues to increase annually.^3^ For women with CHD, pregnancy poses an increased risk of complications for both mother and fetus, as pregnancy places increased demand on the maternal cardiovascular system requiring augmentation of stroke volume, heart rate, cardiac output, plasma volume, and a physiologic hypercoagulable state.^4^ Hemodynamic changes throughout pregnancy and during labor and delivery can be particularly detrimental to patients with cardiovascular diseases due to limited ability to augment cardiac output, difficulty tolerating changes in plasma volume, and adverse effects of a hypercoagulable state on women with a variety of CHD-related conditions.^5^

Major adverse cardiac events related to pregnancy are more common among patients with CHD compared to those without CHD, and the level of risk increases with increasing severity of CHD.^6^ Cardiac impairment is associated with abnormal patterns of uteroplacental doppler flow (UDF) and, in addition to adverse cardiac outcomes, CHD may contribute to other obstetric, fetal, and neonatal complications.^7^ Maternal complications include preterm labor, caesarian delivery, bleeding or thrombosis, gestational hypertension or pre-eclampsia, and death.^1,2,8^ Fetal complications include prematurity, low birth weight, intrauterine growth restriction (IUGR), spontaneous abortion, and stillbirth.^1,2^ The level of risk of both maternal and fetal/morbidity and mortality are dependent on physiology and severity of the CHD as well as social factors such as ability to access specialty care.

Compared to pregnant White women, pregnant Black and Hispanic women have a higher risk of obstetric complications and mortality.^9–11^ These disparities may be related to several factors, including delivery in lower quality hospitals, lack of access to adequate prenatal care, and racial and ethnic discrimination.^10^ Black ethnicity has also been associated with an increased risk of miscarriage compared to White ethnicity,^12^ and Black infants have been found to have significantly worse birth outcomes compared to White infants.^13,14^ Maternal morbidity and mortality are also significantly worse among women living in rural areas compared to those living in urban areas.^15^ Disparities in outcomes may be due to clinical factors such as workforce shortages, and to social determinants such as lack of transportation, lack of paid leave, barriers related to childcare access, food insecurity, and poverty.^16,17^ The inability to access adequate care in rural areas is associated with significantly worse maternal and fetal outcomes.^11,17^

The aim of this study was to compare maternal and fetal complications among pregnant women with CHD residing in Georgia covered under Medicaid by maternal CHD severity, age, socioeconomic deprivation, race, rurality and other characteristics for those with the same insurance coverage.

## METHODS

The study was approved by the Emory University Institutional Review Board on July 10, 2023 (IRB # STUDY00006154) which included a complete waiver of HIPAA authorization and informed consent. The cohort was derived from 173,083 Georgians covered by Georgia Medicaid at some point during 2008-2019 with at least one select CHD-related ICD code.^18^ Those whose date of birth (DOB) was prior to 01/01/1958 or after 01/01/2009 (n=84,408), those without an encounter (n=21,262) and those without a CHD diagnosed during the study period (n=14,423) were excluded. In addition, male Medicaid beneficiaries (24,409), women without a documented pregnancy encounter during the study period (n=23,063), and Medicaid beneficiaries with an ICD diagnostic code 745.5 or Q21.1 in isolation [secundum atrial septal defect (ASD) or patent foramen ovale (PFO)] were not included in the analytic cohort due to lack of specificity of these codes (1,156).^31,32^ Lastly, those whose CHD-related ICD diagnostic codes categorized them as having an “Other” CHD (n=54) or “Other Vascular” condition (n=15) (**Table S1**) (n=69 combined), those with unknown race (n=206), those missing county of residence (n=20) and those whose Social Deprivation Index (SDI) could not be calculated (n=606) were also excluded. The final analytic dataset contained **3,461** women, aged 11-50 years, who experienced at least one pregnancy, and were covered by Georgia Medicaid between 2008-2019; among these women, **5,904** pregnancies were identified over the 12-year study period.

CHD was defined by ICD-9-CM and ICD-10-CM codes (**Table S1**), categorized by native anatomy, and classified as severe and non-severe for analysis. Pregnancy was defined by the Clinical Classification Software (CCS) based on ICD-9-CM, ICD-10-CM, and CPT codes outlined in **Table S2**. The beginning and end of each pregnancy as well as last menstrual period (LMP) and gestational age were estimated using ICD-9-CM codes described in Ailes et al. 2016’s methods, as well as ICD-10-CM codes described in Ailes et al. (2023) methods, with some modifications (**Table S3)**.^19,20^ Subsequent pregnancies were defined by a gap of a minimum of 2 months between the end of one pregnancy and the calculated LMP prior to the next pregnancy.

Outcomes included two types of pregnancy-related complications, **maternal complications (Table S4**) such as obstetric complications or other adverse maternal health effects related to pregnancy, and **fetal complications** (**Table S4**) including any adverse outcomes of the fetus, as well as **fetal death** (fetal demise after 20 weeks gestation), **stillbirth**, **and early pregnancy loss** (spontaneous or therapeutic abortion and ectopic pregnancy, **Table S4**). Having only access to maternal records, neonatal outcomes occurring shortly after birth were unavailable for analysis. Several predictor variables were assessed: race/ethnicity, CHD anatomic severity, geographic region (urban-rural residence), number of pregnancies, age at first documented pregnancy, and social deprivation. Since there were a small number of patients who identified as Asian (n=0), Native Hawaiian/Pacific Islander (n=0), American Indian/Alaska Native (n=6), or other racial groups (n=1), these racial groups were omitted from analysis. CHD anatomy was operationalized using a native anatomic group classification scheme, modified by project clinicians, which classified anatomic defects as: severe (CHD typically requiring intervention in the first year of life), shunt, valve, and shunt with valve. CHD anatomy was collapsed into two groups: severe vs. non-severe for analysis (**Table S1**). Urban-rural residence was determined using the 2013 NCHS (National Center for Health Statistics) Urban-Rural Classification Scheme for Counties where a 6-level county classification was collapsed into a dichotomous variable, urban or rural. Patients were also categorized by Social Deprivation Index (SDI), a composite social determinant of health measure, developed by the Robert Graham Center, Policy Studies in Family Medicine & Primary Care, to estimate the social deprivation of one’s residential area such as county, census tract, or ZCTA.^21–23^ This composite index, ranging from 0.0 to 100.0 and categorized into quintiles with higher SDI scores signifying a higher level of social deprivation, while lower scores indicating a lower level of deprivation, assesses seven salient social determinants of health/demographic characteristics collected by the American Community Survey (i.e., percent living in poverty, percent with less than 12 years of education, percent single-parent households, the percentage living in rented housing units, the percentage living in the overcrowded housing unit, percent of households without a car, and percentage unemployed adults under 65 years of age).^21,23,24^ For the current analyses, when a pregnancy fell across multiple years, the year of the end of the pregnancy was used to identify the appropriate SDI score. Additional covariates included were **number of pregnancies**, which was dichotomized as pregnant once or pregnant two or more times; **age at first pregnancy**, classified into three age groups: 11-19 years, 20-34 years, and 35-50 years; and **singleton pregnancy or higher order pregnancy. Pre-existing comorbid conditions** which included arrhythmia, coronary artery disease (CAD), diabetes mellitus (DM), heart failure (HF), hypertension (HTN), hyperlipidemia, obesity, pulmonary vascular disease and residual structural heart disease, operationalized by ICD-9-CM and ICD-10-CM codes, were analyzed and were collapsed into a composite measure, ‘any’ or ‘none’ (**Table S5**).

## ANALYSIS

All analyses were conducted using SAS version 9.4 statistical software (SAS institute, Cary, NC). Descriptive analysis included frequencies and percentages for all categorical variables. Differences in conditions included in maternal and fetal complications, fetal death and stillbirth, and early pregnancy loss were determined race/ethnic group, geographic residential group, and CHD anatomic severity group using chi-square analysis. In addition, multivariate logistic regression analyses were conducted using the backward elimination approach (BWE) for each of the four outcomes separately, while controlling for select predictors.

## RESULTS

### Cohort Description

For a total of 3,461 women with CHD who experienced at least one pregnancy between 2008-2019, bivariate analyses compared the association of each of the four outcomes with various select demographic characteristics (**Table 1**), and similar bivariate analyses for the 5,904 pregnancies for this cohort are displayed in **Table 2**. Overall, for race and ethnicity, 47.5% identified as non-Hispanic White (nHW), 50.1% identified as non-Hispanic Black (nHB), and only 2.4% of women identified as Hispanic/Latina. Over three quarters of the cohort, 77.4%, resided in urban Georgia areas. A little more than a quarter of the cohort (27.6%) had severe native CHD anatomy, while 72.4% were categorized as non-severe. At least one pre-existing comorbid condition was identified in 20.0% of women with CHD prior to a pregnancy. Compared to nHW women, nHB women were more likely to have comorbid conditions prior to pregnancy (26.6% vs 21.5%, p=0.002), with higher rates of obesity and diabetes driving the difference. The proportion of women with severe CHD (22.2%) who had pre-pregnancy comorbid conditions was similar compared to women with non-severe CHD (24.8%), (p=ns).

**Table 1.**
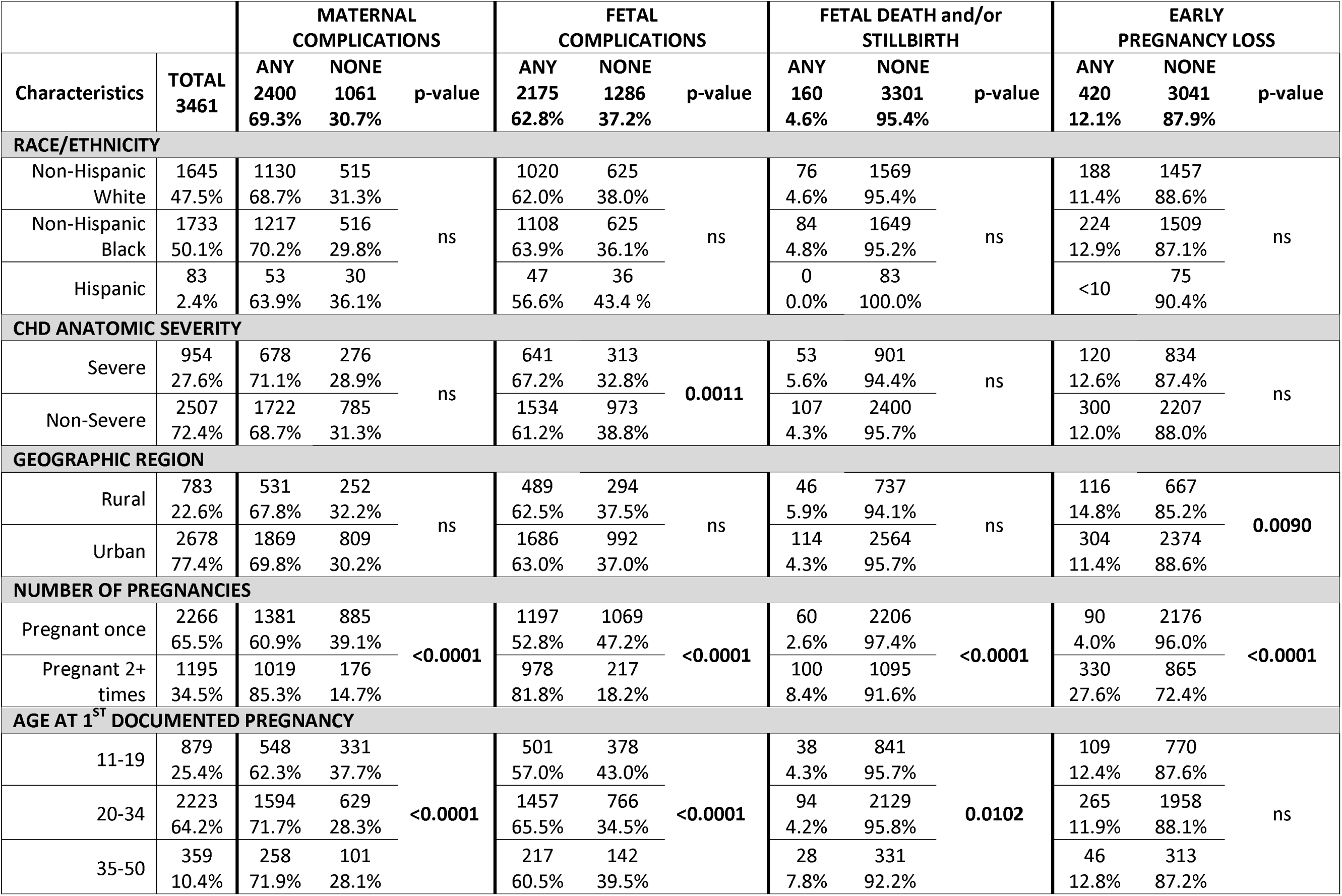
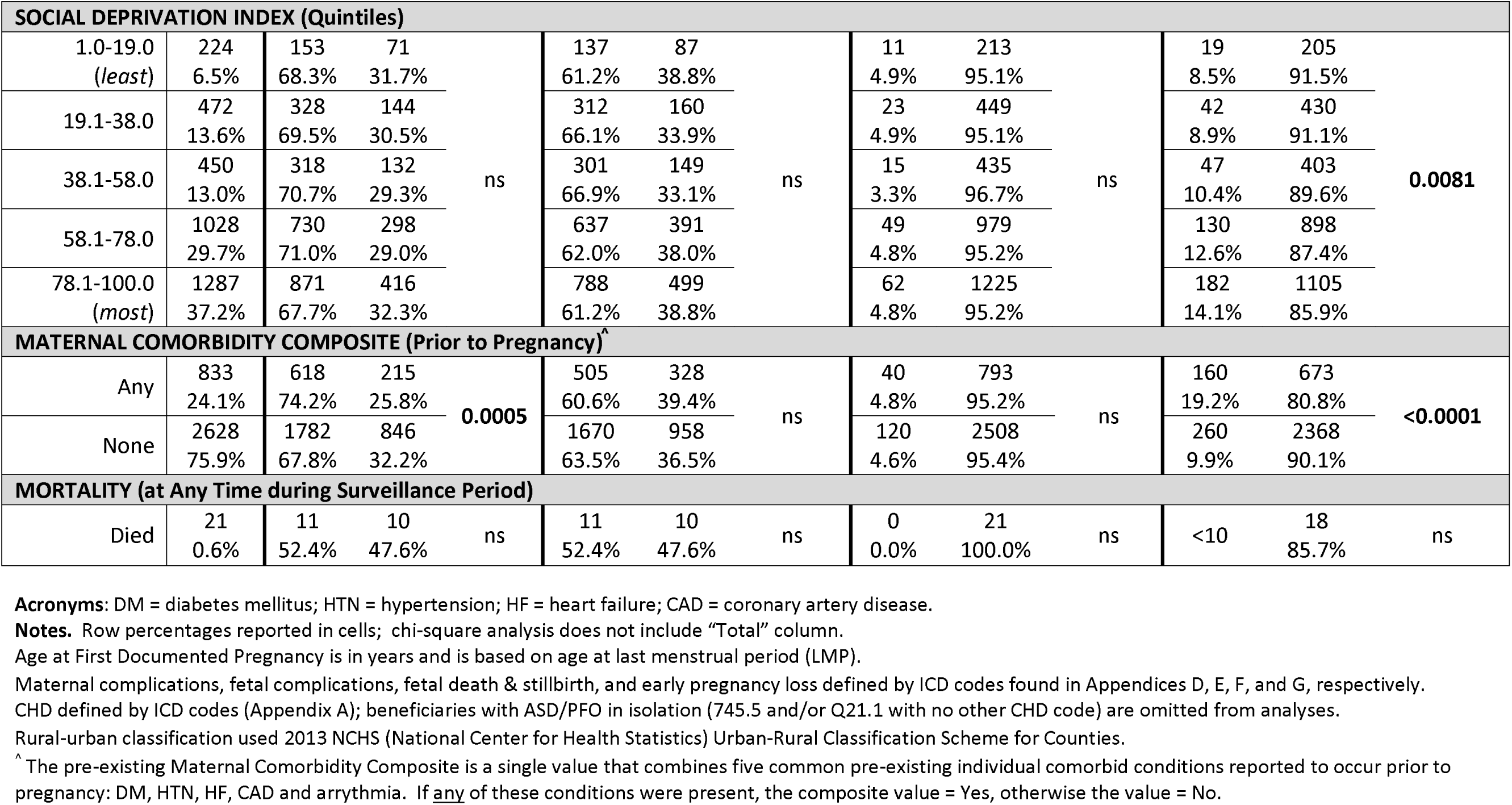
Characteristics of pregnant Medicaid beneficiaries with congenital heart defects (CHD), Total, and for those with and without maternal complications, fetal complications, fetal death and stillbirth, and early pregnancy loss, Georgia 2008-2019.

**Table 2.**
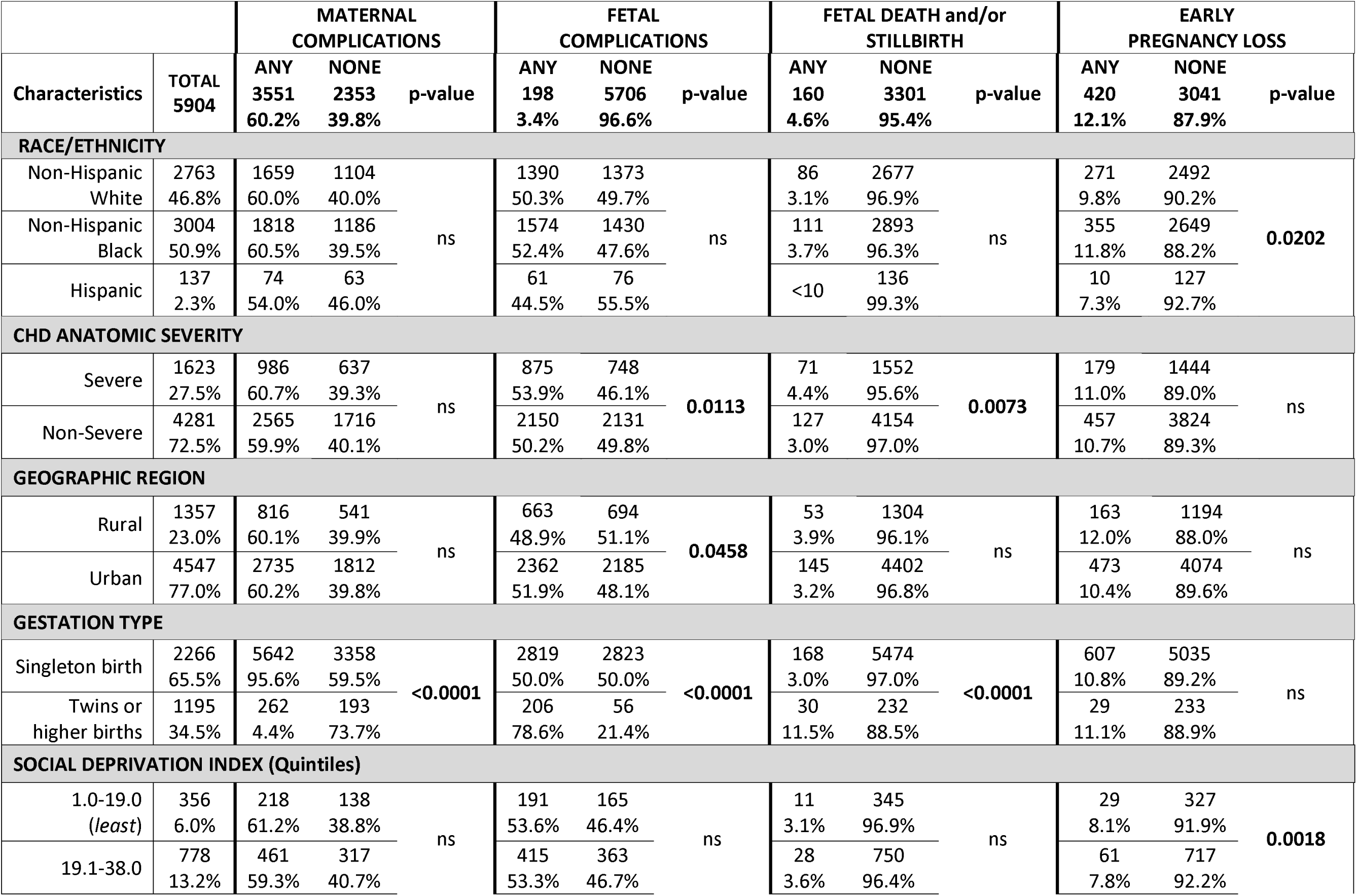
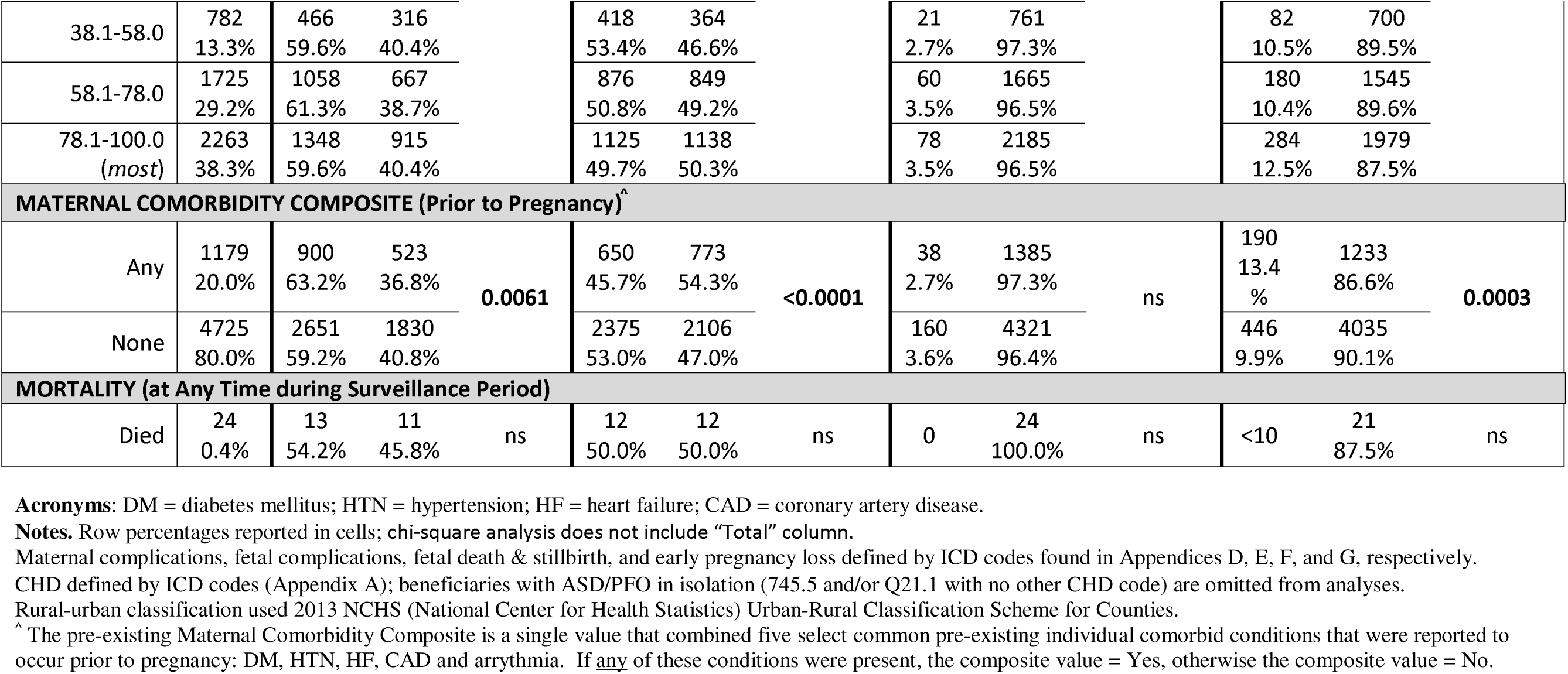
Characteristics of pregnancies among Medicaid beneficiaries with and without maternal complications, fetal complications, neonatal deaths, and early pregnancy loss, Georgia 2008-2019.

### Pregnancy-related maternal and fetal complications

As women may have experienced more than one pregnancy during the surveillance period, the characteristics of each pregnancy are shown in Table 2. Overall, 60.2% of pregnancies had at least one maternal complication, 51.2% had at least one fetal complication, 3.4% ended with fetal death, and 10.8% had early pregnancy loss defined as spontaneous or therapeutic abortion. There were no obstetric-related maternal deaths in the cohort. The specific pregnancy related maternal complications making up the maternal complications outcome, fetal complications making up the fetal complication outcome, fetal death and stillbirth categories as well as early pregnancy loss categories are shown in **Table 3** by CHD severity, race, ethnicity, and rurality. Findings for specific complications are discussed in subheadings in the results section below. Final adjusted multivariate logistic regression models were performed on maternal and fetal complications, fetal death, and early pregnancy loss and included race/ethnicity, CHD anatomic severity, age at first documented pregnancy, social deprivation index (in quintiles), number of gestations, and a comorbid condition composite variable (**Table 4**). Multiple gestations were associated with more adverse outcomes in all four categories. The odds of maternal complications were 2.37 times higher (95% CI 1.64-3.42), odds of fetal complications were 4.29 times higher (95% CI 2.87-6.43), odds of fetal death were 4.18 times higher (95% CI 2.75-6.37), and odds of early pregnancy loss were 2.03 times higher (95% CI 1.43-2.88) in pregnancies with multiple gestations than in singleton pregnancies.

**Table 3.** Maternal complications, fetal complications, fetal death and/or stillbirth, and early pregnancy loss by total and congenital heart defect (CHD) severity, race/ethnicity, and geographic region for pregnant Medicaid beneficiaries with CHD, Georgia 2008-2019.

|  | TOTAL<br>3461 | CHD ANATOMIC SEVERITY |  |  | RACE/ETHNICITY |  |  |  | GEOGRAPHIC REGION |  |  |
| --- | --- | --- | --- | --- | --- | --- | --- | --- | --- | --- | --- |
|  |  | Severe | Non-Severe | p-value | nHW | nHB | Hispanic | p-value | Rural | Urban | p-value |
| **ANY MATERNAL COMPLICATIONS** | **2400**<br>**69.3%** | 678<br>71.1% | 1722<br>68.7% | ns | 1130<br>68.7% | 1217<br>70.2% | 53<br>63.9% | ns | 531<br>67.8% | 1869<br>69.8% | ns |
| **Anemia in Preg (Gestational)** | **745**<br>**21.5%** | 208<br>21.8% | 537<br>21.4% | ns | **286**<br>**17.4%** | **440**<br>**25.4%** | **19**<br>**22.9%** | **<0.0001** | 163<br>20.8% | 582<br>21.7% | ns |
| **Preterm Labor, Threatened (OB)** | **663**<br>**19.2%** | 169<br>17.7% | 494<br>19.7% | ns | 331<br>20.1% | 317<br>18.3% | 15<br>18.1% | ns | **173**<br>**22.1%** | **490**<br>**18.3%** | **0.0175** |
| **Mental Health & Substance Abuse Disorders (Gestational)** | **551**<br>**15.9%** | 163<br>17.1% | 388<br>15.5% | ns | **359**<br>**21.8%** | **189**<br>**10.9%** | **<10** | **<0.0001** | 142<br>18.1% | 409<br>15.3% | **0.0541** |
| **Poly- or Oligohydramnios (OB)** | **541**<br>**15.6%** | **177**<br>**18.5%** | **364**<br>**14.5%** | **0.0035** | 274<br>16.7% | 253<br>14.6% | 14<br>16.9% | ns | 118<br>15.1% | 423<br>15.8% | ns |
| **Hemorrhage (OB)** | **457**<br>**13.2%** | 129<br>13.5% | 328<br>13.1% | ns | 204<br>12.4% | 244<br>14.1% | **<10** | ns | **123**<br>**15.7%** | **334**<br>**12.5%** | **0.0186** |
| **Diabetic Complications of Pregnancy (Gestational)** | **440**<br>**12.7%** | 132<br>13.8% | 308<br>12.3% | ns | 206<br>12.5% | 220<br>12.7% | 14<br>16.9% | ns | 94<br>12.0% | 346<br>12.9% | ns |
| **Premature Rupture of Membrane (OB)** | **405**<br>**11.7%** | 112<br>11.7% | 293<br>11.7% | ns | 172<br>10.5% | 223<br>12.9% | 10<br>12.0% | ns | 89<br>11.4% | 316<br>11.8% | ns |
| **Preeclampsia, Eclampsia, Toxemia & HELLP (OB)** | **392**<br>**11.3%** | 111<br>11.6% | 281<br>11.2% | ns | **155**<br>**9.4%** | **227**<br>**13.1%** | **10**<br>**12.0%** | **0.0033** | 91<br>11.6% | 301<br>11.2% | ns |
| **Hypertensive Complications of Pregnancy (Gestational)** | **221**<br>**6.4%** | 61<br>6.4% | 160<br>6.4% | ns | 101<br>6.1% | 117<br>6.7% | **<10** | ns | 44<br>5.6% | 177<br>6.6% | ns |
| **Composite of Other Maternal Complication<sup>^</sup>** | **467**<br>**13.5%** | 121<br>12.7% | 346<br>13.8% | ns | 209<br>12.7% | 242<br>14.0% | 16<br>19.3% | ns | 93<br>11.9% | 374<br>14.0% | ns |
| **ANY FETAL COMPLICATIONS<sup>^^</sup>** | **2175**<br>**62.8%** | **641**<br>**67.2%** | **1534**<br>**61.2%** | **0.0011** | 1020<br>62.0% | 1108<br>63.9% | 47<br>56.6% | ns | 489<br>62.5% | 1686<br>63.0% | ns |
| **Fetal Distress** | **841**<br>**24.3%** | **260**<br>**27.2%** | **581**<br>**23.1%** | **0.0124** | **346**<br>**21.0%** | **476**<br>**27.5%** | **19**<br>**22.9%** | **<0.0001** | 203<br>25.9% | 638<br>23.8% | ns |
| **Fetal Growth Restriction** | **749**<br>**21.6%** | **263**<br>**27.6%** | **486**<br>**19.4%** | **<0.0001** | 351<br>21.3% | 385<br>22.2% | 13<br>15.7% | ns | 168<br>21.5% | 581<br>21.7% | ns |
| **Preterm delivery** | **711**<br>**20.5%** | **220**<br>**23.1%** | **491**<br>**19.6%** | **0.0237** | 325<br>19.8% | 374<br>21.6% | 12<br>14.5% | ns | 145<br>18.5% | 566<br>21.1% | ns |
| **Excess Fetal Growth** | **255**<br>**7.4%** | **44**<br>**4.6%** | **211**<br>**8.4%** | **<0.0001** | **144**<br>**8.7%** | **105**<br>**6.1%** | **<10** | **0.0112** | 67<br>8.6% | 188<br>7.0% | ns |
| **Placental Complications Affecting Newborn** | **164**<br>**4.7%** | **87**<br>**9.1%** | **77**<br>**3.1%** | **<0.0001** | 76<br>4.6% | 87<br>5.0% | <10 | ns | 31<br>4.0% | 133<br>5.0% | ns |
| **ANY FETAL DEATH AND/OR STILLBIRTH<sup>^^^</sup>** | **160**<br>**4.6%** | 53<br>5.6% | 107<br>4.3% | ns | 76<br>4.6% | 84<br>4.8% | - | ns | 46<br>5.9% | 114<br>4.3% | ns |
| **Fetal Demise, Death or Stillborn (Neonatal)** | **153**<br>**4.4%** | 49<br>5.1% | 104<br>4.1% | ns | 72<br>4.4% | 81<br>4.7% | - | ns | 42<br>5.4% | 111<br>4.1% | ns |
| **ANY EARLY PREGNANCY LOSS** | **420**<br>**12.1%** | 120<br>12.6% | 300<br>12.0% | ns | 1188<br>1.4% | 224<br>12.9% | <10 | ns | **116**<br>**14.8%** | **304**<br>**11.4%** | **0.0090** |
| **Spontaneous Abortion** | **328**<br>**9.5%** | 94<br>9.8% | 234<br>9.3% | ns | 152<br>9.2% | 170<br>9.8% | <10 | ns | **89**<br>**11.4%** | **239**<br>**8.9%** | **0.0402** |
| **Ectopic Pregnancy (Maternal)** | **102**<br>**2.9%** | 30<br>3.1% | 72<br>2.9% | ns | 44<br>2.7% | 55<br>3.2% | <10 | ns | 28<br>3.6% | 74<br>2.8% | ns |
| **Termination of Pregnancy** | **28**<br>**0.8%** | <10 | 19<br>0.8% | ns | <10 | 19<br>1.1% | <10 | ns | <10 | 21<br>0.8% | ns |

**Acronyms:** nHW = non-Hispanic White; nHB = non-Hispanic Black; OB = obstetrics; Multi = multi-gestational; HELLP = hemolysis, elevated liver enzymes, low platelet count.
**Notes.** Column percentages reported.
Maternal Complications, Fetal Complications, Fetal Death and/or Stillbirth, and Early Pregnancy Loss defined by ICD codes in Appendices D, E, F, and G, respectively.
<sup>^</sup> Composite of Other Maternal Complications includes previa, abruption & abnormal placental attachment (OB), pregnancy-related Infections (OB), cervical incompetence & abnormal cervix (OB), thromboembolic complications (DVT, PE, CVA, Thrombosis) (gestational), placental insufficiency (OB), liver & biliary tract disorders of pregnancy (gestational), peripartum cardiomyopathy (gestational), anesthesia complications (OB), malignancy (gestational), obstetric air embolism (OB), amniotic fluid embolism (OB).
<sup>^^</sup> For Any Fetal Complications, fetal hemorrhage is included but not reported here due to infrequency.
<sup>^^^</sup> For Any Fetal Death and/or Stillbirth, fetal death within multi-gestational pregnancy (multi) and liveborn & stillborn (neonatal) are included but not reported here due to infrequency.

**Table 4.** Adjusted Associations of select covariates with maternal and fetal complications of pregnancies, fetal death and stillbirth, and early pregnancy loss among Medicaid beneficiaries with congenital heart disease (CHD), Georgia 2008-2019.

|  | **MATERNAL COMPLICATIONS** |  | **FETAL COMPLICATIONS** |  | **FETAL DEATH & STILLBIRTH** |  | **EARLY PREGNANCY LOSS** |  |
| --- | --- | --- | --- | --- | --- | --- | --- | --- |
|  | aOR | 95% CI | aOR | 95% CI | aOR | 95% CI | aOR | 95% CI |
| **RACE/ETHNICITY** |  |  |  |  |  |  |  |  |
| Non-Hispanic White | 1.00 | -- | 1.00 | -- | 1.00 | -- | 1.00 | -- |
| Non-Hispanic Black | 1.04 | 0.88-1.21 | 1.12 | 0.96-1.30 | 1.01 | 0.72-1.43 | 1.02 | 0.81-1.27 |
| Hispanic | 0.84 | 0.53-1.34 | 0.82 | 0.52-1.29 | -- | -- | 0.87 | 0.41-1.84 |
| **GEOGRAPHIC REGION** |  |  |  |  |  |  |  |  |
| Urban | 1.00 | -- | 1.00 | -- | 1.00 | -- | 1.00 | -- |
| Rural | 0.96 | 0.80-1.16 | 1.07 | 0.90-1.28 | **1.51** | **1.02-2.22** | 1.21 | 0.94-1.55 |
| **CHD ANATOMIC COMPLEXITY** |  |  |  |  |  |  |  |  |
| Severe | 1.08 | 0.91-1.28 | **1.25** | **1.07-1.47** | 1.30 | 0.92-1.84 | 1.09 | 0.86-1.37 |
| Non-Severe | 1.00 | -- | 1.00 | -- | 1.00 | -- | 1.00 | -- |
| **AGE AT 1<sup>ST</sup> DOCUMENTED PREGNANCY** |  |  |  |  |  |  |  |  |
| 11-19 | **0.64** | **0.54-0.76** | **0.72** | **0.62-0.85** | 1.06 | 0.72-1.57 | 0.98 | 0.77-1.25 |
| 20-34 | 1.00 | -- | 1.00 | -- | 1.00 | -- | 1.00 | -- |
| 35-50 | 0.97 | 0.76-1.25 | 0.80 | 0.64-1.01 | **1.93** | **1.24-3.05** | 1.03 | 0.73-1.44 |
| **SOCIAL DEPRIVATION INDEX (Quintiles)** |  |  |  |  |  |  |  |  |
| 1.0-19.0 (least) | 1.00 | -- | 1.00 | -- | 1.00 | -- | 1.00 | -- |
| 19.1-38.0 | 1.05 | 0.74-1.48 | 1.18 | 0.85-1.65 | 0.92 | 0.43-1.94 | 1.05 | 0.59-1.86 |
| 38.1-58.0 | 1.12 | 0.79-1.60 | 1.22 | 0.87-1.72 | 0.57 | 0.25-1.27 | 1.23 | 0.70-2.16 |
| 58.1-78.0 | 1.14 | 0.82-1.57 | 0.97 | 0.71-1.32 | 0.81 | 0.41-1.63 | 1.47 | 0.88-2.47 |
| 78.1-100.0 (most) | 0.96 | 0.69-1.32 | 0.91 | 0.67-1.24 | 0.76 | 0.38-1.53 | 1.60 | 0.95-2.69 |
| **GESTATION TYPE** |  |  |  |  |  |  |  |  |
| Singleton birth | 1.00 | -- | 1.00 | -- | 1.00 | -- | 1.00 | -- |
| Twins or higher births | **2.37** | **1.64-3.42** | **4.29** | **2.87-6.43** | **4.18** | **2.75-6.37** | **2.03** | **1.43-2.88** |
| **MATERNAL COMORBIDITY COMPOSITE (PRIOR TO PREGNANCY)<sup>^</sup>** |  |  |  |  |  |  |  |  |
| None | 1.00 | -- | 1.00 | -- | 1.00 | -- | 1.00 | -- |
| Any | **1.43** | **1.20-1.71** | 0.92 | 0.79-1.09 | 1.06 | 0.73-1.54 | **2.16** | **1.74-2.69** |

**Acronyms:** aOR = adjusted odds ratio; DM = diabetes mellitus; HTN = hypertension; HF = heart failure; CAD = coronary artery disease.
Maternal complications, fetal complications, fetal death & stillbirth, and early pregnancy loss defined by ICD codes found in Appendices D, E, F, and G, respectively.
Social Deprivation Index is a composite measure at ZCTA level based on demographics collected by American Community Survey.
Multivariate analyses adjusted for race/ethnicity, CHD anatomic anatomy, age at 1<sup>st</sup> pregnancy, social deprivation index and gestation type, and presence/absence of select comorbid conditions prior to pregnancy.
<sup>^</sup> The pre-existing Maternal Comorbidity Composite is a single value that combined five select common pre-existing individual comorbid conditions that were reported to occur prior to pregnancy: DM, HTN, HF, CAD and arrhythmia. If any of these conditions were present, the composite value = Yes, otherwise the value is No.

### Early Pregnancy Loss

Initial analysis found that early pregnancy loss was significantly more common among nHB women with CHD (11.8%) compared to their nHW counterparts (9.8%), (p=0.02). More socioeconomic deprivation, measured by SDI quintile was also associated with early pregnancy loss compared to least social deprivation (12.5% vs 8.1%, p=0.002). Having any maternal pre-existing comorbidity was associated with 13.4% early pregnancy loss, compared to 9.9% for those without pre-existing comorbid conditions, p=0.0003. Spontaneous abortion was more common among women at the highest (most) socioeconomic deprivation quintile, occurring in 11.1%, versus 7.1% of the least deprived quintile (p=0.04, data not shown). In multivariate adjusted analysis, women with any maternal pre-existing comorbidity had a more than 2-fold risk of early pregnancy loss (aOR 2.16), as did women with twin or higher order pregnancy (aOR 2.03), Table 4.

### Fetal Complications, Fetal Death and Stillbirth

In this analysis, fetal complications and fetal death were also significantly more common in pregnancies to women with severe CHD compared to pregnancies in women with non-severe CHD (53.9% vs. 50.2%, p=0.0113 and 4.4% vs. 3.0%, p=0.0073 respectively). Fetal complications remained significant for women with severe CHD in multivariate analysis (aOR 1.25). Fetal complications (78.6% vs 50.0%, p<0.0001), and fetal death (11.5% vs 3.0%, p<0.0001) were all more common in multi-gestational pregnancies than in singleton pregnancies. Fetal distress, fetal growth restriction, preterm delivery, excess fetal growth, and placental complications affecting the newborn were all significantly more common in women with severe CHD compared to non-severe CHD (**Table 3**). Preterm delivery was most common among pregnancies to women with severe CHD (23.1% pre-term delivery) compared to women with non-severe CHD (19.6%, p < 0.0001). Fetal distress was the most common fetal complication, occurring in 24.3% of pregnancies. Fetal distress was more common in nHB women whereas excess fetal growth was more common in nHW women (**Table 3**).

In the adjusted analysis (**Table 4**), fetal complications were significantly more common in pregnancies to women with severe CHD vs. non-severe CHD (aOR 1.25, 95% CI 1.07-1.47). Pregnancies to women of advanced maternal age (35-50 years) were found to have nearly 2-fold higher odds of fetal death/stillbirth (aOR 1.93, 95% CI 1.24-3.05) compared to younger age groups. The adjusted analysis also revealed a relationship between geographic region and fetal death, with fetal death being more common among pregnancies in rural areas than those in urban areas (aOR 1.51, 95% CI 1.02-2.22).

### Maternal Pregnancy related Complications

Maternal complications were more common in multi-gestational pregnancies than in singleton pregnancies (73.7% vs 59.5%, p<0.0001). No significant differences in maternal complications were noted for CHD severity, rurality, race/ethnicity, or social deprivation (**Table 2**). The proportion of each of the adverse maternal complications among pregnancies in women with CHD are shown in **Table 3**. Anemia in pregnancy (21.5%) and threatened preterm labor (19.2%) were the most common maternal pregnancy related complications. Poly- or oligo-hydramnios more often complicated pregnancies of women with severe CHD compared to non-severe CHD (18.5 vs 14.5%, p = 0.004) whereas other maternal complications were not significantly different between severe and non-severe CHD.

As shown in **Table 4**, having any of the comorbidities included in the composite comorbidity variable was associated with more maternal complications (aOR 1.43, 95% CI 1.20-1.71) and more early pregnancy loss (aOR 2.16, 95% CI 1.74-2.69). Pregnancies to women who had their first documented pregnancy at a young age (11-19) were found to have lower odds of maternal complications (aOR 0.64, 95% CI 0.54-0.76).

### Pre-existing comorbid conditions

Non-gestational comorbid conditions prior to pregnancy are noted in **Table 5**. Comorbid conditions prior to pregnancy were similar between severe CHD and non-severe CHD groups with the exception of residual structural heart disease which was more common in non-severe CHD (12.8% vs 7.4%, p < 0.0001). Diabetes mellitus, hypertension, heart failure, and obesity were more common in nHB women compared to nHW women prior to pregnancy.

## DISCUSSION

In a cohort of women with CHD who experienced pregnancy during 2008-2019 and were covered by Georgia Medicaid, approximately 60% of pregnancies were associated with at least one maternal complication and approximately 50% had at least one fetal complication. The proportion with severe CHD anatomy in our cohort was higher than predicted for adult US women overall which may be associated with a higher overall maternal and fetal complication rate. We found differences in fetal complications by maternal CHD severity, but did not find significant differences in maternal outcomes by CHD severity among women included in the dataset. The non-severe CHD population had more residual structural heart disease than the severe CHD group, but other pre-existing comorbidities were not significantly different between severe and non-severe CHD.

Our study adds to previous literature by using a population-based dataset with a large number of pregnancies to women with CHD to examine contributors to pregnancy-related outcomes among women with similar insurance coverage. These high maternal and fetal complication rates are consistent with existing literature showing increased rates of adverse maternal outcomes in patients with CHD compared to those without CHD.^6,8,25,26^ A 2022 review found significantly higher rates of cesarian deliveries, miscarriages/stillbirths, and preterm labor among pregnant women with CHD compared to the general obstetric population.^5^ Studies have also reported elevated fetal and neonatal complication rates among pregnancies to women with CHD, most commonly related to inherited CHD, prematurity, and small for gestational age (SGA) neonates.^5^ Severity of CHD was also associated with adverse outcomes, as pregnancies to women with severe CHD were more likely than those with non-severe CHD to have fetal complications. Severe CHD may limit cardiac output augmentation and thus placental perfusion later in pregnancy, contributing to a higher rate of fetal complications including fetal death.

Maternal complications were also high among the entire cohort with no significant differences between severe and non-severe CHD. The high maternal complications among women with non-severe CHD in our cohort may reflect a more complicated CHD population due to residual structural heart disease or other complications. The high prevalence of anemia and hemorrhage may be due to a number of factors including an indication for anticoagulation or antiplatelet therapy for prosthetic valves, arrythmias or stents, or to coagulopathy from intrinsic liver disease^.27^ Our cohort also had higher than average rates of hypertensive disorders of pregnancy, including gestational hypertension, preeclampsia, eclampsia, and Hemolysis, Elevated Liver enzymes and Low Platelets (HELLP) syndrome (**Table 3**). A cross-sectional analysis found that in 2019, 7.57% of all pregnancies nationally were impacted by a hypertensive disorder of pregnancy.^28^ In our cohort, 11.3% of pregnancies were complicated by preeclampsia/toxemia, eclampsia, or HELLP, and another 6.4% was complicated by gestational hypertension. A maladaptive vascular response of varied etiologies can lead to preeclampsia in pregnancy.^29^ Impaired placental perfusion related to limited cardiac output augmentation in pregnancy may contribute to some of the excess cases of preeclampsia compared to the general population.^29^

Of note, 20.5% of pregnancies in our cohort ended in preterm delivery, compared to a national preterm delivery rate of 10.23% in 2019.^30^ This is consistent with prior studies that have shown increased rates of premature birth in pregnancies to women with congenital heart disease.^26^

The high rate of preterm delivery in the CHD population is likely multifactorial. Maternal complications related to underlying cardiovascular disease may necessitate preterm delivery, and the high rate of fetal complications may similarly necessitate preterm delivery in the CHD population.

Geographic area (rurality) was significantly related to fetal death. Possible explanations include distance from birthing centers or lack of access to higher levels of care in the event of pregnancy-related complications. Compared to the state of Georgia, our CHD population was more likely to reside in an urban area (77.4% versus 60.3% for the State of Georgia) and had a higher proportion of severe CHD (27.6%) than predicted for an adult CHD population (predicted 12% of adults with CHD in US have severe CHD).^21^ In the adjusted analysis, race/ethnicity was not associated with adverse maternal or fetal outcomes, which differs from existing literature on pregnancy. Possible explanations for these findings include uniform coverage by Georgia Medicaid among the women in our cohort during their pregnancy, as well as characteristics of the overall cohort. The proportion of women who identified as Black (50.1%) was higher than the proportion of women in the State of Georgia identified as Black by census data (33.2%).^33^ These results suggest the type of insurance coverage women have during pregnancy may play a role in disparities in pregnancy-related outcomes.

Our findings highlight the importance of severe maternal CHD in fetal outcomes, and the high-risk nature for both mother and fetus during pregnancy. Further study is warranted in a larger cohort with commercial or other insurance coverage during pregnancy to understand disparities in outcomes.

### Limitations

All patients in this cohort were covered by Georgia Medicaid, limiting applicability of findings to a similar dataset. Our findings cannot be extrapolated to other states or to individuals with commercial insurance or those who are uninsured. Administrative data limits accurate analysis of physiologic staging. It is possible the non-severe CHD group covered by Georgia Medicaid may have had a more complicated disease course than those with non-severe CHD covered by commercial insurance, which is commonly employer-based in the US, and thus not represented in our dataset. Some of the specified outcomes may also be limited by documentation in an administrative dataset. Studying a larger population may be able to mitigate the effect of this limitation. Future studies should include broader populations with greater geographic diversity, beyond the state of Georgia.

## CONCLUSION

Among women covered by Georgia Medicaid during their pregnancy, pregnancies to women with severe CHD have a higher rate of fetal complications and fetal demise than pregnancies to women with non-severe CHD. Future studies should examine fetal outcomes by specific CHD diagnosis and further evaluate the reasons for adverse fetal outcomes.

## Data Availability

The dataset contains personal identifiers and is not able to be shared

## AUTHOR CONTRIBUTIONS

Shayli Patel: Formulation of research question, writing and editing of manuscript.

Cheryl Raskind-Hood: Statistical analysis, formulation of research question, writing and editing of manuscript.

Travis Wilson: Editing of manuscript

Fred H. Rodriguez III: Formulation of research question, editing of manuscript

Alex Haffner: Data management, analysis

Lindsey Ivey: Editing of manuscript.

Vijaya Kancherla: Formulation of research question

Wendy Book: Funding acquisition, formulation of research question, assistance with analysis, writing and editing of manuscript

## Notes

### Competing Interest Statement

The authors have declared no competing interest.

### Funding Statement

Funding: Centers for Disease Control and Prevention National Center on Birth Defects and Developmental Disabilities, DD19-1902.

### Author Declarations

The study was approved by the Emory University Institutional Review Board on July 10, 2023 (IRB # STUDY00006154) which included a complete waiver of HIPAA authorization and informed consent.

## REFERENCES

1. Niwa K. Adult Congenital Heart Disease with Pregnancy. Korean Circ J. 2018;48:251–276. doi: 10.4070/kcj.2018.0070

2. Raskind-Hood C, Saraf A, Riehle-Colarusso T, Glidewell J, Gurvitz M, Dunn JE, Lui GK, Van Zutphen A, McGarry C, Hogue CJ, et al. Assessing Pregnancy, Gestational Complications, and Co-morbidities in Women With Congenital Heart Defects (Data from ICD-9-CM Codes in 3 US Surveillance Sites). Am J Cardiol. 2020;125:812–819. doi: 10.1016/j.amjcard.2019.12.001

3. Cutshall A, Gourdine A, Bender W, Karuppiah A. Trends in outcomes of pregnancy in patients with congenital heart disease. Curr Opin Anaesthesiol. 2023;36:35–41. doi: 10.1097/ACO.0000000000001208

4. Davis MB, Arendt K, Bello NA, Brown H, Briller J, Epps K, Hollier L, Langen E, Park K, Walsh MN. Team-based care of women with cardiovascular disease from pre-conception through pregnancy and postpartum: JACC focus seminar 1/5. Journal of the American College of Cardiology. 2021;77:1763–1777.

5. Kearney K, Zentner D, Cordina R. Management of Maternal Complex Congenital Heart Disease During Pregnancy. Curr Heart Fail Rep. 2021;18:353–361. doi: 10.1007/s11897-021-00534-x

6. Venkataramani R, Lewis AE, Santos JI, Dhondu HS, Ramakrishna H. Maternal and Fetal Outcomes in Adult Congenital Heart Disease. J Cardiothorac Vasc Anesth. 2022;36:3676–3684. doi: 10.1053/j.jvca.2022.04.040

7. Pieper PG, Balci A, Aarnoudse JG, Kampman MA, Sollie KM, Groen H, Mulder BJ, Oudijk MA, Roos-Hesselink JW, Cornette J, et al. Uteroplacental blood flow, cardiac function, and pregnancy outcome in women with congenital heart disease. Circulation. 2013;128:2478–2487. doi: 10.1161/CIRCULATIONAHA.113.002810

8. Steiner JM, Lokken E, Bayley E, Pechan J, Curtin A, Buber J, Albright C. Cardiac and Pregnancy Outcomes of Pregnant Patients With Congenital Heart Disease According to Risk Classification System. Am J Cardiol. 2021;161:95–101. doi: 10.1016/j.amjcard.2021.08.037

9. Hopkins FW, MacKay AP, Koonin LM, Berg CJ, Irwin M, Atrash HK. Pregnancy-related mortality in Hispanic women in the United States. Obstet Gynecol. 1999;94:747–752. doi: 10.1016/s0029-7844(99)00393-2

10. Howell EA. Reducing Disparities in Severe Maternal Morbidity and Mortality. Clin Obstet Gynecol. 2018;61:387–399. doi: 10.1097/GRF.0000000000000349

11. Shah LM, Varma B, Nasir K, Walsh MN, Blumenthal RS, Mehta LS, Sharma G. Reducing disparities in adverse pregnancy outcomes in the United States. Am Heart J. 2021;242:92–102. doi: 10.1016/j.ahj.2021.08.019

12. Quenby S, Gallos ID, Dhillon-Smith RK, Podesek M, Stephenson MD, Fisher J, Brosens JJ, Brewin J, Ramhorst R, Lucas ES, et al. Miscarriage matters: the epidemiological, physical, psychological, and economic costs of early pregnancy loss. Lancet. 2021;397:1658–1667. doi: 10.1016/S0140-6736(21)00682-6

13. Lu MC, Kotelchuck M, Hogan V, Jones L, Wright K, Halfon N. Closing the Black-White gap in birth outcomes: a life-course approach. Ethn Dis. 2010;20:S2–62-76.

14. Rice WS, Goldfarb SS, Brisendine AE, Burrows S, Wingate MS. Disparities in Infant Mortality by Race Among Hispanic and Non-Hispanic Infants. Matern Child Health J. 2017;21:1581–1588. doi: 10.1007/s10995-017-2290-3

15. Womack LS, Rossen LM, Hirai AH. Urban–rural infant mortality disparities by race and ethnicity and cause of death. American journal of preventive medicine. 2020;58:254–260. doi: 10.1016/j.amepre.2019.09.010

16. Kozhimannil KB, Interrante JD, Henning-Smith C, Admon LK. Rural-Urban Differences In Severe Maternal Morbidity And Mortality In The US, 2007-15. Health Aff (Millwood). 2019;38:2077–2085. doi: 10.1377/hlthaff.2019.00805

17. Anglim AJ, Radke SM. Rural Maternal Health Care Outcomes, Drivers, and Patient Perspectives. Clin Obstet Gynecol. 2022;65:788–800. doi: 10.1097/GRF.0000000000000753

18. Ivey LC, Rodriguez III FH, Shi H, Chong C, Chen J, Raskind-Hood CL, Downing KF, Farr SL, Book WM. Positive Predictive Value of International Classification of Diseases, Ninth Revision, Clinical Modification, and International Classification of Diseases, Tenth Revision, Clinical Modification, Codes for Identification of Congenital Heart Defects. Journal of the American Heart Association. 2023;12:e030821. doi: 10.1161/JAHA.123.030821

19. Ailes EC, Simeone RM, Dawson AL, Petersen EE, Gilboa SM. Using insurance claims data to identify and estimate critical periods in pregnancy: An application to antidepressants. Birth Defects Research Part A: Clinical and Molecular Teratology. 2016;106:927–934. doi: 10.1002/bdra.23573

20. Ailes EC, Zhu W, Clark EA, Huang YA, Lampe MA, Kourtis AP, Reefhuis J, Hoover KW. Identification of pregnancies and their outcomes in healthcare claims data, 2008-2019: An algorithm. PLoS One. 2023;18:e0284893. doi: 10.1371/journal.pone.0284893

21. Social deprivation index (SDI). Robert Graham Center - Policy Studies in Family Medicine & Primary Care. (2018, November 5). Retrieved November 29, 2021, from https://www.graham-center.org/rgc/maps-data-tools/sdi/social-deprivation-index.html.

22. Kirby A, Curtis E, Hlohovsky S, Brown A, O’Donnell C. Pregnancy outcomes and risk evaluation in a contemporary adult congenital heart disease cohort. Heart, Lung and Circulation. 2021;30:1364–1372. doi: 10.1016/j.hlc.2021.03.005

23. Phillips RL, Liaw W, Crampton P, Exeter DJ, Bazemore A, Vickery KD, Petterson S, Carrozza M. How other countries use deprivation indices—and why the United States desperately needs one. Health Affairs. 2016;35:1991–1998. doi: 10.1377/hlthaff.2016.0709

24. Butler DC, Petterson S, Phillips RL, Bazemore AW. Measures of social deprivation that predict health care access and need within a rational area of primary care service delivery. Health Serv Res. 2013;48:539–559. doi: 10.1111/j.1475-6773.2012.01449.x

25. Hoyert D. Maternal Mortality Rates in the United States, 2021. NCHS Health E-Stats. Centers for Disease Control and Prevention (CDC). National Center for Health Statistics. 2023.

26. Ramage K, Grabowska K, Silversides C, Quan H, Metcalfe A. Association of adult congenital heart disease with pregnancy, maternal, and neonatal outcomes. JAMA Network Open. 2019;2:e193667–e193667. doi: 10.1001/jamanetworkopen.2019.3667

27. Majiyagbe OO, Akinsete AM, Adeyemo TA, Salako AO, Ekure EN, Okoromah CA. Coagulation abnormalities in children with uncorrected congenital heart defects seen at a teaching hospital in a developing country. PloS one. 2022;17:e0263948. doi: 10.1371/journal.pone.0263948

28. Bruno AM, Allshouse AA, Metz TD, Theilen LH. Trends in hypertensive disorders of pregnancy in the United States from 1989 to 2020. Obstetrics & Gynecology. 2022;140:83–86. doi: 10.1097/AOG.0000000000004824

29. Jung E, Romero R, Yeo L, Gomez-Lopez N, Chaemsaithong P, Jaovisidha A, Gotsch F, Erez O. The etiology of preeclampsia. American journal of obstetrics and gynecology. 2022;226:S844–S866. doi: 10.1016/j.ajog.2021.11.1356.

30. Martin JA, Hamilton BE, Osterman MJ, Driscoll AK, Drake P. Births: final data for 2017. 2018.

31. Rodriguez FH 3rd, Ephrem G, Gerardin JF, Raskind-Hood C, Hogue C, Book W. The 745.5 issue in code-based, adult congenital heart disease population studies: Relevance to current and future ICD-9-CM and ICD-10-CM studies. Congenit Heart Dis. 2018 Jan;13(1):59–64. doi: 10.1111/chd.12563. Epub 2017 Dec 20. PMID: 29266726.

32. Ivey LC, Rodriguez FH 3rd, Shi H, Chong C, Chen J, Raskind-Hood CL, Downing KF, Farr SL, Book WM. Positive Predictive Value of International Classification of Diseases, Ninth Revision, Clinical Modification, and International Classification of Diseases, Tenth Revision, Clinical Modification, Codes for Identification of Congenital Heart Defects. J Am Heart Assoc. 2023 Aug 15;12(16):e030821. doi: 10.1161/JAHA.123.030821. Epub 2023 Aug 7. PMID: 37548168; PMCID: PMC10492959.

33. https://www.census.gov/quickfacts/fact/table/GA/PST045223

