## Supplemental Table S1 for "Maternal and Fetal Complications Among Pregnant Women with Congenital Heart Disease"

| **Supplemental Table S1: Categorization of Included Congenital Heart Defect Codes** | | |
| --- | --- | --- |
| **ICD Diagnosis Code Label** | **ICD-9-CM** | **ICD-10-CM** |
| **Severe *(15 ICD-9-CM codes and 14 ICD-10-CM codes)*** | | |
| Hypoplastic Left Heart Syndrome (HLHS) | 746.7 | Q23.4 |
| Tricuspid Atresia, stenosis or absence | 746.1 | Q22.4 |
| Hypoplastic right heart syndrome |  | Q22.6 |
| Single ventricle, cor trioculare, double inlet left ventricle | 745.3 | Q20.4 |
| Cor bioulare | 745.7 |  |
| Pulmonary valve atresia or absence | 746.01 | Q22.0 |
| Truncus Arteriosus, Common Truncus | 745.0 | Q20.0 |
| Double outlet right ventricle (DORV) | 745.11 | Q20.1 |
| Double outlet Left ventricle (DOLV) |  | Q20.2 |
| Tetralogy of Fallot | 745.2 | Q21.3 |
| Transposition of the Great arteries (TGA), Complete TGA, dextro-TGA, TGA not otherwise specified, classical TGA | 745.1 | Q20.3 |
|  | 745.10 |  |
|  | 745.19 |  |
| Congenital Corrected transposition of the great arteries (CCTGA), levo-TGA | 745.12 | Q20.5 |
| Endocardial cushion defect | 745.6 | Q21.2 |
| Atrioventricular septal defect | 745.60 |  |
| Complete atrioventricular canal defect (CAVCD), Endocardial cushion defect unspec., Endocardial cushion defect other | 745.69 |  |
| Interrupted aortic arch | 747.11 | Q25.21 |
| Total anomalous pulmonary venous return (TAPVR) | 747.41 | Q26.2 |
| **SHUNT AND VALVE *Cases with no severe code AND with both Shunt AND Valve code*** | | |
| **SHUNT *(7 ICD-9-CM codes & 8 ICD-10 codes) Cases with shunt codes with or without Other group codes, but no severe or valve codes*** | | |
| Ventricular septal defect (VSD) | 745.4 | Q21.0 |
| Secundum atrial septal defect (ASD) | 745.5 | Q21.1 |
| Primum atrial septal defect | 745.61 |  |
| Other specified defect of septal closure, sinus venosus ASD, inferior sinus venosus ASD, superior sinus venosus ASD | 745.8 | Q21.8 |
| Congenital malformation of cardiac septum, unspecified |  | Q21.9 |
| Unspecified defect of septal closure | 745.9 |  |
| Patent ductus arteriosus (PDA) | 747.0 | Q25.0 |
| Aortopulmonary septal defect (AP window) |  | Q21.4 |
| Partial anomalous pulmonary venous return (PAPVR) | 747.42 | Q26.3 |
| Anomalous pulmonary venous connection, unspecified |  | Q26.4 |
| **VALVE *(16 ICD-9-CM codes & 22 ICD-10 codes) Case with valve codes w or wo other group codes & has no codes in severe or shunt*** | | |
| Anomalies of the pulmonary valve | 746.0 |  |
| Pulmonary valve anomaly, unspecified | 746.00 | Q22.3 |
| Pulmonary valve stenosis (PS) | 746.02 | Q22.1 |
| Pulmonary valve anomaly, other, pulmonary valve regurgitation | 746.09 | Q22.2 |
| Ebstein anomaly of the tricuspid valve | 746.2 | Q22.5 |
| Congenital malformations of tricuspid valve |  | Q22.8 |
| Congenital malformation of tricuspid valve, unspecified |  | Q22.9 |
| Aortic valve stenosis (AS) | 746.3 | Q23.0 |
| Aortic insufficiency or bicuspid or unicuspid aortic valve | 746.4 | Q23.1 |
| Other congenital malformations of aortic & mitral valves |  | Q23.8 |
| Congenital malformation of aortic & mitral valves, spec |  | Q23.9 |
| Mitral stenosis or mitral valve abnormalities | 746.5 | Q23.2 |
| Mitral insufficiency, cleft mitral valve | 746.6 | Q23.3 |
| Subaortic stenosis, subaortic membrane | 746.81 | Q24.4 |
| Infundibular or subvalvar pulmonary stenosis | 746.83 | Q24.3 |
| Coarctation of the aorta | 747.1 | Q25.1 |
|  | 747.10 |  |
| Supravalvular aortic stenosis |  | Q25.3 |
| Hypoplasia of the aorta |  | Q25.42 |
| Atresia or stenosis of aorta | 747.22 | Q25.29 |
| Absence and aplasia of aorta |  | Q25.41 |
| Atresia of pulmonary artery |  | Q25.5 |
| Pulmonary artery atresia, coarctation, or hypoplasia of the branch pulmonary arteries | 747.31 | Q25.71 |
| Anomalies of the pulmonary artery, other | 747.39 | Q25.79 |
| **OTHER CHD *(7 ICD-9-CM codes and 8 ICD-10-CM codes) Cases with only "other group" codes and no codes in severe shunt or valve groups*** | | |
| Cor triatriatum | 746.82 | Q24.2 |
| Coronary artery anomaly, anomalous coronary artery, anomalous left coronary artery off of the pulmonary artery (ALCAPA), anomalous right coronary artery off of the pulmonary artery (ARCAPA) | 746.85 | Q24.5 |
| **OTHER VASCULAR *(3 ICD-10-CM codes) Cases with other vascular codes required either code Q25.45 or the combination of Q25.47 and Q25.48 to be included*** | | |
| Double aortic arch |  | Q25.45 |
| Right aortic arch component of Vascular ring |  | Q25.47 |
| Anomalous origin of the subclavian artery component of vascular ring |  | Q25.48 |
