## Supplemental Table S2 for "Maternal and Fetal Complications Among Pregnant Women with Congenital Heart Disease"

| **Supplemental Table S2: ICD-9-CM , ICD-10-CM and CPT Codes to Define Pregnancy (3613 codes)** | | |
| --- | --- | --- |
| **TYPE** | **CODE** | **DESCRIPTION** |
| DX_ICD9 | 631.0 | inapp chg hcg early preg (begin 2011) |
| DX_ICD9 | 631 | oth abn prod conception (end 2011) |
| DX_ICD9 | 633.0 | abdom preg (end 2002) |
| DX_ICD9 | 633.00 | abdom preg w/o intrauterine preg (begin 2002) |
| DX_ICD9 | 633.1 | tubal preg (end 2002) |
| DX_ICD9 | 633.10 | tubal preg w/o intrauterine preg (begin 2002) |
| DX_ICD9 | 633.2 | ovarian preg (end 2002) |
| DX_ICD9 | 633.20 | ovarian preg w/o intrauterine preg (begin 2002) |
| DX_ICD9 | 633.8 | ectopic preg nec (end 2002) |
| DX_ICD9 | 633.80 | ot ectopic preg w/o intrau preg (begin 2002) |
| DX_ICD9 | 633.9 | ectopic preg nos (end 2002) |
| DX_ICD9 | 633.90 | unspec ectopic preg w/o intrauterine preg (begin 2002) |
| DX_ICD9 | 641.31 | coag def hem- delivered |
| DX_ICD9 | 641.33 | coag def hem- antepartum |
| DX_ICD9 | 666.02 | third-stage hem- delivered w p/p |
| DX_ICD9 | 666.04 | third-stage hem- postpartum |
| DX_ICD10 | O11.1 | pre-existing htn with pre-eclampsia, first tri |
| DX_ICD10 | O11.2 | pre-existing htn with pre-eclampsia, second tri |
| DX_ICD10 | O11.3 | pre-existing htn with pre-eclampsia, third tri |
| DX_ICD10 | O11.9 | pre-existing htn with pre-eclampsia, unspec tri |
| PX_ICD9 | 66.62 | salpingectomy with removal of tubal preg |
| PX_ICD9 | 69.01 | dilation & curettage for termination of preg |
| PX_ICD9 | 69.51 | aspiration curettage of uterus for termination of preg |
| PX_ICD9 | 72.0 | low forceps operation |
| PX_ICD9 | 72.1 | low forceps operation with episiotomy |
| PX_ICD9 | 72.21 | mid forceps operation with episiotomy |
| PX_ICD9 | 72.29 | oth mid forceps operation |
| PX_ICD9 | 72.31 | high forceps operation with episiotomy |
| PX_ICD9 | 72.39 | oth high forceps operation |
| PX_ICD9 | 72.4 | forceps rotation of fetal head |
| PX_ICD9 | 72.51 | partial breech extraction with forceps to aftercoming head |
| PX_ICD9 | 72.52 | oth partial breech extraction |
| PX_ICD9 | 72.53 | total breech extraction with forceps to aftercoming head |
| PX_ICD9 | 72.6 | forceps application to aftercoming head |
| PX_ICD9 | 72.71 | vacuum extraction with episiotomy |
| PX_ICD9 | 72.79 | oth vacuum extraction |
| PX_ICD9 | 72.8 | oth specif instrumental delivery |
| PX_ICD9 | 72.9 | unspec instrumental delivery |
| PX_ICD9 | 73.01 | induction of labor by artificial rupt of membranes |
| PX_ICD9 | 73.09 | oth artificial rupt of membranes |
| PX_ICD9 | 73.1 | oth surgical induction of labor |
| PX_ICD9 | 73.21 | internal & combined version wo extraction |
| PX_ICD9 | 73.22 | internal & combined version with extraction |
| PX_ICD9 | 73.3 | failed forceps |
| PX_ICD9 | 73.4 | medical induction of labor |
| PX_ICD9 | 73.51 | manual rotation of fetal head |
| PX_ICD9 | 73.59 | oth total breech extraction |
| PX_ICD9 | 73.6 | episiotomy |
| PX_ICD9 | 73.8 | operations on fetus to facilitate delivery |
| PX_ICD9 | 73.91 | external version assisting delivery |
| PX_ICD9 | 73.92 | replacement of prolapsed umbilical cord |
| PX_ICD9 | 73.93 | incision of cervix to assist delivery |
| PX_ICD9 | 73.94 | pubiotomy to assist delivery |
| PX_ICD9 | 73.99 | oth operations assisting delivery |
| PX_ICD9 | 74.0 | classical cesarean section |
| PX_ICD9 | 74.1 | low cervical cesarean section |
| PX_ICD9 | 74.2 | extraperitoneal cesarean section |
| PX_ICD9 | 74.3 | removal of extratubal ectopic preg |
| PX_ICD9 | 74.4 | cesarean section of oth specif type |
| PX_ICD9 | 74.91 | hysterotomy to terminate preg |
| PX_ICD9 | 74.99 | oth cesarean section of unspec type |
| PX_ICD9 | 75.0 | intra-amniotic injection for abortion |
| DX_ICD9 | 268.2 | osteomalacia, unspec |
| DX_ICD9 | 293.89 | oth spec transient mental disorders due to conds, oth |
| DX_ICD9 | 632 | missed abortion |
| DX_ICD9 | 633.01 | abdom preg w intrauterine preg (begin 2002) |
| DX_ICD9 | 633.11 | tubal preg w intrauterine preg (begin 2002) |
| DX_ICD9 | 633.21 | ovarian preg w intrauterine preg (begin 2002) |
| DX_ICD9 | 633.81 | oth ectopic preg w intrauterine preg (begin 2002) |
| DX_ICD9 | 633.91 | unspec ectopic preg w intrauterine preg (begin 2002) |
| DX_ICD9 | 634.00 | SAB w pelvic infection - unspec |
| DX_ICD9 | 634.01 | SAB w pelvic infection- incomplete |
| DX_ICD9 | 634.02 | SAB w pelvic infection- complete |
| DX_ICD9 | 634.10 | SAB w hem- unspec |
| DX_ICD9 | 634.11 | SAB w hem- incomplete |
| DX_ICD9 | 634.12 | SAB w hem- complete |
| DX_ICD9 | 634.20 | SAB w pelvic dam- unspec |
| DX_ICD9 | 634.21 | SAB w pelvic dam- incomplete |
| DX_ICD9 | 634.22 | SAB w pelvic dam- complete |
| DX_ICD9 | 634.30 | SAB w renal failure- unspec |
| DX_ICD9 | 634.31 | SAB w renal failure- incomplete |
| DX_ICD9 | 634.32 | SAB w renal failure- complete |
| DX_ICD9 | 634.40 | SAB w metabolic disease- unspec |
| DX_ICD9 | 634.41 | SAB w metabolic disease- incomplete |
| DX_ICD9 | 634.42 | SAB w metabolic disease |
| DX_ICD9 | 634.50 | SAB w shock- unspec |
| DX_ICD9 | 634.51 | SAB w shock- incomplete |
| DX_ICD9 | 634.52 | SAB w shock- complete |
| DX_ICD9 | 634.60 | SAB w embol- unspec |
| DX_ICD9 | 634.61 | SAB w embol- incomplete |
| DX_ICD9 | 634.62 | SAB w embol- complete |
| DX_ICD9 | 634.70 | SAB w compl nec- unspec |
| DX_ICD9 | 634.71 | SAB, with oth spec complic, incomplete |
| DX_ICD9 | 634.72 | SAB w compl nec- complete |
| DX_ICD9 | 634.80 | SAB w compl nos- unspec |
| DX_ICD9 | 634.81 | SAB w compl nos- incomplete |
| DX_ICD9 | 634.82 | SAB w compl nos- complete |
| DX_ICD9 | 634.90 | SAB uncompl- unspec |
| DX_ICD9 | 634.91 | SAB uncompl- incomplete |
| DX_ICD9 | 634.92 | SAB uncompl- complete |
| DX_ICD9 | 635.00 | legal abortion w pelvic infection- unspec |
| DX_ICD9 | 635.01 | legal abortion w pelvic infection- incomplete |
| DX_ICD9 | 635.02 | legal abortion w pelvic infection- complete |
| DX_ICD9 | 635.10 | legal abortion w hem- unspec |
| DX_ICD9 | 635.11 | legal abortion w hem- incomplete |
| DX_ICD9 | 635.12 | legal abortion w hem- complete |
| DX_ICD9 | 635.20 | legal abortion w pelvic dam- unspec |
| DX_ICD9 | 635.21 | legal abortion w pelvic dam- incomplete |
| DX_ICD9 | 635.22 | legal abortion w pelvic dam- complete |
| DX_ICD9 | 635.30 | legal abortion w renal failure- unspec |
| DX_ICD9 | 635.31 | legal abortion w renal failure- incomplete |
| DX_ICD9 | 635.32 | legal abortion w renal failure- complete |
| DX_ICD9 | 635.40 | legal abortion w metabolic disease- unspec |
| DX_ICD9 | 635.41 | legal abortion w metabolic disease- incomplete |
| DX_ICD9 | 635.42 | legal abortion w metabolic disease- complete |
| DX_ICD9 | 635.50 | legal abortion w shock- unspec |
| DX_ICD9 | 635.51 | legal abortion w shock- incomplete |
| DX_ICD9 | 635.52 | legal abortion w shock- complete |
| DX_ICD9 | 635.60 | legal abortion w embolism- unspec |
| DX_ICD9 | 635.61 | legal abortion w embolism- incomplete |
| DX_ICD9 | 635.62 | legal abortion w embolism- complete |
| DX_ICD9 | 635.70 | legal abortion w compl nec- unspec |
| DX_ICD9 | 635.71 | legal abortion w compl nec- incomplete |
| DX_ICD9 | 635.72 | legal abortion w compl nec- complete |
| DX_ICD9 | 635.80 | legal abortion w compl nos- unspec |
| DX_ICD9 | 635.81 | legal abortion w compl nos- incomplete |
| DX_ICD9 | 635.82 | legal abortion w compl nos- complete |
| DX_ICD9 | 635.90 | legal abortion uncompl- unspec |
| DX_ICD9 | 635.91 | legal abortion uncompl- incomplete |
| DX_ICD9 | 635.92 | legal abortion uncompl- complete |
| DX_ICD9 | 636.00 | illegal abortion w pelvic infection- unspec |
| DX_ICD9 | 636.01 | illegal abortion w pelvic infection- incomplete |
| DX_ICD9 | 636.02 | illegal abortion w pelvic infection- complete |
| DX_ICD9 | 636.10 | illegal abortion w hem- unspec |
| DX_ICD9 | 636.11 | illegal abortion w hem- incomplete |
| DX_ICD9 | 636.12 | illegal abortion w hem- complete |
| DX_ICD9 | 636.20 | illegal abortion w pelvic dam- unspec |
| DX_ICD9 | 636.21 | illegal abortion w pelvic dam- incomplete |
| DX_ICD9 | 636.22 | illegal abortion w pelvic dam- complete |
| DX_ICD9 | 636.30 | illegal abortion w renal failure- unspec |
| DX_ICD9 | 636.31 | illegal abortion w renal failure- incomplete |
| DX_ICD9 | 636.32 | illegal abortion w renal failure- complete |
| DX_ICD9 | 636.40 | illegal abortion w metabolic disease- unspec |
| DX_ICD9 | 636.41 | illegal abortion w metabolic disease- incomplete |
| DX_ICD9 | 636.42 | illegal abortion w metabolic disease- complete |
| DX_ICD9 | 636.50 | illegal abortion w shock- unspec |
| DX_ICD9 | 636.51 | illegal abortion w shock- incomplete |
| DX_ICD9 | 636.52 | illegal abortion w shock- complete |
| DX_ICD9 | 636.60 | illegal abortion w embolism- unspec |
| DX_ICD9 | 636.61 | illegal abortion w embolism- incomplete |
| DX_ICD9 | 636.62 | illegal abortion w embolism- complete |
| DX_ICD9 | 636.70 | illegal abortion w compl nec- unspec |
| DX_ICD9 | 636.71 | illegal abortion w compl nec- incomplete |
| DX_ICD9 | 636.72 | illegal abortion w compl nec- complete |
| DX_ICD9 | 636.80 | illegal abortion w compl nos- unspec |
| DX_ICD9 | 636.81 | illegal abortion w compl nos- incomplete |
| DX_ICD9 | 636.82 | illegal abortion w compl nos- complete |
| DX_ICD9 | 636.90 | illegal abortion uncompl- unspec |
| DX_ICD9 | 636.91 | illegal abortion uncompl- incomplete |
| DX_ICD9 | 636.92 | illegal abortion uncompl complete |
| DX_ICD9 | 637.00 | abortion nos w pelvic infection- unspec |
| DX_ICD9 | 637.01 | abortion nos w pelvic infection- incomplete |
| DX_ICD9 | 637.02 | abortion nos w pelvic infection- complete |
| DX_ICD9 | 637.10 | abortion nos w hem- unspec |
| DX_ICD9 | 637.11 | abortion nos w hem- incomplete |
| DX_ICD9 | 637.12 | abortion nos w hem- complete |
| DX_ICD9 | 637.20 | abortion nos w pelvic dam- unspec |
| DX_ICD9 | 637.21 | abortion nos w pelvic dam- incomplete |
| DX_ICD9 | 637.22 | abortion nos w pelvic dam- complete |
| DX_ICD9 | 637.30 | abortion nos w renal failure- unspec |
| DX_ICD9 | 637.31 | abortion nos w renal failure- incomplete |
| DX_ICD9 | 637.32 | abortion nos w renal failure- complete |
| DX_ICD9 | 637.40 | abortion nos w metabolic disease- unspec |
| DX_ICD9 | 637.41 | abortion nos w metabolic disease- incomplete |
| DX_ICD9 | 637.42 | abortion nos w metabolic disease- complete |
| DX_ICD9 | 637.50 | abortion nos w shock- unspec |
| DX_ICD9 | 637.51 | abortion nos w shock- incomplete |
| DX_ICD9 | 637.52 | abortion nos w shock- complete |
| DX_ICD9 | 637.60 | abortion nos w embolism- unspec |
| DX_ICD9 | 637.61 | abortion nos w embolism- incomplete |
| DX_ICD9 | 637.62 | abortion nos w embolism- complete |
| DX_ICD9 | 637.70 | abortion nos w compl nec- unspec |
| DX_ICD9 | 637.71 | abortion nos w compl nec- incomplete |
| DX_ICD9 | 637.72 | abortion nos w compl nec- complete |
| DX_ICD9 | 637.80 | abortion nos w compl nos- unspec |
| DX_ICD9 | 637.81 | abortion nos w compl nos- incomplete |
| DX_ICD9 | 637.82 | abortion nos w compl nos- complete |
| DX_ICD9 | 637.90 | ab nos uncomplicat-unsp |
| DX_ICD9 | 637.91 | ab nos uncomplicat-inc |
| DX_ICD9 | 637.92 | ab nos uncomplicat-comp |
| DX_ICD9 | 638.0 | attempted abortion w pelvic infection |
| DX_ICD9 | 638.1 | attempted abortion w hemorrhage |
| DX_ICD9 | 638.2 | attempted abortion w pelvic dam |
| DX_ICD9 | 638.3 | attempted abortion w renal failure |
| DX_ICD9 | 638.4 | attempted abortion w metabolic disease |
| DX_ICD9 | 638.5 | attempted abortion w shock |
| DX_ICD9 | 638.6 | attempted abortion w embolism |
| DX_ICD9 | 638.7 | attempted abortion w compl nec |
| DX_ICD9 | 638.8 | attempted abortion w compl nos |
| DX_ICD9 | 638.9 | attempted abort uncompl |
| DX_ICD9 | 639.0 | post abortion gu infection |
| DX_ICD9 | 639.1 | post abortion hem |
| DX_ICD9 | 639.2 | post abortion pelvic dam |
| DX_ICD9 | 639.3 | post abortion renal failure |
| DX_ICD9 | 639.4 | post abortion metabolic disease |
| DX_ICD9 | 639.6 | post abortion embolism |
| DX_ICD9 | 639.8 | post abortion compl nec |
| DX_ICD9 | 639.9 | post abortion compl nos |
| DX_ICD9 | 640.80 | hem early preg nec- unspec |
| DX_ICD9 | 640.81 | hem early preg nec- delivered |
| DX_ICD9 | 640.83 | hem early preg nec- antepartum |
| DX_ICD9 | 640.90 | hem early preg- unspec |
| DX_ICD9 | 640.91 | hem early preg- delivered |
| DX_ICD9 | 640.93 | hem early preg- antepartum |
| DX_ICD9 | 641.00 | plcnta previa- unspec |
| DX_ICD9 | 641.01 | plcnta previa- delivered |
| DX_ICD9 | 641.03 | plcnta previa- antepartum |
| DX_ICD9 | 641.10 | plcnta previa hem- unspec |
| DX_ICD9 | 641.11 | plcnta previa hem- delivered |
| DX_ICD9 | 641.13 | plcnta previa hem- antepartum |
| DX_ICD9 | 641.20 | prem separation placenta- unspec |
| DX_ICD9 | 641.21 | prem separation placenta- delivered |
| DX_ICD9 | 641.23 | prem separation placenta- antepartum |
| DX_ICD9 | 641.30 | coag def hem- unspec |
| DX_ICD9 | 641.80 | antepartum hem nec- unspec |
| DX_ICD9 | 641.81 | antepartum hem nec- delivered |
| DX_ICD9 | 641.83 | antepartum hem nec- antepartum |
| DX_ICD9 | 641.90 | antepartum hem nos- unspec |
| DX_ICD9 | 641.91 | antepartum hem nos- delivered |
| DX_ICD9 | 641.93 | antepartum hem nos- antepartum |
| DX_ICD9 | 642.03 | essential hypertension- antepartum |
| DX_ICD9 | 642.13 | renal htn antepartum |
| DX_ICD9 | 642.23 | old htn nec- antepartum |
| DX_ICD9 | 642.33 | trans hypertension- antepartum |
| DX_ICD9 | 642.40 | mild/nos preeclampsia- unspec |
| DX_ICD9 | 642.41 | mild/nos preeclampsia- delivered |
| DX_ICD9 | 642.42 | mild preeclampsia- delivered w p/p |
| DX_ICD9 | 642.43 | mild/nos preeclampsia- antepartum |
| DX_ICD9 | 642.44 | mild/nos preeclampsia- p/p |
| DX_ICD9 | 642.50 | severe preeclampsia- unspec |
| DX_ICD9 | 642.51 | severe preeclampsia- delivered |
| DX_ICD9 | 642.52 | severe preeclampsia- delivered w p/p |
| DX_ICD9 | 642.53 | severe preeclampsia- antepartum |
| DX_ICD9 | 642.54 | severe preeclampsia- postpartum |
| DX_ICD9 | 642.60 | eclampsia- unspec |
| DX_ICD9 | 642.61 | eclampsia- delivered |
| DX_ICD9 | 642.62 | eclampsia- delivered w p/p |
| DX_ICD9 | 642.63 | eclampsia- antepartum |
| DX_ICD9 | 642.64 | eclampsia- postpartum |
| DX_ICD9 | 642.70 | toxemia w old hypertension- unspec |
| DX_ICD9 | 642.71 | toxemia w old hypertension- delivered |
| DX_ICD9 | 642.72 | toxemia w old hypertension- delivered w p/p |
| DX_ICD9 | 642.73 | toxemia w old hypertension- antepartum |
| DX_ICD9 | 642.74 | toxemia w old hypertension- postpartum |
| DX_ICD9 | 642.90 | htn in preg nos- unspec |
| DX_ICD9 | 642.93 | htn not othwise specif- antepartum |
| DX_ICD9 | 643.00 | mild hyperem grav-unspec |
| DX_ICD9 | 643.01 | mild hyperem grav-unspec |
| DX_ICD9 | 643.03 | mild hyperem grav-unspec |
| DX_ICD9 | 643.10 | hyperem w metab dis-unsp |
| DX_ICD9 | 643.11 | hyperem w metab dis-unsp |
| DX_ICD9 | 643.13 | hyperem w metab dis-unsp |
| DX_ICD9 | 643.83 | vomit compl preg-antepar |
| DX_ICD9 | 643.93 | vomit of pg nos-antepart |
| DX_ICD9 | 644.00 | threat prem labor- unspec |
| DX_ICD9 | 644.03 | threat prem labor- antepartum |
| DX_ICD9 | 644.10 | threat labor nec- unspec |
| DX_ICD9 | 644.13 | threat labor nec- antepartum |
| DX_ICD9 | 644.20 | early onset delivery unspec |
| DX_ICD9 | 644.21 | early onset delivery- delivered |
| DX_ICD9 | 645.00 | prolonged preg- unspec (begin 1991) |
| DX_ICD9 | 645.01 | prolonged preg- delivered (begin 1991) |
| DX_ICD9 | 645.03 | prolonged preg- antepartum (begin 1991) |
| DX_ICD9 | 645.10 | post term preg- unspec (begin 2000) |
| DX_ICD9 | 645.11 | post term preg- delivered (begin 2000) |
| DX_ICD9 | 645.13 | post term preg- antepartum (begin 2000) |
| DX_ICD9 | 645.20 | prolonged preg- unspec or begin 2000) |
| DX_ICD9 | 645.21 | prolonged preg- del w or w/o antepartum (begin 2000) |
| DX_ICD9 | 645.23 | prolonged preg- antepartum (begin 2000) |
| DX_ICD9 | 646.00 | papyraceous fetus- unspec |
| DX_ICD9 | 646.01 | papyraceous fetus- delivered |
| DX_ICD9 | 646.03 | papyraceous fetus- antepartum |
| DX_ICD9 | 646.10 | edema in preg-unspec |
| DX_ICD9 | 646.12 | edema in preg-del w p/p |
| DX_ICD9 | 646.13 | edema in preg-antepartum |
| DX_ICD9 | 646.14 | edema in preg-postpartum |
| DX_ICD9 | 646.20 | renal dis preg nos-unsp |
| DX_ICD9 | 646.23 | renal dis nos-antepartum |
| DX_ICD9 | 646.33 | habitual abort-antepart |
| DX_ICD9 | 646.43 | neuritis of preg-antepar |
| DX_ICD9 | 646.53 | asy bacteriuria-antepart |
| DX_ICD9 | 646.63 | gu infection-antepartum |
| DX_ICD9 | 646.70 | liver dis in preg-unspec |
| DX_ICD9 | 646.83 | preg compl nec-antepart |
| DX_ICD9 | 646.93 | preg compl nos-antepart |
| DX_ICD9 | 647.00 | syphilis in preg-unspec |
| DX_ICD9 | 647.03 | syphilis-antepartum |
| DX_ICD9 | 647.13 | gonorrhea-antepartum |
| DX_ICD9 | 647.23 | oth vd-antepartum |
| DX_ICD9 | 647.33 | tuberculosis-antepartum |
| DX_ICD9 | 647.43 | malaria-antepartum |
| DX_ICD9 | 647.53 | rubella-antepartum |
| DX_ICD9 | 647.63 | oth viral dis-antepartum |
| DX_ICD9 | 647.83 | infect dis nec-antepart |
| DX_ICD9 | 647.93 | infect nos-antepartum |
| DX_ICD9 | 648.03 | diabetes- antepartum |
| DX_ICD9 | 648.13 | thyroid dysfunc-antepart |
| DX_ICD9 | 648.23 | anemia- antepartum |
| DX_ICD9 | 648.42 | mental disorders of moth |
| DX_ICD9 | 648.53 | congen cv dis-antepartum |
| DX_ICD9 | 648.63 | cv dis nec-antepartum |
| DX_ICD9 | 648.73 | bone disorder-antepartum |
| DX_ICD9 | 648.83 | abnormal glucose- antepartum |
| DX_ICD9 | 648.93 | oth curr cond-antepartum |
| DX_ICD9 | 649.03 | tobacco use dis-antepart (begin 2006) |
| DX_ICD9 | 649.13 | obesity-antepartum (begin 2006) |
| DX_ICD9 | 649.14 | obesity-postpartum (begin 2006) |
| DX_ICD9 | 649.23 | bariatrc surg stat-antep (begin 2006) |
| DX_ICD9 | 649.33 | coagulation def-antepart (begin 2006) |
| DX_ICD9 | 649.43 | epilepsy-antepartum (begin 2006) |
| DX_ICD9 | 649.53 | spotting-antepartum (begin 2006) |
| DX_ICD9 | 649.63 | uterine size des-antepar (begin 2006) |
| DX_ICD9 | 649.73 | cervical shortening-ante (begin 2008) |
| DX_ICD9 | 650 | normal delivery |
| DX_ICD9 | 651.03 | twin preg-antepart |
| DX_ICD9 | 651.13 | trip preg-antepartum |
| DX_ICD9 | 651.23 | quad preg-antepart |
| DX_ICD9 | 651.33 | twins w fetal loss- antepartum (begin 1989) |
| DX_ICD9 | 651.43 | triplets w fetal loss- antepartum (begin 1989) |
| DX_ICD9 | 651.53 | quads w fetal loss- antepartum (begin 1989) |
| DX_ICD9 | 651.63 | multi gest w fetal loss- antepartum (begin 1989) |
| DX_ICD9 | 651.73 | multi gest- fetal reduction- antepartum (begin 2005) |
| DX_ICD9 | 651.83 | multi gest- fetal reduction- antepartum (begin 2005) |
| DX_ICD9 | 652.03 | unstable lie- antepartum |
| DX_ICD9 | 652.13 | cephalic version nos- antepartum |
| DX_ICD9 | 652.23 | breech present- antepartum |
| DX_ICD9 | 652.33 | transverse/ oblique lie- antepartum |
| DX_ICD9 | 652.43 | face/brow present- antepartum |
| DX_ICD9 | 652.53 | high head at term- antepartum |
| DX_ICD9 | 652.63 | multi gest malpresentation- antepartum |
| DX_ICD9 | 652.73 | prolapsed arm- antepartum |
| DX_ICD9 | 652.83 | malposition nec- antepartum |
| DX_ICD9 | 652.93 | malposition nos- antepartum |
| DX_ICD9 | 653.03 | pelv deform nos-antepart |
| DX_ICD9 | 653.13 | contrac pelv nos-antepar |
| DX_ICD9 | 653.23 | inlet contract-antepart |
| DX_ICD9 | 653.33 | outlet contract-antepart |
| DX_ICD9 | 653.43 | fetopel disprop-antepart |
| DX_ICD9 | 653.53 | fetal dispro nos-antepar |
| DX_ICD9 | 653.63 | hydroceph fetus-antepart |
| DX_ICD9 | 653.73 | oth abn fet dispro-antep |
| DX_ICD9 | 653.83 | dispropor nec-antepartum |
| DX_ICD9 | 653.93 | dispropor nos-antepartum |
| DX_ICD9 | 654.03 | congen abn uter-antepart |
| DX_ICD9 | 654.13 | uterine tumor-antepartum |
| DX_ICD9 | 654.33 | retrovert uter-antepart |
| DX_ICD9 | 654.43 | abn uterus nec-antepart |
| DX_ICD9 | 654.53 | cervical incompetence- antepartum |
| DX_ICD9 | 654.63 | abnormal cervix nec- antepartum |
| DX_ICD9 | 654.64 | abnormal cervix nec- postpartum |
| DX_ICD9 | 654.73 | abnorm vagina-antepartum |
| DX_ICD9 | 654.83 | abnormal vulva-antepart |
| DX_ICD9 | 654.93 | abn pelv org nec-antepar (begin 1990) |
| DX_ICD9 | 655.03 | fetal cns malfor-antepar |
| DX_ICD9 | 655.13 | fet chromo abn-antepart |
| DX_ICD9 | 655.23 | famil hered dis-antepart |
| DX_ICD9 | 655.33 | fet damg d/t virus-antep |
| DX_ICD9 | 655.43 | fet damg d/t dis-antepar |
| DX_ICD9 | 655.63 | radiat fet damag-antepar |
| DX_ICD9 | 655.83 | fetal abnorm nec-antepar |
| DX_ICD9 | 655.93 | fetal abnorm nos-antepar |
| DX_ICD9 | 656.03 | fetal-matern hem-antepar |
| DX_ICD9 | 656.13 | rh isoimmunizat-antepart |
| DX_ICD9 | 656.23 | abo isoimmunizat-antepar |
| DX_ICD9 | 656.33 | fetal distress- antepartum |
| DX_ICD9 | 656.40 | intrauterine death- unspec |
| DX_ICD9 | 656.41 | intrauterine death- delivered |
| DX_ICD9 | 656.43 | intrauterine death- antepartum |
| DX_ICD9 | 656.53 | poor fetal growth- antepartum |
| DX_ICD9 | 656.63 | excess fetal growth- antepartum |
| DX_ICD9 | 656.73 | oth placental condition- antepartum |
| DX_ICD9 | 656.83 | fetal/placental problem nec- antepartum |
| DX_ICD9 | 656.91 | fetal/placental problem nos- delivered |
| DX_ICD9 | 656.93 | fetal/placental problem nos- antepartum |
| DX_ICD9 | 657.03 | polyhydramnios- antepartum (begin 1991) |
| DX_ICD9 | 658.03 | oligohydramnios- antepartum |
| DX_ICD9 | 658.13 | premature rupt membrane- antepartum |
| DX_ICD9 | 658.23 | prolong rup memb-antepar |
| DX_ICD9 | 658.33 | artif rupt memb-antepart |
| DX_ICD9 | 658.43 | amniotic infect-antepart |
| DX_ICD9 | 658.83 | amnion prob nec-antepart |
| DX_ICD9 | 658.93 | amnion prob nos-antepart |
| DX_ICD9 | 659.03 | fail mech induct-antepar |
| DX_ICD9 | 659.13 | fail induct nos-antepart |
| DX_ICD9 | 659.23 | pyrexia in labor-antepar |
| DX_ICD9 | 659.33 | septicem in labor-antepa |
| DX_ICD9 | 659.43 | gr& multiparity, antepartum condition or complication |
| DX_ICD9 | 659.53 | elderly primigravida, antepartum condition or complication |
| DX_ICD9 | 659.63 | advan matern age-antepa (begin 1992) |
| DX_ICD9 | 659.73 | abn fetl hrt antepart (begin 1998) |
| DX_ICD9 | 659.83 | compl labor nec-antepart |
| DX_ICD9 | 659.93 | compl labor nos-antepart |
| DX_ICD9 | 660.03 | obstruction/ fetal malposition- antepartum |
| DX_ICD9 | 660.13 | bony pelv obstruc-antepa |
| DX_ICD9 | 660.23 | abn pelv tis obstr-antep |
| DX_ICD9 | 660.33 | persist occiptpost-antep |
| DX_ICD9 | 660.43 | shoulder dystocia-antepa |
| DX_ICD9 | 660.53 | locked twins-antepartum |
| DX_ICD9 | 660.63 | fail trial lab nos-antep |
| DX_ICD9 | 660.73 | fail forceps nos-antepar |
| DX_ICD9 | 660.83 | obstruc labor nec-antepa |
| DX_ICD9 | 660.93 | obstruc labor nos-antepa |
| DX_ICD9 | 661.03 | prim uter inert-antepart |
| DX_ICD9 | 661.13 | sec uterine inert-antepa |
| DX_ICD9 | 661.23 | uterine inert nec-antepa |
| DX_ICD9 | 661.33 | precipitate labor-antepa |
| DX_ICD9 | 661.43 | uter dystocia nos-antepa |
| DX_ICD9 | 661.93 | abnorm labor nos-antepar |
| DX_ICD9 | 662.03 | prolong 1st stage-antepa |
| DX_ICD9 | 662.13 | prolong labor nos-antepa |
| DX_ICD9 | 662.23 | prolong 2nd stage-antepa |
| DX_ICD9 | 662.33 | delay del 2 twin-antepar |
| DX_ICD9 | 663.03 | cord prolapse- antepartum |
| DX_ICD9 | 663.13 | cord around neck- antepartum |
| DX_ICD9 | 663.23 | cord compression nec- antepartum |
| DX_ICD9 | 663.33 | cord entangled nec- antepartum |
| DX_ICD9 | 663.43 | short cord- antepartum |
| DX_ICD9 | 663.53 | vasa previa- antepartum |
| DX_ICD9 | 663.63 | vascular lesion cord- antepartum |
| DX_ICD9 | 663.83 | cord complication nec- antepartum |
| DX_ICD9 | 663.93 | cord complication nos- antepartum |
| DX_ICD9 | 665.00 | prelabor rupt uter-unsp |
| DX_ICD9 | 665.01 | prelabor rupt uterus-del |
| DX_ICD9 | 665.03 | prelab rupt uter-antepar |
| DX_ICD9 | 665.10 | rupt uterus nos-unsp (begin 1992) |
| DX_ICD9 | 665.11 | rupt uterus nos-deliv (begin 1992) |
| DX_ICD9 | 665.12 | rupt uter nos-del w p/p (end 1992) |
| DX_ICD9 | 665.14 | rupt uterus nos-postpart (end 1992) |
| DX_ICD9 | 665.83 | ob trauma nec-antepartum |
| DX_ICD9 | 665.93 | ob trauma nos-antepartum |
| DX_ICD9 | 666.00 | third-stage hem- unspec |
| DX_ICD9 | 668.03 | pulm complicat-antepart |
| DX_ICD9 | 668.13 | heart complic-antepart |
| DX_ICD9 | 668.23 | cns compl in del-antepar |
| DX_ICD9 | 668.83 | anesthesia complication antepartum |
| DX_ICD9 | 668.93 | anesthesia complication- antepartum |
| DX_ICD9 | 669.03 | matern distress-antepar |
| DX_ICD9 | 669.13 | obstetric shock-antepar |
| DX_ICD9 | 669.23 | maternal hypotension- antepartum |
| DX_ICD9 | 669.43 | oth cx of obstet proc- antepartum |
| DX_ICD9 | 669.71 | cesarean delivery nos |
| DX_ICD9 | 669.83 | complication delivery nec- antepartum |
| DX_ICD9 | 669.93 | complication delivery nos- antepartum |
| DX_ICD9 | 671.13 | varicose vulva-antepart |
| DX_ICD9 | 671.23 | thrombophlebit-antepart |
| DX_ICD9 | 671.30 | deep thrombosis antepartum- unspec |
| DX_ICD9 | 671.31 | deep thrombosis antepartum- delivered |
| DX_ICD9 | 671.33 | deep vein thrombosis- antepartum |
| DX_ICD9 | 671.53 | thrombosis nec- antepartum |
| DX_ICD9 | 671.83 | venous compl nec-antepar |
| DX_ICD9 | 671.93 | venous compl nos-antepar |
| DX_ICD9 | 673.03 | obstetric air embolism- antepartum |
| DX_ICD9 | 673.13 | amniotic embol-antepart |
| DX_ICD9 | 673.23 | pulmonary embolism nos- antepartum |
| DX_ICD9 | 673.33 | ob pyemic embol-antepart |
| DX_ICD9 | 673.83 | pulmonary embolism nec- antepartum |
| DX_ICD9 | 674.03 | cerebrovascular disease- antepartum |
| DX_ICD9 | 675.03 | infect nipple-antepartum |
| DX_ICD9 | 675.13 | breast abscess-antepart |
| DX_ICD9 | 675.23 | mastitis-antepartum |
| DX_ICD9 | 675.83 | breast inf nec-antepart |
| DX_ICD9 | 675.93 | breast inf nos-antepart |
| DX_ICD9 | 676.03 | retract nipple-antepart |
| DX_ICD9 | 676.13 | cracked nipple-antepart |
| DX_ICD9 | 676.23 | breast engorge-antepart |
| DX_ICD9 | 676.33 | breast dis nec-antepart |
| DX_ICD9 | 676.63 | galactorrhea-antepartum |
| DX_ICD9 | 677 | late effect of preg birth complic (begin 1994) |
| DX_ICD9 | 678.03 | fetal hematologic-ante (begin 2008) |
| DX_ICD9 | 678.11 | fetal conjoin twins-del (begin 2008) |
| DX_ICD9 | 678.13 | fetal conjoin twins-ante (begin 2008) |
| DX_ICD9 | 679.00 | mat comp in utero-unsp (begin 2008) |
| DX_ICD9 | 679.01 | mat comp in utero-del (begin 2008) |
| DX_ICD9 | 679.02 | mat comp in utro-del-p/p (begin 2008) |
| DX_ICD9 | 679.03 | mat comp in utero-ante (begin 2008) |
| DX_ICD9 | 679.04 | mat comp in utero-p/p (begin 2008) |
| DX_ICD9 | 679.10 | fetal comp in utero-unsp (begin 2008) |
| DX_ICD9 | 679.11 | fetal comp in utero-del (begin 2008) |
| DX_ICD9 | 679.12 | ftl cmp in utro-del-p/p (begin 2008) |
| DX_ICD9 | 679.13 | fetal comp in utero-ante (begin 2008) |
| DX_ICD9 | 679.14 | fetal comp in utero-p/p (begin 2008) |
| DX_ICD9 | 761.4 | ectopic preg affecting newborn |
| DX_ICD9 | 763.8 | compl delivery nec affecting newborn (end 1998) |
| DX_ICD9 | 764.0 | light-for-dates w/o fetal mal (begin 1980 end 1988) |
| DX_ICD9 | 764.09 | light-for-dates 2500+g (begin 1988) |
| DX_ICD9 | 764.1 | light-for-dates w fetal mal (begin 1980 end 1988) |
| DX_ICD9 | 764.19 | light-for-date w mal 2500+g (begin 1988) |
| DX_ICD9 | 764.2 | fetal mal w/o light-for-dates (begin 1980 end 1988) |
| DX_ICD9 | 764.9 | fetal growth retard nos (begin 1980 end 1988) |
| DX_ICD9 | 765.0 | extreme immaturity (begin 1980, end 1988) |
| DX_ICD9 | 765.1 | oth preterm infants (begin 1980, end 1988) |
| DX_ICD9 | 765.29 | 37 or more completed weeks |
| DX_ICD9 | 770.1 | newborn massive aspiration syn (end 2005) |
| DX_ICD9 | 770.8 | post-birth resp prob nec (end 2002) |
| DX_ICD9 | 772.1 | newborn intraventricular hem (end 2001) |
| DX_ICD9 | 779.6 | termination of preg |
| DX_ICD9 | 792.3 | abn find-amniotic fluid |
| PX_CPT | 1965 | anesthesia for incomplete or missed abortion procedures |
| PX_CPT | 1966 | anesthesia for induced abortion procedures |
| PX_CPT | 59120 | surgical tx ect preg ; tubal or ovarian, req salpingect/oophor, ab |
| PX_CPT | 59121 | surgical tx ect preg; tubal or ovarian, wo salpingect/oophor |
| PX_CPT | 59130 | surgical tx ect preg; abdom preg |
| PX_CPT | 59135 | surgical tx ect preg; interstitial, uterine preg req tot hysterect |
| PX_CPT | 59136 | surgi tx ect preg; interstitial, uterine preg w partial resect uterus |
| PX_CPT | 59140 | surgical tx ect preg; cervical, w evacuation |
| PX_CPT | 59150 | laparoscopic tx ect preg; wo salpingectomy/oophorectomy |
| PX_CPT | 59151 | laparoscopic tx ect preg; w salpingectomy/oophorectomy |
| PX_CPT | 59409 | vaginal delivery only (w or wo episiotomy &/or forceps); |
| PX_CPT | 59410 | vag del only (w or wo episiotomy &/or forceps); incl pp care |
| PX_CPT | 59412 | external cephalic version, w or wo tocolysis |
| PX_CPT | 59414 | delivery of plcnta (separate procedure) |
| PX_CPT | 59514 | cesarean delivery only; |
| PX_CPT | 59515 | cesarean delivery only; incl postpartum care |
| PX_CPT | 59525 | subtot or tot hysterect after c-sec del (list sep code for prim pr) |
| PX_CPT | 59612 | vag del only, after prev c-sec delivery (w or wo episioto/forceps) |
| PX_CPT | 59614 | vag del only, after prev c-sec del (w or wo episiotomy/forceps) |
| PX_CPT | 59620 | cesarean del only, after attempt vag del after prev c-sec delivery |
| PX_CPT | 59622 | cesarean del only, after attempt vag del after prev c-sec delivery |
| PX_CPT | 59812 | tx incomplete abortion, any tri, completed surgically |
| PX_CPT | 59820 | tx missed abortion, completed surgically; first tri |
| PX_CPT | 59821 | tx missed abortion, completed surgically; second tri |
| PX_CPT | 59830 | tx septic abortion, completed surgically |
| PX_CPT | 59840 | induced abortion, by dilation & curettage |
| PX_CPT | 59841 | induced abortion, by dilation & evacuation |
| PX_CPT | 59850 | induc abort, by 1+ intra-amnio injects (amnio-inject), incl hosp |
| PX_CPT | 59851 | induc abort, by 1+ intra-amnio injects (amnio-inject), incl hosp |
| PX_CPT | 59852 | induc abort, by 1+ intra-amnio injects (amnio-inject), incl hosp |
| PX_CPT | 59855 | induc abort, 1+ vag supps (prostagl&in) w or wo cerv dilation |
| PX_CPT | 59856 | induced abortion |
| PX_CPT | 59857 | induced abortion |
| PX_ICD10 | 10A00ZZ | abortion of products of conception, open approach |
| PX_ICD10 | 10A03ZZ | abortion of products of conception, percutaneous approach |
| PX_ICD10 | 10A04ZZ | abort products conception, percutaneous endoscopic approach |
| PX_ICD10 | 10A07Z6 | abort products conception, vacuum, via nat or artific opening |
| PX_ICD10 | 10A07ZW | abort products conception, laminaria, via nat or artific open |
| PX_ICD10 | 10A07ZX | abort products concept, abortifacient, via natural or artific open |
| PX_ICD10 | 10A07ZZ | abort products conception, via natural or artific opening |
| PX_ICD10 | 10A08ZZ | abortion products conception, via nat or artific opening endo |
| PX_ICD10 | 10D00Z0 | extract products conception, classical, open approach |
| PX_ICD10 | 10D00Z1 | extract products conception, low cervical, open approach |
| PX_ICD10 | 10D00Z2 | extract products conception, extraperitoneal, open approach |
| PX_ICD10 | 10D07Z3 | extract products conception, low forceps, via nat or artific open |
| PX_ICD10 | 10D07Z4 | extract products conception, mid forceps, via nat or artific open |
| PX_ICD10 | 10D07Z5 | extract products conception, high forceps, via nat or artific open |
| PX_ICD10 | 10D07Z6 | extract products conception, vacuum, via nat or artific opening |
| PX_ICD10 | 10D07Z7 | extract products conception, intern ver, via nat or artific open |
| PX_ICD10 | 10D07Z8 | extract products conception, oth, via nat or artific opening |
| PX_ICD10 | 10D17ZZ | extract products conception, retained, via nat or artific opening |
| PX_ICD10 | 10D18ZZ | extract products concept, retained, via nat or artific open endo |
| PX_ICD10 | 10D27ZZ | extract products concept, ectopic, via natural or artific opening |
| PX_ICD10 | 10D28ZZ | extract prods concept, ectopic, via natural or artific open endo |
| PX_ICD10 | 10E0XZZ | delivery products concept, external approach |
| PX_ICD10 | 10J07ZZ | inspect products of conception, via natural or artific opening |
| PX_ICD10 | 10J20ZZ | inspection of products of conception, ectopic, open approach |
| PX_ICD10 | 10J23ZZ | inspect products conception, ectopic, percutaneous approach |
| PX_ICD10 | 10J24ZZ | inspect prods concept, ectopic, percutan endoscopic approach |
| PX_ICD10 | 10J27ZZ | inspect products concept, ectopic, via nat or artific opening |
| PX_ICD10 | 10J28ZZ | inspect prods concept, ectop, via nat or artific open endoscopic |
| PX_ICD10 | 10J2XZZ | inspect products conception, ectopic, external approach |
| PX_ICD10 | 10S07ZZ | reposition products conception, via natural or artificial opening |
| PX_ICD10 | 10S0XZZ | reposition products conception, external approach |
| PX_ICD10 | 10S20ZZ | reposition products conception, ectopic, open approach |
| PX_ICD10 | 10S23ZZ | reposition products conception, ectopic, percutan approach |
| PX_ICD10 | 10S24ZZ | reposit prods concept, ectopic, percutan endoscopic approach |
| PX_ICD10 | 10S27ZZ | reposit prods concept, ectopic, via nat or artificial opening |
| PX_ICD10 | 10S28ZZ | reposit prods concept, ectopic, via nat or artifi open endoscopic |
| PX_ICD10 | 10T20ZZ | resection of products of conception, ectopic, open approach |
| PX_ICD10 | 10T23ZZ | resect products of conception, ectopic, percutaneous approach |
| PX_ICD10 | 10T23ZZ | resect products of conception, ectopic, percutaneous approach |
| PX_ICD10 | 10T24ZZ | resect products concept, ectopic, percutan endoscop approach |
| PX_ICD10 | 10T24ZZ | resect products concept, ectopic, percutan endoscop approach |
| PX_ICD10 | 10T27ZZ | resect products conception, ectopic, via nat or artificial opening |
| PX_ICD10 | 10T28ZZ | resect products concept, ectopic, via nat or artif open endoscop |
| DX_ICD10 | A34 | obstetrical tetanus |
| DX_ICD10 | F53 | puerperal psychosis |
| DX_ICD10 | M83.0 | puerperal osteomalacia |
| DX_ICD10 | O00 | abdom preg |
| DX_ICD10 | O00.0 | abdom preg |
| DX_ICD10 | O00.00 | abdom preg wo intrauterine preg |
| DX_ICD10 | O00.01 | abdom preg with intrauterine preg |
| DX_ICD10 | O00.1 | tubal preg |
| DX_ICD10 | O00.10 | tubal preg wo intrauterine preg |
| DX_ICD10 | O00.101 | right tubal preg wo intrauterine preg |
| DX_ICD10 | O00.102 | left tubal preg wo intrauterine preg |
| DX_ICD10 | O00.109 | unspec tubal preg wo intrauterine preg |
| DX_ICD10 | O00.11 | tubal preg with intrauterine preg |
| DX_ICD10 | O00.111 | right tubal preg with intrauterine preg |
| DX_ICD10 | O00.112 | left tubal preg with intrauterine preg |
| DX_ICD10 | O00.119 | unspec tubal preg with intrauterine preg |
| DX_ICD10 | O00.2 | ovarian preg |
| DX_ICD10 | O00.20 | ovarian preg wo intrauterine preg |
| DX_ICD10 | O00.201 | right ovarian preg wo intrauterine preg |
| DX_ICD10 | O00.202 | left ovarian preg wo intrauterine preg |
| DX_ICD10 | O00.209 | unspec ovarian preg wo intrauterine preg |
| DX_ICD10 | O00.21 | ovarian preg with intrauterine preg |
| DX_ICD10 | O00.211 | right ovarian preg with intrauterine preg |
| DX_ICD10 | O00.212 | left ovarian preg with intrauterine preg |
| DX_ICD10 | O00.219 | unspec ovarian preg with intrauterine preg |
| DX_ICD10 | O00.8 | oth ectopic preg |
| DX_ICD10 | O00.80 | oth ectopic preg wo intrauterine preg |
| DX_ICD10 | O00.81 | oth ectopic preg with intrauterine preg |
| DX_ICD10 | O00.9 | ectopic preg, unspec |
| DX_ICD10 | O00.90 | unspec ectopic preg wo intrauterine preg |
| DX_ICD10 | O00.91 | unspec ectopic preg with intrauterine preg |
| DX_ICD10 | O02 | oth abnormal products of conception) |
| DX_ICD10 | O02.1 | missed abortion |
| DX_ICD10 | O02.8 | oth specif abnormal products of conception |
| DX_ICD10 | O02.81 | inapprop change in quantitative hcg in early preg |
| DX_ICD10 | O02.89 | oth abnormal products of conception |
| DX_ICD10 | O02.9 | abnormal product of conception, unspec |
| DX_ICD10 | O03 | SAB |
| DX_ICD10 | O03.0 | genital tract & pelvic infection after incomplete SAB |
| DX_ICD10 | O03.1 | delayed or excessive hemorrhage following incomplete SAB |
| DX_ICD10 | O03.2 | embolism following incomplete SAB |
| DX_ICD10 | O03.3 | genital tract & pelvic infection following incomplete SAB |
| DX_ICD10 | O03.30 | unspec complication following incomplete SAB |
| DX_ICD10 | O03.31 | shock following incomplete SAB |
| DX_ICD10 | O03.32 | renal failure following incomplete SAB |
| DX_ICD10 | O03.33 | metabolic disorder following incomplete SAB |
| DX_ICD10 | O03.34 | dam to pelvic organs following incomplete SAB |
| DX_ICD10 | O03.35 | oth venous complic following incomplete SAB |
| DX_ICD10 | O03.36 | cardiac arrest following incomplete SAB |
| DX_ICD10 | O03.37 | sepsis following incomplete SAB |
| DX_ICD10 | O03.38 | urinary tract infection following incomplete SAB |
| DX_ICD10 | O03.39 | incomplete SAB with oth complic |
| DX_ICD10 | O03.4 | incomplete SAB wo complication |
| DX_ICD10 | O03.5 | genital tract & pelvic infect after complete or unspec SAB |
| DX_ICD10 | O03.6 | delayed or excessive hemor after complete or unspec SAB |
| DX_ICD10 | O03.7 | embolism following complete or unspec SAB |
| DX_ICD10 | O03.8 | oth & unspec complic after complete or unspec SAB |
| DX_ICD10 | O03.80 | unspec complication after complete or unspec SAB |
| DX_ICD10 | O03.81 | shock following complete or unspec SAB |
| DX_ICD10 | O03.82 | renal failure following complete or unspec SAB |
| DX_ICD10 | O03.83 | metabolic disorder following complete or unspec SAB |
| DX_ICD10 | O03.84 | dam to pelvic organs after complete or unspec SAB |
| DX_ICD10 | O03.85 | oth venous complic after complete or unspec SAB |
| DX_ICD10 | O03.86 | cardiac arrest following complete or unspec SAB |
| DX_ICD10 | O03.87 | sepsis following complete or unspec SAB |
| DX_ICD10 | O03.88 | urinary tract infection after complete or unspec SAB |
| DX_ICD10 | O03.89 | complete or unspec SAB with oth complic |
| DX_ICD10 | O03.9 | complete or unspec SAB wo complication |
| DX_ICD10 | O04 | complic following (induced) termination of preg |
| DX_ICD10 | O04.5 | genital tract & pelvic infection after (induced) termination preg |
| DX_ICD10 | O04.6 | delayed or excessive hemor after (induced) termination preg |
| DX_ICD10 | O04.7 | embolism following (induced) termination of preg |
| DX_ICD10 | O04.8 | (induced) termination preg with oth & unspec complic |
| DX_ICD10 | O04.80 | (induced) termination preg with unspec complic |
| DX_ICD10 | O04.81 | shock following (induced) termination of preg |
| DX_ICD10 | O04.82 | renal failure following (induced) termination of preg |
| DX_ICD10 | O04.83 | metabolic disorder following (induced) termination of preg |
| DX_ICD10 | O04.84 | dam to pelvic organs following (induced) termination of preg |
| DX_ICD10 | O04.85 | oth venous complic after (induced) termination preg |
| DX_ICD10 | O04.86 | cardiac arrest following (induced) termination of preg |
| DX_ICD10 | O04.87 | sepsis following (induced) termination of preg |
| DX_ICD10 | O04.88 | urinary tract infection following (induced) termination of preg |
| DX_ICD10 | O04.89 | (induced) termination of preg with oth complic |
| DX_ICD10 | O07 | failed attempted termination of preg |
| DX_ICD10 | O07.0 | genital tract & pelvic infect after failed attempt termination preg |
| DX_ICD10 | O07.1 | delayed or excessive hemor after fail attempt termin preg |
| DX_ICD10 | O07.2 | embolism following failed attempted termination of preg |
| DX_ICD10 | O07.3 | failed attempted termin preg with oth & unspec complic |
| DX_ICD10 | O07.30 | failed attempted termination of preg with unspec complic |
| DX_ICD10 | O07.31 | shock following failed attempted termination of preg |
| DX_ICD10 | O07.32 | renal failure following failed attempted termination of preg |
| DX_ICD10 | O07.33 | metabolic disorder after failed attempted termination of preg |
| DX_ICD10 | O07.34 | dam to pelvic organs after failed attempt termin of preg |
| DX_ICD10 | O07.35 | oth venous complic after failed attempted termination of preg |
| DX_ICD10 | O07.36 | cardiac arrest after failed attempted termination of preg |
| DX_ICD10 | O07.37 | sepsis following failed attempted termination of preg |
| DX_ICD10 | O07.38 | urinary tract infection after failed attempted termination of preg |
| DX_ICD10 | O07.39 | failed attempted termination of preg with oth complic |
| DX_ICD10 | O07.4 | failed attempted termination of preg wo complication |
| DX_ICD10 | O08 | complic following ectopic & molar preg |
| DX_ICD10 | O08.0 | genital tract & pelvic infection after ectopic & molar preg |
| DX_ICD10 | O08.1 | delayed or excessive hemor after ectopic & molar preg |
| DX_ICD10 | O08.2 | embolism following ectopic & molar preg |
| DX_ICD10 | O08.4 | renal failure following ectopic & molar preg |
| DX_ICD10 | O08.5 | metabolic disorders following an ectopic & molar preg |
| DX_ICD10 | O08.6 | dam to pelvic organs & tissues after ectopic & molar preg |
| DX_ICD10 | O08.7 | oth venous complic after an ectopic & molar preg |
| DX_ICD10 | O08.8 | oth complic after an ectopic & molar preg |
| DX_ICD10 | O08.81 | cardiac arrest following an ectopic & molar preg |
| DX_ICD10 | O08.82 | sepsis following ectopic & molar preg |
| DX_ICD10 | O08.83 | urinary tract infection following an ectopic & molar preg |
| DX_ICD10 | O08.89 | oth complic following an ectopic & molar preg |
| DX_ICD10 | O08.9 | unspec complication following an ectopic & molar preg |
| DX_ICD10 | O09 | supervision of high risk preg |
| DX_ICD10 | O09.0 | supervis preg with history of infertility, unspec tri |
| DX_ICD10 | O09.00 | supervis preg with history of infertility, unspec tri |
| DX_ICD10 | O09.01 | supervis preg with history of infertility, first tri |
| DX_ICD10 | O09.02 | supervis preg with history of infertility, second tri |
| DX_ICD10 | O09.03 | supervis preg with history of infertility, third tri |
| DX_ICD10 | O09.10 | supervis preg with history of ectopic preg, unspec tri |
| DX_ICD10 | O09.11 | supervis preg with history of ectopic preg, first tri |
| DX_ICD10 | O09.12 | supervis preg with history of ectopic preg, second tri |
| DX_ICD10 | O09.13 | supervis preg with history of ectopic preg, third tri |
| DX_ICD10 | O09.2 | supervis preg with oth poor reproduct or obstetric hx |
| DX_ICD10 | O09.21 | supervis preg with history of pre-term labor |
| DX_ICD10 | O09.211 | supervis preg with history of pre-term labor, first tri |
| DX_ICD10 | O09.212 | supervis preg with history of pre-term labor, second tri |
| DX_ICD10 | O09.213 | supervis preg with history of pre-term labor, third tri |
| DX_ICD10 | O09.219 | supervis preg with history of pre-term labor, unspec tri |
| DX_ICD10 | O09.29 | supervis preg with oth poor reproductive or obstetric hx |
| DX_ICD10 | O09.291 | supervision preg with oth poor reproduct or obstetric hx, first tri |
| DX_ICD10 | O09.292 | supervis preg w oth poor reproducti or obstetric hx, second tri |
| DX_ICD10 | O09.293 | supervis preg w oth poor reproduct or obstetric hx, third tri |
| DX_ICD10 | O09.299 | supervis preg w oth poor reproduct or obstetric hx, unspec tri |
| DX_ICD10 | O09.3 | supervis preg with insufficient antenatal care |
| DX_ICD10 | O09.30 | supervis preg with insufficient antenatal care, unspec tri |
| DX_ICD10 | O09.31 | supervis preg with insufficient antenatal care, first tri |
| DX_ICD10 | O09.32 | supervis preg with insufficient antenatal care, second tri |
| DX_ICD10 | O09.33 | supervis preg with insufficient antenatal care, third tri |
| DX_ICD10 | O09.4 | supervis preg with gr& multiparity |
| DX_ICD10 | O09.40 | supervis preg with gr& multiparity, unspec tri |
| DX_ICD10 | O09.41 | supervis preg with gr& multiparity, first tri |
| DX_ICD10 | O09.42 | supervis preg with gr& multiparity, second tri |
| DX_ICD10 | O09.43 | supervis preg with gr& multiparity, third tri |
| DX_ICD10 | O09.5 | supervision of elderly primigravida & multigravida |
| DX_ICD10 | O09.51 | supervision of elderly primigravida |
| DX_ICD10 | O09.511 | supervision of elderly primigravida, first tri |
| DX_ICD10 | O09.512 | supervision of elderly primigravida, second tri |
| DX_ICD10 | O09.513 | supervision of elderly primigravida, third tri |
| DX_ICD10 | O09.519 | supervision of elderly primigravida, unspec tri |
| DX_ICD10 | O09.52 | supervision of elderly multigravida |
| DX_ICD10 | O09.521 | supervision of elderly multigravida, first tri |
| DX_ICD10 | O09.522 | supervision of elderly multigravida, second tri |
| DX_ICD10 | O09.523 | supervision of elderly multigravida, third tri |
| DX_ICD10 | O09.529 | supervision of elderly multigravida, unspec tri |
| DX_ICD10 | O09.6 | supervision of young primigravida & multigravida |
| DX_ICD10 | O09.61 | supervision of young primigravida |
| DX_ICD10 | O09.611 | supervision of young primigravida, first tri |
| DX_ICD10 | O09.612 | supervision of young primigravida, second tri |
| DX_ICD10 | O09.613 | supervision of young primigravida, third tri |
| DX_ICD10 | O09.619 | supervision of young primigravida, unspec tri |
| DX_ICD10 | O09.62 | supervision of young multigravida |
| DX_ICD10 | O09.621 | supervision of young multigravida, first tri |
| DX_ICD10 | O09.622 | supervision of young multigravida, second tri |
| DX_ICD10 | O09.623 | supervision of young multigravida, third tri |
| DX_ICD10 | O09.629 | supervision of young multigravida, unspec tri |
| DX_ICD10 | O09.7 | supervision of high risk preg due to social problems |
| DX_ICD10 | O09.70 | supervision of high risk preg due to social problems, unspec tri |
| DX_ICD10 | O09.71 | supervision of high risk preg due to social problems, first tri |
| DX_ICD10 | O09.72 | supervision of high risk preg due to social problems, second tri |
| DX_ICD10 | O09.73 | supervision of high risk preg due to social problems, third tri |
| DX_ICD10 | O09.8 | supervision of oth high risk pregnancies |
| DX_ICD10 | O09.81 | supervision preg resulting from assisted reproductive tech |
| DX_ICD10 | O09.811 | supervis preg resulting from assisted reproductive tech, first tri |
| DX_ICD10 | O09.812 | supervis preg resulting from assisted reproduct tech, second tri |
| DX_ICD10 | O09.813 | supervis preg resulting from assisted reproduct tech, third tri |
| DX_ICD10 | O09.819 | supervis preg resulting from assisted reproduct tech, unspec tri |
| DX_ICD10 | O09.82 | supervis preg w hx of in utero procedure during previous preg |
| DX_ICD10 | O09.821 | supervis preg w hx of in utero procedure during prev preg, first tri |
| DX_ICD10 | O09.822 | supervis preg w hx of in utero proc during prev preg, second tri |
| DX_ICD10 | O09.823 | supervis preg w hx of in utero proc during prev preg, third tri |
| DX_ICD10 | O09.829 | supervis preg w hx of in utero proc during prev preg, unspec tri |
| DX_ICD10 | O09.89 | supervision of oth high risk pregnancies |
| DX_ICD10 | O09.891 | supervision of oth high risk pregnancies, first tri |
| DX_ICD10 | O09.892 | supervision of oth high risk pregnancies, second tri |
| DX_ICD10 | O09.893 | supervision of oth high risk pregnancies, third tri |
| DX_ICD10 | O09.899 | supervision of oth high risk pregnancies, unspec tri |
| DX_ICD10 | O09.9 | supervision of high risk preg, unspec |
| DX_ICD10 | O09.90 | supervision of high risk preg, unspec, unspec tri |
| DX_ICD10 | O09.91 | supervision of high risk preg, unspec, first tri |
| DX_ICD10 | O09.92 | supervision of high risk preg, unspec, second tri |
| DX_ICD10 | O09.93 | supervision of high risk preg, unspec, third tri |
| DX_ICD10 | O09.A0 | supervis preg with hx of molar preg, unspec tri |
| DX_ICD10 | O09.A1 | supervis preg with history of molar preg, first tri |
| DX_ICD10 | O09.A2 | supervis preg with history of molar preg, second tri |
| DX_ICD10 | O09.A3 | supervis preg with history of molar preg, third tri |
| DX_ICD10 | O10 | pre-existing htn complic preg, childbirth & the puerperium |
| DX_ICD10 | O10.0 | pre-existing essential htn complic preg, childbirth & puerperium |
| DX_ICD10 | O10.01 | pre-existing essential htn complic preg |
| DX_ICD10 | O10.011 | pre-existing essential htn complic preg, first tri |
| DX_ICD10 | O10.012 | pre-existing essential htn complic preg, second tri |
| DX_ICD10 | O10.013 | pre-existing essential htn complic preg, third tri |
| DX_ICD10 | O10.019 | pre-existing essential htn complic preg, unspec tri |
| DX_ICD10 | O10.02 | pre-existing essential htn complic childbirth |
| DX_ICD10 | O10.03 | pre-existing essential htn complic the puerperium |
| DX_ICD10 | O10.1 | pre-exist hypertens hrt dis complic preg, chldbrth & puerperium |
| DX_ICD10 | O10.11 | pre-existing hypertensive hrt dis complic preg |
| DX_ICD10 | O10.111 | pre-existing hypertensive hrt dis complic preg, first tri |
| DX_ICD10 | O10.112 | pre-existing hypertensive hrt dis complic preg, second tri |
| DX_ICD10 | O10.113 | pre-existing hypertensive hrt dis complic preg, third tri |
| DX_ICD10 | O10.119 | pre-existing hypertensive hrt dis complic preg, unspec tri |
| DX_ICD10 | O10.12 | pre-existing hypertensive heart disease complic childbirth |
| DX_ICD10 | O10.13 | pre-existing hypertens hrt dis complic the puerperium |
| DX_ICD10 | O10.2 | pre-exist hypertens chron kidn dis complic preg, brth & puerperium |
| DX_ICD10 | O10.21 | pre-existing hypertensive chronic kidney disease complic preg |
| DX_ICD10 | O10.211 | pre-exist hypertensive chron kidn dis complic preg, first tri |
| DX_ICD10 | O10.212 | pre-existing hypertensive chron kidn dis complic preg, second tri |
| DX_ICD10 | O10.213 | pre-existing hypertensive chron kidn dis complicg preg, third tri |
| DX_ICD10 | O10.219 | pre-existing hypertensive chronic kidn dis complic preg, unspec tri |
| DX_ICD10 | O10.22 | pre-existing hypertensive chronic kidney disease complic childbirth |
| DX_ICD10 | O10.23 | pre-existing hypertensive chron kidn dis complic the puerperium |
| DX_ICD10 | O10.3 | pre-exist hyptnsive hrt & chron kidn dis compl preg, brth & puerperium |
| DX_ICD10 | O10.31 | pre-existing hypertensive hrt & chron kidn dis complic preg |
| DX_ICD10 | O10.311 | pre-exist hypertensive hrt & chron kidn dis complic preg, first tri |
| DX_ICD10 | O10.312 | pre-existing hypertensive hrt & chron kidn dis complic preg, second tri |
| DX_ICD10 | O10.313 | pre-existing hypertensive hrt & chron kidn dis complic preg, third tri |
| DX_ICD10 | O10.319 | pre-existing hypertensive hrt & chron kidn dis complic preg, unspec tri |
| DX_ICD10 | O10.32 | pre-existing hypertensive hrt & chron kidn dis complic childbirth |
| DX_ICD10 | O10.33 | pre-existing hypertensive hrt & chron kidn dis complic puerperium |
| DX_ICD10 | O10.4 | pre-exist secondary htn complic preg, childbirth & the puerperium |
| DX_ICD10 | O10.41 | pre-existing secondary htn complic preg |
| DX_ICD10 | O10.411 | pre-existing secondary htn complic preg, first tri |
| DX_ICD10 | O10.412 | pre-existing secondary htn complic preg, second tri |
| DX_ICD10 | O10.413 | pre-existing secondary htn complic preg, third tri |
| DX_ICD10 | O10.419 | pre-existing secondary htn complic preg, unspec tri |
| DX_ICD10 | O10.42 | pre-existing secondary htn complic childbirth |
| DX_ICD10 | O10.43 | pre-existing secondary htn complic the puerperium |
| DX_ICD10 | O10.9 | unspec pre-existing htn complic preg, childbirth & the puerperium) |
| DX_ICD10 | O10.91 | unspec pre-existing htn complic preg |
| DX_ICD10 | O10.911 | unspec pre-existing htn complic preg, first tri |
| DX_ICD10 | O10.912 | unspec pre-existing htn complic preg, second tri |
| DX_ICD10 | O10.913 | unspec pre-existing htn complic preg, third tri |
| DX_ICD10 | O10.919 | unspec pre-existing htn complic preg, unspec tri |
| DX_ICD10 | O10.92 | unspec pre-existing htn complic childbirth |
| DX_ICD10 | O10.93 | unspec pre-existing htn complic the puerperium |
| DX_ICD10 | O11 | pre-existing hypertens disorder with superimposed proteinuria |
| DX_ICD10 | O11.4 | pre-existing htn with pre-eclampsia, complic childbirth |
| DX_ICD10 | O11.5 | pre-existing htn with pre-eclampsia, complic the puerperium |
| DX_ICD10 | O12 | gestal [preg-induced] edema & proteinuria wo hypertension |
| DX_ICD10 | O12.0 | gestal edema |
| DX_ICD10 | O12.00 | gestal edema, unspec tri |
| DX_ICD10 | O12.01 | gestal edema, first tri |
| DX_ICD10 | O12.02 | gestal edema, second tri |
| DX_ICD10 | O12.03 | gestal edema, third tri |
| DX_ICD10 | O12.04 | gestal edema, complic childbirth |
| DX_ICD10 | O12.05 | gestal edema, complic the puerperium |
| DX_ICD10 | O12.1 | gestal proteinuria |
| DX_ICD10 | O12.10 | gestal proteinuria, unspec tri |
| DX_ICD10 | O12.11 | gestal proteinuria, first tri |
| DX_ICD10 | O12.12 | gestal proteinuria, second tri |
| DX_ICD10 | O12.13 | gestal proteinuria, third tri |
| DX_ICD10 | O12.14 | gestal proteinuria, complic childbirth |
| DX_ICD10 | O12.15 | gestal proteinuria, complic the puerperium |
| DX_ICD10 | O12.2 | gestal edema with proteinuria |
| DX_ICD10 | O12.20 | gestal edema with proteinuria, unspec tri |
| DX_ICD10 | O12.21 | gestal edema with proteinuria, first tri |
| DX_ICD10 | O12.22 | gestal edema with proteinuria, second tri |
| DX_ICD10 | O12.23 | gestal edema with proteinuria, third tri |
| DX_ICD10 | O12.24 | gestal edema with proteinuria, complic childbirth |
| DX_ICD10 | O12.25 | gestal edema with proteinuria, complic the puerperium |
| DX_ICD10 | O13 | gestal [preg-induced] htn wo significant proteinuria |
| DX_ICD10 | O13.1 | gestal [preg-induced] htn wo significant proteinuria, first tri |
| DX_ICD10 | O13.2 | gestal [preg-induced] htn wo significant proteinuria, second tri |
| DX_ICD10 | O13.3 | gestal [preg-induced] htn wo significant proteinuria, third tri |
| DX_ICD10 | O13.4 | gestal [preg-induced] htn wo significant proteinuria, complic childbirth |
| DX_ICD10 | O13.5 | gest [preg-induced] htn wo signif proteinuria, complic puerperium |
| DX_ICD10 | O13.9 | gest [preg-induced] htn wo significant proteinuria, unspec tri |
| DX_ICD10 | O14 | gestal [preg-induced] htn with significant proteinuria |
| DX_ICD10 | O14.0 | mild pre-eclampsia |
| DX_ICD10 | O14.00 | mild to moderate pre-eclampsia, unspec tri |
| DX_ICD10 | O14.02 | mild to moderate pre-eclampsia, second tri |
| DX_ICD10 | O14.03 | mild to moderate pre-eclampsia, third tri |
| DX_ICD10 | O14.04 | mild to moderate pre-eclampsia, complic childbirth |
| DX_ICD10 | O14.05 | mild to moderate pre-eclampsia, complic the puerperium |
| DX_ICD10 | O14.1 | severe pre-eclampsia |
| DX_ICD10 | O14.10 | severe pre-eclampsia, unspec tri |
| DX_ICD10 | O14.12 | severe pre-eclampsia, second tri |
| DX_ICD10 | O14.13 | severe pre-eclampsia, third tri |
| DX_ICD10 | O14.14 | severe pre-eclampsia complic childbirth |
| DX_ICD10 | O14.15 | severe pre-eclampsia, complic the puerperium |
| DX_ICD10 | O14.2 | hellp syndrome (hellp) |
| DX_ICD10 | O14.20 | hellp syndrome (hellp), unspec tri |
| DX_ICD10 | O14.22 | hellp syndrome (hellp), second tri |
| DX_ICD10 | O14.23 | hellp syndrome (hellp), third tri |
| DX_ICD10 | O14.24 | hellp syndrome, complic childbirth |
| DX_ICD10 | O14.25 | hellp syndrome, complic the puerperium |
| DX_ICD10 | O14.9 | unspec pre-eclampsia |
| DX_ICD10 | O14.90 | unspec pre-eclampsia, unspec tri |
| DX_ICD10 | O14.92 | unspec pre-eclampsia, second tri |
| DX_ICD10 | O14.93 | unspec pre-eclampsia, third tri |
| DX_ICD10 | O14.94 | unspec pre-eclampsia, complic childbirth |
| DX_ICD10 | O14.95 | unspec pre-eclampsia, complic the puerperium |
| DX_ICD10 | O15 | eclampsia |
| DX_ICD10 | O15.0 | eclampsia in preg |
| DX_ICD10 | O15.00 | eclampsia complic preg, unspec tri |
| DX_ICD10 | O15.02 | eclampsia complic preg, second tri |
| DX_ICD10 | O15.03 | eclampsia complic preg, third tri |
| DX_ICD10 | O15.1 | eclampsia complic labor |
| DX_ICD10 | O15.2 | eclampsia complic the puerperium |
| DX_ICD10 | O15.9 | eclampsia, unspec as to time period |
| DX_ICD10 | O16 | unspec maternal hypertension |
| DX_ICD10 | O16.1 | unspec maternal hypertension, first tri |
| DX_ICD10 | O16.2 | unspec maternal hypertension, second tri |
| DX_ICD10 | O16.3 | unspec maternal hypertension, third tri |
| DX_ICD10 | O16.4 | unspec maternal hypertension, complic childbirth |
| DX_ICD10 | O16.5 | unspec maternal hypertension, complic the puerperium |
| DX_ICD10 | O16.9 | unspec maternal hypertension, unspec tri |
| DX_ICD10 | O20 | hemorrhage in early preg |
| DX_ICD10 | O20.0 | threatened abortion |
| DX_ICD10 | O20.8 | oth hemorrhage in early preg |
| DX_ICD10 | O20.9 | hemorrhage in early preg, unspec |
| DX_ICD10 | O21 | excessive vomiting in preg |
| DX_ICD10 | O21.0 | mild hyperemesis gravidarum |
| DX_ICD10 | O21.1 | hyperemesis gravidarum with metabolic disturbance |
| DX_ICD10 | O21.2 | late vomiting of preg |
| DX_ICD10 | O21.8 | oth vomiting complic preg |
| DX_ICD10 | O21.9 | vomiting of preg, unspec |
| DX_ICD10 | O22 | venous complic in preg |
| DX_ICD10 | O22.0 | varicose veins of lower extremity in preg |
| DX_ICD10 | O22.00 | varicose veins of lower extremity in preg, unspec tri |
| DX_ICD10 | O22.01 | varicose veins of lower extremity in preg, first tri |
| DX_ICD10 | O22.02 | varicose veins of lower extremity in preg, second tri |
| DX_ICD10 | O22.03 | varicose veins of lower extremity in preg, third tri |
| DX_ICD10 | O22.1 | genital varices in preg |
| DX_ICD10 | O22.10 | genital varices in preg, unspec tri |
| DX_ICD10 | O22.11 | genital varices in preg, first tri |
| DX_ICD10 | O22.12 | genital varices in preg, second tri |
| DX_ICD10 | O22.13 | genital varices in preg, third tri |
| DX_ICD10 | O22.2 | superficial thrombophlebitis in preg |
| DX_ICD10 | O22.20 | superficial thrombophlebitis in preg, unspec tri |
| DX_ICD10 | O22.21 | superficial thrombophlebitis in preg, first tri |
| DX_ICD10 | O22.22 | superficial thrombophlebitis in preg, second tri |
| DX_ICD10 | O22.23 | superficial thrombophlebitis in preg, third tri |
| DX_ICD10 | O22.3 | deep phlebothrombosis in preg |
| DX_ICD10 | O22.30 | deep phlebothrombosis in preg, unspec tri |
| DX_ICD10 | O22.31 | deep phlebothrombosis in preg, first tri |
| DX_ICD10 | O22.32 | deep phlebothrombosis in preg, second tri |
| DX_ICD10 | O22.33 | deep phlebothrombosis in preg, third tri |
| DX_ICD10 | O22.4 | hemorrhoids in preg |
| DX_ICD10 | O22.40 | cerebral venous thrombosis in preg, unspec tri |
| DX_ICD10 | O22.41 | cerebral venous thrombosis in preg, first tri |
| DX_ICD10 | O22.42 | cerebral venous thrombosis in preg, second tri |
| DX_ICD10 | O22.43 | cerebral venous thrombosis in preg, third tri |
| DX_ICD10 | O22.5 | cerebral venous thrombosis in preg |
| DX_ICD10 | O22.50 | cerebral venous thrombosis in preg, unspec tri |
| DX_ICD10 | O22.51 | cerebral venous thrombosis in preg, first tri |
| DX_ICD10 | O22.52 | cerebral venous thrombosis in preg, second tri |
| DX_ICD10 | O22.53 | cerebral venous thrombosis in preg, third tri |
| DX_ICD10 | O22.8 | oth venous complic in preg |
| DX_ICD10 | O22.8X | oth venous complic in preg |
| DX_ICD10 | O22.8X1 | oth venous complic in preg, first tri |
| DX_ICD10 | O22.8X2 | oth venous complic in preg, second tri |
| DX_ICD10 | O22.8X3 | oth venous complic in preg, third tri |
| DX_ICD10 | O22.8X9 | oth venous complic in preg, unspec tri |
| DX_ICD10 | O22.9 | venous complication in preg, unspec |
| DX_ICD10 | O22.90 | venous complication in preg, unspec, unspec tri |
| DX_ICD10 | O22.91 | venous complication in preg, unspec, first tri |
| DX_ICD10 | O22.92 | venous complication in preg, unspec, second tri |
| DX_ICD10 | O22.93 | venous complication in preg, unspec, third tri |
| DX_ICD10 | O23 | infections of genitourinary tract in preg |
| DX_ICD10 | O23.0 | infections of kidney in preg |
| DX_ICD10 | O23.00 | infections of kidney in preg, unspec tri |
| DX_ICD10 | O23.01 | infections of kidney in preg, first tri |
| DX_ICD10 | O23.02 | infections of kidney in preg, second tri |
| DX_ICD10 | O23.03 | infections of kidney in preg, third tri |
| DX_ICD10 | O23.1 | infections of bladder in preg |
| DX_ICD10 | O23.10 | infections of bladder in preg, unspec tri |
| DX_ICD10 | O23.11 | infections of bladder in preg, first tri |
| DX_ICD10 | O23.12 | infections of bladder in preg, second tri |
| DX_ICD10 | O23.13 | infections of bladder in preg, third tri |
| DX_ICD10 | O23.2 | infections of urethra in preg |
| DX_ICD10 | O23.20 | infections of urethra in preg, unspec tri |
| DX_ICD10 | O23.21 | infections of urethra in preg, first tri |
| DX_ICD10 | O23.22 | infections of urethra in preg, second tri |
| DX_ICD10 | O23.23 | infections of urethra in preg, third tri |
| DX_ICD10 | O23.3 | infections of oth parts of urinary tract in preg |
| DX_ICD10 | O23.30 | infections of oth parts of urinary tract in preg, unspec tri |
| DX_ICD10 | O23.31 | infections of oth parts of urinary tract in preg, first tri |
| DX_ICD10 | O23.32 | infections of oth parts of urinary tract in preg, second tri |
| DX_ICD10 | O23.33 | infections of oth parts of urinary tract in preg, third tri |
| DX_ICD10 | O23.4 | unspec infection of urinary tract in preg |
| DX_ICD10 | O23.40 | unspec infection of urinary tract in preg, unspec tri |
| DX_ICD10 | O23.41 | unspec infection of urinary tract in preg, first tri |
| DX_ICD10 | O23.42 | unspec infection of urinary tract in preg, second tri |
| DX_ICD10 | O23.43 | unspec infection of urinary tract in preg, third tri |
| DX_ICD10 | O23.5 | infections of the genital tract in preg |
| DX_ICD10 | O23.51 | infection of cervix in preg |
| DX_ICD10 | O23.511 | infections of cervix in preg, first tri |
| DX_ICD10 | O23.512 | infections of cervix in preg, second tri |
| DX_ICD10 | O23.513 | infections of cervix in preg, third tri |
| DX_ICD10 | O23.519 | infections of cervix in preg, unspec tri |
| DX_ICD10 | O23.52 | salpingo-oophoritis in preg |
| DX_ICD10 | O23.521 | salpingo-oophoritis in preg, first tri |
| DX_ICD10 | O23.522 | salpingo-oophoritis in preg, second tri |
| DX_ICD10 | O23.523 | salpingo-oophoritis in preg, third tri |
| DX_ICD10 | O23.529 | salpingo-oophoritis in preg, unspec tri |
| DX_ICD10 | O23.59 | infection of oth part of genital tract in preg |
| DX_ICD10 | O23.591 | infection of oth part of genital tract in preg, first tri |
| DX_ICD10 | O23.592 | infection of oth part of genital tract in preg, second tri |
| DX_ICD10 | O23.593 | infection of oth part of genital tract in preg, third tri |
| DX_ICD10 | O23.599 | infection of oth part of genital tract in preg, unspec tri |
| DX_ICD10 | O23.9 | unspec genitourinary tract infection in preg |
| DX_ICD10 | O23.90 | unspec genitourinary tract infection in preg, unspec tri |
| DX_ICD10 | O23.91 | unspec genitourinary tract infection in preg, first tri |
| DX_ICD10 | O23.92 | unspec genitourinary tract infection in preg, second tri |
| DX_ICD10 | O23.93 | unspec genitourinary tract infection in preg, third tri |
| DX_ICD10 | O24 | diabetes mellitus in preg, childbirth, & the puerperium |
| DX_ICD10 | O24.0 | pre-existing dm, type 1, in preg, childbirth & the puerperium |
| DX_ICD10 | O24.01 | pre-existing dm, type 1, in preg |
| DX_ICD10 | O24.011 | pre-existing type 1 diabetes mellitus, in preg, first tri |
| DX_ICD10 | O24.012 | pre-existing type 1 diabetes mellitus, in preg, second tri |
| DX_ICD10 | O24.013 | pre-existing type 1 diabetes mellitus, in preg, third tri |
| DX_ICD10 | O24.019 | pre-existing type 1 diabetes mellitus, in preg, unspec tri |
| DX_ICD10 | O24.02 | pre-existing type 1 diabetes mellitus, in childbirth |
| DX_ICD10 | O24.03 | pre-existing type 1 diabetes mellitus, in the puerperium |
| DX_ICD10 | O24.1 | pre-existing dm, type 2, in preg, childbirth & the puerperium |
| DX_ICD10 | O24.11 | pre-existing diabetes mellitus, type 2, in preg |
| DX_ICD10 | O24.111 | pre-existing type 2 diabetes mellitus, in preg, first tri |
| DX_ICD10 | O24.112 | pre-existing type 2 diabetes mellitus, in preg, second tri |
| DX_ICD10 | O24.113 | pre-existing type 2 diabetes mellitus, in preg, third tri |
| DX_ICD10 | O24.119 | pre-existing type 2 diabetes mellitus, in preg, unspec tri |
| DX_ICD10 | O24.12 | pre-existing type 2 diabetes mellitus, in childbirth |
| DX_ICD10 | O24.13 | pre-existing type 2 diabetes mellitus, in the puerperium |
| DX_ICD10 | O24.3 | unspec pre-existing dm in preg, childbirth & the puerperium) |
| DX_ICD10 | O24.31 | unspec pre-existing diabetes mellitus in preg |
| DX_ICD10 | O24.311 | unspec pre-existing diabetes mellitus in preg, first tri |
| DX_ICD10 | O24.312 | unspec pre-existing diabetes mellitus in preg, second tri |
| DX_ICD10 | O24.313 | unspec pre-existing diabetes mellitus in preg, third tri |
| DX_ICD10 | O24.319 | unspec pre-existing dm in preg, unspec tri |
| DX_ICD10 | O24.32 | unspec pre-existing diabetes mellitus in childbirth |
| DX_ICD10 | O24.33 | unspec pre-existing diabetes mellitus in the puerperium |
| DX_ICD10 | O24.4 | gestal diabetes mellitus |
| DX_ICD10 | O24.41 | gestal diabetes mellitus in preg |
| DX_ICD10 | O24.410 | gestal diabetes mellitus in preg, diet controlled |
| DX_ICD10 | O24.414 | gestal diabetes mellitus in preg, insulin controlled |
| DX_ICD10 | O24.415 | gestal dm in preg, controlled by oral hypoglycemic drugs |
| DX_ICD10 | O24.419 | gestal diabetes mellitus in preg, unspec control |
| DX_ICD10 | O24.42 | gestal diabetes mellitus in childbirth |
| DX_ICD10 | O24.420 | gestal diabetes mellitus in childbirth, diet controlled |
| DX_ICD10 | O24.424 | gestal diabetes mellitus in childbirth, insulin controlled |
| DX_ICD10 | O24.425 | gestal dm in childbirth, controlled by oral hypoglycemic drugs |
| DX_ICD10 | O24.429 | gestal diabetes mellitus in childbirth, unspec control |
| DX_ICD10 | O24.43 | gestal diabetes mellitus in the puerperium |
| DX_ICD10 | O24.430 | gestal diabetes mellitus in the puerperium, diet controlled |
| DX_ICD10 | O24.434 | gestal diabetes mellitus in the puerperium, insulin controlled |
| DX_ICD10 | O24.435 | gestal dm in puerperium, controlled by oral hypoglycemic drugs |
| DX_ICD10 | O24.439 | gestal diabetes mellitus in the puerperium, unspec control |
| DX_ICD10 | O24.8 | oth pre-existing dm in preg, childbirth, & the puerperium |
| DX_ICD10 | O24.81 | oth pre-existing diabetes mellitus in preg |
| DX_ICD10 | O24.811 | oth pre-existing diabetes mellitus in preg, first tri |
| DX_ICD10 | O24.812 | oth pre-existing diabetes mellitus in preg, second tri |
| DX_ICD10 | O24.813 | oth pre-existing diabetes mellitus in preg, third tri |
| DX_ICD10 | O24.819 | oth pre-existing diabetes mellitus in preg, unspec tri |
| DX_ICD10 | O24.82 | oth pre-existing diabetes mellitus in childbirth |
| DX_ICD10 | O24.83 | oth pre-existing diabetes mellitus in the puerperium |
| DX_ICD10 | O24.9 | unspec diabetes mellitus in preg, childbirth & the puerperium |
| DX_ICD10 | O24.91 | unspec diabetes mellitus in preg |
| DX_ICD10 | O24.911 | unspec diabetes mellitus in preg, first tri |
| DX_ICD10 | O24.912 | unspec diabetes mellitus in preg, second tri |
| DX_ICD10 | O24.913 | unspec diabetes mellitus in preg, third tri |
| DX_ICD10 | O24.919 | unspec diabetes mellitus in preg, unspec tri |
| DX_ICD10 | O24.92 | unspec diabetes mellitus in childbirth |
| DX_ICD10 | O24.93 | unspec diabetes mellitus in the puerperium |
| DX_ICD10 | O25 | malnutrition in preg, childbirth & the puerperium |
| DX_ICD10 | O25.1 | malnutrition in preg |
| DX_ICD10 | O25.10 | malnutrition in preg, unspec tri |
| DX_ICD10 | O25.11 | malnutrition in preg, first tri |
| DX_ICD10 | O25.12 | malnutrition in preg, second tri |
| DX_ICD10 | O25.13 | malnutrition in preg, third tri |
| DX_ICD10 | O25.2 | malnutrition in childbirth |
| DX_ICD10 | O25.3 | malnutrition in the puerperium |
| DX_ICD10 | O26 | maternal care for oth conditions predominantly related to preg |
| DX_ICD10 | O26.0 | maternal care for oth conditions predominantly related to preg |
| DX_ICD10 | O26.00 | excessive weight gain in preg, unspec tri |
| DX_ICD10 | O26.01 | excessive weight gain in preg, first tri |
| DX_ICD10 | O26.02 | excessive weight gain in preg, second tri |
| DX_ICD10 | O26.03 | excessive weight gain in preg, third tri |
| DX_ICD10 | O26.1 | low weight gain in preg |
| DX_ICD10 | O26.10 | low weight gain in preg, unspec tri |
| DX_ICD10 | O26.11 | low weight gain in preg, first tri |
| DX_ICD10 | O26.12 | low weight gain in preg, second tri |
| DX_ICD10 | O26.13 | low weight gain in preg, third tri |
| DX_ICD10 | O26.2 | preg care of habitual aborter |
| DX_ICD10 | O26.20 | preg care for patient with recurrent preg loss, unspec tri |
| DX_ICD10 | O26.21 | preg care for patient with recurrent preg loss, first tri |
| DX_ICD10 | O26.22 | preg care for patient with recurrent preg loss, second tri |
| DX_ICD10 | O26.23 | preg care for patient with recurrent preg loss, third tri |
| DX_ICD10 | O26.3 | retained intrauterine contraceptive device in preg |
| DX_ICD10 | O26.30 | retained intrauterine contraceptive device in preg, unspec tri |
| DX_ICD10 | O26.31 | retained intrauterine contraceptive device in preg, first tri |
| DX_ICD10 | O26.32 | retained intrauterine contraceptive device in preg, second tri |
| DX_ICD10 | O26.33 | retained intrauterine contraceptive device in preg, third tri |
| DX_ICD10 | O26.4 | herpes gestis |
| DX_ICD10 | O26.40 | herpes gestis, unspec tri |
| DX_ICD10 | O26.41 | herpes gestis, first tri |
| DX_ICD10 | O26.42 | herpes gestis, second tri |
| DX_ICD10 | O26.43 | herpes gestis, third tri |
| DX_ICD10 | O26.5 | maternal hypotension syndrome |
| DX_ICD10 | O26.50 | maternal hypotension syndrome, unspec tri |
| DX_ICD10 | O26.51 | maternal hypotension syndrome, first tri |
| DX_ICD10 | O26.52 | maternal hypotension syndrome, second tri |
| DX_ICD10 | O26.53 | maternal hypotension syndrome, third tri |
| DX_ICD10 | O26.6 | liver disorders in preg, childbirth & the puerperium |
| DX_ICD10 | O26.61 | liver disorders in preg |
| DX_ICD10 | O26.611 | liver & biliary tract disorders in preg, first tri |
| DX_ICD10 | O26.612 | liver & biliary tract disorders in preg, second tri |
| DX_ICD10 | O26.613 | liver & biliary tract disorders in preg, third tri |
| DX_ICD10 | O26.619 | liver & biliary tract disorders in preg, unspec tri |
| DX_ICD10 | O26.62 | liver & biliary tract disorders in childbirth |
| DX_ICD10 | O26.63 | liver & biliary tract disorders in the puerperium |
| DX_ICD10 | O26.7 | subluxation symphysis (pubis) in preg, chldbrth & puerperium |
| DX_ICD10 | O26.71 | subluxation of symphysis (pubis) in preg |
| DX_ICD10 | O26.711 | subluxation of symphysis (pubis) in preg, first tri |
| DX_ICD10 | O26.712 | subluxation of symphysis (pubis) in preg, second tri |
| DX_ICD10 | O26.713 | subluxation of symphysis (pubis) in preg, third tri |
| DX_ICD10 | O26.719 | subluxation of symphysis (pubis) in preg, unspec tri |
| DX_ICD10 | O26.72 | subluxation of symphysis (pubis) in childbirth |
| DX_ICD10 | O26.73 | subluxation of symphysis (pubis) in the puerperium |
| DX_ICD10 | O26.8 | oth specif preg related conditions |
| DX_ICD10 | O26.81 | preg related exhaustion & fatigue |
| DX_ICD10 | O26.811 | preg related exhaustion & fatigue, first tri |
| DX_ICD10 | O26.812 | preg related exhaustion & fatigue, second tri |
| DX_ICD10 | O26.813 | preg related exhaustion & fatigue, third tri |
| DX_ICD10 | O26.819 | preg related exhaustion & fatigue, unspec tri |
| DX_ICD10 | O26.82 | preg related peripheral neuritis |
| DX_ICD10 | O26.821 | preg related peripheral neuritis, first tri |
| DX_ICD10 | O26.822 | preg related peripheral neuritis, second tri |
| DX_ICD10 | O26.823 | preg related peripheral neuritis, third tri |
| DX_ICD10 | O26.829 | preg related peripheral neuritis, unspec tri |
| DX_ICD10 | O26.83 | preg related renal disease |
| DX_ICD10 | O26.831 | preg related renal disease, first tri |
| DX_ICD10 | O26.832 | preg related renal disease, second tri |
| DX_ICD10 | O26.833 | preg related renal disease, third tri |
| DX_ICD10 | O26.839 | preg related renal disease, unspec tri |
| DX_ICD10 | O26.84 | uterine size-date discrepancy complic preg |
| DX_ICD10 | O26.841 | uterine size-date discrepancy, first tri |
| DX_ICD10 | O26.842 | uterine size-date discrepancy, second tri |
| DX_ICD10 | O26.843 | uterine size-date discrepancy, third tri |
| DX_ICD10 | O26.849 | uterine size-date discrepancy, unspec tri |
| DX_ICD10 | O26.85 | spotting complic preg |
| DX_ICD10 | O26.851 | spotting complic preg, first tri |
| DX_ICD10 | O26.852 | spotting complic preg, second tri |
| DX_ICD10 | O26.853 | spotting complic preg, third tri |
| DX_ICD10 | O26.859 | spotting complic preg, unspec tri |
| DX_ICD10 | O26.86 | pruritic urticarial papules & plaques of preg (puppp) |
| DX_ICD10 | O26.87 | cervical shortening |
| DX_ICD10 | O26.872 | cervical shortening, second tri |
| DX_ICD10 | O26.873 | cervical shortening, third tri |
| DX_ICD10 | O26.879 | cervical shortening, unspec tri |
| DX_ICD10 | O26.89 | oth specif preg related conditions |
| DX_ICD10 | O26.891 | oth specif preg related conditions, first tri |
| DX_ICD10 | O26.892 | oth specif preg related conditions, second tri |
| DX_ICD10 | O26.893 | oth specif preg related conditions, third tri |
| DX_ICD10 | O26.899 | oth specif preg related conditions, unspec tri |
| DX_ICD10 | O26.9 | preg related conditions, unspec |
| DX_ICD10 | O26.90 | preg related conditions, unspec, unspec tri |
| DX_ICD10 | O26.91 | preg related conditions, unspec, first tri |
| DX_ICD10 | O26.92 | preg related conditions, unspec, second tri |
| DX_ICD10 | O26.93 | preg related conditions, unspec, third tri |
| DX_ICD10 | O28.5 | abn chromosomal & genetic finding on antenatal screening moth |
| DX_ICD10 | O28.8 | oth abnormal findings on antenatal screening of moth |
| DX_ICD10 | O28.9 | unspec abnormal findings on antenatal screening of moth |
| DX_ICD10 | O29 | complic of anesthesia during preg |
| DX_ICD10 | O29.0 | pulmonary complic of anesthesia during preg |
| DX_ICD10 | O29.01 | aspiration pneumonitis due to anesthesia during preg |
| DX_ICD10 | O29.011 | aspiration pneumonitis due to anesthesia during preg, first tri |
| DX_ICD10 | O29.012 | aspiration pneumonitis due to anesthesia during preg, second tri |
| DX_ICD10 | O29.013 | aspiration pneumonitis due to anesthesia during preg, third tri |
| DX_ICD10 | O29.019 | aspiration pneumonitis due to anesthesia during preg, unspec tri |
| DX_ICD10 | O29.02 | pressure collapse of lung due to anesthesia during preg |
| DX_ICD10 | O29.021 | pressure collapse of lung due to anesthesia during preg, first tri |
| DX_ICD10 | O29.022 | pressure collapse of lung due to anesthesia during preg, second tri |
| DX_ICD10 | O29.023 | pressure collapse of lung due to anesthesia during preg, third tri |
| DX_ICD10 | O29.029 | pressure collapse of lung due to anesthesia during preg, unspec tri |
| DX_ICD10 | O29.09 | oth pulmonary complic of anesthesia during preg |
| DX_ICD10 | O29.091 | oth pulmonary complic of anesthesia during preg, first tri |
| DX_ICD10 | O29.092 | oth pulmonary complic of anesthesia during preg, second tri |
| DX_ICD10 | O29.093 | oth pulmonary complic of anesthesia during preg, third tri |
| DX_ICD10 | O29.099 | oth pulmonary complic of anesthesia during preg, unspec tri |
| DX_ICD10 | O29.1 | cardiac complic of anesthesia during preg |
| DX_ICD10 | O29.11 | cardiac arrest due to anesthesia during preg |
| DX_ICD10 | O29.111 | cardiac arrest due to anesthesia during preg, first tri |
| DX_ICD10 | O29.112 | cardiac arrest due to anesthesia during preg, second tri |
| DX_ICD10 | O29.113 | cardiac arrest due to anesthesia during preg, third tri |
| DX_ICD10 | O29.119 | cardiac arrest due to anesthesia during preg, unspec tri |
| DX_ICD10 | O29.12 | cardiac failure due to anesthesia during preg |
| DX_ICD10 | O29.121 | cardiac failure due to anesthesia during preg, first tri |
| DX_ICD10 | O29.122 | cardiac failure due to anesthesia during preg, second tri |
| DX_ICD10 | O29.123 | cardiac failure due to anesthesia during preg, third tri |
| DX_ICD10 | O29.129 | cardiac failure due to anesthesia during preg, unspec tri |
| DX_ICD10 | O29.19 | oth cardiac complic of anesthesia during preg |
| DX_ICD10 | O29.191 | oth cardiac complic of anesthesia during preg, first tri |
| DX_ICD10 | O29.192 | oth cardiac complic of anesthesia during preg, second tri |
| DX_ICD10 | O29.193 | oth cardiac complic of anesthesia during preg, third tri |
| DX_ICD10 | O29.199 | oth cardiac complic of anesthesia during preg, unspec tri |
| DX_ICD10 | O29.2 | cns complic of anesthesia during preg |
| DX_ICD10 | O29.21 | cerebral anoxia due to anesthesia during preg |
| DX_ICD10 | O29.211 | cerebral anoxia due to anesthesia during preg, first tri |
| DX_ICD10 | O29.212 | cerebral anoxia due to anesthesia during preg, second tri |
| DX_ICD10 | O29.213 | cerebral anoxia due to anesthesia during preg, third tri |
| DX_ICD10 | O29.219 | cerebral anoxia due to anesthesia during preg, unspec tri |
| DX_ICD10 | O29.29 | oth cns complic of anesthesia during preg |
| DX_ICD10 | O29.291 | oth cns complic of anesthesia during preg, first tri |
| DX_ICD10 | O29.292 | oth cns system complic of anesthesia during preg, second tri |
| DX_ICD10 | O29.293 | oth cns complic of anesthesia during preg, third tri |
| DX_ICD10 | O29.299 | oth cns complic of anesthesia during preg, unspec tri |
| DX_ICD10 | O29.3 | toxic reaction to local anesthesia during preg |
| DX_ICD10 | O29.3X | toxic reaction to local anesthesia during preg |
| DX_ICD10 | O29.3X1 | toxic reaction to local anesthesia during preg, first tri |
| DX_ICD10 | O29.3X2 | toxic reaction to local anesthesia during preg, second tri |
| DX_ICD10 | O29.3X3 | toxic reaction to local anesthesia during preg, third tri |
| DX_ICD10 | O29.3X9 | toxic reaction to local anesthesia during preg, unspec tri |
| DX_ICD10 | O29.4 | spinal & epidural anesthesia induced headache during preg |
| DX_ICD10 | O29.40 | spinal & epidural anesthesia induced headache during preg, unspec tri |
| DX_ICD10 | O29.41 | spinal & epidural anesthesia induced headache during preg, first tri |
| DX_ICD10 | O29.42 | spinal & epidural anesthesia induced headache during preg, second tri |
| DX_ICD10 | O29.43 | spinal & epidural anesthesia induced headache during preg, third tri |
| DX_ICD10 | O29.5 | oth complic of spinal & epidural anesthesia during preg |
| DX_ICD10 | O29.5X | oth complic of spinal & epidural anesthesia during preg |
| DX_ICD10 | O29.5X1 | oth complic of spinal & epidural anesthesia during preg, first tri |
| DX_ICD10 | O29.5X2 | oth complic of spinal & epidural anesthesia during preg, second tri |
| DX_ICD10 | O29.5X3 | oth complic of spinal & epidural anesthesia during preg, third tri |
| DX_ICD10 | O29.5X9 | oth complic of spinal & epidural anesthesia during preg, unspec tri |
| DX_ICD10 | O29.6 | failed or difficult intubation for anesthesia during preg |
| DX_ICD10 | O29.60 | failed or difficult intubation for anesthesia during preg, unspec tri |
| DX_ICD10 | O29.61 | failed or difficult intubation for anesthesia during preg, first tri |
| DX_ICD10 | O29.62 | failed or difficult intubation for anesthesia during preg, second tri |
| DX_ICD10 | O29.63 | failed or difficult intubation for anesthesia during preg, third tri |
| DX_ICD10 | O29.8 | oth complic of anesthesia during preg |
| DX_ICD10 | O29.8X | oth complic of anesthesia during preg |
| DX_ICD10 | O29.8X1 | oth complic of anesthesia during preg, first tri |
| DX_ICD10 | O29.8X2 | oth complic of anesthesia during preg, second tri |
| DX_ICD10 | O29.8X3 | oth complic of anesthesia during preg, third tri |
| DX_ICD10 | O29.8X9 | oth complic of anesthesia during preg, unspec tri |
| DX_ICD10 | O29.9 | unspec complication of anesthesia during preg |
| DX_ICD10 | O29.90 | unspec complication of anesthesia during preg, unspec tri |
| DX_ICD10 | O29.91 | unspec complication of anesthesia during preg, first tri |
| DX_ICD10 | O29.92 | unspec complication of anesthesia during preg, second tri |
| DX_ICD10 | O29.93 | unspec complication of anesthesia during preg, third tri |
| DX_ICD10 | O30 | multi gest |
| DX_ICD10 | O30.0 | twin preg |
| DX_ICD10 | O30.00 | win preg, unspec |
| DX_ICD10 | O30.001 | twin preg, unspec # plcnta & unspec # amniotic sacs, first tri |
| DX_ICD10 | O30.002 | twin preg, unspec # plcnta & unspec # amniotic sacs, second tri |
| DX_ICD10 | O30.003 | twin preg, unspec # plcnta & unspec # amniotic sacs, third tri |
| DX_ICD10 | O30.009 | twin preg, unspec # plcnta & unspec # amniotic sacs, unspec tri |
| DX_ICD10 | O30.01 | twin preg, monoamniotic/monochorionic |
| DX_ICD10 | O30.011 | twin preg, monochorionic/monoamniotic, first tri |
| DX_ICD10 | O30.012 | twin preg, monochorionic/monoamniotic, second tri |
| DX_ICD10 | O30.013 | twin preg, monochorionic/monoamniotic, third tri |
| DX_ICD10 | O30.019 | twin preg, monochorionic/monoamniotic, unspec tri |
| DX_ICD10 | O30.02 | conjoined twins |
| DX_ICD10 | O30.021 | conjoined twin preg, first tri |
| DX_ICD10 | O30.022 | conjoined twin preg, second tri |
| DX_ICD10 | O30.023 | conjoined twin preg, third tri |
| DX_ICD10 | O30.029 | conjoined twin preg, unspec tri |
| DX_ICD10 | O30.03 | twin preg, monochorionic/diamniotic |
| DX_ICD10 | O30.031 | twin preg, monochorionic/diamniotic, first tri |
| DX_ICD10 | O30.032 | twin preg, monochorionic/diamniotic, second tri |
| DX_ICD10 | O30.033 | twin preg, monochorionic/diamniotic, third tri |
| DX_ICD10 | O30.039 | twin preg, monochorionic/diamniotic, unspec tri |
| DX_ICD10 | O30.04 | twin preg, dichorionic/diamniotic |
| DX_ICD10 | O30.041 | twin preg, dichorionic/diamniotic, first tri |
| DX_ICD10 | O30.042 | twin preg, dichorionic/diamniotic, second tri |
| DX_ICD10 | O30.043 | twin preg, dichorionic/diamniotic, third tri |
| DX_ICD10 | O30.049 | twin preg, dichorionic/diamniotic, unspec tri |
| DX_ICD10 | O30.09 | oth twin preg |
| DX_ICD10 | O30.091 | twin preg, unable to determine # plcnta & # amniotic sacs, first tri |
| DX_ICD10 | O30.092 | twin preg, unable to determine # plcnta & # amniotic sacs, second tri |
| DX_ICD10 | O30.093 | twin preg, unable to determine # plcnta & # amniotic sacs, third tri |
| DX_ICD10 | O30.099 | twin preg, unable to determine # plcnta & # amniotic sacs, unspec tri |
| DX_ICD10 | O30.1 | trip preg |
| DX_ICD10 | O30.10 | trip preg, unspec tri |
| DX_ICD10 | O30.101 | trip preg, unspec # plcnta & unspec # amniotic sacs, first tri |
| DX_ICD10 | O30.102 | trip preg, unspec # plcnta & unspec # amniotic sacs, second tri |
| DX_ICD10 | O30.103 | trip preg, unspec # plcnta & unspec # amniotic sacs, third tri |
| DX_ICD10 | O30.109 | trip preg, unspec # plcnta & unspec # amniotic sacs, unspec tri |
| DX_ICD10 | O30.11 | trip preg, first tri |
| DX_ICD10 | O30.111 | trip preg with two or more monochorionic fetuses, first tri |
| DX_ICD10 | O30.112 | trip preg with two or more monochorionic fetuses, second tri |
| DX_ICD10 | O30.113 | trip preg with two or more monochorionic fetuses, third tri |
| DX_ICD10 | O30.119 | trip preg with two or more monochorionic fetuses, unspec tri |
| DX_ICD10 | O30.12 | trip preg, second tri |
| DX_ICD10 | O30.121 | trip preg with two or more monoamniotic fetuses, first tri |
| DX_ICD10 | O30.122 | trip preg with two or more monoamniotic fetuses, second tri |
| DX_ICD10 | O30.123 | trip preg with two or more monoamniotic fetuses, third tri |
| DX_ICD10 | O30.129 | trip preg with two or more monoamniotic fetuses, unspec tri |
| DX_ICD10 | O30.131 | trip preg, trichorionic/triamniotic, first tri |
| DX_ICD10 | O30.132 | trip preg, trichorionic/triamniotic, second tri |
| DX_ICD10 | O30.133 | trip preg, trichorionic/triamniotic, third tri |
| DX_ICD10 | O30.139 | trip preg, trichorionic/triamniotic, unspec tri |
| DX_ICD10 | O30.19 | trip preg, unable to dtrm num plcnta & amnio sacs |
| DX_ICD10 | O30.191 | trip preg, unable to dtrm # plcnta & # amniotic sacs, first tri |
| DX_ICD10 | O30.192 | trip preg, unable to dtrm # plcnta & # amniotic sacs, second tri |
| DX_ICD10 | O30.193 | trip preg, unable to dtrm # plcnta & # amniotic sacs, third tri |
| DX_ICD10 | O30.199 | trip preg, unable dtrm # plcnta & # amniotic sacs, unspec tri |
| DX_ICD10 | O30.2 | quad preg |
| DX_ICD10 | O30.20 | quad preg, unspec tri |
| DX_ICD10 | O30.201 | quad preg, unspec # plcnta & unspec # amnio sacs, first tri |
| DX_ICD10 | O30.202 | quad preg, unspec # plcnta & unspec # amnio sacs, second tri |
| DX_ICD10 | O30.203 | quad preg, unspec # plcnta & unspec # amniotic sacs, third tri |
| DX_ICD10 | O30.209 | quad preg, unspec # plcnta & # amniotic sacs, unspec tri |
| DX_ICD10 | O30.21 | quad preg, first tri |
| DX_ICD10 | O30.211 | quad preg with 2+ monochorionic fetuses, first tri |
| DX_ICD10 | O30.212 | quad preg with 2+ monochorionic fetuses, second tri |
| DX_ICD10 | O30.213 | quad preg with 2+ monochorionic fetuses, third tri |
| DX_ICD10 | O30.219 | quad preg with 2+ monochorionic fetuses, unspec tri |
| DX_ICD10 | O30.22 | quad preg, second tri |
| DX_ICD10 | O30.221 | quad preg with 2+ monoamniotic fetuses, first tri |
| DX_ICD10 | O30.222 | quad preg with 2+ monoamniotic fetuses, second tri |
| DX_ICD10 | O30.223 | quad preg with 2+ monoamniotic fetuses, third tri |
| DX_ICD10 | O30.229 | quad preg with 2+ monoamniotic fetuses, unspec tri |
| DX_ICD10 | O30.231 | quad preg, quadrachorionic/quadra-amniotic, first tri |
| DX_ICD10 | O30.232 | quad preg, quadrachorionic/quadra-amniotic, second tri |
| DX_ICD10 | O30.233 | quad preg, quadrachorionic/quadra-amniotic, third tri |
| DX_ICD10 | O30.239 | quad preg, quadrachorionic/quadra-amniotic, unspec tri |
| DX_ICD10 | O30.29 | quad preg, unable to dtrm num plcnta & amnio sacs |
| DX_ICD10 | O30.291 | quad preg, unable to dtrm # plcnta & # amniotic sacs, first tri |
| DX_ICD10 | O30.292 | quad preg, unable to dtrm # plcnta & # amniotic sacs, second tri |
| DX_ICD10 | O30.293 | quad preg, unable to dtrm # plcnta & # amniotic sacs, third tri |
| DX_ICD10 | O30.299 | quad preg, unable dtrm # plcnta & # amniotic sacs, unspecif tri |
| DX_ICD10 | O30.8 | oth multi gest |
| DX_ICD10 | O30.80 | oth multi gest, unspeci tri |
| DX_ICD10 | O30.801 | oth spec multi gest, unspec # plcnta & unspec # amnio sacs, first tri |
| DX_ICD10 | O30.802 | oth spec multi gest, unspec # plcnta & unspec # amnio sacs, second tri |
| DX_ICD10 | O30.803 | oth spec multi gest, unspec # plcnta & unspec # amnio sacs, third tri |
| DX_ICD10 | O30.809 | oth spec multi gest, unspec # plcnt & unspec # amnio sacs, unspec tri |
| DX_ICD10 | O30.81 | oth multi gest, first tri |
| DX_ICD10 | O30.811 | oth spec multi gest with 2+ monochorionic fetuses, first tri |
| DX_ICD10 | O30.812 | oth spec multi gest with 2+ monochorionic fetuses, second tri |
| DX_ICD10 | O30.813 | oth spec multi gest with 2+ monochorionic fetuses, third tri |
| DX_ICD10 | O30.819 | oth spec multi gest with 2+ monochorionic fetuses, unspec tri |
| DX_ICD10 | O30.82 | oth multi gest, second tri |
| DX_ICD10 | O30.821 | oth spec multi gest with 2+ monoamniotic fetuses, first tri |
| DX_ICD10 | O30.822 | oth spec multi gest with 2+ monoamniotic fetuses, second tri |
| DX_ICD10 | O30.823 | oth spec multi gest with 2+ monoamniotic fetuses, third tri |
| DX_ICD10 | O30.829 | oth spec multi gest with 2+ monoamniotic fetuses, unspec tri |
| DX_ICD10 | O30.831 | oth spec multi gest, # chorions & amnions equal # fetuses, first tri |
| DX_ICD10 | O30.832 | oth spec multi gest, # chorions & amnions equal # fetuses, second tri |
| DX_ICD10 | O30.833 | oth spec multi gest, # chorions & amnions equal # fetuses, third tri |
| DX_ICD10 | O30.839 | oth spec multi gest, # chorions & amnions equal # fetuses, unspec tri |
| DX_ICD10 | O30.89 | oth multi gest, unable to dtrm num plcnta & amnio sacs |
| DX_ICD10 | O30.891 | oth spec multi gest, unable to dtrm # plcnta & # amniotic sacs, first tri |
| DX_ICD10 | O30.892 | oth spec multi gest, unable to dtrm # plcnta & # amnio sacs, second tri |
| DX_ICD10 | O30.893 | oth spec multi gest, unable to dtrm # plcnta & # amnio sacs, third tri |
| DX_ICD10 | O30.899 | oth spec multi gest, unable dtrm # plcnta & # amniotic sacs, unspec tri |
| DX_ICD10 | O30.9 | multi gest, unspec |
| DX_ICD10 | O30.90 | multi gest, unspec, unspec tri |
| DX_ICD10 | O30.91 | multi gest, unspec, first tri |
| DX_ICD10 | O30.92 | multi gest, unspec, second tri |
| DX_ICD10 | O30.93 | multi gest, unspec, third tri |
| DX_ICD10 | O31 | complic specific to multi gest |
| DX_ICD10 | O31.0 | papyraceous fetus |
| DX_ICD10 | O31.00 | papyraceous fetus, unspec tri |
| DX_ICD10 | O31.00X0 | papyraceous fetus, unspec tri, na or unspec |
| DX_ICD10 | O31.00X1 | papyraceous fetus, unspec tri, fetus 1 |
| DX_ICD10 | O31.00X2 | papyraceous fetus, unspec tri, fetus 2 |
| DX_ICD10 | O31.00X3 | papyraceous fetus, unspec tri, fetus 3 |
| DX_ICD10 | O31.00X4 | papyraceous fetus, unspec tri, fetus 4 |
| DX_ICD10 | O31.00X5 | papyraceous fetus, unspec tri, fetus 5 |
| DX_ICD10 | O31.00X9 | papyraceous fetus, unspec tri, oth fetus |
| DX_ICD10 | O31.01 | papyraceous fetus, first tri |
| DX_ICD10 | O31.01X0 | papyraceous fetus, first tri, na or unspec |
| DX_ICD10 | O31.01X1 | papyraceous fetus, first tri, fetus 1 |
| DX_ICD10 | O31.01X2 | papyraceous fetus, first tri, fetus 2 |
| DX_ICD10 | O31.01X3 | papyraceous fetus, first tri, fetus 3 |
| DX_ICD10 | O31.01X4 | papyraceous fetus, first tri, fetus 4 |
| DX_ICD10 | O31.01X5 | papyraceous fetus, first tri, fetus 5 |
| DX_ICD10 | O31.01X9 | papyraceous fetus, first tri, oth fetus |
| DX_ICD10 | O31.02 | papyraceous fetus, second tri |
| DX_ICD10 | O31.02X0 | papyraceous fetus, second tri, na or unspec |
| DX_ICD10 | O31.02X1 | papyraceous fetus, second tri, fetus 1 |
| DX_ICD10 | O31.02X2 | papyraceous fetus, second tri, fetus 2 |
| DX_ICD10 | O31.02X3 | papyraceous fetus, second tri, fetus 3 |
| DX_ICD10 | O31.02X4 | papyraceous fetus, second tri, fetus 4 |
| DX_ICD10 | O31.02X5 | papyraceous fetus, second tri, fetus 5 |
| DX_ICD10 | O31.02X9 | papyraceous fetus, second tri, oth fetus |
| DX_ICD10 | O31.03 | papyraceous fetus, third tri |
| DX_ICD10 | O31.03X0 | papyraceous fetus, third tri, na or unspec |
| DX_ICD10 | O31.03X1 | papyraceous fetus, third tri, fetus 1 |
| DX_ICD10 | O31.03X2 | papyraceous fetus, third tri, fetus 2 |
| DX_ICD10 | O31.03X3 | papyraceous fetus, third tri, fetus 3 |
| DX_ICD10 | O31.03X4 | papyraceous fetus, third tri, fetus 4 |
| DX_ICD10 | O31.03X5 | papyraceous fetus, third tri, fetus 5 |
| DX_ICD10 | O31.03X9 | papyraceous fetus, third tri, oth fetus |
| DX_ICD10 | O31.1 | continuing preg after SAB of 1+ fetus |
| DX_ICD10 | O31.10 | continuing preg after SAB of 1+ fetus, unspec tri |
| DX_ICD10 | O31.10X0 | continuing preg after SAB of 1+ fetus, unspec tri , na or unspec |
| DX_ICD10 | O31.10X1 | continuing preg after SAB of 1+ fetus, unspec tri , fetus 1 |
| DX_ICD10 | O31.10X2 | continuing preg after SAB of 1+ fetus, unspec tri , fetus 2 |
| DX_ICD10 | O31.10X3 | continuing preg after SAB of 1+ fetus, unspec tri , fetus 3 |
| DX_ICD10 | O31.10X4 | continuing preg after SAB of 1+ fetus, unspec tri , fetus 4 |
| DX_ICD10 | O31.10X5 | continuing preg after SAB of 1+ fetus, unspec tri , fetus 5 |
| DX_ICD10 | O31.10X9 | continuing preg after SAB of 1+ fetus, unspec tri , oth fetus |
| DX_ICD10 | O31.11 | continuing preg after SAB of 1+ fetus, first tri |
| DX_ICD10 | O31.11X0 | continuing preg after SAB of 1+ fetus, first tri , na or unspec |
| DX_ICD10 | O31.11X1 | continuing preg after SAB of 1+ fetus, first tri , fetus 1 |
| DX_ICD10 | O31.11X2 | continuing preg after SAB of 1+ fetus, first tri , fetus 2 |
| DX_ICD10 | O31.11X3 | continuing preg after SAB of 1+ fetus, first tri, fetus 3 |
| DX_ICD10 | O31.11X4 | continuing preg after SAB of 1+ fetus, first tri, fetus 4 |
| DX_ICD10 | O31.11X5 | continuing preg after SAB of 1+ fetus, first tri, fetus 5 |
| DX_ICD10 | O31.11X9 | continuing preg after SAB of 1+ fetus, first tri, oth fetus |
| DX_ICD10 | O31.12 | continuing preg after SAB of 1+ fetus, second tri |
| DX_ICD10 | O31.12X0 | contin preg after SAB of 1+ fetus, second tri, na or unspec |
| DX_ICD10 | O31.12X1 | continuing preg after SAB of 1+ fetus, second tri, fetus 1 |
| DX_ICD10 | O31.12X2 | continuing preg after SAB of 1+ fetus, second tri, fetus 2 |
| DX_ICD10 | O31.12X3 | continuing preg after SAB of 1+ fetus, second tri, fetus 3 |
| DX_ICD10 | O31.12X4 | continuing preg after SAB of 1+ fetus, second tri, fetus 4 |
| DX_ICD10 | O31.12X5 | continuing preg after SAB of 1+ fetus, second tri, fetus 5 |
| DX_ICD10 | O31.12X9 | continuing preg after SAB of 1+ fetus, second tri, oth fetus |
| DX_ICD10 | O31.13 | continuing preg after SAB of 1+ fetus, third tri |
| DX_ICD10 | O31.13X0 | continuing preg after SAB of 1+ fetus, third tri, na or unspec |
| DX_ICD10 | O31.13X1 | continuing preg after SAB of 1+ fetus, third tri, fetus 1 |
| DX_ICD10 | O31.13X2 | continuing preg after SAB of 1+ fetus, third tri, fetus 2 |
| DX_ICD10 | O31.13X3 | continuing preg after SAB of 1+ fetus, third tri, fetus 3 |
| DX_ICD10 | O31.13X4 | continuing preg after SAB of 1+ fetus, third tri, fetus 4 |
| DX_ICD10 | O31.13X5 | continuing preg after SAB of 1+ fetus, third tri, fetus 5 |
| DX_ICD10 | O31.13X9 | continuing preg after SAB of 1+ fetus, third tri, oth fetus |
| DX_ICD10 | O31.2 | continuing preg after intrauterine death of 1+ fetus |
| DX_ICD10 | O31.20 | continuing preg after intrauterine death of 1+ fetus, unspec tri |
| DX_ICD10 | O31.20X0 | contin preg after intraut death of 1+ fetus, unspec tri, na or unspec |
| DX_ICD10 | O31.20X1 | contin preg after intraut death of 1+ fetus, unspec tri, fetus 1 |
| DX_ICD10 | O31.20X2 | contin preg after intraut death of 1+ fetus, unspec tri, fetus 2 |
| DX_ICD10 | O31.20X3 | contin preg after intraut death of 1+ fetus, unspec tri, fetus 3 |
| DX_ICD10 | O31.20X4 | contin preg after intraut death of 1+ fetus, unspec tri, fetus 4 |
| DX_ICD10 | O31.20X5 | contin preg after intraut death of 1+ fetus, unspec tri, fetus 5 |
| DX_ICD10 | O31.20X9 | contin preg after intrauterine death of one+ fetus, unspec tri, oth fetus |
| DX_ICD10 | O31.21 | contin preg after intrauterine death of one+ fetus, first tri |
| DX_ICD10 | O31.21X0 | contin preg after intrauterine death of one+ fetus, first tri, na or unspec |
| DX_ICD10 | O31.21X1 | contin preg after intraut death of 1+ fetus, first tri, fetus 1 |
| DX_ICD10 | O31.21X2 | contin preg after intraut death of 1+ fetus, first tri, fetus 2 |
| DX_ICD10 | O31.21X3 | contin preg after intraut death of 1+ fetus, first tri, fetus 3 |
| DX_ICD10 | O31.21X4 | contin preg after intraut death of 1+ fetus, first tri, fetus 4 |
| DX_ICD10 | O31.21X5 | contin preg after intraut death of 1+ fetus, first tri, fetus 5 |
| DX_ICD10 | O31.21X9 | contin preg after intraut death of 1+ fetus, first tri, oth fetus |
| DX_ICD10 | O31.22 | contin preg after intraut death of 1+ fetus, second tri |
| DX_ICD10 | O31.22X0 | contin preg after intraut death of one+ fetus, second tri, na or unspec |
| DX_ICD10 | O31.22X1 | contin preg after intraut death of 1+ fetus, second tri, fetus 1 |
| DX_ICD10 | O31.22X2 | contin preg after intraut death of 1+ fetus, second tri, fetus 2 |
| DX_ICD10 | O31.22X3 | contin preg after intraut death of 1+ fetus, second tri, fetus 3 |
| DX_ICD10 | O31.22X4 | contin preg after intraut death of 1+ fetus, second tri, fetus 4 |
| DX_ICD10 | O31.22X5 | contin preg after intraut death of 1+ fetus, second tri, fetus 5 |
| DX_ICD10 | O31.22X9 | contin preg after intraut death of one+ fetus, second tri, oth fetus |
| DX_ICD10 | O31.23 | contin preg after intraut death of 1+ fetus, third tri |
| DX_ICD10 | O31.23X0 | contin preg after intraut death of one+ fetus, third tri, na or unspec |
| DX_ICD10 | O31.23X1 | contin preg after intraut death of 1+ fetus, third tri, fetus 1 |
| DX_ICD10 | O31.23X2 | contin preg after intraut death of 1+ fetus, third tri, fetus 2 |
| DX_ICD10 | O31.23X3 | contin preg after intraut death of 1+ fetus, third tri, fetus 3 |
| DX_ICD10 | O31.23X4 | contin preg after intraut death of 1+ fetus, third tri, fetus 4 |
| DX_ICD10 | O31.23X5 | contin preg after intraut death of 1+ fetus, third tri, fetus 5 |
| DX_ICD10 | O31.23X9 | contin preg after intraut death of 1+fetus, third tri, oth fetus |
| DX_ICD10 | O31.3 | contin preg after elect fetal reduct of one fetus |
| DX_ICD10 | O31.30 | contin preg after elect fetal reduct of 1+ fetus, unspec tri |
| DX_ICD10 | O31.30X0 | contin preg after elect fetal reduct of 1+ fetus, unspec tri, na or unspec |
| DX_ICD10 | O31.30X1 | contin preg after elect fetal reduct of 1+ fetus, unspec tri, fetus 1 |
| DX_ICD10 | O31.30X2 | contin preg after elect fetal reduct of 1+ fetus, unspec tri, fetus 2 |
| DX_ICD10 | O31.30X3 | contin preg after elect fetal reduct of 1+ fetus, unspec tri, fetus 3 |
| DX_ICD10 | O31.30X4 | contin preg after elect fetal reduct of 1+ fetus, unspec tri, fetus 4 |
| DX_ICD10 | O31.30X5 | contin preg after elect fetal reduct of 1+ fetus, unspec tri, fetus 5 |
| DX_ICD10 | O31.30X9 | contin preg after elect fetal reduct of 1+ fetus, unspec tri, oth fetus |
| DX_ICD10 | O31.31 | contin preg after elect fetal reduct of 1+ fetus, first tri |
| DX_ICD10 | O31.31X0 | contin preg after elect fetal reduct of 1+ fetus, first tri, na or unspec |
| DX_ICD10 | O31.31X1 | contin preg after elect fetal reduct of 1+ fetus, first tri, fetus 1 |
| DX_ICD10 | O31.31X2 | contin preg after elect fetal reduct of 1+ fetus, first tri, fetus 2 |
| DX_ICD10 | O31.31X3 | contin preg after elect fetal reduct of 1+ fetus, first tri, fetus 3 |
| DX_ICD10 | O31.31X4 | contin preg after elect fetal reduct of 1+ fetus, first tri, fetus 4 |
| DX_ICD10 | O31.31X5 | contin preg after elect fetal reduct of 1+ fetus, first tri, fetus 5 |
| DX_ICD10 | O31.31X9 | contin preg after elect fetal reduct of 1+ fetus, first tri, oth fetus |
| DX_ICD10 | O31.32 | contin preg after elect fetal reduct of 1+ fetus, second tri |
| DX_ICD10 | O31.32X0 | contin preg after elect fetal reduct of 1+ fetus, second tri, na or unspec |
| DX_ICD10 | O31.32X1 | contin preg after elect fetal reduct of 1+ fetus, second tri, fetus 1 |
| DX_ICD10 | O31.32X2 | contin preg after elect fetal reduct of 1+ fetus, second tri, fetus 2 |
| DX_ICD10 | O31.32X3 | contin preg after elect fetal reduct of 1+ fetus, second tri, fetus 3 |
| DX_ICD10 | O31.32X4 | contin preg after elect fetal reduct of 1+ fetus, second tri, fetus 4 |
| DX_ICD10 | O31.32X5 | contin preg after elect fetal reduct of 1+ fetus, second tri, fetus 5 |
| DX_ICD10 | O31.32X9 | contin preg after elect fetal reduct of 1+ fetus, second tri, oth fetus |
| DX_ICD10 | O31.33 | contin preg after elect fetal reduct of 1+ fetus, third tri |
| DX_ICD10 | O31.33X0 | contin preg after elect fetal reduct of 1+ fetus, third tri, na or unspec |
| DX_ICD10 | O31.33X1 | contin preg after elect fetal reduct of 1+ fetus, third tri, fetus 1 |
| DX_ICD10 | O31.33X2 | contin preg after elect fetal reduct of 1+ fetus, third tri, fetus 2 |
| DX_ICD10 | O31.33X3 | contin preg after elect fetal reduct of 1+ fetus, third tri, fetus 3 |
| DX_ICD10 | O31.33X4 | contin preg after elect fetal reduct of 1+ fetus, third tri, fetus 4 |
| DX_ICD10 | O31.33X5 | contin preg after elect fetal reduct of 1+ fetus, third tri, fetus 5 |
| DX_ICD10 | O31.33X9 | contin preg after elect fetal reduct of 1+ fetus, third tri, oth fetus |
| DX_ICD10 | O31.8 | oth complic specific to multi gest |
| DX_ICD10 | O31.8X | oth complic specific to multi gest |
| DX_ICD10 | O31.8X1 | oth complic specific to multi gest, first tri |
| DX_ICD10 | O31.8X10 | oth complic specific to multi gest, first tri, na or unspec |
| DX_ICD10 | O31.8X11 | oth complic specific to multi gest, first tri, fetus 1 |
| DX_ICD10 | O31.8X12 | oth complic specific to multi gest, first tri, fetus 2 |
| DX_ICD10 | O31.8X13 | oth complic specific to multi gest, first tri, fetus 3 |
| DX_ICD10 | O31.8X14 | oth complic specific to multi gest, first tri, fetus 4 |
| DX_ICD10 | O31.8X15 | oth complic specific to multi gest, first tri, fetus 5 |
| DX_ICD10 | O31.8X19 | oth complic specific to multi gest, first tri, oth fetus |
| DX_ICD10 | O31.8X2 | oth complic specific to multi gest, second tri |
| DX_ICD10 | O31.8X20 | oth complic specific to multi gest, second tri, na or unspec |
| DX_ICD10 | O31.8X21 | oth complic specific to multi gest, second tri, fetus 1 |
| DX_ICD10 | O31.8X22 | oth complic specific to multi gest, second tri, fetus 2 |
| DX_ICD10 | O31.8X23 | oth complic specific to multi gest, second tri, fetus 3 |
| DX_ICD10 | O31.8X24 | oth complic specific to multi gest, second tri, fetus 4 |
| DX_ICD10 | O31.8X25 | oth complic specific to multi gest, second tri, fetus 5 |
| DX_ICD10 | O31.8X29 | oth complic specific to multi gest, second tri, oth fetus |
| DX_ICD10 | O31.8X3 | oth complic specific to multi gest, third tri |
| DX_ICD10 | O31.8X30 | oth complic specific to multi gest, third tri, na or unspec |
| DX_ICD10 | O31.8X31 | oth complic specific to multi gest, third tri, fetus 1 |
| DX_ICD10 | O31.8X32 | oth complic specific to multi gest, third tri, fetus 2 |
| DX_ICD10 | O31.8X33 | oth complic specific to multi gest, third tri, fetus 3 |
| DX_ICD10 | O31.8X34 | oth complic specific to multi gest, third tri, fetus 4 |
| DX_ICD10 | O31.8X35 | oth complic specific to multi gest, third tri, fetus 5 |
| DX_ICD10 | O31.8X39 | oth complic specific to multi gest, third tri, oth fetus |
| DX_ICD10 | O31.8X9 | oth complic specific to multi gest, unspec tri |
| DX_ICD10 | O31.8X90 | oth complic specific to multi gest, unspec tri, na or unspec |
| DX_ICD10 | O31.8X91 | oth complic specific to multi gest, unspec tri, fetus 1 |
| DX_ICD10 | O31.8X92 | oth complic specific to multi gest, unspec tri, fetus 2 |
| DX_ICD10 | O31.8X93 | oth complic specific to multi gest, unspec tri, fetus 3 |
| DX_ICD10 | O31.8X94 | oth complic specific to multi gest, unspec tri, fetus 4 |
| DX_ICD10 | O31.8X95 | oth complic specific to multi gest, unspec tri, fetus 5 |
| DX_ICD10 | O31.8X99 | oth complic specific to multi gest, unspec tri, oth fetus |
| DX_ICD10 | O32 | maternal care for malpresentation of fetus |
| DX_ICD10 | O32.0 | maternal care for unstable lie |
| DX_ICD10 | O32.0XX0 | maternal care for unstable lie, na or unspec |
| DX_ICD10 | O32.0XX1 | maternal care for unstable lie, fetus 1 |
| DX_ICD10 | O32.0XX2 | maternal care for unstable lie, fetus 2 |
| DX_ICD10 | O32.0XX3 | maternal care for unstable lie, fetus 3 |
| DX_ICD10 | O32.0XX4 | maternal care for unstable lie, fetus 4 |
| DX_ICD10 | O32.0XX5 | maternal care for unstable lie, fetus 5 |
| DX_ICD10 | O32.0XX9 | maternal care for unstable lie, oth fetus |
| DX_ICD10 | O32.1 | maternal care for breech presentation |
| DX_ICD10 | O32.1XX0 | maternal care for breech presentation, na or unspec |
| DX_ICD10 | O32.1XX1 | maternal care for breech presentation, fetus 1 |
| DX_ICD10 | O32.1XX2 | maternal care for breech presentation, fetus 2 |
| DX_ICD10 | O32.1XX3 | maternal care for breech presentation, fetus 3 |
| DX_ICD10 | O32.1XX4 | maternal care for breech presentation, fetus 4 |
| DX_ICD10 | O32.1XX5 | maternal care for breech presentation, fetus 5 |
| DX_ICD10 | O32.1XX9 | maternal care for breech presentation, oth fetus |
| DX_ICD10 | O32.2 | maternal care for transverse & oblique lie |
| DX_ICD10 | O32.2XX0 | maternal care for transverse & oblique lie, na or unspec |
| DX_ICD10 | O32.2XX1 | maternal care for transverse & oblique lie, fetus 1 |
| DX_ICD10 | O32.2XX2 | maternal care for transverse & oblique lie, fetus 2 |
| DX_ICD10 | O32.2XX3 | maternal care for transverse & oblique lie, fetus 3 |
| DX_ICD10 | O32.2XX4 | maternal care for transverse & oblique lie, fetus 4 |
| DX_ICD10 | O32.2XX5 | maternal care for transverse & oblique lie, fetus 5 |
| DX_ICD10 | O32.2XX9 | maternal care for transverse & oblique lie, oth fetus |
| DX_ICD10 | O32.3 | maternal care for face, brow & chin presentation |
| DX_ICD10 | O32.3XX0 | maternal care for face, brow & chin presentation, na or unspec |
| DX_ICD10 | O32.3XX1 | maternal care for face, brow & chin presentation, fetus 1 |
| DX_ICD10 | O32.3XX2 | maternal care for face, brow & chin presentation, fetus 2 |
| DX_ICD10 | O32.3XX3 | maternal care for face, brow & chin presentation, fetus 3 |
| DX_ICD10 | O32.3XX4 | maternal care for face, brow & chin presentation, fetus 4 |
| DX_ICD10 | O32.3XX5 | maternal care for face, brow & chin presentation, fetus 5 |
| DX_ICD10 | O32.3XX9 | maternal care for face, brow & chin presentation, oth fetus |
| DX_ICD10 | O32.4 | maternal care for high head at term |
| DX_ICD10 | O32.4XX0 | maternal care for high head at term, na or unspec |
| DX_ICD10 | O32.4XX1 | maternal care for high head at term, fetus 1 |
| DX_ICD10 | O32.4XX2 | maternal care for high head at term, fetus 2 |
| DX_ICD10 | O32.4XX3 | maternal care for high head at term, fetus 3 |
| DX_ICD10 | O32.4XX4 | maternal care for high head at term, fetus 4 |
| DX_ICD10 | O32.4XX5 | maternal care for high head at term, fetus 5 |
| DX_ICD10 | O32.4XX9 | maternal care for high head at term, oth fetus |
| DX_ICD10 | O32.6 | maternal care for compound presentation |
| DX_ICD10 | O32.6XX0 | maternal care for compound presentation, na or unspec |
| DX_ICD10 | O32.6XX1 | maternal care for compound presentation, fetus 1 |
| DX_ICD10 | O32.6XX2 | maternal care for compound presentation, fetus 2 |
| DX_ICD10 | O32.6XX3 | maternal care for compound presentation, fetus 3 |
| DX_ICD10 | O32.6XX4 | maternal care for compound presentation, fetus 4 |
| DX_ICD10 | O32.6XX5 | maternal care for compound presentation, fetus 5 |
| DX_ICD10 | O32.6XX9 | maternal care for compound presentation, oth fetus |
| DX_ICD10 | O32.8 | maternal care for oth malpresentation of fetus |
| DX_ICD10 | O32.8XX0 | maternal care for oth malpresentation of fetus, na or unspec |
| DX_ICD10 | O32.8XX1 | maternal care for oth malpresentation of fetus, fetus 1 |
| DX_ICD10 | O32.8XX2 | maternal care for oth malpresentation of fetus, fetus 2 |
| DX_ICD10 | O32.8XX3 | maternal care for oth malpresentation of fetus, fetus 3 |
| DX_ICD10 | O32.8XX4 | maternal care for oth malpresentation of fetus, fetus 4 |
| DX_ICD10 | O32.8XX5 | maternal care for oth malpresentation of fetus, fetus 5 |
| DX_ICD10 | O32.8XX9 | maternal care for oth malpresentation of fetus, oth fetus |
| DX_ICD10 | O32.9 | maternal care for malpresentation of fetus, unspec |
| DX_ICD10 | O32.9XX0 | maternal care for malpresentation of fetus, unspec, na or unspec |
| DX_ICD10 | O32.9XX1 | maternal care for malpresentation of fetus, unspec, fetus 1 |
| DX_ICD10 | O32.9XX2 | maternal care for malpresentation of fetus, unspec, fetus 2 |
| DX_ICD10 | O32.9XX3 | maternal care for malpresentation of fetus, unspec, fetus 3 |
| DX_ICD10 | O32.9XX4 | maternal care for malpresentation of fetus, unspec, fetus 4 |
| DX_ICD10 | O32.9XX5 | maternal care for malpresentation of fetus, unspec, fetus 5 |
| DX_ICD10 | O32.9XX9 | maternal care for malpresentation of fetus, unspec, oth fetus |
| DX_ICD10 | O33 | maternal care for disproportion |
| DX_ICD10 | O33.0 | maternal care for disproportion due to deformity of mat pelvic bones |
| DX_ICD10 | O33.1 | maternal care for disproportion due to generally contracted pelvis |
| DX_ICD10 | O33.2 | maternal care for disproportion due to inlet contraction of pelvis |
| DX_ICD10 | O33.3 | matern care for disproprtn due to outlet contrctn of pelvis |
| DX_ICD10 | O33.3XX0 | matern care for disprop due to outlet contraction pelvis, na or unspec |
| DX_ICD10 | O33.3XX1 | matern care for disprop due to outlet contraction pelvis, fetus 1 |
| DX_ICD10 | O33.3XX2 | matern care for disproportion due to outlet contract pelvis, fetus 2 |
| DX_ICD10 | O33.3XX3 | matern care for disproportion due to outlet contract pelvis, fetus 3 |
| DX_ICD10 | O33.3XX4 | matern care for disproportion due to outlet contract pelvis, fetus 4 |
| DX_ICD10 | O33.3XX5 | matern care for disproportion due to outlet contract pelvis, fetus 5 |
| DX_ICD10 | O33.3XX9 | matern care for disproportion due to outlet contract pelvis, oth fetus |
| DX_ICD10 | O33.4 | matern care for disproportion of mixed mat & fetal origin |
| DX_ICD10 | O33.4XX0 | matern care for disproportion of mixed mat & fetal origin, na or unspec |
| DX_ICD10 | O33.4XX1 | matern care for disproportion of mixed mat & fetal origin, fetus 1 |
| DX_ICD10 | O33.4XX2 | maternal care for disproportion of mixed mat & fetal origin, fetus 2 |
| DX_ICD10 | O33.4XX3 | maternal care for disproportion of mixed mat & fetal origin, fetus 3 |
| DX_ICD10 | O33.4XX4 | maternal care for disproportion of mixed mat & fetal origin, fetus 4 |
| DX_ICD10 | O33.4XX5 | maternal care for disproportion of mixed mat & fetal origin, fetus 5 |
| DX_ICD10 | O33.4XX9 | maternal care for disproportion of mixed mat & fetal origin, oth fetus |
| DX_ICD10 | O33.5 | maternal care for disproportion due to unusually lg fetus |
| DX_ICD10 | O33.5XX0 | maternal care for disproportion due to unusually lg fetus, na or unspec |
| DX_ICD10 | O33.5XX1 | maternal care for disproportion due to unusually lg fetus, fetus 1 |
| DX_ICD10 | O33.5XX2 | maternal care for disproportion due to unusually lg fetus, fetus 2 |
| DX_ICD10 | O33.5XX3 | maternal care for disproportion due to unusually lg fetus, fetus 3 |
| DX_ICD10 | O33.5XX4 | maternal care for disproportion due to unusually lg fetus, fetus 4 |
| DX_ICD10 | O33.5XX5 | maternal care for disproportion due to unusually lg fetus, fetus 5 |
| DX_ICD10 | O33.5XX9 | maternal care for disproportion due to unusually lg fetus, oth fetus |
| DX_ICD10 | O33.6 | maternal care for disproportion due to hydrocephalic fetus |
| DX_ICD10 | O33.6XX0 | maternal care for disproportion due to hydroceph fetus, na or unspec |
| DX_ICD10 | O33.6XX1 | maternal care for disproportion due to hydrocephalic fetus, fetus 1 |
| DX_ICD10 | O33.6XX2 | maternal care for disproportion due to hydrocephalic fetus, fetus 2 |
| DX_ICD10 | O33.6XX3 | maternal care for disproportion due to hydrocephalic fetus, fetus 3 |
| DX_ICD10 | O33.6XX4 | maternal care for disproportion due to hydrocephalic fetus, fetus 4 |
| DX_ICD10 | O33.6XX5 | maternal care for disproportion due to hydrocephalic fetus, fetus 5 |
| DX_ICD10 | O33.6XX9 | maternal care for disproportion due to hydrocephalic fetus, oth fetus |
| DX_ICD10 | O33.7 | maternal care for disproportion due to oth fetal deformities |
| DX_ICD10 | O33.7XX0 | maternal care for disproport due to oth fetal deformities, na or unspec |
| DX_ICD10 | O33.7XX1 | maternal care for disproport due to oth fetal deformities, fetus 1 |
| DX_ICD10 | O33.7XX2 | maternal care for disproport due to oth fetal deformities, fetus 2 |
| DX_ICD10 | O33.7XX3 | maternal care for disproport due to oth fetal deformities, fetus 3 |
| DX_ICD10 | O33.7XX4 | maternal care for disproport due to oth fetal deformities, fetus 4 |
| DX_ICD10 | O33.7XX5 | maternal care for disproport due to oth fetal deformities, fetus 5 |
| DX_ICD10 | O33.7XX9 | maternal care for disproport due to oth fetal deformities, oth fetus |
| DX_ICD10 | O33.8 | maternal care for disproportion of oth origin |
| DX_ICD10 | O33.9 | maternal care for disproportion, unspec |
| DX_ICD10 | O34 | maternal care for abnormality of pelvic organs |
| DX_ICD10 | O34.0 | maternal care for congenital malformation of uterus |
| DX_ICD10 | O34.00 | maternal care for unspec congenital malformation of uterus, unspec tri |
| DX_ICD10 | O34.01 | maternal care for unspec congenital malformation of uterus, first tri |
| DX_ICD10 | O34.02 | maternal care for unspec congenital malformation of uterus, second tri |
| DX_ICD10 | O34.03 | maternal care for unspec congenital malformation of uterus, third tri |
| DX_ICD10 | O34.1 | maternal care for benign tumor of corpus uteri |
| DX_ICD10 | O34.10 | maternal care for benign tumor of corpus uteri, unspec tri |
| DX_ICD10 | O34.11 | maternal care for benign tumor of corpus uteri, first tri |
| DX_ICD10 | O34.12 | maternal care for benign tumor of corpus uteri, second tri |
| DX_ICD10 | O34.13 | maternal care for benign tumor of corpus uteri, third tri |
| DX_ICD10 | O34.2 | maternal care due to uterine scar from previous surgery |
| DX_ICD10 | O34.29 | maternal care due to uterine scar from oth previous surgery |
| DX_ICD10 | O34.3 | maternal care for cervical incompetence |
| DX_ICD10 | O34.30 | maternal care for cervical incompetence, unspec tri |
| DX_ICD10 | O34.31 | maternal care for cervical incompetence, first tri |
| DX_ICD10 | O34.32 | maternal care for cervical incompetence, second tri |
| DX_ICD10 | O34.33 | maternal care for cervical incompetence, third tri |
| DX_ICD10 | O34.4 | maternal care for oth abnormalities of cervix |
| DX_ICD10 | O34.40 | maternal care for oth abnormalities of cervix, unspec tri |
| DX_ICD10 | O34.41 | maternal care for oth abnormalities of cervix, first tri |
| DX_ICD10 | O34.42 | maternal care for oth abnormalities of cervix, second tri |
| DX_ICD10 | O34.43 | maternal care for oth abnormalities of cervix, third tri |
| DX_ICD10 | O34.5 | maternal care for oth abnormalities of gravid uterus |
| DX_ICD10 | O34.51 | maternal care for incarceration of gravid uterus |
| DX_ICD10 | O34.511 | maternal care for incarceration of gravid uterus, first tri |
| DX_ICD10 | O34.512 | maternal care for incarceration of gravid uterus, second tri |
| DX_ICD10 | O34.513 | maternal care for incarceration of gravid uterus, third tri |
| DX_ICD10 | O34.519 | maternal care for incarceration of gravid uterus, unspec tri |
| DX_ICD10 | O34.52 | maternal care for prolapse of gravid uterus |
| DX_ICD10 | O34.521 | maternal care for prolapse of gravid uterus, first tri |
| DX_ICD10 | O34.522 | maternal care for prolapse of gravid uterus, second tri |
| DX_ICD10 | O34.523 | maternal care for prolapse of gravid uterus, third tri |
| DX_ICD10 | O34.529 | maternal care for prolapse of gravid uterus, unspec tri |
| DX_ICD10 | O34.53 | maternal care for retroversion of gravid uterus |
| DX_ICD10 | O34.531 | maternal care for retroversion of gravid uterus, first tri |
| DX_ICD10 | O34.532 | maternal care for retroversion of gravid uterus, second tri |
| DX_ICD10 | O34.533 | maternal care for retroversion of gravid uterus, third tri |
| DX_ICD10 | O34.539 | maternal care for retroversion of gravid uterus, unspec tri |
| DX_ICD10 | O34.59 | maternal care for oth abnormalities of gravid uterus |
| DX_ICD10 | O34.591 | maternal care for oth abnormalities of gravid uterus, first tri |
| DX_ICD10 | O34.592 | maternal care for oth abnormalities of gravid uterus, second tri |
| DX_ICD10 | O34.593 | maternal care for oth abnormalities of gravid uterus, third tri |
| DX_ICD10 | O34.599 | maternal care for oth abnormalities of gravid uterus, unspec tri |
| DX_ICD10 | O34.6 | maternal care for abnormality of vagina |
| DX_ICD10 | O34.60 | maternal care for abnormality of vagina, unspec tri |
| DX_ICD10 | O34.61 | maternal care for abnormality of vagina, first tri |
| DX_ICD10 | O34.62 | maternal care for abnormality of vagina, second tri |
| DX_ICD10 | O34.63 | maternal care for abnormality of vagina, third tri |
| DX_ICD10 | O34.7 | maternal care for abnormality of vulva & perineum |
| DX_ICD10 | O34.70 | maternal care for abnormality of vulva & perineum, unspec tri |
| DX_ICD10 | O34.71 | maternal care for abnormality of vulva & perineum, first tri |
| DX_ICD10 | O34.72 | maternal care for abnormality of vulva & perineum, second tri |
| DX_ICD10 | O34.73 | maternal care for abnormality of vulva & perineum, third tri |
| DX_ICD10 | O34.8 | maternal care for oth abnormalities of pelvic organs |
| DX_ICD10 | O34.80 | maternal care for oth abnormalities of pelvic organs, unspec tri |
| DX_ICD10 | O34.81 | maternal care for oth abnormalities of pelvic organs, first tri |
| DX_ICD10 | O34.82 | maternal care for oth abnormalities of pelvic organs, second tri |
| DX_ICD10 | O34.83 | maternal care for oth abnormalities of pelvic organs, third tri |
| DX_ICD10 | O34.9 | maternal care for abnormality of pelvic organ, unspec |
| DX_ICD10 | O34.90 | maternal care for abnormality of pelvic organ, unspec, unspec tri |
| DX_ICD10 | O34.91 | maternal care for abnormality of pelvic organ, unspec, first tri |
| DX_ICD10 | O34.92 | maternal care for abnormality of pelvic organ, unspec, second tri |
| DX_ICD10 | O34.93 | maternal care for abnormality of pelvic organ, unspec, third tri |
| DX_ICD10 | O35 | maternal care for known or suspected fetal abnormality & dam |
| DX_ICD10 | O35.0 | maternal care for (suspected) cns malformation in fetus |
| DX_ICD10 | O35.0XX0 | maternal care for (suspected) cns malformation in fetus, na or unspec |
| DX_ICD10 | O35.0XX1 | maternal care for (suspected) cns malformation in fetus, fetus 1 |
| DX_ICD10 | O35.0XX2 | maternal care for (suspected) cns malformation in fetus, fetus 2 |
| DX_ICD10 | O35.0XX3 | maternal care for (suspected) cns malformation in fetus, fetus 3 |
| DX_ICD10 | O35.0XX4 | maternal care for (suspected) cns malformation in fetus, fetus 4 |
| DX_ICD10 | O35.0XX5 | maternal care for (suspected) cns malformation in fetus, fetus 5 |
| DX_ICD10 | O35.0XX9 | maternal care for (suspected) cns malformation in fetus, oth fetus |
| DX_ICD10 | O35.1 | maternal care for (suspected) chromosomal abnormality in fetus |
| DX_ICD10 | O35.1XX0 | mat care for (suspected) chromosomal abn in fetus, na or unspec |
| DX_ICD10 | O35.1XX1 | maternal care for (suspected) chromosomal abn in fetus, fetus 1 |
| DX_ICD10 | O35.1XX2 | maternal care for (suspected) chromosomal abn in fetus, fetus 2 |
| DX_ICD10 | O35.1XX3 | maternal care for (suspected) chromosomal abn in fetus, fetus 3 |
| DX_ICD10 | O35.1XX4 | maternal care for (suspected) chromosomal abn in fetus, fetus 4 |
| DX_ICD10 | O35.1XX5 | maternal care for (suspected) chromosomal abn in fetus, fetus 5 |
| DX_ICD10 | O35.1XX9 | maternal care for (suspected) chromosomal abn in fetus, oth fetus |
| DX_ICD10 | O35.2 | maternal care for (suspected) hereditary dis in fetus |
| DX_ICD10 | O35.2XX0 | maternal care for (suspected) hereditary dis in fetus, na or unspec |
| DX_ICD10 | O35.2XX1 | maternal care for (suspected) hereditary dis in fetus, fetus 1 |
| DX_ICD10 | O35.2XX2 | maternal care for (suspected) hereditary dis in fetus, fetus 2 |
| DX_ICD10 | O35.2XX3 | maternal care for (suspected) hereditary dis in fetus, fetus 3 |
| DX_ICD10 | O35.2XX4 | maternal care for (suspected) hereditary dis in fetus, fetus 4 |
| DX_ICD10 | O35.2XX5 | maternal care for (suspected) hereditary dis in fetus, fetus 5 |
| DX_ICD10 | O35.2XX9 | maternal care for (suspected) hereditary dis in fetus, oth fetus |
| DX_ICD10 | O35.3 | maternal care for (suspected) dam to fetus from viral dis in moth |
| DX_ICD10 | O35.3XX0 | mat care for (suspect) dam to fetus from viral dis in moth, na or unspec |
| DX_ICD10 | O35.3XX1 | mat care for (suspected) dam to fetus from viral dis in moth, fetus 1 |
| DX_ICD10 | O35.3XX2 | mat care for (suspected) dam to fetus from viral dis in moth, fetus 2 |
| DX_ICD10 | O35.3XX3 | mat care for (suspected) dam to fetus from viral dis in moth, fetus 3 |
| DX_ICD10 | O35.3XX4 | mat care for (suspected) dam to fetus from viral dis in moth, fetus 4 |
| DX_ICD10 | O35.3XX5 | mat care for (suspected) dam to fetus from viral dis in moth, fetus 5 |
| DX_ICD10 | O35.3XX9 | mat care for (suspected) dam to fetus from viral dis in moth, oth fetus |
| DX_ICD10 | O35.4 | mat care for (suspected) dam to fetus from alcohol |
| DX_ICD10 | O35.4XX0 | mat care for (suspected) dam to fetus from alcohol, na or unspec |
| DX_ICD10 | O35.4XX1 | mat care for (suspected) dam to fetus from alcohol, fetus 1 |
| DX_ICD10 | O35.4XX2 | mat care for (suspected) dam to fetus from alcohol, fetus 2 |
| DX_ICD10 | O35.4XX3 | maternal care for (suspected) dam to fetus from alcohol, fetus 3 |
| DX_ICD10 | O35.4XX4 | maternal care for (suspected) dam to fetus from alcohol, fetus 4 |
| DX_ICD10 | O35.4XX5 | maternal care for (suspected) dam to fetus from alcohol, fetus 5 |
| DX_ICD10 | O35.4XX9 | maternal care for (suspected) dam to fetus from alcohol, oth fetus |
| DX_ICD10 | O35.5 | maternal care for (suspected) dam to fetus by drugs |
| DX_ICD10 | O35.5XX0 | maternal care for (suspected) dam to fetus by drugs, na or unspec |
| DX_ICD10 | O35.5XX1 | maternal care for (suspected) dam to fetus by drugs, fetus 1 |
| DX_ICD10 | O35.5XX2 | maternal care for (suspected) dam to fetus by drugs, fetus 2 |
| DX_ICD10 | O35.5XX3 | maternal care for (suspected) dam to fetus by drugs, fetus 3 |
| DX_ICD10 | O35.5XX4 | maternal care for (suspected) dam to fetus by drugs, fetus 4 |
| DX_ICD10 | O35.5XX5 | maternal care for (suspected) dam to fetus by drugs, fetus 5 |
| DX_ICD10 | O35.5XX9 | maternal care for (suspected) dam to fetus by drugs, oth fetus |
| DX_ICD10 | O35.6 | maternal care for (suspected) dam to fetus by radiation |
| DX_ICD10 | O35.6XX0 | maternal care for (suspected) dam to fetus by radiation, na or unspec |
| DX_ICD10 | O35.6XX1 | maternal care for (suspected) dam to fetus by radiation, fetus 1 |
| DX_ICD10 | O35.6XX2 | maternal care for (suspected) dam to fetus by radiation, fetus 2 |
| DX_ICD10 | O35.6XX3 | maternal care for (suspected) dam to fetus by radiation, fetus 3 |
| DX_ICD10 | O35.6XX4 | maternal care for (suspected) dam to fetus by radiation, fetus 4 |
| DX_ICD10 | O35.6XX5 | maternal care for (suspected) dam to fetus by radiation, fetus 5 |
| DX_ICD10 | O35.6XX9 | maternal care for (suspected) dam to fetus by radiation, oth fetus |
| DX_ICD10 | O35.7 | maternal care for (suspected) dam to fetus by oth med proc |
| DX_ICD10 | O35.7XX0 | mat care for (suspect) dam to fetus by oth med proc, na or unspec |
| DX_ICD10 | O35.7XX1 | mat care for (suspect) dam to fetus by oth med proc, fetus 1 |
| DX_ICD10 | O35.7XX2 | maternal care for (suspected) dam to fetus by oth med proc, fetus 2 |
| DX_ICD10 | O35.7XX3 | maternal care for (suspected) dam to fetus by oth med proc, fetus 3 |
| DX_ICD10 | O35.7XX4 | maternal care for (suspected) dam to fetus by oth med proc, fetus 4 |
| DX_ICD10 | O35.7XX5 | maternal care for (suspected) dam to fetus by oth med proc, fetus 5 |
| DX_ICD10 | O35.7XX9 | maternal care for (suspected) dam to fetus by oth med proc, oth fetus |
| DX_ICD10 | O35.8 | maternal care for oth (suspected) fetal abn & dam |
| DX_ICD10 | O35.8XX0 | maternal care for oth (suspected) fetal abn & dam, na or unspec |
| DX_ICD10 | O35.8XX1 | maternal care for oth (suspected) fetal abn & dam, fetus 1 |
| DX_ICD10 | O35.8XX2 | maternal care for oth (suspected) fetal abn & dam, fetus 2 |
| DX_ICD10 | O35.8XX3 | maternal care for oth (suspected) fetal abn & dam, fetus 3 |
| DX_ICD10 | O35.8XX4 | maternal care for oth (suspected) fetal abn & dam, fetus 4 |
| DX_ICD10 | O35.8XX5 | maternal care for oth (suspected) fetal abn & dam, fetus 5 |
| DX_ICD10 | O35.8XX9 | maternal care for oth (suspected) fetal abn & dam, oth fetus |
| DX_ICD10 | O35.9 | maternal care for (suspected) fetal abn & dam, unspec |
| DX_ICD10 | O35.9XX0 | maternal care for (suspected) fetal abn & dam, unspec, na or unspec |
| DX_ICD10 | O35.9XX1 | maternal care for (suspected) fetal abn & dam, unspec, fetus 1 |
| DX_ICD10 | O35.9XX2 | maternal care for (suspected) fetal abn & dam, unspec, fetus 2 |
| DX_ICD10 | O35.9XX3 | maternal care for (suspected) fetal abn & dam, unspec, fetus 3 |
| DX_ICD10 | O35.9XX4 | maternal care for (suspected) fetal abn & dam, unspec, fetus 4 |
| DX_ICD10 | O35.9XX5 | maternal care for (suspected) fetal abn & dam, unspec, fetus 5 |
| DX_ICD10 | O35.9XX9 | maternal care for (suspected) fetal abn & dam, unspec, oth fetus |
| DX_ICD10 | O36 | maternal care for oth fetal problems |
| DX_ICD10 | O36.0 | maternal care for rhesus isoimmunization |
| DX_ICD10 | O36.01 | maternal care for anti-d [rh] antibodies |
| DX_ICD10 | O36.011 | maternal care for anti-d [rh] antibodies, first tri |
| DX_ICD10 | O36.0110 | maternal care for anti-d [rh] antibodies, first tri, na or unspec |
| DX_ICD10 | O36.0111 | maternal care for anti-d [rh] antibodies, first tri, fetus 1 |
| DX_ICD10 | O36.0112 | maternal care for anti-d [rh] antibodies, first tri, fetus 2 |
| DX_ICD10 | O36.0113 | maternal care for anti-d [rh] antibodies, first tri, fetus 3 |
| DX_ICD10 | O36.0114 | maternal care for anti-d [rh] antibodies, first tri, fetus 4 |
| DX_ICD10 | O36.0115 | maternal care for anti-d [rh] antibodies, first tri, fetus 5 |
| DX_ICD10 | O36.0119 | maternal care for anti-d [rh] antibodies, first tri, oth fetus |
| DX_ICD10 | O36.012 | maternal care for anti-d [rh] antibodies, second tri |
| DX_ICD10 | O36.0120 | maternal care for anti-d [rh] antibodies, second tri, na or unspec |
| DX_ICD10 | O36.0121 | maternal care for anti-d [rh] antibodies, second tri, fetus 1 |
| DX_ICD10 | O36.0122 | maternal care for anti-d [rh] antibodies, second tri, fetus 2 |
| DX_ICD10 | O36.0123 | maternal care for anti-d [rh] antibodies, second tri, fetus 3 |
| DX_ICD10 | O36.0124 | maternal care for anti-d [rh] antibodies, second tri, fetus 4 |
| DX_ICD10 | O36.0125 | maternal care for anti-d [rh] antibodies, second tri, fetus 5 |
| DX_ICD10 | O36.0129 | maternal care for anti-d [rh] antibodies, second tri, oth fetus |
| DX_ICD10 | O36.013 | maternal care for anti-d [rh] antibodies, third tri |
| DX_ICD10 | O36.0130 | maternal care for anti-d [rh] antibodies, third tri, na or unspec |
| DX_ICD10 | O36.0131 | maternal care for anti-d [rh] antibodies, third tri, fetus 1 |
| DX_ICD10 | O36.0132 | maternal care for anti-d [rh] antibodies, third tri, fetus 2 |
| DX_ICD10 | O36.0133 | maternal care for anti-d [rh] antibodies, third tri, fetus 3 |
| DX_ICD10 | O36.0134 | maternal care for anti-d [rh] antibodies, third tri, fetus 4 |
| DX_ICD10 | O36.0135 | maternal care for anti-d [rh] antibodies, third tri, fetus 5 |
| DX_ICD10 | O36.0139 | maternal care for anti-d [rh] antibodies, third tri, oth fetus |
| DX_ICD10 | O36.019 | maternal care for anti-d [rh] antibodies, unspec tri |
| DX_ICD10 | O36.0190 | maternal care for anti-d [rh] antibodies, unspec tri, na or unspec |
| DX_ICD10 | O36.0191 | maternal care for anti-d [rh] antibodies, unspec tri, fetus 1 |
| DX_ICD10 | O36.0192 | maternal care for anti-d [rh] antibodies, unspec tri, fetus 2 |
| DX_ICD10 | O36.0193 | maternal care for anti-d [rh] antibodies, unspec tri, fetus 3 |
| DX_ICD10 | O36.0194 | maternal care for anti-d [rh] antibodies, unspec tri, fetus 4 |
| DX_ICD10 | O36.0195 | maternal care for anti-d [rh] antibodies, unspec tri, fetus 5 |
| DX_ICD10 | O36.0199 | maternal care for anti-d [rh] antibodies, unspec tri, oth fetus |
| DX_ICD10 | O36.09 | maternal care for oth rhesus isoimmunization |
| DX_ICD10 | O36.091 | maternal care for oth rhesus isoimmunization, first tri |
| DX_ICD10 | O36.0910 | maternal care for oth rhesus isoimmunization, first tri, na or unspec |
| DX_ICD10 | O36.0911 | maternal care for oth rhesus isoimmunization, first tri, fetus 1 |
| DX_ICD10 | O36.0912 | maternal care for oth rhesus isoimmunization, first tri, fetus 2 |
| DX_ICD10 | O36.0913 | maternal care for oth rhesus isoimmunization, first tri, fetus 3 |
| DX_ICD10 | O36.0914 | maternal care for oth rhesus isoimmunization, first tri, fetus 4 |
| DX_ICD10 | O36.0915 | maternal care for oth rhesus isoimmunization, first tri, fetus 5 |
| DX_ICD10 | O36.0919 | maternal care for oth rhesus isoimmunization, first tri, oth fetus |
| DX_ICD10 | O36.092 | maternal care for oth rhesus isoimmunization, second tri |
| DX_ICD10 | O36.0920 | maternal care for oth rhesus isoimmunization, second tri, na or unspec |
| DX_ICD10 | O36.0921 | maternal care for oth rhesus isoimmunization, second tri, fetus 1 |
| DX_ICD10 | O36.0922 | maternal care for oth rhesus isoimmunization, second tri, fetus 2 |
| DX_ICD10 | O36.0923 | maternal care for oth rhesus isoimmunization, second tri, fetus 3 |
| DX_ICD10 | O36.0924 | maternal care for oth rhesus isoimmunization, second tri, fetus 4 |
| DX_ICD10 | O36.0925 | maternal care for oth rhesus isoimmunization, second tri, fetus 5 |
| DX_ICD10 | O36.0929 | maternal care for oth rhesus isoimmunization, second tri, oth fetus |
| DX_ICD10 | O36.093 | maternal care for oth rhesus isoimmunization, third tri |
| DX_ICD10 | O36.0930 | maternal care for oth rhesus isoimmunization, third tri, na or unspec |
| DX_ICD10 | O36.0931 | maternal care for oth rhesus isoimmunization, third tri, fetus 1 |
| DX_ICD10 | O36.0932 | maternal care for oth rhesus isoimmunization, third tri, fetus 2 |
| DX_ICD10 | O36.0933 | maternal care for oth rhesus isoimmunization, third tri, fetus 3 |
| DX_ICD10 | O36.0934 | maternal care for oth rhesus isoimmunization, third tri, fetus 4 |
| DX_ICD10 | O36.0935 | maternal care for oth rhesus isoimmunization, third tri, fetus 5 |
| DX_ICD10 | O36.0939 | maternal care for oth rhesus isoimmunization, third tri, oth fetus |
| DX_ICD10 | O36.099 | maternal care for oth rhesus isoimmunization, unspec tri |
| DX_ICD10 | O36.0990 | maternal care for oth rhesus isoimmunization, unspec tri, na or unspec |
| DX_ICD10 | O36.0991 | maternal care for oth rhesus isoimmunization, unspec tri, fetus 1 |
| DX_ICD10 | O36.0992 | maternal care for oth rhesus isoimmunization, unspec tri, fetus 2 |
| DX_ICD10 | O36.0993 | maternal care for oth rhesus isoimmunization, unspec tri, fetus 3 |
| DX_ICD10 | O36.0994 | maternal care for oth rhesus isoimmunization, unspec tri, fetus 4 |
| DX_ICD10 | O36.0995 | maternal care for oth rhesus isoimmunization, unspec tri, fetus 5 |
| DX_ICD10 | O36.0999 | maternal care for oth rhesus isoimmunization, unspec tri, oth fetus |
| DX_ICD10 | O36.1 | maternal care for oth isoimmunization |
| DX_ICD10 | O36.11 | maternal care for anti-a sensitization |
| DX_ICD10 | O36.111 | maternal care for anti-a sensitization, first tri |
| DX_ICD10 | O36.1110 | maternal care for anti-a sensitization, first tri, na or unspec |
| DX_ICD10 | O36.1111 | maternal care for anti-a sensitization, first tri, fetus 1 |
| DX_ICD10 | O36.1112 | maternal care for anti-a sensitization, first tri, fetus 2 |
| DX_ICD10 | O36.1113 | maternal care for anti-a sensitization, first tri, fetus 3 |
| DX_ICD10 | O36.1114 | maternal care for anti-a sensitization, first tri, fetus 4 |
| DX_ICD10 | O36.1115 | maternal care for anti-a sensitization, first tri, fetus 5 |
| DX_ICD10 | O36.1119 | maternal care for anti-a sensitization, first tri, oth fetus |
| DX_ICD10 | O36.112 | maternal care for anti-a sensitization, second tri |
| DX_ICD10 | O36.1120 | maternal care for anti-a sensitization, second tri, na or unspec |
| DX_ICD10 | O36.1121 | maternal care for anti-a sensitization, second tri, fetus 1 |
| DX_ICD10 | O36.1122 | maternal care for anti-a sensitization, second tri, fetus 2 |
| DX_ICD10 | O36.1123 | maternal care for anti-a sensitization, second tri, fetus 3 |
| DX_ICD10 | O36.1124 | maternal care for anti-a sensitization, second tri, fetus 4 |
| DX_ICD10 | O36.1125 | maternal care for anti-a sensitization, second tri, fetus 5 |
| DX_ICD10 | O36.1129 | maternal care for anti-a sensitization, second tri, oth fetus |
| DX_ICD10 | O36.113 | maternal care for anti-a sensitization, third tri |
| DX_ICD10 | O36.1130 | maternal care for anti-a sensitization, third tri, na or unspec |
| DX_ICD10 | O36.1131 | maternal care for anti-a sensitization, third tri, fetus 1 |
| DX_ICD10 | O36.1132 | maternal care for anti-a sensitization, third tri, fetus 2 |
| DX_ICD10 | O36.1133 | maternal care for anti-a sensitization, third tri, fetus 3 |
| DX_ICD10 | O36.1134 | maternal care for anti-a sensitization, third tri, fetus 4 |
| DX_ICD10 | O36.1135 | maternal care for anti-a sensitization, third tri, fetus 5 |
| DX_ICD10 | O36.1139 | maternal care for anti-a sensitization, third tri, oth fetus |
| DX_ICD10 | O36.119 | maternal care for anti-a sensitization, unspec tri |
| DX_ICD10 | O36.1190 | maternal care for anti-a sensitization, unspec tri, na or unspec |
| DX_ICD10 | O36.1191 | maternal care for anti-a sensitization, unspec tri, fetus 1 |
| DX_ICD10 | O36.1192 | maternal care for anti-a sensitization, unspec tri, fetus 2 |
| DX_ICD10 | O36.1193 | maternal care for anti-a sensitization, unspec tri, fetus 3 |
| DX_ICD10 | O36.1194 | maternal care for anti-a sensitization, unspec tri, fetus 4 |
| DX_ICD10 | O36.1195 | maternal care for anti-a sensitization, unspec tri, fetus 5 |
| DX_ICD10 | O36.1199 | maternal care for anti-a sensitization, unspec tri, oth fetus |
| DX_ICD10 | O36.19 | maternal care for oth isoimmunization |
| DX_ICD10 | O36.191 | maternal care for oth isoimmunization, first tri |
| DX_ICD10 | O36.1910 | maternal care for oth isoimmunization, first tri, na or unspec |
| DX_ICD10 | O36.1911 | maternal care for oth isoimmunization, first tri, fetus 1 |
| DX_ICD10 | O36.1912 | maternal care for oth isoimmunization, first tri, fetus 2 |
| DX_ICD10 | O36.1913 | maternal care for oth isoimmunization, first tri, fetus 3 |
| DX_ICD10 | O36.1914 | maternal care for oth isoimmunization, first tri, fetus 4 |
| DX_ICD10 | O36.1915 | maternal care for oth isoimmunization, first tri, fetus 5 |
| DX_ICD10 | O36.1919 | maternal care for oth isoimmunization, first tri, oth fetus |
| DX_ICD10 | O36.192 | maternal care for oth isoimmunization, second tri |
| DX_ICD10 | O36.1920 | maternal care for oth isoimmunization, second tri, na or unspec |
| DX_ICD10 | O36.1921 | maternal care for oth isoimmunization, second tri, fetus 1 |
| DX_ICD10 | O36.1922 | maternal care for oth isoimmunization, second tri, fetus 2 |
| DX_ICD10 | O36.1923 | maternal care for oth isoimmunization, second tri, fetus 3 |
| DX_ICD10 | O36.1924 | maternal care for oth isoimmunization, second tri, fetus 4 |
| DX_ICD10 | O36.1925 | maternal care for oth isoimmunization, second tri, fetus 5 |
| DX_ICD10 | O36.1929 | maternal care for oth isoimmunization, second tri, oth fetus |
| DX_ICD10 | O36.193 | maternal care for oth isoimmunization, third tri |
| DX_ICD10 | O36.1930 | maternal care for oth isoimmunization, third tri, na or unspec |
| DX_ICD10 | O36.1931 | maternal care for oth isoimmunization, third tri, fetus 1 |
| DX_ICD10 | O36.1932 | maternal care for oth isoimmunization, third tri, fetus 2 |
| DX_ICD10 | O36.1933 | maternal care for oth isoimmunization, third tri, fetus 3 |
| DX_ICD10 | O36.1934 | maternal care for oth isoimmunization, third tri, fetus 4 |
| DX_ICD10 | O36.1935 | maternal care for oth isoimmunization, third tri, fetus 5 |
| DX_ICD10 | O36.1939 | maternal care for oth isoimmunization, third tri, oth fetus |
| DX_ICD10 | O36.199 | maternal care for oth isoimmunization, unspec tri |
| DX_ICD10 | O36.1990 | maternal care for oth isoimmunization, unspec tri, na or unspec |
| DX_ICD10 | O36.1991 | maternal care for oth isoimmunization, unspec tri, fetus 1 |
| DX_ICD10 | O36.1992 | maternal care for oth isoimmunization, unspec tri, fetus 2 |
| DX_ICD10 | O36.1993 | maternal care for oth isoimmunization, unspec tri, fetus 3 |
| DX_ICD10 | O36.1994 | maternal care for oth isoimmunization, unspec tri, fetus 4 |
| DX_ICD10 | O36.1995 | maternal care for oth isoimmunization, unspec tri, fetus 5 |
| DX_ICD10 | O36.1999 | maternal care for oth isoimmunization, unspec tri, oth fetus |
| DX_ICD10 | O36.2 | maternal care for hydrops fetalis |
| DX_ICD10 | O36.20 | maternal care for hydrops fetalis, unspec tri |
| DX_ICD10 | O36.20X0 | maternal care for hydrops fetalis, unspec tri, na or unspec |
| DX_ICD10 | O36.20X1 | maternal care for hydrops fetalis, unspec tri, fetus 1 |
| DX_ICD10 | O36.20X2 | maternal care for hydrops fetalis, unspec tri, fetus 2 |
| DX_ICD10 | O36.20X3 | maternal care for hydrops fetalis, unspec tri, fetus 3 |
| DX_ICD10 | O36.20X4 | maternal care for hydrops fetalis, unspec tri, fetus 4 |
| DX_ICD10 | O36.20X5 | maternal care for hydrops fetalis, unspec tri, fetus 5 |
| DX_ICD10 | O36.20X9 | maternal care for hydrops fetalis, unspec tri, oth fetus |
| DX_ICD10 | O36.21 | maternal care for hydrops fetalis, first tri |
| DX_ICD10 | O36.21X0 | maternal care for hydrops fetalis, first tri, na or unspec |
| DX_ICD10 | O36.21X1 | maternal care for hydrops fetalis, first tri, fetus 1 |
| DX_ICD10 | O36.21X2 | maternal care for hydrops fetalis, first tri, fetus 2 |
| DX_ICD10 | O36.21X3 | maternal care for hydrops fetalis, first tri, fetus 3 |
| DX_ICD10 | O36.21X4 | maternal care for hydrops fetalis, first tri, fetus 4 |
| DX_ICD10 | O36.21X5 | maternal care for hydrops fetalis, first tri, fetus 5 |
| DX_ICD10 | O36.21X9 | maternal care for hydrops fetalis, first tri, oth fetus |
| DX_ICD10 | O36.22 | maternal care for hydrops fetalis, second tri |
| DX_ICD10 | O36.22X0 | maternal care for hydrops fetalis, second tri, na or unspec |
| DX_ICD10 | O36.22X1 | maternal care for hydrops fetalis, second tri, fetus 1 |
| DX_ICD10 | O36.22X2 | maternal care for hydrops fetalis, second tri, fetus 2 |
| DX_ICD10 | O36.22X3 | maternal care for hydrops fetalis, second tri, fetus 3 |
| DX_ICD10 | O36.22X4 | maternal care for hydrops fetalis, second tri, fetus 4 |
| DX_ICD10 | O36.22X5 | maternal care for hydrops fetalis, second tri, fetus 5 |
| DX_ICD10 | O36.22X9 | maternal care for hydrops fetalis, second tri, oth fetus |
| DX_ICD10 | O36.23 | maternal care for hydrops fetalis, third tri |
| DX_ICD10 | O36.23X0 | maternal care for hydrops fetalis, third tri, na or unspec |
| DX_ICD10 | O36.23X1 | maternal care for hydrops fetalis, third tri, fetus 1 |
| DX_ICD10 | O36.23X2 | maternal care for hydrops fetalis, third tri, fetus 2 |
| DX_ICD10 | O36.23X3 | maternal care for hydrops fetalis, third tri, fetus 3 |
| DX_ICD10 | O36.23X4 | maternal care for hydrops fetalis, third tri, fetus 4 |
| DX_ICD10 | O36.23X5 | maternal care for hydrops fetalis, third tri, fetus 5 |
| DX_ICD10 | O36.23X9 | maternal care for hydrops fetalis, third tri, oth fetus |
| DX_ICD10 | O36.4 | maternal care for intrauterine death |
| DX_ICD10 | O36.4XX0 | maternal care for intrauterine death, na or unspec |
| DX_ICD10 | O36.4XX1 | maternal care for intrauterine death, fetus 1 |
| DX_ICD10 | O36.4XX2 | maternal care for intrauterine death, fetus 2 |
| DX_ICD10 | O36.4XX3 | maternal care for intrauterine death, fetus 3 |
| DX_ICD10 | O36.4XX4 | maternal care for intrauterine death, fetus 4 |
| DX_ICD10 | O36.4XX5 | maternal care for intrauterine death, fetus 5 |
| DX_ICD10 | O36.4XX9 | maternal care for intrauterine death, oth fetus |
| DX_ICD10 | O36.5 | maternal care for known or suspected poor fetal growth |
| DX_ICD10 | O36.51 | maternal care for known or suspected placental insufficiency |
| DX_ICD10 | O36.511 | maternal care for known or suspected placental insufficiency, first tri |
| DX_ICD10 | O36.5110 | maternal care for known/suspect plcntal insuff, first tri, na or unspec |
| DX_ICD10 | O36.5111 | maternal care for known/suspect plcntal insuffic, first tri, fetus 1 |
| DX_ICD10 | O36.5112 | maternal care for known/suspect plcental insuffic, first tri, fetus 2 |
| DX_ICD10 | O36.5113 | maternal care for known/suspect plcental insuffic, first tri, fetus 3 |
| DX_ICD10 | O36.5114 | maternal care for known/suspect plcental insuffic, first tri, fetus 4 |
| DX_ICD10 | O36.5115 | maternal care for known/suspect plcntal insuffic, first tri, fetus 5 |
| DX_ICD10 | O36.5119 | maternal care for known/suspect plcntal insuffic, first tri, oth fetus |
| DX_ICD10 | O36.512 | maternal care for known/suspect plcntal insufficiency, second tri |
| DX_ICD10 | O36.5120 | mat care for known/suspect plcntal insuffic, second tri, na or unspec |
| DX_ICD10 | O36.5121 | mat care for known/suspect plcntal insuffic, second tri, fetus 1 |
| DX_ICD10 | O36.5122 | mat care for known/suspect plcntal insuffic, second tri, fetus 2 |
| DX_ICD10 | O36.5123 | mat care for known/suspect plcntal insuffic, second tri, fetus 3 |
| DX_ICD10 | O36.5124 | mat care for known/suspect plcntal insuffic, second tri, fetus 4 |
| DX_ICD10 | O36.5125 | mat care for known/suspect plcntal insuffic, second tri, fetus 5 |
| DX_ICD10 | O36.5129 | mat care for known/suspect plcntal insuffic, second tri, oth fetus |
| DX_ICD10 | O36.513 | mat care for known/suspect plcntal insuffic, third tri |
| DX_ICD10 | O36.5130 | mat care for known/suspect plcntal insuffic, third tri, na or unspec |
| DX_ICD10 | O36.5131 | mat care for known/suspect plcntal insuffic, third tri, fetus 1 |
| DX_ICD10 | O36.5132 | mat care for known/suspect plcntal insuffic, third tri, fetus 2 |
| DX_ICD10 | O36.5133 | mat care for known/suspect plcntal insuffic, third tri, fetus 3 |
| DX_ICD10 | O36.5134 | mat care for known/suspect plcntal insuffic, third tri, fetus 4 |
| DX_ICD10 | O36.5135 | mat care for known/suspect plcntal insuffic, third tri, fetus 5 |
| DX_ICD10 | O36.5139 | mat care for known/suspect plcntal insuffic, third tri, oth fetus |
| DX_ICD10 | O36.519 | mat care for known/suspect plcntal insuffic, unspec tri |
| DX_ICD10 | O36.5190 | mat care for known/suspect plcntal insuffic, unspec tri, na or unspec |
| DX_ICD10 | O36.5191 | mat care for known/suspect plcntal insuffic, unspec tri, fetus 1 |
| DX_ICD10 | O36.5192 | mat care for known/suspect plcntal insuffic, unspec tri, fetus 2 |
| DX_ICD10 | O36.5193 | mat care for known/suspect plcntal insuffic, unspec tri, fetus 3 |
| DX_ICD10 | O36.5194 | mat care for known/suspect plcntal insuffic, unspec tri, fetus 4 |
| DX_ICD10 | O36.5195 | mat care for known/suspect plcntal insuffic, unspec tri, fetus 5 |
| DX_ICD10 | O36.5199 | mat care for known/suspect plcntal insuffic, unspec tri, oth fetus |
| DX_ICD10 | O36.59 | mat care for known/suspect poor fetal growth |
| DX_ICD10 | O36.591 | mat care for known/suspect poor fetal growth, first tri |
| DX_ICD10 | O36.5910 | mat care for known/suspect fetal growth, first tri, na or unspec |
| DX_ICD10 | O36.5911 | mat care for known/suspect poor fetal growth, first tri, fetus 1 |
| DX_ICD10 | O36.5912 | mat care for known/suspect poor fetal growth, first tri, fetus 2 |
| DX_ICD10 | O36.5913 | mat care for known/suspect poor fetal growth, first tri, fetus 3 |
| DX_ICD10 | O36.5914 | mat care for known/suspect poor fetal growth, first tri, fetus 4 |
| DX_ICD10 | O36.5915 | mat care for known/suspect poor fetal growth, first tri, fetus 5 |
| DX_ICD10 | O36.5919 | mat care for known/suspect poor fetal growth, first tri, oth fetus |
| DX_ICD10 | O36.592 | mat care for known/suspect poor fetal growth, second tri |
| DX_ICD10 | O36.5920 | mat care for known/suspect poor fetal growth, second tri, na or unspec |
| DX_ICD10 | O36.5921 | mat care for known/suspect poor fetal growth, second tri, fetus 1 |
| DX_ICD10 | O36.5922 | mat care for known/suspect poor fetal growth, second tri, fetus 2 |
| DX_ICD10 | O36.5923 | mat care for known/suspect poor fetal growth, second tri, fetus 3 |
| DX_ICD10 | O36.5924 | mat care for known/suspect poor fetal growth, second tri, fetus 4 |
| DX_ICD10 | O36.5925 | mat care for known/suspect poor fetal growth, second tri, fetus 5 |
| DX_ICD10 | O36.5929 | mat care for known/suspect poor fetal growth, second tri, oth fetus |
| DX_ICD10 | O36.593 | mat care for known/suspect poor fetal growth, third tri |
| DX_ICD10 | O36.5930 | mat care for known/suspect poor fetal growth, third tri, na or unspec |
| DX_ICD10 | O36.5931 | mat care for known/suspect poor fetal growth, third tri, fetus 1 |
| DX_ICD10 | O36.5932 | mat care for known/suspect poor fetal growth, third tri, fetus 2 |
| DX_ICD10 | O36.5933 | mat care for known/suspect poor fetal growth, third tri, fetus 3 |
| DX_ICD10 | O36.5934 | mat care for known/suspect poor fetal growth, third tri, fetus 4 |
| DX_ICD10 | O36.5935 | mat care for known/suspect poor fetal growth, third tri, fetus 5 |
| DX_ICD10 | O36.5939 | mat care for known/suspect poor fetal growth, third tri, oth fetus |
| DX_ICD10 | O36.599 | mat care for known/suspect poor fetal growth, unspec tri |
| DX_ICD10 | O36.5990 | mat care for known/suspect poor fetal growth, unspec tri, na or unspec |
| DX_ICD10 | O36.5991 | mat care for known/suspect poor fetal growth, unspec tri, fetus 1 |
| DX_ICD10 | O36.5992 | mat care for known/suspect poor fetal growth, unspec tri, fetus 2 |
| DX_ICD10 | O36.5993 | mat care for known/suspect poor fetal growth, unspec tri, fetus 3 |
| DX_ICD10 | O36.5994 | mat care for known/suspect poor fetal growth, unspec tri, fetus 4 |
| DX_ICD10 | O36.5995 | mat care for known/suspect poor fetal growth, unspec tri, fetus 5 |
| DX_ICD10 | O36.5999 | mat care for known/suspect poor fetal growth, unspec tri, oth fetus |
| DX_ICD10 | O36.6 | maternal care for excessive fetal growth |
| DX_ICD10 | O36.60 | maternal care for excessive fetal growth, unspec tri |
| DX_ICD10 | O36.60X0 | maternal care for excessive fetal growth, unspec tri, na or unspec |
| DX_ICD10 | O36.60X1 | maternal care for excessive fetal growth, unspec tri, fetus 1 |
| DX_ICD10 | O36.60X2 | maternal care for excessive fetal growth, unspec tri, fetus 2 |
| DX_ICD10 | O36.60X3 | maternal care for excessive fetal growth, unspec tri, fetus 3 |
| DX_ICD10 | O36.60X4 | maternal care for excessive fetal growth, unspec tri, fetus 4 |
| DX_ICD10 | O36.60X5 | maternal care for excessive fetal growth, unspec tri, fetus 5 |
| DX_ICD10 | O36.60X9 | maternal care for excessive fetal growth, unspec tri, oth fetus |
| DX_ICD10 | O36.61 | maternal care for excessive fetal growth, first tri |
| DX_ICD10 | O36.61X0 | maternal care for excessive fetal growth, first tri, na or unspec |
| DX_ICD10 | O36.61X1 | maternal care for excessive fetal growth, first tri, fetus 1 |
| DX_ICD10 | O36.61X2 | maternal care for excessive fetal growth, first tri, fetus 2 |
| DX_ICD10 | O36.61X3 | maternal care for excessive fetal growth, first tri, fetus 3 |
| DX_ICD10 | O36.61X4 | maternal care for excessive fetal growth, first tri, fetus 4 |
| DX_ICD10 | O36.61X5 | maternal care for excessive fetal growth, first tri, fetus 5 |
| DX_ICD10 | O36.61X9 | maternal care for excessive fetal growth, first tri, oth fetus |
| DX_ICD10 | O36.62 | maternal care for excessive fetal growth, second tri |
| DX_ICD10 | O36.62X0 | maternal care for excessive fetal growth, second tri, na or unspec |
| DX_ICD10 | O36.62X1 | maternal care for excessive fetal growth, second tri, fetus 1 |
| DX_ICD10 | O36.62X2 | maternal care for excessive fetal growth, second tri, fetus 2 |
| DX_ICD10 | O36.62X3 | maternal care for excessive fetal growth, second tri, fetus 3 |
| DX_ICD10 | O36.62X4 | maternal care for excessive fetal growth, second tri, fetus 4 |
| DX_ICD10 | O36.62X5 | maternal care for excessive fetal growth, second tri, fetus 5 |
| DX_ICD10 | O36.62X9 | maternal care for excessive fetal growth, second tri, oth fetus |
| DX_ICD10 | O36.63 | maternal care for excessive fetal growth, third tri |
| DX_ICD10 | O36.63X0 | maternal care for excessive fetal growth, third tri, na or unspec |
| 333DX_ICD10 | O36.63X1 | maternal care for excessive fetal growth, third tri, fetus 1 |
| DX_ICD10 | O36.63X2 | maternal care for excessive fetal growth, third tri, fetus 2 |
| DX_ICD10 | O36.63X3 | maternal care for excessive fetal growth, third tri, fetus 3 |
| DX_ICD10 | O36.63X4 | maternal care for excessive fetal growth, third tri, fetus 4 |
| DX_ICD10 | O36.63X5 | maternal care for excessive fetal growth, third tri, fetus 5 |
| DX_ICD10 | O36.63X9 | maternal care for excessive fetal growth, third tri, oth fetus |
| DX_ICD10 | O36.7 | maternal care for viable fetus in abdom preg |
| DX_ICD10 | O36.70 | maternal care for viable fetus in abdom preg, unspec tri |
| DX_ICD10 | O36.70X0 | maternal care for viable fetus in abdom preg, unspec tri, na or unspec |
| DX_ICD10 | O36.70X1 | maternal care for viable fetus in abdom preg, unspec tri, fetus 1 |
| DX_ICD10 | O36.70X2 | maternal care for viable fetus in abdom preg, unspec tri, fetus 2 |
| DX_ICD10 | O36.70X3 | maternal care for viable fetus in abdom preg, unspec tri, fetus 3 |
| DX_ICD10 | O36.70X4 | maternal care for viable fetus in abdom preg, unspec tri, fetus 4 |
| DX_ICD10 | O36.70X5 | maternal care for viable fetus in abdom preg, unspec tri, fetus 5 |
| DX_ICD10 | O36.70X9 | maternal care for viable fetus in abdom preg, unspec tri, oth fetus |
| DX_ICD10 | O36.71 | maternal care for viable fetus in abdom preg, first tri |
| DX_ICD10 | O36.71X0 | maternal care for viable fetus in abdom preg, first tri, na or unspec |
| DX_ICD10 | O36.71X1 | maternal care for viable fetus in abdom preg, first tri, fetus 1 |
| DX_ICD10 | O36.71X2 | maternal care for viable fetus in abdom preg, first tri, fetus 2 |
| DX_ICD10 | O36.71X3 | maternal care for viable fetus in abdom preg, first tri, fetus 3 |
| DX_ICD10 | O36.71X4 | maternal care for viable fetus in abdom preg, first tri, fetus 4 |
| DX_ICD10 | O36.71X5 | maternal care for viable fetus in abdom preg, first tri, fetus 5 |
| DX_ICD10 | O36.71X9 | maternal care for viable fetus in abdom preg, first tri, oth fetus |
| DX_ICD10 | O36.72 | maternal care for viable fetus in abdom preg, second tri |
| DX_ICD10 | O36.72X0 | maternal care for viable fetus in abdom preg, second tri, na or unspec |
| DX_ICD10 | O36.72X1 | maternal care for viable fetus in abdom preg, second tri, fetus 1 |
| DX_ICD10 | O36.72X2 | maternal care for viable fetus in abdom preg, second tri, fetus 2 |
| DX_ICD10 | O36.72X3 | maternal care for viable fetus in abdom preg, second tri, fetus 3 |
| DX_ICD10 | O36.72X4 | maternal care for viable fetus in abdom preg, second tri, fetus 4 |
| DX_ICD10 | O36.72X5 | maternal care for viable fetus in abdom preg, second tri, fetus 5 |
| DX_ICD10 | O36.72X9 | maternal care for viable fetus in abdom preg, second tri, oth fetus |
| DX_ICD10 | O36.73 | maternal care for viable fetus in abdom preg, third tri |
| DX_ICD10 | O36.73X0 | maternal care for viable fetus in abdom preg, third tri, na or unspec |
| DX_ICD10 | O36.73X1 | maternal care for viable fetus in abdom preg, third tri, fetus 1 |
| DX_ICD10 | O36.73X2 | maternal care for viable fetus in abdom preg, third tri, fetus 2 |
| DX_ICD10 | O36.73X3 | maternal care for viable fetus in abdom preg, third tri, fetus 3 |
| DX_ICD10 | O36.73X4 | maternal care for viable fetus in abdom preg, third tri, fetus 4 |
| DX_ICD10 | O36.73X5 | maternal care for viable fetus in abdom preg, third tri, fetus 5 |
| DX_ICD10 | O36.73X9 | maternal care for viable fetus in abdom preg, third tri, oth fetus |
| DX_ICD10 | O36.8 | maternal care for oth specif fetal problems |
| DX_ICD10 | O36.80 | preg with inconclusive fetal viability |
| DX_ICD10 | O36.80X0 | preg with inconclusive fetal viability, na or unspec |
| DX_ICD10 | O36.80X1 | preg with inconclusive fetal viability, fetus 1 |
| DX_ICD10 | O36.80X2 | preg with inconclusive fetal viability, fetus 2 |
| DX_ICD10 | O36.80X3 | preg with inconclusive fetal viability, fetus 3 |
| DX_ICD10 | O36.80X4 | preg with inconclusive fetal viability, fetus 4 |
| DX_ICD10 | O36.80X5 | preg with inconclusive fetal viability, fetus 5 |
| DX_ICD10 | O36.80X9 | preg with inconclusive fetal viability, oth fetus |
| DX_ICD10 | O36.81 | decreased fetal movements |
| DX_ICD10 | O36.812 | decreased fetal movements, second tri |
| DX_ICD10 | O36.8120 | decreased fetal movements, second tri, na or unspec |
| DX_ICD10 | O36.8121 | decreased fetal movements, second tri, fetus 1 |
| DX_ICD10 | O36.8122 | decreased fetal movements, second tri, fetus 2 |
| DX_ICD10 | O36.8123 | decreased fetal movements, second tri, fetus 3 |
| DX_ICD10 | O36.8124 | decreased fetal movements, second tri, fetus 4 |
| DX_ICD10 | O36.8125 | decreased fetal movements, second tri, fetus 5 |
| DX_ICD10 | O36.8129 | decreased fetal movements, second tri, oth fetus |
| DX_ICD10 | O36.813 | decreased fetal movements, third tri |
| DX_ICD10 | O36.8130 | decreased fetal movements, third tri, na or unspec |
| DX_ICD10 | O36.8131 | decreased fetal movements, third tri, fetus 1 |
| DX_ICD10 | O36.8132 | decreased fetal movements, third tri, fetus 2 |
| DX_ICD10 | O36.8133 | decreased fetal movements, third tri, fetus 3 |
| DX_ICD10 | O36.8134 | decreased fetal movements, third tri, fetus 4 |
| DX_ICD10 | O36.8135 | decreased fetal movements, third tri, fetus 5 |
| DX_ICD10 | O36.8139 | decreased fetal movements, third tri, oth fetus |
| DX_ICD10 | O36.819 | decreased fetal movements, unspec tri |
| DX_ICD10 | O36.8190 | decreased fetal movements, unspec tri, na or unspec |
| DX_ICD10 | O36.8191 | decreased fetal movements, unspec tri, fetus 1 |
| DX_ICD10 | O36.8192 | decreased fetal movements, unspec tri, fetus 2 |
| DX_ICD10 | O36.8193 | decreased fetal movements, unspec tri, fetus 3 |
| DX_ICD10 | O36.8194 | decreased fetal movements, unspec tri, fetus 4 |
| DX_ICD10 | O36.8195 | decreased fetal movements, unspec tri, fetus 5 |
| DX_ICD10 | O36.8199 | decreased fetal movements, unspec tri, oth fetus |
| DX_ICD10 | O36.82 | fetal anemia & thrombocytopenia |
| DX_ICD10 | O36.821 | fetal anemia & thrombocytopenia, first tri |
| DX_ICD10 | O36.8210 | fetal anemia & thrombocytopenia, first tri, na or unspec |
| DX_ICD10 | O36.8211 | fetal anemia & thrombocytopenia, first tri, fetus 1 |
| DX_ICD10 | O36.8212 | fetal anemia & thrombocytopenia, first tri, fetus 2 |
| DX_ICD10 | O36.8213 | fetal anemia & thrombocytopenia, first tri, fetus 3 |
| DX_ICD10 | O36.8214 | fetal anemia & thrombocytopenia, first tri, fetus 4 |
| DX_ICD10 | O36.8215 | fetal anemia & thrombocytopenia, first tri, fetus 5 |
| DX_ICD10 | O36.8219 | fetal anemia & thrombocytopenia, first tri, oth fetus |
| DX_ICD10 | O36.822 | fetal anemia & thrombocytopenia, second tri |
| DX_ICD10 | O36.8220 | fetal anemia & thrombocytopenia, second tri, na or unspec |
| DX_ICD10 | O36.8221 | fetal anemia & thrombocytopenia, second tri, fetus 1 |
| DX_ICD10 | O36.8222 | fetal anemia & thrombocytopenia, second tri, fetus 2 |
| DX_ICD10 | O36.8223 | fetal anemia & thrombocytopenia, second tri, fetus 3 |
| DX_ICD10 | O36.8224 | fetal anemia & thrombocytopenia, second tri, fetus 4 |
| DX_ICD10 | O36.8225 | fetal anemia & thrombocytopenia, second tri, fetus 5 |
| DX_ICD10 | O36.8229 | fetal anemia & thrombocytopenia, second tri, oth fetus |
| DX_ICD10 | O36.823 | fetal anemia & thrombocytopenia, third tri |
| DX_ICD10 | O36.8230 | fetal anemia & thrombocytopenia, third tri, na or unspec |
| DX_ICD10 | O36.8231 | fetal anemia & thrombocytopenia, third tri, fetus 1 |
| DX_ICD10 | O36.8232 | fetal anemia & thrombocytopenia, third tri, fetus 2 |
| DX_ICD10 | O36.8233 | fetal anemia & thrombocytopenia, third tri, fetus 3 |
| DX_ICD10 | O36.8234 | fetal anemia & thrombocytopenia, third tri, fetus 4 |
| DX_ICD10 | O36.8235 | fetal anemia & thrombocytopenia, third tri, fetus 5 |
| DX_ICD10 | O36.8239 | fetal anemia & thrombocytopenia, third tri, oth fetus |
| DX_ICD10 | O36.829 | fetal anemia & thrombocytopenia, unspec tri |
| DX_ICD10 | O36.8290 | fetal anemia & thrombocytopenia, unspec tri, na or unspec |
| DX_ICD10 | O36.8291 | fetal anemia & thrombocytopenia, unspec tri, fetus 1 |
| DX_ICD10 | O36.8292 | fetal anemia & thrombocytopenia, unspec tri, fetus 2 |
| DX_ICD10 | O36.8293 | fetal anemia & thrombocytopenia, unspec tri, fetus 3 |
| DX_ICD10 | O36.8294 | fetal anemia & thrombocytopenia, unspec tri, fetus 4 |
| DX_ICD10 | O36.8295 | fetal anemia & thrombocytopenia, unspec tri, fetus 5 |
| DX_ICD10 | O36.8299 | fetal anemia & thrombocytopenia, unspec tri, oth fetus |
| DX_ICD10 | O36.8310 | maternal care for abn fetal hrt rate or rhythm, first tri, na or unspec |
| DX_ICD10 | O36.8311 | maternal care for abn fetal hrt rate or rhythm, first tri, fetus 1 |
| DX_ICD10 | O36.8312 | maternal care for abn fetal hrt rate or rhythm, first tri, fetus 2 |
| DX_ICD10 | O36.8313 | maternal care for abn fetal hrt rate or rhythm, first tri, fetus 3 |
| DX_ICD10 | O36.8314 | maternal care for abn fetal hrt rate or rhythm, first tri, fetus 4 |
| DX_ICD10 | O36.8315 | maternal care for abn fetal hrt rate or rhythm, first tri, fetus 5 |
| DX_ICD10 | O36.8319 | maternal care for abn fetal hrt rate or rhythm, first tri, oth fetus |
| DX_ICD10 | O36.8320 | maternal care for abn fetal hrt rate or rhythm, second tri, na or unspec |
| DX_ICD10 | O36.8321 | maternal care for abn fetal hrt rate or rhythm, second tri, fetus 1 |
| DX_ICD10 | O36.8322 | maternal care for abn fetal hrt rate or rhythm, second tri, fetus 2 |
| DX_ICD10 | O36.8323 | maternal care for abn fetal hrt rate or rhythm, second tri, fetus 3 |
| DX_ICD10 | O36.8324 | maternal care for abn fetal hrt rate or rhythm, second tri, fetus 4 |
| DX_ICD10 | O36.8325 | maternal care for abn fetal hrt rate or rhythm, second tri, fetus 5 |
| DX_ICD10 | O36.8329 | maternal care for abn fetal hrt rate or rhythm, second tri, oth fetus |
| DX_ICD10 | O36.8330 | maternal care for abn fetal hrt rate or rhythm, third tri, na or unspec |
| DX_ICD10 | O36.8331 | maternal care for abn fetal hrt rate or rhythm, third tri, fetus 1 |
| DX_ICD10 | O36.8332 | maternal care for abn fetal hrt rate or rhythm, third tri, fetus 2 |
| DX_ICD10 | O36.8333 | maternal care for abn fetal hrt rate or rhythm, third tri, fetus 3 |
| DX_ICD10 | O36.8334 | maternal care for abn fetal hrt rate or rhythm, third tri, fetus 4 |
| DX_ICD10 | O36.8335 | maternal care for abn fetal hrt rate or rhythm, third tri, fetus 5 |
| DX_ICD10 | O36.8339 | maternal care for abn fetal hrt rate or rhythm, third tri, oth fetus |
| DX_ICD10 | O36.8390 | maternal care for abn fetal hrt rate or rhythm, unspec tri, na or unspec |
| DX_ICD10 | O36.8391 | maternal care for abn fetal hrt rate or rhythm, unspec tri, fetus 1 |
| DX_ICD10 | O36.8392 | maternal care for abn fetal hrt rate or rhythm, unspec tri, fetus 2 |
| DX_ICD10 | O36.8393 | maternal care for abn fetal hrt rate or rhythm, unspec tri, fetus 3 |
| DX_ICD10 | O36.8394 | maternal care for abn fetal hrt rate or rhythm, unspec tri, fetus 4 |
| DX_ICD10 | O36.8395 | maternal care for abn fetal hrt rate or rhythm, unspec tri, fetus 5 |
| DX_ICD10 | O36.8399 | maternal care for abn fetal hrt rate or rhythm, unspec tri, oth fetus |
| DX_ICD10 | O36.89 | maternal care for oth specif fetal problems |
| DX_ICD10 | O36.891 | maternal care for oth specif fetal problems, first tri |
| DX_ICD10 | O36.8910 | maternal care for oth specif fetal problems, first tri, na or unspec |
| DX_ICD10 | O36.8911 | maternal care for oth specif fetal problems, first tri, fetus 1 |
| DX_ICD10 | O36.8912 | maternal care for oth specif fetal problems, first tri, fetus 2 |
| DX_ICD10 | O36.8913 | maternal care for oth specif fetal problems, first tri, fetus 3 |
| DX_ICD10 | O36.8914 | maternal care for oth specif fetal problems, first tri, fetus 4 |
| DX_ICD10 | O36.8915 | maternal care for oth specif fetal problems, first tri, fetus 5 |
| DX_ICD10 | O36.8919 | maternal care for oth specif fetal problems, first tri, oth fetus |
| DX_ICD10 | O36.892 | maternal care for oth specif fetal problems, second tri |
| DX_ICD10 | O36.8920 | maternal care for oth spec fetal problems, second tri, na or unspec |
| DX_ICD10 | O36.8921 | maternal care for oth specif fetal problems, second tri, fetus 1 |
| DX_ICD10 | O36.8922 | maternal care for oth specif fetal problems, second tri, fetus 2 |
| DX_ICD10 | O36.8923 | maternal care for oth specif fetal problems, second tri, fetus 3 |
| DX_ICD10 | O36.8924 | maternal care for oth specif fetal problems, second tri, fetus 4 |
| DX_ICD10 | O36.8925 | maternal care for oth specif fetal problems, second tri, fetus 5 |
| DX_ICD10 | O36.8929 | maternal care for oth specif fetal problems, second tri, oth fetus |
| DX_ICD10 | O36.893 | maternal care for oth specif fetal problems, third tri |
| DX_ICD10 | O36.8930 | maternal care for oth specif fetal problems, third tri, na or unspec |
| DX_ICD10 | O36.8931 | maternal care for oth specif fetal problems, third tri, fetus 1 |
| DX_ICD10 | O36.8932 | maternal care for oth specif fetal problems, third tri, fetus 2 |
| DX_ICD10 | O36.8933 | maternal care for oth specif fetal problems, third tri, fetus 3 |
| DX_ICD10 | O36.8934 | maternal care for oth specif fetal problems, third tri, fetus 4 |
| DX_ICD10 | O36.8935 | maternal care for oth specif fetal problems, third tri, fetus 5 |
| DX_ICD10 | O36.8939 | maternal care for oth specif fetal problems, third tri, oth fetus |
| DX_ICD10 | O36.899 | maternal care for oth specif fetal problems, unspec tri |
| DX_ICD10 | O36.8990 | maternal care for oth specif fetal problems, unspec tri, na or unspec |
| DX_ICD10 | O36.8991 | maternal care for oth specif fetal problems, unspec tri, fetus 1 |
| DX_ICD10 | O36.8992 | maternal care for oth specif fetal problems, unspec tri, fetus 2 |
| DX_ICD10 | O36.8993 | maternal care for oth specif fetal problems, unspec tri, fetus 3 |
| DX_ICD10 | O36.8994 | maternal care for oth specif fetal problems, unspec tri, fetus 4 |
| DX_ICD10 | O36.8995 | maternal care for oth specif fetal problems, unspec tri, fetus 5 |
| DX_ICD10 | O36.8999 | maternal care for oth specif fetal problems, unspec tri, oth fetus |
| DX_ICD10 | O36.9 | maternal care for fetal problem, unspec |
| DX_ICD10 | O36.90 | maternal care for fetal problem, unspec, unspec tri |
| DX_ICD10 | O36.90X0 | maternal care for fetal problem, unspec, unspec tri, na or unspec |
| DX_ICD10 | O36.90X1 | maternal care for fetal problem, unspec, unspec tri, fetus 1 |
| DX_ICD10 | O36.90X2 | maternal care for fetal problem, unspec, unspec tri, fetus 2 |
| DX_ICD10 | O36.90X3 | maternal care for fetal problem, unspec, unspec tri, fetus 3 |
| DX_ICD10 | O36.90X4 | maternal care for fetal problem, unspec, unspec tri, fetus 4 |
| DX_ICD10 | O36.90X5 | maternal care for fetal problem, unspec, unspec tri, fetus 5 |
| DX_ICD10 | O36.90X9 | maternal care for fetal problem, unspec, unspec tri, oth fetus |
| DX_ICD10 | O36.91 | maternal care for fetal problem, unspec, first tri |
| DX_ICD10 | O36.91X0 | maternal care for fetal problem, unspec, first tri, na or unspec |
| DX_ICD10 | O36.91X1 | maternal care for fetal problem, unspec, first tri, fetus 1 |
| DX_ICD10 | O36.91X2 | maternal care for fetal problem, unspec, first tri, fetus 2 |
| DX_ICD10 | O36.91X3 | maternal care for fetal problem, unspec, first tri, fetus 3 |
| DX_ICD10 | O36.91X4 | maternal care for fetal problem, unspec, first tri, fetus 4 |
| DX_ICD10 | O36.91X5 | maternal care for fetal problem, unspec, first tri, fetus 5 |
| DX_ICD10 | O36.91X9 | maternal care for fetal problem, unspec, first tri, oth fetus |
| DX_ICD10 | O36.92 | maternal care for fetal problem, unspec, second tri |
| DX_ICD10 | O36.92X0 | maternal care for fetal problem, unspec, second tri, na or unspec |
| DX_ICD10 | O36.92X1 | maternal care for fetal problem, unspec, second tri, fetus 1 |
| DX_ICD10 | O36.92X2 | maternal care for fetal problem, unspec, second tri, fetus 2 |
| DX_ICD10 | O36.92X3 | maternal care for fetal problem, unspec, second tri, fetus 3 |
| DX_ICD10 | O36.92X4 | maternal care for fetal problem, unspec, second tri, fetus 4 |
| DX_ICD10 | O36.92X5 | maternal care for fetal problem, unspec, second tri, fetus 5 |
| DX_ICD10 | O36.92X9 | maternal care for fetal problem, unspec, second tri, oth fetus |
| DX_ICD10 | O36.93 | maternal care for fetal problem, unspec, third tri |
| DX_ICD10 | O36.93X0 | maternal care for fetal problem, unspec, third tri, na or unspec |
| DX_ICD10 | O36.93X1 | maternal care for fetal problem, unspec, third tri, fetus 1 |
| DX_ICD10 | O36.93X2 | maternal care for fetal problem, unspec, third tri, fetus 2 |
| DX_ICD10 | O36.93X3 | maternal care for fetal problem, unspec, third tri, fetus 3 |
| DX_ICD10 | O36.93X4 | maternal care for fetal problem, unspec, third tri, fetus 4 |
| DX_ICD10 | O36.93X5 | maternal care for fetal problem, unspec, third tri, fetus 5 |
| DX_ICD10 | O36.93X9 | maternal care for fetal problem, unspec, third tri, oth fetus |
| DX_ICD10 | O40 | polyhydramnios |
| DX_ICD10 | O40.1 | polyhydramnios, first tri |
| DX_ICD10 | O40.1XX0 | polyhydramnios, first tri, na or unspec |
| DX_ICD10 | O40.1XX1 | polyhydramnios, first tri, fetus 1 |
| DX_ICD10 | O40.1XX2 | polyhydramnios, first tri, fetus 2 |
| DX_ICD10 | O40.1XX3 | polyhydramnios, first tri, fetus 3 |
| DX_ICD10 | O40.1XX4 | polyhydramnios, first tri, fetus 4 |
| DX_ICD10 | O40.1XX5 | polyhydramnios, first tri, fetus 5 |
| DX_ICD10 | O40.1XX9 | polyhydramnios, first tri, oth fetus |
| DX_ICD10 | O40.2 | polyhydramnios, second tri |
| DX_ICD10 | O40.2XX0 | polyhydramnios, second tri, na or unspec |
| DX_ICD10 | O40.2XX1 | polyhydramnios, second tri, fetus 1 |
| DX_ICD10 | O40.2XX2 | polyhydramnios, second tri, fetus 2 |
| DX_ICD10 | O40.2XX3 | polyhydramnios, second tri, fetus 3 |
| DX_ICD10 | O40.2XX4 | polyhydramnios, second tri, fetus 4 |
| DX_ICD10 | O40.2XX5 | polyhydramnios, second tri, fetus 5 |
| DX_ICD10 | O40.2XX9 | polyhydramnios, second tri, oth fetus |
| DX_ICD10 | O40.3 | polyhydramnios, third tri |
| DX_ICD10 | O40.3XX0 | polyhydramnios, third tri, na or unspec |
| DX_ICD10 | O40.3XX1 | polyhydramnios, third tri, fetus 1 |
| DX_ICD10 | O40.3XX2 | polyhydramnios, third tri, fetus 2 |
| DX_ICD10 | O40.3XX3 | polyhydramnios, third tri, fetus 3 |
| DX_ICD10 | O40.3XX4 | polyhydramnios, third tri, fetus 4 |
| DX_ICD10 | O40.3XX5 | polyhydramnios, third tri, fetus 5 |
| DX_ICD10 | O40.3XX9 | polyhydramnios, third tri, oth fetus |
| DX_ICD10 | O40.9 | polyhydramnios, unspec tri |
| DX_ICD10 | O40.9XX0 | polyhydramnios, unspec tri, na or unspec |
| DX_ICD10 | O40.9XX1 | polyhydramnios, unspec tri, fetus 1 |
| DX_ICD10 | O40.9XX2 | polyhydramnios, unspec tri, fetus 2 |
| DX_ICD10 | O40.9XX3 | polyhydramnios, unspec tri, fetus 3 |
| DX_ICD10 | O40.9XX4 | polyhydramnios, unspec tri, fetus 4 |
| DX_ICD10 | O40.9XX5 | polyhydramnios, unspec tri, fetus 5 |
| DX_ICD10 | O40.9XX9 | polyhydramnios, unspec tri, oth fetus |
| DX_ICD10 | O41 | oth disorders of amniotic fluid & membranes |
| DX_ICD10 | O41.0 | oligohydramnios |
| DX_ICD10 | O41.00 | oligohydramnios, unspec tri |
| DX_ICD10 | O41.00X0 | oligohydramnios, unspec tri, na or unspec |
| DX_ICD10 | O41.00X1 | oligohydramnios, unspec tri, fetus 1 |
| DX_ICD10 | O41.00X2 | oligohydramnios, unspec tri, fetus 2 |
| DX_ICD10 | O41.00X3 | oligohydramnios, unspec tri, fetus 3 |
| DX_ICD10 | O41.00X4 | oligohydramnios, unspec tri, fetus 4 |
| DX_ICD10 | O41.00X5 | oligohydramnios, unspec tri, fetus 5 |
| DX_ICD10 | O41.00X9 | oligohydramnios, unspec tri, oth fetus |
| DX_ICD10 | O41.01 | oligohydramnios, first tri |
| DX_ICD10 | O41.01X0 | oligohydramnios, first tri, na or unspec |
| DX_ICD10 | O41.01X1 | oligohydramnios, first tri, fetus 1 |
| DX_ICD10 | O41.01X2 | oligohydramnios, first tri, fetus 2 |
| DX_ICD10 | O41.01X3 | oligohydramnios, first tri, fetus 3 |
| DX_ICD10 | O41.01X4 | oligohydramnios, first tri, fetus 4 |
| DX_ICD10 | O41.01X5 | oligohydramnios, first tri, fetus 5 |
| DX_ICD10 | O41.01X9 | oligohydramnios, first tri, oth fetus |
| DX_ICD10 | O41.02 | oligohydramnios, second tri |
| DX_ICD10 | O41.02X0 | oligohydramnios, second tri, na or unspec |
| DX_ICD10 | O41.02X1 | oligohydramnios, second tri, fetus 1 |
| DX_ICD10 | O41.02X2 | oligohydramnios, second tri, fetus 2 |
| DX_ICD10 | O41.02X3 | oligohydramnios, second tri, fetus 3 |
| DX_ICD10 | O41.02X4 | oligohydramnios, second tri, fetus 4 |
| DX_ICD10 | O41.02X5 | oligohydramnios, second tri, fetus 5 |
| DX_ICD10 | O41.02X9 | oligohydramnios, second tri, oth fetus |
| DX_ICD10 | O41.03 | oligohydramnios, third tri |
| DX_ICD10 | O41.03X0 | oligohydramnios, third tri, na or unspec |
| DX_ICD10 | O41.03X1 | oligohydramnios, third tri, fetus 1 |
| DX_ICD10 | O41.03X2 | oligohydramnios, third tri, fetus 2 |
| DX_ICD10 | O41.03X3 | oligohydramnios, third tri, fetus 3 |
| DX_ICD10 | O41.03X4 | oligohydramnios, third tri, fetus 4 |
| DX_ICD10 | O41.03X5 | oligohydramnios, third tri, fetus 5 |
| DX_ICD10 | O41.03X9 | oligohydramnios, third tri, oth fetus |
| DX_ICD10 | O41.1 | infection of amnio sac & membranes |
| DX_ICD10 | O41.10 | infection of amnio sac & membranes, unspec |
| DX_ICD10 | O41.101 | infection of amnio sac & membranes, unspec, first tri |
| DX_ICD10 | O41.1010 | infection of amnio sac & membranes, unspec, first tri, na or unspec |
| DX_ICD10 | O41.1011 | infection of amnio sac & membranes, unspec, first tri, fetus 1 |
| DX_ICD10 | O41.1012 | infection of amnio sac & membranes, unspec, first tri, fetus 2 |
| DX_ICD10 | O41.1013 | infection of amnio sac & membranes, unspec, first tri, fetus 3 |
| DX_ICD10 | O41.1014 | infection of amnio sac & membranes, unspec, first tri, fetus 4 |
| DX_ICD10 | O41.1015 | infection of amnio sac & membranes, unspec, first tri, fetus 5 |
| DX_ICD10 | O41.1019 | infection of amnio sac & membranes, unspec, first tri, oth fetus |
| DX_ICD10 | O41.102 | infection of amnio sac & membranes, unspec, second tri |
| DX_ICD10 | O41.1020 | infection of amnio sac & membranes, unspec, second tri, na or unspec |
| DX_ICD10 | O41.1021 | infection of amnio sac & membranes, unspec, second tri, fetus 1 |
| DX_ICD10 | O41.1022 | infection of amnio sac & membranes, unspec, second tri, fetus 2 |
| DX_ICD10 | O41.1023 | infection of amnio sac & membranes, unspec, second tri, fetus 3 |
| DX_ICD10 | O41.1024 | infection of amnio sac & membranes, unspec, second tri, fetus 4 |
| DX_ICD10 | O41.1025 | infection of amnio sac & membranes, unspec, second tri, fetus 5 |
| DX_ICD10 | O41.1029 | infection of amnio sac & membranes, unspec, second tri, oth fetus |
| DX_ICD10 | O41.103 | infection of amnio sac & membranes, unspec, third tri |
| DX_ICD10 | O41.1030 | infection of amnio sac & membranes, unspec, third tri, na or unspec |
| DX_ICD10 | O41.1031 | infection of amnio sac & membranes, unspec, third tri, fetus 1 |
| DX_ICD10 | O41.1032 | infection of amnio sac & membranes, unspec, third tri, fetus 2 |
| DX_ICD10 | O41.1033 | infection of amnio sac & membranes, unspec, third tri, fetus 3 |
| DX_ICD10 | O41.1034 | infection of amnio sac & membranes, unspec, third tri, fetus 4 |
| DX_ICD10 | O41.1035 | infection of amnio sac & membranes, unspec, third tri, fetus 5 |
| DX_ICD10 | O41.1039 | infection of amnio sac & membranes, unspec, third tri, oth fetus |
| DX_ICD10 | O41.109 | infection of amnio sac & membranes, unspec, unspec tri |
| DX_ICD10 | O41.1090 | infection of amnio sac & membranes, unspec, unspec tri, na or unspec |
| DX_ICD10 | O41.1091 | infection of amnio sac & membranes, unspec, unspec tri, fetus 1 |
| DX_ICD10 | O41.1092 | infection of amnio sac & membranes, unspec, unspec tri, fetus 2 |
| DX_ICD10 | O41.1093 | infection of amnio sac & membranes, unspec, unspec tri, fetus 3 |
| DX_ICD10 | O41.1094 | infection of amnio sac & membranes, unspec, unspec tri, fetus 4 |
| DX_ICD10 | O41.1095 | infection of amnio sac & membranes, unspec, unspec tri, fetus 5 |
| DX_ICD10 | O41.1099 | infection of amnio sac & membranes, unspec, unspec tri, oth fetus |
| DX_ICD10 | O41.12 | chorioamnionitis |
| DX_ICD10 | O41.121 | chorioamnionitis, first tri |
| DX_ICD10 | O41.1210 | chorioamnionitis, first tri, na or unspec |
| DX_ICD10 | O41.1211 | chorioamnionitis, first tri, fetus 1 |
| DX_ICD10 | O41.1212 | chorioamnionitis, first tri, fetus 2 |
| DX_ICD10 | O41.1213 | chorioamnionitis, first tri, fetus 3 |
| DX_ICD10 | O41.1214 | chorioamnionitis, first tri, fetus 4 |
| DX_ICD10 | O41.1215 | chorioamnionitis, first tri, fetus 5 |
| DX_ICD10 | O41.1219 | chorioamnionitis, first tri, oth fetus |
| DX_ICD10 | O41.122 | chorioamnionitis, second tri |
| DX_ICD10 | O41.1220 | chorioamnionitis, second tri, na or unspec |
| DX_ICD10 | O41.1221 | chorioamnionitis, second tri, fetus 1 |
| DX_ICD10 | O41.1222 | chorioamnionitis, second tri, fetus 2 |
| DX_ICD10 | O41.1223 | chorioamnionitis, second tri, fetus 3 |
| DX_ICD10 | O41.1224 | chorioamnionitis, second tri, fetus 4 |
| DX_ICD10 | O41.1225 | chorioamnionitis, second tri, fetus 5 |
| DX_ICD10 | O41.1229 | chorioamnionitis, second tri, oth fetus |
| DX_ICD10 | O41.123 | chorioamnionitis, second tri |
| DX_ICD10 | O41.1230 | chorioamnionitis, third tri, na or unspec |
| DX_ICD10 | O41.1231 | chorioamnionitis, third tri, fetus 1 |
| DX_ICD10 | O41.1232 | chorioamnionitis, third tri, fetus 2 |
| DX_ICD10 | O41.1233 | chorioamnionitis, third tri, fetus 3 |
| DX_ICD10 | O41.1234 | chorioamnionitis, third tri, fetus 4 |
| DX_ICD10 | O41.1235 | chorioamnionitis, third tri, fetus 5 |
| DX_ICD10 | O41.1239 | chorioamnionitis, third tri, oth fetus |
| DX_ICD10 | O41.129 | chorioamnionitis, unspec tri |
| DX_ICD10 | O41.1290 | chorioamnionitis, unspec tri, na or unspec |
| DX_ICD10 | O41.1291 | chorioamnionitis, unspec tri, fetus 1 |
| DX_ICD10 | O41.1292 | chorioamnionitis, unspec tri, fetus 2 |
| DX_ICD10 | O41.1293 | chorioamnionitis, unspec tri, fetus 3 |
| DX_ICD10 | O41.1294 | chorioamnionitis, unspec tri, fetus 4 |
| DX_ICD10 | O41.1295 | chorioamnionitis, unspec tri, fetus 5 |
| DX_ICD10 | O41.1299 | chorioamnionitis, unspec tri, oth fetus |
| DX_ICD10 | O41.14 | placentitis |
| DX_ICD10 | O41.141 | placentitis, first tri |
| DX_ICD10 | O41.1410 | placentitis, first tri, na or unspec |
| DX_ICD10 | O41.1411 | placentitis, first tri, fetus 1 |
| DX_ICD10 | O41.1412 | placentitis, first tri, fetus 2 |
| DX_ICD10 | O41.1413 | placentitis, first tri, fetus 3 |
| DX_ICD10 | O41.1414 | placentitis, first tri, fetus 4 |
| DX_ICD10 | O41.1415 | placentitis, first tri, fetus 5 |
| DX_ICD10 | O41.1419 | placentitis, first tri, oth fetus |
| DX_ICD10 | O41.142 | placentitis, second tri |
| DX_ICD10 | O41.1420 | placentitis, second tri, na or unspec |
| DX_ICD10 | O41.1421 | placentitis, second tri, fetus 1 |
| DX_ICD10 | O41.1422 | placentitis, second tri, fetus 2 |
| DX_ICD10 | O41.1423 | placentitis, second tri, fetus 3 |
| DX_ICD10 | O41.1424 | placentitis, second tri, fetus 4 |
| DX_ICD10 | O41.1425 | placentitis, second tri, fetus 5 |
| DX_ICD10 | O41.1429 | placentitis, second tri, oth fetus |
| DX_ICD10 | O41.143 | placentitis, third tri |
| DX_ICD10 | O41.1430 | placentitis, third tri, na or unspec |
| DX_ICD10 | O41.1431 | placentitis, third tri, fetus 1 |
| DX_ICD10 | O41.1432 | placentitis, third tri, fetus 2 |
| DX_ICD10 | O41.1433 | placentitis, third tri, fetus 3 |
| DX_ICD10 | O41.1434 | placentitis, third tri, fetus 4 |
| DX_ICD10 | O41.1435 | placentitis, third tri, fetus 5 |
| DX_ICD10 | O41.1439 | placentitis, third tri, oth fetus |
| DX_ICD10 | O41.149 | placentitis, unspec tri |
| DX_ICD10 | O41.1490 | placentitis, unspec tri, na or unspec |
| DX_ICD10 | O41.1491 | placentitis, unspec tri, fetus 1 |
| DX_ICD10 | O41.1492 | placentitis, unspec tri, fetus 2 |
| DX_ICD10 | O41.1493 | placentitis, unspec tri, fetus 3 |
| DX_ICD10 | O41.1494 | placentitis, unspec tri, fetus 4 |
| DX_ICD10 | O41.1495 | placentitis, unspec tri, fetus 5 |
| DX_ICD10 | O41.1499 | placentitis, unspec tri, oth fetus |
| DX_ICD10 | O41.8 | oth specif disorders of amniotic fluid & membranes |
| DX_ICD10 | O41.8X | oth specif disorders of amniotic fluid & membranes |
| DX_ICD10 | O41.8X1 | oth specif disorders of amniotic fluid & membranes, first tri |
| DX_ICD10 | O41.8X10 | oth specif disorders of amnio fluid & membranes, first tri, na or unspec |
| DX_ICD10 | O41.8X11 | oth specif disorders of amniotic fluid & membranes, first tri, fetus 1 |
| DX_ICD10 | O41.8X12 | oth specif disorders of amniotic fluid & membranes, first tri, fetus 2 |
| DX_ICD10 | O41.8X13 | oth specif disorders of amniotic fluid & membranes, first tri, fetus 3 |
| DX_ICD10 | O41.8X14 | oth specif disorders of amniotic fluid & membranes, first tri, fetus 4 |
| DX_ICD10 | O41.8X15 | oth specif disorders of amniotic fluid & membranes, first tri, fetus 5 |
| DX_ICD10 | O41.8X19 | oth specif disorders of amniotic fluid & membranes, first tri, oth fetus |
| DX_ICD10 | O41.8X2 | oth specif disorders of amniotic fluid & membranes, second tri |
| DX_ICD10 | O41.8X20 | oth specif dis of amniotic fluid & membranes, second tri, na or unspec |
| DX_ICD10 | O41.8X21 | oth specif disorders of amniotic fluid & membranes, second tri, fetus 1 |
| DX_ICD10 | O41.8X22 | oth specif disorders of amniotic fluid & membranes, second tri, fetus 2 |
| DX_ICD10 | O41.8X23 | oth specif disorders of amniotic fluid & membranes, second tri, fetus 3 |
| DX_ICD10 | O41.8X24 | oth specif disorders of amniotic fluid & membranes, second tri, fetus 4 |
| DX_ICD10 | O41.8X25 | oth specif disorders of amniotic fluid & membranes, second tri, fetus 5 |
| DX_ICD10 | O41.8X29 | oth specif disorders of amnio fluid & membranes, second tri, oth fetus |
| DX_ICD10 | O41.8X3 | oth specif disorders of amniotic fluid & membranes, third tri |
| DX_ICD10 | O41.8X30 | oth specif disorders of amnio fluid & membr, third tri, na or unspec |
| DX_ICD10 | O41.8X31 | oth specif disorders of amniotic fluid & membranes, third tri, fetus 1 |
| DX_ICD10 | O41.8X32 | oth specif disorders of amniotic fluid & membranes, third tri, fetus 2 |
| DX_ICD10 | O41.8X33 | oth specif disorders of amniotic fluid & membranes, third tri, fetus 3 |
| DX_ICD10 | O41.8X34 | oth specif disorders of amniotic fluid & membranes, third tri, fetus 4 |
| DX_ICD10 | O41.8X35 | oth specif disorders of amniotic fluid & membranes, third tri, fetus 5 |
| DX_ICD10 | O41.8X39 | oth specif disorders of amniotic fluid & membranes, third tri, oth fetus |
| DX_ICD10 | O41.8X9 | oth specif disorders of amniotic fluid & membranes, unspec tri |
| DX_ICD10 | O41.8X90 | oth specif dis of amniotic fluid & membranes, unspec tri, na or unspec |
| DX_ICD10 | O41.8X91 | oth specif disorders of amniotic fluid & membranes, unspec tri, fetus 1 |
| DX_ICD10 | O41.8X92 | oth specif disorders of amniotic fluid & membranes, unspec tri, fetus 2 |
| DX_ICD10 | O41.8X93 | oth specif disorders of amniotic fluid & membranes, unspec tri, fetus 3 |
| DX_ICD10 | O41.8X94 | oth specif disorders of amniotic fluid & membranes, unspec tri, fetus 4 |
| DX_ICD10 | O41.8X95 | oth specif disorders of amniotic fluid & membranes, unspec tri, fetus 5 |
| DX_ICD10 | O41.8X99 | oth specif disorders of amnio fluid & membran, unspec tri, oth fetus |
| DX_ICD10 | O41.9 | disorder of amniotic fluid & membranes, unspec |
| DX_ICD10 | O41.90 | disorder of amniotic fluid & membranes, unspec, unspec tri |
| DX_ICD10 | O41.90X0 | disorder of amnio fluid & membran, unspec, unspec tri, na or unspec |
| DX_ICD10 | O41.90X1 | disorder of amniotic fluid & membranes, unspec, unspec tri, fetus 1 |
| DX_ICD10 | O41.90X2 | disorder of amniotic fluid & membranes, unspec, unspec tri, fetus 2 |
| DX_ICD10 | O41.90X3 | disorder of amniotic fluid & membranes, unspec, unspec tri, fetus 3 |
| DX_ICD10 | O41.90X4 | disorder of amniotic fluid & membranes, unspec, unspec tri, fetus 4 |
| DX_ICD10 | O41.90X5 | disorder of amniotic fluid & membranes, unspec, unspec tri, fetus 5 |
| DX_ICD10 | O41.90X9 | disorder of amniotic fluid & membranes, unspec, unspec tri, oth fetus |
| DX_ICD10 | O41.91 | disorder of amniotic fluid & membranes, unspec, first tri |
| DX_ICD10 | O41.91X0 | disorder of amniotic fluid & membranes, unspec, first tri, na or unspec |
| DX_ICD10 | O41.91X1 | disorder of amniotic fluid & membranes, unspec, first tri, fetus 1 |
| DX_ICD10 | O41.91X2 | disorder of amniotic fluid & membranes, unspec, first tri, fetus 2 |
| DX_ICD10 | O41.91X3 | disorder of amniotic fluid & membranes, unspec, first tri, fetus 3 |
| DX_ICD10 | O41.91X4 | disorder of amniotic fluid & membranes, unspec, first tri, fetus 4 |
| DX_ICD10 | O41.91X5 | disorder of amniotic fluid & membranes, unspec, first tri, fetus 5 |
| DX_ICD10 | O41.91X9 | disorder of amniotic fluid & membranes, unspec, first tri, oth fetus |
| DX_ICD10 | O41.92 | disorder of amniotic fluid & membranes, unspec, second tri |
| DX_ICD10 | O41.92X0 | disorder of amnio fluid & membranes, unspec, second tri, na or unspec |
| DX_ICD10 | O41.92X1 | disorder of amniotic fluid & membranes, unspec, second tri, fetus 1 |
| DX_ICD10 | O41.92X2 | disorder of amniotic fluid & membranes, unspec, second tri, fetus 2 |
| DX_ICD10 | O41.92X3 | disorder of amniotic fluid & membranes, unspec, second tri, fetus 3 |
| DX_ICD10 | O41.92X4 | disorder of amniotic fluid & membranes, unspec, second tri, fetus 4 |
| DX_ICD10 | O41.92X5 | disorder of amniotic fluid & membranes, unspec, second tri, fetus 5 |
| DX_ICD10 | O41.92X9 | disorder of amniotic fluid & membranes, unspec, second tri, oth fetus |
| DX_ICD10 | O41.93 | disorder of amniotic fluid & membranes, unspec, third tri |
| DX_ICD10 | O41.93X0 | disorder of amniotic fluid & membranes, unspec, third tri, na or unspec |
| DX_ICD10 | O41.93X1 | disorder of amniotic fluid & membranes, unspec, third tri, fetus 1 |
| DX_ICD10 | O41.93X2 | disorder of amniotic fluid & membranes, unspec, third tri, fetus 2 |
| DX_ICD10 | O41.93X3 | disorder of amniotic fluid & membranes, unspec, third tri, fetus 3 |
| DX_ICD10 | O41.93X4 | disorder of amniotic fluid & membranes, unspec, third tri, fetus 4 |
| DX_ICD10 | O41.93X5 | disorder of amniotic fluid & membranes, unspec, third tri, fetus 5 |
| DX_ICD10 | O41.93X9 | disorder of amniotic fluid & membranes, unspec, third tri, oth fetus |
| DX_ICD10 | O42 | premature rupt of membranes |
| DX_ICD10 | O42.0 | premature rupt of membran, onset of labor w/in 24 hrs of rupt |
| DX_ICD10 | O42.00 | premat rupt membran, onset labor w/in 24 hrs rupt, unspec wk gest |
| DX_ICD10 | O42.01 | preterm premat rupt membran, onset labor w/in 24 hrs rupt) |
| DX_ICD10 | O42.011 | preterm premat rupt membran, onset labor w/in 24 hrs rupt, first tri |
| DX_ICD10 | O42.012 | preterm premat rupt membran, onset labor w/in 24 hrs rupt, second tri |
| DX_ICD10 | O42.013 | preterm premat rupt membran, onset labor w/in 24 hrs rupt, third tri |
| DX_ICD10 | O42.019 | preterm premat rupt membran, onset labor w/in 24 hrs rupt, unspec tri |
| DX_ICD10 | O42.02 | full-term premat rupt membran, onset labor w/in 24 hrs rupt |
| DX_ICD10 | O42.1 | premat rupt membrane, onset labor 24+ hrs after rupt |
| DX_ICD10 | O42.10 | premat rupt membran, onset lab 24+ hrs after rupt, unspec wks gest |
| DX_ICD10 | O42.11 | pt premat rupt membran, onset labor 24+ hrs after rupt, first tri) |
| DX_ICD10 | O42.111 | pt premat rupt membran, onset labor 24+ hrs after rupt, first tri |
| DX_ICD10 | O42.112 | pt premat rupt membran, onset labor 24+ hrs after rupt, second tri |
| DX_ICD10 | O42.113 | pt premat rupt membran, onset labor 24+ hrs after rupt, third tri |
| DX_ICD10 | O42.119 | pt premat rupt membran, onset labor 24+ hrs after rupt, unspec tri |
| DX_ICD10 | O42.12 | full-term premat rupt membran, onset labor 24+ hrs after rupt |
| DX_ICD10 | O42.9 | premat rupt membran, unspec length rupt & onset labor |
| DX_ICD10 | O42.90 | premat rupt memb, unspec len rupt & lab, unspec wks gest |
| DX_ICD10 | O42.91 | pt premat rupt of mem, unspec len rupt & labor) |
| DX_ICD10 | O42.911 | pt premat rupt mem, unspec len rupt & labor, unspec wks gest, first tri |
| DX_ICD10 | O42.912 | pt prem rupt mem, unspec len rupt & labor, unspec wks gest,sec tri |
| DX_ICD10 | O42.913 | pt prem rupt mem, unspec len rupt & labor, unspec wks gest, third tri |
| DX_ICD10 | O42.919 | pt prem rupt mem, unspec len rupt & lab, unspec wks gest, unspec tri |
| DX_ICD10 | O42.92 | ft prem rupt membran, unspec length rupt & labor |
| DX_ICD10 | O43 | plcntal dis |
| DX_ICD10 | O43.0 | placental transfusion syndromes |
| DX_ICD10 | O43.01 | fetomaternal placental transfusion syndrome |
| DX_ICD10 | O43.011 | fetomaternal placental transfusion syndrome, first tri |
| DX_ICD10 | O43.012 | fetomaternal placental transfusion syndrome, second tri |
| DX_ICD10 | O43.013 | fetomaternal placental transfusion syndrome, third tri |
| DX_ICD10 | O43.019 | fetomaternal placental transfusion syndrome, unspec tri |
| DX_ICD10 | O43.02 | fetus-to-fetus placental transfusion syndrome |
| DX_ICD10 | O43.021 | fetus-to-fetus placental transfusion syndrome, first tri |
| DX_ICD10 | O43.022 | fetus-to-fetus placental transfusion syndrome, second tri |
| DX_ICD10 | O43.023 | fetus-to-fetus placental transfusion syndrome, third tri |
| DX_ICD10 | O43.029 | fetus-to-fetus placental transfusion syndrome, unspec tri |
| DX_ICD10 | O43.1 | malformation of placenta |
| DX_ICD10 | O43.10 | malformation of placenta, unspec |
| DX_ICD10 | O43.101 | malformation of placenta, unspec, first tri |
| DX_ICD10 | O43.102 | malformation of placenta, unspec, second tri |
| DX_ICD10 | O43.103 | malformation of placenta, unspec, third tri |
| DX_ICD10 | O43.109 | malformation of placenta, unspec, unspec tri |
| DX_ICD10 | O43.11 | circumvallate placenta |
| DX_ICD10 | O43.111 | circumvallate placenta, first tri |
| DX_ICD10 | O43.112 | circumvallate placenta, second tri |
| DX_ICD10 | O43.113 | circumvallate placenta, third tri |
| DX_ICD10 | O43.119 | circumvallate placenta, unspec tri |
| DX_ICD10 | O43.12 | velamentous insertion of umbilical cord |
| DX_ICD10 | O43.121 | velamentous insertion of umbilical cord, first tri |
| DX_ICD10 | O43.122 | velamentous insertion of umbilical cord, second tri |
| DX_ICD10 | O43.123 | velamentous insertion of umbilical cord, third tri |
| DX_ICD10 | O43.129 | velamentous insertion of umbilical cord, unspec tri |
| DX_ICD10 | O43.19 | oth malformation of placenta |
| DX_ICD10 | O43.191 | oth malformation of placenta, first tri |
| DX_ICD10 | O43.192 | oth malformation of placenta, second tri |
| DX_ICD10 | O43.193 | oth malformation of placenta, third tri |
| DX_ICD10 | O43.199 | oth malformation of placenta, unspec tri |
| DX_ICD10 | O43.2 | morbidly adherent placenta |
| DX_ICD10 | O43.21 | plcnta accreta |
| DX_ICD10 | O43.211 | plcnta accreta, first tri |
| DX_ICD10 | O43.212 | plcnta accreta, second tri |
| DX_ICD10 | O43.213 | plcnta accreta, third tri |
| DX_ICD10 | O43.219 | plcnta accreta, unspec tri |
| DX_ICD10 | O43.22 | plcnta increta |
| DX_ICD10 | O43.221 | plcnta increta, first tri |
| DX_ICD10 | O43.222 | plcnta increta, second tri |
| DX_ICD10 | O43.223 | plcnta increta, third tri |
| DX_ICD10 | O43.229 | plcnta increta, unspec tri |
| DX_ICD10 | O43.23 | plcnta percreta |
| DX_ICD10 | O43.231 | plcnta percreta, first tri |
| DX_ICD10 | O43.232 | plcnta percreta, second tri |
| DX_ICD10 | O43.233 | plcnta percreta, third tri |
| DX_ICD10 | O43.239 | plcnta percreta, unspec tri |
| DX_ICD10 | O43.8 | oth placental disorders |
| DX_ICD10 | O43.81 | placental infarction |
| DX_ICD10 | O43.811 | placental infarction, first tri |
| DX_ICD10 | O43.812 | placental infarction, second tri |
| DX_ICD10 | O43.813 | placental infarction, third tri |
| DX_ICD10 | O43.819 | placental infarction, unspec tri |
| DX_ICD10 | O43.89 | oth placental disorders |
| DX_ICD10 | O43.891 | oth placental disorders, first tri |
| DX_ICD10 | O43.892 | oth placental disorders, second tri |
| DX_ICD10 | O43.893 | oth placental disorders, third tri |
| DX_ICD10 | O43.899 | oth placental disorders, unspec tri |
| DX_ICD10 | O43.9 | unspec placental disorder |
| DX_ICD10 | O43.90 | unspec placental disorder, unspec tri |
| DX_ICD10 | O43.91 | unspec placental disorder, first tri |
| DX_ICD10 | O43.92 | unspec placental disorder, second tri |
| DX_ICD10 | O43.93 | unspec placental disorder, third tri |
| DX_ICD10 | O44 | plcnta previa |
| DX_ICD10 | O44.0 | plcnta previa specif as wo hemorrhage |
| DX_ICD10 | O44.00 | complete plcnta previa nos or wo hemorrhage, unspec tri |
| DX_ICD10 | O44.01 | complete plcnta previa nos or wo hemorrhage, first tri |
| DX_ICD10 | O44.02 | complete plcnta previa nos or wo hemorrhage, second tri |
| DX_ICD10 | O44.03 | complete plcnta previa nos or wo hemorrhage, third tri |
| DX_ICD10 | O44.1 | plcnta previa with hemorrhage coagulation defect, unspec, second tri |
| DX_ICD10 | O44.10 | complete plcnta previa with hemorrhage, unspec tri |
| DX_ICD10 | O44.11 | complete plcnta previa with hemorrhage, first tri |
| DX_ICD10 | O44.12 | complete plcnta previa with hemorrhage, second tri |
| DX_ICD10 | O44.13 | complete plcnta previa with hemorrhage, third tri |
| DX_ICD10 | O44.20 | partial plcnta previa nos or wo hemorrhage, unspec tri |
| DX_ICD10 | O44.21 | partial plcnta previa nos or wo hemorrhage, first tri |
| DX_ICD10 | O44.22 | partial plcnta previa nos or wo hemorrhage, second tri |
| DX_ICD10 | O44.23 | partial plcnta previa nos or wo hemorrhage, third tri |
| DX_ICD10 | O44.30 | partial plcnta previa with hemorrhage, unspec tri |
| DX_ICD10 | O44.31 | partial plcnta previa with hemorrhage, first tri |
| DX_ICD10 | O44.32 | partial plcnta previa with hemorrhage, second tri |
| DX_ICD10 | O44.33 | partial plcnta previa with hemorrhage, third tri |
| DX_ICD10 | O44.40 | low lying plcnta nos or wo hemorrhage, unspec tri |
| DX_ICD10 | O44.41 | low lying plcnta nos or wo hemorrhage, first tri |
| DX_ICD10 | O44.42 | low lying plcnta nos or wo hemorrhage, second tri |
| DX_ICD10 | O44.43 | low lying plcnta nos or wo hemorrhage, third tri |
| DX_ICD10 | O44.50 | low lying plcnta with hemorrhage, unspec tri |
| DX_ICD10 | O44.51 | low lying plcnta with hemorrhage, first tri |
| DX_ICD10 | O44.52 | low lying plcnta with hemorrhage, second tri |
| DX_ICD10 | O44.53 | low lying plcnta with hemorrhage, third tri |
| DX_ICD10 | O45 | premat sep plcnta [abruptio placentae] |
| DX_ICD10 | O45.0 | premat sep plcnta with coagulation defect |
| DX_ICD10 | O45.00 | premat sep plcnta w coagulation defect, unsp |
| DX_ICD10 | O45.001 | premat sep plcnta with coagulation defect, unspec, first tri |
| DX_ICD10 | O45.002 | premat sep plcnta with coagulation defect, unspec, second tri |
| DX_ICD10 | O45.003 | premat sep plcnta with coagulation defect, unspec, third tri |
| DX_ICD10 | O45.009 | premat sep plcnta with coagulation defect, unspec, unspec tri |
| DX_ICD10 | O45.01 | premat sep plcnta with afibrinogenemia |
| DX_ICD10 | O45.011 | premat sep plcnta with afibrinogenemia, first tri |
| DX_ICD10 | O45.012 | premat sep plcnta with afibrinogenemia, second tri |
| DX_ICD10 | O45.013 | premat sep plcnta with afibrinogenemia, third tri |
| DX_ICD10 | O45.019 | premat sep plcnta with afibrinogenemia, unspec tri |
| DX_ICD10 | O45.02 | premat sep plcnta w dissem intravasc coagul |
| DX_ICD10 | O45.021 | premat sep plcnta w dissem intravasc coagul, first tri |
| DX_ICD10 | O45.022 | premat sep plcnta intravasc coagul, second tri |
| DX_ICD10 | O45.023 | premat sep plcnta w dissem intravasc coagul, third tri |
| DX_ICD10 | O45.029 | premat sep plcnta w dissem intravasc coagul, unspec tri |
| DX_ICD10 | O45.09 | premat sep plcnta with oth coagulation defect |
| DX_ICD10 | O45.091 | premat sep plcnta with oth coagulation defect, first tri |
| DX_ICD10 | O45.092 | premat sep plcnta with oth coagulation defect, second tri |
| DX_ICD10 | O45.093 | premat sep plcnta with oth coagulation defect, third tri |
| DX_ICD10 | O45.099 | premat sep plcnta with oth coagulation defect, unspec tri |
| DX_ICD10 | O45.8 | oth premat sep placenta |
| DX_ICD10 | O45.8X | oth premat sep placenta |
| DX_ICD10 | O45.8X1 | oth premat sep placenta, first tri |
| DX_ICD10 | O45.8X2 | oth premat sep placenta, second tri |
| DX_ICD10 | O45.8X3 | oth premat sep placenta, third tri |
| DX_ICD10 | O45.8X9 | oth premat sep placenta, unspec tri |
| DX_ICD10 | O45.9 | premat sep placenta, unspec abruptio placentae nos |
| DX_ICD10 | O45.90 | premat sep placenta, unspec, unspec tri |
| DX_ICD10 | O45.91 | premat sep placenta, unspec, first tri |
| DX_ICD10 | O45.92 | premat sep placenta, unspec, second tri |
| DX_ICD10 | O45.93 | premat sep placenta, unspec, third tri |
| DX_ICD10 | O46 | antepartum hemorrhage, not elsewhere classified |
| DX_ICD10 | O46.0 | antepartum hemorrhage with coagulation defect |
| DX_ICD10 | O46.00 | antepartum hemorrhage with coagulation defect, unspec |
| DX_ICD10 | O46.001 | antepartum hemorrhage with coagulation defect, unspec, first tri |
| DX_ICD10 | O46.002 | antepartum hemorrhage with coagulation defect, unspec, second tri |
| DX_ICD10 | O46.003 | antepartum hemorrhage with coagulation defect, unspec, third tri |
| DX_ICD10 | O46.009 | antepartum hemorrhage with coagulation defect, unspec, unspec tri |
| DX_ICD10 | O46.01 | antepartum hemorrhage with afibrinogenemia |
| DX_ICD10 | O46.011 | antepartum hemorrhage with afibrinogenemia, first tri |
| DX_ICD10 | O46.012 | antepartum hemorrhage with afibrinogenemia, second tri |
| DX_ICD10 | O46.013 | antepartum hemorrhage with afibrinogenemia, third tri |
| DX_ICD10 | O46.019 | antepartum hemorrhage with afibrinogenemia, unspec tri |
| DX_ICD10 | O46.02 | antepartum hemorrhage w dissem intravasc coagul |
| DX_ICD10 | O46.021 | antepartum hemorrhage w dissem intravasc coagul, first tri |
| DX_ICD10 | O46.022 | antepartum hemorrhage w dissem intravasc coagul, second tri |
| DX_ICD10 | O46.023 | antepartum hemorrhage w dissem intravasc coagul, third tri |
| DX_ICD10 | O46.029 | antepartum hemorrhage w dissem intravasc coagul, unspec tri |
| DX_ICD10 | O46.09 | antepartum hemorrhage with oth coagulation defect |
| DX_ICD10 | O46.091 | antepartum hemorrhage with oth coagulation defect, first tri |
| DX_ICD10 | O46.092 | antepartum hemorrhage with oth coagulation defect, second tri |
| DX_ICD10 | O46.093 | antepartum hemorrhage with oth coagulation defect, third tri |
| DX_ICD10 | O46.099 | antepartum hemorrhage with oth coagulation defect, unspec tri |
| DX_ICD10 | O46.8 | oth antepartum hemorrhage |
| DX_ICD10 | O46.8X | oth antepartum hemorrhage |
| DX_ICD10 | O46.8X1 | oth antepartum hemorrhage, first tri |
| DX_ICD10 | O46.8X2 | oth antepartum hemorrhage, second tri |
| DX_ICD10 | O46.8X3 | oth antepartum hemorrhage, third tri |
| DX_ICD10 | O46.8X9 | oth antepartum hemorrhage, unspec tri |
| DX_ICD10 | O46.9 | antepartum hemorrhage, unspec |
| DX_ICD10 | O46.90 | antepartum hemorrhage, unspec, unspec tri |
| DX_ICD10 | O46.91 | antepartum hemorrhage, unspec, first tri |
| DX_ICD10 | O46.92 | antepartum hemorrhage, unspec, second tri |
| DX_ICD10 | O46.93 | antepartum hemorrhage, unspec, third tri |
| DX_ICD10 | O47 | false labor |
| DX_ICD10 | O47.0 | false labor before 37 completed weeks of gest |
| DX_ICD10 | O47.00 | false labor before 37 completed weeks of gest, unspec tri |
| DX_ICD10 | O47.02 | false labor before 37 completed weeks of gest, second tri |
| DX_ICD10 | O47.03 | false labor before 37 completed weeks of gest, third tri |
| DX_ICD10 | O47.1 | false labor at or after 37 completed weeks of gest |
| DX_ICD10 | O47.9 | false labor, unspec |
| DX_ICD10 | O48 | late preg |
| DX_ICD10 | O48.0 | post-term preg |
| DX_ICD10 | O48.1 | prolonged preg |
| DX_ICD10 | O60 | preterm labor |
| DX_ICD10 | O60.0 | preterm labor wo delivery |
| DX_ICD10 | O60.00 | preterm labor wo delivery, unspec tri |
| DX_ICD10 | O60.02 | preterm labor wo delivery, second tri |
| DX_ICD10 | O60.03 | preterm labor wo delivery, third tri |
| DX_ICD10 | O60.1 | preterm labor with preterm delivery |
| DX_ICD10 | O60.10 | preterm labor with preterm delivery, unspec tri |
| DX_ICD10 | O60.10X0 | preterm labor with preterm delivery, unspec tri, na or unspec |
| DX_ICD10 | O60.10X1 | preterm labor with preterm delivery, unspec tri, fetus 1 |
| DX_ICD10 | O60.10X2 | preterm labor with preterm delivery, unspec tri, fetus 2 |
| DX_ICD10 | O60.10X3 | preterm labor with preterm delivery, unspec tri, fetus 3 |
| DX_ICD10 | O60.10X4 | preterm labor with preterm delivery, unspec tri, fetus 4 |
| DX_ICD10 | O60.10X5 | preterm labor with preterm delivery, unspec tri, fetus 5 |
| DX_ICD10 | O60.10X9 | preterm labor with preterm delivery, unspec tri, oth fetus |
| DX_ICD10 | O60.12 | preterm labor second tri with preterm delivery second tri |
| DX_ICD10 | O60.12X0 | preterm labor second tri with preterm delivery second tri, na or unspec |
| DX_ICD10 | O60.12X1 | preterm labor second tri with preterm delivery second tri, fetus 1 |
| DX_ICD10 | O60.12X2 | preterm labor second tri with preterm delivery second tri, fetus 2 |
| DX_ICD10 | O60.12X3 | preterm labor second tri with preterm delivery second tri, fetus 3 |
| DX_ICD10 | O60.12X4 | preterm labor second tri with preterm delivery second tri, fetus 4 |
| DX_ICD10 | O60.12X5 | preterm labor second tri with preterm delivery second tri, fetus 5 |
| DX_ICD10 | O60.12X9 | preterm labor second tri with preterm delivery second tri, oth fetus |
| DX_ICD10 | O60.13 | preterm labor second tri with preterm delivery third tri |
| DX_ICD10 | O60.13X0 | preterm labor second tri with preterm delivery third tri, na or unspec |
| DX_ICD10 | O60.13X1 | preterm labor second tri with preterm delivery third tri, fetus 1 |
| DX_ICD10 | O60.13X2 | preterm labor second tri with preterm delivery third tri, fetus 2 |
| DX_ICD10 | O60.13X3 | preterm labor second tri with preterm delivery third tri, fetus 3 |
| DX_ICD10 | O60.13X4 | preterm labor second tri with preterm delivery third tri, fetus 4 |
| DX_ICD10 | O60.13X5 | preterm labor second tri with preterm delivery third tri, fetus 5 |
| DX_ICD10 | O60.13X9 | preterm labor second tri with preterm delivery third tri, oth fetus |
| DX_ICD10 | O60.14 | preterm labor third tri with preterm delivery third tri |
| DX_ICD10 | O60.14X0 | preterm labor third tri with preterm delivery third tri, na or unspec |
| DX_ICD10 | O60.14X1 | preterm labor third tri with preterm delivery third tri, fetus 1 |
| DX_ICD10 | O60.14X2 | preterm labor third tri with preterm delivery third tri, fetus 2 |
| DX_ICD10 | O60.14X3 | preterm labor third tri with preterm delivery third tri, fetus 3 |
| DX_ICD10 | O60.14X4 | preterm labor third tri with preterm delivery third tri, fetus 4 |
| DX_ICD10 | O60.14X5 | preterm labor third tri with preterm delivery third tri, fetus 5 |
| DX_ICD10 | O60.14X9 | preterm labor third tri with preterm delivery third tri, oth fetus |
| DX_ICD10 | O60.2 | term delivery with preterm labor |
| DX_ICD10 | O60.20 | term delivery with preterm labor, unspec tri |
| DX_ICD10 | O60.20X0 | term delivery with preterm labor, unspec tri, na or unspec |
| DX_ICD10 | O60.20X1 | term delivery with preterm labor, unspec tri, fetus 1 |
| DX_ICD10 | O60.20X2 | term delivery with preterm labor, unspec tri, fetus 2 |
| DX_ICD10 | O60.20X3 | term delivery with preterm labor, unspec tri, fetus 3 |
| DX_ICD10 | O60.20X4 | term delivery with preterm labor, unspec tri, fetus 4 |
| DX_ICD10 | O60.20X5 | term delivery with preterm labor, unspec tri, fetus 5 |
| DX_ICD10 | O60.20X9 | term delivery with preterm labor, unspec tri, oth fetus |
| DX_ICD10 | O60.22 | term delivery with preterm labor, second tri |
| DX_ICD10 | O60.22X0 | term delivery with preterm labor, second tri, na or unspec |
| DX_ICD10 | O60.22X1 | term delivery with preterm labor, second tri, fetus 1 |
| DX_ICD10 | O60.22X2 | term delivery with preterm labor, second tri, fetus 2 |
| DX_ICD10 | O60.22X3 | term delivery with preterm labor, second tri, fetus 3 |
| DX_ICD10 | O60.22X4 | term delivery with preterm labor, second tri, fetus 4 |
| DX_ICD10 | O60.22X5 | term delivery with preterm labor, second tri, fetus 5 |
| DX_ICD10 | O60.22X9 | term delivery with preterm labor, second tri, oth fetus |
| DX_ICD10 | O60.23 | term delivery with preterm labor, third tri |
| DX_ICD10 | O60.23X0 | term delivery with preterm labor, third tri, na or unspec |
| DX_ICD10 | O60.23X1 | term delivery with preterm labor, third tri, fetus 1 |
| DX_ICD10 | O60.23X2 | term delivery with preterm labor, third tri, fetus 2 |
| DX_ICD10 | O60.23X3 | term delivery with preterm labor, third tri, fetus 3 |
| DX_ICD10 | O60.23X4 | term delivery with preterm labor, third tri, fetus 4 |
| DX_ICD10 | O60.23X5 | term delivery with preterm labor, third tri, fetus 5 |
| DX_ICD10 | O60.23X9 | term delivery with preterm labor, third tri, oth fetus |
| DX_ICD10 | O61 | failed induction of labor |
| DX_ICD10 | O61.0 | failed medical induction of labor |
| DX_ICD10 | O61.1 | failed instrumental induction of labor |
| DX_ICD10 | O61.8 | oth failed induction of labor |
| DX_ICD10 | O61.9 | failed induction of labor, unspec |
| DX_ICD10 | O62 | abnormalities of forces of labor |
| DX_ICD10 | O62.0 | primary inadequate contractions |
| DX_ICD10 | O62.1 | secondary uterine inertia |
| DX_ICD10 | O62.2 | oth uterine inertia |
| DX_ICD10 | O62.3 | precipitate labor |
| DX_ICD10 | O62.4 | hypertonic, incoordinate, & prolonged uterine contractions |
| DX_ICD10 | O62.8 | oth abnormalities of forces of labor |
| DX_ICD10 | O62.9 | abnormality of forces of labor, unspec |
| DX_ICD10 | O63 | long labor |
| DX_ICD10 | O63.0 | prolonged first stage (of labor) |
| DX_ICD10 | O63.1 | prolonged second stage (of labor) |
| DX_ICD10 | O63.2 | delayed delivery of second twin, triplet, etc. |
| DX_ICD10 | O63.9 | long labor, unspec |
| DX_ICD10 | O64 | obstructed labor due to malposition & malpresentation of fetus |
| DX_ICD10 | O64.0 | obstructed labor due to incomplete rotation of fetal head |
| DX_ICD10 | O64.0XX0 | obstructed labor due to incomplete rotation of fetal head, na or unspec |
| DX_ICD10 | O64.0XX1 | obstructed labor due to incomplete rotation of fetal head, fetus 1 |
| DX_ICD10 | O64.0XX2 | obstructed labor due to incomplete rotation of fetal head, fetus 2 |
| DX_ICD10 | O64.0XX3 | obstructed labor due to incomplete rotation of fetal head, fetus 3 |
| DX_ICD10 | O64.0XX4 | obstructed labor due to incomplete rotation of fetal head, fetus 4 |
| DX_ICD10 | O64.0XX5 | obstructed labor due to incomplete rotation of fetal head, fetus 5 |
| DX_ICD10 | O64.0XX9 | obstructed labor due to incomplete rotation of fetal head, oth fetus |
| DX_ICD10 | O64.1 | obstructed labor due to breech presentation |
| DX_ICD10 | O64.1XX0 | obstructed labor due to breech presentation, na or unspec |
| DX_ICD10 | O64.1XX1 | obstructed labor due to breech presentation, fetus 1 |
| DX_ICD10 | O64.1XX2 | obstructed labor due to breech presentation, fetus 2 |
| DX_ICD10 | O64.1XX3 | obstructed labor due to breech presentation, fetus 3 |
| DX_ICD10 | O64.1XX4 | obstructed labor due to breech presentation, fetus 4 |
| DX_ICD10 | O64.1XX5 | obstructed labor due to breech presentation, fetus 5 |
| DX_ICD10 | O64.1XX9 | obstructed labor due to breech presentation, oth fetus |
| DX_ICD10 | O64.2 | obstructed labor due to face presentation |
| DX_ICD10 | O64.2XX0 | obstructed labor due to face presentation, na or unspec |
| DX_ICD10 | O64.2XX1 | obstructed labor due to face presentation, fetus 1 |
| DX_ICD10 | O64.2XX2 | obstructed labor due to face presentation, fetus 2 |
| DX_ICD10 | O64.2XX3 | obstructed labor due to face presentation, fetus 3 |
| DX_ICD10 | O64.2XX4 | obstructed labor due to face presentation, fetus 4 |
| DX_ICD10 | O64.2XX5 | obstructed labor due to face presentation, fetus 5 |
| DX_ICD10 | O64.2XX9 | obstructed labor due to face presentation, oth fetus |
| DX_ICD10 | O64.3 | obstructed labor due to brow presentation |
| DX_ICD10 | O64.3XX0 | obstructed labor due to brow presentation, na or unspec |
| DX_ICD10 | O64.3XX1 | obstructed labor due to brow presentation, fetus 1 |
| DX_ICD10 | O64.3XX2 | obstructed labor due to brow presentation, fetus 2 |
| DX_ICD10 | O64.3XX3 | obstructed labor due to brow presentation, fetus 3 |
| DX_ICD10 | O64.3XX4 | obstructed labor due to brow presentation, fetus 4 |
| DX_ICD10 | O64.3XX5 | obstructed labor due to brow presentation, fetus 5 |
| DX_ICD10 | O64.3XX9 | obstructed labor due to brow presentation, oth fetus |
| DX_ICD10 | O64.4 | obstructed labor due to shoulder presentation |
| DX_ICD10 | O64.4XX0 | obstructed labor due to shoulder presentation, na or unspec |
| DX_ICD10 | O64.4XX1 | obstructed labor due to shoulder presentation, fetus 1 |
| DX_ICD10 | O64.4XX2 | obstructed labor due to shoulder presentation, fetus 2 |
| DX_ICD10 | O64.4XX3 | obstructed labor due to shoulder presentation, fetus 3 |
| DX_ICD10 | O64.4XX4 | obstructed labor due to shoulder presentation, fetus 4 |
| DX_ICD10 | O64.4XX5 | obstructed labor due to shoulder presentation, fetus 5 |
| DX_ICD10 | O64.4XX9 | obstructed labor due to shoulder presentation, oth fetus |
| DX_ICD10 | O64.5 | obstructed labor due to compound presentation |
| DX_ICD10 | O64.5XX0 | obstructed labor due to compound presentation, na or unspec |
| DX_ICD10 | O64.5XX1 | obstructed labor due to compound presentation, fetus 1 |
| DX_ICD10 | O64.5XX2 | obstructed labor due to compound presentation, fetus 2 |
| DX_ICD10 | O64.5XX3 | obstructed labor due to compound presentation, fetus 3 |
| DX_ICD10 | O64.5XX4 | obstructed labor due to compound presentation, fetus 4 |
| DX_ICD10 | O64.5XX5 | obstructed labor due to compound presentation, fetus 5 |
| DX_ICD10 | O64.5XX9 | obstructed labor due to compound presentation, oth fetus |
| DX_ICD10 | O64.8 | obstructed labor due to oth malposition & malpresentation |
| DX_ICD10 | O64.8XX0 | obstructed labor due to oth malpos & malpresentation, na or unspec |
| DX_ICD10 | O64.8XX1 | obstructed labor due to oth malposition & malpresentation, fetus 1 |
| DX_ICD10 | O64.8XX2 | obstructed labor due to oth malposition & malpresentation, fetus 2 |
| DX_ICD10 | O64.8XX3 | obstructed labor due to oth malposition & malpresentation, fetus 3 |
| DX_ICD10 | O64.8XX4 | obstructed labor due to oth malposition & malpresentation, fetus 4 |
| DX_ICD10 | O64.8XX5 | obstructed labor due to oth malposition & malpresentation, fetus 5 |
| DX_ICD10 | O64.8XX9 | obstructed labor due to oth malposition & malpresentation, oth fetus |
| DX_ICD10 | O64.9 | obstructed labor due to malposition & malpresentation, unspec |
| DX_ICD10 | O64.9XX0 | obstructed labor due to malpos & malpresent, unspec, na or unspec |
| DX_ICD10 | O64.9XX1 | obstructed labor due to malposition & malpresent, unspec, fetus 1 |
| DX_ICD10 | O64.9XX2 | obstructed labor due to malposition & malpresent, unspec, fetus 2 |
| DX_ICD10 | O64.9XX3 | obstructed labor due to malposition & malpresent, unspec, fetus 3 |
| DX_ICD10 | O64.9XX4 | obstructed labor due to malposition & malpresent, unspec, fetus 4 |
| DX_ICD10 | O64.9XX5 | obstructed labor due to malposition & malpresent, unspec, fetus 5 |
| DX_ICD10 | O64.9XX9 | obstructed labor due to malposition & malpresent, unspec, oth fetus |
| DX_ICD10 | O65 | obstructed labor due to maternal pelvic abnormality |
| DX_ICD10 | O65.0 | obstructed labor due to deformed pelvis |
| DX_ICD10 | O65.1 | obstructed labor due to generally contracted pelvis |
| DX_ICD10 | O65.2 | obstructed labor due to pelvic inlet contraction |
| DX_ICD10 | O65.3 | obstructed labor due to pelvic outlet & mid-cavity contraction |
| DX_ICD10 | O65.4 | obstructed labor due to fetopelvic disproportion, unspec |
| DX_ICD10 | O65.5 | obstructed labor due to abnormality of maternal pelvic organs |
| DX_ICD10 | O65.8 | obstructed labor due to oth maternal pelvic abnormalities |
| DX_ICD10 | O65.9 | obstructed labor due to maternal pelvic abnormality, unspec |
| DX_ICD10 | O66 | oth obstructed labor |
| DX_ICD10 | O66.0 | obstructed labor due to shoulder dystocia |
| DX_ICD10 | O66.1 | obstructed labor due to locked twins |
| DX_ICD10 | O66.2 | obstructed labor due to unusually large fetus |
| DX_ICD10 | O66.3 | obstructed labor due to oth abnormalities of fetus |
| DX_ICD10 | O66.4 | failed trial of labor |
| DX_ICD10 | O66.40 | failed trial of labor, unspec |
| DX_ICD10 | O66.41 | failed attempted vaginal birth after previous cesarean delivery |
| DX_ICD10 | O66.5 | attempted application of vacuum extractor & forceps |
| DX_ICD10 | O66.6 | obstructed labor due to oth multi fetuses |
| DX_ICD10 | O66.8 | oth specif obstructed labor |
| DX_ICD10 | O66.9 | obstructed labor, unspec |
| DX_ICD10 | O67 | lab & del complic by intrapartum hemor, not elsewhere classified |
| DX_ICD10 | O67.0 | intrapartum hemorrhage with coagulation defect |
| DX_ICD10 | O67.8 | oth intrapartum hemorrhage |
| DX_ICD10 | O67.9 | intrapartum hemorrhage, unspec |
| DX_ICD10 | O68 | l&d complic by abnormality of fetal acid-base balance |
| DX_ICD10 | O69 | l&d complic by umbilical cord complic |
| DX_ICD10 | O69.0 | l&d complic by prolapse of cord |
| DX_ICD10 | O69.0XX0 | l&d complic by prolapse of cord, na or unspec |
| DX_ICD10 | O69.0XX1 | l&d complic by prolapse of cord, fetus 1 |
| DX_ICD10 | O69.0XX2 | l&d complic by prolapse of cord, fetus 2 |
| DX_ICD10 | O69.0XX3 | l&d complic by prolapse of cord, fetus 3 |
| DX_ICD10 | O69.0XX4 | l&d complic by prolapse of cord, fetus 4 |
| DX_ICD10 | O69.0XX5 | l&d complic by prolapse of cord, fetus 5 |
| DX_ICD10 | O69.0XX9 | l&d complic by prolapse of cord, oth fetus |
| DX_ICD10 | O69.1 | l&d complic by cord around neck, with compression |
| DX_ICD10 | O69.1XX0 | l&d complic by cord around neck, w compression, na or unspec |
| DX_ICD10 | O69.1XX1 | l&d complic by cord around neck, w compression, fetus 1 |
| DX_ICD10 | O69.1XX2 | l&d complic by cord around neck, w compression, fetus 2 |
| DX_ICD10 | O69.1XX3 | l&d complic by cord around neck, w compression, fetus 3 |
| DX_ICD10 | O69.1XX4 | l&d complic by cord around neck, with compression, fetus 4 |
| DX_ICD10 | O69.1XX5 | l&d complic by cord around neck, w compression, fetus 5 |
| DX_ICD10 | O69.1XX9 | l&d complic by cord around neck, w compression, oth fetus |
| DX_ICD10 | O69.2 | l&d complic by oth cord entanglement, w compression |
| DX_ICD10 | O69.2XX0 | l&d complic by oth cord entanglement, w compression, na or unspec |
| DX_ICD10 | O69.2XX1 | l&d complic by oth cord entanglement, w compression, fetus 1 |
| DX_ICD10 | O69.2XX2 | l&d complic by oth cord entanglement, w compression, fetus 2 |
| DX_ICD10 | O69.2XX3 | l&d complic by oth cord entanglement, w compression, fetus 3 |
| DX_ICD10 | O69.2XX4 | l&d complic by oth cord entanglement, w compression, fetus 4 |
| DX_ICD10 | O69.2XX5 | l&d complic by oth cord entanglement, w compression, fetus 5 |
| DX_ICD10 | O69.2XX9 | l&d complic by oth cord entanglement, w compression, oth fetus |
| DX_ICD10 | O69.3 | l&d complic by short cord |
| DX_ICD10 | O69.3XX0 | l&d complic by short cord, na or unspec |
| DX_ICD10 | O69.3XX1 | l&d complic by short cord, fetus 1 |
| DX_ICD10 | O69.3XX2 | l&d complic by short cord, fetus 2 |
| DX_ICD10 | O69.3XX3 | l&d complic by short cord, fetus 3 |
| DX_ICD10 | O69.3XX4 | l&d complic by short cord, fetus 4 |
| DX_ICD10 | O69.3XX5 | l&d complic by short cord, fetus 5 |
| DX_ICD10 | O69.3XX9 | l&d complic by short cord, oth fetus |
| DX_ICD10 | O69.4 | l&d complic by vasa previa |
| DX_ICD10 | O69.4XX0 | l&d complic by vasa previa, na or unspec |
| DX_ICD10 | O69.4XX1 | l&d complic by vasa previa, fetus 1 |
| DX_ICD10 | O69.4XX2 | l&d complic by vasa previa, fetus 2 |
| DX_ICD10 | O69.4XX3 | l&d complic by vasa previa, fetus 3 |
| DX_ICD10 | O69.4XX4 | l&d complic by vasa previa, fetus 4 |
| DX_ICD10 | O69.4XX5 | l&d complic by vasa previa, fetus 5 |
| DX_ICD10 | O69.4XX9 | l&d complic by vasa previa, oth fetus |
| DX_ICD10 | O69.5 | l&d complic by vascular lesion of cord |
| DX_ICD10 | O69.5XX0 | l&d complic by vascular lesion of cord, na or unspec |
| DX_ICD10 | O69.5XX1 | l&d complic by vascular lesion of cord, fetus 1 |
| DX_ICD10 | O69.5XX2 | l&d complic by vascular lesion of cord, fetus 2 |
| DX_ICD10 | O69.5XX3 | l&d complic by vascular lesion of cord, fetus 3 |
| DX_ICD10 | O69.5XX4 | l&d complic by vascular lesion of cord, fetus 4 |
| DX_ICD10 | O69.5XX5 | l&d complic by vascular lesion of cord, fetus 5 |
| DX_ICD10 | O69.5XX9 | l&d complic by vascular lesion of cord, oth fetus |
| DX_ICD10 | O69.8 | l&d complic by oth cord complic |
| DX_ICD10 | O69.81 | l&d complic by cord around neck, wo compression |
| DX_ICD10 | O69.81X0 | l&d complic by cord around neck, wo compression, na or unspec |
| DX_ICD10 | O69.81X1 | l&d complic by cord around neck, wo compression, fetus 1 |
| DX_ICD10 | O69.81X2 | l&d complic by cord around neck, wo compression, fetus 2 |
| DX_ICD10 | O69.81X3 | l&d complic by cord around neck, wo compression, fetus 3 |
| DX_ICD10 | O69.81X4 | l&d complic by cord around neck, wo compression, fetus 4 |
| DX_ICD10 | O69.81X5 | l&d complic by cord around neck, wo compression, fetus 5 |
| DX_ICD10 | O69.81X9 | l&d complic by cord around neck, wo compression, oth fetus |
| DX_ICD10 | O69.82 | l&d complic by oth cord entanglement, wo compression |
| DX_ICD10 | O69.82X0 | l&d complic by oth cord entanglement, wo compression, na or unspec |
| DX_ICD10 | O69.82X1 | l&d complic by oth cord entanglement, wo compression, fetus 1 |
| DX_ICD10 | O69.82X2 | l&d complic by oth cord entanglement, wo compression, fetus 2 |
| DX_ICD10 | O69.82X3 | l&d complic by oth cord entanglement, wo compression, fetus 3 |
| DX_ICD10 | O69.82X4 | l&d complic by oth cord entanglement, wo compression, fetus 4 |
| DX_ICD10 | O69.82X5 | l&d complic by oth cord entanglement, wo compression, fetus 5 |
| DX_ICD10 | O69.82X9 | l&d complic by oth cord entanglement, wo compression, oth fetus |
| DX_ICD10 | O69.89 | l&d complic by oth cord complic |
| DX_ICD10 | O69.89X0 | l&d complic by oth cord complic, na or unspec |
| DX_ICD10 | O69.89X1 | l&d complic by oth cord complic, fetus 1 |
| DX_ICD10 | O69.89X2 | l&d complic by oth cord complic, fetus 2 |
| DX_ICD10 | O69.89X3 | l&d complic by oth cord complic, fetus 3 |
| DX_ICD10 | O69.89X4 | l&d complic by oth cord complic, fetus 4 |
| DX_ICD10 | O69.89X5 | l&d complic by oth cord complic, fetus 5 |
| DX_ICD10 | O69.89X9 | l&d complic by oth cord complic, oth fetus |
| DX_ICD10 | O69.9 | l&d complic by cord complication, unspec |
| DX_ICD10 | O69.9XX0 | l&d complic by cord complication, unspec, na or unspec |
| DX_ICD10 | O69.9XX1 | l&d complic by cord complication, unspec, fetus 1 |
| DX_ICD10 | O69.9XX2 | l&d complic by cord complication, unspec, fetus 2 |
| DX_ICD10 | O69.9XX3 | l&d complic by cord complication, unspec, fetus 3 |
| DX_ICD10 | O69.9XX4 | l&d complic by cord complication, unspec, fetus 4 |
| DX_ICD10 | O69.9XX5 | l&d complic by cord complication, unspec, fetus 5 |
| DX_ICD10 | O69.9XX9 | l&d complic by cord complication, unspec, oth fetus |
| DX_ICD10 | O70 | perineal laceration during delivery |
| DX_ICD10 | O70.0 | first degree perineal laceration during delivery |
| DX_ICD10 | O70.1 | second degree perineal laceration during delivery |
| DX_ICD10 | O70.2 | third degree perineal laceration during delivery |
| DX_ICD10 | O70.20 | third degree perineal laceration during delivery, unspec |
| DX_ICD10 | O70.21 | third degree perineal laceration during delivery, iiia |
| DX_ICD10 | O70.22 | third degree perineal laceration during delivery, iiib |
| DX_ICD10 | O70.23 | third degree perineal laceration during delivery, iiic |
| DX_ICD10 | O70.3 | fourth degree perineal laceration during delivery |
| DX_ICD10 | O70.4 | anal sphincter tear complic del, not assoc w third degree laceration |
| DX_ICD10 | O70.9 | perineal laceration during delivery, unspec |
| DX_ICD10 | O71 | oth obstetric trauma |
| DX_ICD10 | O71.0 | rupt of uterus (spontaneous) before onset of labor) |
| DX_ICD10 | O71.00 | rupt of uterus before onset of labor, unspec tri |
| DX_ICD10 | O71.02 | rupt of uterus before onset of labor, second tri |
| DX_ICD10 | O71.03 | rupt of uterus before onset of labor, third tri |
| DX_ICD10 | O71.1 | rupt of uterus during labor |
| DX_ICD10 | O71.2 | postpartum inversion of uterus |
| DX_ICD10 | O71.3 | obstetric laceration of cervix |
| DX_ICD10 | O71.4 | obstetric high vaginal laceration alone |
| DX_ICD10 | O71.5 | oth obstetric injury to pelvic organs |
| DX_ICD10 | O71.6 | obstetric dam to pelvic joints & ligaments |
| DX_ICD10 | O71.7 | obstetric hematoma of pelvis |
| DX_ICD10 | O71.8 | oth specif obstetric trauma |
| DX_ICD10 | O71.81 | laceration of uterus, not elsewhere classified |
| DX_ICD10 | O71.82 | oth specif trauma to perineum & vulva |
| DX_ICD10 | O71.89 | oth specif obstetric trauma |
| DX_ICD10 | O71.9 | obstetric trauma, unspec |
| DX_ICD10 | O72 | postpartum hemorrhage |
| DX_ICD10 | O72.0 | third-stage hemorrhage |
| DX_ICD10 | O72.1 | oth immediate postpartum hemorrhage |
| DX_ICD10 | O72.2 | delayed & secondary postpartum hemorrhage |
| DX_ICD10 | O72.3 | postpartum coagulation defects |
| DX_ICD10 | O73 | retained plcnta & membranes, wo hemorrhage |
| DX_ICD10 | O73.0 | retained plcnta wo hemorrhage |
| DX_ICD10 | O73.1 | retained portions of plcnta & membranes, wo hemorrhage |
| DX_ICD10 | O74 | complic of anesthesia during labor & delivery |
| DX_ICD10 | O74.0 | aspiration pneumonitis due to anesthesia during labor & delivery |
| DX_ICD10 | O74.1 | oth pulmonary complic of anesthesia during labor & delivery |
| DX_ICD10 | O74.2 | cardiac complic of anesthesia during labor & delivery |
| DX_ICD10 | O74.3 | cns complic of anesthesia during labor & delivery |
| DX_ICD10 | O74.4 | toxic reaction to local anesthesia during labor & delivery |
| DX_ICD10 | O74.5 | spinal & epidural anesthesia-induced headache during labor & delivery |
| DX_ICD10 | O74.6 | oth complic of spinal & epidural anesthesia during labor & delivery |
| DX_ICD10 | O74.7 | failed or difficult intubation for anesthesia during labor & delivery |
| DX_ICD10 | O74.8 | oth complic of anesthesia during labor & delivery |
| DX_ICD10 | O74.9 | complication of anesthesia during labor & delivery, unspec |
| DX_ICD10 | O75 | oth complic of labor & delivery, not elsewhere classified |
| DX_ICD10 | O75.0 | maternal distress during labor & delivery |
| DX_ICD10 | O75.1 | shock during or following labor & delivery |
| DX_ICD10 | O75.2 | pyrexia during labor, not elsewhere classified |
| DX_ICD10 | O75.3 | oth infection during labor |
| DX_ICD10 | O75.4 | oth complic of obstetric surgery & procedures |
| DX_ICD10 | O75.5 | delayed delivery after artificial rupt of membranes |
| DX_ICD10 | O75.8 | oth specif complic of labor & delivery |
| DX_ICD10 | O75.81 | maternal exhaustion complic labor & delivery |
| DX_ICD10 | O75.82 | onset lab after 37 wks but before 39 wks, w del by (planned) c-section |
| DX_ICD10 | O75.89 | oth specif complic of labor & delivery |
| DX_ICD10 | O75.9 | complication of labor & delivery, unspec |
| DX_ICD10 | O76 | abnormality in fetal heart rate & rhythm complic labor & delivery |
| DX_ICD10 | O77 | oth fetal stress complic labor & delivery |
| DX_ICD10 | O77.0 | l&d complic by meconium in amniotic fluid |
| DX_ICD10 | O77.1 | fetal stress in labor or delivery due to drug administration |
| DX_ICD10 | O77.8 | l&d complic by oth evidence of fetal stress |
| DX_ICD10 | O77.9 | l&d complic by fetal stress, unspec |
| DX_ICD10 | O80 | encounter for full-term uncomplic delivery |
| DX_ICD10 | O82 | encounter for cesarean delivery wo indication |
| DX_ICD10 | O85 | puerperal sepsis |
| DX_ICD10 | O86 | oth puerperal infections |
| DX_ICD10 | O86.0 | infection of obstetric surgical wound |
| DX_ICD10 | O86.00 | infection of obstetric surgical wound, unspec |
| DX_ICD10 | O86.01 | infection of obstetric surgical wound, superficial incisional site |
| DX_ICD10 | O86.02 | infection of obstetric surgical wound, deep incisional site |
| DX_ICD10 | O86.03 | infection of obstetric surgical wound, organ & space site |
| DX_ICD10 | O86.04 | sepsis following an obstetrical procedure |
| DX_ICD10 | O86.09 | infection of obstetric surgical wound, oth surgical site |
| DX_ICD10 | O86.1 | oth infection of genital tract following delivery |
| DX_ICD10 | O86.11 | cervicitis following delivery |
| DX_ICD10 | O86.12 | endometritis following delivery |
| DX_ICD10 | O86.13 | vaginitis following delivery |
| DX_ICD10 | O86.19 | oth infection of genital tract following delivery |
| DX_ICD10 | O86.2 | urinary tract infection following delivery |
| DX_ICD10 | O86.20 | urinary tract infection following delivery, unspec |
| DX_ICD10 | O86.21 | infection of kidney following delivery |
| DX_ICD10 | O86.22 | infection of bladder following delivery |
| DX_ICD10 | O86.29 | oth urinary tract infection following delivery |
| DX_ICD10 | O86.4 | pyrexia of unknown origin following delivery |
| DX_ICD10 | O86.8 | oth specif puerperal infections |
| DX_ICD10 | O86.81 | puerperal septic thrombophlebitis |
| DX_ICD10 | O86.89 | oth specif puerperal infections |
| DX_ICD10 | O87 | venous complic in the puerperium |
| DX_ICD10 | O87.0 | superficial thrombophlebitis in the puerperium |
| DX_ICD10 | O87.1 | deep phlebothrombosis in the puerperium |
| DX_ICD10 | O87.2 | hemorrhoids in the puerperium |
| DX_ICD10 | O87.3 | cerebral venous thrombosis in the puerperium |
| DX_ICD10 | O87.4 | varicose veins of lower extremity in the puerperium |
| DX_ICD10 | O87.8 | oth venous complic in the puerperium |
| DX_ICD10 | O87.9 | venous complication in the puerperium, unspec |
| DX_ICD10 | O88 | obstetric embolism |
| DX_ICD10 | O88.0 | obstetric air embolism |
| DX_ICD10 | O88.01 | obstetric air embolism in preg |
| DX_ICD10 | O88.011 | air embolism in preg, first tri |
| DX_ICD10 | O88.012 | air embolism in preg, second tri |
| DX_ICD10 | O88.013 | air embolism in preg, third tri |
| DX_ICD10 | O88.019 | air embolism in preg, unspec tri |
| DX_ICD10 | O88.02 | air embolism in childbirth |
| DX_ICD10 | O88.03 | air embolism in the puerperium |
| DX_ICD10 | O88.1 | amniotic fluid embolism |
| DX_ICD10 | O88.11 | amniotic fluid embolism in preg |
| DX_ICD10 | O88.111 | amniotic fluid embolism in preg, first tri |
| DX_ICD10 | O88.112 | amniotic fluid embolism in preg, second tri |
| DX_ICD10 | O88.113 | amniotic fluid embolism in preg, third tri |
| DX_ICD10 | O88.119 | amniotic fluid embolism in preg, unspec tri |
| DX_ICD10 | O88.12 | amniotic fluid embolism in childbirth |
| DX_ICD10 | O88.13 | amniotic fluid embolism in the puerperium |
| DX_ICD10 | O88.2 | obstetric thromboembolism |
| DX_ICD10 | O88.21 | thromboembolism in preg |
| DX_ICD10 | O88.211 | thromboembolism in preg, first tri |
| DX_ICD10 | O88.212 | thromboembolism in preg, second tri |
| DX_ICD10 | O88.213 | thromboembolism in preg, third tri |
| DX_ICD10 | O88.219 | thromboembolism in preg, unspec tri |
| DX_ICD10 | O88.22 | thromboembolism in childbirth |
| DX_ICD10 | O88.23 | thromboembolism in the puerperium |
| DX_ICD10 | O88.3 | obstetric pyemic & septic embolism |
| DX_ICD10 | O88.31 | pyemic & septic embolism in preg |
| DX_ICD10 | O88.311 | pyemic & septic embolism in preg, first tri |
| DX_ICD10 | O88.312 | pyemic & septic embolism in preg, second tri |
| DX_ICD10 | O88.313 | pyemic & septic embolism in preg, third tri |
| DX_ICD10 | O88.319 | pyemic & septic embolism in preg, unspec tri |
| DX_ICD10 | O88.32 | pyemic & septic embolism in childbirth |
| DX_ICD10 | O88.33 | pyemic & septic embolism in the puerperium |
| DX_ICD10 | O88.8 | oth obstetric embolism |
| DX_ICD10 | O88.81 | oth embolism in preg |
| DX_ICD10 | O88.811 | oth embolism in preg, first tri |
| DX_ICD10 | O88.812 | oth embolism in preg, second tri |
| DX_ICD10 | O88.813 | oth embolism in preg, third tri |
| DX_ICD10 | O88.819 | oth embolism in preg, unspec tri |
| DX_ICD10 | O88.82 | oth embolism in childbirth |
| DX_ICD10 | O88.83 | oth embolism in the puerperium |
| DX_ICD10 | O89 | complic of anesthesia during the puerperium |
| DX_ICD10 | O89.0 | pulmonary complic of anesthesia during the puerperium |
| DX_ICD10 | O89.01 | aspiration pneumonitis due to anesthesia during the puerperium |
| DX_ICD10 | O89.09 | oth pulmonary complic of anesthesia during the puerperium |
| DX_ICD10 | O89.1 | cardiac complic of anesthesia during the puerperium |
| DX_ICD10 | O89.2 | cns complic of anesthesia during the puerperium |
| DX_ICD10 | O89.3 | toxic reaction to local anesthesia during the puerperium |
| DX_ICD10 | O89.4 | spinal & epidural anesthesia-induced headache during the puerperium |
| DX_ICD10 | O89.5 | oth complic of spinal & epidural anesthesia during the puerperium |
| DX_ICD10 | O89.6 | failed or difficult intubation for anesthesia during the puerperium |
| DX_ICD10 | O89.8 | oth complic of anesthesia during the puerperium |
| DX_ICD10 | O89.9 | complication of anesthesia during the puerperium, unspec |
| DX_ICD10 | O90 | complic of the puerperium, not elsewhere classified |
| DX_ICD10 | O90.0 | disruption of cesarean delivery wound |
| DX_ICD10 | O90.1 | disruption of perineal obstetric wound |
| DX_ICD10 | O90.2 | hematoma of obstetric wound |
| DX_ICD10 | O90.3 | peripartum cardiomyopathy |
| DX_ICD10 | O90.4 | postpartum acute kidney failure |
| DX_ICD10 | O90.5 | postpartum thyroiditis |
| DX_ICD10 | O90.6 | postpartum mood disturbance |
| DX_ICD10 | O90.7 | complic of the puerperium that are not classified elsewhere |
| DX_ICD10 | O90.8 | oth complic of the puerperium, not elsewhere classified |
| DX_ICD10 | O90.81 | anemia of the puerperium |
| DX_ICD10 | O90.89 | oth complic of the puerperium, not elsewhere classified |
| DX_ICD10 | O90.9 | complication of the puerperium, unspec |
| DX_ICD10 | O91 | infections of breast associated with preg, the puerperium & lactation |
| DX_ICD10 | O91.0 | infection of nipple associated with preg, the puerperium & lactation |
| DX_ICD10 | O91.01 | infection of nipple associated with preg |
| DX_ICD10 | O91.011 | infection of nipple associated with preg, first tri |
| DX_ICD10 | O91.012 | infection of nipple associated with preg, second tri |
| DX_ICD10 | O91.013 | infection of nipple associated with preg, third tri |
| DX_ICD10 | O91.019 | infection of nipple associated with preg, unspec tri |
| DX_ICD10 | O91.02 | infection of nipple associated with the puerperium |
| DX_ICD10 | O91.03 | infection of nipple associated with lactation |
| DX_ICD10 | O91.1 | abscess of breast associated with preg, the puerperium & lactation |
| DX_ICD10 | O91.11 | abscess of breast associated with preg |
| DX_ICD10 | O91.111 | abscess of breast associated with preg, first tri |
| DX_ICD10 | O91.112 | abscess of breast associated with preg, second tri |
| DX_ICD10 | O91.113 | abscess of breast associated with preg, third tri |
| DX_ICD10 | O91.119 | abscess of breast associated with preg, unspec tri |
| DX_ICD10 | O91.12 | abscess of breast associated with the puerperium |
| DX_ICD10 | O91.13 | abscess of breast associated with lactation |
| DX_ICD10 | O91.2 | nonpurulent mastitis associated with preg, the puerperium & lactation |
| DX_ICD10 | O91.21 | nonpurulent mastitis associated with preg |
| DX_ICD10 | O91.211 | nonpurulent mastitis associated with preg, first tri |
| DX_ICD10 | O91.212 | nonpurulent mastitis associated with preg, second tri |
| DX_ICD10 | O91.213 | nonpurulent mastitis associated with preg, third tri |
| DX_ICD10 | O91.219 | nonpurulent mastitis associated with preg, unspec tri |
| DX_ICD10 | O91.22 | nonpurulent mastitis associated with the puerperium |
| DX_ICD10 | O91.23 | nonpurulent mastitis associated with lactation |
| DX_ICD10 | O92 | oth dis breast & dis lactation assoc w preg & puerperium |
| DX_ICD10 | O92.0 | retracted nipple associated with preg, the puerperium, & lactation |
| DX_ICD10 | O92.01 | retracted nipple associated with preg |
| DX_ICD10 | O92.011 | retracted nipple associated with preg, first tri |
| DX_ICD10 | O92.012 | retracted nipple associated with preg, second tri |
| DX_ICD10 | O92.013 | retracted nipple associated with preg, third tri |
| DX_ICD10 | O92.019 | retracted nipple associated with preg, unspec tri |
| DX_ICD10 | O92.02 | retracted nipple associated with the puerperium |
| DX_ICD10 | O92.03 | retracted nipple associated with lactation |
| DX_ICD10 | O92.1 | cracked nipple associated with preg, the puerperium, & lactation |
| DX_ICD10 | O92.11 | cracked nipple associated with preg |
| DX_ICD10 | O92.111 | cracked nipple associated with preg, first tri |
| DX_ICD10 | O92.112 | cracked nipple associated with preg, second tri |
| DX_ICD10 | O92.113 | cracked nipple associated with preg, third tri |
| DX_ICD10 | O92.119 | cracked nipple associated with preg, unspec tri |
| DX_ICD10 | O92.12 | cracked nipple associated with the puerperium |
| DX_ICD10 | O92.13 | cracked nipple associated with lactation |
| DX_ICD10 | O92.2 | oth & unspec dis breast assoc w preg & the puerperium |
| DX_ICD10 | O92.20 | unspec disorder of breast associated with preg & the puerperium |
| DX_ICD10 | O92.29 | oth disorders of breast associated with preg & the puerperium |
| DX_ICD10 | O92.5 | suppressed lactation |
| DX_ICD10 | O92.6 | galactorrhea |
| DX_ICD10 | O94 | sequelae of complication of preg, childbirth, & the puerperium |
| DX_ICD10 | O98 | mat infect & parasit diss but complic preg, brth & puerperium |
| DX_ICD10 | O98.0 | tuberculosis complic preg, childbirth & the puerperium |
| DX_ICD10 | O98.01 | tuberculosis complic preg |
| DX_ICD10 | O98.011 | tuberculosis complic preg, first tri |
| DX_ICD10 | O98.012 | tuberculosis complic preg, second tri |
| DX_ICD10 | O98.013 | tuberculosis complic preg, third tri |
| DX_ICD10 | O98.019 | tuberculosis complic preg, unspec tri |
| DX_ICD10 | O98.02 | tuberculosis complic childbirth |
| DX_ICD10 | O98.03 | tuberculosis complic the puerperium |
| DX_ICD10 | O98.1 | syphilis complic preg, childbirth & the puerperium |
| DX_ICD10 | O98.11 | syphilis complic preg |
| DX_ICD10 | O98.111 | syphilis complic preg, first tri |
| DX_ICD10 | O98.112 | syphilis complic preg, second tri |
| DX_ICD10 | O98.113 | syphilis complic preg, third tri |
| DX_ICD10 | O98.119 | syphilis complic preg, unspec tri |
| DX_ICD10 | O98.12 | syphilis complic childbirth |
| DX_ICD10 | O98.13 | syphilis complic the puerperium |
| DX_ICD10 | O98.2 | gonorrhea complic preg, childbirth & the puerperium |
| DX_ICD10 | O98.21 | gonorrhea complic preg |
| DX_ICD10 | O98.211 | gonorrhea complic preg, first tri |
| DX_ICD10 | O98.212 | gonorrhea complic preg, second tri |
| DX_ICD10 | O98.213 | gonorrhea complic preg, third tri |
| DX_ICD10 | O98.219 | gonorrhea complic preg, unspec tri |
| DX_ICD10 | O98.22 | gonorrhea complic childbirth |
| DX_ICD10 | O98.23 | gonorrhea complic the puerperium |
| DX_ICD10 | O98.3 | oth infect predom sex transm complic preg, chldbrth & puerperium |
| DX_ICD10 | O98.31 | oth infect predom sex transm complic preg |
| DX_ICD10 | O98.311 | oth infect predom sex transm complic preg, first tri |
| DX_ICD10 | O98.312 | oth infect predom sex transm complic preg, second tri |
| DX_ICD10 | O98.313 | oth infect predom sex transm complic preg, third tri |
| DX_ICD10 | O98.319 | oth infect predom sex transm complic preg, unspec tri |
| DX_ICD10 | O98.32 | oth infect predom sex transm complic childbirth |
| DX_ICD10 | O98.33 | oth infections with a predom sex transmission complica puerperium |
| DX_ICD10 | O98.4 | viral hepatitis complic preg, childbirth & the puerperium |
| DX_ICD10 | O98.41 | viral hepatitis complic preg |
| DX_ICD10 | O98.411 | viral hepatitis complic preg, first tri |
| DX_ICD10 | O98.412 | viral hepatitis complic preg, second tri |
| DX_ICD10 | O98.413 | viral hepatitis complic preg, third tri |
| DX_ICD10 | O98.419 | viral hepatitis complic preg, unspec tri |
| DX_ICD10 | O98.42 | viral hepatitis complic childbirth |
| DX_ICD10 | O98.43 | viral hepatitis complic the puerperium |
| DX_ICD10 | O98.5 | oth viral dis complic preg, childbirth & the puerperium |
| DX_ICD10 | O98.51 | oth viral dis complic preg |
| DX_ICD10 | O98.511 | oth viral diss complic preg, first tri |
| DX_ICD10 | O98.512 | oth viral diseases complic preg, second tri |
| DX_ICD10 | O98.513 | oth viral diseases complic preg, third tri |
| DX_ICD10 | O98.519 | oth viral diseases complic preg, unspec tri |
| DX_ICD10 | O98.52 | oth viral diseases complic childbirth |
| DX_ICD10 | O98.53 | oth viral diseases complic the puerperium |
| DX_ICD10 | O98.6 | protozoal diseases complic preg, childbirth & the puerperium |
| DX_ICD10 | O98.61 | protozoal diseases complic preg |
| DX_ICD10 | O98.611 | protozoal diseases complic preg, first tri |
| DX_ICD10 | O98.612 | protozoal diseases complic preg, second tri |
| DX_ICD10 | O98.613 | protozoal diseases complic preg, third tri |
| DX_ICD10 | O98.619 | protozoal diseases complic preg, unspec tri |
| DX_ICD10 | O98.62 | protozoal diseases complic childbirth |
| DX_ICD10 | O98.63 | protozoal diseases complic the puerperium |
| DX_ICD10 | O98.7 | hiv disease complic preg, childbirth & the puerperium |
| DX_ICD10 | O98.71 | hiv disease complic preg |
| DX_ICD10 | O98.711 | hiv disease complic preg, first tri |
| DX_ICD10 | O98.712 | hiv disease complic preg, second tri |
| DX_ICD10 | O98.713 | hiv disease complic preg, third tri |
| DX_ICD10 | O98.719 | hiv disease complic preg, unspec tri |
| DX_ICD10 | O98.72 | hiv disease complic childbirth |
| DX_ICD10 | O98.73 | hiv disease complic the puerperium |
| DX_ICD10 | O98.8 | oth mat infect & parasit dis complic preg, childbirth & the puerperium |
| DX_ICD10 | O98.81 | oth mat infect & parasit diseases complic preg |
| DX_ICD10 | O98.811 | oth mat infect & parasit diseases complic preg, first tri |
| DX_ICD10 | O98.812 | oth mat infect & parasit diseases complic preg, second tri |
| DX_ICD10 | O98.813 | oth mat infect & parasit diseases complic preg, third tri |
| DX_ICD10 | O98.819 | oth mat infect & parasit diseases complic preg, unspec tri |
| DX_ICD10 | O98.82 | oth mat infect & parasit diseases complic childbirth |
| DX_ICD10 | O98.83 | oth mat infect & parasit diseases complic the puerperium |
| DX_ICD10 | O98.9 | unspec mat infect & parasit dis complic preg, chldbrth & puerperium |
| DX_ICD10 | O98.91 | unspec mat infect & parasit disease complic preg |
| DX_ICD10 | O98.911 | unspec mat infect & parasit disease complic preg, first tri |
| DX_ICD10 | O98.912 | unspec mat infect & parasit disease complic preg, second tri |
| DX_ICD10 | O98.913 | unspec mat infect & parasit disease complic preg, third tri |
| DX_ICD10 | O98.919 | unspec mat infect & parasit disease complic preg, unspec tri |
| DX_ICD10 | O98.92 | unspec mat infect & parasit disease complic childbirth |
| DX_ICD10 | O98.93 | unspec mat infect & parasit disease complic the puerperium |
| DX_ICD10 | O99 | oth mat dis but complic preg, childbirth & the puerperium |
| DX_ICD10 | O99.0 | anemia complic preg, childbirth & the puerperium |
| DX_ICD10 | O99.01 | anemia complic preg |
| DX_ICD10 | O99.011 | anemia complic preg, first tri |
| DX_ICD10 | O99.012 | anemia complic preg, second tri |
| DX_ICD10 | O99.013 | anemia complic preg, third tri |
| DX_ICD10 | O99.019 | anemia complic preg, unspec tri |
| DX_ICD10 | O99.02 | anemia complic childbirth |
| DX_ICD10 | O99.03 | anemia complic the puerperium |
| DX_ICD10 | O99.1 | oth dis blood/immune compl preg/childbrth |
| DX_ICD10 | O99.11 | oth dis blood & bloodforming org & imm mech compl preg |
| DX_ICD10 | O99.111 | oth dis blood & blood-form orgs & dis imm mech compl preg, first tri |
| DX_ICD10 | O99.112 | oth dis blood & blood-form orgs & dis imm mech compl preg, sec tri |
| DX_ICD10 | O99.113 | oth dis blood & blood-form orgs & dis imm mech compl preg, third tri |
| DX_ICD10 | O99.119 | oth dis bld & blood-form orgs & dis imm mech compl preg, unspec tri |
| DX_ICD10 | O99.12 | oth dis blood & blood-form orgs & dis imm mech complic chldbrth |
| DX_ICD10 | O99.13 | oth dis blood & blood-form orgs & dis imm mech complic puerperium |
| DX_ICD10 | O99.2 | endocrine, nutrit & metabol dis complic preg, chldbrth & puerperium |
| DX_ICD10 | O99.21 | obesity complic preg, childbirth, & the puerperium |
| DX_ICD10 | O99.210 | obesity complic preg, unspec tri |
| DX_ICD10 | O99.211 | obesity complic preg, first tri |
| DX_ICD10 | O99.212 | obesity complic preg, second tri |
| DX_ICD10 | O99.213 | obesity complic preg, third tri |
| DX_ICD10 | O99.214 | obesity complic childbirth |
| DX_ICD10 | O99.215 | obesity complic the puerperium |
| DX_ICD10 | O99.28 | oth endo, nutrit & metab dis complic preg, childbirth & the puerperium |
| DX_ICD10 | O99.280 | endo, nutrit & metabolic dis complic preg, unspec tri |
| DX_ICD10 | O99.281 | endo, nutrit & metabolic dis complic preg, first tri |
| DX_ICD10 | O99.282 | endo, nutrit & metabolic dis complic preg, second tri |
| DX_ICD10 | O99.283 | endo, nutrit & metabolic dis complic preg, third tri |
| DX_ICD10 | O99.284 | endocrine, nutritional & metabolic diseases complic childbirth |
| DX_ICD10 | O99.285 | endocrine, nutritional & metabolic diseases complic puerperium |
| DX_ICD10 | O99.3 | mental dis & nervous sys complicg preg, childbirth & the puerperium |
| DX_ICD10 | O99.31 | alcohol use complic preg,childbirth, & the puerperium |
| DX_ICD10 | O99.310 | alcohol use complic preg, unspec tri |
| DX_ICD10 | O99.311 | alcohol use complic preg, first tri |
| DX_ICD10 | O99.312 | alcohol use complic preg, second tri |
| DX_ICD10 | O99.313 | alcohol use complic preg, third tri |
| DX_ICD10 | O99.314 | alcohol use complic childbirth |
| DX_ICD10 | O99.315 | alcohol use complic the puerperium |
| DX_ICD10 | O99.32 | drug use complic preg, childbirth, & the puerperium |
| DX_ICD10 | O99.320 | drug use complic preg, unspec tri |
| DX_ICD10 | O99.321 | drug use complic preg, first tri |
| DX_ICD10 | O99.322 | drug use complic preg, second tri |
| DX_ICD10 | O99.323 | drug use complic preg, third tri |
| DX_ICD10 | O99.324 | drug use complic childbirth |
| DX_ICD10 | O99.325 | drug use complic the puerperium |
| DX_ICD10 | O99.33 | smoking (tobacco) complic preg, childbirth, & the puerperium |
| DX_ICD10 | O99.330 | smoking (tobacco) complic preg, unspec tri |
| DX_ICD10 | O99.331 | smoking (tobacco) complic preg, first tri |
| DX_ICD10 | O99.332 | smoking (tobacco) complic preg, second tri |
| DX_ICD10 | O99.333 | smoking (tobacco) complic preg, third tri |
| DX_ICD10 | O99.334 | smoking (tobacco) complic childbirth |
| DX_ICD10 | O99.335 | smoking (tobacco) complic the puerperium |
| DX_ICD10 | O99.34 | oth mental disorders complic preg, childbirth, & puerperium |
| DX_ICD10 | O99.340 | oth mental disorders complic preg, unspec tri |
| DX_ICD10 | O99.341 | oth mental disorders complic preg, first tri |
| DX_ICD10 | O99.342 | oth mental disorders complic preg, second tri |
| DX_ICD10 | O99.343 | oth mental disorders complic preg, third tri |
| DX_ICD10 | O99.344 | oth mental disorders complic childbirth |
| DX_ICD10 | O99.345 | oth mental disorders complic the puerperium |
| DX_ICD10 | O99.35 | dis nervous system complic preg, childbirth, & puerperium |
| DX_ICD10 | O99.350 | diseases of the nervous system complic preg, unspec tri |
| DX_ICD10 | O99.351 | diseases of the nervous system complic preg, first tri |
| DX_ICD10 | O99.352 | diseases of the nervous system complic preg, second tri |
| DX_ICD10 | O99.353 | diseases of the nervous system complic preg, third tri |
| DX_ICD10 | O99.354 | diseases of the nervous system complic childbirth |
| DX_ICD10 | O99.355 | diseases of the nervous system complic the puerperium |
| DX_ICD10 | O99.4 | dis circulatory system complic preg, childbirth & the puerperium |
| DX_ICD10 | O99.41 | diseases of the circulatory system complic preg |
| DX_ICD10 | O99.411 | diseases of the circulatory system complic preg, first tri |
| DX_ICD10 | O99.412 | diseases of the circulatory system complic preg, second tri |
| DX_ICD10 | O99.413 | diseases of the circulatory system complic preg, third tri |
| DX_ICD10 | O99.419 | diseases of the circulatory system complic preg, unspec tri |
| DX_ICD10 | O99.42 | diseases of the circulatory system complic childbirth |
| DX_ICD10 | O99.43 | diseases of the circulatory system complic the puerperium |
| DX_ICD10 | O99.5 | diseases respiratory sys complic preg, childbirth & puerperium |
| DX_ICD10 | O99.51 | diseases of the respiratory sys complic preg |
| DX_ICD10 | O99.511 | diseases of the respiratory system complic preg, first tri |
| DX_ICD10 | O99.512 | diseases of the respiratory sys complic preg, second tri |
| DX_ICD10 | O99.513 | diseases of the respiratory sys complic preg, third tri |
| DX_ICD10 | O99.519 | diseases of the respiratory sys complic preg, unspec tri |
| DX_ICD10 | O99.52 | diseases of the respiratory sys complic childbirth |
| DX_ICD10 | O99.53 | diseases of the respiratory sys complic the puerperium |
| DX_ICD10 | O99.6 | diseases of the digestive sys complic preg, childbirth & the puerperium |
| DX_ICD10 | O99.61 | diseases of the digestive sys complic preg |
| DX_ICD10 | O99.611 | diseases of the digestive sys complic preg, first tri |
| DX_ICD10 | O99.612 | diseases of the digestive sys complic preg, second tri |
| DX_ICD10 | O99.613 | diseases of the digestive sys complic preg, third tri |
| DX_ICD10 | O99.619 | diseases of the digestive sys complic preg, unspec tri |
| DX_ICD10 | O99.62 | diseases of the digestive sys complic childbirth |
| DX_ICD10 | O99.63 | diseases of the digestive sys complic the puerperium |
| DX_ICD10 | O99.7 | dis skin & subcutan tissue complic preg, childbirth & the puerperium |
| DX_ICD10 | O99.71 | diseases of the skin & subcutaneous tissue complic preg |
| DX_ICD10 | O99.711 | diseases of the skin & subcutaneous tissue complic preg, first tri |
| DX_ICD10 | O99.712 | diseases of the skin & subcutaneous tissue complic preg, second tri |
| DX_ICD10 | O99.713 | diseases of the skin & subcutaneous tissue complic preg, third tri |
| DX_ICD10 | O99.719 | diseases of the skin & subcutaneous tissue complic preg, unspec tri |
| DX_ICD10 | O99.72 | diseases of the skin & subcutaneous tissue complic childbirth |
| DX_ICD10 | O99.73 | diseases of the skin & subcutaneous tissue complic the puerperium |
| DX_ICD10 | O99.8 | oth specif dis & conds complic preg, childbirth & the puerperium |
| DX_ICD10 | O99.81 | abnormal glucose complic preg, childbirth & the puerperium |
| DX_ICD10 | O99.810 | abnormal glucose complic preg |
| DX_ICD10 | O99.814 | abnormal glucose complic childbirth |
| DX_ICD10 | O99.815 | abnormal glucose complic the puerperium |
| DX_ICD10 | O99.82 | strept b carrier state complic preg, childbirth & the puerperium) |
| DX_ICD10 | O99.820 | streptococcus b carrier state complic preg |
| DX_ICD10 | O99.824 | streptococcus b carrier state complic childbirth |
| DX_ICD10 | O99.825 | streptococcus b carrier state complic the puerperium |
| DX_ICD10 | O99.83 | oth infection carrier state complic preg, childbirth & the puerperium |
| DX_ICD10 | O99.830 | oth infection carrier state complic preg |
| DX_ICD10 | O99.834 | oth infection carrier state complic childbirth |
| DX_ICD10 | O99.835 | oth infection carrier state complic the puerperium |
| DX_ICD10 | O99.84 | bariatric surgery status complic preg, childbirth & the puerperium |
| DX_ICD10 | O99.840 | bariatric surgery status complic preg, unspec tri |
| DX_ICD10 | O99.841 | bariatric surgery status complic preg, first tri |
| DX_ICD10 | O99.842 | bariatric surgery status complic preg, second tri |
| DX_ICD10 | O99.843 | bariatric surgery status complic preg, third tri |
| DX_ICD10 | O99.844 | bariatric surgery status complic childbirth |
| DX_ICD10 | O99.845 | bariatric surgery status complic the puerperium |
| DX_ICD10 | O99.89 | oth specif dis & conditions complic preg, childbirth & the puerperium |
| DX_ICD10 | O99.891 | oth specif diseases & conditions complic preg |
| DX_ICD10 | O99.892 | oth specif diseases & conditions complic childbirth |
| DX_ICD10 | O99.893 | oth specif diseases & conditions complic puerperium |
| DX_ICD10 | O9A | mat malig neo, traum injur complic preg, chldbrth & puerperium |
| DX_ICD10 | O9A.1 | malignant neoplasm complic preg, childbirth & the puerperium |
| DX_ICD10 | O9A.11 | malignant neoplasm complic preg |
| DX_ICD10 | O9A.111 | malignant neoplasm complic preg, first tri |
| DX_ICD10 | O9A.112 | malignant neoplasm complic preg, second tri |
| DX_ICD10 | O9A.113 | malignant neoplasm complic preg, third tri |
| DX_ICD10 | O9A.119 | malignant neoplasm complic preg, unspec tri |
| DX_ICD10 | O9A.12 | malignant neoplasm complic childbirth |
| DX_ICD10 | O9A.13 | malignant neoplasm complic the puerperium |
| DX_ICD10 | O9A.2 | inj, poison & oth conseq complic preg, birth & puerperium |
| DX_ICD10 | O9A.21 | inj, poison & oth conseq external causes complic preg |
| DX_ICD10 | O9A.211 | inj, poison & oth conseq external causes complic preg, first tri |
| DX_ICD10 | O9A.212 | inj, poison & oth conseq external causes complic, second tri |
| DX_ICD10 | O9A.213 | inj, poison & oth conseq external causes complic, third tri |
| DX_ICD10 | O9A.219 | inj, poison & oth conseq external causes complic preg, unspec tri |
| DX_ICD10 | O9A.22 | inj, poison & oth conseq external causes complic childbirth |
| DX_ICD10 | O9A.23 | inj, poison & oth conseq external causes complic the puerperium |
| DX_ICD10 | O9A.3 | physical abuse complic preg, childbirth & the puerperium |
| DX_ICD10 | O9A.31 | physical abuse complic preg |
| DX_ICD10 | O9A.311 | physical abuse complic preg, first tri |
| DX_ICD10 | O9A.312 | physical abuse complic preg, second tri |
| DX_ICD10 | O9A.313 | physical abuse complic preg, third tri |
| DX_ICD10 | O9A.319 | physical abuse complic preg, unspec tri |
| DX_ICD10 | O9A.32 | physical abuse complic childbirth |
| DX_ICD10 | O9A.33 | physical abuse complic the puerperium |
| DX_ICD10 | O9A.4 | sexual abuse complic preg, childbirth & the puerperium |
| DX_ICD10 | O9A.41 | sexual abuse complic preg |
| DX_ICD10 | O9A.411 | sexual abuse complic preg, first tri |
| DX_ICD10 | O9A.412 | sexual abuse complic preg, second tri |
| DX_ICD10 | O9A.413 | sexual abuse complic preg, third tri |
| DX_ICD10 | O9A.419 | sexual abuse complic preg, unspec tri |
| DX_ICD10 | O9A.42 | sexual abuse complic childbirth |
| DX_ICD10 | O9A.43 | sexual abuse complic the puerperium |
| DX_ICD10 | O9A.5 | psychological abuse complic preg, childbirth & the puerperium |
| DX_ICD10 | O9A.51 | psychological abuse complic preg |
| DX_ICD10 | O9A.511 | psychological abuse complic preg, first tri |
| DX_ICD10 | O9A.512 | psychological abuse complic preg, second tri |
| DX_ICD10 | O9A.513 | psychological abuse complic preg, third tri |
| DX_ICD10 | O9A.519 | psychological abuse complic preg, unspec tri |
| DX_ICD10 | O9A.52 | psychological abuse complic childbirth |
| DX_ICD10 | O9A.53 | psychological abuse complic the puerperium |
| DX_ICD9 | V22.0 | supervis normal 1st preg |
| DX_ICD9 | V22.1 | supervis oth normal preg |
| DX_ICD9 | V22.2 | preg state- incidental |
| DX_ICD9 | V23.0 | supervision of high-risk preg with history of infertility |
| DX_ICD9 | V23.1 | supervision of high-risk preg with history of trophoblastic disease |
| DX_ICD9 | V23.2 | preg w hx of abortion |
| DX_ICD9 | V23.3 | supervision of high-risk preg with gr& multiparity |
| DX_ICD9 | V23.4 | preg w poor obstetric hx (end 2002) |
| DX_ICD9 | V23.41 | preg with history of pre-term labor (begin 2002) |
| DX_ICD9 | V23.42 | preg with history of ectopic preg |
| DX_ICD9 | V23.49 | preg with oth poor obstetric history |
| DX_ICD9 | V23.5 | supervision of high-risk preg with oth poor reproductive history |
| DX_ICD9 | V23.7 | supervision of high-risk preg with insufficient prenatal care |
| DX_ICD9 | V23.8 | suprv high-risk preg nec (end 1998) |
| DX_ICD9 | V23.81 | supervision of high-risk preg with elderly primigravida |
| DX_ICD9 | V23.82 | multigrav (35 or older) (begin 1998) |
| DX_ICD9 | V23.83 | primigravida (less than 16) (begin 1998) |
| DX_ICD9 | V23.84 | multigravida (less than 16) (begin 1998) |
| DX_ICD9 | V23.85 | pregnt-assist repro tech (begin 2008) |
| DX_ICD9 | V23.86 | preg-hx in utro prev prg (begin 2008) |
| DX_ICD9 | V23.87 | preg w incon fetl viabil (begin 2011) |
| DX_ICD9 | V23.89 | oth hi risk preg (begin 1998) |
| DX_ICD9 | V23.9 | suprv high-risk preg nos |
| DX_ICD9 | V24.0 | postpart care after del |
| DX_ICD9 | V24.1 | postpart care-lactation |
| DX_ICD9 | V24.2 | rout postpart follow-up |
| DX_ICD9 | V27.0 | deliver-single liveborn |
| DX_ICD9 | V27.1 | deliver- single stillborn |
| DX_ICD9 | V27.2 | outcome of delivery, twins, both liveborn |
| DX_ICD9 | V27.3 | outcome of delivery, twins, 1 liveborn & 1 stillborn |
| DX_ICD9 | V27.4 | outcome of delivery, twins, both stillborn |
| DX_ICD9 | V27.5 | outcome of delivery, oth multi birth, all liveborn |
| DX_ICD9 | V27.6 | outcome of delivery, oth multi birth, some liveborn |
| DX_ICD9 | V27.7 | deliver- multi births- all stillborn |
| DX_ICD9 | V27.9 | outcome of delivery nos |
| DX_ICD9 | V30.0 | single liveborn infant, delivered vaginally |
| DX_ICD9 | V30.00 | single liveborn born in hospital delivered wo cesarean section |
| DX_ICD9 | V30.01 | single liveborn, born in hospital, delivered by cesarean section |
| DX_ICD9 | V30.1 | single liveborn, born in hospital, delivered by cesarean section |
| DX_ICD9 | V30.2 | single liveborn, born outside hospital & not hospitalized |
| DX_ICD9 | V31.0 | twin, mate live born- in hospital |
| DX_ICD9 | V31.00 | twin birth, mate liveborn, born in hospital, delivered w/o c-section |
| DX_ICD9 | V31.01 | twin birth, mate liveborn, born in hospital, delivered by c-section |
| DX_ICD9 | V31.1 | twin birth, mate liveborn, born before admission to hospital |
| DX_ICD9 | V31.2 | twin birth, mate liveborn, born outside hospital & not hospitalized |
| DX_ICD9 | V32.0 | twin, mate stillborn- in hospital |
| DX_ICD9 | V32.00 | twin birth, mate stillborn, born in hospital, delivered w/o c-section |
| DX_ICD9 | V32.01 | twin birth, mate stillborn, born in hospital, delivered by c-section |
| DX_ICD9 | V32.1 | twin birth, mate stillborn, born before admission to hospital |
| DX_ICD9 | V32.2 | twin birth, mate stillborn, born outside hospital & not hoseltalized |
| DX_ICD9 | V33 | twin nos |
| DX_ICD9 | V33.0 | twin nos- in hospital |
| DX_ICD9 | V33.00 | twin birth, unspec if mate LB or SB, born in hospital, del wo c-section |
| DX_ICD9 | V33.01 | twin birth, unspec if mate LB or SB, born in hospital, del by c-section |
| DX_ICD9 | V33.1 | twin birth, unspec if mate LB or SB, born before admission to hospital |
| DX_ICD9 | V33.2 | twin birth, unspec if mate LB or SB, born outside hospl & not hosp |
| DX_ICD9 | V34.0 | oth multi liveborn- in hospital |
| DX_ICD9 | V34.00 | oth multi birth (3+), mates all LB, born in hosp, del w/o c-sect |
| DX_ICD9 | V34.01 | oth multi birth (3+), mates all LB, born in hosp, del by c-sect |
| DX_ICD9 | V34.1 | oth multibirth (3+), mates all LB, born before admiss to hosp |
| DX_ICD9 | V34.2 | oth multi birth (3+), mates all LB, born outside hosp & not hosp |
| DX_ICD9 | V35.0 | oth multi stillborn- in hospital |
| DX_ICD9 | V35.00 | oth multi birth (3+), mates all SB, born in hosp, del w/o c-section |
| DX_ICD9 | V35.01 | oth multi birth (3+), mates all SB, born in hosp, del by c-section |
| DX_ICD9 | V35.1 | oth multi birth (3+), mates all SB, born before admiss to hospl |
| DX_ICD9 | V35.2 | oth multi birth (3+), mates all SB, born outside of hosp & not hosp |
| DX_ICD9 | V36.0 | multi LB/SB- in hosp |
| DX_ICD9 | V36.00 | oth mult birth (3+), mates LB & SB, born in hosp, delivered w/o c-sect |
| DX_ICD9 | V36.01 | oth mult birth (3+), mates LB & SB, born in hosp, delivered w/o c-sect |
| DX_ICD9 | V36.1 | oth mult birth (3+), mates LB & SB, born before admis to hosp |
| DX_ICD9 | V36.2 | oth mult birth (3+), mates LB & SB, born outside hosp & not hosp |
| DX_ICD9 | V37 | oth/unspec mult births |
| DX_ICD9 | V37.0 | multi birth nos- in hosp |
| DX_ICD9 | V37.00 | oth mult brth (3+), unspec if mates LB or SB, born hosp, del w/o c-sect |
| DX_ICD9 | V37.01 | oth mult brth (3+), unspec if mates LB or SB, born hosp del by c-sect |
| DX_ICD9 | V37.1 | oth mult brth (3+), unspec if mates LB or SB, born b4 admiss to hosp |
| DX_ICD9 | V37.2 | oth mult brth (3+), unspec if mates LB or SB, born outside of hosp |
| DX_ICD9 | V39.0 | LB nos- in hospital |
| DX_ICD9 | V39.00 | LB, unspec if single, twin or multi, born in hosp, del w/o c-section |
| DX_ICD9 | V39.01 | LB, unspec if single, twin or multi, born in hosp, del by c-section |
| DX_ICD9 | V39.1 | LB, unspec if single, twin, or multi, born b4 admission to hosp |
| DX_ICD9 | V39.2 | LB, unspec if single, twin or multi, born outside hosp & not hosp |
| DX_ICD9 | V72.4 | preg test-positive (begin 2005) |
| DX_ICD9 | V72.42 | preg test-positive (begin 2005) |
| DX_ICD9 | V91.00 | twin gest-plac/sac nos (begin 2010) |
| DX_ICD9 | V91.01 | twin gest-monochr/monoam (begin 2010) |
| DX_ICD9 | V91.02 | twin gest-monochr/diamni (begin 2010) |
| DX_ICD9 | V91.03 | twin gest-dich/diamniotc (begin 2010) |
| DX_ICD9 | V91.09 | twin gest-plac/sac undet (begin 2010) |
| DX_ICD9 | V91.10 | tripl gest-plac/sac nos (begin 2010) |
| DX_ICD9 | V91.11 | trip gest 2+ monochor (begin 2010) |
| DX_ICD9 | V91.12 | trip gest 2+ monoamn (begin 2010) |
| DX_ICD9 | V91.19 | tripl gest-plac/sac und (begin 2010) |
| DX_ICD9 | V91.20 | quad gest-plac/sac nos (begin 2010) |
| DX_ICD9 | V91.21 | quad gest 2+ monochorion (begin 2010) |
| DX_ICD9 | V91.22 | quad gest 2+ monoamniotc (begin 2010) |
| DX_ICD9 | V91.29 | quad gest-plac/sac undet (begin 2010) |
| DX_ICD9 | V91.90 | mult gest-plac/sac nos (begin 2010) |
| DX_ICD9 | V91.91 | mult gest 2+ monochr nec (begin 2010) |
| DX_ICD9 | V91.92 | mult gest 2+ monoamn nec (begin 2010) |
| DX_ICD9 | V91.99 | mult gest-plac/sac undet (begin 2010) |
| DX_ICD10 | Z32 | encounter for preg test & childbirth & childcare instruction |
| DX_ICD10 | Z32.0 | encounter for preg test |
| DX_ICD10 | Z32.00 | encounter for preg test, result unknown) |
| DX_ICD10 | Z32.01 | encounter for preg test, result positive |
| DX_ICD10 | Z32.2 | encounter for childbirth instruction |
| DX_ICD10 | Z32.3 | encounter for childcare instruction |
| DX_ICD10 | Z33 | pregnant state |
| DX_ICD10 | Z33.1 | pregnant state, incidental |
| DX_ICD10 | Z33.2 | encounter for elect termination of preg |
| DX_ICD10 | Z33.3 | pregnant state, gestal carrier |
| DX_ICD10 | Z34 | encounter for supervision of normal preg |
| DX_ICD10 | Z34.0 | encounter for supervision of normal first preg |
| DX_ICD10 | Z34.00 | encounter for supervision of normal first preg, unspec tri |
| DX_ICD10 | Z34.01 | encounter for supervision of normal first preg, first tri |
| DX_ICD10 | Z34.02 | encounter for supervision of normal first preg, second tri |
| DX_ICD10 | Z34.03 | encounter for supervision of normal first preg, third tri |
| DX_ICD10 | Z34.8 | encounter for supervision of oth normal preg |
| DX_ICD10 | Z34.80 | encounter for supervision of oth normal preg, unspec tri |
| DX_ICD10 | Z34.81 | encounter for supervision of oth normal preg, first tri |
| DX_ICD10 | Z34.82 | encounter for supervision of oth normal preg, second tri |
| DX_ICD10 | Z34.83 | encounter for supervision of oth normal preg, third tri |
| DX_ICD10 | Z34.9 | encounter for supervision of normal preg, unspec |
| DX_ICD10 | Z34.90 | encounter for supervision of normal preg, unspec, unspec tri |
| DX_ICD10 | Z34.91 | encounter for supervision of normal preg, unspec, first tri |
| DX_ICD10 | Z34.92 | encounter for supervision of normal preg, unspec, second tri |
| DX_ICD10 | Z34.93 | encounter for supervision of normal preg, unspec, third tri |
| DX_ICD10 | Z36 | encounter for antenatal screening of moth |
| DX_ICD10 | Z36.0 | encounter for antenatal screening for chromosomal anomalies |
| DX_ICD10 | Z36.1 | encounter for antenatal screening for raised alphafetoprotein level |
| DX_ICD10 | Z36.2 | encounter for oth antenatal screening follow-up |
| DX_ICD10 | Z36.3 | encounter for antenatal screening for malformations |
| DX_ICD10 | Z36.4 | encounter for antenatal screening for fetal growth retardation |
| DX_ICD10 | Z36.5 | encounter for antenatal screening for isoimmunization |
| DX_ICD10 | Z36.81 | encounter for antenatal screening for hydrops fetalis |
| DX_ICD10 | Z36.82 | encounter for antenatal screening for nuchal translucency |
| DX_ICD10 | Z36.83 | encounter for fetal screening for congenital cardiac abnormalities |
| DX_ICD10 | Z36.84 | encounter for antenatal screening for fetal lung maturity |
| DX_ICD10 | Z36.85 | encounter for antenatal screening for streptococcus b |
| DX_ICD10 | Z36.86 | encounter for antenatal screening for cervical length |
| DX_ICD10 | Z36.87 | encounter for antenatal screening for uncertain dates |
| DX_ICD10 | Z36.88 | encounter for antenatal screening for fetal macrosomia |
| DX_ICD10 | Z36.89 | encounter for oth specif antenatal screening |
| DX_ICD10 | Z36.8A | encounter for antenatal screening for oth genetic defects |
| DX_ICD10 | Z36.9 | encounter for antenatal screening, unspec |
| DX_ICD10 | Z37 | outcome of delivery |
| DX_ICD10 | Z37.0 | single live birth |
| DX_ICD10 | Z37.1 | single stillbirth |
| DX_ICD10 | Z37.2 | twins, both liveborn |
| DX_ICD10 | Z37.3 | twins, one liveborn & one stillborn |
| DX_ICD10 | Z37.4 | twins, both stillborn |
| DX_ICD10 | Z37.5 | oth multi births, all liveborn |
| DX_ICD10 | Z37.50 | multi births, unspec, all liveborn |
| DX_ICD10 | Z37.51 | triplets, all liveborn |
| DX_ICD10 | Z37.52 | quadlets, all liveborn |
| DX_ICD10 | Z37.53 | quintuplets, all liveborn |
| DX_ICD10 | Z37.54 | sextuplets, all liveborn |
| DX_ICD10 | Z37.59 | oth multi births, all liveborn |
| DX_ICD10 | Z37.6 | oth multi births, some liveborn |
| DX_ICD10 | Z37.60 | multi births, unspec, some liveborn |
| DX_ICD10 | Z37.61 | triplets, some liveborn |
| DX_ICD10 | Z37.62 | quadlets, some liveborn |
| DX_ICD10 | Z37.63 | quintuplets, some liveborn |
| DX_ICD10 | Z37.64 | sextuplets, some liveborn |
| DX_ICD10 | Z37.69 | oth multi births, some liveborn |
| DX_ICD10 | Z37.7 | oth multi births, all stillborn |
| DX_ICD10 | Z37.9 | outcome of delivery, unspec |
| DX_ICD10 | Z38 | liveborn infants according to place of birth & type of delivery |
| DX_ICD10 | Z38.0 | single liveborn infant, born in hospital |
| DX_ICD10 | Z38.00 | single liveborn infant, delivered vaginally |
| DX_ICD10 | Z38.01 | single liveborn infant, delivered by cesarean |
| DX_ICD10 | Z38.1 | single liveborn infant, born outside hospital |
| DX_ICD10 | Z38.2 | single liveborn infant, unspec as to place of birth |
| DX_ICD10 | Z38.3 | twin liveborn infant, born in hospital |
| DX_ICD10 | Z38.30 | twin liveborn infant, delivered vaginally |
| DX_ICD10 | Z38.31 | twin liveborn infant, delivered by cesarean |
| DX_ICD10 | Z38.4 | twin liveborn infant, born outside hospital |
| DX_ICD10 | Z38.5 | twin liveborn infant, unspec as to place of birth |
| DX_ICD10 | Z38.6 | oth multi liveborn infant, born in hospital |
| DX_ICD10 | Z38.61 | trip liveborn infant, delivered vaginally |
| DX_ICD10 | Z38.62 | trip liveborn infant, delivered by cesarean |
| DX_ICD10 | Z38.63 | quad liveborn infant, delivered vaginally |
| DX_ICD10 | Z38.64 | quad liveborn infant, delivered by cesarean |
| DX_ICD10 | Z38.65 | quintuplet liveborn infant, delivered vaginally |
| DX_ICD10 | Z38.66 | quintuplet liveborn infant, delivered by cesarean |
| DX_ICD10 | Z38.68 | oth multi liveborn infant, delivered vaginally |
| DX_ICD10 | Z38.69 | oth multi liveborn infant, delivered by cesarean |
| DX_ICD10 | Z38.7 | oth multi liveborn infant, born outside hospital |
| DX_ICD10 | Z38.8 | oth multi liveborn infant, unspec as to place of birth |
| DX_ICD10 | Z39 | encounter for maternal postpartum care & examination |
| DX_ICD10 | Z39.0 | encounter for care & examination of moth immediately after delivery |
| DX_ICD10 | Z39.1 | encounter for care & examination of lactating moth |
| DX_ICD10 | Z39.2 | encounter for routine postpartum follow-up |
| DX_ICD10 | Z3A | weeks of gest |
| DX_ICD10 | Z3A.0 | weeks of gest of preg, unsp or less than 10 weeks |
| DX_ICD10 | Z3A.00 | weeks of gest of preg not specif |
| DX_ICD10 | Z3A.01 | less than 8 weeks gest of preg |
| DX_ICD10 | Z3A.08 | 8 weeks gest of preg |
| DX_ICD10 | Z3A.09 | 9 weeks gest of preg |
| DX_ICD10 | Z3A.1 | weeks of gest of preg, weeks 10-19 |
| DX_ICD10 | Z3A.10 | 10 weeks gest of preg |
| DX_ICD10 | Z3A.11 | 11 weeks gest of preg |
| DX_ICD10 | Z3A.12 | 12 weeks gest of preg |
| DX_ICD10 | Z3A.13 | 13 weeks gest of preg |
| DX_ICD10 | Z3A.14 | 14 weeks gest of preg |
| DX_ICD10 | Z3A.15 | 15 weeks gest of preg |
| DX_ICD10 | Z3A.16 | 16 weeks gest of preg |
| DX_ICD10 | Z3A.17 | 17 weeks gest of preg |
| DX_ICD10 | Z3A.18 | 18 weeks gest of preg |
| DX_ICD10 | Z3A.19 | 19 weeks gest of preg |
| DX_ICD10 | Z3A.2 | weeks of gest of preg, weeks 20-29 |
| DX_ICD10 | Z3A.20 | 20 weeks gest of preg |
| DX_ICD10 | Z3A.21 | 21 weeks gest of preg |
| DX_ICD10 | Z3A.22 | 22 weeks gest of preg |
| DX_ICD10 | Z3A.23 | 23 weeks gest of preg |
| DX_ICD10 | Z3A.24 | 24 weeks gest of preg |
| DX_ICD10 | Z3A.25 | 25 weeks gest of preg |
| DX_ICD10 | Z3A.26 | 26 weeks gest of preg |
| DX_ICD10 | Z3A.27 | 27 weeks gest of preg |
| DX_ICD10 | Z3A.28 | 28 weeks gest of preg |
| DX_ICD10 | Z3A.29 | 29 weeks gest of preg |
| DX_ICD10 | Z3A.3 | weeks of gest of preg, weeks 30-39 |
| DX_ICD10 | Z3A.30 | 30 weeks gest of preg |
| DX_ICD10 | Z3A.31 | 31 weeks gest of preg |
| DX_ICD10 | Z3A.32 | 32 weeks gest of preg |
| DX_ICD10 | Z3A.33 | 33 weeks gest of preg |
| DX_ICD10 | Z3A.34 | 34 weeks gest of preg |
| DX_ICD10 | Z3A.35 | 35 weeks gest of preg |
| DX_ICD10 | Z3A.36 | 36 weeks gest of preg |
| DX_ICD10 | Z3A.37 | 37 weeks gest of preg |
| DX_ICD10 | Z3A.38 | 38 weeks gest of preg |
| DX_ICD10 | Z3A.39 | 39 weeks gest of preg |
| DX_ICD10 | Z3A.4 | weeks of gest of preg, weeks 40 or greater |
| DX_ICD10 | Z3A.40 | 40 weeks gest of preg |
| DX_ICD10 | Z3A.41 | 41 weeks gest of preg |
| DX_ICD10 | Z3A.42 | 42 weeks gest of preg |
| DX_ICD10 | Z3A.49 | greater than 42 weeks gest of preg |
