## Supplemental Table S3 for "Maternal and Fetal Complications Among Pregnant Women with Congenital Heart Disease"

| **Supplemental Table S3: ICD and CPT Codes to define last menstrual period to determine beginning and end of each pregnancy (1750 codes)** | | | |
| --- | --- | --- | --- |
| TYPE | CODE | DESCRIPTION | GESTATIONAL AGE (weeks) |
| DX_ICD9 | 631.0 | inapp chg hcg early preg (begin 2011) | 8 |
| DX_ICD9 | 631 | oth abn prod conception (end 2011) | 8 |
| DX_ICD9 | 631.8 | oth abn prod conception (begin 2011) | 8 |
| DX_ICD9 | 633.0 | abdominal pregnancy (end 2002) | 8 |
| DX_ICD9 | 633.00 | abdominal pregnancy w/o intrauterine pregnancy (begin 2002) | 8 |
| DX_ICD9 | 633.01 | abdominal pregnancy w intrauterine pregnancy (begin 2002) | 8 |
| DX_ICD9 | 633.1 | tubal pregnancy (end 2002) | 8 |
| DX_ICD9 | 633.10 | tubal pregnancy w/o intrauterine pregnancy (begin 2002) | 8 |
| DX_ICD9 | 633.11 | tubal pregnancy w intrauterine pregnancy (begin 2002) | 8 |
| DX_ICD9 | 633.2 | ovarian pregnancy (end 2002) | 8 |
| DX_ICD9 | 633.20 | ovarian pregnancy w/o intrauterine pregnancy (begin 2002) | 8 |
| DX_ICD9 | 633.21 | ovarian pregnancy w intrauterine pregnancy (begin 2002) | 8 |
| DX_ICD9 | 633.8 | ectopic pregnancy nec (end 2002) | 8 |
| DX_ICD9 | 633.80 | ot ectopic pregnancy w/o intrau preg (begin 2002) | 8 |
| DX_ICD9 | 633.81 | ot ectopic pregnancy w intrauterine pregnancy (begin 2002) | 8 |
| DX_ICD9 | 633.9 | ectopic pregnancy nos (end 2002) | 8 |
| DX_ICD9 | 633.90 | unspec ectopic pregnancy w/o intrauterine preg (begin 2002) | 8 |
| DX_ICD9 | 633.91 | unspec ectopic pregnancy w intrauterine preg (begin 2002) | 8 |
| DX_ICD9 | 634.00 | spontaneous abortion w pelvic infection - unspecified | 8 |
| DX_ICD9 | 634.01 | spontaneous abortion w pelvic infection- incomplete | 8 |
| DX_ICD9 | 634.02 | spontaneous abortion w pelvic infection- complete | 8 |
| DX_ICD9 | 634.10 | spontaneous abortion w hem- unspecified | 8 |
| DX_ICD9 | 634.11 | spontaneous abortion w hem- incomplete | 8 |
| DX_ICD9 | 634.12 | spontaneous abortion w hem- complete | 8 |
| DX_ICD9 | 634.20 | spontaneous abortion w pelvic damage- unspecified | 8 |
| DX_ICD9 | 634.21 | spontaneous abortion w pelvic damage- incomplete | 8 |
| DX_ICD9 | 634.22 | spontaneous abortion w pelvic damage- complete | 8 |
| DX_ICD9 | 634.30 | spontaneous abortion w renal failure- unspecified | 8 |
| DX_ICD9 | 634.31 | spontaneous abortion w renal failure- incomplete | 8 |
| DX_ICD9 | 634.32 | spontaneous abortion w renal failure- complete | 8 |
| DX_ICD9 | 634.40 | spontaneous abortion w metabolic disease- unspecified | 8 |
| DX_ICD9 | 634.41 | spontaneous abortion w metabolic disease- incomplete | 8 |
| DX_ICD9 | 634.42 | spontaneous abortion w metabolic disease | 8 |
| DX_ICD9 | 634.50 | spontaneous abortion w shock- unspecified | 8 |
| DX_ICD9 | 634.51 | spontaneous abortion w shock- incomplete | 8 |
| DX_ICD9 | 634.52 | spontaneous abortion w shock- complete | 8 |
| DX_ICD9 | 634.60 | spontaneous abortion w embol- unspecified | 8 |
| DX_ICD9 | 634.61 | spontaneous abortion w embol- incomplete | 8 |
| DX_ICD9 | 634.62 | spontaneous abortion w embol- complete | 8 |
| DX_ICD9 | 634.70 | spontaneous abortion w compl nec- unspecified 63470 | 8 |
| DX_ICD9 | 634.71 | spontaneous abortion, with other specified complications, incomplete | 8 |
| DX_ICD9 | 634.72 | spontaneous abortion w compl nec- complete | 8 |
| DX_ICD9 | 634.80 | spontaneous abortion w compl nos- unspecified | 8 |
| DX_ICD9 | 634.81 | spontaneous abortion w compl nos- incomplete | 8 |
| DX_ICD9 | 634.82 | spontaneous abortion w compl nos- complete | 8 |
| DX_ICD9 | 634.90 | spontaneous abortion uncompl- unspecified | 8 |
| DX_ICD9 | 634.91 | spontaneous abortion uncompl- incomplete | 8 |
| DX_ICD9 | 634.92 | spontaneous abortion uncompl- complete | 8 |
| DX_ICD9 | 761.4 | ectopic pregnancy affecting newborn | 8 |
| DX_ICD9 | 779.6 | termination of pregnancy | 8 |
| PX_ICD9 | 66.62 | salpingectomy with removal of tubal pregnancy | 8 |
| PX_ICD9 | 74.3 | removal of extratubal ectopic pregnancy | 8 |
| DX_ICD10 | O00 | abdominal pregnancy | 8 |
| DX_ICD10 | O00.0 | abdominal pregnancy | 8 |
| DX_ICD10 | O00.00 | abdominal pregnancy without intrauterine pregnancy | 8 |
| DX_ICD10 | O00.01 | abdominal pregnancy with intrauterine pregnancy | 8 |
| DX_ICD10 | O00.1 | tubal pregnancy | 8 |
| DX_ICD10 | O00.10 | tubal pregnancy without intrauterine pregnancy | 8 |
| DX_ICD10 | O00.101 | right tubal pregnancy without intrauterine pregnancy | 8 |
| DX_ICD10 | O00.102 | left tubal pregnancy without intrauterine pregnancy | 8 |
| DX_ICD10 | O00.109 | unspecified tubal pregnancy without intrauterine pregnancy | 8 |
| DX_ICD10 | O00.11 | tubal pregnancy with intrauterine pregnancy | 8 |
| DX_ICD10 | O00.111 | right tubal pregnancy with intrauterine pregnancy | 8 |
| DX_ICD10 | O00.112 | left tubal pregnancy with intrauterine pregnancy | 8 |
| DX_ICD10 | O00.119 | unspecified tubal pregnancy with intrauterine pregnancy | 8 |
| DX_ICD10 | O00.2 | ovarian pregnancy | 8 |
| DX_ICD10 | O00.20 | ovarian pregnancy without intrauterine pregnancy | 8 |
| DX_ICD10 | O00.201 | right ovarian pregnancy without intrauterine pregnancy | 8 |
| DX_ICD10 | O00.202 | left ovarian pregnancy without intrauterine pregnancy | 8 |
| DX_ICD10 | O00.209 | unspecified ovarian pregnancy without intrauterine pregnancy | 8 |
| DX_ICD10 | O00.21 | ovarian pregnancy with intrauterine pregnancy | 8 |
| DX_ICD10 | O00.211 | right ovarian pregnancy with intrauterine pregnancy | 8 |
| DX_ICD10 | O00.212 | left ovarian pregnancy with intrauterine pregnancy | 8 |
| DX_ICD10 | O00.219 | unspecified ovarian pregnancy with intrauterine pregnancy | 8 |
| DX_ICD10 | O00.8 | other ectopic pregnancy | 8 |
| DX_ICD10 | O00.80 | other ectopic pregnancy without intrauterine pregnancy | 8 |
| DX_ICD10 | O00.81 | other ectopic pregnancy with intrauterine pregnancy | 8 |
| DX_ICD10 | O00.9 | ectopic pregnancy, unspecified | 8 |
| DX_ICD10 | O00.90 | unspecified ectopic pregnancy without intrauterine pregnancy | 8 |
| DX_ICD10 | O00.91 | unspecified ectopic pregnancy with intrauterine pregnancy | 8 |
| DX_ICD10 | O03 | spontaneous abortion | 8 |
| DX_ICD10 | O03.0 | genital tract and pelvic infection after incomplete spontaneous abortion | 8 |
| DX_ICD10 | O03.1 | delayed or excessive hemorrhage after incomplete spontaneous abortion | 8 |
| DX_ICD10 | O03.2 | embolism following incomplete spontaneous abortion | 8 |
| DX_ICD10 | O03.3 | genital tract and pelvic infection after incomplete spontaneous abortion | 8 |
| DX_ICD10 | O03.30 | unspecified complication following incomplete spontaneous abortion | 8 |
| DX_ICD10 | O03.31 | shock following incomplete spontaneous abortion | 8 |
| DX_ICD10 | O03.32 | renal failure following incomplete spontaneous abortion | 8 |
| DX_ICD10 | O03.33 | metabolic disorder following incomplete spontaneous abortion | 8 |
| DX_ICD10 | O03.34 | damage to pelvic organs following incomplete spontaneous abortion | 8 |
| DX_ICD10 | O03.35 | other venous complications following incomplete spontaneous abortion | 8 |
| DX_ICD10 | O03.36 | cardiac arrest following incomplete spontaneous abortion | 8 |
| DX_ICD10 | O03.37 | sepsis following incomplete spontaneous abortion | 8 |
| DX_ICD10 | O03.38 | urinary tract infection following incomplete spontaneous abortion | 8 |
| DX_ICD10 | O03.39 | incomplete spontaneous abortion with other complications | 8 |
| DX_ICD10 | O03.4 | incomplete spontaneous abortion without complication | 8 |
| DX_ICD10 | O03.5 | genital tract and pelvic infection after complete or unspec spontan abortion | 8 |
| DX_ICD10 | O03.6 | delayed or excessive hemorrhage after complete or unspec spontan abortion | 8 |
| DX_ICD10 | O03.7 | embolism following complete or unspecified spontaneous abortion | 8 |
| DX_ICD10 | O03.8 | other and unspec complications after complete or unspec spontan abortion | 8 |
| DX_ICD10 | O03.80 | Unspec complication after complete or unspecified spontaneous abortion | 8 |
| DX_ICD10 | O03.81 | shock following complete or unspecified spontaneous abortion | 8 |
| DX_ICD10 | O03.82 | renal failure following complete or unspecified spontaneous abortion | 8 |
| DX_ICD10 | O03.83 | metabolic disorder following complete or unspecified spontaneous abortion | 8 |
| DX_ICD10 | O03.84 | damage to pelvic organs after complete or unspecified spontaneous abortion | 8 |
| DX_ICD10 | O03.85 | other venous complications after complete or unspec spontaneous abortion | 8 |
| DX_ICD10 | O03.86 | cardiac arrest following complete or unspecified spontaneous abortion | 8 |
| DX_ICD10 | O03.87 | sepsis following complete or unspecified spontaneous abortion | 8 |
| DX_ICD10 | O03.88 | urinary tract infection after complete or unspec spontaneous abortion | 8 |
| DX_ICD10 | O03.89 | complete or unspecified spontaneous abortion with other complications | 8 |
| DX_ICD10 | O03.9 | complete or unspecified spontaneous abortion without complication | 8 |
| DX_ICD10 | O08 | complications following ectopic and molar pregnancy | 8 |
| DX_ICD10 | Z3A.01 | less than 8 weeks gestation of pregnancy | 8 |
| DX_ICD10 | Z3A.08 | 8 weeks gestation of pregnancy | 8 |
| PX_ICD10 | 10D27ZZ | extraction of products of conception, ectopic, via natural or artificial opening | 8 |
| PX_ICD10 | 10D28ZZ | extraction of products of concept, ectopic, via natural or artif open endoscop | 8 |
| PX_ICD10 | 10J20ZZ | inspection of products of conception, ectopic, open approach | 8 |
| PX_ICD10 | 10J23ZZ | inspection of products of conception, ectopic, percutaneous approach | 8 |
| PX_ICD10 | 10J24ZZ | inspection of products of concept, ectopic, percutan endoscopic approach | 8 |
| PX_ICD10 | 10J27ZZ | inspection of products of conception, ectopic, via natural or artificial opening | 8 |
| PX_ICD10 | 10J28ZZ | inspect of products of concept, ectopic, via natural or artif open endoscopic | 8 |
| PX_ICD10 | 10J2XZZ | inspection of products of conception, ectopic, external approach | 8 |
| PX_ICD10 | 10S20ZZ | reposition products of conception, ectopic, open approach | 8 |
| PX_ICD10 | 10S23ZZ | reposition products of conception, ectopic, percutaneous approach | 8 |
| PX_ICD10 | 10S24ZZ | reposition products of conception, ectopic, percutan endoscopic approach | 8 |
| PX_ICD10 | 10S27ZZ | reposition products of conception, ectopic, via natural or artificial opening | 8 |
| PX_ICD10 | 10S28ZZ | reposition products of concept, ectopic, via natural or artif open endoscopic | 8 |
| PX_ICD10 | 10T20ZZ | resection of products of conception, ectopic, open approach | 8 |
| PX_ICD10 | 10T23ZZ | resection of products of conception, ectopic, percutaneous approach | 8 |
| PX_ICD10 | 10T24ZZ | resection of products of conception, ectopic, percutan endoscopic approach | 8 |
| PX_ICD10 | 10T27ZZ | resection of products of conception, ectopic, via natural or artificial opening | 8 |
| PX_ICD10 | 10T28ZZ | resect of products of concept, ectopic, via natural or artif open endoscopic | 8 |
| PX_CPT | 59120 | surg tx ectop preg; tubal or ovarian, req salpingectomy +/- oophorectomy, ab | 8 |
| PX_CPT | 59121 | surgical tx ectop preg; tubal or ovarian, wo salpingectomy +/- oophorectomy | 8 |
| PX_CPT | 59130 | surgical treatment of ectopic pregnancy; abdominal pregnancy | 8 |
| PX_CPT | 59135 | surgical tx ectop preg; interstitial, uterine pregnancy req total hysterectomy | 8 |
| PX_CPT | 59136 | surgical tx ectop preg; interstitial, uterine preg w partial resection of uterus | 8 |
| PX_CPT | 59140 | surgical treatment of ectopic pregnancy; cervical, with evacuation | 8 |
| PX_CPT | 59150 | laparoscopic txf ectop preg; wo salpingectomy and/or oophorectomy | 8 |
| PX_CPT | 59151 | laparoscopic tx ectop preg; w salpingectomy and/or oophorectomy | 8 |
| DX_ICD10 | Z3A.09 | 9 weeks gestation of pregnancy | 9 |
| DX_ICD9 | 635.00 | legal abortion w pelvic infection- unspecified | 10 |
| DX_ICD9 | 635.01 | legal abortion w pelvic infection- incomplete | 10 |
| DX_ICD9 | 635.02 | legal abortion w pelvic infection- complete | 10 |
| DX_ICD9 | 635.10 | legal abortion w hem- unspecified | 10 |
| DX_ICD9 | 635.11 | legal abortion w hem- incomplete | 10 |
| DX_ICD9 | 635.12 | legal abortion w hem- complete | 10 |
| DX_ICD9 | 635.20 | legal abortion w pelvic damage- unspecified | 10 |
| DX_ICD9 | 635.21 | legal abortion w pelvic damage- incomplete | 10 |
| DX_ICD9 | 635.22 | legal abortion w pelvic damage- complete | 10 |
| DX_ICD9 | 635.30 | legal abortion w renal failure- unspecified | 10 |
| DX_ICD9 | 635.31 | legal abortion w renal failure- incomplete | 10 |
| DX_ICD9 | 635.32 | legal abortion w renal failure- complete | 10 |
| DX_ICD9 | 635.40 | legal abortion w metabolic disease- unspecified | 10 |
| DX_ICD9 | 635.41 | legal abortion w metabolic disease- incomplete | 10 |
| DX_ICD9 | 635.42 | legal abortion w metabolic disease- complete | 10 |
| DX_ICD9 | 635.50 | legal abortion w shock- unspecified | 10 |
| DX_ICD9 | 635.51 | legal abortion w shock- incomplete | 10 |
| DX_ICD9 | 635.52 | legal abortion w shock- complete | 10 |
| DX_ICD9 | 635.60 | legal abortion w embolism- unspecified | 10 |
| DX_ICD9 | 635.61 | legal abortion w embolism- incomplete | 10 |
| DX_ICD9 | 635.62 | legal abortion w embolism- complete | 10 |
| DX_ICD9 | 635.70 | legal abortion w compl nec- unspecified | 10 |
| DX_ICD9 | 635.71 | legal abortion w compl nec- incomplete | 10 |
| DX_ICD9 | 635.72 | legal abortion w compl nec- complete | 10 |
| DX_ICD9 | 635.80 | legal abortion w compl nos- unspecified | 10 |
| DX_ICD9 | 635.81 | legal abortion w compl nos- incomplete | 10 |
| DX_ICD9 | 635.82 | legal abortion w compl nos- complete | 10 |
| DX_ICD9 | 635.90 | legal abortion uncompl- unspecified | 10 |
| DX_ICD9 | 635.91 | legal abortion uncompl- incomplete | 10 |
| DX_ICD9 | 635.92 | legal abortion uncompl- complete | 10 |
| DX_ICD9 | 636.00 | illegal abortion w pelvic infection- unspecified | 10 |
| DX_ICD9 | 636.01 | illegal abortion w pelvic infection- incomplete | 10 |
| DX_ICD9 | 636.02 | illegal abortion w pelvic infection- complete | 10 |
| DX_ICD9 | 636.10 | illegal abortion w hem- unspecified | 10 |
| DX_ICD9 | 636.11 | illegal abortion w hem- incomplete | 10 |
| DX_ICD9 | 636.12 | illegal abortion w hem- complete | 10 |
| DX_ICD9 | 636.20 | illegal abortion w pelvic damage- unspecified | 10 |
| DX_ICD9 | 636.21 | illegal abortion w pelvic damage- incomplete | 10 |
| DX_ICD9 | 636.22 | illegal abortion w pelvic damage- complete | 10 |
| DX_ICD9 | 636.30 | illegal abortion w renal failure- unspecified | 10 |
| DX_ICD9 | 636.31 | illegal abortion w renal failure- incomplete | 10 |
| DX_ICD9 | 636.32 | illegal abortion w renal failure- complete | 10 |
| DX_ICD9 | 636.40 | illegal abortion w metabolic disease- unspecified | 10 |
| DX_ICD9 | 636.41 | illegal abortion w metabolic disease- incomplete | 10 |
| DX_ICD9 | 636.42 | illegal abortion w metabolic disease- complete | 10 |
| DX_ICD9 | 636.50 | illegal abortion w shock- unspecified | 10 |
| DX_ICD9 | 636.51 | illegal abortion w shock- incomplete | 10 |
| DX_ICD9 | 636.52 | illegal abortion w shock- complete | 10 |
| DX_ICD9 | 636.60 | illegal abortion w embolism- unspecified | 10 |
| DX_ICD9 | 636.61 | illegal abortion w embolism- incomplete | 10 |
| DX_ICD9 | 636.62 | illegal abortion w embolism- complete | 10 |
| DX_ICD9 | 636.70 | illegal abortion w compl nec- unspecified | 10 |
| DX_ICD9 | 636.71 | illegal abortion w compl nec- incomplete | 10 |
| DX_ICD9 | 636.72 | illegal abortion w compl nec- complete | 10 |
| DX_ICD9 | 636.80 | illegal abortion w compl nos- unspecified | 10 |
| DX_ICD9 | 636.81 | illegal abortion w compl nos- incomplete | 10 |
| DX_ICD9 | 636.82 | illegal abortion w compl nos- complete | 10 |
| DX_ICD9 | 636.90 | illegal abortion uncompl- unspecified | 10 |
| DX_ICD9 | 636.91 | illegal abortion uncompl- incomplete | 10 |
| DX_ICD9 | 636.92 | illegal abortion uncompl complete | 10 |
| DX_ICD9 | 637.00 | abortion nos w pelvic infection- unspecified | 10 |
| DX_ICD9 | 637.01 | abortion nos w pelvic infection- incomplete | 10 |
| DX_ICD9 | 637.02 | abortion nos w pelvic infection- complete | 10 |
| DX_ICD9 | 637.10 | abortion nos w hem- unspecified | 10 |
| DX_ICD9 | 637.11 | abortion nos w hem- incomplete | 10 |
| DX_ICD9 | 637.12 | abortion nos w hem- complete | 10 |
| DX_ICD9 | 637.20 | abortion nos w pelvic damage- unspecified | 10 |
| DX_ICD9 | 637.21 | abortion nos w pelvic damage- incomplete | 10 |
| DX_ICD9 | 637.22 | abortion nos w pelvic damage- complete | 10 |
| DX_ICD9 | 637.30 | abortion nos w renal failure- unspecified | 10 |
| DX_ICD9 | 637.31 | abortion nos w renal failure- incomplete | 10 |
| DX_ICD9 | 637.32 | abortion nos w renal failure- complete | 10 |
| DX_ICD9 | 637.40 | abortion nos w metabolic disease- unspecified | 10 |
| DX_ICD9 | 637.41 | abortion nos w metabolic disease- incomplete | 10 |
| DX_ICD9 | 637.42 | abortion nos w metabolic disease- complete | 10 |
| DX_ICD9 | 637.50 | abortion nos w shock- unspecified | 10 |
| DX_ICD9 | 637.51 | abortion nos w shock- incomplete | 10 |
| DX_ICD9 | 637.52 | abortion nos w shock- complete | 10 |
| DX_ICD9 | 637.60 | abortion nos w embolism- unspecified | 10 |
| DX_ICD9 | 637.61 | abortion nos w embolism- incomplete | 10 |
| DX_ICD9 | 637.62 | abortion nos w embolism- complete | 10 |
| DX_ICD9 | 637.70 | abortion nos w compl nec- unspecified | 10 |
| DX_ICD9 | 637.71 | abortion nos w compl nec- incomplete | 10 |
| DX_ICD9 | 637.72 | abortion nos w compl nec- complete | 10 |
| DX_ICD9 | 637.80 | abortion nos w compl nos- unspecified | 10 |
| DX_ICD9 | 637.81 | abortion nos w compl nos- incomplete | 10 |
| DX_ICD9 | 637.82 | abortion nos w compl nos- complete | 10 |
| DX_ICD9 | 637.90 | ab nos uncomplicat-unsp | 10 |
| DX_ICD9 | 637.91 | ab nos uncomplicat-inc | 10 |
| DX_ICD9 | 637.92 | ab nos uncomplicat-comp | 10 |
| DX_ICD9 | 638.0 | attempted abortion w pelvic infection | 10 |
| DX_ICD9 | 638.1 | attempted abortion w hemorrhage | 10 |
| DX_ICD9 | 638.2 | attempted abortion w pelvic damage | 10 |
| DX_ICD9 | 638.3 | attempted abortion w renal failure | 10 |
| DX_ICD9 | 638.4 | attempted abortion w metabolic disease | 10 |
| DX_ICD9 | 638.5 | attempted abortion w shock | 10 |
| DX_ICD9 | 638.7 | attempted abortion w compl nec | 10 |
| DX_ICD9 | 638.8 | attempted abortion w compl nos | 10 |
| DX_ICD9 | 638.9 | attempted abort uncompl | 10 |
| DX_ICD9 | 639.5 | post abortion shock | 10 |
| PX_ICD9 | 69.01 | dilation and curettage for termination of pregnancy | 10 |
| PX_ICD9 | 69.51 | aspiration curettage of uterus for termination of pregnancy | 10 |
| DX_ICD10 | O02.0 | blighted ovum and nonhydatidiform mole | 10 |
| DX_ICD10 | O04 | complications following (induced) termination of pregnancy | 10 |
| DX_ICD10 | O04.5 | genital tract & pelvic infection after (induced) termination of preg | 10 |
| DX_ICD10 | O04.6 | delayed or excessive hemorrhage after (induced) termination of preg | 10 |
| DX_ICD10 | O04.7 | embolism following (induced) termination of pregnancy | 10 |
| DX_ICD10 | O04.8 | (induced) termination of pregnancy with other and unspecified complications | 10 |
| DX_ICD10 | O04.80 | (induced) termination of pregnancy with unspecified complications | 10 |
| DX_ICD10 | O04.81 | shock following (induced) termination of pregnancy | 10 |
| DX_ICD10 | O04.82 | renal failure following (induced) termination of pregnancy | 10 |
| DX_ICD10 | O04.83 | metabolic disorder following (induced) termination of pregnancy | 10 |
| DX_ICD10 | O04.84 | damage to pelvic organs following (induced) termination of pregnancy | 10 |
| DX_ICD10 | O04.85 | other venous complications following (induced) termination of pregnancy | 10 |
| DX_ICD10 | O04.86 | cardiac arrest following (induced) termination of pregnancy | 10 |
| DX_ICD10 | O04.87 | sepsis following (induced) termination of pregnancy | 10 |
| DX_ICD10 | O04.88 | urinary tract infection following (induced) termination of pregnancy | 10 |
| DX_ICD10 | O04.89 | (induced) termination of pregnancy with other complications | 10 |
| DX_ICD10 | O07.0 | genital tract & pelvic infection after failed attempted termination of pregnancy | 10 |
| DX_ICD10 | O07.1 | delayed or excessive hemorrhage after failed attempted termination of preg | 10 |
| DX_ICD10 | O07.30 | failed attempted termination of pregnancy with unspecified complications | 10 |
| DX_ICD10 | O07.31 | shock following failed attempted termination of pregnancy | 10 |
| DX_ICD10 | O07.32 | renal failure following failed attempted termination of pregnancy | 10 |
| DX_ICD10 | O07.33 | metabolic disorder following failed attempted termination of pregnancy | 10 |
| DX_ICD10 | O07.34 | damage to pelvic organs following failed attempted termination of pregnancy | 10 |
| DX_ICD10 | O07.35 | other venous complications after failed attempted termination of pregnancy | 10 |
| DX_ICD10 | O07.36 | cardiac arrest following failed attempted termination of pregnancy | 10 |
| DX_ICD10 | O07.37 | sepsis following failed attempted termination of pregnancy | 10 |
| DX_ICD10 | O07.38 | urinary tract infection following failed attempted termination of pregnancy | 10 |
| DX_ICD10 | O07.39 | failed attempted termination of pregnancy with other complications | 10 |
| DX_ICD10 | O07.4 | failed attempted termination of pregnancy without complication | 10 |
| DX_ICD10 | O08.0 | genital tract and pelvic infection following ectopic and molar pregnancy | 10 |
| DX_ICD10 | O08.1 | delayed or excessive hemorrhage following ectopic and molar pregnancy | 10 |
| DX_ICD10 | O08.2 | embolism following ectopic and molar pregnancy | 10 |
| DX_ICD10 | O08.3 | shock following ectopic and molar pregnancy | 10 |
| DX_ICD10 | O08.4 | renal failure following ectopic and molar pregnancy | 10 |
| DX_ICD10 | O08.5 | metabolic disorders following an ectopic and molar pregnancy | 10 |
| DX_ICD10 | O08.6 | damage to pelvic organs and tissues following an ectopic and molar pregnancy | 10 |
| DX_ICD10 | O08.7 | other venous complications following an ectopic and molar pregnancy | 10 |
| DX_ICD10 | O08.8 | other complications following an ectopic and molar pregnancy | 10 |
| DX_ICD10 | O08.81 | cardiac arrest following an ectopic and molar pregnancy | 10 |
| DX_ICD10 | O08.82 | sepsis following ectopic and molar pregnancy | 10 |
| DX_ICD10 | O08.83 | urinary tract infection following an ectopic and molar pregnancy | 10 |
| DX_ICD10 | O08.89 | other complications following an ectopic and molar pregnancy | 10 |
| DX_ICD10 | O08.9 | unspecified complication following an ectopic and molar pregnancy | 10 |
| DX_ICD10 | Z33.2 | encounter for elective termination of pregnancy | 10 |
| DX_ICD10 | Z3A.10 | 10 weeks gestation of pregnancy | 10 |
| PX_ICD10 | 10A03ZZ | abortion of products of conception, percutaneous approach | 10 |
| PX_ICD10 | 10A04ZZ | abortion of products of conception, percutaneous endoscopic approach | 10 |
| PX_ICD10 | 10A07Z6 | abortion of products of conception, vacuum, via natural or artificial opening | 10 |
| PX_ICD10 | 10A07ZX | abortion of products of concept, abortifacient, via natural or artificial opening | 10 |
| PX_ICD10 | 10A07ZZ | abortion of products of conception, via natural or artificial opening | 10 |
| PX_ICD10 | 10A08ZZ | abortion of products of conception, via natural or artificial opening endoscopic | 10 |
| PX_CPT | 1966 | anesthesia for induced abortion procedures | 10 |
| PX_CPT | 59840 | induced abortion, by dilation and curettage | 10 |
| PX_CPT | 59856 | induced abortion | 10 |
| PX_CPT | 59857 | induced abortion | 10 |
| DX_ICD10 | Z3A.11 | 11 weeks gestation of pregnancy | 11 |
| DX_ICD9 | 632 | missed abortion | 12 |
| DX_ICD9 | 768.3 | fetal distress first noted during labor and delivery, in liveborn infant | 12 |
| DX_ICD10 | O01.0 | classical hydatidiform mole | 12 |
| DX_ICD10 | O01.1 | incomplete and partial hydatidiform mole | 12 |
| DX_ICD10 | O01.9 | hydatidiform mole, unspecified | 12 |
| DX_ICD10 | O02.1 | missed abortion | 12 |
| DX_ICD10 | O09.01 | supervision of pregnancy with history of infertility, first trimester | 12 |
| DX_ICD10 | O09.11 | supervision of pregnancy with history of ectopic pregnancy, first trimester | 12 |
| DX_ICD10 | O09.211 | supervision of pregnancy with history of pre-term labor, first trimester | 12 |
| DX_ICD10 | O09.291 | supervision of preg with other poor reproductive or obstetric history, first tri | 12 |
| DX_ICD10 | O09.31 | supervision of pregnancy with insufficient antenatal care, first trimester | 12 |
| DX_ICD10 | O09.41 | supervision of pregnancy with grand multiparity, first trimester | 12 |
| DX_ICD10 | O09.511 | supervision of elderly primigravida, first trimester | 12 |
| DX_ICD10 | O09.521 | supervision of elderly multigravida, first trimester | 12 |
| DX_ICD10 | O09.611 | supervision of young primigravida, first trimester | 12 |
| DX_ICD10 | O09.621 | supervision of young multigravida, first trimester | 12 |
| DX_ICD10 | O09.71 | supervision of high risk pregnancy due to social problems, first trimester | 12 |
| DX_ICD10 | O09.811 | supervision of preg resulting from assisted reproductive technology, first tri | 12 |
| DX_ICD10 | O09.821 | supervision of preg w hx of in utero procedure during prior preg, first trimester | 12 |
| DX_ICD10 | O09.891 | supervision of other high risk pregnancies, first trimester | 12 |
| DX_ICD10 | O09.91 | supervision of high risk pregnancy, unspecified, first trimester | 12 |
| DX_ICD10 | O09.A1 | supervision of pregnancy with history of molar pregnancy, first trimester | 12 |
| DX_ICD10 | O10.011 | pre-existing essential hypertension complicating pregnancy, first trimester | 12 |
| DX_ICD10 | O10.111 | pre-existing hypertensive heart disease complicating pregnancy, first trimester | 12 |
| DX_ICD10 | O10.211 | pre-existing hypertensive chronic kidney disease complicating preg, first tri | 12 |
| DX_ICD10 | O10.311 | pre-existing hypertensive heart & chronic kidney dis complicating preg, first tri | 12 |
| DX_ICD10 | O10.911 | unspecified pre-existing hypertension complicating pregnancy, first trimester | 12 |
| DX_ICD10 | O11.1 | pre-existing hypertension with pre-eclampsia, first trimester | 12 |
| DX_ICD10 | O11.1 | pre-existing hypertension with pre-eclampsia, first trimester | 12 |
| DX_ICD10 | O12.01 | gestational edema, first trimester | 12 |
| DX_ICD10 | O12.11 | gestational proteinuria, first trimester | 12 |
| DX_ICD10 | O12.21 | gestational edema with proteinuria, first trimester | 12 |
| DX_ICD10 | O13.1 | gestational [preg-induced] hypertension without significant proteinuria, first tri | 12 |
| DX_ICD10 | O16.1 | unspecified maternal hypertension, first trimester | 12 |
| DX_ICD10 | O22.01 | varicose veins of lower extremity in pregnancy, first trimester | 12 |
| DX_ICD10 | O22.11 | genital varices in pregnancy, first trimester | 12 |
| DX_ICD10 | O22.21 | superficial thrombophlebitis in pregnancy, first trimester | 12 |
| DX_ICD10 | O22.31 | deep phlebothrombosis in pregnancy, first trimester | 12 |
| DX_ICD10 | O22.41 | cerebral venous thrombosis in pregnancy, first trimester | 12 |
| DX_ICD10 | O22.51 | cerebral venous thrombosis in pregnancy, first trimester | 12 |
| DX_ICD10 | O22.8X1 | other venous complications in pregnancy, first trimester | 12 |
| DX_ICD10 | O22.91 | venous complication in pregnancy, unspecified, first trimester | 12 |
| DX_ICD10 | O23.01 | infections of kidney in pregnancy, first trimester | 12 |
| DX_ICD10 | O23.11 | infections of bladder in pregnancy, first trimester | 12 |
| DX_ICD10 | O23.21 | infections of urethra in pregnancy, first trimester | 12 |
| DX_ICD10 | O23.31 | infections of other parts of urinary tract in pregnancy, first trimester | 12 |
| DX_ICD10 | O23.41 | unspecified infection of urinary tract in pregnancy, first trimester | 12 |
| DX_ICD10 | O23.511 | infections of cervix in pregnancy, first trimester | 12 |
| DX_ICD10 | O23.521 | salpingo-oophoritis in pregnancy, first trimester | 12 |
| DX_ICD10 | O23.591 | infection of other part of genital tract in pregnancy, first trimester | 12 |
| DX_ICD10 | O23.91 | unspecified genitourinary tract infection in pregnancy, first trimester | 12 |
| DX_ICD10 | O24.011 | pre-existing type 1 diabetes mellitus, in pregnancy, first trimester | 12 |
| DX_ICD10 | O24.111 | pre-existing type 2 diabetes mellitus, in pregnancy, first trimester | 12 |
| DX_ICD10 | O24.311 | unspecified pre-existing diabetes mellitus in pregnancy, first trimester | 12 |
| DX_ICD10 | O24.811 | other pre-existing diabetes mellitus in pregnancy, first trimester | 12 |
| DX_ICD10 | O24.911 | unspecified diabetes mellitus in pregnancy, first trimester | 12 |
| DX_ICD10 | O25.11 | malnutrition in pregnancy, first trimester | 12 |
| DX_ICD10 | O26.01 | excessive weight gain in pregnancy, first trimester | 12 |
| DX_ICD10 | O26.11 | low weight gain in pregnancy, first trimester | 12 |
| DX_ICD10 | O26.21 | pregnancy care for patient with recurrent pregnancy loss, first trimester | 12 |
| DX_ICD10 | O26.31 | retained intrauterine contraceptive device in pregnancy, first trimester | 12 |
| DX_ICD10 | O26.41 | herpes gestationis, first trimester | 12 |
| DX_ICD10 | O26.51 | maternal hypotension syndrome, first trimester | 12 |
| DX_ICD10 | O26.611 | liver and biliary tract disorders in pregnancy, first trimester | 12 |
| DX_ICD10 | O26.711 | subluxation of symphysis (pubis) in pregnancy, first trimester | 12 |
| DX_ICD10 | O26.811 | pregnancy related exhaustion and fatigue, first trimester | 12 |
| DX_ICD10 | O26.821 | pregnancy related peripheral neuritis, first trimester | 12 |
| DX_ICD10 | O26.831 | pregnancy related renal disease, first trimester | 12 |
| DX_ICD10 | O26.841 | uterine size-date discrepancy, first trimester | 12 |
| DX_ICD10 | O26.851 | spotting complicating pregnancy, first trimester | 12 |
| DX_ICD10 | O26.891 | other specified pregnancy related conditions, first trimester | 12 |
| DX_ICD10 | O26.91 | pregnancy related conditions, unspecified, first trimester | 12 |
| DX_ICD10 | O29.011 | aspiration pneumonitis due to anesthesia during pregnancy, first trimester | 12 |
| DX_ICD10 | O29.021 | pressure collapse of lung due to anesthesia during pregnancy, first trimester | 12 |
| DX_ICD10 | O29.091 | other pulmonary complications of anesthesia during pregnancy, first trimester | 12 |
| DX_ICD10 | O29.111 | cardiac arrest due to anesthesia during pregnancy, first trimester | 12 |
| DX_ICD10 | O29.121 | cardiac failure due to anesthesia during pregnancy, first trimester | 12 |
| DX_ICD10 | O29.191 | other cardiac complications of anesthesia during pregnancy, first trimester | 12 |
| DX_ICD10 | O29.211 | cerebral anoxia due to anesthesia during pregnancy, first trimester | 12 |
| DX_ICD10 | O29.291 | other CNS complications of anesthesia during pregnancy, first trimester | 12 |
| DX_ICD10 | O29.3X1 | toxic reaction to local anesthesia during pregnancy, first trimester | 12 |
| DX_ICD10 | O29.41 | spinal & epidural anesthesia induced headache during preg, first trimester | 12 |
| DX_ICD10 | O29.5X1 | other complications of spinal and epidural anesthesia during preg, first tri | 12 |
| DX_ICD10 | O29.61 | failed or difficult intubation for anesthesia during pregnancy, first trimester | 12 |
| DX_ICD10 | O29.8X1 | other complications of anesthesia during pregnancy, first trimester | 12 |
| DX_ICD10 | O29.91 | unspecified complication of anesthesia during pregnancy, first trimester | 12 |
| DX_ICD10 | O30.001 | twin preg, unspec # placenta & unspec # amniotic sacs, first trimester | 12 |
| DX_ICD10 | O30.011 | twin pregnancy, monochorionic/monoamniotic, first trimester | 12 |
| DX_ICD10 | O30.021 | conjoined twin pregnancy, first trimester | 12 |
| DX_ICD10 | O30.031 | twin pregnancy, monochorionic/diamniotic, first trimester | 12 |
| DX_ICD10 | O30.041 | twin pregnancy, dichorionic/diamniotic, first trimester | 12 |
| DX_ICD10 | O30.091 | twin preg, unable to determine # placenta & # amniotic sacs, first trimester | 12 |
| DX_ICD10 | O30.101 | triplet preg, unspec # placenta and unspec # amniotic sacs, first trimester | 12 |
| DX_ICD10 | O30.111 | triplet pregnancy with two or more monochorionic fetuses, first trimester | 12 |
| DX_ICD10 | O30.121 | triplet pregnancy with two or more monoamniotic fetuses, first trimester | 12 |
| DX_ICD10 | O30.131 | triplet pregnancy, trichorionic/triamniotic, first trimester | 12 |
| DX_ICD10 | O30.191 | triplet preg, unable to determine # placenta & # amniotic sacs, first trimester | 12 |
| DX_ICD10 | O30.201 | quadruplet preg, unspec # placenta & unspec # amniotic sacs, first trimester | 12 |
| DX_ICD10 | O30.211 | quadruplet pregnancy with two or more monochorionic fetuses, first trimester | 12 |
| DX_ICD10 | O30.221 | quadruplet pregnancy with two or more monoamniotic fetuses, first trimester | 12 |
| DX_ICD10 | O30.231 | quadruplet pregnancy, quadrachorionic/quadra-amniotic, first trimester | 12 |
| DX_ICD10 | O30.291 | quadruplet preg, unable to determine # placenta & # amniotic sacs, first tri | 12 |
| DX_ICD10 | O30.801 | oth spec multi gestation, unspec # placenta & unspec #amniotic sacs, first tri | 12 |
| DX_ICD10 | O30.811 | other spec multi gestation w 2+ monochorionic fetuses, first trimester | 12 |
| DX_ICD10 | O30.821 | other spec multi gestation w 2+ monoamniotic fetuses, first trimester | 12 |
| DX_ICD10 | O30.831 | other spec multi gestation, # chorions & amnions both equal to # fetuses, first tri | 12 |
| DX_ICD10 | O30.891 | oth spec multi gestation, unable to determine # placenta & # amniotic sacs, first tri | 12 |
| DX_ICD10 | O30.91 | multiple gestation, unspecified, first trimester | 12 |
| DX_ICD10 | O31.01X0 | papyraceous fetus, first trimester, not applicable or unspecified | 12 |
| DX_ICD10 | O31.01X1 | papyraceous fetus, first trimester, fetus 1 | 12 |
| DX_ICD10 | O31.01X2 | papyraceous fetus, first trimester, fetus 2 | 12 |
| DX_ICD10 | O31.01X3 | papyraceous fetus, first trimester, fetus 3 | 12 |
| DX_ICD10 | O31.01X4 | papyraceous fetus, first trimester, fetus 4 | 12 |
| DX_ICD10 | O31.01X5 | papyraceous fetus, first trimester, fetus 5 | 12 |
| DX_ICD10 | O31.01X9 | papyraceous fetus, first trimester, other fetus | 12 |
| DX_ICD10 | O31.11X1 | continuing preg after spontan abortion of one fetus or more, first trimester, fetus 1 | 12 |
| DX_ICD10 | O31.11X2 | continuing preg after spontan abortion of one fetus or more, first trimester, fetus 2 | 12 |
| DX_ICD10 | O31.11X3 | continuing preg after spontan abortion of one fetus or more, first trimester, fetus 3 | 12 |
| DX_ICD10 | O31.11X4 | continuing preg after spontan abortion of one fetus or more, first trimester, fetus 4 | 12 |
| DX_ICD10 | O31.11X5 | continuing preg after spontan abortion of one fetus or more, first trimester, fetus 5 | 12 |
| DX_ICD10 | O31.11X9 | continuing preg after spontan abortion of one fetus or more, first trimester, oth fetus | 12 |
| DX_ICD10 | O31.21X0 | continuing preg after intrauterine death of one fetus+, first trimester, n/a nspecified | 12 |
| DX_ICD10 | O31.21X1 | continuing preg after intrauterine death of one fetus or more, first trimester, fetus 1 | 12 |
| DX_ICD10 | O31.21X2 | continuing preg after intrauterine death of one fetus or more, first trimester, fetus 2 | 12 |
| DX_ICD10 | O31.21X3 | continuing preg after intrauterine death of one fetus or more, first trimester, fetus 3 | 12 |
| DX_ICD10 | O31.21X4 | continuing preg after intrauterine death of one fetus or more, first trimester, fetus 4 | 12 |
| DX_ICD10 | O31.21X5 | continuing preg after intrauterine death of one fetus or more, first trimester, fetus 5 | 12 |
| DX_ICD10 | O31.21X9 | continuing preg after intrauterine death of one fetus+, first trimester, oth fetus | 12 |
| DX_ICD10 | O31.31X0 | continuing preg after elective fetal reduct 1 fetus+, first trimester, n/a or unspec | 12 |
| DX_ICD10 | O31.31X1 | continuing preg after elective fetal reduct 1 fetus+, first trimester, fetus 1 | 12 |
| DX_ICD10 | O31.31X2 | continuing preg after elective fetal reduct of 1 fetus or more, first trimester, fetus 2 | 12 |
| DX_ICD10 | O31.31X3 | continuing preg after elective fetal reduct of 1 fetus or more, first trimester, fetus 3 | 12 |
| DX_ICD10 | O31.31X4 | continuing preg after elective fetal reduct of 1 fetus or more, first trimester, fetus 4 | 12 |
| DX_ICD10 | O31.31X5 | continuing preg after elective fetal reduct of 1 fetus or more, first trimester, fetus 5 | 12 |
| DX_ICD10 | O31.31X9 | continuing preg after elective fetal reduct of 1 fetus or more, first tri, oth fetus | 12 |
| DX_ICD10 | O31.8X10 | oth complic spec to multi gestation, first trimester, not applicable or unspec | 12 |
| DX_ICD10 | O31.8X11 | other complications specific to multiple gestation, first trimester, fetus 1 | 12 |
| DX_ICD10 | O31.8X12 | other complications specific to multiple gestation, first trimester, fetus 2 | 12 |
| DX_ICD10 | O31.8X13 | other complications specific to multiple gestation, first trimester, fetus 3 | 12 |
| DX_ICD10 | O31.8X14 | other complications specific to multiple gestation, first trimester, fetus 4 | 12 |
| DX_ICD10 | O31.8X15 | other complications specific to multiple gestation, first trimester, fetus 5 | 12 |
| DX_ICD10 | O31.8X19 | other complications specific to multiple gestation, first trimester, other fetus | 12 |
| DX_ICD10 | O34.01 | maternal care for unspecified congenital malformation of uterus, first trimester | 12 |
| DX_ICD10 | O34.11 | maternal care for benign tumor of corpus uteri, first trimester | 12 |
| DX_ICD10 | O34.31 | maternal care for cervical incompetence, first trimester | 12 |
| DX_ICD10 | O34.41 | maternal care for other abnormalities of cervix, first trimester | 12 |
| DX_ICD10 | O34.511 | maternal care for incarceration of gravid uterus, first trimester | 12 |
| DX_ICD10 | O34.521 | maternal care for prolapse of gravid uterus, first trimester | 12 |
| DX_ICD10 | O34.531 | maternal care for retroversion of gravid uterus, first trimester | 12 |
| DX_ICD10 | O34.591 | maternal care for other abnormalities of gravid uterus, first trimester | 12 |
| DX_ICD10 | O34.61 | maternal care for abnormality of vagina, first trimester | 12 |
| DX_ICD10 | O34.71 | maternal care for abnormality of vulva and perineum, first trimester | 12 |
| DX_ICD10 | O34.81 | maternal care for other abnormalities of pelvic organs, first trimester | 12 |
| DX_ICD10 | O34.91 | maternal care for abnormality of pelvic organ, unspecified, first trimester | 12 |
| DX_ICD10 | O36.0110 | maternal care for anti-d [rh] antibodies, first trimester, not applicable or unspecified | 12 |
| DX_ICD10 | O36.0111 | maternal care for anti-d [rh] antibodies, first trimester, fetus 1 | 12 |
| DX_ICD10 | O36.0112 | maternal care for anti-d [rh] antibodies, first trimester, fetus 2 | 12 |
| DX_ICD10 | O36.0113 | maternal care for anti-d [rh] antibodies, first trimester, fetus 3 | 12 |
| DX_ICD10 | O36.0114 | maternal care for anti-d [rh] antibodies, first trimester, fetus 4 | 12 |
| DX_ICD10 | O36.0115 | maternal care for anti-d [rh] antibodies, first trimester, fetus 5 | 12 |
| DX_ICD10 | O36.0119 | maternal care for anti-d [rh] antibodies, first trimester, other fetus | 12 |
| DX_ICD10 | O36.0910 | maternal care for oth rhesus isoimmunization, first tri, not applicable or unspec | 12 |
| DX_ICD10 | O36.0911 | maternal care for other rhesus isoimmunization, first trimester, fetus 1 | 12 |
| DX_ICD10 | O36.0912 | maternal care for other rhesus isoimmunization, first trimester, fetus 2 | 12 |
| DX_ICD10 | O36.0913 | maternal care for other rhesus isoimmunization, first trimester, fetus 3 | 12 |
| DX_ICD10 | O36.0914 | maternal care for other rhesus isoimmunization, first trimester, fetus 4 | 12 |
| DX_ICD10 | O36.0915 | maternal care for other rhesus isoimmunization, first trimester, fetus 5 | 12 |
| DX_ICD10 | O36.0919 | maternal care for other rhesus isoimmunization, first trimester, other fetus | 12 |
| DX_ICD10 | O36.1110 | maternal care for anti-a sensitization, first trimester, not applicable or unspecified | 12 |
| DX_ICD10 | O36.1111 | maternal care for anti-a sensitization, first trimester, fetus 1 | 12 |
| DX_ICD10 | O36.1112 | maternal care for anti-a sensitization, first trimester, fetus 2 | 12 |
| DX_ICD10 | O36.1113 | maternal care for anti-a sensitization, first trimester, fetus 3 | 12 |
| DX_ICD10 | O36.1114 | maternal care for anti-a sensitization, first trimester, fetus 4 | 12 |
| DX_ICD10 | O36.1115 | maternal care for anti-a sensitization, first trimester, fetus 5 | 12 |
| DX_ICD10 | O36.1119 | maternal care for anti-a sensitization, first trimester, other fetus | 12 |
| DX_ICD10 | O36.1910 | maternal care for other isoimmunization, first trimester, not applicable or unspec | 12 |
| DX_ICD10 | O36.1911 | maternal care for other isoimmunization, first trimester, fetus 1 | 12 |
| DX_ICD10 | O36.1912 | maternal care for other isoimmunization, first trimester, fetus 2 | 12 |
| DX_ICD10 | O36.1913 | maternal care for other isoimmunization, first trimester, fetus 3 | 12 |
| DX_ICD10 | O36.1914 | maternal care for other isoimmunization, first trimester, fetus 4 | 12 |
| DX_ICD10 | O36.1915 | maternal care for other isoimmunization, first trimester, fetus 5 | 12 |
| DX_ICD10 | O36.1919 | maternal care for other isoimmunization, first trimester, other fetus | 12 |
| DX_ICD10 | O36.21X0 | maternal care for hydrops fetalis, first trimester, not applicable or unspecified | 12 |
| DX_ICD10 | O36.21X1 | maternal care for hydrops fetalis, first trimester, fetus 1 | 12 |
| DX_ICD10 | O36.21X2 | maternal care for hydrops fetalis, first trimester, fetus 2 | 12 |
| DX_ICD10 | O36.21X3 | maternal care for hydrops fetalis, first trimester, fetus 3 | 12 |
| DX_ICD10 | O36.21X4 | maternal care for hydrops fetalis, first trimester, fetus 4 | 12 |
| DX_ICD10 | O36.21X5 | maternal care for hydrops fetalis, first trimester, fetus 5 | 12 |
| DX_ICD10 | O36.21X9 | maternal care for hydrops fetalis, first trimester, other fetus | 12 |
| DX_ICD10 | O36.5110 | maternal care for known or suspected placental insufficiency, first tri, n/a or unspec | 12 |
| DX_ICD10 | O36.5111 | maternal care for known or suspected placental insufficiency, first tri, fetus 1 | 12 |
| DX_ICD10 | O36.5112 | maternal care for known or suspected placental insufficiency, first tri, fetus 2 | 12 |
| DX_ICD10 | O36.5113 | maternal care for known or suspected placental insufficiency, first tri, fetus 3 | 12 |
| DX_ICD10 | O36.5114 | maternal care for known or suspected placental insufficiency, first tri, fetus 4 | 12 |
| DX_ICD10 | O36.5115 | maternal care for known or suspected placental insufficiency, first tri, fetus 5 | 12 |
| DX_ICD10 | O36.5119 | maternal care for known or suspected placental insufficiency, first tri, other fetus | 12 |
| DX_ICD10 | O36.5910 | maternal care for other known or suspected poor fetal growth, first tri, n/a or unspec | 12 |
| DX_ICD10 | O36.5911 | maternal care for other known or suspected poor fetal growth, first tri, fetus 1 | 12 |
| DX_ICD10 | O36.5912 | maternal care for other known or suspected poor fetal growth, first tri, fetus 2 | 12 |
| DX_ICD10 | O36.5913 | maternal care for other known or suspected poor fetal growth, first tri, fetus 3 | 12 |
| DX_ICD10 | O36.5914 | maternal care for other known or suspected poor fetal growth, first tri, fetus 4 | 12 |
| DX_ICD10 | O36.5915 | maternal care for other known or suspected poor fetal growth, first tri, fetus 5 | 12 |
| DX_ICD10 | O36.5919 | maternal care for other known or suspected poor fetal growth, first tri, other fetus | 12 |
| DX_ICD10 | O36.61X0 | maternal care for excessive fetal growth, first tri, n/a or unspec | 12 |
| DX_ICD10 | O36.61X1 | maternal care for excessive fetal growth, first trimester, fetus 1 | 12 |
| DX_ICD10 | O36.61X2 | maternal care for excessive fetal growth, first trimester, fetus 2 | 12 |
| DX_ICD10 | O36.61X3 | maternal care for excessive fetal growth, first trimester, fetus 3 | 12 |
| DX_ICD10 | O36.61X4 | maternal care for excessive fetal growth, first trimester, fetus 4 | 12 |
| DX_ICD10 | O36.61X5 | maternal care for excessive fetal growth, first trimester, fetus 5 | 12 |
| DX_ICD10 | O36.61X9 | maternal care for excessive fetal growth, first trimester, other fetus | 12 |
| DX_ICD10 | O36.71X0 | maternal care for viable fetus in abdominal preg, first tri, n/a or unspec | 12 |
| DX_ICD10 | O36.71X1 | maternal care for viable fetus in abdominal pregnancy, first trimester, fetus 1 | 12 |
| DX_ICD10 | O36.71X2 | maternal care for viable fetus in abdominal pregnancy, first trimester, fetus 2 | 12 |
| DX_ICD10 | O36.71X3 | maternal care for viable fetus in abdominal pregnancy, first trimester, fetus 3 | 12 |
| DX_ICD10 | O36.71X4 | maternal care for viable fetus in abdominal pregnancy, first trimester, fetus 4 | 12 |
| DX_ICD10 | O36.71X5 | maternal care for viable fetus in abdominal pregnancy, first trimester, fetus 5 | 12 |
| DX_ICD10 | O36.71X9 | maternal care for viable fetus in abdominal pregnancy, first trimester, other fetus | 12 |
| DX_ICD10 | O36.8210 | fetal anemia and thrombocytopenia, first trimester, not applicable or unspecified | 12 |
| DX_ICD10 | O36.8211 | fetal anemia and thrombocytopenia, first trimester, fetus 1 | 12 |
| DX_ICD10 | O36.8212 | fetal anemia and thrombocytopenia, first trimester, fetus 2 | 12 |
| DX_ICD10 | O36.8213 | fetal anemia and thrombocytopenia, first trimester, fetus 3 | 12 |
| DX_ICD10 | O36.8214 | fetal anemia and thrombocytopenia, first trimester, fetus 4 | 12 |
| DX_ICD10 | O36.8215 | fetal anemia and thrombocytopenia, first trimester, fetus 5 | 12 |
| DX_ICD10 | O36.8219 | fetal anemia and thrombocytopenia, first trimester, other fetus | 12 |
| DX_ICD10 | O36.8310 | maternal care for abnormalities of fetal heart rate or rhythm, first tri, n/a or unspec | 12 |
| DX_ICD10 | O36.8311 | maternal care for abnormalities of fetal heart rate or rhythm, first trimester, fetus 1 | 12 |
| DX_ICD10 | O36.8312 | maternal care for abnormalities of fetal heart rate or rhythm, first trimester, fetus 2 | 12 |
| DX_ICD10 | O36.8313 | maternal care for abnormalities of fetal heart rate or rhythm, first trimester, fetus 3 | 12 |
| DX_ICD10 | O36.8314 | maternal care for abnormalities of fetal heart rate or rhythm, first trimester, fetus 4 | 12 |
| DX_ICD10 | O36.8315 | maternal care for abnormalities of fetal heart rate or rhythm, first trimester, fetus 5 | 12 |
| DX_ICD10 | O36.8319 | maternal care for abnormalities of fetal heart rate or rhythm, first tri, other fetus | 12 |
| DX_ICD10 | O36.8910 | maternal care for other specified fetal problems, first tri, n/a or unspec | 12 |
| DX_ICD10 | O36.8911 | maternal care for other specified fetal problems, first trimester, fetus 1 | 12 |
| DX_ICD10 | O36.8912 | maternal care for other specified fetal problems, first trimester, fetus 2 | 12 |
| DX_ICD10 | O36.8913 | maternal care for other specified fetal problems, first trimester, fetus 3 | 12 |
| DX_ICD10 | O36.8914 | maternal care for other specified fetal problems, first trimester, fetus 4 | 12 |
| DX_ICD10 | O36.8915 | maternal care for other specified fetal problems, first trimester, fetus 5 | 12 |
| DX_ICD10 | O36.8919 | maternal care for other specified fetal problems, first trimester, other fetus | 12 |
| DX_ICD10 | O36.91X0 | maternal care for fetal problem, unspec, first tri, not applicable or unspecified | 12 |
| DX_ICD10 | O36.91X1 | maternal care for fetal problem, unspecified, first trimester, fetus 1 | 12 |
| DX_ICD10 | O36.91X2 | maternal care for fetal problem, unspecified, first trimester, fetus 2 | 12 |
| DX_ICD10 | O36.91X3 | maternal care for fetal problem, unspecified, first trimester, fetus 3 | 12 |
| DX_ICD10 | O36.91X4 | maternal care for fetal problem, unspecified, first trimester, fetus 4 | 12 |
| DX_ICD10 | O36.91X5 | maternal care for fetal problem, unspecified, first trimester, fetus 5 | 12 |
| DX_ICD10 | O36.91X9 | maternal care for fetal problem, unspecified, first trimester, other fetus | 12 |
| DX_ICD10 | O40.1XX0 | polyhydramnios, first trimester, not applicable or unspecified | 12 |
| DX_ICD10 | O40.1XX1 | polyhydramnios, first trimester, fetus 1 | 12 |
| DX_ICD10 | O40.1XX2 | polyhydramnios, first trimester, fetus 2 | 12 |
| DX_ICD10 | O40.1XX3 | polyhydramnios, first trimester, fetus 3 | 12 |
| DX_ICD10 | O40.1XX4 | polyhydramnios, first trimester, fetus 4 | 12 |
| DX_ICD10 | O40.1XX5 | polyhydramnios, first trimester, fetus 5 | 12 |
| DX_ICD10 | O40.1XX9 | polyhydramnios, first trimester, other fetus | 12 |
| DX_ICD10 | O41.01X0 | oligohydramnios, first trimester, not applicable or unspecified | 12 |
| DX_ICD10 | O41.01X1 | oligohydramnios, first trimester, fetus 1 | 12 |
| DX_ICD10 | O41.01X2 | oligohydramnios, first trimester, fetus 2 | 12 |
| DX_ICD10 | O41.01X3 | oligohydramnios, first trimester, fetus 3 | 12 |
| DX_ICD10 | O41.01X4 | oligohydramnios, first trimester, fetus 4 | 12 |
| DX_ICD10 | O41.01X5 | oligohydramnios, first trimester, fetus 5 | 12 |
| DX_ICD10 | O41.01X9 | oligohydramnios, first trimester, other fetus | 12 |
| DX_ICD10 | O41.1010 | infection of amniotic sac & membranes, unspec, first trimester, n/a or unspec | 12 |
| DX_ICD10 | O41.1011 | infection of amniotic sac and membranes, unspecified, first trimester, fetus 1 | 12 |
| DX_ICD10 | O41.1012 | infection of amniotic sac and membranes, unspecified, first trimester, fetus 2 | 12 |
| DX_ICD10 | O41.1013 | infection of amniotic sac and membranes, unspecified, first trimester, fetus 3 | 12 |
| DX_ICD10 | O41.1014 | infection of amniotic sac and membranes, unspecified, first trimester, fetus 4 | 12 |
| DX_ICD10 | O41.1015 | infection of amniotic sac and membranes, unspecified, first trimester, fetus 5 | 12 |
| DX_ICD10 | O41.1019 | infection of amniotic sac and membranes, unspecified, first trimester, other fetus | 12 |
| DX_ICD10 | O41.1210 | chorioamnionitis, first trimester, not applicable or unspecified | 12 |
| DX_ICD10 | O41.1211 | chorioamnionitis, first trimester, fetus 1 | 12 |
| DX_ICD10 | O41.1212 | chorioamnionitis, first trimester, fetus 2 | 12 |
| DX_ICD10 | O41.1213 | chorioamnionitis, first trimester, fetus 3 | 12 |
| DX_ICD10 | O41.1214 | chorioamnionitis, first trimester, fetus 4 | 12 |
| DX_ICD10 | O41.1215 | chorioamnionitis, first trimester, fetus 5 | 12 |
| DX_ICD10 | O41.1219 | chorioamnionitis, first trimester, other fetus | 12 |
| DX_ICD10 | O41.1410 | placentitis, first trimester, not applicable or unspecified | 12 |
| DX_ICD10 | O41.1411 | placentitis, first trimester, fetus 1 | 12 |
| DX_ICD10 | O41.1412 | placentitis, first trimester, fetus 2 | 12 |
| DX_ICD10 | O41.1413 | placentitis, first trimester, fetus 3 | 12 |
| DX_ICD10 | O41.1414 | placentitis, first trimester, fetus 4 | 12 |
| DX_ICD10 | O41.1415 | placentitis, first trimester, fetus 5 | 12 |
| DX_ICD10 | O41.1419 | placentitis, first trimester, other fetus | 12 |
| DX_ICD10 | O41.8X10 | oth spec disorders of amniotic fluid and membranes, first trimester, n/a or unspec | 12 |
| DX_ICD10 | O41.8X11 | other specified disorders of amniotic fluid and membranes, first trimester, fetus 1 | 12 |
| DX_ICD10 | O41.8X12 | other specified disorders of amniotic fluid and membranes, first trimester, fetus 2 | 12 |
| DX_ICD10 | O41.8X13 | other specified disorders of amniotic fluid and membranes, first trimester, fetus 3 | 12 |
| DX_ICD10 | O41.8X14 | other specified disorders of amniotic fluid and membranes, first trimester, fetus 4 | 12 |
| DX_ICD10 | O41.8X15 | other specified disorders of amniotic fluid and membranes, first trimester, fetus 5 | 12 |
| DX_ICD10 | O41.8X19 | oth specified disorders of amniotic fluid and membranes, first trimester, other fetus | 12 |
| DX_ICD10 | O41.91X0 | disorder of amniotic fluid and membranes, unspecified, first tri, n/a or unspec | 12 |
| DX_ICD10 | O41.91X1 | disorder of amniotic fluid and membranes, unspecified, first trimester, fetus 1 | 12 |
| DX_ICD10 | O41.91X2 | disorder of amniotic fluid and membranes, unspecified, first trimester, fetus 2 | 12 |
| DX_ICD10 | O41.91X3 | disorder of amniotic fluid and membranes, unspecified, first trimester, fetus 3 | 12 |
| DX_ICD10 | O41.91X4 | disorder of amniotic fluid and membranes, unspecified, first trimester, fetus 4 | 12 |
| DX_ICD10 | O41.91X5 | disorder of amniotic fluid and membranes, unspecified, first trimester, fetus 5 | 12 |
| DX_ICD10 | O41.91X9 | disorder of amniotic fluid and membranes, unspecified, first trimester, other fetus | 12 |
| DX_ICD10 | O42.011 | preterm premature rupture membranes, onset labor w in 24 hours of rupture, first tri | 12 |
| DX_ICD10 | O42.111 | preterm premature rupture membranes, onset labor > than 24 hrs after rupture, 1 tri | 12 |
| DX_ICD10 | O42.911 | preterm premat rupt membranes, unspec length of time btwn rupture & labor, 1^st^ tri | 12 |
| DX_ICD10 | O43.011 | fetomaternal placental transfusion syndrome, first trimester | 12 |
| DX_ICD10 | O43.021 | fetus-to-fetus placental transfusion syndrome, first trimester | 12 |
| DX_ICD10 | O43.101 | malformation of placenta, unspecified, first trimester | 12 |
| DX_ICD10 | O43.111 | circumvallate placenta, first trimester | 12 |
| DX_ICD10 | O43.121 | velamentous insertion of umbilical cord, first trimester | 12 |
| DX_ICD10 | O43.191 | other malformation of placenta, first trimester | 12 |
| DX_ICD10 | O43.211 | placenta accreta, first trimester | 12 |
| DX_ICD10 | O43.221 | placenta increta, first trimester | 12 |
| DX_ICD10 | O43.231 | placenta percreta, first trimester | 12 |
| DX_ICD10 | O43.811 | placental infarction, first trimester | 12 |
| DX_ICD10 | O43.891 | other placental disorders, first trimester | 12 |
| DX_ICD10 | O43.91 | unspecified placental disorder, first trimester | 12 |
| DX_ICD10 | O44.01 | complete placenta previa nos or without hemorrhage, first trimester | 12 |
| DX_ICD10 | O44.11 | complete placenta previa with hemorrhage, first trimester | 12 |
| DX_ICD10 | O44.21 | partial placenta previa nos or without hemorrhage, first trimester | 12 |
| DX_ICD10 | O44.31 | partial placenta previa with hemorrhage, first trimester | 12 |
| DX_ICD10 | O44.41 | low lying placenta nos or without hemorrhage, first trimester | 12 |
| DX_ICD10 | O44.51 | low lying placenta with hemorrhage, first trimester | 12 |
| DX_ICD10 | O45.001 | premature separation of placenta with coagulation defect, unspecified, first trimester | 12 |
| DX_ICD10 | O45.011 | premature separation of placenta w afibrinogenemia, first trimester | 12 |
| DX_ICD10 | O45.021 | premature separation of placenta w disseminated intravascular coagulation, first tri | 12 |
| DX_ICD10 | O45.091 | premature separation of placenta w other coagulation defect, first trimester | 12 |
| DX_ICD10 | O45.8X1 | other premature separation placenta, first trimester | 12 |
| DX_ICD10 | O45.91 | premature separation of placenta, unspecified, first trimester | 12 |
| DX_ICD10 | O46.001 | antepartum hemorrhage with coagulation defect, unspecified, first trimester | 12 |
| DX_ICD10 | O46.011 | antepartum hemorrhage with afibrinogenemia, first trimester | 12 |
| DX_ICD10 | O46.021 | antepartum hemorrhage with disseminated intravascular coagulation, first trimester | 12 |
| DX_ICD10 | O46.091 | antepartum hemorrhage with other coagulation defect, first trimester | 12 |
| DX_ICD10 | O46.8X1 | other antepartum hemorrhage, first trimester | 12 |
| DX_ICD10 | O46.91 | antepartum hemorrhage, unspecified, first trimester | 12 |
| DX_ICD10 | O63.0 | prolonged first stage (of labor) | 12 |
| DX_ICD10 | O70.0 | first degree perineal laceration during delivery | 12 |
| DX_ICD10 | O88.011 | air embolism in pregnancy, first trimester | 12 |
| DX_ICD10 | O88.111 | amniotic fluid embolism in pregnancy, first trimester | 12 |
| DX_ICD10 | O88.211 | thromboembolism in pregnancy, first trimester | 12 |
| DX_ICD10 | O88.311 | pyemic and septic embolism in pregnancy, first trimester | 12 |
| DX_ICD10 | O88.811 | other embolism in pregnancy, first trimester | 12 |
| DX_ICD10 | O91.011 | infection of nipple associated with pregnancy, first trimester | 12 |
| DX_ICD10 | O91.111 | abscess of breast associated with pregnancy, first trimester | 12 |
| DX_ICD10 | O91.211 | nonpurulent mastitis associated with pregnancy, first trimester | 12 |
| DX_ICD10 | O92.011 | retracted nipple associated with pregnancy, first trimester | 12 |
| DX_ICD10 | O92.111 | cracked nipple associated with pregnancy, first trimester | 12 |
| DX_ICD10 | O98.011 | tuberculosis complicating pregnancy, first trimester | 12 |
| DX_ICD10 | O98.111 | syphilis complicating pregnancy, first trimester | 12 |
| DX_ICD10 | O98.211 | gonorrhea complicating pregnancy, first trimester | 12 |
| DX_ICD10 | O98.311 | oth infections w a predominantly sexual mode transmission complicating preg, first tri | 12 |
| DX_ICD10 | O98.411 | viral hepatitis complicating pregnancy, first trimester | 12 |
| DX_ICD10 | O98.511 | other viral diseases complicating pregnancy, first trimester | 12 |
| DX_ICD10 | O98.611 | protozoal diseases complicating pregnancy, first trimester | 12 |
| DX_ICD10 | O98.711 | human immunodeficiency virus [hiv] disease complicating pregnancy, first trimester | 12 |
| DX_ICD10 | O98.811 | other maternal infectious and parasitic diseases complicating pregnancy, first tri | 12 |
| DX_ICD10 | O98.911 | unspec maternal infectious and parasitic disease complicating pregnancy, first tri | 12 |
| DX_ICD10 | O99.011 | anemia complicating pregnancy, first trimester | 12 |
| DX_ICD10 | O99.111 | oth dis of blood & blood-form organs & certain dis immune mech complic preg, first tri | 12 |
| DX_ICD10 | O99.211 | obesity complicating pregnancy, first trimester | 12 |
| DX_ICD10 | O99.281 | endocrine, nutritional and metabolic diseases complicating pregnancy, first trimester | 12 |
| DX_ICD10 | O99.311 | alcohol use complicating pregnancy, first trimester | 12 |
| DX_ICD10 | O99.321 | drug use complicating pregnancy, first trimester | 12 |
| DX_ICD10 | O99.331 | smoking (tobacco) complicating pregnancy, first trimester | 12 |
| DX_ICD10 | O99.341 | other mental disorders complicating pregnancy, first trimester | 12 |
| DX_ICD10 | O99.351 | diseases of the nervous system complicating pregnancy, first trimester | 12 |
| DX_ICD10 | O99.411 | diseases of the circulatory system complicating pregnancy, first trimester | 12 |
| DX_ICD10 | O99.511 | diseases of the respiratory system complicating pregnancy, first trimester | 12 |
| DX_ICD10 | O99.611 | diseases of the digestive system complicating pregnancy, first trimester | 12 |
| DX_ICD10 | O99.711 | diseases of the skin and subcutaneous tissue complicating pregnancy, first trimester | 12 |
| DX_ICD10 | O99.841 | bariatric surgery status complicating pregnancy, first trimester | 12 |
| DX_ICD10 | O9A.111 | malignant neoplasm complicating pregnancy, first trimester | 12 |
| DX_ICD10 | O9A.211 | injury, poisoning & certain oth consequences of external causes complic preg, first tri | 12 |
| DX_ICD10 | O9A.311 | physical abuse complicating pregnancy, first trimester | 12 |
| DX_ICD10 | O9A.411 | sexual abuse complicating pregnancy, first trimester | 12 |
| DX_ICD10 | O9A.511 | psychological abuse complicating pregnancy, first trimester | 12 |
| DX_ICD10 | Z34.00 | encounter for supervision of normal first pregnancy, unspecified trimester | 12 |
| DX_ICD10 | Z34.01 | encounter for supervision of normal first pregnancy, first trimester | 12 |
| DX_ICD10 | Z34.81 | encounter for supervision of other normal pregnancy, first trimester | 12 |
| DX_ICD10 | Z34.91 | encounter for supervision of normal pregnancy, unspecified, first trimester | 12 |
| DX_ICD10 | Z3A.12 | 12 weeks gestation of pregnancy | 12 |
| PX_CPT | 1965 | anesthesia for incomplete or missed abortion procedures | 12 |
| PX_CPT | 59820 | treatment of missed abortion, completed surgically; first trimester | 12 |
| DX_ICD10 | Z3A.13 | 13 weeks gestation of pregnancy | 13 |
| DX_ICD10 | Z3A.14 | 14 weeks gestation of pregnancy | 14 |
| DX_ICD10 | Z3A.15 | 15 weeks gestation of pregnancy | 15 |
| DX_ICD10 | Z3A.16 | 16 weeks gestation of pregnancy | 16 |
| DX_ICD10 | Z3A.17 | 17 weeks gestation of pregnancy | 17 |
| DX_ICD10 | Z3A.18 | 18 weeks gestation of pregnancy | 18 |
| DX_ICD10 | Z3A.19 | 19 weeks gestation of pregnancy | 19 |
| DX_ICD10 | Z3A.20 | 20 weeks gestation of pregnancy | 20 |
| PX_ICD10 | 10A07ZW | abortion of products of conception, laminaria, via natural or artificial opening | 20 |
| DX_ICD10 | O30.102 | triplet pregnancy, unspec # placenta & unspec # amniotic sacs, second trimester | 21 |
| DX_ICD10 | O30.202 | quadruplet preg, unspec # placenta & unspec # amniotic sacs, second trimester | 21 |
| DX_ICD10 | Z3A.21 | 21 weeks gestation of pregnancy | 21 |
| PX_CPT | 59821 | treatment of missed abortion, completed surgically; second trimester | 21 |
| DX_ICD9 | 765.01 | extreme immaturity < 500g (begin 1988) | 22 |
| DX_ICD9 | 765.11 | preterm nec < 500g (begin 1988) | 22 |
| DX_ICD10 | P07.01 | extremely low birth weight newborn, less than 500 grams | 22 |
| DX_ICD10 | P07.01 | extremely low birth weight newborn, less than 500 grams | 22 |
| DX_ICD10 | P07.21 | extreme immaturity of newborn, gestational age less than 23 completed weeks | 22 |
| DX_ICD10 | Z3A.22 | 22 weeks gestation of pregnancy | 22 |
| DX_ICD9 | 765.21 | less than 24 completed weeks of gestation (begin 2002) | 23 |
| DX_ICD10 | O09.02 | supervision of pregnancy with history of infertility, second trimester | 23 |
| DX_ICD10 | O09.12 | supervision of pregnancy with history of ectopic pregnancy, second trimester | 23 |
| DX_ICD10 | O09.212 | supervision of pregnancy with history of pre-term labor, second trimester | 23 |
| DX_ICD10 | O09.292 | supervision of pregnancy w oth poor reproductive or obstetric history, second trimester | 23 |
| DX_ICD10 | O09.32 | supervision of pregnancy with insufficient antenatal care, second trimester | 23 |
| DX_ICD10 | O09.42 | supervision of pregnancy with grand multiparity, second trimester | 23 |
| DX_ICD10 | O09.512 | supervision of elderly primigravida, second trimester | 23 |
| DX_ICD10 | O09.522 | supervision of elderly multigravida, second trimester | 23 |
| DX_ICD10 | O09.612 | supervision of young primigravida, second trimester | 23 |
| DX_ICD10 | O09.622 | supervision of young multigravida, second trimester | 23 |
| DX_ICD10 | O09.72 | supervision of high risk pregnancy due to social problems, second trimester | 23 |
| DX_ICD10 | O09.812 | supervision of pregnancy resulting from assisted reproductive technology, second tri | 23 |
| DX_ICD10 | O09.822 | supervision of preg w history of in utero procedure during previous pregnancy, second tri | 23 |
| DX_ICD10 | O09.892 | supervision of other high risk pregnancies, second trimester | 23 |
| DX_ICD10 | O09.92 | supervision of high risk pregnancy, unspecified, second trimester | 23 |
| DX_ICD10 | O09.A2 | supervision of pregnancy with history of molar pregnancy, second trimester | 23 |
| DX_ICD10 | O10.012 | pre-existing essential hypertension complicating pregnancy, second trimester | 23 |
| DX_ICD10 | O10.112 | pre-existing hypertensive heart disease complicating pregnancy, second trimester | 23 |
| DX_ICD10 | O10.212 | pre-existing hypertensive chronic kidney disease complicating preg, second trimester | 23 |
| DX_ICD10 | O10.312 | pre-existing hypertensive heart & chronic kidney disease complicating preg, second tri | 23 |
| DX_ICD10 | O10.411 | pre-existing secondary hypertension complicating pregnancy, first trimester | 23 |
| DX_ICD10 | O10.412 | pre-existing secondary hypertension complicating pregnancy, second trimester | 23 |
| DX_ICD10 | O10.419 | pre-existing secondary hypertension complicating pregnancy, unspecified trimester | 23 |
| DX_ICD10 | O10.42 | pre-existing secondary hypertension complicating childbirth | 23 |
| DX_ICD10 | O10.43 | pre-existing secondary hypertension complicating the puerperium | 23 |
| DX_ICD10 | O10.912 | unspecified pre-existing hypertension complicating pregnancy, second trimester | 23 |
| DX_ICD10 | O11.2 | pre-existing hypertension with pre-eclampsia, second trimester | 23 |
| DX_ICD10 | O11.2 | pre-existing hypertension with pre-eclampsia, second trimester | 23 |
| DX_ICD10 | O12.02 | gestational edema, second trimester | 23 |
| DX_ICD10 | O12.12 | gestational proteinuria, second trimester | 23 |
| DX_ICD10 | O12.22 | gestational edema with proteinuria, second trimester | 23 |
| DX_ICD10 | O13.2 | gestational [pregnancy-induced] hypertension without significant proteinuria, second tri | 23 |
| DX_ICD10 | O14.02 | mild to moderate pre-eclampsia, second trimester | 23 |
| DX_ICD10 | O14.12 | severe pre-eclampsia, second trimester | 23 |
| DX_ICD10 | O14.22 | hellp syndrome (hellp), second trimester | 23 |
| DX_ICD10 | O14.92 | unspecified pre-eclampsia, second trimester | 23 |
| DX_ICD10 | O15.02 | eclampsia complicating pregnancy, second trimester | 23 |
| DX_ICD10 | O16.2 | unspecified maternal hypertension, second trimester | 23 |
| DX_ICD10 | O22.02 | varicose veins of lower extremity in pregnancy, second trimester | 23 |
| DX_ICD10 | O22.12 | genital varices in pregnancy, second trimester | 23 |
| DX_ICD10 | O22.22 | superficial thrombophlebitis in pregnancy, second trimester | 23 |
| DX_ICD10 | O22.32 | deep phlebothrombosis in pregnancy, second trimester | 23 |
| DX_ICD10 | O22.42 | cerebral venous thrombosis in pregnancy, second trimester | 23 |
| DX_ICD10 | O22.52 | cerebral venous thrombosis in pregnancy, second trimester | 23 |
| DX_ICD10 | O22.8X2 | other venous complications in pregnancy, second trimester | 23 |
| DX_ICD10 | O22.92 | venous complication in pregnancy, unspecified, second trimester | 23 |
| DX_ICD10 | O23.02 | infections of kidney in pregnancy, second trimester | 23 |
| DX_ICD10 | O23.12 | infections of bladder in pregnancy, second trimester | 23 |
| DX_ICD10 | O23.22 | infections of urethra in pregnancy, second trimester | 23 |
| DX_ICD10 | O23.32 | infections of other parts of urinary tract in pregnancy, second trimester | 23 |
| DX_ICD10 | O23.42 | unspecified infection of urinary tract in pregnancy, second trimester | 23 |
| DX_ICD10 | O23.512 | infections of cervix in pregnancy, second trimester | 23 |
| DX_ICD10 | O23.522 | salpingo-oophoritis in pregnancy, second trimester | 23 |
| DX_ICD10 | O23.592 | infection of other part of genital tract in pregnancy, second trimester | 23 |
| DX_ICD10 | O23.92 | unspecified genitourinary tract infection in pregnancy, second trimester | 23 |
| DX_ICD10 | O24.012 | pre-existing type 1 diabetes mellitus, in pregnancy, second trimester | 23 |
| DX_ICD10 | O24.112 | pre-existing type 2 diabetes mellitus, in pregnancy, second trimester | 23 |
| DX_ICD10 | O24.312 | unspecified pre-existing diabetes mellitus in pregnancy, second trimester | 23 |
| DX_ICD10 | O24.812 | other pre-existing diabetes mellitus in pregnancy, second trimester | 23 |
| DX_ICD10 | O24.912 | unspecified diabetes mellitus in pregnancy, second trimester | 23 |
| DX_ICD10 | O25.12 | malnutrition in pregnancy, second trimester | 23 |
| DX_ICD10 | O26.02 | excessive weight gain in pregnancy, second trimester | 23 |
| DX_ICD10 | O26.12 | low weight gain in pregnancy, second trimester | 23 |
| DX_ICD10 | O26.22 | pregnancy care for patient with recurrent pregnancy loss, second trimester | 23 |
| DX_ICD10 | O26.32 | retained intrauterine contraceptive device in pregnancy, second trimester | 23 |
| DX_ICD10 | O26.42 | herpes gestationis, second trimester | 23 |
| DX_ICD10 | O26.52 | maternal hypotension syndrome, second trimester | 23 |
| DX_ICD10 | O26.612 | liver and biliary tract disorders in pregnancy, second trimester | 23 |
| DX_ICD10 | O26.712 | subluxation of symphysis (pubis) in pregnancy, second trimester | 23 |
| DX_ICD10 | O26.812 | pregnancy related exhaustion and fatigue, second trimester | 23 |
| DX_ICD10 | O26.822 | pregnancy related peripheral neuritis, second trimester | 23 |
| DX_ICD10 | O26.832 | pregnancy related renal disease, second trimester | 23 |
| DX_ICD10 | O26.842 | uterine size-date discrepancy, second trimester | 23 |
| DX_ICD10 | O26.852 | spotting complicating pregnancy, second trimester | 23 |
| DX_ICD10 | O26.872 | cervical shortening, second trimester | 23 |
| DX_ICD10 | O26.892 | other specified pregnancy related conditions, second trimester | 23 |
| DX_ICD10 | O26.92 | pregnancy related conditions, unspecified, second trimester | 23 |
| DX_ICD10 | O29.012 | aspiration pneumonitis due to anesthesia during pregnancy, second trimester | 23 |
| DX_ICD10 | O29.022 | pressure collapse of lung due to anesthesia during pregnancy, second trimester | 23 |
| DX_ICD10 | O29.092 | other pulmonary complications of anesthesia during pregnancy, second trimester | 23 |
| DX_ICD10 | O29.112 | cardiac arrest due to anesthesia during pregnancy, second trimester | 23 |
| DX_ICD10 | O29.122 | cardiac failure due to anesthesia during pregnancy, second trimester | 23 |
| DX_ICD10 | O29.192 | other cardiac complications of anesthesia during pregnancy, second trimester | 23 |
| DX_ICD10 | O29.212 | cerebral anoxia due to anesthesia during pregnancy, second trimester | 23 |
| DX_ICD10 | O29.292 | other central nervous system complications of anesthesia during pregnancy, second tri | 23 |
| DX_ICD10 | O29.3X2 | toxic reaction to local anesthesia during pregnancy, second trimester | 23 |
| DX_ICD10 | O29.42 | spinal and epidural anesthesia induced headache during pregnancy, second trimester | 23 |
| DX_ICD10 | O29.5X2 | other complications of spinal and epidural anesthesia during preg, second trimester | 23 |
| DX_ICD10 | O29.62 | failed or difficult intubation for anesthesia during pregnancy, second trimester | 23 |
| DX_ICD10 | O29.8X2 | other complications of anesthesia during pregnancy, second trimester | 23 |
| DX_ICD10 | O29.92 | unspecified complication of anesthesia during pregnancy, second trimester | 23 |
| DX_ICD10 | O30.002 | twin preg, unspec # placenta & unspec # amniotic sacs, second trimester | 23 |
| DX_ICD10 | O30.012 | twin pregnancy, monochorionic/monoamniotic, second trimester | 23 |
| DX_ICD10 | O30.022 | conjoined twin pregnancy, second trimester | 23 |
| DX_ICD10 | O30.032 | twin pregnancy, monochorionic/diamniotic, second trimester | 23 |
| DX_ICD10 | O30.042 | twin pregnancy, dichorionic/diamniotic, second trimester | 23 |
| DX_ICD10 | O30.092 | twin pregnancy, unable to determine # placenta & # amniotic sacs, second tri | 23 |
| DX_ICD10 | O30.112 | triplet pregnancy with two or more monochorionic fetuses, second trimester | 23 |
| DX_ICD10 | O30.122 | triplet pregnancy with two or more monoamniotic fetuses, second trimester | 23 |
| DX_ICD10 | O30.132 | triplet pregnancy, trichorionic/triamniotic, second trimester | 23 |
| DX_ICD10 | O30.192 | triplet preg, unable to determine # placenta & # amniotic sacs, second tri | 23 |
| DX_ICD10 | O30.212 | quadruplet pregnancy with two or more monochorionic fetuses, second trimester | 23 |
| DX_ICD10 | O30.222 | quadruplet pregnancy with two or more monoamniotic fetuses, second trimester | 23 |
| DX_ICD10 | O30.232 | quadruplet pregnancy, quadrachorionic/quadra-amniotic, second trimester | 23 |
| DX_ICD10 | O30.292 | quadruplet pregnancy, unable to determine # placenta & # amniotic sacs, second tri | 23 |
| DX_ICD10 | O30.802 | other spec multi gestation, unspec # placenta & unspec # amniotic sacs, second tri | 23 |
| DX_ICD10 | O30.812 | other specified multiple gestation with two or more monochorionic fetuses, second tri | 23 |
| DX_ICD10 | O30.822 | other specified multiple gestation with two or more monoamniotic fetuses, second tri | 23 |
| DX_ICD10 | O30.832 | oth spec multi gestation, # chorions & amnions are equal to # fetuses, second tri | 23 |
| DX_ICD10 | O30.892 | oth spec multiple gestation, unable to determine # placenta & # amniotic sacs, sec tri | 23 |
| DX_ICD10 | O30.92 | multiple gestation, unspecified, second trimester | 23 |
| DX_ICD10 | O31.02X0 | papyraceous fetus, second trimester, not applicable or unspecified | 23 |
| DX_ICD10 | O31.02X1 | papyraceous fetus, second trimester, fetus 1 | 23 |
| DX_ICD10 | O31.02X2 | papyraceous fetus, second trimester, fetus 2 | 23 |
| DX_ICD10 | O31.02X3 | papyraceous fetus, second trimester, fetus 3 | 23 |
| DX_ICD10 | O31.02X4 | papyraceous fetus, second trimester, fetus 4 | 23 |
| DX_ICD10 | O31.02X5 | papyraceous fetus, second trimester, fetus 5 | 23 |
| DX_ICD10 | O31.02X9 | papyraceous fetus, second trimester, other fetus | 23 |
| DX_ICD10 | O31.12X0 | continuing preg after spontan abortion of 1 fetus+, second n/a or unspec | 23 |
| DX_ICD10 | O31.12X1 | continuing preg after spontan abortion of 1 fetus+, second trimester, fetus 1 | 23 |
| DX_ICD10 | O31.12X2 | continuing preg after spontan abortion of 1 fetus+, second trimester, fetus 2 | 23 |
| DX_ICD10 | O31.12X3 | continuing preg after spontan abortion of 1 fetus+, second trimester, fetus 3 | 23 |
| DX_ICD10 | O31.12X4 | continuing preg after spontan abortion of 1 fetus+, second trimester, fetus 4 | 23 |
| DX_ICD10 | O31.12X5 | continuing preg after spontan abortion of 1 fetus+, second trimester, fetus 5 | 23 |
| DX_ICD10 | O31.12X9 | continuing pregnancy after spontan abortion of 1 fetus+, second tri, oth fetus | 23 |
| DX_ICD10 | O31.22X0 | continuing preg after intrauterine death of 1 fetus+, second trimester, n/a or unspeci | 23 |
| DX_ICD10 | O31.22X1 | continuing preg after intrauterine death of one fetus or more, second trimester, fetus 1 | 23 |
| DX_ICD10 | O31.22X2 | continuing preg after intrauterine death of one fetus or more, second trimester, fetus 2 | 23 |
| DX_ICD10 | O31.22X3 | continuing preg after intrauterine death of one fetus or more, second trimester, fetus 3 | 23 |
| DX_ICD10 | O31.22X4 | continuing preg after intrauterine death of one fetus or more, second trimester, fetus 4 | 23 |
| DX_ICD10 | O31.22X5 | continuing preg after intrauterine death of one fetus or more, second trimester, fetus 5 | 23 |
| DX_ICD10 | O31.22X9 | continuing preg after intrauterine death of one fetus or more, second trim, oth fetus | 23 |
| DX_ICD10 | O31.32X0 | continuing preg after elective fetal reduction of one fetus+, second tri, n/a or unspec | 23 |
| DX_ICD10 | O31.32X1 | continuing preg after elective fetal reduction of 1 fetus+, second tri, fetus 1 | 23 |
| DX_ICD10 | O31.32X2 | continuing preg after elective fetal reduction of 1 fetus+, second tri, fetus 2 | 23 |
| DX_ICD10 | O31.32X3 | continuing preg after elective fetal reduction of 1 fetus+, second tri, fetus 3 | 23 |
| DX_ICD10 | O31.32X4 | continuing preg after elective fetal reduction of 1 fetus+, second tri, fetus 4 | 23 |
| DX_ICD10 | O31.32X5 | continuing preg after elective fetal reduction of 1 fetus+, second tri, fetus 5 | 23 |
| DX_ICD10 | O31.32X9 | continuing pregnancy after elective fetal reduction of one fetus+, second tri, oth fetus | 23 |
| DX_ICD10 | O31.8X20 | other complications specific to multiple gestation, second tri, n/a or unspec | 23 |
| DX_ICD10 | O31.8X21 | other complications specific to multiple gestation, second trimester, fetus 1 | 23 |
| DX_ICD10 | O31.8X22 | other complications specific to multiple gestation, second trimester, fetus 2 | 23 |
| DX_ICD10 | O31.8X23 | other complications specific to multiple gestation, second trimester, fetus 3 | 23 |
| DX_ICD10 | O31.8X24 | other complications specific to multiple gestation, second trimester, fetus 4 | 23 |
| DX_ICD10 | O31.8X25 | other complications specific to multiple gestation, second trimester, fetus 5 | 23 |
| DX_ICD10 | O31.8X29 | other complications specific to multiple gestation, second trimester, other fetus | 23 |
| DX_ICD10 | O34.02 | maternal care for unspecified congenital malformation of uterus, second trimester | 23 |
| DX_ICD10 | O34.12 | maternal care for benign tumor of corpus uteri, second trimester | 23 |
| DX_ICD10 | O34.32 | maternal care for cervical incompetence, second trimester | 23 |
| DX_ICD10 | O34.42 | maternal care for other abnormalities of cervix, second trimester | 23 |
| DX_ICD10 | O34.512 | maternal care for incarceration of gravid uterus, second trimester | 23 |
| DX_ICD10 | O34.522 | maternal care for prolapse of gravid uterus, second trimester | 23 |
| DX_ICD10 | O34.532 | maternal care for retroversion of gravid uterus, second trimester | 23 |
| DX_ICD10 | O34.592 | maternal care for other abnormalities of gravid uterus, second trimester | 23 |
| DX_ICD10 | O34.62 | maternal care for abnormality of vagina, second trimester | 23 |
| DX_ICD10 | O34.72 | maternal care for abnormality of vulva and perineum, second trimester | 23 |
| DX_ICD10 | O34.82 | maternal care for other abnormalities of pelvic organs, second trimester | 23 |
| DX_ICD10 | O34.92 | maternal care for abnormality of pelvic organ, unspecified, second trimester | 23 |
| DX_ICD10 | O36.0120 | maternal care for anti-d [rh] antibodies, second trimester, not applicable or unspecified | 23 |
| DX_ICD10 | O36.0121 | maternal care for anti-d [rh] antibodies, second trimester, fetus 1 | 23 |
| DX_ICD10 | O36.0122 | maternal care for anti-d [rh] antibodies, second trimester, fetus 2 | 23 |
| DX_ICD10 | O36.0123 | maternal care for anti-d [rh] antibodies, second trimester, fetus 3 | 23 |
| DX_ICD10 | O36.0124 | maternal care for anti-d [rh] antibodies, second trimester, fetus 4 | 23 |
| DX_ICD10 | O36.0125 | maternal care for anti-d [rh] antibodies, second trimester, fetus 5 | 23 |
| DX_ICD10 | O36.0129 | maternal care for anti-d [rh] antibodies, second trimester, other fetus | 23 |
| DX_ICD10 | O36.0920 | maternal care for other rhesus isoimmunization, second tri, not applicable or unspec | 23 |
| DX_ICD10 | O36.0921 | maternal care for other rhesus isoimmunization, second trimester, fetus 1 | 23 |
| DX_ICD10 | O36.0922 | maternal care for other rhesus isoimmunization, second trimester, fetus 2 | 23 |
| DX_ICD10 | O36.0923 | maternal care for other rhesus isoimmunization, second trimester, fetus 3 | 23 |
| DX_ICD10 | O36.0924 | maternal care for other rhesus isoimmunization, second trimester, fetus 4 | 23 |
| DX_ICD10 | O36.0925 | maternal care for other rhesus isoimmunization, second trimester, fetus 5 | 23 |
| DX_ICD10 | O36.0929 | maternal care for other rhesus isoimmunization, second trimester, other fetus | 23 |
| DX_ICD10 | O36.1120 | maternal care for anti-a sensitization, second trimester, not applicable or unspecified | 23 |
| DX_ICD10 | O36.1121 | maternal care for anti-a sensitization, second trimester, fetus 1 | 23 |
| DX_ICD10 | O36.1122 | maternal care for anti-a sensitization, second trimester, fetus 2 | 23 |
| DX_ICD10 | O36.1123 | maternal care for anti-a sensitization, second trimester, fetus 3 | 23 |
| DX_ICD10 | O36.1124 | maternal care for anti-a sensitization, second trimester, fetus 4 | 23 |
| DX_ICD10 | O36.1125 | maternal care for anti-a sensitization, second trimester, fetus 5 | 23 |
| DX_ICD10 | O36.1129 | maternal care for anti-a sensitization, second trimester, other fetus | 23 |
| DX_ICD10 | O36.1920 | maternal care for other isoimmunization, second trimester, not applicable or unspec | 23 |
| DX_ICD10 | O36.1921 | maternal care for other isoimmunization, second trimester, fetus 1 | 23 |
| DX_ICD10 | O36.1922 | maternal care for other isoimmunization, second trimester, fetus 2 | 23 |
| DX_ICD10 | O36.1923 | maternal care for other isoimmunization, second trimester, fetus 3 | 23 |
| DX_ICD10 | O36.1924 | maternal care for other isoimmunization, second trimester, fetus 4 | 23 |
| DX_ICD10 | O36.1925 | maternal care for other isoimmunization, second trimester, fetus 5 | 23 |
| DX_ICD10 | O36.1929 | maternal care for other isoimmunization, second trimester, other fetus | 23 |
| DX_ICD10 | O36.22X0 | maternal care for hydrops fetalis, second trimester, not applicable or unspecified | 23 |
| DX_ICD10 | O36.22X1 | maternal care for hydrops fetalis, second trimester, fetus 1 | 23 |
| DX_ICD10 | O36.22X2 | maternal care for hydrops fetalis, second trimester, fetus 2 | 23 |
| DX_ICD10 | O36.22X3 | maternal care for hydrops fetalis, second trimester, fetus 3 | 23 |
| DX_ICD10 | O36.22X4 | maternal care for hydrops fetalis, second trimester, fetus 4 | 23 |
| DX_ICD10 | O36.22X5 | maternal care for hydrops fetalis, second trimester, fetus 5 | 23 |
| DX_ICD10 | O36.22X9 | maternal care for hydrops fetalis, second trimester, other fetus | 23 |
| DX_ICD10 | O36.5120 | maternal care for known or suspected placental insufficiency, second tri, n/a or unspec | 23 |
| DX_ICD10 | O36.5121 | maternal care for known or suspected placental insufficiency, second trimester, fetus 1 | 23 |
| DX_ICD10 | O36.5122 | maternal care for known or suspected placental insufficiency, second trimester, fetus 2 | 23 |
| DX_ICD10 | O36.5123 | maternal care for known or suspected placental insufficiency, second trimester, fetus 3 | 23 |
| DX_ICD10 | O36.5124 | maternal care for known or suspected placental insufficiency, second trimester, fetus 4 | 23 |
| DX_ICD10 | O36.5125 | maternal care for known or suspected placental insufficiency, second trimester, fetus 5 | 23 |
| DX_ICD10 | O36.5129 | maternal care for known or suspected placental insufficiency, second tri, other fetus | 23 |
| DX_ICD10 | O36.5920 | maternal care for other known or suspected poor fetal growth, second tri, n/a or unspec | 23 |
| DX_ICD10 | O36.5921 | maternal care for other known or suspected poor fetal growth, second trimester, fetus 1 | 23 |
| DX_ICD10 | O36.5922 | maternal care for other known or suspected poor fetal growth, second trimester, fetus 2 | 23 |
| DX_ICD10 | O36.5923 | maternal care for other known or suspected poor fetal growth, second trimester, fetus 3 | 23 |
| DX_ICD10 | O36.5924 | maternal care for other known or suspected poor fetal growth, second trimester, fetus 4 | 23 |
| DX_ICD10 | O36.5925 | maternal care for other known or suspected poor fetal growth, second trimester, fetus 5 | 23 |
| DX_ICD10 | O36.5929 | maternal care for other known or suspected poor fetal growth, second tri, oth fetus | 23 |
| DX_ICD10 | O36.62X0 | maternal care for excessive fetal growth, second trimester, not applicable or unspec | 23 |
| DX_ICD10 | O36.62X1 | maternal care for excessive fetal growth, second trimester, fetus 1 | 23 |
| DX_ICD10 | O36.62X2 | maternal care for excessive fetal growth, second trimester, fetus 2 | 23 |
| DX_ICD10 | O36.62X3 | maternal care for excessive fetal growth, second trimester, fetus 3 | 23 |
| DX_ICD10 | O36.62X4 | maternal care for excessive fetal growth, second trimester, fetus 4 | 23 |
| DX_ICD10 | O36.62X5 | maternal care for excessive fetal growth, second trimester, fetus 5 | 23 |
| DX_ICD10 | O36.62X9 | maternal care for excessive fetal growth, second trimester, other fetus | 23 |
| DX_ICD10 | O36.72X0 | maternal care for viable fetus in abdominal pregnancy, second trimester, n/a or unspec | 23 |
| DX_ICD10 | O36.72X1 | maternal care for viable fetus in abdominal pregnancy, second trimester, fetus 1 | 23 |
| DX_ICD10 | O36.72X2 | maternal care for viable fetus in abdominal pregnancy, second trimester, fetus 2 | 23 |
| DX_ICD10 | O36.72X3 | maternal care for viable fetus in abdominal pregnancy, second trimester, fetus 3 | 23 |
| DX_ICD10 | O36.72X4 | maternal care for viable fetus in abdominal pregnancy, second trimester, fetus 4 | 23 |
| DX_ICD10 | O36.72X5 | maternal care for viable fetus in abdominal pregnancy, second trimester, fetus 5 | 23 |
| DX_ICD10 | O36.72X9 | maternal care for viable fetus in abdominal pregnancy, second trimester, other fetus | 23 |
| DX_ICD10 | O36.8120 | decreased fetal movements, second trimester, not applicable or unspecified | 23 |
| DX_ICD10 | O36.8121 | decreased fetal movements, second trimester, fetus 1 | 23 |
| DX_ICD10 | O36.8122 | decreased fetal movements, second trimester, fetus 2 | 23 |
| DX_ICD10 | O36.8123 | decreased fetal movements, second trimester, fetus 3 | 23 |
| DX_ICD10 | O36.8124 | decreased fetal movements, second trimester, fetus 4 | 23 |
| DX_ICD10 | O36.8125 | decreased fetal movements, second trimester, fetus 5 | 23 |
| DX_ICD10 | O36.8129 | decreased fetal movements, second trimester, other fetus | 23 |
| DX_ICD10 | O36.8220 | fetal anemia and thrombocytopenia, second trimester, not applicable or unspecified | 23 |
| DX_ICD10 | O36.8221 | fetal anemia and thrombocytopenia, second trimester, fetus 1 | 23 |
| DX_ICD10 | O36.8222 | fetal anemia and thrombocytopenia, second trimester, fetus 2 | 23 |
| DX_ICD10 | O36.8223 | fetal anemia and thrombocytopenia, second trimester, fetus 3 | 23 |
| DX_ICD10 | O36.8224 | fetal anemia and thrombocytopenia, second trimester, fetus 4 | 23 |
| DX_ICD10 | O36.8225 | fetal anemia and thrombocytopenia, second trimester, fetus 5 | 23 |
| DX_ICD10 | O36.8229 | fetal anemia and thrombocytopenia, second trimester, other fetus | 23 |
| DX_ICD10 | O36.8320 | maternal care for abnormalities of fetal heart rate or rhythm, second tri, n/a or unspec | 23 |
| DX_ICD10 | O36.8321 | maternal care for abnormalities of fetal heart rate or rhythm, second trimester, fetus 1 | 23 |
| DX_ICD10 | O36.8322 | maternal care for abnormalities of fetal heart rate or rhythm, second trimester, fetus 2 | 23 |
| DX_ICD10 | O36.8323 | maternal care for abnormalities of fetal heart rate or rhythm, second trimester, fetus 3 | 23 |
| DX_ICD10 | O36.8324 | maternal care for abnormalities of fetal heart rate or rhythm, second trimester, fetus 4 | 23 |
| DX_ICD10 | O36.8325 | maternal care for abnormalities of fetal heart rate or rhythm, second trimester, fetus 5 | 23 |
| DX_ICD10 | O36.8329 | maternal care for abnormalities of the fetal heart rate or rhythm, second tri, oth fetus | 23 |
| DX_ICD10 | O36.8920 | maternal care for oth spec fetal problems, second tri, n/a or unspec | 23 |
| DX_ICD10 | O36.8921 | maternal care for other specified fetal problems, second trimester, fetus 1 | 23 |
| DX_ICD10 | O36.8922 | maternal care for other specified fetal problems, second trimester, fetus 2 | 23 |
| DX_ICD10 | O36.8923 | maternal care for other specified fetal problems, second trimester, fetus 3 | 23 |
| DX_ICD10 | O36.8924 | maternal care for other specified fetal problems, second trimester, fetus 4 | 23 |
| DX_ICD10 | O36.8925 | maternal care for other specified fetal problems, second trimester, fetus 5 | 23 |
| DX_ICD10 | O36.8929 | maternal care for other specified fetal problems, second trimester, oth fetus | 23 |
| DX_ICD10 | O36.92X0 | maternal care for fetal problem, unspecified, second trimester, n/a or unspec | 23 |
| DX_ICD10 | O36.92X1 | maternal care for fetal problem, unspecified, second trimester, fetus 1 | 23 |
| DX_ICD10 | O36.92X2 | maternal care for fetal problem, unspecified, second trimester, fetus 2 | 23 |
| DX_ICD10 | O36.92X3 | maternal care for fetal problem, unspecified, second trimester, fetus 3 | 23 |
| DX_ICD10 | O36.92X4 | maternal care for fetal problem, unspecified, second trimester, fetus 4 | 23 |
| DX_ICD10 | O36.92X5 | maternal care for fetal problem, unspecified, second trimester, fetus 5 | 23 |
| DX_ICD10 | O36.92X9 | maternal care for fetal problem, unspecified, second trimester, other fetus | 23 |
| DX_ICD10 | O40.2XX0 | polyhydramnios, second trimester, not applicable or unspecified | 23 |
| DX_ICD10 | O40.2XX1 | polyhydramnios, second trimester, fetus 1 | 23 |
| DX_ICD10 | O40.2XX2 | polyhydramnios, second trimester, fetus 2 | 23 |
| DX_ICD10 | O40.2XX3 | polyhydramnios, second trimester, fetus 3 | 23 |
| DX_ICD10 | O40.2XX4 | polyhydramnios, second trimester, fetus 4 | 23 |
| DX_ICD10 | O40.2XX5 | polyhydramnios, second trimester, fetus 5 | 23 |
| DX_ICD10 | O40.2XX9 | polyhydramnios, second trimester, other fetus | 23 |
| DX_ICD10 | O41.02X0 | oligohydramnios, second trimester, not applicable or unspecified | 23 |
| DX_ICD10 | O41.02X1 | oligohydramnios, second trimester, fetus 1 | 23 |
| DX_ICD10 | O41.02X2 | oligohydramnios, second trimester, fetus 2 | 23 |
| DX_ICD10 | O41.02X3 | oligohydramnios, second trimester, fetus 3 | 23 |
| DX_ICD10 | O41.02X4 | oligohydramnios, second trimester, fetus 4 | 23 |
| DX_ICD10 | O41.02X5 | oligohydramnios, second trimester, fetus 5 | 23 |
| DX_ICD10 | O41.02X9 | oligohydramnios, second trimester, other fetus | 23 |
| DX_ICD10 | O41.1020 | infection of amniotic sac and membranes, unspecified, second trimester, n/a or unspec | 23 |
| DX_ICD10 | O41.1021 | infection of amniotic sac and membranes, unspecified, second trimester, fetus 1 | 23 |
| DX_ICD10 | O41.1022 | infection of amniotic sac and membranes, unspecified, second trimester, fetus 2 | 23 |
| DX_ICD10 | O41.1023 | infection of amniotic sac and membranes, unspecified, second trimester, fetus 3 | 23 |
| DX_ICD10 | O41.1024 | infection of amniotic sac and membranes, unspecified, second trimester, fetus 4 | 23 |
| DX_ICD10 | O41.1025 | infection of amniotic sac and membranes, unspecified, second trimester, fetus 5 | 23 |
| DX_ICD10 | O41.1029 | infection of amniotic sac and membranes, unspecified, second trimester, other fetus | 23 |
| DX_ICD10 | O41.1220 | chorioamnionitis, second trimester, not applicable or unspecified | 23 |
| DX_ICD10 | O41.1221 | chorioamnionitis, second trimester, fetus 1 | 23 |
| DX_ICD10 | O41.1222 | chorioamnionitis, second trimester, fetus 2 | 23 |
| DX_ICD10 | O41.1223 | chorioamnionitis, second trimester, fetus 3 | 23 |
| DX_ICD10 | O41.1224 | chorioamnionitis, second trimester, fetus 4 | 23 |
| DX_ICD10 | O41.1225 | chorioamnionitis, second trimester, fetus 5 | 23 |
| DX_ICD10 | O41.1229 | chorioamnionitis, second trimester, other fetus | 23 |
| DX_ICD10 | O41.1420 | placentitis, second trimester, not applicable or unspecified | 23 |
| DX_ICD10 | O41.1421 | placentitis, second trimester, fetus 1 | 23 |
| DX_ICD10 | O41.1422 | placentitis, second trimester, fetus 2 | 23 |
| DX_ICD10 | O41.1423 | placentitis, second trimester, fetus 3 | 23 |
| DX_ICD10 | O41.1424 | placentitis, second trimester, fetus 4 | 23 |
| DX_ICD10 | O41.1425 | placentitis, second trimester, fetus 5 | 23 |
| DX_ICD10 | O41.1429 | placentitis, second trimester, oth fetus | 23 |
| DX_ICD10 | O41.8X20 | other specified disorders of amniotic fluid and membranes, second tri, n/a or unspec | 23 |
| DX_ICD10 | O41.8X21 | other specified disorders of amniotic fluid and membranes, second trimester, fetus 1 | 23 |
| DX_ICD10 | O41.8X22 | other specified disorders of amniotic fluid and membranes, second trimester, fetus 2 | 23 |
| DX_ICD10 | O41.8X23 | other specified disorders of amniotic fluid and membranes, second trimester, fetus 3 | 23 |
| DX_ICD10 | O41.8X24 | other specified disorders of amniotic fluid and membranes, second trimester, fetus 4 | 23 |
| DX_ICD10 | O41.8X25 | other specified disorders of amniotic fluid and membranes, second trimester, fetus 5 | 23 |
| DX_ICD10 | O41.8X29 | other specified disorders of amniotic fluid and membranes, second trimester, oth fetus | 23 |
| DX_ICD10 | O41.92X0 | disorder of amniotic fluid and membranes, unspec, second trimester, n/a or unspec | 23 |
| DX_ICD10 | O41.92X1 | disorder of amniotic fluid and membranes, unspecified, second trimester, fetus 1 | 23 |
| DX_ICD10 | O41.92X2 | disorder of amniotic fluid and membranes, unspecified, second trimester, fetus 2 | 23 |
| DX_ICD10 | O41.92X3 | disorder of amniotic fluid and membranes, unspecified, second trimester, fetus 3 | 23 |
| DX_ICD10 | O41.92X4 | disorder of amniotic fluid and membranes, unspecified, second trimester, fetus 4 | 23 |
| DX_ICD10 | O41.92X5 | disorder of amniotic fluid and membranes, unspecified, second trimester, fetus 5 | 23 |
| DX_ICD10 | O41.92X9 | disorder of amniotic fluid and membranes, unspecified, second trimester, other fetus | 23 |
| DX_ICD10 | O42.012 | preterm premature rupture of membranes, onset of labor w in 24 hrs rupture, second tri | 23 |
| DX_ICD10 | O42.112 | preterm premature rupture of membranes, onset labor 24 hrs+ after rupture, sec tri | 23 |
| DX_ICD10 | O42.912 | preterm premat rupt membranes, unspec length time btwn rupture & laborr, second tri | 23 |
| DX_ICD10 | O43.012 | fetomaternal placental transfusion syndrome, second trimester | 23 |
| DX_ICD10 | O43.022 | fetus-to-fetus placental transfusion syndrome, second trimester | 23 |
| DX_ICD10 | O43.102 | malformation of placenta, unspecified, second trimester | 23 |
| DX_ICD10 | O43.112 | circumvallate placenta, second trimester | 23 |
| DX_ICD10 | O43.122 | velamentous insertion of umbilical cord, second trimester | 23 |
| DX_ICD10 | O43.192 | other malformation of placenta, second trimester | 23 |
| DX_ICD10 | O43.212 | placenta accreta, second trimester | 23 |
| DX_ICD10 | O43.222 | placenta increta, second trimester | 23 |
| DX_ICD10 | O43.232 | placenta percreta, second trimester | 23 |
| DX_ICD10 | O43.812 | placental infarction, second trimester | 23 |
| DX_ICD10 | O43.892 | other placental disorders, second trimester | 23 |
| DX_ICD10 | O43.92 | unspecified placental disorder, second trimester | 23 |
| DX_ICD10 | O44.02 | complete placenta previa nos or without hemorrhage, second trimester | 23 |
| DX_ICD10 | O44.12 | complete placenta previa with hemorrhage, second trimester | 23 |
| DX_ICD10 | O44.22 | partial placenta previa nos or without hemorrhage, second trimester | 23 |
| DX_ICD10 | O44.32 | partial placenta previa with hemorrhage, second trimester | 23 |
| DX_ICD10 | O44.42 | low lying placenta nos or without hemorrhage, second trimester | 23 |
| DX_ICD10 | O44.52 | low lying placenta with hemorrhage, second trimester | 23 |
| DX_ICD10 | O45.002 | premature separation of placenta with coagulation defect, unspecified, second tri | 23 |
| DX_ICD10 | O45.012 | premature separation of placenta with afibrinogenemia, second trimester | 23 |
| DX_ICD10 | O45.022 | premature separation of placenta with disseminated intravascular coagul, second tri | 23 |
| DX_ICD10 | O45.092 | premature separation of placenta with other coagulation defect, second trimester | 23 |
| DX_ICD10 | O45.8X2 | other premature separation of placenta, second trimester | 23 |
| DX_ICD10 | O45.92 | premature separation of placenta, unspecified, second trimester | 23 |
| DX_ICD10 | O46.002 | antepartum hemorrhage with coagulation defect, unspecified, second trimester | 23 |
| DX_ICD10 | O46.012 | antepartum hemorrhage with afibrinogenemia, second trimester | 23 |
| DX_ICD10 | O46.022 | antepartum hemorrhage with disseminated intravascular coagulation, second trimester | 23 |
| DX_ICD10 | O46.092 | antepartum hemorrhage with other coagulation defect, second trimester | 23 |
| DX_ICD10 | O46.8X2 | other antepartum hemorrhage, second trimester | 23 |
| DX_ICD10 | O46.92 | antepartum hemorrhage, unspecified, second trimester | 23 |
| DX_ICD10 | O60.02 | preterm labor without delivery, second trimester | 23 |
| DX_ICD10 | O60.12X0 | preterm labor second trimester with preterm delivery second trimester, n/a or unspec | 23 |
| DX_ICD10 | O60.12X1 | preterm labor second trimester with preterm delivery second trimester, fetus 1 | 23 |
| DX_ICD10 | O60.12X2 | preterm labor second trimester with preterm delivery second trimester, fetus 2 | 23 |
| DX_ICD10 | O60.12X3 | preterm labor second trimester with preterm delivery second trimester, fetus 3 | 23 |
| DX_ICD10 | O60.12X4 | preterm labor second trimester with preterm delivery second trimester, fetus 4 | 23 |
| DX_ICD10 | O60.12X5 | preterm labor second trimester with preterm delivery second trimester, fetus 5 | 23 |
| DX_ICD10 | O60.12X9 | preterm labor second trimester with preterm delivery second trimester, other fetus | 23 |
| DX_ICD10 | O60.22X0 | term delivery with preterm labor, second trimester, not applicable or unspecified | 23 |
| DX_ICD10 | O60.22X1 | term delivery with preterm labor, second trimester, fetus 1 | 23 |
| DX_ICD10 | O60.22X2 | term delivery with preterm labor, second trimester, fetus 2 | 23 |
| DX_ICD10 | O60.22X3 | term delivery with preterm labor, second trimester, fetus 3 | 23 |
| DX_ICD10 | O60.22X4 | term delivery with preterm labor, second trimester, fetus 4 | 23 |
| DX_ICD10 | O60.22X5 | term delivery with preterm labor, second trimester, fetus 5 | 23 |
| DX_ICD10 | O60.22X9 | term delivery with preterm labor, second trimester, other fetus | 23 |
| DX_ICD10 | O62.1 | secondary uterine inertia | 23 |
| DX_ICD10 | O63.1 | prolonged second stage (of labor) | 23 |
| DX_ICD10 | O63.2 | delayed delivery of second twin, triplet, etc. | 23 |
| DX_ICD10 | O70.1 | second degree perineal laceration during delivery | 23 |
| DX_ICD10 | O71.02 | rupture of uterus before onset of labor, second trimester | 23 |
| DX_ICD10 | O72.2 | delayed and secondary postpartum hemorrhage | 23 |
| DX_ICD10 | O88.012 | air embolism in pregnancy, second trimester | 23 |
| DX_ICD10 | O88.112 | amniotic fluid embolism in pregnancy, second trimester | 23 |
| DX_ICD10 | O88.212 | thromboembolism in pregnancy, second trimester | 23 |
| DX_ICD10 | O88.312 | pyemic and septic embolism in pregnancy, second trimester | 23 |
| DX_ICD10 | O88.812 | other embolism in pregnancy, second trimester | 23 |
| DX_ICD10 | O91.012 | infection of nipple associated with pregnancy, second trimester | 23 |
| DX_ICD10 | O91.112 | abscess of breast associated with pregnancy, second trimester | 23 |
| DX_ICD10 | O91.212 | nonpurulent mastitis associated with pregnancy, second trimester | 23 |
| DX_ICD10 | O92.012 | retracted nipple associated with pregnancy, second trimester | 23 |
| DX_ICD10 | O92.112 | cracked nipple associated with pregnancy, second trimester | 23 |
| DX_ICD10 | O98.012 | tuberculosis complicating pregnancy, second trimester | 23 |
| DX_ICD10 | O98.112 | syphilis complicating pregnancy, second trimester | 23 |
| DX_ICD10 | O98.212 | gonorrhea complicating pregnancy, second trimester | 23 |
| DX_ICD10 | O98.312 | other infections w a predominantly sexual mode of transmission complic preg, sec tri | 23 |
| DX_ICD10 | O98.412 | viral hepatitis complicating pregnancy, second trimester | 23 |
| DX_ICD10 | O98.512 | other viral diseases complicating pregnancy, second trimester | 23 |
| DX_ICD10 | O98.612 | protozoal diseases complicating pregnancy, second trimester | 23 |
| DX_ICD10 | O98.712 | human immunodeficiency virus [hiv] disease complicating pregnancy, second trimester | 23 |
| DX_ICD10 | O98.812 | other maternal infectious and parasitic diseases complicating pregnancy, second tri | 23 |
| DX_ICD10 | O98.912 | unspecified maternal infectious and parasitic disease complicating preg, second tri | 23 |
| DX_ICD10 | O99.012 | anemia complicating pregnancy, second trimester | 23 |
| DX_ICD10 | O99.112 | other dis blood & blood-form organs & certain dis immune mech complic preg, sec tri | 23 |
| DX_ICD10 | O99.212 | obesity complicating pregnancy, second trimester | 23 |
| DX_ICD10 | O99.282 | endocrine, nutritional and metabolic diseases complicating pregnancy, second tri | 23 |
| DX_ICD10 | O99.312 | alcohol use complicating pregnancy, second trimester | 23 |
| DX_ICD10 | O99.322 | drug use complicating pregnancy, second trimester | 23 |
| DX_ICD10 | O99.332 | smoking (tobacco) complicating pregnancy, second trimester | 23 |
| DX_ICD10 | O99.342 | other mental disorders complicating pregnancy, second trimester | 23 |
| DX_ICD10 | O99.352 | diseases of the nervous system complicating pregnancy, second trimester | 23 |
| DX_ICD10 | O99.412 | diseases of the circulatory system complicating pregnancy, second trimester | 23 |
| DX_ICD10 | O99.512 | diseases of the respiratory system complicating pregnancy, second trimester | 23 |
| DX_ICD10 | O99.612 | diseases of the digestive system complicating pregnancy, second trimester | 23 |
| DX_ICD10 | O99.712 | diseases of the skin and subcutaneous tissue complicating pregnancy, second tri | 23 |
| DX_ICD10 | O99.842 | bariatric surgery status complicating pregnancy, second trimester | 23 |
| DX_ICD10 | O9A.112 | malignant neoplasm complicating pregnancy, second trimester | 23 |
| DX_ICD10 | O9A.212 | injury, poisoning & certain other consequences of external causes complic preg, sec tri | 23 |
| DX_ICD10 | O9A.312 | physical abuse complicating pregnancy, second trimester | 23 |
| DX_ICD10 | O9A.412 | sexual abuse complicating pregnancy, second trimester | 23 |
| DX_ICD10 | O9A.512 | psychological abuse complicating pregnancy, second trimester | 23 |
| DX_ICD10 | P07.22 | extreme immaturity of newborn, gestational age 23 completed weeks | 23 |
| DX_ICD10 | Z34.02 | encounter for supervision of normal first pregnancy, second trimester | 23 |
| DX_ICD10 | Z34.82 | encounter for supervision of other normal pregnancy, second trimester | 23 |
| DX_ICD10 | Z34.92 | encounter for supervision of normal pregnancy, unspecified, second trimester | 23 |
| DX_ICD10 | Z3A.23 | 23 weeks gestation of pregnancy | 23 |
| DX_ICD9 | 765.22 | 24 completed wks gestation | 24 |
| DX_ICD10 | P07.23 | extreme immaturity of newborn, gestational age 24 completed weeks | 24 |
| DX_ICD10 | Z3A.24 | 24 weeks gestation of pregnancy | 24 |
| DX_ICD9 | 765.02 | extreme immaturity 500-749g (begin 1988) | 25 |
| DX_ICD9 | 765.12 | preterm nec 500-749g (begin 1988) | 25 |
| DX_ICD10 | P07.02 | extremely low birth weight newborn, 500-749 grams | 25 |
| DX_ICD10 | P07.02 | extremely low birth weight newborn, 500-749 grams | 25 |
| DX_ICD10 | P07.24 | extreme immaturity of newborn, gestational age 25 completed weeks | 25 |
| DX_ICD10 | Z3A.25 | 25 weeks gestation of pregnancy | 25 |
| DX_ICD9 | 765.23 | 25-26 completed wks gestation | 26 |
| DX_ICD10 | P07.25 | extreme immaturity of newborn, gestational age 26 completed weeks | 26 |
| DX_ICD10 | Z3A.26 | 26 weeks gestation of pregnancy | 26 |
| DX_ICD9 | 765.03 | extreme immaturity 750-999g (begin 1988) | 27 |
| DX_ICD9 | 765.13 | preterm nec 750-999g (begin 1988) | 27 |
| DX_ICD10 | P07.03 | extremely low birth weight newborn, 750-999 grams | 27 |
| DX_ICD10 | P07.26 | extreme immaturity of newborn, gestational age 27 completed weeks | 27 |
| DX_ICD10 | Z3A.27 | 27 weeks gestation of pregnancy | 27 |
| DX_ICD9 | 765.00 | extreme immaturity weight nos | 28 |
| DX_ICD9 | 765.0 | extreme immaturity (begin 1980, end 1988) | 28 |
| DX_ICD9 | 765.24 | 27-28 completed wks gestation | 28 |
| DX_ICD10 | P07.31 | preterm newborn, gestational age 28 completed weeks | 28 |
| DX_ICD10 | Z3A.28 | 28 weeks gestation of pregnancy | 28 |
| DX_ICD9 | 765.04 | extreme immaturity 1000-1249g (begin 1988) | 29 |
| DX_ICD9 | 765.14 | preterm nec 1000-1249g (begin 1988) | 29 |
| DX_ICD10 | P07.14 | other low birth weight newborn, 1000-1249 grams | 29 |
| DX_ICD10 | P07.14 | other low birth weight newborn, 1000-1249 grams | 29 |
| DX_ICD10 | P07.32 | preterm newborn, gestational age 29 completed weeks | 29 |
| DX_ICD10 | Z3A.29 | 29 weeks gestation of pregnancy | 29 |
| DX_ICD9 | 651.31 | twins w fetal loss & retention of 1 fetus, delivered, w or w/o antepartum | 30 |
| DX_ICD9 | 651.41 | triplets w fetal loss & retention of 1+ fetus(es), delivered, w or w/o antepartum | 30 |
| DX_ICD9 | 651.51 | quadruplet w fetal loss & retention of 1+ fetus(es), delivered, w or w/o antepartum | 30 |
| DX_ICD9 | 765.05 | extreme immaturity 1250-1499g (begin 1988) | 30 |
| DX_ICD9 | 765.15 | preterm nec 1250-1499g (begin 1988) | 30 |
| DX_ICD9 | 765.25 | 29-30 completed wks gestation | 30 |
| DX_ICD9 | V27.1 | deliver- single stillborn | 30 |
| DX_ICD9 | V27.3 | outcome of delivery, twins, 1 liveborn & 1 stillborn | 30 |
| DX_ICD9 | V27.4 | outcome of delivery, twins, both stillborn | 30 |
| DX_ICD9 | V27.6 | outcome of delivery, other multiple birth, some liveborn | 30 |
| DX_ICD9 | V27.7 | deliver- multiple births- all stillborn | 30 |
| DX_ICD9 | V32.0 | twin, mate stillborn- in hospital | 30 |
| DX_ICD9 | V32.00 | twin birth, mate stillborn, born in hospital, delivered w/o c-section | 30 |
| DX_ICD9 | V32.01 | twin birth, mate stillborn, born in hospital, delivered by c-section | 30 |
| DX_ICD9 | V32.1 | twin birth, mate stillborn, born before admission to hospital | 30 |
| DX_ICD9 | V32.2 | twin birth, mate stillborn, born outside hospital and not hospitalized | 30 |
| DX_ICD9 | V35.0 | other multiple stillborn- in hospital | 30 |
| DX_ICD9 | V35.00 | other multiple birth (3+), mates all stillborn, born in hospital, delivered w/o c-section | 30 |
| DX_ICD9 | V35.01 | other multiple birth (3+), mates all stillborn, born in hospital, delivered by c-section | 30 |
| DX_ICD9 | V35.1 | oth multiple birth (three or more), mates all stillborn, born before admission to hospital | 30 |
| DX_ICD9 | V35.2 | oth multi birth (3+), mates all stillborn, born outside of hospital & not hospitalized | 30 |
| DX_ICD9 | V36.0 | multiple liveborn/stillborn- in hospital | 30 |
| DX_ICD9 | V36.00 | other multiple birth (3+), mates LB & stillborn, born in hosp, delivered w/o c-section | 30 |
| DX_ICD9 | V36.01 | other multiple birth (3+), mates LB & stillborn, born in hospital, delivered w/o c-section | 30 |
| DX_ICD9 | V36.1 | other multiple birth (3+), mates LB and stillborn, born before admission to hospital | 30 |
| DX_ICD10 | P07.15 | other low birth weight newborn, 1250-1499 grams | 30 |
| DX_ICD10 | P07.33 | preterm newborn, gestational age 30 completed weeks | 30 |
| DX_ICD10 | P95 | stillbirth | 30 |
| DX_ICD10 | Z37.1 | single stillbirth | 30 |
| DX_ICD10 | Z37.3 | twins, one liveborn and one stillborn | 30 |
| DX_ICD10 | Z37.4 | twins, both stillborn | 30 |
| DX_ICD10 | Z37.6 | other multiple births, some liveborn | 30 |
| DX_ICD10 | Z37.60 | multiple births, unspecified, some liveborn | 30 |
| DX_ICD10 | Z37.61 | triplets, some liveborn | 30 |
| DX_ICD10 | Z37.62 | quadruplets, some liveborn | 30 |
| DX_ICD10 | Z37.63 | quintuplets, some liveborn | 30 |
| DX_ICD10 | Z37.64 | sextuplets, some liveborn | 30 |
| DX_ICD10 | Z37.69 | other multiple births, some liveborn | 30 |
| DX_ICD10 | Z37.7 | other multiple births, all stillborn | 30 |
| DX_ICD10 | Z3A.30 | 30 weeks gestation of pregnancy | 30 |
| DX_ICD9 | 651.21 | quadruplet pregnancy, delivered, w or w/o- antepartum | 31 |
| DX_ICD9 | 765.06 | extreme immaturity 1500-1749g (begin 1988) | 31 |
| DX_ICD9 | 765.16 | preterm nec 1500- 1749g (begin 1988) | 31 |
| DX_ICD10 | P07.16 | other low birth weight newborn, 1500-1749 grams | 31 |
| DX_ICD10 | P07.16 | other low birth weight newborn, 1500-1749 grams | 31 |
| DX_ICD10 | P07.34 | preterm newborn, gestational age 31 completed weeks | 31 |
| DX_ICD10 | Z3A.31 | 31 weeks gestation of pregnancy | 31 |
| DX_ICD9 | 765.07 | extreme immaturity 1750-1999g (begin 1988) | 32 |
| DX_ICD9 | 765.17 | preterm nec1750-1999g (begin 1988) | 32 |
| DX_ICD9 | 765.26 | 31-32 completed wks gestation | 32 |
| DX_ICD10 | P07.17 | other low birth weight newborn, 1750-1999 grams | 32 |
| DX_ICD10 | P07.35 | preterm newborn, gestational age 32 completed weeks | 32 |
| DX_ICD10 | Z3A.32 | 32 weeks gestation of pregnancy | 32 |
| DX_ICD9 | 651.11 | triplet pregnancy, delivered, w or w/o- antepartum | 33 |
| DX_ICD9 | V27.5 | outcome of delivery, other multiple birth, all liveborn | 33 |
| DX_ICD9 | V34.0 | other multiple liveborn- in hospital | 33 |
| DX_ICD9 | V34.00 | other multiple birth (3+), mates all liveborn, born in hospital, delivered w/o c-section | 33 |
| DX_ICD9 | V34.01 | other multiple birth (3+), mates all liveborn, born in hospital, delivered by c-section | 33 |
| DX_ICD9 | V34.1 | other multiple birth (3+), mates all liveborn, born before admission to hospital | 33 |
| DX_ICD9 | V34.2 | other multiple birth (3+), mates all liveborn, born outside hospital and not hospitalized | 33 |
| DX_ICD9 | V36.2 | other multiple birth (3+), mates LB & stillborn, born outside hospital & not hospitalized | 33 |
| DX_ICD9 | V37 | other/unspecified multiple births | 33 |
| DX_ICD9 | V37.0 | multiple birth nos- in hospital | 33 |
| DX_ICD9 | V37.00 | oth multi birth (3+), unspec if mates LB or stillborn, born in hospital, del w/o c-section | 33 |
| DX_ICD9 | V37.01 | oth multi birth (3+), unspec if mates LB or stillborn, born in hospital, del by c-section | 33 |
| DX_ICD9 | V37.1 | oth multi birth (3+), unspec whether mates LB or stillborn, before admission to hospital | 33 |
| DX_ICD9 | V37.2 | oth multi birth (3+), unspec whether mates liveborn or stillborn, born outside of hospital | 33 |
| DX_ICD10 | P07.36 | preterm newborn, gestational age 33 completed weeks | 33 |
| DX_ICD10 | Z37.5 | other multiple births, all liveborn | 33 |
| DX_ICD10 | Z37.50 | multiple births, unspecified, all liveborn | 33 |
| DX_ICD10 | Z37.51 | triplets, all liveborn | 33 |
| DX_ICD10 | Z37.52 | quadruplets, all liveborn | 33 |
| DX_ICD10 | Z37.53 | quintuplets, all liveborn | 33 |
| DX_ICD10 | Z37.54 | sextuplets, all liveborn | 33 |
| DX_ICD10 | Z37.59 | other multiple births, all liveborn | 33 |
| DX_ICD10 | Z38.61 | triplet liveborn infant, delivered vaginally | 33 |
| DX_ICD10 | Z38.62 | triplet liveborn infant, delivered by cesarean | 33 |
| DX_ICD10 | Z38.63 | quadruplet liveborn infant, delivered vaginally | 33 |
| DX_ICD10 | Z38.64 | quadruplet liveborn infant, delivered by cesarean | 33 |
| DX_ICD10 | Z38.65 | quintuplet liveborn infant, delivered vaginally | 33 |
| DX_ICD10 | Z38.66 | quintuplet liveborn infant, delivered by cesarean | 33 |
| DX_ICD10 | Z38.68 | other multiple liveborn infant, delivered vaginally | 33 |
| DX_ICD10 | Z38.69 | other multiple liveborn infant, delivered by cesarean | 33 |
| DX_ICD10 | Z38.7 | other multiple liveborn infant, born outside hospital | 33 |
| DX_ICD10 | Z38.8 | other multiple liveborn infant, unspecified as to place of birth | 33 |
| DX_ICD10 | Z3A.33 | 33 weeks gestation of pregnancy | 33 |
| DX_ICD9 | 644.21 | early onset delivery- delivered | 34 |
| DX_ICD9 | 765.08 | extreme immaturity 2000-2499g (begin 1988) | 34 |
| DX_ICD9 | 765.1 | other preterm infants (begin 1980, end 1988) | 34 |
| DX_ICD9 | 765.18 | preterm nec 2000-2499g (begin 1988) | 34 |
| DX_ICD9 | 765.27 | 33-34 completed wks gestation | 34 |
| DX_ICD10 | O30.103 | triplet preg, unspec #f placenta and unspecified number of amniotic sacs, third tri | 34 |
| DX_ICD10 | O30.203 | quadruplet preg, unspec # placenta & unspecified number of amniotic sacs, third tri | 34 |
| DX_ICD10 | P07.18 | other low birth weight newborn, 2000-2499 grams | 34 |
| DX_ICD10 | P07.37 | preterm newborn, gestational age 34 completed weeks | 34 |
| DX_ICD10 | Z3A.34 | 34 weeks gestation of pregnancy | 34 |
| DX_ICD9 | 765.09 | extreme immaturity 2500+g (begin 1988) | 35 |
| DX_ICD9 | 765.19 | preterm nec 2500+g (begin 1988) | 35 |
| DX_ICD10 | P07.30 | preterm newborn, unspecified weeks of gestation | 35 |
| DX_ICD10 | P07.38 | preterm newborn, gestational age 35 completed weeks | 35 |
| DX_ICD10 | Z3A.35 | 35 weeks gestation of pregnancy | 35 |
| DX_ICD9 | 651.01 | twin pregnancy, delivered, w or w/o- antepartum | 36 |
| DX_ICD9 | 666.00 | third-stage hem- unspecified | 36 |
| DX_ICD9 | 666.02 | third-stage hem- delivered w p/p | 36 |
| DX_ICD9 | 666.04 | third-stage hem- postpartum | 36 |
| DX_ICD9 | 678.11 | fetal conjoin twins-del (begin 2008) | 36 |
| DX_ICD9 | 765.28 | 35-36 completed wks gestation (begin 2000) | 36 |
| DX_ICD9 | V27.2 | outcome of delivery, twins, both liveborn | 36 |
| DX_ICD9 | V31.0 | twin, mate live born- in hospital | 36 |
| DX_ICD9 | V31.00 | twin birth, mate liveborn, born in hospital, delivered w/o c-section | 36 |
| DX_ICD9 | V31.01 | twin birth, mate liveborn, born in hospital, delivered by c-section | 36 |
| DX_ICD9 | V31.1 | twin birth, mate liveborn, born before admission to hospital | 36 |
| DX_ICD9 | V31.2 | twin birth, mate liveborn, born outside hospital and not hospitalized | 36 |
| DX_ICD9 | V33 | twin nos | 36 |
| DX_ICD9 | V33.0 | twin nos- in hospital | 36 |
| DX_ICD9 | V33.00 | twin birth, unspec whether mate LB or stillborn, born in hospital, delivered w/o c-section | 36 |
| DX_ICD9 | V33.01 | twin birth, unspec whether mate LB or stillborn, born in hospital, delivered by c-section | 36 |
| DX_ICD9 | V33.1 | twin birth, unspec whether mate LB or stillborn, born before admission to hospital | 36 |
| DX_ICD9 | V33.2 | twin birth, unspec whether mate LB or stillborn, born outside hosp & not hospitalized | 36 |
| PX_ICD9 | 73.21 | internal and combined version without extraction | 36 |
| PX_ICD9 | 73.91 | external version assisting delivery | 36 |
| DX_ICD10 | O09.03 | supervision of pregnancy with history of infertility, third trimester | 36 |
| DX_ICD10 | O09.13 | supervision of pregnancy with history of ectopic pregnancy, third trimester | 36 |
| DX_ICD10 | O09.213 | supervision of pregnancy with history of pre-term labor, third trimester | 36 |
| DX_ICD10 | O09.293 | supervision of preg with other poor reproductive or obstetric hx, third trimester | 36 |
| DX_ICD10 | O09.33 | supervision of pregnancy with insufficient antenatal care, third trimester | 36 |
| DX_ICD10 | O09.43 | supervision of pregnancy with grand multiparity, third trimester | 36 |
| DX_ICD10 | O09.513 | supervision of elderly primigravida, third trimester | 36 |
| DX_ICD10 | O09.523 | supervision of elderly multigravida, third trimester | 36 |
| DX_ICD10 | O09.613 | supervision of young primigravida, third trimester | 36 |
| DX_ICD10 | O09.623 | supervision of young multigravida, third trimester | 36 |
| DX_ICD10 | O09.73 | supervision of high risk pregnancy due to social problems, third trimester | 36 |
| DX_ICD10 | O09.813 | supervision of preg resulting from assisted reproductive technology, third trimester | 36 |
| DX_ICD10 | O09.823 | supervision of pregnancy with history of in utero procedure during previous preg, third tri | 36 |
| DX_ICD10 | O09.893 | supervision of other high risk pregnancies, third trimester | 36 |
| DX_ICD10 | O09.93 | supervision of high risk pregnancy, unspecified, third trimester | 36 |
| DX_ICD10 | O09.A3 | supervision of pregnancy with history of molar pregnancy, third trimester | 36 |
| DX_ICD10 | O10.013 | pre-existing essential hypertension complicating pregnancy, third trimester | 36 |
| DX_ICD10 | O10.113 | pre-existing hypertensive heart disease complicating pregnancy, third trimester | 36 |
| DX_ICD10 | O10.213 | pre-existing hypertensive chronic kidney disease complicating preg, third trimester | 36 |
| DX_ICD10 | O10.313 | pre-existing hypertensive heart & chronic kidney disease complicating preg, third tri | 36 |
| DX_ICD10 | O10.413 | pre-existing secondary hypertension complicating pregnancy, third trimester | 36 |
| DX_ICD10 | O10.913 | unspecified pre-existing hypertension complicating pregnancy, third trimester | 36 |
| DX_ICD10 | O11.3 | pre-existing hypertension with pre-eclampsia, third trimester | 36 |
| DX_ICD10 | O12.03 | gestational edema, third trimester | 36 |
| DX_ICD10 | O12.13 | gestational proteinuria, third trimester | 36 |
| DX_ICD10 | O12.23 | gestational edema with proteinuria, third trimester | 36 |
| DX_ICD10 | O13.3 | gestational [preg-induced] hypertension without significant proteinuria, third trimester | 36 |
| DX_ICD10 | O14.03 | mild to moderate pre-eclampsia, third trimester | 36 |
| DX_ICD10 | O14.13 | severe pre-eclampsia, third trimester | 36 |
| DX_ICD10 | O14.23 | hellp syndrome (hellp), third trimester | 36 |
| DX_ICD10 | O14.93 | unspecified pre-eclampsia, third trimester | 36 |
| DX_ICD10 | O15.03 | eclampsia complicating pregnancy, third trimester | 36 |
| DX_ICD10 | O16.3 | unspecified maternal hypertension, third trimester | 36 |
| DX_ICD10 | O22.03 | varicose veins of lower extremity in pregnancy, third trimester | 36 |
| DX_ICD10 | O22.13 | genital varices in pregnancy, third trimester | 36 |
| DX_ICD10 | O22.23 | superficial thrombophlebitis in pregnancy, third trimester | 36 |
| DX_ICD10 | O22.33 | deep phlebothrombosis in pregnancy, third trimester | 36 |
| DX_ICD10 | O22.43 | cerebral venous thrombosis in pregnancy, third trimester | 36 |
| DX_ICD10 | O22.53 | cerebral venous thrombosis in pregnancy, third trimester | 36 |
| DX_ICD10 | O22.8X3 | other venous complications in pregnancy, third trimester | 36 |
| DX_ICD10 | O22.93 | venous complication in pregnancy, unspecified, third trimester | 36 |
| DX_ICD10 | O23.03 | infections of kidney in pregnancy, third trimester | 36 |
| DX_ICD10 | O23.13 | infections of bladder in pregnancy, third trimester | 36 |
| DX_ICD10 | O23.23 | infections of urethra in pregnancy, third trimester | 36 |
| DX_ICD10 | O23.33 | infections of other parts of urinary tract in pregnancy, third trimester | 36 |
| DX_ICD10 | O23.43 | unspecified infection of urinary tract in pregnancy, third trimester | 36 |
| DX_ICD10 | O23.513 | infections of cervix in pregnancy, third trimester | 36 |
| DX_ICD10 | O23.523 | salpingo-oophoritis in pregnancy, third trimester | 36 |
| DX_ICD10 | O23.593 | infection of other part of genital tract in pregnancy, third trimester | 36 |
| DX_ICD10 | O23.93 | unspecified genitourinary tract infection in pregnancy, third trimester | 36 |
| DX_ICD10 | O24.013 | pre-existing type 1 diabetes mellitus, in pregnancy, third trimester | 36 |
| DX_ICD10 | O24.113 | pre-existing type 2 diabetes mellitus, in pregnancy, third trimester | 36 |
| DX_ICD10 | O24.313 | unspecified pre-existing diabetes mellitus in pregnancy, third trimester | 36 |
| DX_ICD10 | O24.813 | other pre-existing diabetes mellitus in pregnancy, third trimester | 36 |
| DX_ICD10 | O24.913 | unspecified diabetes mellitus in pregnancy, third trimester | 36 |
| DX_ICD10 | O25.13 | malnutrition in pregnancy, third trimester | 36 |
| DX_ICD10 | O26.03 | excessive weight gain in pregnancy, third trimester | 36 |
| DX_ICD10 | O26.13 | low weight gain in pregnancy, third trimester | 36 |
| DX_ICD10 | O26.23 | pregnancy care for patient with recurrent pregnancy loss, third trimester | 36 |
| DX_ICD10 | O26.33 | retained intrauterine contraceptive device in pregnancy, third trimester | 36 |
| DX_ICD10 | O26.43 | herpes gestationis, third trimester | 36 |
| DX_ICD10 | O26.53 | maternal hypotension syndrome, third trimester | 36 |
| DX_ICD10 | O26.613 | liver and biliary tract disorders in pregnancy, third trimester | 36 |
| DX_ICD10 | O26.713 | subluxation of symphysis (pubis) in pregnancy, third trimester | 36 |
| DX_ICD10 | O26.813 | pregnancy related exhaustion and fatigue, third trimester | 36 |
| DX_ICD10 | O26.823 | pregnancy related peripheral neuritis, third trimester | 36 |
| DX_ICD10 | O26.833 | pregnancy related renal disease, third trimester | 36 |
| DX_ICD10 | O26.843 | uterine size-date discrepancy, third trimester | 36 |
| DX_ICD10 | O26.853 | spotting complicating pregnancy, third trimester | 36 |
| DX_ICD10 | O26.873 | cervical shortening, third trimester | 36 |
| DX_ICD10 | O26.893 | other specified pregnancy related conditions, third trimester | 36 |
| DX_ICD10 | O26.93 | pregnancy related conditions, unspecified, third trimester | 36 |
| DX_ICD10 | O29.013 | aspiration pneumonitis due to anesthesia during pregnancy, third trimester | 36 |
| DX_ICD10 | O29.023 | pressure collapse of lung due to anesthesia during pregnancy, third trimester | 36 |
| DX_ICD10 | O29.093 | other pulmonary complications of anesthesia during pregnancy, third trimester | 36 |
| DX_ICD10 | O29.113 | cardiac arrest due to anesthesia during pregnancy, third trimester | 36 |
| DX_ICD10 | O29.123 | cardiac failure due to anesthesia during pregnancy, third trimester | 36 |
| DX_ICD10 | O29.193 | other cardiac complications of anesthesia during pregnancy, third trimester | 36 |
| DX_ICD10 | O29.213 | cerebral anoxia due to anesthesia during pregnancy, third trimester | 36 |
| DX_ICD10 | O29.293 | other CNS complications of anesthesia during pregnancy, third tri | 36 |
| DX_ICD10 | O29.3X3 | toxic reaction to local anesthesia during pregnancy, third trimester | 36 |
| DX_ICD10 | O29.43 | spinal and epidural anesthesia induced headache during pregnancy, third trimester | 36 |
| DX_ICD10 | O29.5X3 | other complications of spinal and epidural anesthesia during pregnancy, third trimester | 36 |
| DX_ICD10 | O29.63 | failed or difficult intubation for anesthesia during pregnancy, third trimester | 36 |
| DX_ICD10 | O29.8X3 | other complications of anesthesia during pregnancy, third trimester | 36 |
| DX_ICD10 | O29.93 | unspecified complication of anesthesia during pregnancy, third trimester | 36 |
| DX_ICD10 | O30.003 | twin preg, unspec # placenta & unspecified number of amniotic sacs, third trimester | 36 |
| DX_ICD10 | O30.013 | twin pregnancy, monochorionic/monoamniotic, third trimester | 36 |
| DX_ICD10 | O30.023 | conjoined twin pregnancy, third trimester | 36 |
| DX_ICD10 | O30.033 | twin pregnancy, monochorionic/diamniotic, third trimester | 36 |
| DX_ICD10 | O30.043 | twin pregnancy, dichorionic/diamniotic, third trimester | 36 |
| DX_ICD10 | O30.093 | twin preg, unable to determine # placenta and number of amniotic sacs, third trimester | 36 |
| DX_ICD10 | O30.113 | triplet pregnancy with two or more monochorionic fetuses, third trimester | 36 |
| DX_ICD10 | O30.123 | triplet pregnancy with two or more monoamniotic fetuses, third trimester | 36 |
| DX_ICD10 | O30.133 | triplet pregnancy, trichorionic/triamniotic, third trimester | 36 |
| DX_ICD10 | O30.193 | triplet pregnancy, unable to determine # placenta & # amniotic sacs, third tri | 36 |
| DX_ICD10 | O30.213 | quadruplet pregnancy with two or more monochorionic fetuses, third trimester | 36 |
| DX_ICD10 | O30.223 | quadruplet pregnancy with two or more monoamniotic fetuses, third trimester | 36 |
| DX_ICD10 | O30.233 | quadruplet pregnancy, quadrachorionic/quadra-amniotic, third trimester | 36 |
| DX_ICD10 | O30.293 | quadruplet pregnancy, unable to determine # placenta & # amniotic sacs, third tri | 36 |
| DX_ICD10 | O30.803 | other specified multiple gestation, unspec # placenta & unspec# amniotic sacs, third tri | 36 |
| DX_ICD10 | O30.813 | other spec multiple gestation with two or more monochorionic fetuses, third tri | 36 |
| DX_ICD10 | O30.823 | other spec multiple gestation with two or more monoamniotic fetuses, third tri | 36 |
| DX_ICD10 | O30.833 | other specified multiple gestation, # chorions & amnions are equal to # fetuses, third tri | 36 |
| DX_ICD10 | O30.893 | other spec multi gestation, unable to determine # placenta & # amniotic sacs, third tri | 36 |
| DX_ICD10 | O30.93 | multiple gestation, unspecified, third trimester | 36 |
| DX_ICD10 | O31.03X0 | papyraceous fetus, third trimester, not applicable or unspecified | 36 |
| DX_ICD10 | O31.03X1 | papyraceous fetus, third trimester, fetus 1 | 36 |
| DX_ICD10 | O31.03X2 | papyraceous fetus, third trimester, fetus 2 | 36 |
| DX_ICD10 | O31.03X3 | papyraceous fetus, third trimester, fetus 3 | 36 |
| DX_ICD10 | O31.03X4 | papyraceous fetus, third trimester, fetus 4 | 36 |
| DX_ICD10 | O31.03X5 | papyraceous fetus, third trimester, fetus 5 | 36 |
| DX_ICD10 | O31.03X9 | papyraceous fetus, third trimester, other fetus | 36 |
| DX_ICD10 | O31.13X0 | continuing preg after spontaneous abortion of 1 fetus+, third tri, n/a or unspec | 36 |
| DX_ICD10 | O31.13X1 | continuing preg after spontaneous abortion of one fetus or more, third trimester, fetus 1 | 36 |
| DX_ICD10 | O31.13X2 | continuing preg after spontaneous abortion of one fetus or more, third trimester, fetus 2 | 36 |
| DX_ICD10 | O31.13X3 | continuing preg after spontaneous abortion of one fetus or more, third trimester, fetus 3 | 36 |
| DX_ICD10 | O31.13X4 | continuing preg after spontaneous abortion of one fetus or more, third trimester, fetus 4 | 36 |
| DX_ICD10 | O31.13X5 | continuing preg after spontaneous abortion of one fetus or more, third trimester, fetus 5 | 36 |
| DX_ICD10 | O31.13X9 | continuing preg after spontan abortion of 1 fetus+, third tri, other fetus | 36 |
| DX_ICD10 | O31.23X0 | continuing preg after intrauterine death of 1 fetus+, third tri, n/a or unspecified | 36 |
| DX_ICD10 | O31.23X1 | continuing preg after intrauterine death of one fetus or more, third trimester, fetus 1 | 36 |
| DX_ICD10 | O31.23X2 | continuing preg after intrauterine death of 1 fetus+, third trimester, fetus 2 | 36 |
| DX_ICD10 | O31.23X3 | continuing preg after intrauterine death of 1 fetus+, third trimester, fetus 3 | 36 |
| DX_ICD10 | O31.23X4 | continuing preg after intrauterine death of 1 fetus+, third trimester, fetus 4 | 36 |
| DX_ICD10 | O31.23X5 | continuing preg after intrauterine death of 1 fetus+, third trimester, fetus 5 | 36 |
| DX_ICD10 | O31.23X9 | continuing preg after intrauterine death of 1 fetus+, third trimester, other fetus | 36 |
| DX_ICD10 | O31.33X0 | continuing preg after elective fetal reduction of 1 fetus+, third tri, n/a or unspecified | 36 |
| DX_ICD10 | O31.33X1 | continuing preg after elective fetal reduction of 1 fetus+, third trimester, fetus 1 | 36 |
| DX_ICD10 | O31.33X2 | continuing preg after elective fetal reduction of 1 fetus+, third trimester, fetus 2 | 36 |
| DX_ICD10 | O31.33X3 | continuing preg after elective fetal reduction of 1 fetus+, third trimester, fetus 3 | 36 |
| DX_ICD10 | O31.33X4 | continuing preg after elective fetal reduction of 1 fetus+, third trimester, fetus 4 | 36 |
| DX_ICD10 | O31.33X5 | continuing preg after elective fetal reduction of 1 fetus+, third trimester, fetus 5 | 36 |
| DX_ICD10 | O31.33X9 | continuing preg after elective fetal reduction of 1 fetus+, third trimester, other fetus | 36 |
| DX_ICD10 | O31.8X30 | other complications spec to multi gestation, third tri, n/a or unspec | 36 |
| DX_ICD10 | O31.8X31 | other complications specific to multiple gestation, third trimester, fetus 1 | 36 |
| DX_ICD10 | O31.8X32 | other complications specific to multiple gestation, third trimester, fetus 2 | 36 |
| DX_ICD10 | O31.8X33 | other complications specific to multiple gestation, third trimester, fetus 3 | 36 |
| DX_ICD10 | O31.8X34 | other complications specific to multiple gestation, third trimester, fetus 4 | 36 |
| DX_ICD10 | O31.8X35 | other complications specific to multiple gestation, third trimester, fetus 5 | 36 |
| DX_ICD10 | O31.8X39 | other complications specific to multiple gestation, third trimester, other fetus | 36 |
| DX_ICD10 | O34.03 | maternal care for unspecified congenital malformation of uterus, third trimester | 36 |
| DX_ICD10 | O34.13 | maternal care for benign tumor of corpus uteri, third trimester | 36 |
| DX_ICD10 | O34.33 | maternal care for cervical incompetence, third trimester | 36 |
| DX_ICD10 | O34.43 | maternal care for other abnormalities of cervix, third trimester | 36 |
| DX_ICD10 | O34.513 | maternal care for incarceration of gravid uterus, third trimester | 36 |
| DX_ICD10 | O34.523 | maternal care for prolapse of gravid uterus, third trimester | 36 |
| DX_ICD10 | O34.533 | maternal care for retroversion of gravid uterus, third trimester | 36 |
| DX_ICD10 | O34.593 | maternal care for other abnormalities of gravid uterus, third trimester | 36 |
| DX_ICD10 | O34.63 | maternal care for abnormality of vagina, third trimester | 36 |
| DX_ICD10 | O34.73 | maternal care for abnormality of vulva and perineum, third trimester | 36 |
| DX_ICD10 | O34.83 | maternal care for other abnormalities of pelvic organs, third trimester | 36 |
| DX_ICD10 | O34.93 | maternal care for abnormality of pelvic organ, unspecified, third trimester | 36 |
| DX_ICD10 | O36.0130 | maternal care for anti-d [rh] antibodies, third trimester, n/a or unspec | 36 |
| DX_ICD10 | O36.0131 | maternal care for anti-d [rh] antibodies, third trimester, fetus 1 | 36 |
| DX_ICD10 | O36.0132 | maternal care for anti-d [rh] antibodies, third trimester, fetus 2 | 36 |
| DX_ICD10 | O36.0133 | maternal care for anti-d [rh] antibodies, third trimester, fetus 3 | 36 |
| DX_ICD10 | O36.0134 | maternal care for anti-d [rh] antibodies, third trimester, fetus 4 | 36 |
| DX_ICD10 | O36.0135 | maternal care for anti-d [rh] antibodies, third trimester, fetus 5 | 36 |
| DX_ICD10 | O36.0139 | maternal care for anti-d [rh] antibodies, third trimester, oth fetus | 36 |
| DX_ICD10 | O36.0930 | maternal care for other rhesus isoimmunization, third tri, n/a or unspec | 36 |
| DX_ICD10 | O36.0931 | maternal care for other rhesus isoimmunization, third trimester, fetus 1 | 36 |
| DX_ICD10 | O36.0932 | maternal care for other rhesus isoimmunization, third trimester, fetus 2 | 36 |
| DX_ICD10 | O36.0933 | maternal care for other rhesus isoimmunization, third trimester, fetus 3 | 36 |
| DX_ICD10 | O36.0934 | maternal care for other rhesus isoimmunization, third trimester, fetus 4 | 36 |
| DX_ICD10 | O36.0935 | maternal care for other rhesus isoimmunization, third trimester, fetus 5 | 36 |
| DX_ICD10 | O36.0939 | maternal care for other rhesus isoimmunization, third trimester, oth fetus | 36 |
| DX_ICD10 | O36.1130 | maternal care for anti-a sensitization, third trimester, n/a or unspec | 36 |
| DX_ICD10 | O36.1131 | maternal care for anti-a sensitization, third trimester, fetus 1 | 36 |
| DX_ICD10 | O36.1132 | maternal care for anti-a sensitization, third trimester, fetus 2 | 36 |
| DX_ICD10 | O36.1133 | maternal care for anti-a sensitization, third trimester, fetus 3 | 36 |
| DX_ICD10 | O36.1134 | maternal care for anti-a sensitization, third trimester, fetus 4 | 36 |
| DX_ICD10 | O36.1135 | maternal care for anti-a sensitization, third trimester, fetus 5 | 36 |
| DX_ICD10 | O36.1139 | maternal care for anti-a sensitization, third trimester, oth fetus | 36 |
| DX_ICD10 | O36.1930 | maternal care for other isoimmunization, third trimester, n/a or unspec | 36 |
| DX_ICD10 | O36.1931 | maternal care for other isoimmunization, third trimester, fetus 1 | 36 |
| DX_ICD10 | O36.1932 | maternal care for other isoimmunization, third trimester, fetus 2 | 36 |
| DX_ICD10 | O36.1933 | maternal care for other isoimmunization, third trimester, fetus 3 | 36 |
| DX_ICD10 | O36.1934 | maternal care for other isoimmunization, third trimester, fetus 4 | 36 |
| DX_ICD10 | O36.1935 | maternal care for other isoimmunization, third trimester, fetus 5 | 36 |
| DX_ICD10 | O36.1939 | maternal care for other isoimmunization, third trimester, oth fetus | 36 |
| DX_ICD10 | O36.23X0 | maternal care for hydrops fetalis, third trimester, n/a or unspec | 36 |
| DX_ICD10 | O36.23X1 | maternal care for hydrops fetalis, third trimester, fetus 1 | 36 |
| DX_ICD10 | O36.23X2 | maternal care for hydrops fetalis, third trimester, fetus 2 | 36 |
| DX_ICD10 | O36.23X3 | maternal care for hydrops fetalis, third trimester, fetus 3 | 36 |
| DX_ICD10 | O36.23X4 | maternal care for hydrops fetalis, third trimester, fetus 4 | 36 |
| DX_ICD10 | O36.23X5 | maternal care for hydrops fetalis, third trimester, fetus 5 | 36 |
| DX_ICD10 | O36.23X9 | maternal care for hydrops fetalis, third trimester, oth fetus | 36 |
| DX_ICD10 | O36.5130 | maternal care for known or suspected placental insufficiency, third tri, n/a or unspec | 36 |
| DX_ICD10 | O36.5131 | maternal care for known or suspected placental insufficiency, third trimester, fetus 1 | 36 |
| DX_ICD10 | O36.5132 | maternal care for known or suspected placental insufficiency, third trimester, fetus 2 | 36 |
| DX_ICD10 | O36.5133 | maternal care for known or suspected placental insufficiency, third trimester, fetus 3 | 36 |
| DX_ICD10 | O36.5134 | maternal care for known or suspected placental insufficiency, third trimester, fetus 4 | 36 |
| DX_ICD10 | O36.5135 | maternal care for known or suspected placental insufficiency, third trimester, fetus 5 | 36 |
| DX_ICD10 | O36.5139 | maternal care for known or suspected placental insufficiency, third trimester, oth fetus | 36 |
| DX_ICD10 | O36.5930 | maternal care for other known or suspected poor fetal growth, third tri, n/a or unspec | 36 |
| DX_ICD10 | O36.5931 | maternal care for other known or suspected poor fetal growth, third trimester, fetus 1 | 36 |
| DX_ICD10 | O36.5932 | maternal care for other known or suspected poor fetal growth, third trimester, fetus 2 | 36 |
| DX_ICD10 | O36.5933 | maternal care for other known or suspected poor fetal growth, third trimester, fetus 3 | 36 |
| DX_ICD10 | O36.5934 | maternal care for other known or suspected poor fetal growth, third trimester, fetus 4 | 36 |
| DX_ICD10 | O36.5935 | maternal care for other known or suspected poor fetal growth, third trimester, fetus 5 | 36 |
| DX_ICD10 | O36.5939 | maternal care for other known or suspected poor fetal growth, third trimester, oth fetus | 36 |
| DX_ICD10 | O36.63X0 | maternal care for excessive fetal growth, third trimester, n/a or unspec | 36 |
| DX_ICD10 | O36.63X1 | maternal care for excessive fetal growth, third trimester, fetus 1 | 36 |
| DX_ICD10 | O36.63X2 | maternal care for excessive fetal growth, third trimester, fetus 2 | 36 |
| DX_ICD10 | O36.63X3 | maternal care for excessive fetal growth, third trimester, fetus 3 | 36 |
| DX_ICD10 | O36.63X4 | maternal care for excessive fetal growth, third trimester, fetus 4 | 36 |
| DX_ICD10 | O36.63X5 | maternal care for excessive fetal growth, third trimester, fetus 5 | 36 |
| DX_ICD10 | O36.63X9 | maternal care for excessive fetal growth, third trimester, other fetus | 36 |
| DX_ICD10 | O36.73X0 | maternal care for viable fetus in abdominal pregnancy, third trimester, n/a or unspec | 36 |
| DX_ICD10 | O36.73X1 | maternal care for viable fetus in abdominal pregnancy, third trimester, fetus 1 | 36 |
| DX_ICD10 | O36.73X2 | maternal care for viable fetus in abdominal pregnancy, third trimester, fetus 2 | 36 |
| DX_ICD10 | O36.73X3 | maternal care for viable fetus in abdominal pregnancy, third trimester, fetus 3 | 36 |
| DX_ICD10 | O36.73X4 | maternal care for viable fetus in abdominal pregnancy, third trimester, fetus 4 | 36 |
| DX_ICD10 | O36.73X5 | maternal care for viable fetus in abdominal pregnancy, third trimester, fetus 5 | 36 |
| DX_ICD10 | O36.73X9 | maternal care for viable fetus in abdominal pregnancy, third trimester, oth fetus | 36 |
| DX_ICD10 | O36.8130 | decreased fetal movements, third trimester, n/a or unspec | 36 |
| DX_ICD10 | O36.8131 | decreased fetal movements, third trimester, fetus 1 | 36 |
| DX_ICD10 | O36.8132 | decreased fetal movements, third trimester, fetus 2 | 36 |
| DX_ICD10 | O36.8133 | decreased fetal movements, third trimester, fetus 3 | 36 |
| DX_ICD10 | O36.8134 | decreased fetal movements, third trimester, fetus 4 | 36 |
| DX_ICD10 | O36.8135 | decreased fetal movements, third trimester, fetus 5 | 36 |
| DX_ICD10 | O36.8139 | decreased fetal movements, third trimester, other fetus | 36 |
| DX_ICD10 | O36.8230 | fetal anemia and thrombocytopenia, third trimester, n/a or unspec | 36 |
| DX_ICD10 | O36.8231 | fetal anemia and thrombocytopenia, third trimester, fetus 1 | 36 |
| DX_ICD10 | O36.8232 | fetal anemia and thrombocytopenia, third trimester, fetus 2 | 36 |
| DX_ICD10 | O36.8233 | fetal anemia and thrombocytopenia, third trimester, fetus 3 | 36 |
| DX_ICD10 | O36.8234 | fetal anemia and thrombocytopenia, third trimester, fetus 4 | 36 |
| DX_ICD10 | O36.8235 | fetal anemia and thrombocytopenia, third trimester, fetus 5 | 36 |
| DX_ICD10 | O36.8239 | fetal anemia and thrombocytopenia, third trimester, oth fetus | 36 |
| DX_ICD10 | O36.8330 | maternal care for abnormalities of fetal hrt rate or rhythm, third trimester, n/a or unspec | 36 |
| DX_ICD10 | O36.8331 | maternal care for abnormalities of fetal hrt rate or rhythm, third trimester, fetus 1 | 36 |
| DX_ICD10 | O36.8332 | maternal care for abnormalities of fetal hrt rate or rhythm, third trimester, fetus 2 | 36 |
| DX_ICD10 | O36.8333 | maternal care for abnormalities of fetal hrt rate or rhythm, third trimester, fetus 3 | 36 |
| DX_ICD10 | O36.8334 | maternal care for abnormalities of fetal hrt rate or rhythm, third trimester, fetus 4 | 36 |
| DX_ICD10 | O36.8335 | maternal care for abnormalities of fetal hrt rate or rhythm, third trimester, fetus 5 | 36 |
| DX_ICD10 | O36.8339 | maternal care for abnormalities of fetal hrt rate or rhythm, third trimester, oth fetus | 36 |
| DX_ICD10 | O36.8930 | maternal care for other spec fetal problems, third trimester, n/a or unspec | 36 |
| DX_ICD10 | O36.8931 | maternal care for other specified fetal problems, third trimester, fetus 1 | 36 |
| DX_ICD10 | O36.8932 | maternal care for other specified fetal problems, third trimester, fetus 2 | 36 |
| DX_ICD10 | O36.8933 | maternal care for other specified fetal problems, third trimester, fetus 3 | 36 |
| DX_ICD10 | O36.8934 | maternal care for other specified fetal problems, third trimester, fetus 4 | 36 |
| DX_ICD10 | O36.8935 | maternal care for other specified fetal problems, third trimester, fetus 5 | 36 |
| DX_ICD10 | O36.8939 | maternal care for other specified fetal problems, third trimester, oth fetus | 36 |
| DX_ICD10 | O36.93X0 | maternal care for fetal problem, unspecified, third trimester, n/a or unspec | 36 |
| DX_ICD10 | O36.93X1 | maternal care for fetal problem, unspecified, third trimester, fetus 1 | 36 |
| DX_ICD10 | O36.93X2 | maternal care for fetal problem, unspecified, third trimester, fetus 2 | 36 |
| DX_ICD10 | O36.93X3 | maternal care for fetal problem, unspecified, third trimester, fetus 3 | 36 |
| DX_ICD10 | O36.93X4 | maternal care for fetal problem, unspecified, third trimester, fetus 4 | 36 |
| DX_ICD10 | O36.93X5 | maternal care for fetal problem, unspecified, third trimester, fetus 5 | 36 |
| DX_ICD10 | O36.93X9 | maternal care for fetal problem, unspecified, third trimester, oth fetus | 36 |
| DX_ICD10 | O40.3XX0 | polyhydramnios, third trimester, n/a or unspec | 36 |
| DX_ICD10 | O40.3XX1 | polyhydramnios, third trimester, fetus 1 | 36 |
| DX_ICD10 | O40.3XX2 | polyhydramnios, third trimester, fetus 2 | 36 |
| DX_ICD10 | O40.3XX3 | polyhydramnios, third trimester, fetus 3 | 36 |
| DX_ICD10 | O40.3XX4 | polyhydramnios, third trimester, fetus 4 | 36 |
| DX_ICD10 | O40.3XX5 | polyhydramnios, third trimester, fetus 5 | 36 |
| DX_ICD10 | O40.3XX9 | polyhydramnios, third trimester, oth fetus | 36 |
| DX_ICD10 | O41.03X0 | oligohydramnios, third trimester, n/a or unspec | 36 |
| DX_ICD10 | O41.03X1 | oligohydramnios, third trimester, fetus 1 | 36 |
| DX_ICD10 | O41.03X2 | oligohydramnios, third trimester, fetus 2 | 36 |
| DX_ICD10 | O41.03X3 | oligohydramnios, third trimester, fetus 3 | 36 |
| DX_ICD10 | O41.03X4 | oligohydramnios, third trimester, fetus 4 | 36 |
| DX_ICD10 | O41.03X5 | oligohydramnios, third trimester, fetus 5 | 36 |
| DX_ICD10 | O41.03X9 | oligohydramnios, third trimester, oth fetus | 36 |
| DX_ICD10 | O41.1030 | infection of amniotic sac and membranes, unspec, third tri, n/a or unspec | 36 |
| DX_ICD10 | O41.1031 | infection of amniotic sac and membranes, unspecified, third trimester, fetus 1 | 36 |
| DX_ICD10 | O41.1032 | infection of amniotic sac and membranes, unspecified, third trimester, fetus 2 | 36 |
| DX_ICD10 | O41.1033 | infection of amniotic sac and membranes, unspecified, third trimester, fetus 3 | 36 |
| DX_ICD10 | O41.1034 | infection of amniotic sac and membranes, unspecified, third trimester, fetus 4 | 36 |
| DX_ICD10 | O41.1035 | infection of amniotic sac and membranes, unspecified, third trimester, fetus 5 | 36 |
| DX_ICD10 | O41.1039 | infection of amniotic sac and membranes, unspecified, third trimester, oth fetus | 36 |
| DX_ICD10 | O41.1230 | chorioamnionitis, third trimester, n/a or unspec | 36 |
| DX_ICD10 | O41.1231 | chorioamnionitis, third trimester, fetus 1 | 36 |
| DX_ICD10 | O41.1232 | chorioamnionitis, third trimester, fetus 2 | 36 |
| DX_ICD10 | O41.1233 | chorioamnionitis, third trimester, fetus 3 | 36 |
| DX_ICD10 | O41.1234 | chorioamnionitis, third trimester, fetus 4 | 36 |
| DX_ICD10 | O41.1235 | chorioamnionitis, third trimester, fetus 5 | 36 |
| DX_ICD10 | O41.1239 | chorioamnionitis, third trimester, oth fetus | 36 |
| DX_ICD10 | O41.1430 | placentitis, third trimester, n/a or unspec | 36 |
| DX_ICD10 | O41.1431 | placentitis, third trimester, fetus 1 | 36 |
| DX_ICD10 | O41.1432 | placentitis, third trimester, fetus 2 | 36 |
| DX_ICD10 | O41.1433 | placentitis, third trimester, fetus 3 | 36 |
| DX_ICD10 | O41.1434 | placentitis, third trimester, fetus 4 | 36 |
| DX_ICD10 | O41.1435 | placentitis, third trimester, fetus 5 | 36 |
| DX_ICD10 | O41.1439 | placentitis, third trimester, oth fetus | 36 |
| DX_ICD10 | O41.8X30 | other specified disorders of amniotic fluid and membranes, third tri, n/a or unspec | 36 |
| DX_ICD10 | O41.8X31 | other specified disorders of amniotic fluid and membranes, third trimester, fetus 1 | 36 |
| DX_ICD10 | O41.8X32 | other specified disorders of amniotic fluid and membranes, third trimester, fetus 2 | 36 |
| DX_ICD10 | O41.8X33 | other specified disorders of amniotic fluid and membranes, third trimester, fetus 3 | 36 |
| DX_ICD10 | O41.8X34 | other specified disorders of amniotic fluid and membranes, third trimester, fetus 4 | 36 |
| DX_ICD10 | O41.8X35 | other specified disorders of amniotic fluid and membranes, third trimester, fetus 5 | 36 |
| DX_ICD10 | O41.8X39 | other specified disorders of amniotic fluid and membranes, third trimester, oth fetus | 36 |
| DX_ICD10 | O41.93X0 | disorder of amniotic fluid and membranes, unspec, third tri, n/a or unspecified | 36 |
| DX_ICD10 | O41.93X1 | disorder of amniotic fluid and membranes, unspecified, third trimester, fetus 1 | 36 |
| DX_ICD10 | O41.93X2 | disorder of amniotic fluid and membranes, unspecified, third trimester, fetus 2 | 36 |
| DX_ICD10 | O41.93X3 | disorder of amniotic fluid and membranes, unspecified, third trimester, fetus 3 | 36 |
| DX_ICD10 | O41.93X4 | disorder of amniotic fluid and membranes, unspecified, third trimester, fetus 4 | 36 |
| DX_ICD10 | O41.93X5 | disorder of amniotic fluid and membranes, unspecified, third trimester, fetus 5 | 36 |
| DX_ICD10 | O41.93X9 | disorder of amniotic fluid and membranes, unspecified, third trimester, oth fetus | 36 |
| DX_ICD10 | O42.013 | preterm premature rupture of membranes, onset of labor w in 24 hrs of rupture, third tri | 36 |
| DX_ICD10 | O42.113 | preterm premature rupture of membranes, onset of labor 24+ hrs after rupture, third tri | 36 |
| DX_ICD10 | O42.913 | preterm premat rupt membranes, unspec to length time btwn rupt & labor labor, 3^rd^ tri | 36 |
| DX_ICD10 | O43.013 | fetomaternal placental transfusion syndrome, third trimester | 36 |
| DX_ICD10 | O43.023 | fetus-to-fetus placental transfusion syndrome, third trimester | 36 |
| DX_ICD10 | O43.103 | malformation of placenta, unspecified, third trimester | 36 |
| DX_ICD10 | O43.113 | circumvallate placenta, third trimester | 36 |
| DX_ICD10 | O43.123 | velamentous insertion of umbilical cord, third trimester | 36 |
| DX_ICD10 | O43.193 | other malformation of placenta, third trimester | 36 |
| DX_ICD10 | O43.213 | placenta accreta, third trimester | 36 |
| DX_ICD10 | O43.223 | placenta increta, third trimester | 36 |
| DX_ICD10 | O43.233 | placenta percreta, third trimester | 36 |
| DX_ICD10 | O43.813 | placental infarction, third trimester | 36 |
| DX_ICD10 | O43.893 | other placental disorders, third trimester | 36 |
| DX_ICD10 | O43.93 | unspecified placental disorder, third trimester | 36 |
| DX_ICD10 | O44.03 | complete placenta previa nos or without hemorrhage, third trimester | 36 |
| DX_ICD10 | O44.13 | complete placenta previa with hemorrhage, third trimester | 36 |
| DX_ICD10 | O44.23 | partial placenta previa nos or without hemorrhage, third trimester | 36 |
| DX_ICD10 | O44.33 | partial placenta previa with hemorrhage, third trimester | 36 |
| DX_ICD10 | O44.43 | low lying placenta nos or without hemorrhage, third trimester | 36 |
| DX_ICD10 | O44.53 | low lying placenta with hemorrhage, third trimester | 36 |
| DX_ICD10 | O45.003 | premature separation of placenta with coagulation defect, unspecified, third trimester | 36 |
| DX_ICD10 | O45.013 | premature separation of placenta with afibrinogenemia, third trimester | 36 |
| DX_ICD10 | O45.023 | premature separation of placenta with disseminated intravascular coagulation, third tri | 36 |
| DX_ICD10 | O45.093 | premature separation of placenta with other coagulation defect, third trimester | 36 |
| DX_ICD10 | O45.8X3 | other premature separation of placenta, third trimester | 36 |
| DX_ICD10 | O45.93 | premature separation of placenta, unspecified, third trimester | 36 |
| DX_ICD10 | O46.003 | antepartum hemorrhage with coagulation defect, unspecified, third trimester | 36 |
| DX_ICD10 | O46.013 | antepartum hemorrhage with afibrinogenemia, third trimester | 36 |
| DX_ICD10 | O46.023 | antepartum hemorrhage with disseminated intravascular coagulation, third trimester | 36 |
| DX_ICD10 | O46.093 | antepartum hemorrhage with other coagulation defect, third trimester | 36 |
| DX_ICD10 | O46.8X3 | other antepartum hemorrhage, third trimester | 36 |
| DX_ICD10 | O46.93 | antepartum hemorrhage, unspecified, third trimester | 36 |
| DX_ICD10 | O60.03 | preterm labor without delivery, third trimester | 36 |
| DX_ICD10 | O60.13X0 | preterm labor second trimester with preterm delivery third tri, n/a or unspec | 36 |
| DX_ICD10 | O60.13X1 | preterm labor second trimester with preterm delivery third trimester, fetus 1 | 36 |
| DX_ICD10 | O60.13X2 | preterm labor second trimester with preterm delivery third trimester, fetus 2 | 36 |
| DX_ICD10 | O60.13X3 | preterm labor second trimester with preterm delivery third trimester, fetus 3 | 36 |
| DX_ICD10 | O60.13X4 | preterm labor second trimester with preterm delivery third trimester, fetus 4 | 36 |
| DX_ICD10 | O60.13X5 | preterm labor second trimester with preterm delivery third trimester, fetus 5 | 36 |
| DX_ICD10 | O60.13X9 | preterm labor second trimester with preterm delivery third trimester, oth fetus | 36 |
| DX_ICD10 | O60.14X0 | preterm labor third trimester with preterm delivery third tri, n/a or unspec | 36 |
| DX_ICD10 | O60.14X1 | preterm labor third trimester with preterm delivery third trimester, fetus 1 | 36 |
| DX_ICD10 | O60.14X2 | preterm labor third trimester with preterm delivery third trimester, fetus 2 | 36 |
| DX_ICD10 | O60.14X3 | preterm labor third trimester with preterm delivery third trimester, fetus 3 | 36 |
| DX_ICD10 | O60.14X4 | preterm labor third trimester with preterm delivery third trimester, fetus 4 | 36 |
| DX_ICD10 | O60.14X5 | preterm labor third trimester with preterm delivery third trimester, fetus 5 | 36 |
| DX_ICD10 | O60.14X9 | preterm labor third trimester with preterm delivery third trimester, oth fetus | 36 |
| DX_ICD10 | O60.23X0 | term delivery with preterm labor, third trimester, n/a or unspec | 36 |
| DX_ICD10 | O60.23X1 | term delivery with preterm labor, third trimester, fetus 1 | 36 |
| DX_ICD10 | O60.23X2 | term delivery with preterm labor, third trimester, fetus 2 | 36 |
| DX_ICD10 | O60.23X3 | term delivery with preterm labor, third trimester, fetus 3 | 36 |
| DX_ICD10 | O60.23X4 | term delivery with preterm labor, third trimester, fetus 4 | 36 |
| DX_ICD10 | O60.23X5 | term delivery with preterm labor, third trimester, fetus 5 | 36 |
| DX_ICD10 | O60.23X9 | term delivery with preterm labor, third trimester, oth fetus | 36 |
| DX_ICD10 | O70.2 | third degree perineal laceration during delivery | 36 |
| DX_ICD10 | O70.20 | third degree perineal laceration during delivery, unspec | 36 |
| DX_ICD10 | O70.21 | third degree perineal laceration during delivery, iiia | 36 |
| DX_ICD10 | O70.22 | third degree perineal laceration during delivery, iiib | 36 |
| DX_ICD10 | O70.23 | third degree perineal laceration during delivery, iiic | 36 |
| DX_ICD10 | O70.4 | anal sphincter tear complicating delivery, not associated with third degree laceration | 36 |
| DX_ICD10 | O71.03 | rupture of uterus before onset of labor, third trimester | 36 |
| DX_ICD10 | O72.0 | third-stage hemorrhage | 36 |
| DX_ICD10 | O88.013 | air embolism in pregnancy, third trimester | 36 |
| DX_ICD10 | O88.113 | amniotic fluid embolism in pregnancy, third trimester | 36 |
| DX_ICD10 | O88.213 | thromboembolism in pregnancy, third trimester | 36 |
| DX_ICD10 | O88.313 | pyemic and septic embolism in pregnancy, third trimester | 36 |
| DX_ICD10 | O88.813 | other embolism in pregnancy, third trimester | 36 |
| DX_ICD10 | O91.013 | infection of nipple associated with pregnancy, third trimester | 36 |
| DX_ICD10 | O91.113 | abscess of breast associated with pregnancy, third trimester | 36 |
| DX_ICD10 | O91.213 | nonpurulent mastitis associated with pregnancy, third trimester | 36 |
| DX_ICD10 | O92.013 | retracted nipple associated with pregnancy, third trimester | 36 |
| DX_ICD10 | O92.113 | cracked nipple associated with pregnancy, third trimester | 36 |
| DX_ICD10 | O98.013 | tuberculosis complicating pregnancy, third trimester | 36 |
| DX_ICD10 | O98.113 | syphilis complicating pregnancy, third trimester | 36 |
| DX_ICD10 | O98.213 | gonorrhea complicating pregnancy, third trimester | 36 |
| DX_ICD10 | O98.313 | other infections w a predominantly sexual mode of transmission complic preg, third tri | 36 |
| DX_ICD10 | O98.413 | viral hepatitis complicating pregnancy, third trimester | 36 |
| DX_ICD10 | O98.513 | other viral diseases complicating pregnancy, third trimester | 36 |
| DX_ICD10 | O98.613 | protozoal diseases complicating pregnancy, third trimester | 36 |
| DX_ICD10 | O98.713 | human immunodeficiency virus [hiv] disease complicating pregnancy, third trimester | 36 |
| DX_ICD10 | O98.813 | other maternal infectious and parasitic diseases complicating pregnancy, third tri | 36 |
| DX_ICD10 | O98.913 | unspecified maternal infectious and parasitic disease complicating pregnancy, third tri | 36 |
| DX_ICD10 | O99.013 | anemia complicating pregnancy, third trimester | 36 |
| DX_ICD10 | O99.113 | oth dis blood & blood-forming organs & certain dis immune mech complic preg, third tri | 36 |
| DX_ICD10 | O99.213 | obesity complicating pregnancy, third trimester | 36 |
| DX_ICD10 | O99.283 | endocrine, nutritional and metabolic diseases complicating pregnancy, third trimester | 36 |
| DX_ICD10 | O99.313 | alcohol use complicating pregnancy, third trimester | 36 |
| DX_ICD10 | O99.323 | drug use complicating pregnancy, third trimester | 36 |
| DX_ICD10 | O99.333 | smoking (tobacco) complicating pregnancy, third trimester | 36 |
| DX_ICD10 | O99.343 | other mental disorders complicating pregnancy, third trimester | 36 |
| DX_ICD10 | O99.353 | diseases of the nervous system complicating pregnancy, third trimester | 36 |
| DX_ICD10 | O99.413 | diseases of the circulatory system complicating pregnancy, third trimester | 36 |
| DX_ICD10 | O99.513 | diseases of the respiratory system complicating pregnancy, third trimester | 36 |
| DX_ICD10 | O99.613 | diseases of the digestive system complicating pregnancy, third trimester | 36 |
| DX_ICD10 | O99.713 | diseases of the skin and subcutaneous tissue complicating pregnancy, third trimester | 36 |
| DX_ICD10 | O99.843 | bariatric surgery status complicating pregnancy, third trimester | 36 |
| DX_ICD10 | O9A.113 | malignant neoplasm complicating pregnancy, third trimester | 36 |
| DX_ICD10 | O9A.213 | injury, poisoning & certain oth consequences of external causes complicpreg, third tri | 36 |
| DX_ICD10 | O9A.313 | physical abuse complicating pregnancy, third trimester | 36 |
| DX_ICD10 | O9A.413 | sexual abuse complicating pregnancy, third trimester | 36 |
| DX_ICD10 | O9A.513 | psychological abuse complicating pregnancy, third trimester | 36 |
| DX_ICD10 | P07.39 | preterm newborn, gestational age 36 completed weeks | 36 |
| DX_ICD10 | Z34.03 | encounter for supervision of normal first pregnancy, third trimester | 36 |
| DX_ICD10 | Z34.83 | encounter for supervision of other normal pregnancy, third trimester | 36 |
| DX_ICD10 | Z34.93 | encounter for supervision of normal pregnancy, unspecified, third trimester | 36 |
| DX_ICD10 | Z37.2 | twins, both liveborn | 36 |
| DX_ICD10 | Z38.3 | twin liveborn infant, born in hospital | 36 |
| DX_ICD10 | Z38.30 | twin liveborn infant, delivered vaginally | 36 |
| DX_ICD10 | Z38.31 | twin liveborn infant, delivered by cesarean | 36 |
| DX_ICD10 | Z38.4 | twin liveborn infant, born outside hospital | 36 |
| DX_ICD10 | Z38.5 | twin liveborn infant, unspecified as to place of birth | 36 |
| DX_ICD10 | Z3A.36 | 36 weeks gestation of pregnancy | 36 |
| PX_CPT | 59412 | external cephalic version, with or without tocolysis | 36 |
| DX_ICD10 | O47.00 | false labor before 37 completed weeks of gestation, unspecified trimester | 37 |
| DX_ICD10 | O47.02 | false labor before 37 completed weeks of gestation, second trimester | 37 |
| DX_ICD10 | O47.03 | false labor before 37 completed weeks of gestation, third trimester | 37 |
| DX_ICD10 | O47.1 | false labor at or after 37 completed weeks of gestation | 37 |
| DX_ICD10 | Z3A.37 | 37 weeks gestation of pregnancy | 37 |
| DX_ICD9 | 649.81 | spon labr w plan c/s-del (begin 2011) | 38 |
| DX_ICD9 | 649.82 | lbr w plan c/s-del w p/p (begin 2011) | 38 |
| DX_ICD10 | O75.82 | onset (spontan) labor aft 37 wks gestat but b4 39 wks gestat, w del by (planned) c-sect | 38 |
| DX_ICD10 | Z3A.38 | 38 weeks gestation of pregnancy | 38 |
| DX_ICD9 | 650 | normal delivery | 39 |
| DX_ICD9 | 656.91 | fetal/placental problem nos- delivered | 39 |
| DX_ICD9 | 669.71 | cesarean delivery nos | 39 |
| DX_ICD9 | 765.29 | 37 or more completed weeks | 39 |
| DX_ICD9 | V27.0 | deliver-single liveborn | 39 |
| DX_ICD9 | V30.0 | single liveborn infant, delivered vaginally | 39 |
| DX_ICD9 | V30.00 | single liveborn born in hospital delivered without cesarean section | 39 |
| DX_ICD9 | V30.01 | single liveborn, born in hospital, delivered by cesarean section | 39 |
| DX_ICD9 | V30.1 | single liveborn, born in hospital, delivered by cesarean section | 39 |
| DX_ICD9 | V30.2 | single liveborn, born outside hospital and not hospitalized | 39 |
| DX_ICD9 | V39.0 | liveborn nos- in hospital | 39 |
| DX_ICD9 | V39.00 | liveborn, unspec if single, twin or multiple, born in hospital, delivered w/o c-section | 39 |
| DX_ICD9 | V39.01 | liveborn, unspec if single, twin or multiple, born in hospital, delivered by c-section | 39 |
| DX_ICD9 | V39.1 | liveborn, unspec whether single, twin, or multiple, born before admission to hospital | 39 |
| DX_ICD9 | V39.2 | liveborn, unspec whether single, twin or multi, born outside hosp & not hospitalized | 39 |
| PX_ICD9 | 72.0 | low forceps operation | 39 |
| PX_ICD9 | 72.1 | low forceps operation with episiotomy | 39 |
| PX_ICD9 | 72.21 | mid forceps operation with episiotomy | 39 |
| PX_ICD9 | 72.29 | other mid forceps operation | 39 |
| PX_ICD9 | 72.31 | high forceps operation with episiotomy | 39 |
| PX_ICD9 | 72.39 | other high forceps operation | 39 |
| PX_ICD9 | 72.4 | forceps rotation of fetal head | 39 |
| PX_ICD9 | 72.51 | partial breech extraction with forceps to aftercoming head | 39 |
| PX_ICD9 | 72.52 | other partial breech extraction | 39 |
| PX_ICD9 | 72.53 | total breech extraction with forceps to aftercoming head | 39 |
| PX_ICD9 | 72.6 | forceps application to aftercoming head | 39 |
| PX_ICD9 | 72.71 | vacuum extraction with episiotomy | 39 |
| PX_ICD9 | 72.79 | other vacuum extraction | 39 |
| PX_ICD9 | 72.8 | other specified instrumental delivery | 39 |
| PX_ICD9 | 72.9 | unspecified instrumental delivery | 39 |
| PX_ICD9 | 73.01 | induction of labor by artificial rupture of membranes | 39 |
| PX_ICD9 | 73.09 | other artificial rupture of membranes | 39 |
| PX_ICD9 | 73.1 | other surgical induction of labor | 39 |
| PX_ICD9 | 73.22 | internal and combined version with extraction | 39 |
| PX_ICD9 | 73.3 | failed forceps | 39 |
| PX_ICD9 | 73.4 | medical induction of labor | 39 |
| PX_ICD9 | 73.51 | manual rotation of fetal head | 39 |
| PX_ICD9 | 73.59 | other total breech extraction | 39 |
| PX_ICD9 | 73.6 | episiotomy | 39 |
| PX_ICD9 | 73.8 | operations on fetus to facilitate delivery | 39 |
| PX_ICD9 | 73.92 | replacement of prolapsed umbilical cord | 39 |
| PX_ICD9 | 73.93 | incision of cervix to assist delivery | 39 |
| PX_ICD9 | 73.94 | pubiotomy to assist delivery | 39 |
| PX_ICD9 | 73.99 | other operations assisting delivery | 39 |
| PX_ICD9 | 74.0 | classical cesarean section | 39 |
| PX_ICD9 | 74.1 | low cervical cesarean section | 39 |
| PX_ICD9 | 74.2 | extraperitoneal cesarean section | 39 |
| PX_ICD9 | 74.4 | cesarean section of other specified type | 39 |
| PX_ICD9 | 74.99 | other cesarean section of unspecified type | 39 |
| DX_ICD10 | O80 | encounter for full-term uncomplicated delivery | 39 |
| DX_ICD10 | O82 | encounter for cesarean delivery without indication | 39 |
| DX_ICD10 | Z37.0 | single live birth | 39 |
| DX_ICD10 | Z38.0 | single liveborn infant, born in hospital | 39 |
| DX_ICD10 | Z38.00 | single liveborn infant, delivered vaginally | 39 |
| DX_ICD10 | Z38.01 | single liveborn infant, delivered by cesarean | 39 |
| DX_ICD10 | Z38.1 | single liveborn infant, born outside hospital | 39 |
| DX_ICD10 | Z38.2 | single liveborn infant, unspecified as to place of birth | 39 |
| DX_ICD10 | Z3A.39 | 39 weeks gestation of pregnancy | 39 |
| PX_ICD10 | 10E0XZZ | delivery of products of conception, external approach | 39 |
| PX_CPT | 59409 | vaginal delivery only (with or without episiotomy and/or forceps); | 39 |
| PX_CPT | 59410 | vaginal delivery only (w or wo episiotomy and/or forceps); including postpartum care | 39 |
| PX_CPT | 59414 | delivery of placenta (separate procedure) | 39 |
| PX_CPT | 59514 | cesarean delivery only; | 39 |
| PX_CPT | 59515 | cesarean delivery only; including postpartum care | 39 |
| PX_CPT | 59525 | subtot or total hysterectomy after c-sect delivery (list sep in addition to code for primary | 39 |
| PX_CPT | 59612 | vaginal delivery only, after previous cesarean del (w or wo episiotomy and/or forceps); | 39 |
| PX_CPT | 59614 | vaginal delivery only, after previous c-sect delivery (w or wo episiotomy and/or forceps); | 39 |
| PX_CPT | 59620 | cesarean delivery only, after attempted vag delivery after previous cesarean delivery; | 39 |
| PX_CPT | 59622 | cesarean delivery only, following attempted vaginal delivery after previous cesarean del; | 39 |
| DX_ICD10 | Z3A.40 | 40 weeks gestation of pregnancy | 40 |
| DX_ICD10 | Z3A.41 | 41 weeks gestation of pregnancy | 41 |
| DX_ICD9 | 645.11 | post term pregnancy- delivered (begin 2000) | 42 |
| DX_ICD9 | 645.21 | prolonged pregnancy- del w or w/o antepartum (begin 2000) | 42 |
| DX_ICD9 | 645.23 | prolonged pregnancy- antepartum (begin 2000) | 42 |
| DX_ICD9 | 766.21 | post-term infant | 42 |
| DX_ICD9 | 766.22 | prolonged gestation of infant | 42 |
| DX_ICD10 | O48.1 | prolonged pregnancy | 42 |
| DX_ICD10 | P08.21 | post-term newborn | 42 |
| DX_ICD10 | P08.22 | prolonged gestation of newborn | 42 |
| DX_ICD10 | Z3A.42 | 42 weeks gestation of pregnancy | 42 |
| DX_ICD10 | Z3A.49 | greater than 42 weeks gestation of pregnancy | 43 |
