## Supplemental S4 for "Maternal and Fetal Complications Among Pregnant Women with Congenital Heart Disease"

| **Supplemental Table S4: ICD Codes to Define Four Outcomes: Maternal Complications, Fetal Complications, Fetal Death and/or Stillbirth, and Early Pregnancy Loss.** | | | |
| --- | --- | --- | --- |
| **MATERNAL COMPLICATIONS (900 codes) [21 Conditions]** | | | |
|  | **TYPE** | **CODE** | **DESCRIPTION** |
| **Amniotic Fluid Embolism (OB)** 5 ICD-9 DX 8 ICD-10 DX **13 total** | DX_ICD9 | 673.1 | amniotic embolism-unspecified |
|  | DX_ICD9 | 673.11 | amniotic embolism-delivered |
|  | DX_ICD9 | 673.12 | amniotic embolism-delivered w postpartum complication |
|  | DX_ICD9 | 673.13 | amniotic embolism-antepartum |
|  | DX_ICD9 | 673.14 | amniotic embolism-postpartum |
|  | DX_ICD10 | O88.1 | amniotic fluid embolism |
|  | DX_ICD10 | O88.11 | amniotic fluid embolism in preg |
|  | DX_ICD10 | O88.111 | amniotic fluid embolism in preg, first tri |
|  | DX_ICD10 | O88.112 | amniotic fluid embolism in preg, second tri |
|  | DX_ICD10 | O88.113 | amniotic fluid embolism in preg, third tri |
|  | DX_ICD10 | O88.119 | amniotic fluid embolism in preg, unspec tri |
|  | DX_ICD10 | O88.12 | amniotic fluid embolism in childbirth |
|  | DX_ICD10 | O88.13 | amniotic fluid embolism in the puerperium |
| **Anemia in Preg (Gestational)** 4 ICD-9 DX 9 ICD-10 DX **13 total** | DX_ICD9 | 648.20 | anemia in pregnancy-unspecified |
|  | DX_ICD9 | 648.21 | anemia-delivered |
|  | DX_ICD9 | 648.22 | anemia-delivered w postpartum complication |
|  | DX_ICD9 | 648.24 | anemia-postpartum |
|  | DX_ICD10 | O90.81 | anemia of the puerperium |
|  | DX_ICD10 | O99.0 | anemia complicating pregnancy, childbirth and the puerperium |
|  | DX_ICD10 | O99.01 | anemia complicating pregnancy |
|  | DX_ICD10 | O99.011 | anemia complicating pregnancy, first tri |
|  | DX_ICD10 | O99.012 | anemia complicating pregnancy, second tri |
|  | DX_ICD10 | O99.013 | anemia complicating pregnancy, third tri |
|  | DX_ICD10 | O99.019 | anemia complicating pregnancy, unspec tri |
|  | DX_ICD10 | O99.02 | anemia complicating childbirth |
|  | DX_ICD10 | O99.03 | anemia complicating the puerperium |
| **Anesthesia Complications (OB)** 25 ICD-9 DX 100 ICD-10 DX **125 total** | DX_ICD9 | 668.00 | pulmonary complication in delivery-unspecified |
|  | DX_ICD9 | 668.01 | pulmonary complication in delivery |
|  | DX_ICD9 | 668.02 | pulmonary complication-delivered w postpartum complication |
|  | DX_ICD9 | 668.03 | pulmonary complication-antepartum |
|  | DX_ICD9 | 668.04 | pulmonary complication-postpartum |
|  | DX_ICD9 | 668.10 | heart complication in delivery-unspecified |
|  | DX_ICD9 | 668.11 | heart complication in delivery |
|  | DX_ICD9 | 668.12 | heart complication-delivered w postpartum complication |
|  | DX_ICD9 | 668.13 | heart complic-antepartum |
|  | DX_ICD9 | 668.14 | heart complic-postpartum |
|  | DX_ICD9 | 668.20 | cns compl labor/del-unspecified |
|  | DX_ICD9 | 668.21 | cns compl lab/del-deliv |
|  | DX_ICD9 | 668.22 | cns complic-delivered w postpartum complication |
|  | DX_ICD9 | 668.23 | cns compl in del-antepartum |
|  | DX_ICD9 | 668.24 | cns compl in del-postpartum |
|  | DX_ICD9 | 668.80 | anesthesia complication delivery nec- unspecified |
|  | DX_ICD9 | 668.81 | anesthesia complication nec-delivered |
|  | DX_ICD9 | 668.82 | anesthesia complication nec-delivered w postpartum complication |
|  | DX_ICD9 | 668.83 | anesthesia complication antepartum |
|  | DX_ICD9 | 668.84 | anesthesia complication postpartum |
|  | DX_ICD9 | 668.90 | anesthesia complication delivered nos-unspecified |
|  | DX_ICD9 | 668.91 | anesthesia complication nos-delivered |
|  | DX_ICD9 | 668.92 | anesthesia complication nos-delivered w postpartum complication |
|  | DX_ICD9 | 668.93 | anesthesia complication- antepartum |
|  | DX_ICD9 | 668.94 | anesthesia complication- postpartum |
|  | DX_ICD10 | O29 | complications of anesthesia during pregnancy |
|  | DX_ICD10 | O29.0 | pulmonary complications of anesthesia during pregnancy |
|  | DX_ICD10 | O29.01 | aspiration pneumonitis due to anesthesia during pregnancy |
|  | DX_ICD10 | O29.011 | aspiration pneumonitis due to anesthesia during preg, first tri |
|  | DX_ICD10 | O29.012 | aspiration pneumonitis due to anesthesia during preg, second tri |
|  | DX_ICD10 | O29.013 | aspiration pneumonitis due to anesthesia during preg, third tri |
|  | DX_ICD10 | O29.019 | aspiration pneumonitis due to anesthesia during preg, unspec tri |
|  | DX_ICD10 | O29.02 | pressure collapse lung due to anesthesia during pregnancy |
|  | DX_ICD10 | O29.021 | pressure collapse lung due to anesthesia during preg, first tri |
|  | DX_ICD10 | O29.022 | pressure collapse lung due to anesthesia during preg, second tri |
|  | DX_ICD10 | O29.023 | pressure collapse lung due to anesthesia during preg, third tri |
|  | DX_ICD10 | O29.029 | pressure collapse lung due to anesthesia during preg, unspec tri |
|  | DX_ICD10 | O29.09 | other pulmonary complications anesthesia during pregnancy |
|  | DX_ICD10 | O29.091 | other pulmonary complications anesthesia during preg, first tri |
|  | DX_ICD10 | O29.092 | other pulmonary complications anesthesia during preg, second tri |
|  | DX_ICD10 | O29.093 | other pulmonary complications anesthesia during preg, third tri |
|  | DX_ICD10 | O29.099 | other pulmonary complications anesthesia during preg, unspec tri |
|  | DX_ICD10 | O29.1 | cardiac complications anesthesia during pregnancy |
|  | DX_ICD10 | O29.11 | cardiac arrest due to anesthesia during pregnancy |
|  | DX_ICD10 | O29.111 | cardiac arrest due to anesthesia during pregnancy, first tri |
|  | DX_ICD10 | O29.112 | cardiac arrest due to anesthesia during pregnancy, second tri |
|  | DX_ICD10 | O29.113 | cardiac arrest due to anesthesia during pregnancy, third tri |
|  | DX_ICD10 | O29.119 | cardiac arrest due to anesthesia during preg, unspec tri |
|  | DX_ICD10 | O29.12 | cardiac failure due to anesthesia during pregnancy |
|  | DX_ICD10 | O29.121 | cardiac failure due to anesthesia during preg, first tri |
|  | DX_ICD10 | O29.122 | cardiac failure due to anesthesia during preg, second tri |
|  | DX_ICD10 | O29.123 | cardiac failure due to anesthesia during preg, third tri |
|  | DX_ICD10 | O29.129 | cardiac failure due to anesthesia during preg, unspec tri |
|  | DX_ICD10 | O29.19 | other cardiac complications of anesthesia during preg |
|  | DX_ICD10 | O29.191 | other cardiac complications of anesthesia during preg, first tri |
|  | DX_ICD10 | O29.192 | other cardiac complications of anesthesia during preg, second tri |
|  | DX_ICD10 | O29.193 | other cardiac complications of anesthesia during preg, third tri |
|  | DX_ICD10 | O29.199 | other cardiac complications of anesthesia during preg, unspec tri |
|  | DX_ICD10 | O29.2 | central nervous system complications of anesthesia during preg |
|  | DX_ICD10 | O29.21 | cerebral anoxia due to anesthesia during preg |
|  | DX_ICD10 | O29.211 | cerebral anoxia due to anesthesia during preg, first tri |
|  | DX_ICD10 | O29.212 | cerebral anoxia due to anesthesia during preg, second tri |
|  | DX_ICD10 | O29.213 | cerebral anoxia due to anesthesia during preg, third tri |
|  | DX_ICD10 | O29.219 | cerebral anoxia due to anesthesia during preg, unspec tri |
|  | DX_ICD10 | O29.29 | other cns complications of anesthesia during preg |
|  | DX_ICD10 | O29.291 | other cns complications of anesthesia during preg, first tri |
|  | DX_ICD10 | O29.292 | other cns complications of anesthesia during preg, second tri |
|  | DX_ICD10 | O29.293 | other cns complications of anesthesia during preg, third tri |
|  | DX_ICD10 | O29.299 | other cns complications of anesthesia during preg, unspec tri |
|  | DX_ICD10 | O29.3 | toxic reaction to local anesthesia during preg |
|  | DX_ICD10 | O29.3X | toxic reaction to local anesthesia during preg |
|  | DX_ICD10 | O29.3X1 | toxic reaction to local anesthesia during preg, first tri |
|  | DX_ICD10 | O29.3X2 | toxic reaction to local anesthesia during preg, second tri |
|  | DX_ICD10 | O29.3X3 | toxic reaction to local anesthesia during preg, third tri |
|  | DX_ICD10 | O29.3X9 | toxic reaction to local anesthesia during preg, unspec tri |
|  | DX_ICD10 | O29.4 | spinal & epidural anesthesia induced headache during preg |
|  | DX_ICD10 | O29.40 | spinal & epidural anesthesia headache during preg, unspec tri |
|  | DX_ICD10 | O29.41 | spinal & epidural anesthesia headache during preg, first tri |
|  | DX_ICD10 | O29.42 | spinal & epidural anesthesia headache during preg, second tri |
|  | DX_ICD10 | O29.43 | spinal & epidural anesthesia headache during preg, third tri |
|  | DX_ICD10 | O29.5 | other complic of spinal & epidural anesthesia during preg |
|  | DX_ICD10 | O29.5X | other complic of spinal & epidural anesthesia during preg |
|  | DX_ICD10 | O29.5X1 | other complic of spinal & epidural anesthesia during preg, first tri |
|  | DX_ICD10 | O29.5X2 | other complic of spinal & epidural anesthesia during preg, second tri |
|  | DX_ICD10 | O29.5X3 | other complic of spinal & epidural anesthesia during preg, third tri |
|  | DX_ICD10 | O29.5X9 | other complic of spinal & epidural anesthesia during preg, unspec tri |
|  | DX_ICD10 | O29.6 | failed or difficult intubation for anesthesia during preg |
|  | DX_ICD10 | O29.60 | failed or difficult intubation for anesthesia during preg, unspec tri |
|  | DX_ICD10 | O29.61 | failed or difficult intubation for anesthesia during preg, first tri |
|  | DX_ICD10 | O29.62 | failed or difficult intubation for anesthesia during preg, second tri |
|  | DX_ICD10 | O29.63 | failed or difficult intubation for anesthesia during preg, third tri |
|  | DX_ICD10 | O29.8 | other complications of anesthesia during preg |
|  | DX_ICD10 | O29.8X | other complications of anesthesia during preg |
|  | DX_ICD10 | O29.8X1 | other complications of anesthesia during preg, first tri |
|  | DX_ICD10 | O29.8X2 | other complications of anesthesia during preg, second tri |
|  | DX_ICD10 | O29.8X3 | other complications of anesthesia during preg, third tri |
|  | DX_ICD10 | O29.8X9 | other complications of anesthesia during preg, unspec tri |
|  | DX_ICD10 | O29.9 | unspec complication of anesthesia during preg |
|  | DX_ICD10 | O29.90 | unspec complication of anesthesia during preg, unspec tri |
|  | DX_ICD10 | O29.91 | unspec complication of anesthesia during preg, first tri |
|  | DX_ICD10 | O29.92 | unspec complication of anesthesia during preg, second tri |
|  | DX_ICD10 | O29.93 | unspec complication of anesthesia during preg, third tri |
|  | DX_ICD10 | O74 | complications of anesthesia during labor and delivery |
|  | DX_ICD10 | O74.0 | aspiration pneumonitis due to anesthesia during labor and delivery |
|  | DX_ICD10 | O74.1 | other pulmonary complic of anesthesia during labor and delivery |
|  | DX_ICD10 | O74.2 | cardiac complications of anesthesia during labor and delivery |
|  | DX_ICD10 | O74.3 | cns complications of anesthesia during labor & delivery |
|  | DX_ICD10 | O74.4 | toxic reaction to local anesthesia during labor & delivery |
|  | DX_ICD10 | O74.5 | spinal & epidural anesthesia-induced headache during labor & del |
|  | DX_ICD10 | O74.6 | other complic of spinal & epidural anesthesia during labor & del |
|  | DX_ICD10 | O74.7 | failed or difficult intubation for anesthesia during labor and delivery |
|  | DX_ICD10 | O74.8 | other complications of anesthesia during labor and delivery |
|  | DX_ICD10 | O74.9 | complication of anesthesia during labor and delivery, unspec |
|  | DX_ICD10 | O89 | complications of anesthesia during the puerperium |
|  | DX_ICD10 | O89.0 | pulmonary complications of anesthesia during the puerperium |
|  | DX_ICD10 | O89.01 | aspiration pneumonitis due to anesthesia during the puerperium |
|  | DX_ICD10 | O89.09 | other pulmonary complications of anesthesia during the puerperium |
|  | DX_ICD10 | O89.1 | cardiac complications of anesthesia during the puerperium |
|  | DX_ICD10 | O89.2 | CNS complications of anesthesia during the puerperium |
|  | DX_ICD10 | O89.3 | toxic reaction to local anesthesia during the puerperium |
|  | DX_ICD10 | O89.4 | spinal & epidural anesthesia-induced headache during puerperium |
|  | DX_ICD10+ | O89.5 | other complic of spinal & epidural anesthesia during puerperium |
|  | DX_ICD10 | O89.6 | failed or difficult intubation for anesthesia during puerperium |
|  | DX_ICD10 | O89.8 | other complications of anesthesia during the puerperium |
|  | DX_ICD10 | O89.9 | complication of anesthesia during the puerperium, unspec |
| **Cervical Incompetence and Abnormal Cervix (OB)** 10 ICD-9 DX 8 ICD-10 DX **18 total** | DX_ICD9 | 654.50 | cervical incompetence preg-unspecified |
|  | DX_ICD9 | 654.51 | cervical incompetence-delivered |
|  | DX_ICD9 | 654.52 | cervical incompetence-delivered w postpartum complication |
|  | DX_ICD9 | 654.53 | cervical incompetence-antepartum |
|  | DX_ICD9 | 654.54 | cervical incompetence-postpartum |
|  | DX_ICD9 | 654.60 | abnormal cervix nec preg-unspecified |
|  | DX_ICD9 | 654.61 | abnormal cervix nec-delivered |
|  | DX_ICD9 | 654.62 | abnormal cervix nec-delivered w postpartum complication |
|  | DX_ICD9 | 654.63 | abnormal cervix nec-antepartum |
|  | DX_ICD9 | 654.64 | abnormal cervix nec-postpartum |
|  | DX_ICD10 | O34.30 | mat care for cervical incompetence, unspecified tri |
|  | DX_ICD10 | O34.31 | mat care for cervical incompetence, first tri |
|  | DX_ICD10 | O34.32 | mat care for cervical incompetence, second tri |
|  | DX_ICD10 | O34.33 | mat care for cervical incompetence, third tri |
|  | DX_ICD10 | O34.40 | mat care for other abnormalities of cervix, unspec tri |
|  | DX_ICD10 | O34.41 | mat care for other abnormalities of cervix, first tri |
|  | DX_ICD10 | O34.42 | mat care for other abnormalities of cervix, second tri |
|  | DX_ICD10 | O34.43 | mat care for other abnormalities of cervix, third tri |
| **Diabetic Complications of Preg (Gestational)** 11 ICD-9 DX 31 ICD-10 DX **42 total** | DX_ICD9 | 648.00 | diabetes in preg- unspecified |
|  | DX_ICD9 | 648.01 | diabetes-delivered |
|  | DX_ICD9 | 648.02 | Diabetes- delivered w postpartum complication |
|  | DX_ICD9 | 648.03 | diabetes- antepartum |
|  | DX_ICD9 | 648.04 | diabetes- postpartum |
|  | DX_ICD9 | 648.80 | abnormal glucose in preg- unspec |
|  | DX_ICD9 | 648.81 | abnormal glucose tolerance- delivery |
|  | DX_ICD9 | 648.82 | abnormal glucose- delivered w postpartum complication |
|  | DX_ICD9 | 648.83 | abnormal glucose-antepartum |
|  | DX_ICD9 | 648.84 | abnormal glucose-postpartum |
|  | DX_ICD9 | 775.0 | syndrome of "infant of a diabetic mother" |
|  | DX_ICD10 | O24.319 | unspec pre-existing DM in pregnancy, unspec tri |
|  | DX_ICD10 | O24.4 | gestational DM |
|  | DX_ICD10 | O24.41 | gestational DM in pregnancy |
|  | DX_ICD10 | O24.410 | gestational DM in pregnancy, diet controlled |
|  | DX_ICD10 | O24.414 | gestational DM in pregnancy, insulin controlled |
|  | DX_ICD10 | O24.415 | gestational DM in preg, controlled by oral hypoglycemic drugs |
|  | DX_ICD10 | O24.419 | gestational DM in pregnancy, unspecified control |
|  | DX_ICD10 | O24.42 | gestational DM in childbirth |
|  | DX_ICD10 | O24.420 | gestational DM in childbirth, diet controlled |
|  | DX_ICD10 | O24.424 | gestational DM in childbirth, insulin controlled |
|  | DX_ICD10 | O24.425 | gestational DM in childbirth, controlled by oral hypoglycemic drugs |
|  | DX_ICD10 | O24.429 | gestational DM in childbirth, unspec control |
|  | DX_ICD10 | O24.43 | gestational DM in the puerperium |
|  | DX_ICD10 | O24.430 | gestational DM in the puerperium, diet controlled |
|  | DX_ICD10 | O24.434 | gestational DM in the puerperium, insulin controlled |
|  | DX_ICD10 | O24.435 | gestat DM in puerperium, controlled by oral hypoglycemic drugs |
|  | DX_ICD10 | O24.439 | gestat DM in the puerperium, unspec control |
|  | DX_ICD10 | O24.9 | unspecified DM in pregnancy, childbirth & puerperium |
|  | DX_ICD10 | O24.91 | unspecified DM in pregnancy |
|  | DX_ICD10 | O24.911 | unspecified DM in pregnancy, first tri |
|  | DX_ICD10 | O24.912 | unspecified DM in pregnancy, second tri |
|  | DX_ICD10 | O24.913 | unspecified DM in pregnancy, third tri |
|  | DX_ICD10 | O24.919 | unspecified DM in preg, unspec tri |
|  | DX_ICD10 | O24.92 | unspecified DM in childbirth |
|  | DX_ICD10 | O24.93 | unspecified DM in the puerperium |
|  | DX_ICD10 | O99.81 | abnormal glucose complicating preg, childbirth and the puerperium |
|  | DX_ICD10 | O99.810 | abnormal glucose complicating pregnancy |
|  | DX_ICD10 | O99.814 | abnormal glucose complicating childbirth |
|  | DX_ICD10 | O99.815 | abnormal glucose complicating the puerperium |
|  | DX_ICD10 | P70.0 | syndrome of infant of mother with gestational diabetes |
|  | DX_ICD10 | P70.1 | syndrome of infant of a diabetic mother |
| **Hemorrhage (OB)** 24 ICD-9 DX 48 ICD-10 DX 72 total | DX_ICD9 | 640.80 | hemorrhage early pregnancy nec-unspecified |
|  | DX_ICD9 | 640.81 | hemorrhage early pregnancy nec-delivered |
|  | DX_ICD9 | 640.83 | hemorrhage early preg nec-antepartum |
|  | DX_ICD9 | 640.90 | hemorrhage early preg-unspecified |
|  | DX_ICD9 | 640.91 | hemorrhage early preg-delivered |
|  | DX_ICD9 | 640.93 | hemorrhage early preg-antepartum |
|  | DX_ICD9 | 641.30 | coag def hemorrhage-unspecified |
|  | DX_ICD9 | 641.31 | coag def hemorrhage-delivered |
|  | DX_ICD9 | 641.33 | coag def hemorrhage-antepartum |
|  | DX_ICD9 | 641.80 | antepartum hemorrhage nec-unspecified |
|  | DX_ICD9 | 641.81 | antepartum hemorrhage nec-delivered |
|  | DX_ICD9 | 641.83 | antepartum hemorrhage nec-antepartum |
|  | DX_ICD9 | 641.90 | antepartum hemorrhage nos-unspecified |
|  | DX_ICD9 | 641.91 | antepartum hemorrhage nos-delivered |
|  | DX_ICD9 | 641.93 | antepartum hemorrhage nos-antepartum |
|  | DX_ICD9 | 666.00 | third-stage hemorrhage-unspecified |
|  | DX_ICD9 | 666.02 | third-stage hemorrhage-delivered w postpartum complication |
|  | DX_ICD9 | 666.04 | third-stage hemorrhage postpartum |
|  | DX_ICD9 | 666.10 | postpartum hemorrhage nec-unspecified |
|  | DX_ICD9 | 666.12 | postpartum hemorrhage nec-delivered w postpartum complication |
|  | DX_ICD9 | 666.14 | postpartum hemorrhage nec-postpartum |
|  | DX_ICD9 | 666.20 | delay p/p hemorrhage-unspecified |
|  | DX_ICD9 | 666.22 | delay p/p hemorrhage-delivered w postpartum complication |
|  | DX_ICD9 | 666.24 | delay p/p hemorrhage-postpartum |
|  | DX_ICD10 | O20 | hemorrhage in early pregnancy |
|  | DX_ICD10 | O20.8 | other hemorrhage in early pregnancy |
|  | DX_ICD10 | O20.9 | hemorrhage in early preg, unspecified |
|  | DX_ICD10 | O44.50 | low lying placenta w hemorrhage, unspecified tri |
|  | DX_ICD10 | O44.51 | low lying placenta w hemorrhage, first tri |
|  | DX_ICD10 | O44.52 | low lying placenta w hemorrhage, second tri |
|  | DX_ICD10 | O44.53 | low lying placenta w hemorrhage, third tri |
|  | DX_ICD10 | O46 | antepartum hemorrhage, not elsewhere classified |
|  | DX_ICD10 | O46.0 | antepartum hemorrhage w coagul defect |
|  | DX_ICD10 | O46.00 | antepartum hemorrhage w coagul defect, unspecified |
|  | DX_ICD10 | O46.001 | antepartum hemorrhage w coagul defect, unspecified, first tri |
|  | DX_ICD10 | O46.002 | antepartum hemorrhage w coagul defect, unspecified, second tri |
|  | DX_ICD10 | O46.003 | antepartum hemorrhage w coagulation defect, unspecified, third tri |
|  | DX_ICD10 | O46.009 | antepartum hemorrhage w coagul defect, unspecified, unspec tri |
|  | DX_ICD10 | O46.01 | antepartum hemorrhage w afibrinogenemia |
|  | DX_ICD10 | O46.011 | antepartum hemorrhage w afibrinogenemia, first tri |
|  | DX_ICD10 | O46.012 | antepartum hemorrhage w afibrinogenemia, second tri |
|  | DX_ICD10 | O46.013 | antepartum hemorrhage w afibrinogenemia, third tri |
|  | DX_ICD10 | O46.019 | antepartum hemorrhage w afibrinogenemia, unspec tri |
|  | DX_ICD10 | O46.02 | antepartum hemorrhage w disseminated intravasc coagul |
|  | DX_ICD10 | O46.021 | antepartum hemorrhage w disseminated intravasc coagul, first tri |
|  | DX_ICD10 | O46.022 | antepartum hemorrhage w disseminated intravasc coagul, second tri |
|  | DX_ICD10 | O46.023 | antepartum hemorrhage w disseminated intravasc coagul, third tri |
|  | DX_ICD10 | O46.029 | antepartum hemorrhage w disseminated intravasc coagul, unspec tri |
|  | DX_ICD10 | O46.09 | antepartum hemorrhage w oth coagulation defect |
|  | DX_ICD10 | O46.091 | antepartum hemorrhage w oth coagulation defect, first tri |
|  | DX_ICD10 | O46.092 | antepartum hemorrhage w oth coagulation defect, second tri |
|  | DX_ICD10 | O46.093 | antepartum hemorrhage w oth coagulation defect, third tri |
|  | DX_ICD10 | O46.099 | antepartum hemorrhage w oth coagulation defect, unspec tri |
|  | DX_ICD10 | O46.8 | other antepartum hemorrhage |
|  | DX_ICD10 | O46.8X | other antepartum hemorrhage |
|  | DX_ICD10 | O46.8X1 | other antepartum hemorrhage, first tri |
|  | DX_ICD10 | O46.8X2 | other antepartum hemorrhage, second tri |
|  | DX_ICD10 | O46.8X3 | other antepartum hemorrhage, third tri |
|  | DX_ICD10 | O46.8X9 | other antepartum hemorrhage, unspec tri |
|  | DX_ICD10 | O46.9 | antepartum hemorrhage, unspecified |
|  | DX_ICD10 | O46.90 | antepartum hemorrhage, unspecified, unspecified tri |
|  | DX_ICD10 | O46.91 | antepartum hemorrhage, unspecified, first tri |
|  | DX_ICD10 | O46.92 | antepartum hemorrhage, unspecified, second tri |
|  | DX_ICD10 | O46.93 | antepartum hemorrhage, unspecified, third tri |
|  | DX_ICD10 | O67 | labor & delivery complic by intrapart hemor, not elsewhere classified |
|  | DX_ICD10 | O67.0 | intrapartum hemorrhage w coagulation defect |
|  | DX_ICD10 | O67.8 | other intrapartum hemorrhage |
|  | DX_ICD10 | O67.9 | intrapartum hemorrhage, unspecified |
|  | DX_ICD10 | O72 | postpartum hemorrhage |
|  | DX_ICD10 | O72.0 | third-stage hemorrhage |
|  | DX_ICD10 | O72.1 | other immediate postpartum hemorrhage |
|  | DX_ICD10 | O72.2 | delayed and secondary postpartum hemorrhage |
| **Hypertensive Complications of Preg (Gestational)** 2 ICD-9 DX 6 ICD-10 DX **8 total** | DX_ICD9 | 642.32 | trans hypertension-delivered w postpartum complication |
|  | DX_ICD9 | 642.34 | trans hypertension-postpartum |
|  | DX_ICD10 | O13.1 | gestational [preg-induc] hypertension wo signif proteinuria, first tri |
|  | DX_ICD10 | O13.2 | gestational [preg-induc] htn wo signif proteinuria, second tri |
|  | DX_ICD10 | O13.3 | gestational [preg-induc] htn wo significant proteinuria, third tri |
|  | DX_ICD10 | O13.4 | gestational [preg-induced] htn wo signif proteinuria, complic birth |
|  | DX_ICD10 | O13.5 | gestat [preg-induced] htn wo signif proteinuria, complic puerperium |
| **Liver and Biliary Tract Disorders of Preg (Gestational)** 3 ICD-9 DX 8 ICD-10 DX **11 total** | DX_ICD9 | 646.70 | liver dis in preg-unspecified |
|  | DX_ICD9 | 646.71 | liver disorder-delivered |
|  | DX_ICD9 | 646.73 | liver disorder-antepartum |
|  | DX_ICD10 | O26.6 | liver disorders in preg, childbirth and the puerperium |
|  | DX_ICD10 | O26.61 | liver disorders in pregnancy |
|  | DX_ICD10 | O26.611 | liver and biliary tract disorders in pregnancy, first tri |
|  | DX_ICD10 | O26.612 | liver and biliary tract disorders in pregnancy, second tri |
|  | DX_ICD10 | O26.613 | liver and biliary tract disorders in pregnancy, third tri |
|  | DX_ICD10 | O26.619 | liver and biliary tract disorders in pregnancy, unspecified tri |
|  | DX_ICD10 | O26.62 | liver and biliary tract disorders in childbirth |
|  | DX_ICD10 | O26.63 | liver and biliary tract disorders in the puerperium |
| **Malignancy (Gestational)** 9 ICD-10 DX **9 total** | DX_ICD10 | O9A | mat malig neo, inj & abuse; complic preg, childbirth puerperium |
|  | DX_ICD10 | O9A.1 | malignant neoplasm complicating preg, childbirth and the puerperium |
|  | DX_ICD10 | O9A.11 | malignant neoplasm complicating pregnancy |
|  | DX_ICD10 | O9A.111 | malignant neoplasm complicating pregnancy, first tri |
|  | DX_ICD10 | O9A.112 | malignant neoplasm complicating pregnancy, second tri |
|  | DX_ICD10 | O9A.113 | malignant neoplasm complicating pregnancy, third tri |
|  | DX_ICD10 | O9A.119 | malignant neoplasm complicating preg, unspec tri |
|  | DX_ICD10 | O9A.12 | malignant neoplasm complicating childbirth |
|  | DX_ICD10 | O9A.13 | malignant neoplasm complicating the puerperium |
| **Mental Health and Substance Abuse Disorders (Gestational)** 20 ICD-9 DX 37 ICD-10 DX **57 total** | DX_ICD9 | 648.30 | drug dependency in pregnancy unspecified |
|  | DX_ICD9 | 648.31 | drug dependency in pregnancy delivered/ antepartum |
|  | DX_ICD9 | 648.32 | drug dependency in pregnancy delivered/ postpartum |
|  | DX_ICD9 | 648.33 | drug dependency in pregnancy/antepartum |
|  | DX_ICD9 | 648.34 | drug dependency in pregnancy/postpartum |
|  | DX_ICD9 | 648.40 | mental disorders in pregnancy unspecified |
|  | DX_ICD9 | 648.41 | mental disorders in preg delivered/antepartum |
|  | DX_ICD9 | 648.42 | mental disorders of mother |
|  | DX_ICD9 | 648.43 | mental disorders in preg antepartum |
|  | DX_ICD9 | 648.44 | mental disorders in preg postpartum |
|  | DX_ICD9 | 649.00 | tobacco use disorder-unspecified (begin 2006) |
|  | DX_ICD9 | 649.01 | tobacco use disorder-delivered (begin 2006) |
|  | DX_ICD9 | 649.02 | tobacco use disorder-del-p/p (begin 2006) |
|  | DX_ICD9 | 649.03 | tobacco use disorder-antepartum (begin 2006) |
|  | DX_ICD9 | 649.04 | tobacco use disorder-postpartum (begin 2006) |
|  | DX_ICD9 | 760.72 | narcotics affecting fetus or newborn via placenta or breast milk |
|  | DX_ICD9 | 760.73 | hallucinogenic agents affecting fetus or NB via placenta or brst milk |
|  | DX_ICD9 | 760.74 | anti-infectives affecting fetus or newborn via placenta or breast milk |
|  | DX_ICD9 | 760.75 | cocaine affecting fetus or newborn via placenta or breast milk |
|  | DX_ICD9 | 779.5 | drug withdrawal syndrome in newborn |
|  | DX_ICD10 | O90.6 | postpartum mood disturbance |
|  | DX_ICD10 | O99.3 | mental dis of nervous system complic preg, childbirth puerperium |
|  | DX_ICD10 | O99.31 | alcohol use complicating preg, childbirth, and the puerperium |
|  | DX_ICD10 | O99.310 | alcohol use complicating preg, unspec tri |
|  | DX_ICD10 | O99.311 | alcohol use complicating preg, first tri |
|  | DX_ICD10 | O99.312 | alcohol use complicating preg, second tri |
|  | DX_ICD10 | O99.313 | alcohol use complicating preg, third tri |
|  | DX_ICD10 | O99.314 | alcohol use complicating childbirth |
|  | DX_ICD10 | O99.315 | alcohol use complicating the puerperium |
|  | DX_ICD10 | O99.32 | drug use complicating preg, childbirth, and the puerperium |
|  | DX_ICD10 | O99.320 | drug use complicating preg, unspec tri |
|  | DX_ICD10 | O99.321 | drug use complicating preg, first tri |
|  | DX_ICD10 | O99.322 | drug use complicating preg, second tri |
|  | DX_ICD10 | O99.323 | drug use complicating preg, third tri |
|  | DX_ICD10 | O99.324 | drug use complicating childbirth |
|  | DX_ICD10 | O99.325 | drug use complicating the puerperium |
|  | DX_ICD10 | O99.33 | smoking (tobacco) complicating preg, childbirth, and the puerperium |
|  | DX_ICD10 | O99.330 | smoking (tobacco) complicating preg, unspec tri |
|  | DX_ICD10 | O99.331 | smoking (tobacco) complicating preg, first tri |
|  | DX_ICD10 | O99.332 | smoking (tobacco) complicating preg, second tri |
|  | DX_ICD10 | O99.333 | smoking (tobacco) complicating preg, third tri |
|  | DX_ICD10 | O99.334 | smoking (tobacco) complicating childbirth |
|  | DX_ICD10 | O99.335 | smoking (tobacco) complicating the puerperium |
|  | DX_ICD10 | O99.34 | other mental disorders complic preg, childbirth and puerperium |
|  | DX_ICD10 | O99.340 | other mental disorders complicating preg, unspec tri |
|  | DX_ICD10 | O99.341 | other mental disorders complicating preg, first tri |
|  | DX_ICD10 | O99.342 | other mental disorders complicating preg, second tri |
|  | DX_ICD10 | O99.343 | other mental disorders complicating preg, third tri |
|  | DX_ICD10 | O99.344 | other mental disorders complicating childbirth |
|  | DX_ICD10 | O99.345 | other mental disorders complicating the puerperium |
|  | DX_ICD10 | P04.2 | newborn affected by mat use of tobacco |
|  | DX_ICD10 | P04.3 | newborn affected by mat use of alcohol |
|  | DX_ICD10 | P04.4 | newborn (suspected) affect by mat use of drugs of addiction |
|  | DX_ICD10 | P04.41 | newborn affected by mat use of cocaine |
|  | DX_ICD10 | P04.49 | newborn affected by mat use of other drugs of addiction |
|  | DX_ICD10 | P96.1 | neonatal withdrawal symptoms from mat use of drugs of addiction |
|  | DX_ICD10 | P96.2 | withdrawal symptoms from therapeutic use of drugs in newborn |
| **Obstetric Air Embolism (OB)** 5 ICD-9 DX 8 ICD-10 DX **13 total** | DX_ICD9 | 673.00 | obstetric air embolism-unspecified |
|  | DX_ICD9 | 673.01 | obstetric air embolism-delivered |
|  | DX_ICD9 | 673.02 | obstetric air embolism-delivered w postpartum complication |
|  | DX_ICD9 | 673.03 | obstetric air embolism-antepartum |
|  | DX_ICD9 | 673.04 | obstetric air embolism-postpartum |
|  | DX_ICD10 | O88.0 | obstetric air embolism |
|  | DX_ICD10 | O88.01 | obstetric air embolism in pregnancy |
|  | DX_ICD10 | O88.011 | air embolism in preg, first tri |
|  | DX_ICD10 | O88.012 | air embolism in preg, second tri |
|  | DX_ICD10 | O88.013 | air embolism in preg, third tri |
|  | DX_ICD10 | O88.019 | air embolism in preg, unspecified tri |
|  | DX_ICD10 | O88.02 | air embolism in childbirth |
|  | DX_ICD10 | O88.03 | air embolism in the puerperium |
| **Peripartum Cardiomyopathy (Gestational)** 5 ICD-9 DX 2 ICD-10 DX **7 total** | DX_ICD9 | 674.50 | peripartum cardiomyopathy- unspecified |
|  | DX_ICD9 | 674.51 | peripartum cardiomyopathy- unspecified |
|  | DX_ICD9 | 674.52 | peripartum cardiomyopathy- unspecified |
|  | DX_ICD9 | 674.53 | peripartum cardiomyopathy- unspecified |
|  | DX_ICD9 | 674.54 | peripartum cardiomyopathy- postpartum or complication |
|  | DX_ICD10 | O90.3 | peripartum cardiomyopathy |
|  | DX_ICD10 | O90.7 | complications of the puerperium that are not classified elsewhere |
| **Placental Insufficiency (OB)** 2 ICD-9 DX 31 ICD-10 DX **33 total** | DX_ICD9 | 656.50 | poor fetal growth- unspec |
|  | DX_ICD9 | 762.2 | abnormal placenta nec/nos affecting newborn |
|  | DX_ICD10 | O36.51 | maternal care for known or suspected placental insufficiency |
|  | DX_ICD10 | O36.511 | mat care for known or suspected placental insuff, first tri |
|  | DX_ICD10 | O36.5111 | mat care for known or suspected placental insuff, first tri, fetus 1 |
|  | DX_ICD10 | O36.5112 | mat care for known or suspected placental insuff, first tri, fetus 2 |
|  | DX_ICD10 | O36.5113 | mat care for known or suspected placental insuff, first tri, fetus 3 |
|  | DX_ICD10 | O36.5114 | mat care for known or suspected placental insuff, first tri, fetus 4 |
|  | DX_ICD10 | O36.5115 | mat care for known or suspected placental insuff, first tri, fetus 5 |
|  | DX_ICD10 | O36.5119 | mat care for known or suspected placental insuff, first tri, oth fetus |
|  | DX_ICD10 | O36.512 | mat care for known or suspected placental insuff, sec tri |
|  | DX_ICD10 | O36.5121 | mat care for known or suspected placental insuff, sec tri, fetus 1 |
|  | DX_ICD10 | O36.5122 | mat care for known or suspected placental insuff, sec tri, fetus 2 |
|  | DX_ICD10 | O36.5123 | mat care for known or suspected placental insuff, sec tri, fetus 3 |
|  | DX_ICD10 | O36.5124 | mat care for known or suspected placental insuff, sec tri, fetus 4 |
|  | DX_ICD10 | O36.5125 | mat care for known or suspected placental insuff, sec tri, fetus 5 |
|  | DX_ICD10 | O36.5129 | mat care for known or suspected placental insuff, sec tri, oth fetus |
|  | DX_ICD10 | O36.513 | mat care for known or suspected placental insuff, third tri |
|  | DX_ICD10 | O36.5131 | mat care for known or suspected placental insuff, third tri, fetus 1 |
|  | DX_ICD10 | O36.5132 | mat care for known or suspected placental insuff, third tri, fetus 2 |
|  | DX_ICD10 | O36.5133 | mat care for known or suspected placental insuff, third tri, fetus 3 |
|  | DX_ICD10 | O36.5134 | mat care for known or suspected placental insuff, third tri, fetus 4 |
|  | DX_ICD10 | O36.5135 | mat care for known or suspected placental insuff, third tri, fetus 5 |
|  | DX_ICD10 | O36.5139 | mat care for known or suspected placental insuff, third tri, oth fetus |
|  | DX_ICD10 | O36.519 | mat care for known or suspected placental insuff, unspec tri |
|  | DX_ICD10 | O36.5190 | mat care for known/suspect placental insuff, unspec tri, n/a or unspec |
|  | DX_ICD10 | O36.5191 | mat care for known or suspected placental insuff, unspec tri, fetus 1 |
|  | DX_ICD10 | O36.5192 | mat care for known or suspected placental insuff, unspec tri, fetus 2 |
|  | DX_ICD10 | O36.5193 | mat care for known or suspected placental insuff, unspec tri, fetus 3 |
|  | DX_ICD10 | O36.5194 | mat care for known or suspected placental insuff, unspec tri, fetus 4 |
|  | DX_ICD10 | O36.5195 | mat care for known or suspected placental insuff, unspec tri, fetus 5 |
|  | DX_ICD10 | O36.5199 | mat care for known/suspect placental insuff, unspec tri, oth fetus |
|  | DX_ICD10 | P02.29 | NB affect by oth morphol & functional abnormal of placenta |
| **Poly- or Oligohydramnios (OB)** 8 ICD-9 DX 69 ICD-10 DX **77 total** | DX_ICD9 | 657.00 | polyhydramnios-unspecified (begin 1991) |
|  | DX_ICD9 | 657.01 | polyhydramnios-delivered (begin 1991) |
|  | DX_ICD9 | 657.03 | polyhydramnios-antepartum (begin 1991) |
|  | DX_ICD9 | 658.00 | oligohydramnios-unspecified |
|  | DX_ICD9 | 658.01 | oligohydramnios-delivered 65801 |
|  | DX_ICD9 | 658.03 | oligohydramnios-antepartum |
|  | DX_ICD9 | 761.2 | oligohydramnios aff nb |
|  | DX_ICD9 | 761.3 | polyhydramnios aff nb |
|  | DX_ICD10 | O40 | polyhydramnios |
|  | DX_ICD10 | O40.1 | polyhydramnios, first tri |
|  | DX_ICD10 | O40.1XX0 | polyhydramnios, first tri, n/a or unspecified |
|  | DX_ICD10 | O40.1XX1 | polyhydramnios, first tri, fetus 1 |
|  | DX_ICD10 | O40.1XX2 | polyhydramnios, first tri, fetus 2 |
|  | DX_ICD10 | O40.1XX3 | polyhydramnios, first tri, fetus 3 |
|  | DX_ICD10 | O40.1XX4 | polyhydramnios, first tri, fetus 4 |
|  | DX_ICD10 | O40.1XX5 | polyhydramnios, first tri, fetus 5 |
|  | DX_ICD10 | O40.1XX9 | polyhydramnios, first tri, oth fetus |
|  | DX_ICD10 | O40.2 | polyhydramnios, second tri |
|  | DX_ICD10 | O40.2XX0 | polyhydramnios, second tri, n/a or unspec |
|  | DX_ICD10 | O40.2XX1 | polyhydramnios, second tri, fetus 1 |
|  | DX_ICD10 | O40.2XX2 | polyhydramnios, second tri, fetus 2 |
|  | DX_ICD10 | O40.2XX3 | polyhydramnios, second tri, fetus 3 |
|  | DX_ICD10 | O40.2XX4 | polyhydramnios, second tri, fetus 4 |
|  | DX_ICD10 | O40.2XX5 | polyhydramnios, second tri, fetus 5 |
|  | DX_ICD10 | O40.2XX9 | polyhydramnios, second tri, oth fetus |
|  | DX_ICD10 | O40.3 | polyhydramnios, third tri |
|  | DX_ICD10 | O40.3XX0 | polyhydramnios, third tri, n/a or unspec |
|  | DX_ICD10 | O40.3XX1 | polyhydramnios, third tri, fetus 1 |
|  | DX_ICD10 | O40.3XX2 | polyhydramnios, third tri, fetus 2 |
|  | DX_ICD10 | O40.3XX3 | polyhydramnios, third tri, fetus 3 |
|  | DX_ICD10 | O40.3XX4 | polyhydramnios, third tri, fetus 4 |
|  | DX_ICD10 | O40.3XX5 | polyhydramnios, third tri, fetus 5 |
|  | DX_ICD10 | O40.3XX9 | polyhydramnios, third tri, oth fetus |
|  | DX_ICD10 | O40.9 | polyhydramnios, unspec tri |
|  | DX_ICD10 | O40.9XX0 | polyhydramnios, unspec tri, n/a or unspec |
|  | DX_ICD10 | O40.9XX1 | polyhydramnios, unspec tri, fetus 1 |
|  | DX_ICD10 | O40.9XX2 | polyhydramnios, unspec tri, fetus 2 |
|  | DX_ICD10 | O40.9XX3 | polyhydramnios, unspec tri, fetus 3 |
|  | DX_ICD10 | O40.9XX4 | polyhydramnios, unspec tri, fetus 4 |
|  | DX_ICD10 | O40.9XX5 | polyhydramnios, unspec tri, fetus 5 |
|  | DX_ICD10 | O40.9XX9 | polyhydramnios, unspec tri, oth fetus |
|  | DX_ICD10 | O41 | other disorders of amniotic fluid & membranes |
|  | DX_ICD10 | O41.0 | oligohydramnios |
|  | DX_ICD10 | O41.00 | oligohydramnios, unspec tri |
|  | DX_ICD10 | O41.00X0 | oligohydramnios, unspec tri, n/a or unspec |
|  | DX_ICD10 | O41.00X1 | oligohydramnios, unspec tri, fetus 1 |
|  | DX_ICD10 | O41.00X2 | oligohydramnios, unspec tri, fetus 2 |
|  | DX_ICD10 | O41.00X3 | oligohydramnios, unspec tri, fetus 3 |
|  | DX_ICD10 | O41.00X4 | oligohydramnios, unspec tri, fetus 4 |
|  | DX_ICD10 | O41.00X5 | oligohydramnios, unspec tri, fetus 5 |
|  | DX_ICD10 | O41.00X9 | oligohydramnios, unspec tri, oth fetus |
|  | DX_ICD10 | O41.01 | oligohydramnios, first tri |
|  | DX_ICD10 | O41.01X0 | oligohydramnios, first tri, n/a or unspecified |
|  | DX_ICD10 | O41.01X1 | oligohydramnios, first tri, fetus 1 |
|  | DX_ICD10 | O41.01X2 | oligohydramnios, first tri, fetus 2 |
|  | DX_ICD10 | O41.01X3 | oligohydramnios, first tri, fetus 3 |
|  | DX_ICD10 | O41.01X4 | oligohydramnios, first tri, fetus 4 |
|  | DX_ICD10 | O41.01X5 | oligohydramnios, first tri, fetus 5 |
|  | DX_ICD10 | O41.01X9 | oligohydramnios, first tri, oth fetus |
|  | DX_ICD10 | O41.02 | oligohydramnios, second tri |
|  | DX_ICD10 | O41.02X0 | oligohydramnios, second tri, n/a or unspec |
|  | DX_ICD10 | O41.02X1 | oligohydramnios, second tri, fetus 1 |
|  | DX_ICD10 | O41.02X2 | oligohydramnios, second tri, fetus 2 |
|  | DX_ICD10 | O41.02X3 | oligohydramnios, second tri, fetus 3 |
|  | DX_ICD10 | O41.02X4 | oligohydramnios, second tri, fetus 4 |
|  | DX_ICD10 | O41.02X5 | oligohydramnios, second tri, fetus 5 |
|  | DX_ICD10 | O41.02X9 | oligohydramnios, second tri, oth fetus |
|  | DX_ICD10 | O41.03 | oligohydramnios, third tri |
|  | DX_ICD10 | O41.03X0 | oligohydramnios, third tri, n/a or unspec |
|  | DX_ICD10 | O41.03X1 | oligohydramnios, third tri, fetus 1 |
|  | DX_ICD10 | O41.03X2 | oligohydramnios, third tri, fetus 2 |
|  | DX_ICD10 | O41.03X3 | oligohydramnios, third tri, fetus 3 |
|  | DX_ICD10 | O41.03X4 | oligohydramnios, third tri, fetus 4 |
|  | DX_ICD10 | O41.03X5 | oligohydramnios, third tri, fetus 5 |
|  | DX_ICD10 | O41.03X9 | oligohydramnios, third tri, oth fetus |
|  | DX_ICD10 | P01.2 | newborn affected by oligohydramnios |
|  | DX_ICD10 | P01.3 | newborn affected by polyhydramnios |
| **Preeclampsia, Eclampsia, Toxemia and HELLP (OB)** 20 ICD-9 DX 37 ICD-10 DX **57 total** | DX_ICD9 | 642.40 | mild/nos preeclampsia-unspecified |
|  | DX_ICD9 | 642.41 | mild/nos preeclampsia-delivered |
|  | DX_ICD9 | 642.42 | mild preeclampsia-delivered w postpartum complication |
|  | DX_ICD9 | 642.43 | mild/nos preeclampsia-antepartum |
|  | DX_ICD9 | 642.44 | mild/nos preeclampsia-p/p |
|  | DX_ICD9 | 642.50 | severe preeclampsia-unspecified |
|  | DX_ICD9 | 642.51 | severe preeclampsia-delivered |
|  | DX_ICD9 | 642.52 | severe preeclampsia-delivered w postpartum complication |
|  | DX_ICD9 | 642.53 | severe preeclampsia-antepartum |
|  | DX_ICD9 | 642.54 | severe preeclampsia-postpartum |
|  | DX_ICD9 | 642.60 | eclampsia-unspec |
|  | DX_ICD9 | 642.61 | eclampsia-delivered |
|  | DX_ICD9 | 642.62 | eclampsia-delivered w postpartum complication |
|  | DX_ICD9 | 642.63 | eclampsia-antepartum |
|  | DX_ICD9 | 642.64 | eclampsia-postpartum |
|  | DX_ICD9 | 642.70 | toxemia w old hypertension-unspec |
|  | DX_ICD9 | 642.71 | toxemia w old hypertension-delivered |
|  | DX_ICD9 | 642.72 | toxemia w old hypertension-delivered w p/p |
|  | DX_ICD9 | 642.73 | toxemia w old hypertension-antepartum |
|  | DX_ICD9 | 642.74 | toxemia w old hypertension-postpartum |
|  | DX_ICD10 | O11.1 | pre-existing hypertension w pre-eclampsia, first tri |
|  | DX_ICD10 | O11.2 | pre-existing hypertension w pre-eclampsia, second tri |
|  | DX_ICD10 | O11.3 | pre-existing hypertension w pre-eclampsia, third tri |
|  | DX_ICD10 | O11.9 | pre-existing hypertension w pre-eclampsia, unspec tri |
|  | DX_ICD10 | O14 | gestational [preg-induced] hypertension w signif proteinuria |
|  | DX_ICD10 | O14.0 | mild pre-eclampsia |
|  | DX_ICD10 | O14.00 | mild to moderate pre-eclampsia, unspec tri |
|  | DX_ICD10 | O14.02 | mild to moderate pre-eclampsia, second tri |
|  | DX_ICD10 | O14.03 | mild to moderate pre-eclampsia, third tri |
|  | DX_ICD10 | O14.04 | mild to moderate pre-eclampsia, complicating childbirth |
|  | DX_ICD10 | O14.05 | mild to moderate pre-eclampsia, complicating the puerperium |
|  | DX_ICD10 | O14.1 | severe pre-eclampsia |
|  | DX_ICD10 | O14.10 | severe pre-eclampsia, unspec tri |
|  | DX_ICD10 | O14.12 | severe pre-eclampsia, second tri |
|  | DX_ICD10 | O14.13 | severe pre-eclampsia, third tri |
|  | DX_ICD10 | O14.14 | severe pre-eclampsia complicating childbirth |
|  | DX_ICD10 | O14.15 | severe pre-eclampsia, complicating the puerperium |
|  | DX_ICD10 | O14.2 | hellp syndrome (hellp) |
|  | DX_ICD10 | O14.20 | hellp syndrome (hellp), unspec tri |
|  | DX_ICD10 | O14.22 | hellp syndrome (hellp), second tri |
|  | DX_ICD10 | O14.23 | hellp syndrome (hellp), third tri |
|  | DX_ICD10 | O14.24 | hellp syndrome, complicating childbirth |
|  | DX_ICD10 | O14.25 | hellp syndrome, complicating the puerperium |
|  | DX_ICD10 | O14.9 | unspec pre-eclampsia |
|  | DX_ICD10 | O14.90 | unspec pre-eclampsia, unspec tri |
|  | DX_ICD10 | O14.92 | unspec pre-eclampsia, second tri |
|  | DX_ICD10 | O14.93 | unspec pre-eclampsia, third tri |
|  | DX_ICD10 | O14.94 | unspec pre-eclampsia, complicating childbirth |
|  | DX_ICD10 | O14.95 | unspec pre-eclampsia, complicating the puerperium |
|  | DX_ICD10 | O15 | eclampsia |
|  | DX_ICD10 | O15.0 | eclampsia in pregnancy |
|  | DX_ICD10 | O15.00 | eclampsia complicating pregnancy, unspecified tri |
|  | DX_ICD10 | O15.02 | eclampsia complicating pregnancy, second tri |
|  | DX_ICD10 | O15.03 | eclampsia complicating preg, third tri |
|  | DX_ICD10 | O15.1 | eclampsia complicating labor |
|  | DX_ICD10 | O15.2 | eclampsia complicating the puerperium |
|  | DX_ICD10 | O15.9 | eclampsia, unspec as to time period |
| **Preg Related Infections (OB)** 7 ICD-9 DX 110 ICD-10 DX **117 total** | DX_ICD9 | 639.0 | post abortion gu infection |
|  | DX_ICD9 | 670.00 | major puerp infect-unspecified (begin 1991) |
|  | DX_ICD9 | 670.02 | major puerp inf-delivered w postpartum complication (begin 1991) |
|  | DX_ICD9 | 670.04 | major puerp inf-postpart (begin 1991) |
|  | DX_ICD9 | 674.30 | ob surg compl nec-unspecified |
|  | DX_ICD9 | 674.32 | ob surg compl-delivered w postpartum complication |
|  | DX_ICD9 | 674.34 | ob surg comp nec-postpartum |
|  | DX_ICD10 | A34 | obstetrical tetanus |
|  | DX_ICD10 | O41.1 | infection of amnio sac & membranes |
|  | DX_ICD10 | O41.10 | infection of amnio sac & memb, unspec |
|  | DX_ICD10 | O41.101 | infection of amnio sac & memb, unspec, first tri |
|  | DX_ICD10 | O41.1010 | infection of amnio sac & memb, unspec, first tri, n/a or unspec |
|  | DX_ICD10 | O41.1011 | infection of amnio sac & memb, unspec, first tri, fetus 1 |
|  | DX_ICD10 | O41.1012 | infection of amnio sac & memb, unspec, first tri, fetus 2 |
|  | DX_ICD10 | O41.1013 | infection of amnio sac & memb, unspec, first tri, fetus 3 |
|  | DX_ICD10 | O41.1014 | infection of amnio sac & memb, unspec, first tri, fetus 4 |
|  | DX_ICD10 | O41.1015 | infection of amnio sac & memb, unspec, first tri, fetus 5 |
|  | DX_ICD10 | O41.1019 | infection of amnio sac & memb, unspec, first tri, oth fetus |
|  | DX_ICD10 | O41.102 | infection of amnio sac & memb, unspec, second tri |
|  | DX_ICD10 | O41.1020 | infect of amnio sac & memb, unspec, second tri, n/a or unspec |
|  | DX_ICD10 | O41.1021 | infection of amnio sac & memb, unspec, second tri, fetus 1 |
|  | DX_ICD10 | O41.1022 | infection of amnio sac & memb, unspec, second tri, fetus 2 |
|  | DX_ICD10 | O41.1023 | infection of amnio sac & memb, unspec, second tri, fetus 3 |
|  | DX_ICD10 | O41.1024 | infection of amnio sac & memb, unspec, second tri, fetus 4 |
|  | DX_ICD10 | O41.1025 | infection of amnio sac & memb, unspec, second tri, fetus 5 |
|  | DX_ICD10 | O41.1029 | infection of amnio sac & memb, unspec, second tri, oth fetus |
|  | DX_ICD10 | O41.103 | infection of amnio sac & memb, unspec, third tri |
|  | DX_ICD10 | O41.1030 | infection of amnio sac & memb, unspec, third tri, n/a or unspec |
|  | DX_ICD10 | O41.1031 | infection of amnio sac & memb, unspec, third tri, fetus 1 |
|  | DX_ICD10 | O41.1032 | infection of amnio sac & memb, unspec, third tri, fetus 2 |
|  | DX_ICD10 | O41.1033 | infection of amnio sac & memb, unspec, third tri, fetus 3 |
|  | DX_ICD10 | O41.1034 | infection of amnio sac & memb, unspec, third tri, fetus 4 |
|  | DX_ICD10 | O41.1035 | infection of amnio sac & memb, unspec, third tri, fetus 5 |
|  | DX_ICD10 | O41.1039 | infection of amnio sac & memb, unspec, third tri, oth fetus |
|  | DX_ICD10 | O41.109 | infection of amnio sac & memb, unspec, unspec tri |
|  | DX_ICD10 | O41.1090 | infect of amnio sac & memb, unspec, unspec tri, n/a or unspec |
|  | DX_ICD10 | O41.1091 | infection of amnio sac & memb, unspec, unspec tri, fetus 1 |
|  | DX_ICD10 | O41.1092 | infection of amnio sac & memb, unspec, unspec tri, fetus 2 |
|  | DX_ICD10 | O41.1093 | infection of amnio sac & memb, unspec, unspec tri, fetus 3 |
|  | DX_ICD10 | O41.1094 | infection of amnio sac & memb, unspec, unspec tri, fetus 4 |
|  | DX_ICD10 | O41.1095 | infection of amnio sac & memb, unspec, unspec tri, fetus 5 |
|  | DX_ICD10 | O41.1099 | infection of amnio sac & memb, unspec, unspec tri, oth fetus |
|  | DX_ICD10 | O41.12 | chorioamnionitis |
|  | DX_ICD10 | O41.121 | chorioamnionitis, first tri |
|  | DX_ICD10 | O41.1210 | chorioamnionitis, first tri, n/a or unspec |
|  | DX_ICD10 | O41.1211 | chorioamnionitis, first tri, fetus 1 |
|  | DX_ICD10 | O41.1212 | chorioamnionitis, first tri, fetus 2 |
|  | DX_ICD10 | O41.1213 | chorioamnionitis, first tri, fetus 3 |
|  | DX_ICD10 | O41.1214 | chorioamnionitis, first tri, fetus 4 |
|  | DX_ICD10 | O41.1215 | chorioamnionitis, first tri, fetus 5 |
|  | DX_ICD10 | O41.1219 | chorioamnionitis, first tri, oth fetus |
|  | DX_ICD10 | O41.122 | chorioamnionitis, second tri |
|  | DX_ICD10 | O41.1220 | chorioamnionitis, second tri, n/a or unspec |
|  | DX_ICD10 | O41.1221 | chorioamnionitis, second tri, fetus 1 |
|  | DX_ICD10 | O41.1222 | chorioamnionitis, second tri, fetus 2 |
|  | DX_ICD10 | O41.1223 | chorioamnionitis, second tri, fetus 3 |
|  | DX_ICD10 | O41.1224 | chorioamnionitis, second tri, fetus 4 |
|  | DX_ICD10 | O41.1225 | chorioamnionitis, second tri, fetus 5 |
|  | DX_ICD10 | O41.1229 | chorioamnionitis, second tri, oth fetus |
|  | DX_ICD10 | O41.123 | chorioamnionitis, second tri |
|  | DX_ICD10 | O41.1230 | chorioamnionitis, third tri, n/a or unspec |
|  | DX_ICD10 | O41.1231 | chorioamnionitis, third tri, fetus 1 |
|  | DX_ICD10 | O41.1232 | chorioamnionitis, third tri, fetus 2 |
|  | DX_ICD10 | O41.1233 | chorioamnionitis, third tri, fetus 3 |
|  | DX_ICD10 | O41.1234 | chorioamnionitis, third tri, fetus 4 |
|  | DX_ICD10 | O41.1235 | chorioamnionitis, third tri, fetus 5 |
|  | DX_ICD10 | O41.1239 | chorioamnionitis, third tri, oth fetus |
|  | DX_ICD10 | O41.129 | chorioamnionitis, unspec tri |
|  | DX_ICD10 | O41.1290 | chorioamnionitis, unspec tri, not applicable or unspec |
|  | DX_ICD10 | O41.1291 | chorioamnionitis, unspec tri, fetus 1 |
|  | DX_ICD10 | O41.1292 | chorioamnionitis, unspec tri, fetus 2 |
|  | DX_ICD10 | O41.1293 | chorioamnionitis, unspec tri, fetus 3 |
|  | DX_ICD10 | O41.1294 | chorioamnionitis, unspec tri, fetus 4 |
|  | DX_ICD10 | O41.1295 | chorioamnionitis, unspec tri, fetus 5 |
|  | DX_ICD10 | O41.1299 | chorioamnionitis, unspec tri, oth fetus |
|  | DX_ICD10 | O41.14 | placentitis |
|  | DX_ICD10 | O41.141 | placentitis, first tri |
|  | DX_ICD10 | O41.1410 | placentitis, first tri, n/a or unspec |
|  | DX_ICD10 | O41.1411 | placentitis, first tri, fetus 1 |
|  | DX_ICD10 | O41.1412 | placentitis, first tri, fetus 2 |
|  | DX_ICD10 | O41.1413 | placentitis, first tri, fetus 3 |
|  | DX_ICD10 | O41.1414 | placentitis, first tri, fetus 4 |
|  | DX_ICD10 | O41.1415 | placentitis, first tri, fetus 5 |
|  | DX_ICD10 | O41.1419 | placentitis, first tri, oth fetus |
|  | DX_ICD10 | O41.142 | placentitis, second tri |
|  | DX_ICD10 | O41.1420 | placentitis, second tri, n/a or unspec |
|  | DX_ICD10 | O41.1421 | placentitis, second tri, fetus 1 |
|  | DX_ICD10 | O41.1422 | placentitis, second tri, fetus 2 |
|  | DX_ICD10 | O41.1423 | placentitis, second tri, fetus 3 |
|  | DX_ICD10 | O41.1424 | placentitis, second tri, fetus 4 |
|  | DX_ICD10 | O41.1425 | placentitis, second tri, fetus 5 |
|  | DX_ICD10 | O41.1429 | placentitis, second tri, oth fetus |
|  | DX_ICD10 | O41.143 | placentitis, third tri |
|  | DX_ICD10 | O41.1430 | placentitis, third tri, n/a or unspec |
|  | DX_ICD10 | O41.1431 | placentitis, third tri, fetus 1 |
|  | DX_ICD10 | O41.1432 | placentitis, third tri, fetus 2 |
|  | DX_ICD10 | O41.1433 | placentitis, third tri, fetus 3 |
|  | DX_ICD10 | O41.1434 | placentitis, third tri, fetus 4 |
|  | DX_ICD10 | O41.1435 | placentitis, third tri, fetus 5 |
|  | DX_ICD10 | O41.1439 | placentitis, third tri, oth fetus |
|  | DX_ICD10 | O41.149 | placentitis, unspec tri |
|  | DX_ICD10 | O41.1490 | placentitis, unspec tri, n/a or unspec |
|  | DX_ICD10 | O41.1491 | placentitis, unspec tri, fetus 1 |
|  | DX_ICD10 | O41.1492 | placentitis, unspec tri, fetus 2 |
|  | DX_ICD10 | O41.1493 | placentitis, unspec tri, fetus 3 |
|  | DX_ICD10 | O41.1494 | placentitis, unspec tri, fetus 4 |
|  | DX_ICD10 | O41.1495 | placentitis, unspec tri, fetus 5 |
|  | DX_ICD10 | O41.1499 | placentitis, unspec tri, oth fetus |
|  | DX_ICD10 | O85 | puerperal sepsis |
|  | DX_ICD10 | O86 | other puerperal infections |
|  | DX_ICD10 | O86.0 | infection of obstetric surgical wound |
|  | DX_ICD10 | O86.00 | infection of obstetric surgical wound, unspec |
|  | DX_ICD10 | O86.01 | infection of obstetric surgical wound, superficial incisional site |
|  | DX_ICD10 | O86.02 | infection of obstetric surgical wound, deep incisional site |
|  | DX_ICD10 | O86.03 | infection of obstetric surgical wound, organ and space site |
|  | DX_ICD10 | O86.04 | sepsis following an obstetrical procedure |
|  | DX_ICD10 | O86.09 | infection of obstetric surgical wound, other surgical site |
| **Premature Rupture of Membrane (OB)** 7 ICD-9 DX 26 ICD-10 DX **33 total** | DX_ICD9 | 658.10 | premature rupture membrane- unspec |
|  | DX_ICD9 | 658.11 | premature rupture membrane- delivered |
|  | DX_ICD9 | 658.13 | premature rupture membrane- antepartum |
|  | DX_ICD9 | 658.20 | prolong rupt membrane-unspecified |
|  | DX_ICD9 | 658.21 | prolong rupt membrane-delivered |
|  | DX_ICD9 | 658.23 | prolong rup membrane-antepartum |
|  | DX_ICD9 | 761.1 | premature rupture membrane aff nb |
|  | DX_ICD10 | O42 | premature rupture of membranes |
|  | DX_ICD10 | O42.0 | premature rupture of membranes, onset of labor w in 24 hrs of rupt |
|  | DX_ICD10 | O42.00 | premat rupt membranes, labor w in 24 hrs of rupt, unspec wks gestat |
|  | DX_ICD10 | O42.01 | preterm premature rupt membranes, labor w in 24 hrs of rupture |
|  | DX_ICD10 | O42.011 | preterm premature ruptur membranes, labor w in 24 hrs rupt, first tri |
|  | DX_ICD10 | O42.012 | preterm premature rupt membranes, labor w in 24 hrs rupt, second tri |
|  | DX_ICD10 | O42.013 | preterm premature rupt membranes, labor w in 24 hrs rupt, third tri |
|  | DX_ICD10 | O42.019 | preterm premat rupt membranes, labor w in 24 hrs of rupt, unspec tri |
|  | DX_ICD10 | O42.02 | full-term premature rupt membranes, labor w in 24 hrs of rupture |
|  | DX_ICD10 | O42.1 | premature rupt membranes, labor 24+ hrs after rupture |
|  | DX_ICD10 | O42.10 | premat rupt membranes, labor 24+ hrs after rupt, unspec wks gestat |
|  | DX_ICD10 | O42.11 | preterm premat rupt membranes, labor 24+ hrs after rupture, first tri |
|  | DX_ICD10 | O42.111 | preterm premat rupt membranes, laborl 24+ hrs after rupture, first tri |
|  | DX_ICD10 | O42.112 | preterm premat rupt membranes, labor 24+ hrs after rupture, sec tri |
|  | DX_ICD10 | O42.113 | preterm premat rupt membranes, labor 24+ hrs after rupture, third tri |
|  | DX_ICD10 | O42.119 | preterm premat rupt membranes, labor 24+hrs after rupt, unspec tri |
|  | DX_ICD10 | O42.12 | full-term premat rupt membranes, labor 24+ hrs after rupt |
|  | DX_ICD10 | O42.9 | premat rupture of membranes, unspec len rupture & labor |
|  | DX_ICD10 | O42.90 | premat rupt membr len time rupt & labor, unspec wks gestat |
|  | DX_ICD10 | O42.91 | preterm premat rupt membr, unspec len rupt & labor |
|  | DX_ICD10 | O42.911 | preterm premat rupt membr, unspec len rupt &labor, first tri |
|  | DX_ICD10 | O42.912 | preterm premat rupt membr, unspec len rupt & labor, sec tri |
|  | DX_ICD10 | O42.913 | preterm premat rupt membr, unspec len rupt & labor, third tri |
|  | DX_ICD10 | O42.919 | preterm premat rupt membr, unspec len rupt & labor, unspec tri |
|  | DX_ICD10 | O42.92 | full-term premat rupt membranes, unspec len rupt & labor |
|  | DX_ICD10 | P01.1 | NB affected by premature rupture of membranes |
| **Preterm Labor, Threatened (OB)** 2 ICD-9 DX 30 ICD-10 DX **32 total** | DX_ICD9 | 644.00 | threat prem labor- unspec |
|  | DX_ICD9 | 644.03 | threat prem labor- antepartum |
|  | DX_ICD10 | O60 | preterm labor |
|  | DX_ICD10 | O60.0 | preterm labor wo delivery |
|  | DX_ICD10 | O60.00 | preterm labor wo delivery, unspec tri |
|  | DX_ICD10 | O60.02 | preterm labor wo delivery, second tri |
|  | DX_ICD10 | O60.03 | preterm labor wo delivery, third tri |
|  | DX_ICD10 | O60.2 | term delivery w preterm labor |
|  | DX_ICD10 | O60.20 | term delivery w preterm labor, unspec tri |
|  | DX_ICD10 | O60.20X0 | term delivery w preterm labor, unspec tri, n/a or unspec |
|  | DX_ICD10 | O60.20X1 | term delivery w preterm labor, unspec tri, fetus 1 |
|  | DX_ICD10 | O60.20X2 | term delivery w preterm labor, unspec tri, fetus 2 |
|  | DX_ICD10 | O60.20X3 | term delivery w preterm labor, unspec tri, fetus 3 |
|  | DX_ICD10 | O60.20X4 | term delivery w preterm labor, unspec tri, fetus 4 |
|  | DX_ICD10 | O60.20X5 | term delivery w preterm labor, unspec tri, fetus 5 |
|  | DX_ICD10 | O60.20X9 | term delivery w preterm labor, unspec tri, oth fetus |
|  | DX_ICD10 | O60.22 | term delivery with preterm labor, second tri |
|  | DX_ICD10 | O60.22X0 | term delivery with preterm labor, second tri, n/a or unspec |
|  | DX_ICD10 | O60.22X1 | term delivery with preterm labor, second tri, fetus 1 |
|  | DX_ICD10 | O60.22X2 | term delivery with preterm labor, second tri, fetus 2 |
|  | DX_ICD10 | O60.22X3 | term delivery with preterm labor, second tri, fetus 3 |
|  | DX_ICD10 | O60.22X4 | term delivery with preterm labor, second tri, fetus 4 |
|  | DX_ICD10 | O60.22X5 | term delivery with preterm labor, second tri, fetus 5 |
|  | DX_ICD10 | O60.22X9 | term delivery with preterm labor, second tri, oth fetus |
|  | DX_ICD10 | O60.23 | term delivery with preterm labor, third tri |
|  | DX_ICD10 | O60.23X0 | term delivery with preterm labor, third tri, n/a or unspec |
|  | DX_ICD10 | O60.23X1 | term delivery with preterm labor, third tri, fetus 1 |
|  | DX_ICD10 | O60.23X2 | term delivery with preterm labor, third tri, fetus 2 |
|  | DX_ICD10 | O60.23X3 | term delivery with preterm labor, third tri, fetus 3 |
|  | DX_ICD10 | O60.23X4 | term delivery with preterm labor, third tri, fetus 4 |
|  | DX_ICD10 | O60.23X5 | term delivery with preterm labor, third tri, fetus 5 |
|  | DX_ICD10 | O60.23X9 | term delivery with preterm labor, third tri, oth fetus |
| **Previa, abruption, and abnormal placental** **attachment (OB)**  16 ICD-9 DX 73 ICD-10 DX **89 total** | DX_ICD9 | 641.00 | placenta previa- unspec |
|  | DX_ICD9 | 641.01 | placenta previa- delivered |
|  | DX_ICD9 | 641.03 | placenta previa- antepartum |
|  | DX_ICD9 | 641.10 | placenta previa hem- unspec |
|  | DX_ICD9 | 641.11 | placenta previa hem- delivered |
|  | DX_ICD9 | 641.13 | placenta previa hem- antepartum |
|  | DX_ICD9 | 641.20 | prem sep placenta- unspec |
|  | DX_ICD9 | 641.21 | prem sep placenta- delivered |
|  | DX_ICD9 | 641.23 | prem sep placenta- antepartum |
|  | DX_ICD9 | 641.31 | coag def hem- delivered |
|  | DX_ICD9 | 641.33 | coag def hem- antepartum |
|  | DX_ICD9 | 666.02 | third-stage hem- delivered w p/p |
|  | DX_ICD9 | 666.04 | third-stage hem- postpartum |
|  | DX_ICD9 | 667.00 | retain placenta nos-unsp |
|  | DX_ICD9 | 667.02 | retnd plac nos-del w p/p |
|  | DX_ICD9 | 667.04 | retain plac nos-postpart |
|  | DX_ICD10 | O43.21 | placenta accreta |
|  | DX_ICD10 | O43.211 | placenta accreta, first tri |
|  | DX_ICD10 | O43.212 | placenta accreta, second tri |
|  | DX_ICD10 | O43.213 | placenta accreta, third tri |
|  | DX_ICD10 | O43.219 | placenta accreta, unspec tri |
|  | DX_ICD10 | O43.22 | placenta increta |
|  | DX_ICD10 | O43.221 | placenta increta, first tri |
|  | DX_ICD10 | O43.222 | placenta increta, second tri |
|  | DX_ICD10 | O43.223 | placenta increta, third tri |
|  | DX_ICD10 | O43.229 | placenta increta, unspec tri |
|  | DX_ICD10 | O43.23 | placenta percreta |
|  | DX_ICD10 | O43.231 | placenta percreta, first tri |
|  | DX_ICD10 | O43.232 | placenta percreta, second tri |
|  | DX_ICD10 | O43.233 | placenta percreta, third tri |
|  | DX_ICD10 | O43.239 | placenta percreta, unspec tri |
|  | DX_ICD10 | O44 | placenta previa |
|  | DX_ICD10 | O44.0 | placenta previa specif as without hemorrhage |
|  | DX_ICD10 | O44.00 | complete plcnta previa nos or wo hemorrhage, unspec tri |
|  | DX_ICD10 | O44.01 | complete placenta previa nos or without hemorrhage, first tri |
|  | DX_ICD10 | O44.02 | complete placenta previa nos or wo hemorrhage, second tri |
|  | DX_ICD10 | O44.03 | complete placenta previa nos or without hemorrhage, third tri |
|  | DX_ICD10 | O44.1 | plcnta previa with hemorrhage coagulation defect, unspec, second tri |
|  | DX_ICD10 | O44.10 | complete placenta previa with hemorrhage, unspec tri |
|  | DX_ICD10 | O44.11 | complete placenta previa with hemorrhage, first tri |
|  | DX_ICD10 | O44.12 | complete placenta previa with hemorrhage, second tri |
|  | DX_ICD10 | O44.13 | complete placenta previa with hemorrhage, third tri |
|  | DX_ICD10 | O44.20 | partial placenta previa nos or wo hemorrhage, unspec tri |
|  | DX_ICD10 | O44.21 | partial placenta previa nos or wo hemorrhage, first tri |
|  | DX_ICD10 | O44.22 | partial placenta previa nos or wo hemorrhage, second tri |
|  | DX_ICD10 | O44.23 | partial placenta previa nos or wo hemorrhage, third tri |
|  | DX_ICD10 | O44.30 | partial placenta previa w hemorrhage, unspec tri |
|  | DX_ICD10 | O44.31 | partial placenta previa w hemorrhage, first tri |
|  | DX_ICD10 | O44.32 | partial placenta previa w hemorrhage, second tri |
|  | DX_ICD10 | O44.33 | partial placenta previa w hemorrhage, third tri |
|  | DX_ICD10 | O45 | premature sep of placenta [abruptio placentae] |
|  | DX_ICD10 | O45.0 | premature sep of placenta w coagulation defect |
|  | DX_ICD10 | O45.00 | premature sep of placenta w coagulation defect, unsp |
|  | DX_ICD10 | O45.001 | premature sep of placenta with coagulation defect, unspec, first tri |
|  | DX_ICD10 | O45.002 | premature sep of placenta with coagulation defect, unspec, second tri |
|  | DX_ICD10 | O45.003 | premature sep of placenta with coagulation defect, unspec, third tri |
|  | DX_ICD10 | O45.009 | premature sep of placenta with coagulation defect, unspec, unspec tri |
|  | DX_ICD10 | O45.01 | premature sep of placenta with afibrinogenemia |
|  | DX_ICD10 | O45.011 | premature sep of placenta with afibrinogenemia, first tri |
|  | DX_ICD10 | O45.012 | premature sep of placenta with afibrinogenemia, second tri |
|  | DX_ICD10 | O45.013 | premature sep of placenta with afibrinogenemia, third tri |
|  | DX_ICD10 | O45.019 | premature sep of placenta with afibrinogenemia, unspec tri |
|  | DX_ICD10 | O45.02 | premature sep of plcnta with disseminated intravascular coagulation |
|  | DX_ICD10 | O45.021 | premature sep of plcnta w disseminated intravascular coagul, first tri |
|  | DX_ICD10 | O45.022 | premature sep of plcnta w disseminated intravasc coagul, second tri |
|  | DX_ICD10 | O45.023 | premature sep of plcnta w disseminated intravascular coagul, third tri |
|  | DX_ICD10 | O45.029 | premature sep of plcnta w disseminated intravasc coagul unspec tri |
|  | DX_ICD10 | O45.09 | premature sep of plcnta w other coagul defect |
|  | DX_ICD10 | O45.091 | premature sep of placenta w other coagul defect, first tri |
|  | DX_ICD10 | O45.092 | premature sep of placenta w other coagul defect, second tri |
|  | DX_ICD10 | O45.093 | premature sep of placenta with other coagul defect, third tri |
|  | DX_ICD10 | O45.099 | premature sep of placenta with other coagul defect, unspec tri |
|  | DX_ICD10 | O45.8 | other premature sep of placenta |
|  | DX_ICD10 | O45.8X | other premature sep of placenta |
|  | DX_ICD10 | O45.8X1 | other premature sep of placenta, first tri |
|  | DX_ICD10 | O45.8X2 | other premature sep of placenta, second tri |
|  | DX_ICD10 | O45.8X3 | other premature sep of placenta, third tri |
|  | DX_ICD10 | O45.8X9 | other premature sep of placenta, unspec tri |
|  | DX_ICD10 | O45.9 | premature sep of placenta, unspec abruptio placentae nos |
|  | DX_ICD10 | O45.90 | premature sep of placenta, unspec, unspec tri |
|  | DX_ICD10 | O45.91 | premature sep of placenta, unspec, first tri |
|  | DX_ICD10 | O45.92 | premature sep of placenta, unspec, second tri |
|  | DX_ICD10 | O45.93 | premature sep of placenta, unspec, third tri |
|  | DX_ICD10 | O69.4XX1 | labor and delivery complicated by vasa previa, fetus 1 |
|  | DX_ICD10 | O69.4XX2 | labor and delivery complicated by vasa previa, fetus 2 |
|  | DX_ICD10 | O69.4XX3 | labor and delivery complicated by vasa previa, fetus 3 |
|  | DX_ICD10 | O69.4XX4 | labor and delivery complicated by vasa previa, fetus 4 |
|  | DX_ICD10 | O69.4XX5 | labor and delivery complicated by vasa previa, fetus 5 |
|  | DX_ICD10 | O69.4XX9 | labor and delivery complicated by vasa previa, other fetus |
| **Thromboembolic Complications (DVT, PE, CVA, Thrombosis) (Gestational)** 31 ICD-9 DX 43 ICD-10 DX **74 total** | DX_ICD9 | 671.30 | deep thrombosis antepartum- unspec |
|  | DX_ICD9 | 671.31 | deep thrombosis antepartum- delivered |
|  | DX_ICD9 | 671.33 | deep vein thrombosis- antepartum |
|  | DX_ICD9 | 671.40 | deep thrombosis postpartum- unspec |
|  | DX_ICD9 | 671.42 | thrombosis postpartum- delivered w p/p |
|  | DX_ICD9 | 671.44 | deep vein thrombosis- postpartum |
|  | DX_ICD9 | 671.50 | thrombosis nec preg- unspec |
|  | DX_ICD9 | 671.51 | thrombosis nec- delivered |
|  | DX_ICD9 | 671.52 | thrombosis nec- delivered w p/p |
|  | DX_ICD9 | 671.53 | thrombosis nec- antepartum |
|  | DX_ICD9 | 671.54 | thrombosis nec- postpartum |
|  | DX_ICD9 | 671.80 | ven compl preg nec-unsp |
|  | DX_ICD9 | 671.81 | venous compl nec-deliver |
|  | DX_ICD9 | 671.82 | ven comp nec-deliv w p/p |
|  | DX_ICD9 | 671.83 | venous compl nec-antepar |
|  | DX_ICD9 | 671.84 | venous compl nec-postpar |
|  | DX_ICD9 | 673.20 | ob pulmonary embolism nos- unspec |
|  | DX_ICD9 | 673.21 | pulmonary embolism nos- delivery |
|  | DX_ICD9 | 673.22 | pulmonary embolism nos- delivery w p/p |
|  | DX_ICD9 | 673.23 | pulmonary embolism nos- antepartum |
|  | DX_ICD9 | 673.24 | pulmonary embolism nos- postpartum |
|  | DX_ICD9 | 673.30 | ob pyemic embol-unspec |
|  | DX_ICD9 | 673.31 | ob pyemic embol-deliver |
|  | DX_ICD9 | 673.32 | ob pyem embol-del w p/p |
|  | DX_ICD9 | 673.33 | ob pyemic embol-antepart |
|  | DX_ICD9 | 673.34 | ob pyemic embol-postpart |
|  | DX_ICD9 | 673.80 | pulmonary embolism nec- delivered |
|  | DX_ICD9 | 673.81 | pulmonary embolism nec- delivered w p/p |
|  | DX_ICD9 | 673.82 | pulmonary embolism nec- delivered w p/p |
|  | DX_ICD9 | 673.83 | pulmonary embolism nec- antepartum |
|  | DX_ICD9 | 673.84 | pulmonary embolism nec- postpartum |
|  | DX_ICD10 | O22.3 | deep phlebothrombosis in preg |
|  | DX_ICD10 | O22.30 | deep phlebothrombosis in preg, unspec tri |
|  | DX_ICD10 | O22.31 | deep phlebothrombosis in preg, first tri |
|  | DX_ICD10 | O22.32 | deep phlebothrombosis in preg, second tri |
|  | DX_ICD10 | O22.33 | deep phlebothrombosis in preg, third tri |
|  | DX_ICD10 | O22.4 | hemorrhoids in preg |
|  | DX_ICD10 | O22.40 | cerebral venous thrombosis in preg, unspec tri |
|  | DX_ICD10 | O22.41 | cerebral venous thrombosis in preg, first tri |
|  | DX_ICD10 | O22.42 | cerebral venous thrombosis in preg, second tri |
|  | DX_ICD10 | O22.43 | cerebral venous thrombosis in preg, third tri |
|  | DX_ICD10 | O22.5 | cerebral venous thrombosis in preg |
|  | DX_ICD10 | O22.50 | cerebral venous thrombosis in preg, unspec tri |
|  | DX_ICD10 | O22.51 | cerebral venous thrombosis in preg, first tri |
|  | DX_ICD10 | O22.52 | cerebral venous thrombosis in preg, second tri |
|  | DX_ICD10 | O22.53 | cerebral venous thrombosis in preg, third tri |
|  | DX_ICD10 | O87.1 | deep phlebothrombosis in the puerperium |
|  | DX_ICD10 | O87.2 | hemorrhoids in the puerperium |
|  | DX_ICD10 | O87.3 | cerebral venous thrombosis in the puerperium |
|  | DX_ICD10 | O88 | obstetric embolism |
|  | DX_ICD10 | O88.2 | obstetric thromboembolism |
|  | DX_ICD10 | O88.21 | thromboembolism in preg |
|  | DX_ICD10 | O88.211 | thromboembolism in preg, first tri |
|  | DX_ICD10 | O88.212 | thromboembolism in preg, second tri |
|  | DX_ICD10 | O88.213 | thromboembolism in preg, third tri |
|  | DX_ICD10 | O88.219 | thromboembolism in preg, unspec tri |
|  | DX_ICD10 | O88.22 | thromboembolism in childbirth |
|  | DX_ICD10 | O88.23 | thromboembolism in the puerperium |
|  | DX_ICD10 | O88.3 | obstetric pyemic and septic embolism |
|  | DX_ICD10 | O88.31 | pyemic and septic embolism in preg |
|  | DX_ICD10 | O88.311 | pyemic and septic embolism in preg, first tri |
|  | DX_ICD10 | O88.312 | pyemic and septic embolism in preg, second tri |
|  | DX_ICD10 | O88.313 | pyemic and septic embolism in preg, third tri |
|  | DX_ICD10 | O88.319 | pyemic and septic embolism in preg, unspec tri |
|  | DX_ICD10 | O88.32 | pyemic and septic embolism in childbirth |
|  | DX_ICD10 | O88.33 | pyemic and septic embolism in the puerperium |
|  | DX_ICD10 | O88.8 | other obstetric embolism |
|  | DX_ICD10 | O88.81 | other embolism in preg |
|  | DX_ICD10 | O88.811 | other embolism in preg, first tri |
|  | DX_ICD10 | O88.812 | other embolism in preg, second tri |
|  | DX_ICD10 | O88.813 | other embolism in preg, third tri |
|  | DX_ICD10 | O88.819 | other embolism in preg, unspec tri |
|  | DX_ICD10 | O88.82 | other embolism in childbirth |
|  | DX_ICD10 | O88.83 | other embolism in the puerperium |
| **Maternal Death**  1 ICD-9 DX 1 ICD-10 DX **2 Total** | DX_ICD9 | 761.6 | mat death aff nb |
|  | DX_ICD10 | P01.6 | newborn affected by mat death |

| **FETAL COMPLICATIONS (366 codes) [6 Conditions]** | | | |
| --- | --- | --- | --- |
|  | **TYPE** | **CODE** | **DESCRIPTION** |
| **Excess Fetal growth** 10 ICD-9 DX 37 ICD-10 DX **47 total** | DX_ICD9 | 656.60 | excess fetal growth- unspec |
|  | DX_ICD9 | 656.61 | excess fetal growth- delivered |
|  | DX_ICD9 | 656.63 | excess fetal growth- antepartum |
|  | DX_ICD9 | 676.60 | exceptionally large baby |
|  | DX_ICD9 | 676.61 | heavy-for-date infant nec |
|  | DX_ICD9 | 676.62 | galactorrhea-del w p/p |
|  | DX_ICD9 | 676.63 | galactorrhea-antepartum |
|  | DX_ICD9 | 676.64 | galactorrhea-postpartum |
|  | DX_ICD9 | 766.0 | exceptionally large baby |
|  | DX_ICD9 | 766.1 | oth "heavy-for-dates" infants |
|  | DX_ICD10 | O36.6 | mat care excessive fetal growth |
|  | DX_ICD10 | O36.60 | mat care excessive fetal growth, unspec tri |
|  | DX_ICD10 | O36.60X0 | mat care excessive fetal growth, unspec tri, n/a or unspec |
|  | DX_ICD10 | O36.60X1 | mat care excessive fetal growth, unspec tri, fetus 1 |
|  | DX_ICD10 | O36.60X2 | mat care excessive fetal growth, unspec tri, fetus 2 |
|  | DX_ICD10 | O36.60X3 | mat care excessive fetal growth, unspec tri, fetus 3 |
|  | DX_ICD10 | O36.60X4 | mat care excessive fetal growth, unspec tri, fetus 4 |
|  | DX_ICD10 | O36.60X5 | mat care excessive fetal growth, unspec tri, fetus 5 |
|  | DX_ICD10 | O36.60X9 | mat care excessive fetal growth, unspec tri, oth fetus |
|  | DX_ICD10 | O36.61 | mat care excessive fetal growth, first tri |
|  | DX_ICD10 | O36.61X0 | mat care excessive fetal growth, first tri, n/a or unspec |
|  | DX_ICD10 | O36.61X1 | mat care excessive fetal growth, first tri, fetus 1 |
|  | DX_ICD10 | O36.61X2 | mat care excessive fetal growth, first tri, fetus 2 |
|  | DX_ICD10 | O36.61X3 | mat care excessive fetal growth, first tri, fetus 3 |
|  | DX_ICD10 | O36.61X4 | mat care excessive fetal growth, first tri, fetus 4 |
|  | DX_ICD10 | O36.61X5 | mat care excessive fetal growth, first tri, fetus 5 |
|  | DX_ICD10 | O36.61X9 | mat care excessive fetal growth, first tri, oth fetus |
|  | DX_ICD10 | O36.62 | mat care excessive fetal growth, sec tri |
|  | DX_ICD10 | O36.62X0 | mat care excessive fetal growth, sec tri, n/a or unspec |
|  | DX_ICD10 | O36.62X1 | mat care excessive fetal growth, sec tri, fetus 1 |
|  | DX_ICD10 | O36.62X2 | mat care excessive fetal growth, sec tri, fetus 2 |
|  | DX_ICD10 | O36.62X3 | mat care excessive fetal growth, sec tri, fetus 3 |
|  | DX_ICD10 | O36.62X4 | mat care excessive fetal growth, sec tri, fetus 4 |
|  | DX_ICD10 | O36.62X5 | mat care excessive fetal growth, sec tri, fetus 5 |
|  | DX_ICD10 | O36.62X9 | mat care excessive fetal growth, sec tri, oth fetus |
|  | DX_ICD10 | O36.63 | mat care excessive fetal growth, third tri |
|  | DX_ICD10 | O36.63X0 | mat care excessive fetal growth, third tri, n/a or unspec |
|  | DX_ICD10 | O36.63X1 | mat care excessive fetal growth, third tri, fetus 1 |
|  | DX_ICD10 | O36.63X2 | mat care excessive fetal growth, third tri, fetus 2 |
|  | DX_ICD10 | O36.63X3 | mat care excessive fetal growth, third tri, fetus 3 |
|  | DX_ICD10 | O36.63X4 | mat care excessive fetal growth, third tri, fetus 4 |
|  | DX_ICD10 | O36.63X5 | mat care excessive fetal growth, third tri, fetus 5 |
|  | DX_ICD10 | O36.63X9 | mat care excessive fetal growth, third tri, oth fetus |
|  | DX_ICD10 | O92.6 | galactorrhea |
|  | DX_ICD10 | P08 | dis of NB related to long gestational & high bw |
|  | DX_ICD10 | P08.0 | exceptionally large NB baby |
|  | DX_ICD10 | P08.1 | oth heavy for gestational age NB |
| **Fetal Distress** 13 ICD-9 DX 15 ICD-10 DX **28 total** | DX_ICD9 | 656.30 | fetal distress- unspecified |
|  | DX_ICD9 | 656.31 | fetal distress- delivered |
|  | DX_ICD9 | 656.33 | fetal distress- antepartum |
|  | DX_ICD9 | 659.70 | abn fetal heart nos (begin 1998) |
|  | DX_ICD9 | 659.71 | abn fetal heart deliv (begin 1998) |
|  | DX_ICD9 | 659.73 | abn fetal heart antepartum (begin 1998) |
|  | DX_ICD9 | 763.81 | abnorm in fetal heart rate or rhythm before onset of labor |
|  | DX_ICD9 | 763.82 | abnormal in fetal heart rate or rhythm dur labor |
|  | DX_ICD9 | 763.83 | abnormal in fetal heart rate or rhythm, unspec to time |
|  | DX_ICD9 | 763.89 | oth spec complic of labor and delivery affecting fetus or NB |
|  | DX_ICD9 | 768.2 | fetal distress before onset of labor, in liveborn infant |
|  | DX_ICD9 | 768.3 | fetal distress first noted dur labor & delivery, in LB infant |
|  | DX_ICD9 | 779.84 | meconium staining |
|  | DX_ICD10 | O68 | labor & delivery complic by abnorm fetal acid-base balance |
|  | DX_ICD10 | O76 | abnorm fetal heart rate & rhythm complic labor & del |
|  | DX_ICD10 | O77 | oth fetal stress complic labor and delivery |
|  | DX_ICD10 | O77.1 | fetal stress in labor or del due to drug admin |
|  | DX_ICD10 | O77.8 | labor & delivery complic by oth evidence of fetal stress |
|  | DX_ICD10 | O77.9 | labor & delivery complic by fetal stress, unspec |
|  | DX_ICD10 | P03.81 | NB (suspected) affect by abnorm fetal (intrauter) hrt rate or rhythm |
|  | DX_ICD10 | P03.810 | NB affect by abnorm fetal (intrauterine) hrt rate or rhythm b4 labor |
|  | DX_ICD10 | P03.811 | NB affect by abnorm fetal (intrauterine) hrt rate or rhythm dur labor |
|  | DX_ICD10 | P03.819 | NB affect by abnorm fetal (intrauter) hrt rate or rhythm, unspec time |
|  | DX_ICD10 | P03.82 | meconium passage during delivery |
|  | DX_ICD10 | P19.0 | metabolic acidemia in NB first noted b4 labor |
|  | DX_ICD10 | P19.1 | metabolic acidemia in NB first noted dur labor |
|  | DX_ICD10 | P19.2 | metabolic acidemia noted at birth |
|  | DX_ICD10 | P96.83 | meconium staining |
| **Fetal growth Restriction** 53 ICD-9 DX 67 ICD-10 DX **120 total** | DX_ICD9 | 656.50 | poor fetal growth- unspec |
|  | DX_ICD9 | 656.51 | poor fetal growth- delivered |
|  | DX_ICD9 | 656.53 | poor fetal growth- antepartum |
|  | DX_ICD9 | 764.00 | light-for- dates weight nos (begin 1988) |
|  | DX_ICD9 | 764.0 | light-for-dates w/o fetal mal (begin 1980 end 1988) |
|  | DX_ICD9 | 764.01 | light-for- dates < 500g (begin 1988) |
|  | DX_ICD9 | 764.02 | light-for- dates 500-749g (begin 1988) |
|  | DX_ICD9 | 764.03 | light-for- dates 750-999g (begin 1988) |
|  | DX_ICD9 | 764.04 | light-for- dates 1000-1249g (begin 1988) |
|  | DX_ICD9 | 764.05 | light-for- dates 1250-1499g (begin 1988) |
|  | DX_ICD9 | 764.06 | light-for- dates 1500-1749g (begin 1988) |
|  | DX_ICD9 | 764.07 | light-for- dates 1750-1999g (begin 1988) |
|  | DX_ICD9 | 764.08 | light-for- dates 2000-2499g (begin 1988) |
|  | DX_ICD9 | 764.09 | light-for-dates 2500+g (begin 1988) |
|  | DX_ICD9 | 764.10 | light-for-date w mal weight nos (begin 1988) |
|  | DX_ICD9 | 764.1 | light-for-dates w fetal mal (begin 1980 end 1988) |
|  | DX_ICD9 | 764.11 | light-for-date w mal <500g (begin 1988) |
|  | DX_ICD9 | 764.12 | light-for-date w mal 500-749g (begin 1988) |
|  | DX_ICD9 | 764.13 | light-for-date w mal 750-999g (begin 1988) |
|  | DX_ICD9 | 764.14 | light-for-date w mal 1000-1249g (begin 1988) |
|  | DX_ICD9 | 764.15 | light-for-date w mal 1250-1499g (begin 1988) |
|  | DX_ICD9 | 764.16 | light-for-date w mal 1500-1749g (begin 1988) |
|  | DX_ICD9 | 764.17 | light-for-date w mal 1750-1999g (begin 1988) |
|  | DX_ICD9 | 764.18 | light-for-date w mal 2000-2499g (begin 1988) |
|  | DX_ICD9 | 764.19 | light-for-date w mal 2500+g (begin 1988) |
|  | DX_ICD9 | 764.20 | fetal malnutrition weight nos (begin 1988) |
|  | DX_ICD9 | 764.2 | fetal mal w/o light-for-dates (begin 1980 end 1988) |
|  | DX_ICD9 | 764.21 | fetal malnutrition < 500g (begin 1988) |
|  | DX_ICD9 | 764.22 | fetal malnutrition 500-749g (begin 1988) |
|  | DX_ICD9 | 764.23 | fetal malnutrition 750-999g (begin 1988) |
|  | DX_ICD9 | 764.24 | fetal malnutrition 1000-1249g (begin 1988) |
|  | DX_ICD9 | 764.25 | fetal malnutrition 1250-1499g (begin 1988) |
|  | DX_ICD9 | 764.26 | fetal malnutrition 1500-1749g (begin 1988) |
|  | DX_ICD9 | 764.27 | fetal malnutrition 1750- 1999g (begin 1988) |
|  | DX_ICD9 | 764.28 | fetal malnutrition 2000-2499g (begin 1988) |
|  | DX_ICD9 | 764.29 | fetal malnutrition 2500+g (begin 1988) |
|  | DX_ICD9 | 764.90 | fetal growth retardation weight nos (begin 1988) |
|  | DX_ICD9 | 764.9 | fetal growth retard nos (begin 1980 end 1988) |
|  | DX_ICD9 | 764.91 | fetal growth retardation < 500g (begin 1988) |
|  | DX_ICD9 | 764.92 | fetal growth retardation 500-749g (begin 1988) |
|  | DX_ICD9 | 764.93 | fetal growth retardation 750-999g (begin 1988) |
|  | DX_ICD9 | 764.94 | fetal growth retardation 1000-1249g (begin 1988) |
|  | DX_ICD9 | 764.95 | fetal growth retardation 1250-1499g (begin 1988) |
|  | DX_ICD9 | 764.96 | fetal growth retardation 1500-1749g (begin 1988) |
|  | DX_ICD9 | 764.97 | fetal growth retardation 1750-1999g (begin 1988) |
|  | DX_ICD9 | 764.98 | fetal growth retardation 2000- 2499g (begin 1988) |
|  | DX_ICD9 | 764.99 | fetal growth retardation 2500+g (begin 1988) |
|  | DX_ICD9 | V21.30 | lbw unspec (begin 2000) |
|  | DX_ICD9 | V21.31 | lbw < 500g (begin 2000) |
|  | DX_ICD9 | V21.32 | lbw 500-999g (begin 2000) |
|  | DX_ICD9 | V21.33 | lbw 1000-1499g (begin 2000) |
|  | DX_ICD9 | V21.34 | lbw 1500-1999g (begin 2000) |
|  | DX_ICD9 | V21.35 | lbw 2000-2500g (begin 2000) |
|  | DX_ICD10 | O36.5 | mat care known/susp poor fetal growth |
|  | DX_ICD10 | O36.5110 | mat care known/susp placental insuffic, first tri, n/a or unspec |
|  | DX_ICD10 | O36.5120 | mat care known/susp placental insuffic, sec tri, n n/a or unspec |
|  | DX_ICD10 | O36.5130 | mat care known/susp placental insuffic, third tri, n/a or unspec |
|  | DX_ICD10 | O36.59 | mat care oth known/susp poor fetal growth |
|  | DX_ICD10 | O36.591 | mat care oth known/susp poor fetal growth, first tri |
|  | DX_ICD10 | O36.5910 | mat care oth known/susp poor fetal growth, first tri, n/a or unspec |
|  | DX_ICD10 | O36.5911 | mat care oth known/susp poor fetal growth, first tri, fetus 1 |
|  | DX_ICD10 | O36.5912 | mat care oth known/susp poor fetal growth, first tri, fetus 2 |
|  | DX_ICD10 | O36.5913 | mat care oth known/susp poor fetal growth, first tri, fetus 3 |
|  | DX_ICD10 | O36.5914 | mat care oth known/susp poor fetal growth, first tri, fetus 4 |
|  | DX_ICD10 | O36.5915 | mat care oth known/susp poor fetal growth, first tri, fetus 5 |
|  | DX_ICD10 | O36.5919 | mat care oth known/susp poor fetal growth, first tri, oth fetus |
|  | DX_ICD10 | O36.592 | mat care oth known/susp poor fetal growth, sec tri |
|  | DX_ICD10 | O36.5920 | mat care oth known/susp poor fetal growth, sec tri, n/a or unspec |
|  | DX_ICD10 | O36.5921 | mat care oth known/susp poor fetal growth, sec tri, fetus 1 |
|  | DX_ICD10 | O36.5922 | mat care oth known/susp poor fetal growth, sec tri, fetus 2 |
|  | DX_ICD10 | O36.5923 | mat care oth known/susp poor fetal growth, sec tri, fetus 3 |
|  | DX_ICD10 | O36.5924 | mat care oth known/susp poor fetal growth, sec tri, fetus 4 |
|  | DX_ICD10 | O36.5925 | mat care oth known/susp poor fetal growth, sec tri, fetus 5 |
|  | DX_ICD10 | O36.5929 | mat care oth known/susp poor fetal growth, sec tri, oth fetus |
|  | DX_ICD10 | O36.593 | mat care oth known/susp poor fetal growth, third tri |
|  | DX_ICD10 | O36.5930 | mat care oth known/susp poor fetal growth, third tri, n/a or unspec |
|  | DX_ICD10 | O36.5931 | mat care oth known/susp poor fetal growth, third tri, fetus 1 |
|  | DX_ICD10 | O36.5932 | mat care oth known/susp poor fetal growth, third tri, fetus 2 |
|  | DX_ICD10 | O36.5933 | mat care oth known/susp poor fetal growth, third tri, fetus 3 |
|  | DX_ICD10 | O36.5934 | mat care oth known/susp poor fetal growth, third tri, fetus 4 |
|  | DX_ICD10 | O36.5935 | mat care oth known/susp poor fetal growth, third tri, fetus 5 |
|  | DX_ICD10 | O36.5939 | mat care oth known/susp poor fetal growth, third tri, oth fetus |
|  | DX_ICD10 | O36.599 | mat care oth known/susp poor fetal growth, unspec tri |
|  | DX_ICD10 | O36.5990 | mat care oth known/susp poor fetal growth, unspec tri, n/a or unspec |
|  | DX_ICD10 | O36.5991 | mat care oth known/susp poor fetal growth, unspec tri, fetus 1 |
|  | DX_ICD10 | O36.5992 | mat care oth known/susp poor fetal growth, unspec tri, fetus 2 |
|  | DX_ICD10 | O36.5993 | mat care oth known/susp poor fetal growth, unspec tri, fetus 3 |
|  | DX_ICD10 | O36.5994 | mat care oth known/susp poor fetal growth, unspec tri, fetus 4 |
|  | DX_ICD10 | O36.5995 | mat care oth known/susp poor fetal growth, unspec tri, fetus 5 |
|  | DX_ICD10 | O36.5999 | mat care oth known/susp poor fetal growth, unspec tri, oth fetus |
|  | DX_ICD10 | P05 | disorders of NB related to slow fetal growth & fetal malnutr |
|  | DX_ICD10 | P05.0 | NB light for gestational age NB light-for-dates |
|  | DX_ICD10 | P05.00 | NB light for gestational age, unspec weight |
|  | DX_ICD10 | P05.01 | NB light for gestational age, less than 500 grams |
|  | DX_ICD10 | P05.02 | NB light for gestational age, 500-749 grams |
|  | DX_ICD10 | P05.03 | NB light for gestational age, 750-999 grams |
|  | DX_ICD10 | P05.04 | NB light for gestational age, 1000-1249 grams |
|  | DX_ICD10 | P05.05 | NB light for gestational age, 1250-1499 grams |
|  | DX_ICD10 | P05.06 | NB light for gestational age, 1500-1749 grams |
|  | DX_ICD10 | P05.07 | NB light for gestational age, 1750-1999 grams |
|  | DX_ICD10 | P05.08 | NB light for gestational ional age, 2000-2499 grams |
|  | DX_ICD10 | P05.1 | NB small for gestational age |
|  | DX_ICD10 | P05.10 | NB small for gestational age, unspec weight |
|  | DX_ICD10 | P05.11 | NB small for gestational age, less than 500 grams |
|  | DX_ICD10 | P05.12 | NB small for gestational age, 500-749 grams |
|  | DX_ICD10 | P05.13 | NB small for gestational age, 750-999 grams |
|  | DX_ICD10 | P05.14 | NB small for gestational age, 1000-1249 grams |
|  | DX_ICD10 | P05.15 | NB small for gestational age, 1250-1499 grams |
|  | DX_ICD10 | P05.16 | NB small for gestational age, 1500-1749 grams |
|  | DX_ICD10 | P05.17 | NB small for gestational age, 1750-1999 grams |
|  | DX_ICD10 | P05.18 | NB small for gestational age, 2000-2499 grams |
|  | DX_ICD10 | P05.2 | NB affect by fetal (intrauterine) malnut not light/small for gestat age |
|  | DX_ICD10 | P05.9 | NB affected by slow intrauterine growth, unspec |
|  | DX_ICD10 | P07 | dis of NB related to short gestation & lbw, not elsewhere class |
|  | DX_ICD10 | P07.01 | extremely lbw NB, less than 500 grams |
|  | DX_ICD10 | P07.02 | extremely lbw NB, 500-749 grams |
|  | DX_ICD10 | P07.10 | oth lbw NB, unspec weight |
|  | DX_ICD10 | P07.14 | oth lbw NB, 1000-1249 grams |
|  | DX_ICD10 | P07.16 | oth lbw NB, 1500-1749 grams |
|  | DX_ICD10 | P07.18 | oth lbw NB, 2000-2499 grams |
| **Fetal Hemorrhage** 9 ICD-9 DX 19 ICD-10 DX **28 total** | DX_ICD9 | 772.0 | fetal blood loss nec |
|  | DX_ICD9 | 772.3 | post-birth umbilical hemorrhage |
|  | DX_ICD9 | 772.4 | NB gastrointestinal hemorrhage |
|  | DX_ICD9 | 772.5 | NB adrenal hemorrhage |
|  | DX_ICD9 | 772.6 | NB cutaneous hemorrhage |
|  | DX_ICD9 | 772.8 | neonatal hemorrhage nec |
|  | DX_ICD9 | 772.9 | neonatal hemorrhage nos |
|  | DX_ICD9 | 776.0 | NB hemorrhagic disease |
|  | DX_ICD9 | 776.5 | congenital anemia |
|  | DX_ICD10 | P50.0 | NB affect by intrauterine (fetal) blood loss from vasa previa |
|  | DX_ICD10 | P50.1 | NB affect by intrauterine (fetal) blood loss from rupt cord |
|  | DX_ICD10 | P50.2 | NB affect by intrauterine (fetal) blood loss from placenta |
|  | DX_ICD10 | P50.3 | NB affect by hemorrhage into co-twin |
|  | DX_ICD10 | P50.4 | NB affect by hemorrhage into mat circulation |
|  | DX_ICD10 | P50.5 | NB affect by intrauterine (fetal) blood loss - cut end co-twins cord |
|  | DX_ICD10 | P50.8 | NB affect by oth intrauterine (fetal) blood loss |
|  | DX_ICD10 | P50.9 | NB affect by intrauterine (fetal) blood loss, unspec |
|  | DX_ICD10 | P51 | umbilical hemorrhage of NB |
|  | DX_ICD10 | P51.0 | massive umbilical hemorrhage of NB |
|  | DX_ICD10 | P51.8 | oth umbilical hemorrhages of NB |
|  | DX_ICD10 | P51.9 | umbilical hemorrhage of NB, unspec |
|  | DX_ICD10 | P53 | hemorrhagic disease of NB |
|  | DX_ICD10 | P54.3 | oth neonatal gastrointestinal hemorrhage |
|  | DX_ICD10 | P54.4 | neonatal adrenal hemorrhage |
|  | DX_ICD10 | P54.5 | neonatal cutaneous hemorrhage |
|  | DX_ICD10 | P54.8 | oth spec neonatal hemorrhages |
|  | DX_ICD10 | P54.9 | neonatal hemorrhage, unspec |
|  | DX_ICD10 | P61.3 | congenital anemia from fetal blood loss |
| **Placental Complications Affecting Newborn** 4 ICD-9 DX 8 ICD-10 DX **12 total** | DX_ICD9 | 656.70 | oth placental condition- unspec |
|  | DX_ICD9 | 762.0 | placenta previa affecting NB |
|  | DX_ICD9 | 762.1 | placenta hem nec affecting NB |
|  | DX_ICD9 | 762.3 | placental transfusion syndrome |
|  | DX_ICD10 | O43.199 | oth malform of placenta, unspec tri |
|  | DX_ICD10 | P02 | NB (suspected) affect by complicating placenta, cord & membranes |
|  | DX_ICD10 | P02.0 | NB affect by placenta previa |
|  | DX_ICD10 | P02.1 | NB affect by oth forms placental sep & hemorrhage |
|  | DX_ICD10 | P02.2 | NB (suspected) affect by oth & unspec morph & funct abnorm plcnta |
|  | DX_ICD10 | P02.20 | NB affect by unspec morph & functional abnorm of placenta |
|  | DX_ICD10 | P02.3 | NB affect by placental transfusion syndromes |
|  | DX_ICD10 | P04 | NB (suspected) affect by noxious subst transmit via plcnta/brst milk |
| **Preterm delivery** 33 ICD-9 DX 65 ICD-10 DX **98 total** | DX_ICD9 | 644.20 | early onset delivery, unspec |
|  | DX_ICD9 | 644.21 | early onset delivery- delivered |
|  | DX_ICD9 | 765.00 | extreme immaturity weight nos |
|  | DX_ICD9 | 765.0 | extreme immaturity (begin 1980, end 1988) |
|  | DX_ICD9 | 765.01 | extreme immaturity < 500g (begin 1988) |
|  | DX_ICD9 | 765.02 | extreme immaturity 500-749g (begin 1988) |
|  | DX_ICD9 | 765.03 | extreme immaturity 750-999g (begin 1988) |
|  | DX_ICD9 | 765.04 | extreme immaturity 1000-1249g (begin 1988) |
|  | DX_ICD9 | 765.05 | extreme immaturity 1250-1499g (begin 1988) |
|  | DX_ICD9 | 765.06 | extreme immaturity 1500-1749g (begin 1988) |
|  | DX_ICD9 | 765.07 | extreme immaturity 1750-1999g (begin 1988) |
|  | DX_ICD9 | 765.08 | extreme immaturity 2000-2499g (begin 1988) |
|  | DX_ICD9 | 765.09 | extreme immaturity 2500+g (begin 1988) |
|  | DX_ICD9 | 765.10 | preterm infant nec weight nos |
|  | DX_ICD9 | 765.1 | oth preterm infants (begin 1980, end 1988) |
|  | DX_ICD9 | 765.11 | preterm nec < 500g (begin 1988) |
|  | DX_ICD9 | 765.12 | preterm nec 500-749g (begin 1988) |
|  | DX_ICD9 | 765.13 | preterm nec 750-999g (begin 1988) |
|  | DX_ICD9 | 765.14 | preterm nec 1000-1249g (begin 1988) |
|  | DX_ICD9 | 765.15 | preterm nec 1250-1499g (begin 1988) |
|  | DX_ICD9 | 765.16 | preterm nec 1500- 1749g (begin 1988) |
|  | DX_ICD9 | 765.17 | preterm nec1750-1999g (begin 1988) |
|  | DX_ICD9 | 765.18 | preterm nec 2000-2499g (begin 1988) |
|  | DX_ICD9 | 765.19 | preterm nec 2500+g (begin 1988) |
|  | DX_ICD9 | 765.20 | unspec wks of gestation |
|  | DX_ICD9 | 765.21 | less than 24 completed wks of gestation (begin 2002) |
|  | DX_ICD9 | 765.22 | 24 completed wks gestation |
|  | DX_ICD9 | 765.23 | 25-26 completed wks gestation |
|  | DX_ICD9 | 765.24 | 27-28 completed wks gestation |
|  | DX_ICD9 | 765.25 | 29-30 completed wks gestation |
|  | DX_ICD9 | 765.26 | 31-32 completed wks gestation |
|  | DX_ICD9 | 765.27 | 33-34 completed wks gestation |
|  | DX_ICD9 | 765.28 | 35-36 completed wks gestation (begin 2000) |
|  | DX_ICD10 | O60.1 | preterm labor w preterm del |
|  | DX_ICD10 | O60.10 | preterm labor w preterm del, unspec tri |
|  | DX_ICD10 | O60.10X0 | preterm labor w preterm del, unspec tri, n/a or unspec |
|  | DX_ICD10 | O60.10X1 | preterm labor w preterm del, unspec tri, fetus 1 |
|  | DX_ICD10 | O60.10X2 | preterm labor w preterm del, unspec tri, fetus 2 |
|  | DX_ICD10 | O60.10X3 | preterm labor w preterm del, unspec tri, fetus 3 |
|  | DX_ICD10 | O60.10X4 | preterm labor w preterm del, unspec tri, fetus 4 |
|  | DX_ICD10 | O60.10X5 | preterm labor w preterm del, unspec tri, fetus 5 |
|  | DX_ICD10 | O60.10X9 | preterm labor w preterm del, unspec tri, oth fetus |
|  | DX_ICD10 | O60.12 | preterm labor sec tri w preterm del sec tri |
|  | DX_ICD10 | O60.12X0 | preterm labor sec tri w preterm del sec tri, n/a or unspec |
|  | DX_ICD10 | O60.12X1 | preterm labor sec tri w preterm del sec tri, fetus 1 |
|  | DX_ICD10 | O60.12X2 | preterm labor sec tri w preterm del sec tri, fetus 2 |
|  | DX_ICD10 | O60.12X3 | preterm labor sec tri w preterm del sec tri, fetus 3 |
|  | DX_ICD10 | O60.12X4 | preterm labor sec tri w preterm del sec tri, fetus 4 |
|  | DX_ICD10 | O60.12X5 | preterm labor sec tri w preterm del sec tri, fetus 5 |
|  | DX_ICD10 | O60.12X9 | preterm labor sec tri w preterm del sec tri, oth fetus |
|  | DX_ICD10 | O60.13 | preterm labor sec tri w preterm del third tri |
|  | DX_ICD10 | O60.13X0 | preterm labor sec tri w preterm del third tri, n/a or unspec |
|  | DX_ICD10 | O60.13X1 | preterm labor sec tri w preterm del third tri, fetus 1 |
|  | DX_ICD10 | O60.13X2 | preterm labor sec tri w preterm del third tri, fetus 2 |
|  | DX_ICD10 | O60.13X3 | preterm labor sec tri w preterm del third tri, fetus 3 |
|  | DX_ICD10 | O60.13X4 | preterm labor sec tri w preterm del third tri, fetus 4 |
|  | DX_ICD10 | O60.13X5 | preterm labor sec tri w preterm del third tri, fetus 5 |
|  | DX_ICD10 | O60.13X9 | preterm labor sec tri w preterm del third tri, oth fetus |
|  | DX_ICD10 | O60.14 | preterm labor third tri w preterm del third tri |
|  | DX_ICD10 | O60.14X0 | preterm labor third tri w preterm del third tri, n/a or unspec |
|  | DX_ICD10 | O60.14X1 | preterm labor third tri w preterm del third tri, fetus 1 |
|  | DX_ICD10 | O60.14X2 | preterm labor third tri w preterm del third tri, fetus 2 |
|  | DX_ICD10 | O60.14X3 | preterm labor third tri w preterm del third tri, fetus 3 |
|  | DX_ICD10 | O60.14X4 | preterm labor third tri w preterm del third tri, fetus 4 |
|  | DX_ICD10 | O60.14X5 | preterm labor third tri w preterm del third tri, fetus 5 |
|  | DX_ICD10 | O60.14X9 | preterm labor third tri w preterm del third tri, oth fetus |
|  | DX_ICD10 | O92.5 | suppressed lactation |
|  | DX_ICD10 | P07.0 | extremely lbw NB |
|  | DX_ICD10 | P07.00 | extremely lbw NB, unspec weight |
|  | DX_ICD10 | P07.01 | extremely lbw NB, less than 500 grams |
|  | DX_ICD10 | P07.02 | extremely lbw NB, 500-749 grams |
|  | DX_ICD10 | P07.03 | extremely lbw NB, 750-999 grams |
|  | DX_ICD10 | P07.1 | oth lbw NB |
|  | DX_ICD10 | P07.10 | oth lbw NB, unspec weight |
|  | DX_ICD10 | P07.14 | oth lbw NB, 1000-1249 grams |
|  | DX_ICD10 | P07.15 | oth lbw NB, 1250-1499 grams |
|  | DX_ICD10 | P07.16 | oth lbw NB, 1500-1749 grams |
|  | DX_ICD10 | P07.17 | oth lbw NB, 1750-1999 grams |
|  | DX_ICD10 | P07.18 | oth lbw NB, 2000-2499 grams |
|  | DX_ICD10 | P07.2 | extreme immaturity of NB |
|  | DX_ICD10 | P07.20 | extreme immaturity of NB, unspecified wks of gestation |
|  | DX_ICD10 | P07.21 | extreme immaturity of NB, gestational age less than 23 wks |
|  | DX_ICD10 | P07.22 | extreme immaturity of NB, gestational age 23 completed wks |
|  | DX_ICD10 | P07.23 | extreme immaturity of NB, gestational age 24 completed wks |
|  | DX_ICD10 | P07.24 | extreme immaturity of NB, gestational age 25 completed wks |
|  | DX_ICD10 | P07.25 | extreme immaturity of NB, gestational age 26 completed wks |
|  | DX_ICD10 | P07.26 | extreme immaturity of NB, gestational age 27 completed wks |
|  | DX_ICD10 | P07.3 | oth preterm NB |
|  | DX_ICD10 | P07.30 | preterm NB, unspecified wks of gestation |
|  | DX_ICD10 | P07.31 | preterm NB, gestational age 28 completed wks |
|  | DX_ICD10 | P07.32 | preterm NB, gestational age 29 completed wks |
|  | DX_ICD10 | P07.33 | preterm NB, gestational age 30 completed wks |
|  | DX_ICD10 | P07.34 | preterm NB, gestational age 31 completed wks |
|  | DX_ICD10 | P07.35 | preterm NB, gestational age 32 completed wks |
|  | DX_ICD10 | P07.36 | preterm NB, gestational age 33 completed wks |
|  | DX_ICD10 | P07.37 | preterm NB, gestational age 34 completed wks |
|  | DX_ICD10 | P07.38 | preterm NB, gestational age 35 completed wks |
|  | DX_ICD10 | P07.39 | preterm NB, gestational age 36 completed wks |

| **FETAL DEATH AND/OR STILLBIRTH (91 codes) [2 Conditions]** | | | |
| --- | --- | --- | --- |
|  | **TYPE** | **CODE** | **DESCRIPTION** |
| **Fetal Death** 9 ICD-9 DX 74 ICD-10 DX **83 Total** | DX_ICD9 | 646.00 | papyraceous fetus-unspecified |
|  | DX_ICD9 | 646.01 | papyraceous fetus-delivered |
|  | DX_ICD9 | 646.03 | papyraceous fetus-antepartum |
|  | DX_ICD9 | 656.40 | intrauterine death-unspecified |
|  | DX_ICD9 | 656.41 | intrauterine death-delivered |
|  | DX_ICD9 | 656.43 | intrauterine death-antepartum |
|  | DX_ICD9 | 768.0 | fetal death- anoxia nos |
|  | DX_ICD9 | 768.1 | fetal death- anoxia during labor |
|  | DX_ICD9 | 770.88 | fetal death- anoxia nos |
|  | DX_ICD10 | O31.0 | papyraceous fetus |
|  | DX_ICD10 | O31.00 | papyraceous fetus, unspecified tri |
|  | DX_ICD10 | O31.00X0 | papyraceous fetus, unspecified tri, not applicable or unspecified |
|  | DX_ICD10 | O31.00X1 | papyraceous fetus, unspecified tri, fetus 1 |
|  | DX_ICD10 | O31.00X2 | papyraceous fetus, unspecified tri, fetus 2 |
|  | DX_ICD10 | O31.00X3 | papyraceous fetus, unspecified tri, fetus 3 |
|  | DX_ICD10 | O31.00X4 | papyraceous fetus, unspecified tri, fetus 4 |
|  | DX_ICD10 | O31.00X5 | papyraceous fetus, unspecified tri, fetus 5 |
|  | DX_ICD10 | O31.00X9 | papyraceous fetus, unspecified tri, other fetus |
|  | DX_ICD10 | O31.01 | papyraceous fetus, first tri |
|  | DX_ICD10 | O31.01X0 | papyraceous fetus, first tri, not applicable or unspecified |
|  | DX_ICD10 | O31.01X1 | papyraceous fetus, first tri, fetus 1 |
|  | DX_ICD10 | O31.01X2 | papyraceous fetus, first tri, fetus 2 |
|  | DX_ICD10 | O31.01X3 | papyraceous fetus, first tri, fetus 3 |
|  | DX_ICD10 | O31.01X4 | papyraceous fetus, first tri, fetus 4 |
|  | DX_ICD10 | O31.01X5 | papyraceous fetus, first tri, fetus 5 |
|  | DX_ICD10 | O31.01X9 | papyraceous fetus, first tri, other fetus |
|  | DX_ICD10 | O31.02 | papyraceous fetus, second tri |
|  | DX_ICD10 | O31.02X0 | papyraceous fetus, second tri, not applicable or unspecified |
|  | DX_ICD10 | O31.02X1 | papyraceous fetus, second tri, fetus 1 |
|  | DX_ICD10 | O31.02X2 | papyraceous fetus, second tri, fetus 2 |
|  | DX_ICD10 | O31.02X3 | papyraceous fetus, second tri, fetus 3 |
|  | DX_ICD10 | O31.02X4 | papyraceous fetus, second tri, fetus 4 |
|  | DX_ICD10 | O31.02X5 | papyraceous fetus, second tri, fetus 5 |
|  | DX_ICD10 | O31.02X9 | papyraceous fetus, second tri, other fetus |
|  | DX_ICD10 | O31.03 | papyraceous fetus, third tri |
|  | DX_ICD10 | O31.03X0 | papyraceous fetus, third tri, not applicable or unspecif |
|  | DX_ICD10 | O31.03X1 | papyraceous fetus, third tri, fetus 1 |
|  | DX_ICD10 | O31.03X2 | papyraceous fetus, third tri, fetus 2 |
|  | DX_ICD10 | O31.03X3 | papyraceous fetus, third tri, fetus 3 |
|  | DX_ICD10 | O31.03X4 | papyraceous fetus, third tri, fetus 4 |
|  | DX_ICD10 | O31.03X5 | papyraceous fetus, third tri, fetus 5 |
|  | DX_ICD10 | O31.03X9 | papyraceous fetus, third tri, other fetus |
|  | DX_ICD10 | O31.2 | continuing preg after intrauterine death 1+ fetus |
|  | DX_ICD10 | O31.20 | continuing preg after intrauterine death 1+ fetus, unspecified tri |
|  | DX_ICD10 | O31.20X0 | contin preg after intrauter death 1+ fetus, unspecif tri, na or unspecif |
|  | DX_ICD10 | O31.20X1 | continuing preg after intrauterine death 1+ fetus, unspecif tri, fetus 1 |
|  | DX_ICD10 | O31.20X2 | continuing preg after intrauterine death 1+ fetus, unspecif tri, fetus 2 |
|  | DX_ICD10 | O31.20X3 | continuing preg after intrauterine death 1+ fetus, unspecif tri, fetus 3 |
|  | DX_ICD10 | O31.20X4 | continuing preg after intrauterine death 1+ fetus, unspecif tri, fetus 4 |
|  | DX_ICD10 | O31.20X5 | continuing preg after intrauterine death 1+ fetus, unspecif tri, fetus 5 |
|  | DX_ICD10 | O31.20X9 | contin preg after intrauter death 1+ fetus, unspecified tri, other fetus |
|  | DX_ICD10 | O31.21 | continuing preg after intrauterine death 1+ fetus, first tri |
|  | DX_ICD10 | O31.21X0 | contin preg after intrauter death 1+ fetus, first tri, na or unspecified |
|  | DX_ICD10 | O31.21X1 | continuing preg after intrauterine death 1+ fetus, first tri, fetus 1 |
|  | DX_ICD10 | O31.21X2 | continuing preg after intrauterine death 1+ fetus, first tri, fetus 2 |
|  | DX_ICD10 | O31.21X3 | continuing preg after intrauterine death 1+ fetus, first tri, fetus 3 |
|  | DX_ICD10 | O31.21X4 | continuing preg after intrauterine death 1+ fetus, first tri, fetus 4 |
|  | DX_ICD10 | O31.21X5 | continuing preg after intrauterine death 1+ fetus, first tri, fetus 5 |
|  | DX_ICD10 | O31.21X9 | continuing preg after intrauterine death 1+ fetus, first tri, other fetus |
|  | DX_ICD10 | O31.22 | continuing preg after intrauterine death 1+ fetus, second tri |
|  | DX_ICD10 | O31.22X0 | contin preg after intrauter death 1+ fetus, second tri, na or unspecified |
|  | DX_ICD10 | O31.22X1 | continuing preg after intrauterine death 1+ fetus, second tri, fetus 1 |
|  | DX_ICD10 | O31.22X2 | continuing preg after intrauterine death 1+ fetus, second tri, fetus 2 |
|  | DX_ICD10 | O31.22X3 | continuing preg after intrauterine death 1+ fetus, second tri, fetus 3 |
|  | DX_ICD10 | O31.22X4 | continuing preg after intrauterine death 1+ fetus, second tri, fetus 4 |
|  | DX_ICD10 | O31.22X5 | continuing preg after intrauterine death 1+ fetus, second tri, fetus 5 |
|  | DX_ICD10 | O31.22X9 | continuing preg after intrauterine death 1+ fetus, second tri, other fetus |
|  | DX_ICD10 | O31.23 | continuing preg after intrauterine death 1+ fetus, third tri |
|  | DX_ICD10 | O31.23X0 | continuing preg after intrauter death 1+ fetus, third tri, na or unspecif |
|  | DX_ICD10 | O31.23X1 | continuing preg after intrauterine death 1+ fetus, third tri, fetus 1 |
|  | DX_ICD10 | O31.23X2 | continuing preg after intrauterine death 1+ fetus, third tri, fetus 2 |
|  | DX_ICD10 | O31.23X3 | continuing preg after intrauterine death 1+ fetus, third tri, fetus 3 |
|  | DX_ICD10 | O31.23X4 | continuing preg after intrauterine death 1+ fetus, third tri, fetus 4 |
|  | DX_ICD10 | O31.23X5 | continuing preg after intrauterine death 1+ fetus, third tri, fetus 5 |
|  | DX_ICD10 | O31.23X9 | continuing preg after intrauterine death 1+ fetus, third tri, other fetus |
|  | DX_ICD10 | O36.4 | mat care for intrauterine death |
|  | DX_ICD10 | O36.4XX0 | mat care for intrauterine death, not applicable or unspecified |
|  | DX_ICD10 | O36.4XX1 | mat care for intrauterine death, fetus 1 |
|  | DX_ICD10 | O36.4XX2 | mat care for intrauterine death, fetus 2 |
|  | DX_ICD10 | O36.4XX3 | mat care for intrauterine death, fetus 3 |
|  | DX_ICD10 | O36.4XX4 | mat care for intrauterine death, fetus 4 |
|  | DX_ICD10 | O36.4XX5 | mat care for intrauterine death, fetus 5 |
|  | DX_ICD10 | O36.4XX9 | mat care for intrauterine death, other fetus |
| **Stillborn** 4 ICD-9 DX 4 ICD-10 DX **8 Total** | DX_ICD9 | V27.1 | deliver- single stillborn |
|  | DX_ICD9 | V27.4 | outcome of delivery, twins, both stillborn |
|  | DX_ICD9 | V27.7 | deliver- multiple births- all stillborn |
|  | DX_ICD9 | V35.0 | other multiple stillborn- in hosp |
|  | DX_ICD10 | P95 | stillbirth |
|  | DX_ICD10 | Z37.1 | single stillbirth |
|  | DX_ICD10 | Z37.4 | twins, both stillborn |
|  | DX_ICD10 | Z37.7 | other multiple births, all stillborn |

| **EARLY PREGNANCY LOSS (421 codes) [3 Conditions]** | | | |
| --- | --- | --- | --- |
|  | **TYPE** | **CODE** | **DESCRIPTION** |
| **Ectopic Preg**  24 ICD-9 DX 2 ICD-9 PX 76 ICD-10 DX 7 CPT 20 ICD-10 PX  **129 Total** | DX_ICD9 | 633.0 | abdominal preg (end 2002) |
|  | DX_ICD9 | 633.00 | abdominal preg w/o intrauterine preg (begin 2002) |
|  | DX_ICD9 | 633.01 | abdominal preg w intrauterine preg (begin 2002) |
|  | DX_ICD9 | 633.1 | tubal preg (end 2002) |
|  | DX_ICD9 | 633.10 | tubal preg w/o intrauterine preg (begin 2002) |
|  | DX_ICD9 | 633.11 | tubal preg w intrauterine preg (begin 2002) |
|  | DX_ICD9 | 633.2 | ovarian preg (end 2002) |
|  | DX_ICD9 | 633.20 | ovarian preg w/o intrauterine preg (begin 2002) |
|  | DX_ICD9 | 633.21 | ovarian preg w intrauterine preg (begin 2002) |
|  | DX_ICD9 | 633.8 | ectopic preg nec (end 2002) |
|  | DX_ICD9 | 633.80 | other ectopic preg w/o intrau preg (begin 2002) |
|  | DX_ICD9 | 633.81 | other ectopic preg w intrauterine preg (begin 2002) |
|  | DX_ICD9 | 633.9 | ectopic preg nos (end 2002) |
|  | DX_ICD9 | 633.90 | unspec ectopic preg w/o intrauterine preg (begin 2002) |
|  | DX_ICD9 | 633.91 | unspec ectopic preg w intrauterine preg (begin 2002) |
|  | DX_ICD9 | 639.1 | post abortion hem |
|  | DX_ICD9 | 639.2 | post abortion pelvic damage |
|  | DX_ICD9 | 639.3 | post abortion renal failure |
|  | DX_ICD9 | 639.4 | post abortion metabolic disease |
|  | DX_ICD9 | 639.5 | post abortion shock |
|  | DX_ICD9 | 639.6 | post abortion embolism |
|  | DX_ICD9 | 639.8 | post abortion complication nec |
|  | DX_ICD9 | 639.9 | post abortion complication nos |
|  | DX_ICD9 | 761.4 | ectopic preg affecting newborn |
|  | PX_ICD9 | 66.62 | salpingectomy with removal of tubal preg |
|  | PX_ICD9 | 74.3 | removal of extratubal ectopic preg |
|  | DX_ICD10 | O00 | abdominal preg |
|  | DX_ICD10 | O00.0 | abdominal preg |
|  | DX_ICD10 | O00.00 | abdominal preg without intrauterine preg |
|  | DX_ICD10 | O00.01 | abdominal preg with intrauterine preg |
|  | DX_ICD10 | O00.1 | tubal preg |
|  | DX_ICD10 | O00.10 | tubal preg wo intrauterine preg |
|  | DX_ICD10 | O00.101 | right tubal preg wo intrauterine preg |
|  | DX_ICD10 | O00.102 | left tubal preg wo intrauterine preg |
|  | DX_ICD10 | O00.109 | unspecif tubal preg without intrauterine preg |
|  | DX_ICD10 | O00.11 | tubal preg with intrauterine preg |
|  | DX_ICD10 | O00.111 | right tubal preg with intrauterine preg |
|  | DX_ICD10 | O00.112 | left tubal preg with intrauterine preg |
|  | DX_ICD10 | O00.119 | unspecified tubal preg with intrauterine preg |
|  | DX_ICD10 | O00.2 | ovarian preg |
|  | DX_ICD10 | O00.20 | ovarian preg wo intrauterine preg |
|  | DX_ICD10 | O00.201 | right ovarian preg wo intrauterine preg |
|  | DX_ICD10 | O00.202 | left ovarian preg wo intrauterine preg |
|  | DX_ICD10 | O00.209 | unspecified ovarian preg wo intrauterine preg |
|  | DX_ICD10 | O00.21 | ovarian preg with intrauterine preg |
|  | DX_ICD10 | O00.211 | right ovarian preg with intrauterine preg |
|  | DX_ICD10 | O00.212 | left ovarian preg with intrauterine preg |
|  | DX_ICD10 | O00.219 | unspecified ovarian preg with intrauterine preg |
|  | DX_ICD10 | O00.8 | other ectopic preg |
|  | DX_ICD10 | O00.80 | other ectopic preg without intrauterine preg |
|  | DX_ICD10 | O00.81 | other ectopic preg with intrauterine preg |
|  | DX_ICD10 | O00.9 | ectopic preg, unspecified |
|  | DX_ICD10 | O00.90 | unspecified ectopic preg wo intrauterine preg |
|  | DX_ICD10 | O00.91 | unspecified ectopic preg w intrauterine preg |
|  | DX_ICD10 | O08 | complications following ectopic and molar preg |
|  | DX_ICD10 | O08.0 | genital tract and pelvic infection following ectopic and molar preg |
|  | DX_ICD10 | O08.1 | delayed or excessive hemorrhage following ectopic and molar preg |
|  | DX_ICD10 | O08.2 | embolism following ectopic and molar preg |
|  | DX_ICD10 | O08.3 | shock following ectopic and molar preg |
|  | DX_ICD10 | O08.4 | renal failure following ectopic and molar preg |
|  | DX_ICD10 | O08.5 | metabolic disorders following an ectopic and molar preg |
|  | DX_ICD10 | O08.6 | damage to pelvic organs and tissue after an ectopic and molar preg |
|  | DX_ICD10 | O08.7 | other venous complications following an ectopic and molar preg |
|  | DX_ICD10 | O08.8 | other complications following an ectopic and molar preg |
|  | DX_ICD10 | O08.81 | cardiac arrest following an ectopic and molar preg |
|  | DX_ICD10 | O08.82 | sepsis following ectopic and molar preg |
|  | DX_ICD10 | O08.83 | urinary tract infection following an ectopic and molar preg |
|  | DX_ICD10 | O08.89 | other complications following an ectopic and molar preg |
|  | DX_ICD10 | O08.9 | unspecified complication following an ectopic and molar preg |
|  | DX_ICD10 | O36.7 | mat care for viable fetus in abdominal preg |
|  | DX_ICD10 | O36.70 | mat care for viable& fetus in abdominal preg, unspecified tri |
|  | DX_ICD10 | O36.70X0 | mat care for viable fetus in abdominal preg, unspecif tri, na or unspecif |
|  | DX_ICD10 | O36.70X1 | mat care for viable fetus in abdominal preg, unspecified tri, fetus 1 |
|  | DX_ICD10 | O36.70X2 | mat care for viable fetus in abdominal preg, unspecified tri, fetus 2 |
|  | DX_ICD10 | O36.70X3 | mat care for viable fetus in abdominal preg, unspecified tri, fetus 3 |
|  | DX_ICD10 | O36.70X4 | mat care for viable fetus in abdominal preg, unspecified tri, fetus 4 |
|  | DX_ICD10 | O36.70X5 | mat care for viable fetus in abdominal preg, unspecified tri, fetus 5 |
|  | DX_ICD10 | O36.70X9 | mat care for viable fetus in abdominal preg, unspecified tri, other fetus |
|  | DX_ICD10 | O36.71 | mat care for viable fetus in abdominal preg, first tri |
|  | DX_ICD10 | O36.71X0 | mat care for viable fetus in abdominal preg, first tri, na or unspecified |
|  | DX_ICD10 | O36.71X1 | mat care for viable fetus in abdominal preg, first tri, fetus 1 |
|  | DX_ICD10 | O36.71X2 | mat care for viable fetus in abdominal preg, first tri, fetus 2 |
|  | DX_ICD10 | O36.71X3 | mat care for viable fetus in abdominal preg, first tri, fetus 3 |
|  | DX_ICD10 | O36.71X4 | mat care for viable fetus in abdominal preg, first tri, fetus 4 |
|  | DX_ICD10 | O36.71X5 | mat care for viable fetus in abdominal preg, first tri, fetus 5 |
|  | DX_ICD10 | O36.71X9 | mat care for viable fetus in abdominal preg, first tri, other fetus |
|  | DX_ICD10 | O36.72 | mat care for viable fetus in abdominal preg, second tri |
|  | DX_ICD10 | O36.72X0 | mat care for viable fetus in abdomin preg, second tri, na or unspecified |
|  | DX_ICD10 | O36.72X1 | mat care for viable fetus in abdominal preg, second tri, fetus 1 |
|  | DX_ICD10 | O36.72X2 | mat care for viable fetus in abdominal preg, second tri, fetus 2 |
|  | DX_ICD10 | O36.72X3 | mat care for viable fetus in abdominal preg, second tri, fetus 3 |
|  | DX_ICD10 | O36.72X4 | mat care for viable fetus in abdominal preg, second tri, fetus 4 |
|  | DX_ICD10 | O36.72X5 | mat care for viable fetus in abdominal preg, second tri, fetus 5 |
|  | DX_ICD10 | O36.72X9 | mat care for viable fetus in abdominal preg, second tri, other fetus |
|  | DX_ICD10 | O36.73 | mat care for viable fetus in abdominal preg, third tri |
|  | DX_ICD10 | O36.73X0 | mat care for viable fetus in abdominal preg, third tri, na or unspecif |
|  | DX_ICD10 | O36.73X1 | mat care for viable fetus in abdominal preg, third tri, fetus 1 |
|  | DX_ICD10 | O36.73X2 | mat care for viable fetus in abdominal preg, third tri, fetus 2 |
|  | DX_ICD10 | O36.73X3 | mat care for viable fetus in abdominal preg, third tri, fetus 3 |
|  | DX_ICD10 | O36.73X4 | mat care for viable fetus in abdominal preg, third tri, fetus 4 |
|  | DX_ICD10 | O36.73X5 | mat care for viable fetus in abdominal preg, third tri, fetus 5 |
|  | DX_ICD10 | O36.73X9 | mat care for viable fetus in abdominal preg, third tri, other fetus |
|  | PX_ICD10 | 10D27ZZ | extraction products concept, ectopic, via nat or artific opening |
|  | PX_ICD10 | 10D28ZZ | extraction products concept, ectop, via nat or artif opening endoscop |
|  | PX_ICD10 | 10J20ZZ | inspection of products conception, ectopic, open approach |
|  | PX_ICD10 | 10J23ZZ | inspection of products conception, ectopic, percutaneous approach |
|  | PX_ICD10 | 10J24ZZ | inspection of products concept ectopic, percutan endoscopic approach |
|  | PX_ICD10 | 10J27ZZ | inspection of products conception, ectopic, via nat or artific opening |
|  | PX_ICD10 | 10J28ZZ | inspect products concept, ectopic, via nat or artific opening endoscopic |
|  | PX_ICD10 | 10J2XZZ | inspect products of concept, ectopic, external approach |
|  | PX_ICD10 | 10S20ZZ | reposit products of concept, ectopic, open approach |
|  | PX_ICD10 | 10S23ZZ | reposit products of concept, ectopic, percutaneous approach |
|  | PX_ICD10 | 10S24ZZ | reposit products concept, ectopic, percutaneous endoscopic approach |
|  | PX_ICD10 | 10S27ZZ | reposit products conception, ectopic, via natural or artif opening |
|  | PX_ICD10 | 10S28ZZ | reposit products conception, ectopic, via natural or artif opening endos |
|  | PX_ICD10 | 10T20ZZ | resect products conception, ectopic, open approach |
|  | PX_ICD10 | 10T23ZZ | resect products concept, ectopic, percutaneous approach |
|  | PX_ICD10 | 10T23ZZ | resection products concept, ectopic, percutaneous approach |
|  | PX_ICD10 | 10T24ZZ | resection products concept, ectopic, percutaneous endoscopic approach |
|  | PX_ICD10 | 10T24ZZ | resection products concept, ectopic, percutaneous endoscopic approach |
|  | PX_ICD10 | 10T27ZZ | resection products conception, ectopic, via nat or artificial opening |
|  | PX_ICD10 | 10T28ZZ | resect products concept, ectopic, via natural or artificial opening endo |
|  | PX_CPT | 59120 | surg tx of ectop preg; tub or ovarian, req salpingect &/or oophorect ab |
|  | PX_CPT | 59121 | surg tx ectop preg; tubal or ovarian, wo salpingect and/or oophorect |
|  | PX_CPT | 59130 | surgical tx of ectopic preg; abdominal preg |
|  | PX_CPT | 59135 | surgical tx of ectop preg; interstitial, uterine preg req tot hysterectomy |
|  | PX_CPT | 59136 | surg tx of ectop preg; interstitial, uterine preg w partial resect uterus |
|  | PX_CPT | 59140 | surgical tx of ectopic preg; cervical, with evacuation |
|  | PX_CPT | 59150 | laparoscopic tx of ectopic preg; wo salpingect and/or oophorectomy |
| **Spontaneous Abortion (SAB)**  42 ICD-9 DX 64 ICD-10 DX 5 CPT **111 Total** | DX_ICD9 | 634.00 | SAB w pelvic infection-unspecified |
|  | DX_ICD9 | 634.01 | SAB w pelvic infection-incomplete |
|  | DX_ICD9 | 634.02 | SAB w pelvic infection-complete |
|  | DX_ICD9 | 634.10 | SAB w hemorrhage-unspecified |
|  | DX_ICD9 | 634.11 | SAB w hemorrhage-incomplete |
|  | DX_ICD9 | 634.12 | SAB w hemorrhage complete |
|  | DX_ICD9 | 634.20 | SAB w pelvic damage-unspecified |
|  | DX_ICD9 | 634.21 | SAB w pelvic damage-incomplete |
|  | DX_ICD9 | 634.22 | SAB w pelvic damage-complete |
|  | DX_ICD9 | 634.30 | SAB w renal failure-unspecified |
|  | DX_ICD9 | 634.31 | SAB w renal failure-incomplete |
|  | DX_ICD9 | 634.32 | SAB w renal failure-complete |
|  | DX_ICD9 | 634.40 | SAB w metabolic disease-unspecified |
|  | DX_ICD9 | 634.41 | SAB w metabolic disease-incomplete |
|  | DX_ICD9 | 634.42 | SAB w metabolic disease |
|  | DX_ICD9 | 634.50 | SAB w shock-unspecified |
|  | DX_ICD9 | 634.51 | SAB w shock-incomplete |
|  | DX_ICD9 | 634.52 | SAB w shock-complete |
|  | DX_ICD9 | 634.60 | SAB w embolism-unspecified |
|  | DX_ICD9 | 634.61 | SAB w embolism-incomplete |
|  | DX_ICD9 | 634.62 | SAB w embolism-complete |
|  | DX_ICD9 | 634.70 | SAB w complication nec-unspecified |
|  | DX_ICD9 | 634.71 | SAB, w other specified complications, incomplete |
|  | DX_ICD9 | 634.72 | SAB w complication nec-complete |
|  | DX_ICD9 | 634.80 | SAB w complication nos-unspecified |
|  | DX_ICD9 | 634.81 | SAB w complication nos-incomplete |
|  | DX_ICD9 | 634.82 | SAB w complication nos-complete |
|  | DX_ICD9 | 634.90 | SAB uncomplicating-unspecified |
|  | DX_ICD9 | 634.91 | SAB uncomplicated-incomplete |
|  | DX_ICD9 | 634.92 | SAB uncomplicated-complete |
|  | DX_ICD9 | 651.30 | twins w fetal loss-unspecified(begin 1989) |
|  | DX_ICD9 | 651.31 | twins w fetal loss & retent 1 fetus, delivered, w or w/o antepartum |
|  | DX_ICD9 | 651.33 | twins w fetal loss-antepartum (begin 1989) |
|  | DX_ICD9 | 651.40 | triplets w fetal loss-unspecified (begin 1989) |
|  | DX_ICD9 | 651.41 | triplets w fetal loss & retent 1+ fetus(es), del, w or w/o antepartum |
|  | DX_ICD9 | 651.43 | triplets w fetal loss-antepartum (begin 1989) |
|  | DX_ICD9 | 634.70 | SAB w complication nec-unspecified 63470 |
|  | DX_ICD9 | 634.71 | SAB, w other specified complications, incomplete |
|  | DX_ICD9 | 634.72 | SAB w complication nec-complete |
|  | DX_ICD9 | 634.80 | SAB w complication nos-unspecified |
|  | DX_ICD9 | 634.81 | SAB w complication nos-incomplete |
|  | DX_ICD9 | 634.82 | SAB w complication nos-complete |
|  | DX_ICD9 | 634.90 | SAB complication-unspecified |
|  | DX_ICD9 | 634.91 | SAB complication-incomplete |
|  | DX_ICD9 | 634.92 | SAB complication-complete |
|  | DX_ICD9 | 651.30 | twins w fetal loss-unspecif (begin 1989) |
|  | DX_ICD9 | 651.31 | twins w fetal loss & retent 1 fetus, del, w or w/o antepartum |
|  | DX_ICD9 | 651.33 | twins w fetal loss-antepartum (begin 1989) |
|  | DX_ICD9 | 651.40 | triplets w fetal loss-unspecif (begin 1989) |
|  | DX_ICD9 | 651.41 | triplets w fetal loss & retent of 1+ fetus(es), del, w or w/o antepartum |
|  | DX_ICD9 | 651.43 | triplets w fetal loss-antepartum (begin 1989) |
|  | DX_ICD9 | 651.50 | quads w fetal loss-unspecified (begin 1989) |
|  | DX_ICD9 | 651.51 | quadruplet w fetal loss & retent 1+ fetus(es), del, w or w/o antepartum |
|  | DX_ICD9 | 651.53 | quads w fetal loss-antepartum (begin 1989) |
|  | DX_ICD9 | 651.60 | multiple gestation w fetal loss-unspecified (begin 1989) |
|  | DX_ICD9 | 651.61 | multiple gestation w fetal loss-del (begin 1989) |
|  | DX_ICD9 | 651.63 | multiple gestation w fetal loss-antepartum (begin 1989) |
|  | DX_ICD10 | O03 | SAB |
|  | DX_ICD10 | O03.0 | genital tract and pelvic infection after incomplete SAB |
|  | DX_ICD10 | O03.1 | delayed or excessive hemorrhage after incomplete SAB |
|  | DX_ICD10 | O03.2 | embolism following incomplete SAB |
|  | DX_ICD10 | O03.3 | genital tract and pelvic infection following incomplete SAB |
|  | DX_ICD10 | O03.30 | unspecified complication following incomplete SAB |
|  | DX_ICD10 | O03.31 | shock following incomplete SAB |
|  | DX_ICD10 | O03.32 | renal failure following incomplete SAB |
|  | DX_ICD10 | O03.33 | metabolic disorder following incomplete SAB |
|  | DX_ICD10 | O03.34 | damage to pelvic organs following incomplete SAB |
|  | DX_ICD10 | O03.35 | other venous complications following incomplete SAB |
|  | DX_ICD10 | O03.36 | cardiac arrest following incomplete SAB |
|  | DX_ICD10 | O03.37 | sepsis following incomplete SAB |
|  | DX_ICD10 | O03.38 | urinary tract infection following incomplete SAB |
|  | DX_ICD10 | O03.39 | incomplete SAB with other complications |
|  | DX_ICD10 | O03.4 | incomplete SAB without complication |
|  | DX_ICD10 | O03.5 | genital tract and pelvic infection following complete or unspecif SAB |
|  | DX_ICD10 | O03.6 | delayed or excessive hemorrhage following complete or unspecif SAB |
|  | DX_ICD10 | O03.7 | embolism following complete or unspecified SAB |
|  | DX_ICD10 | O03.8 | other and unspecif complications following complete or unspecif SAB |
|  | DX_ICD10 | O03.80 | unspecified complication following complete or unspecified SAB |
|  | DX_ICD10 | O03.81 | shock following complete or unspecified SAB |
|  | DX_ICD10 | O03.82 | renal failure following complete or unspecified SAB |
|  | DX_ICD10 | O03.83 | metabolic disorder following complete or unspecified SAB |
|  | DX_ICD10 | O03.84 | damage to pelvic organs following complete or unspecified SAB |
|  | DX_ICD10 | O03.85 | other venous complications following complete or unspecified SAB |
|  | DX_ICD10 | O03.86 | cardiac arrest following complete or unspecified SAB |
|  | DX_ICD10 | O03.87 | sepsis following complete or unspecified SAB |
|  | DX_ICD10 | O03.88 | urinary tract infection following complete or unspecified SAB |
|  | DX_ICD10 | O03.89 | complete or unspecified SAB with other complications |
|  | DX_ICD10 | O03.9 | complete or unspecified SAB without complication |
|  | DX_ICD10 | O31.1 | continuing preg after SAB 1+ fetus |
|  | DX_ICD10 | O31.10 | continuing preg after SAB 1+ fetus, unspecified tri |
|  | DX_ICD10 | O31.10X0 | continuing preg after SAB 1+ fetus, unspecified tri, na or unspecified |
|  | DX_ICD10 | O31.10X1 | continuing preg after SAB 1+ fetus, unspecified tri, fetus 1 |
|  | DX_ICD10 | O31.10X2 | continuing preg after SAB 1+ fetus, unspecified tri, fetus 2 |
|  | DX_ICD10 | O31.10X3 | continuing preg after SAB 1+ fetus, unspecified tri, fetus 3 |
|  | DX_ICD10 | O31.10X4 | continuing preg after SAB 1+ fetus, unspecified tri, fetus 4 |
|  | DX_ICD10 | O31.10X5 | continuing preg after SAB 1+ fetus, unspecified tri, fetus 5 |
|  | DX_ICD10 | O31.10X9 | continuing preg after SAB 1+ fetus, unspecified tri, other fetus |
|  | DX_ICD10 | O31.11 | continuing preg after SAB 1+ fetus, first tri |
|  | DX_ICD10 | O31.11X0 | continuing preg after SAB 1+ fetus, first tri, na or unspecified |
|  | DX_ICD10 | O31.11X1 | continuing preg after SAB 1+ fetus, first tri, fetus 1 |
|  | DX_ICD10 | O31.11X2 | continuing preg after SAB 1+ fetus, first tri, fetus 2 |
|  | DX_ICD10 | O31.11X3 | continuing preg after SAB 1+ fetus, first tri, fetus 3 |
|  | DX_ICD10 | O31.11X4 | continuing preg after SAB 1+ fetus, first tri, fetus 4 |
|  | DX_ICD10 | O31.11X5 | continuing preg after SAB 1+ fetus, first tri, fetus 5 |
|  | DX_ICD10 | O31.11X9 | continuing preg after SAB 1+ fetus, first tri, other fetus |
|  | DX_ICD10 | O31.12 | continuing preg after SAB 1+ fetus, second tri |
|  | DX_ICD10 | O31.12X0 | continuing preg after SAB 1+ fetus, second tri, na or unspecified |
|  | DX_ICD10 | O31.12X1 | continuing preg after SAB 1+ fetus, second tri, fetus 1 |
|  | DX_ICD10 | O31.12X2 | continuing preg after SAB 1+ fetus, second tri, fetus 2 |
|  | DX_ICD10 | O31.12X3 | continuing preg after SAB 1+ fetus, second tri, fetus 3 |
|  | DX_ICD10 | O31.12X4 | continuing preg after SAB 1+ fetus, second tri, fetus 4 |
|  | DX_ICD10 | O31.12X5 | continuing preg after SAB 1+ fetus, second tri, fetus 5 |
|  | DX_ICD10 | O31.12X9 | continuing preg after SAB 1+ fetus, second tri, other fetus |
|  | DX_ICD10 | O31.13 | continuing preg after SAB 1+ fetus, third tri |
|  | DX_ICD10 | O31.13X0 | continuing preg after SAB 1+ fetus, third tri, na or unspecified |
|  | DX_ICD10 | O31.13X1 | continuing preg after SAB 1+ fetus, third tri, fetus 1 |
|  | DX_ICD10 | O31.13X2 | continuing preg after SAB 1+ fetus, third tri, fetus 2 |
|  | DX_ICD10 | O31.13X3 | continuing preg after SAB 1+ fetus, third tri, fetus 3 |
|  | DX_ICD10 | O31.13X4 | continuing preg after SAB 1+ fetus, third tri, fetus 4 |
|  | DX_ICD10 | O31.13X5 | continuing preg after SAB 1+ fetus, third tri, fetus 5 |
|  | DX_ICD10 | O31.13X9 | continuing preg after SAB 1+ fetus, third tri, other fetus |
|  | PX_CPT | 1965 | anesthesia for incomplete or missed abortion procedures |
|  | PX_CPT | 59812 | tx of incomplete abortion, any tri, completed surgically |
|  | PX_CPT | 59820 | tx of missed abortion, completed surgically; first tri |
|  | PX_CPT | 59821 | tx of missed abortion, completed surgically; second tri |
|  | PX_CPT | 59830 | tx of septic abortion, completed surgically |
| **Therapeutic Abortion** 95 ICD-9 DX 4 ICD-9 PX 64 ICD-10 DX 8 ICD-10 PX 10 CPT  **181 Total** | DX_ICD9 | 635.00 | legal abortion w pelvic infection-unspecified |
|  | DX_ICD9 | 635.01 | legal abortion w pelvic infection-incomplete |
|  | DX_ICD9 | 635.02 | legal abortion w pelvic infection-complete |
|  | DX_ICD9 | 635.10 | legal abortion w hemorrhage-unspecified |
|  | DX_ICD9 | 635.11 | legal abortion w hemorrhage-incomplete |
|  | DX_ICD9 | 635.12 | legal abortion w hemorrhage-complete |
|  | DX_ICD9 | 635.20 | legal abortion w pelvic damage-unspecified |
|  | DX_ICD9 | 635.21 | legal abortion w pelvic damage-incomplete |
|  | DX_ICD9 | 635.22 | legal abortion w pelvic damage-complete |
|  | DX_ICD9 | 635.30 | legal abortion w renal failure-unspecified |
|  | DX_ICD9 | 635.31 | legal abortion w renal failure-incomplete |
|  | DX_ICD9 | 635.32 | legal abortion w renal failure-complete |
|  | DX_ICD9 | 635.40 | legal abortion w metabolic disease-unspecified |
|  | DX_ICD9 | 635.41 | legal abortion w metabolic disease-incomplete |
|  | DX_ICD9 | 635.42 | legal abortion w metabolic disease-complete |
|  | DX_ICD9 | 635.50 | legal abortion w shock-unspecified |
|  | DX_ICD9 | 635.51 | legal abortion w shock-incomplete |
|  | DX_ICD9 | 635.52 | legal abortion w shock-complete |
|  | DX_ICD9 | 635.60 | legal abortion w embolism-unspecified |
|  | DX_ICD9 | 635.61 | legal abortion w embolism-incomplete |
|  | DX_ICD9 | 635.62 | legal abortion w embolism-complete |
|  | DX_ICD9 | 635.70 | legal abortion w complication nec-unspecified |
|  | DX_ICD9 | 635.71 | legal abortion w complication nec-incomplete |
|  | DX_ICD9 | 635.72 | legal abortion w complication nec-complete |
|  | DX_ICD9 | 635.80 | legal abortion w complication nos-unspecified |
|  | DX_ICD9 | 635.81 | legal abortion w complication nos-incomplete |
|  | DX_ICD9 | 635.82 | legal abortion w complication nos-complete |
|  | DX_ICD9 | 636.00 | illegal abortion w pelvic infection-unspecified |
|  | DX_ICD9 | 636.01 | illegal abortion w pelvic infection-incomplete |
|  | DX_ICD9 | 636.02 | illegal abortion w pelvic infection-complete |
|  | DX_ICD9 | 636.10 | illegal abortion w hemorrhage-unspecified |
|  | DX_ICD9 | 636.11 | illegal abortion w hemorrhage-incomplete |
|  | DX_ICD9 | 636.12 | illegal abortion w hemorrhage-complete |
|  | DX_ICD9 | 636.20 | illegal abortion w pelvic damage-unspecified |
|  | DX_ICD9 | 636.21 | illegal abortion w pelvic damage-incomplete |
|  | DX_ICD9 | 636.22 | illegal abortion w pelvic damage-complete |
|  | DX_ICD9 | 636.30 | illegal abortion w renal failure-unspecified |
|  | DX_ICD9 | 636.31 | illegal abortion w renal failure-incomplete |
|  | DX_ICD9 | 636.32 | illegal abortion w renal failure-complete |
|  | DX_ICD9 | 636.40 | illegal abortion w metabolic disease-unspecified |
|  | DX_ICD9 | 636.41 | illegal abortion w metabolic disease-incomplete |
|  | DX_ICD9 | 636.42 | illegal abortion w metabolic disease-complete |
|  | DX_ICD9 | 636.50 | illegal abortion w shock-unspecified |
|  | DX_ICD9 | 636.51 | illegal abortion w shock-incomplete |
|  | DX_ICD9 | 636.52 | illegal abortion w shock-complete |
|  | DX_ICD9 | 636.60 | illegal abortion w embolism-unspecified |
|  | DX_ICD9 | 636.61 | illegal abortion w embolism-incomplete |
|  | DX_ICD9 | 636.62 | illegal abortion w embolism-complete |
|  | DX_ICD9 | 636.70 | illegal abortion w complication nec-unspecified |
|  | DX_ICD9 | 636.71 | illegal abortion w complication nec-incomplete |
|  | DX_ICD9 | 636.72 | illegal abortion w complication nec-complete |
|  | DX_ICD9 | 636.80 | illegal abortion w complication nos-unspecified |
|  | DX_ICD9 | 636.81 | illegal abortion w complication nos-incomplete |
|  | DX_ICD9 | 636.82 | illegal abortion w complication nos-complete |
|  | DX_ICD9 | 637.00 | abortion nos w pelvic infection-unspecified |
|  | DX_ICD9 | 637.01 | abortion nos w pelvic infection-incomplete |
|  | DX_ICD9 | 637.02 | abortion nos w pelvic infection-complete |
|  | DX_ICD9 | 637.10 | abortion nos w hemorrhage-unspecified |
|  | DX_ICD9 | 637.11 | abortion nos w hemorrhage-incomplete |
|  | DX_ICD9 | 637.12 | abortion nos w hemorrhage-complete |
|  | DX_ICD9 | 637.20 | abortion nos w pelvic damage-unspecified |
|  | DX_ICD9 | 637.21 | abortion nos w pelvic damage-incomplete |
|  | DX_ICD9 | 637.22 | abortion nos w pelvic damage-complete |
|  | DX_ICD9 | 637.30 | abortion nos w renal failure-unspecified |
|  | DX_ICD9 | 637.31 | abortion nos w renal failure-incomplete |
|  | DX_ICD9 | 637.32 | abortion nos w renal failure-complete |
|  | DX_ICD9 | 637.40 | abortion nos w metabolic disease-unspecified |
|  | DX_ICD9 | 637.41 | abortion nos w metabolic disease-incomplete |
|  | DX_ICD9 | 637.42 | abortion nos w metabolic disease-complete |
|  | DX_ICD9 | 637.50 | abortion nos w shock-unspecified |
|  | DX_ICD9 | 637.51 | abortion nos w shock-incomplete |
|  | DX_ICD9 | 637.52 | abortion nos w shock- complete |
|  | DX_ICD9 | 637.60 | abortion nos w embolism-unspecified |
|  | DX_ICD9 | 637.61 | abortion nos w embolism-incomplete |
|  | DX_ICD9 | 637.62 | abortion nos w embolism-complete |
|  | DX_ICD9 | 637.70 | abortion nos w complication nec unspecified |
|  | DX_ICD9 | 637.71 | abortion nos w complication nec-incomplete |
|  | DX_ICD9 | 637.72 | abortion nos w complication nec-complete |
|  | DX_ICD9 | 637.80 | abortion nos w complication nos-unspecified |
|  | DX_ICD9 | 637.81 | abortion nos w complication nos-incomplete |
|  | DX_ICD9 | 637.82 | abortion nos w complication nos-complete |
|  | DX_ICD9 | 638.0 | attempted abortion w pelvic infection |
|  | DX_ICD9 | 638.1 | attempted abortion w hemorrhage |
|  | DX_ICD9 | 638.2 | attempted abortion w pelvic damage |
|  | DX_ICD9 | 638.3 | attempted abortion w renal failure |
|  | DX_ICD9 | 638.4 | attempted abortion w metabolic disease |
|  | DX_ICD9 | 638.5 | attempted abortion w shock |
|  | DX_ICD9 | 638.6 | attempted abortion w embolism |
|  | DX_ICD9 | 638.7 | attempted abortion w complication nec |
|  | DX_ICD9 | 638.8 | attempted abortion w complication nos |
|  | DX_ICD9 | 638.9 | attempted abort uncomplication |
|  | DX_ICD9 | 651.70 | multiple gestation-fetal reduction-unspecified (begin 2005) |
|  | DX_ICD9 | 651.71 | multiple gestation-fetal reduction-delivered (begin 2005) |
|  | DX_ICD9 | 651.73 | multiple gestation-fetal reduction-antepartum (begin 2005) |
|  | DX_ICD9 | 779.6 | termination of pregnancy |
|  | PX_ICD9 | 69.01 | dilation and curettage for termination of pregnancy |
|  | PX_ICD9 | 69.51 | aspiration curettage of uterus for termination of pregnancy |
|  | PX_ICD9 | 74.91 | hysterotomy to terminate preg |
|  | PX_ICD9 | 75.0 | intra-amniotic injection for abortion |
|  | DX_ICD10 | O04 | complications following (induced) termination of preg |
|  | DX_ICD10 | O04.5 | genital tract and pelvic infection after (induced) termination of preg |
|  | DX_ICD10 | O04.6 | delayed or excessive hemorrhage after (induced) termination of preg |
|  | DX_ICD10 | O04.7 | embolism following (induced) termination of pregnancy |
|  | DX_ICD10 | O04.8 | (induced) termination of preg w other and unspecified complications |
|  | DX_ICD10 | O04.80 | (induced) termination of preg w unspecified complications |
|  | DX_ICD10 | O04.81 | shock following (induced) termination of pregnancy |
|  | DX_ICD10 | O04.82 | renal failure following (induced) termination of pregnancy |
|  | DX_ICD10 | O04.83 | metabolic disorder following (induced) termination of pregnancy |
|  | DX_ICD10 | O04.84 | damage to pelvic organs following (induced) termination of pregnancy |
|  | DX_ICD10 | O04.85 | other venous complications following (induced) termination of preg |
|  | DX_ICD10 | O04.86 | cardiac arrest following (induced) termination of pregnancy |
|  | DX_ICD10 | O04.87 | sepsis following (induced) termination of pregnancy |
|  | DX_ICD10 | O04.88 | urinary tract infection following (induced) termination of pregnancy |
|  | DX_ICD10 | O04.89 | (induced) termination of preg with other complications |
|  | DX_ICD10 | O07 | failed attempted termination of pregnancy |
|  | DX_ICD10 | O07.0 | genital tract and pelvic infection after failed attempted termin preg |
|  | DX_ICD10 | O07.1 | delayed or excessive hemorrhage after failed attempted termin preg |
|  | DX_ICD10 | O07.2 | embolism following failed attempted termination of pregnancy |
|  | DX_ICD10 | O07.3 | failed attempted termin preg w other and unspecified complications |
|  | DX_ICD10 | O07.30 | failed attempted termination of preg w unspecified complications |
|  | DX_ICD10 | O07.31 | shock following failed attempted termination pregnancy |
|  | DX_ICD10 | O07.32 | renal failure following failed attempted termination of pregnancy |
|  | DX_ICD10 | O07.33 | metabolic disorder after failed attempted termination of pregnancy |
|  | DX_ICD10 | O07.34 | damage to pelvic organs after failed attempted termination of preg |
|  | DX_ICD10 | O07.35 | other venous complications following failed attempted termin preg |
|  | DX_ICD10 | O07.36 | cardiac arrest following failed attempted termination of pregnancy |
|  | DX_ICD10 | O07.37 | sepsis following failed attempted termination of pregnancy |
|  | DX_ICD10 | O07.38 | urinary tract infection following failed attempted termination of preg |
|  | DX_ICD10 | O07.39 | failed attempted termination of pregnancy w other complications |
|  | DX_ICD10 | O07.4 | failed attempted termination of pregnancy wo complication |
|  | DX_ICD10 | O31.3 | continuing preg after elective fetal reduct 1+ fetus |
|  | DX_ICD10 | O31.30 | contin preg after elective fetal reduct 1+ fetus, unspec tri |
|  | DX_ICD10 | O31.30X0 | contin preg after elective fetal reduct 1+ fetus, unspec tri, na or unspec |
|  | DX_ICD10 | O31.30X1 | contin preg after elective fetal reduct 1+ fetus, unspec tri, fetus 1 |
|  | DX_ICD10 | O31.30X2 | contin preg after elective fetal reduct 1+ fetus, unspec tri, fetus 2 |
|  | DX_ICD10 | O31.30X3 | contin preg after elective fetal reduct 1+ fetus, unspec tri, fetus 3 |
|  | DX_ICD10 | O31.30X4 | contin preg after elective fetal reduct 1+ fetus, unspec tri, fetus 4 |
|  | DX_ICD10 | O31.30X5 | contin preg after elective fetal reduct 1+ fetus, unspec tri, fetus 5 |
|  | DX_ICD10 | O31.30X9 | contin preg after elective fetal reduct 1+ fetus, unspec tri, other fetus |
|  | DX_ICD10 | O31.31 | contin preg after elective fetal reduct 1+ fetus, first tri |
|  | DX_ICD10 | O31.31X0 | contin preg after elective fetal reduct 1+ fetus, first tri, na or unspecif |
|  | DX_ICD10 | O31.31X1 | contin preg after elective fetal reduct 1+ fetus, first tri, fetus 1 |
|  | DX_ICD10 | O31.31X3 | continuing preg after elective fetal reduct 1+ fetus, first tri, fetus 3 |
|  | DX_ICD10 | O31.31X4 | continuing preg after elective fetal reduct 1+ fetus, first tri, fetus 4 |
|  | DX_ICD10 | O31.31X5 | continuing preg after elective fetal reduct 1+ fetus, first tri, fetus 5 |
|  | DX_ICD10 | O31.31X9 | continuing preg after elective fetal reduct 1+ fetus, first tri, other fetus |
|  | DX_ICD10 | O31.32 | continuing preg after elective fetal reduct 1+ fetus, second tri |
|  | DX_ICD10 | O31.32X0 | contin preg after elect fetal reduct 1+ fetus, second tri, na or unspecif |
|  | DX_ICD10 | O31.32X1 | continuing preg after elective fetal reduct 1+ fetus, second tri, fetus 1 |
|  | DX_ICD10 | O31.32X2 | continuing preg after elective fetal reduct 1+ fetus, second tri, fetus 2 |
|  | DX_ICD10 | O31.32X3 | continuing preg after elective fetal reduct 1+ fetus, second tri, fetus 3 |
|  | DX_ICD10 | O31.32X4 | continuing preg after elective fetal reduct 1+ fetus, second tri, fetus 4 |
|  | DX_ICD10 | O31.32X5 | continuing preg after elective fetal reduct 1+ fetus, second tri, fetus 5 |
|  | DX_ICD10 | O31.32X9 | contin preg after elective fetal reduct 1+ fetus, second tri, other fetus |
|  | DX_ICD10 | O31.33 | continuing preg after elective fetal reduct 1+ fetus, third tri |
|  | DX_ICD10 | O31.33X0 | contin preg after elective fetal reduct 1+ fetus, third tri, na or unspecif |
|  | DX_ICD10 | O31.33X1 | continuing preg after elective fetal reduct 1+ fetus, third tri, fetus 1 |
|  | DX_ICD10 | O31.33X2 | continuing preg after elective fetal reduct 1+ fetus, third tri, fetus 2 |
|  | DX_ICD10 | O31.33X3 | continuing preg after elective fetal reduct 1+ fetus, third tri, fetus 3 |
|  | DX_ICD10 | O31.33X4 | continuing preg after elective fetal reduct 1+ fetus, third tri, fetus 4 |
|  | DX_ICD10 | O31.33X5 | continuing preg after elective fetal reduct 1+ fetus, third tri, fetus 5 |
|  | DX_ICD10 | O31.33X9 | continuing preg after elective fetal reduct 1+ fetus, third tri, other fetus |
|  | PX_ICD10 | 10A00ZZ | abortion of products of conception, open approach |
|  | PX_ICD10 | 10A03ZZ | abortion of products of conception, percutaneous approach |
|  | PX_ICD10 | 10A04ZZ | abortion of products of conception, percutaneous endoscopic approach |
|  | PX_ICD10 | 10A07Z6 | abortion products conception, vacuum, via natural or artificial opening |
|  | PX_ICD10 | 10A07ZW | abortion products conception, laminaria, via nat or artific opening |
|  | PX_ICD10 | 10A07ZX | abortion products conception, abortifacient, via nat or artific opening |
|  | PX_ICD10 | 10A07ZZ | abortion of products of conception, via natural or artificial opening |
|  | PX_ICD10 | 10A08ZZ | abort products concept, via nat or artific opening endoscopic |
|  | PX_CPT | 1966 | anesthesia for induc abort procedures |
|  | PX_CPT | 59151 | laparoscopic tx of ectop preg; with salpingect and/or oophorect |
|  | PX_CPT | 59840 | induc abort, by dilation and curettage |
|  | PX_CPT | 59841 | induc abort, by dilation and evacuation |
|  | PX_CPT | 59850 | induc abort, by 1+ intra-amnio injects (amnio-injections), incl hosp ad |
|  | PX_CPT | 59851 | induc abort, by 1+ intra-amnio injects (amnio-injections), incl hosp ad |
|  | PX_CPT | 59852 | induc abort, by 1+intra-amnio injects (amnio-injections), incl hosp ad |
|  | PX_CPT | 59855 | induc abort, by 1+ vag suppositor (eg, prostaglandin) w or wo cerv dila |
|  | PX_CPT | 59856 | induc abort |
|  | PX_CPT | 59857 | induc abort |

|  | TYPE | CODE | DESCRIPTION |
| --- | --- | --- | --- |
