## Supplemental Table S5 for "Maternal and Fetal Complications Among Pregnant Women with Congenital Heart Disease"

| **Supplemental Table S5: ICD Codes to define maternal non-gestational comorbidities (840 codes)** | | | |
| --- | --- | --- | --- |
|  | **TYPE** | **CODE** | **DESCRIPTION** |
| **Arrhythmia** 42 ICD-9 DX 52 ICD-10 DX **94 Total** | DX_ICD9 | 426.0 | atriovent block complete |
|  | DX_ICD9 | 426.10 | atriovent block nos |
|  | DX_ICD9 | 426.11 | atriovent block-1st degr |
|  | DX_ICD9 | 426.12 | atrioven block-mobitz ii |
|  | DX_ICD9 | 426.13 | av block-2nd degree nec |
|  | DX_ICD9 | 426.5 | bundle branch block nos |
|  | DX_ICD9 | 426.51 | rt bbb/lft post fasc blk |
|  | DX_ICD9 | 426.52 | rt bbb/lft ant fasc blk |
|  | DX_ICD9 | 426.53 | bilat bb block nec |
|  | DX_ICD9 | 426.54 | trifascicular block |
|  | DX_ICD9 | 426.6 | other heart block |
|  | DX_ICD9 | 426.7 | anomalous av excitation |
|  | DX_ICD9 | 426.81 | lown-ganong-levine synd |
|  | DX_ICD9 | 426.82 | long qt syndrome (begin 2005) |
|  | DX_ICD9 | 426.89 | conduction disorder nec |
|  | DX_ICD9 | 426.9 | conduction disorder nos |
|  | DX_ICD9 | 427.0 | parox atrial tachycardia |
|  | DX_ICD9 | 427.1 | parox ventric tachycard |
|  | DX_ICD9 | 427.31 | atrial fibrillation |
|  | DX_ICD9 | 427.32 | atrial flutter |
|  | DX_ICD9 | 427.41 | ventricular fibrillation |
|  | DX_ICD9 | 427.42 | ventricular flutter |
|  | DX_ICD9 | 427.5 | cardiac arrest |
|  | DX_ICD9 | 427.61 | atrial premature beats |
|  | DX_ICD9 | 427.81 | sinoatrial node dysfunct |
|  | DX_ICD9 | 427.89 | cardiac dysrhythmias nec |
|  | DX_ICD9 | 427.9 | cardiac dysrhythmia nos |
|  | DX_ICD9 | 746.86 | congenital heart block |
|  | DX_ICD9 | 798.0 | sudden infant death synd |
|  | DX_ICD9 | 798.1 | instantaneous death |
|  | DX_ICD9 | 798.2 | death within 24 hr sympt |
|  | DX_ICD9 | 798.9 | unattended death |
|  | DX_ICD9 | V12.53 | hx sudden cardiac arrest (begin 2007) |
|  | DX_ICD9 | V45.0 | cardiac pacemaker status (end 1994) |
|  | DX_ICD9 | V45.00 | cardiac device nos in situ (begin 1994) |
|  | DX_ICD9 | V45.01 | cardiac pacem in situ (begin 1994) |
|  | DX_ICD9 | V45.02 | auto implant debril in situ (begin 1994) |
|  | DX_ICD9 | V45.09 | oth cardiac device nec in situ (begin 1994) |
|  | DX_ICD9 | V53.3 | adjust cardiac pacemaker (end 1994) |
|  | DX_ICD9 | V53.31 | adjust cardiac pacemaker (begin 1994) |
|  | DX_ICD9 | V53.32 | adjust auto implant debril (begin 1994) |
|  | DX_ICD9 | V53.39 | adjust oth cardiac device (begin 1994) |
|  | DX_ICD10 | I44.1 | atrioventricular block, second degree |
|  | DX_ICD10 | I44.2 | atrioventricular block, complete |
|  | DX_ICD10 | I44.30 | unspecified atrioventricular block |
|  | DX_ICD10 | I44.39 | other atrioventricular block |
|  | DX_ICD10 | I44.4 | left anterior fascicular block |
|  | DX_ICD10 | I44.5 | left posterior fascicular block |
|  | DX_ICD10 | I44.60 | unspecified fascicular block |
|  | DX_ICD10 | I44.69 | other fascicular block |
|  | DX_ICD10 | I44.7 | left bundle-branch block, unspecified |
|  | DX_ICD10 | I45.0 | right fascicular block |
|  | DX_ICD10 | I45.10 | unspecified right bundle-branch block |
|  | DX_ICD10 | I45.19 | other right bundle-branch block |
|  | DX_ICD10 | I45.2 | bifascicular block |
|  | DX_ICD10 | I45.3 | trifascicular block |
|  | DX_ICD10 | I45.4 | nonspecific intraventricular block |
|  | DX_ICD10 | I45.5 | other specified heart block |
|  | DX_ICD10 | I45.6 | pre-excitation syndrome |
|  | DX_ICD10 | I45.81 | long qt syndrome |
|  | DX_ICD10 | I45.89 | other specified conduction disorders |
|  | DX_ICD10 | I45.9 | conduction disorder, unspecified |
|  | DX_ICD10 | I46.2 | cardiac arrest due to underlying cardiac condition |
|  | DX_ICD10 | I46.8 | cardiac arrest due to other underlying condition |
|  | DX_ICD10 | I46.9 | cardiac arrest, cause unspecified |
|  | DX_ICD10 | I47.0 | re-entry ventricular arrhythmia |
|  | DX_ICD10 | I47.1 | supraventricular tachycardia |
|  | DX_ICD10 | I47.2 | ventricular tachycardia |
|  | DX_ICD10 | I48.0 | paroxysmal atrial fibrillation |
|  | DX_ICD10 | I48.1 | persistent atrial fibrillation |
|  | DX_ICD10 | I48.11 | longstanding persistent atrial fibrillation |
|  | DX_ICD10 | I48.19 | other persistent atrial fibrillation |
|  | DX_ICD10 | I48.2 | chronic atrial fibrillation |
|  | DX_ICD10 | I48.20 | chronic atrial fibrillation, unspecified |
|  | DX_ICD10 | I48.21 | permanent atrial fibrillation |
|  | DX_ICD10 | I48.3 | typical atrial flutter |
|  | DX_ICD10 | I48.4 | atypical atrial flutter |
|  | DX_ICD10 | I48.91 | unspecified atrial fibrillation |
|  | DX_ICD10 | I48.92 | unspecified atrial flutter |
|  | DX_ICD10 | I49.01 | ventricular fibrillation |
|  | DX_ICD10 | I49.02 | ventricular flutter |
|  | DX_ICD10 | I49.2 | junctional premature depolarization |
|  | DX_ICD10 | I49.5 | sick sinus syndrome |
|  | DX_ICD10 | I49.8 | other specified cardiac arrhythmias |
|  | DX_ICD10 | I49.9 | cardiac arrhythmia, unspecified |
|  | DX_ICD10 | Q24.6 | congenital heart block |
|  | DX_ICD10 | Z45.010 | encounter for check & test cardiac pacemaker pulse generator [battery] |
|  | DX_ICD10 | Z45.018 | encounter for adjust & mgt oth part of cardiac pacemaker |
|  | DX_ICD10 | Z45.02 | encounter for adjust & mgt of automatic implantable cardiac defibrillator |
|  | DX_ICD10 | Z45.09 | encounter for adjustment and management of other cardiac device |
|  | DX_ICD10 | Z86.74 | personal history of sudden cardiac arrest |
|  | DX_ICD10 | Z95.0 | presence of cardiac pacemaker |
|  | DX_ICD10 | Z95.810 | presence of automatic (implantable) cardiac defibrillator |
|  | DX_ICD10 | Z95.9 | presence of cardiac and vascular implant and graft, unspecified |
| **Coronary Artery Disease** 48 ICD-9 DX 76 ICD-10 DX **124 Total** | DX_ICD9 | 410.0 | ami anterolateral wall (begin 1980 end 1989) |
|  | DX_ICD9 | 410.00 | ami anterolateral;unspec (begin 1989) |
|  | DX_ICD9 | 41001 | ami anterolateral- init (begin 1989) |
|  | DX_ICD9 | 410.02 | ami anterolateral;subseq (begin 1989) |
|  | DX_ICD9 | 410.1 | ami anterior wall nec (begin 1980 end 1989) |
|  | DX_ICD9 | 410.10 | ami anterior wall;unspec (begin 1989) |
|  | DX_ICD9 | 410.11 | ami anterior wall- init (begin 1989) |
|  | DX_ICD9 | 410.12 | ami anterior wall;subseq (begin 1989) |
|  | DX_ICD9 | 410.2 | ami inferolateral wall (begin 1980 end 1989) |
|  | DX_ICD9 | 410.20 | ami inferolateral;unspec (begin 1989) |
|  | DX_ICD9 | 410.21 | ami inferolateral- init (begin 1989) |
|  | DX_ICD9 | 410.22 | ami inferolateral;subseq (begin 1989) |
|  | DX_ICD9 | 410.3 | ami inferoposterior wall (begin 1980 end 1989) |
|  | DX_ICD9 | 410.30 | ami inferopost- unspec (begin 1989) |
|  | DX_ICD9 | 410.31 | ami inferopost- initial (begin 1989) |
|  | DX_ICD9 | 410.32 | ami inferopost- subseq (begin 1989) |
|  | DX_ICD9 | 410.4 | ami inferior wall nec (begin 1980 end 1989) |
|  | DX_ICD9 | 410.40 | ami inferior wall;unspec (begin 1989) |
|  | DX_ICD9 | 410.41 | ami inferior wall- init (begin 1989) |
|  | DX_ICD9 | 410.42 | ami inferior wall;subseq (begin 1989) |
|  | DX_ICD9 | 410.5 | ami lateral wall nec (begin 1980 end 1989) |
|  | DX_ICD9 | 410.50 | ami lateral nec- unspec (begin 1989) |
|  | DX_ICD9 | 410.51 | ami lateral nec- initial (begin 1989) |
|  | DX_ICD9 | 410.52 | ami lateral nec- subseq (begin 1989) |
|  | DX_ICD9 | 410.6 | true posterior infarct (begin 1980 end 1989) |
|  | DX_ICD9 | 410.60 | true post infarct;unspec (begin 1989) |
|  | DX_ICD9 | 410.61 | true post infarct- init (begin 1989) |
|  | DX_ICD9 | 410.62 | true post infarct;subseq (begin 1989) |
|  | DX_ICD9 | 410.7 | subendocardial infarct (begin 1980 end 1989) |
|  | DX_ICD9 | 410.70 | subendo infarct- unspec (begin 1989) |
|  | DX_ICD9 | 410.71 | subendo infarct- initial (begin 1989) |
|  | DX_ICD9 | 410.72 | subendo infarct- subseq (begin 1989) |
|  | DX_ICD9 | 410.8 | myocardial infarct nec (begin 1980 end 1989) |
|  | DX_ICD9 | 410.80 | ami nec- unspecified (begin 1989) |
|  | DX_ICD9 | 410.81 | ami nec- initial (begin 1989) |
|  | DX_ICD9 | 410.82 | ami nec- subsequent (begin 1989) |
|  | DX_ICD9 | 410.9 | myocardial infarct nos (begin 1980 end 1989) |
|  | DX_ICD9 | 410.90 | ami nos- unspecified (begin 1989) |
|  | DX_ICD9 | 410.91 | ami nos- initial (begin 1989) |
|  | DX_ICD9 | 410.92 | ami nos- subsequent (begin 1989) |
|  | DX_ICD9 | 411.0 | post mi syndrome |
|  | DX_ICD9 | 412 | old myocardial infarct |
|  | DX_ICD9 | 414.02 | coronary athero autolog vein (begin 1994) |
|  | DX_ICD9 | 429.71 | acq cardiac septl defect (begin 1989) |
|  | DX_ICD9 | 429.79 | other sequelae of mi nec (begin 1989) |
|  | DX_ICD9 | 429.2 | ascvd |
|  | DX_ICD9 | V45.81 | aortocoronary bypass |
|  | DX_ICD9 | V45.82 | ptca status (begin 1994) |
|  | DX_ICD10 | I20.0 | unstable angina |
|  | DX_ICD10 | I21.01 | st elevat (stemi) myocardial infarction left main coronary artery |
|  | DX_ICD10 | I21.02 | st elevat (stemi) myocardial infarction left anter descend coronary artery |
|  | DX_ICD10 | I21.09 | st elevat (stemi) myocardial infarction oth coronary artery of anterior wall |
|  | DX_ICD10 | I21.11 | st elevation (stemi) myocardial infarction right coronary artery |
|  | DX_ICD10 | I21.19 | st elevation (stemi) myocardial infarction oth coronary artery of inferior wall |
|  | DX_ICD10 | I21.21 | st elevation (stemi) myocardial infarction left circumflex coronary artery |
|  | DX_ICD10 | I21.29 | st elevation (stemi) myocardial infarction other sites |
|  | DX_ICD10 | I21.3 | st elevation (stemi) myocardial infarction of unspec site |
|  | DX_ICD10 | I21.4 | non-st elevation (nstemi) myocardial infarction |
|  | DX_ICD10 | I21.9 | acute myocardial infarction, unspecified |
|  | DX_ICD10 | I21.A1 | myocardial infarction type 2 |
|  | DX_ICD10 | I21.A9 | other myocardial infarction type |
|  | DX_ICD10 | I22.0 | subsequent st elevation (stemi) myocardial infarction of anterior wall |
|  | DX_ICD10 | I22.1 | subsequent st elevation (stemi) myocardial infarction of inferior wall |
|  | DX_ICD10 | I22.2 | subsequent non-st elevation (nstemi) myocardial infarction |
|  | DX_ICD10 | I22.8 | subsequent st elevation (stemi) myocardial infarction of other sites |
|  | DX_ICD10 | I22.9 | subsequent st elevation (stemi) myocardial infarction of unspecified site |
|  | DX_ICD10 | I23.0 | hemopericardium as current complic after acute myocardial infarction |
|  | DX_ICD10 | I23.1 | ASD as current complic after acute myocardial infarction |
|  | DX_ICD10 | I23.2 | VSD as current complic after acute myocardial infarction |
|  | DX_ICD10 | I23.3 | rupt cardiac wall wo hemopericardium after acute MI |
|  | DX_ICD10 | I23.6 | thrombosis of atrium, auricular appendage, ventricle complic acute MI |
|  | DX_ICD10 | I23.7 | postinfarction angina |
|  | DX_ICD10 | I23.8 | other current complications following acute myocardial infarction |
|  | DX_ICD10 | I24.0 | acute coronary thrombosis not resulting in myocardial infarction |
|  | DX_ICD10 | I24.1 | dresslers syndrome |
|  | DX_ICD10 | I24.8 | other forms of acute ischemic heart disease |
|  | DX_ICD10 | I24.9 | acute ischemic hrt disease, unspecified |
|  | DX_ICD10 | I25.10 | atheroscler hrt disease of native coronary artery without angina pectoris |
|  | DX_ICD10 | I25.110 | atheroscler hrt disease of native coronary artery w unstable angina pectoris |
|  | DX_ICD10 | I25.111 | atheroscler hrt dis native coronary artery w angina pectoris w doc spasm |
|  | DX_ICD10 | I25.118 | atheroscler hrt dis native coronary artery w oth forms of angina pectoris |
|  | DX_ICD10 | I25.119 | atheroscler hrt dis native coronary artery w unspec angina pectoris |
|  | DX_ICD10 | I25.2 | old myocardial infarction |
|  | DX_ICD10 | I25.41 | coronary artery aneurysm |
|  | DX_ICD10 | I25.42 | coronary artery dissection |
|  | DX_ICD10 | I25.5 | ischemic cardiomyopathy |
|  | DX_ICD10 | I25.6 | silent myocardial ischemia |
|  | DX_ICD10 | I25.700 | atheroscler coron art bypass graft(s), unspec, w unstable angina pectoris |
|  | DX_ICD10 | I25.701 | atheroscler coron art bypass graft(s), unspec, w angina pect w doc spasm |
|  | DX_ICD10 | I25.708 | atheroscler coron art bypass graft(s), unspec,w oth forms angina pectoris |
|  | DX_ICD10 | I25.709 | atheroscler coron art bypass graft(s), unspec, w unspec angina pectoris |
|  | DX_ICD10 | I25.710 | atheroscler autolog vein coron art bypass graft(s) w unstable angina pect |
|  | DX_ICD10 | I25.711 | atheroscl autolog vein coron art bypass graft(s) w angin pect w doc spasm |
|  | DX_ICD10 | I25.718 | atheroscler autolog vein coron art bypass graft(s) w oth forms angina pect |
|  | DX_ICD10 | I25.719 | atheroscler autologous vein coron art bypass graft(s) w unspec angin pect |
|  | DX_ICD10 | I25.720 | atheroscler autolog art coron art bypass graft(s) w unstable angina pectoris |
|  | DX_ICD10 | I25.721 | atheroscler autolog art coron art bypass graft(s) w angin pect w doc spasm |
|  | DX_ICD10 | I25.728 | atheroscler autolog art coron art bypass graft(s) w oth forms angina pect |
|  | DX_ICD10 | I25.729 | atheroscler autolog art coron art bypass graft(s) w unspec angina pectoris |
|  | DX_ICD10 | I25.730 | atheroscler nonautolog bio coron art bypass graft(s) w unstable angin pect |
|  | DX_ICD10 | I25.731 | atheroscler nonautolog bio cor art bypas graft(s) w angin pect w doc spasm |
|  | DX_ICD10 | I25.738 | atheroscler nonautolog bio coron art bypass graft(s) w oth forms angin pect |
|  | DX_ICD10 | I25.739 | atheroscler nonautolog bio coron art bypass graft(s) w unspec angina pect |
|  | DX_ICD10 | I25.790 | atheroscler oth coron art bypass graft(s) w unstable angina pectoris |
|  | DX_ICD10 | I25.791 | atheroscler oth coron art bypass graft(s) w angina pectoris w doc spasm |
|  | DX_ICD10 | I25.798 | atheroscler oth coron art bypass graft(s) w oth forms angina pectoris |
|  | DX_ICD10 | I25.799 | atheroscler other coron art bypass graft(s) w unspec angina pectoris |
|  | DX_ICD10 | I25.810 | atheroscler coron art bypass graft(s) wo angina pectoris |
|  | DX_ICD10 | I25.811 | atheroscler native coron art transplanted hrt wo angina pectoris |
|  | DX_ICD10 | I25.82 | chronic total occlusion of coronary artery |
|  | DX_ICD10 | I25.83 | coronary atherosclerosis due to lipid rich plaque |
|  | DX_ICD10 | I25.84 | coronary atherosclerosis due to calcified coronary lesion |
|  | DX_ICD10 | I25.89 | other forms of chronic ischemic heart disease |
|  | DX_ICD10 | I25.9 | chronic ischemic heart disease, unspecified |
|  | DX_ICD10 | I51.0 | cardiac septal defect, acquired |
|  | DX_ICD10 | I51.3 | intracardiac thrombosis, not elsewhere classified |
|  | DX_ICD10 | I70.0 | atherosclerosis of aorta |
|  | DX_ICD10 | T82.211D | breakdown (mechanical) of coron art bypass graft, subseq encounter |
|  | DX_ICD10 | T82.212D | displacement of coronary artery bypass graft, subsequent encounter |
|  | DX_ICD10 | T82.213D | leakage coron art bypass graft, subseq encounter |
|  | DX_ICD10 | T82.218D | oth mechanical complic coronary artery bypass graft, subseq encounter |
|  | DX_ICD10 | Z95.1 | presence of aortocoronary bypass graft |
|  | DX_ICD10 | Z95.5 | presence of coronary angioplasty implant and graft |
|  | DX_ICD10 | Z98.61 | coronary angioplasty status |
| **Diabetes** 47 ICD-9 DX 400 ICD-10 DX **447 Total** | DX_ICD9 | 249.00 | sec dm wo cmp nt st uncn (begin 2008) |
|  | DX_ICD9 | 249.10 | sec dm keto nt st uncntr (begin 2008) |
|  | DX_ICD9 | 249.20 | sec dm hpros nt st uncnr (begin 2008) |
|  | DX_ICD9 | 249.30 | sec dm ot cma nt st uncn (begin 2008) |
|  | DX_ICD9 | 249.40 | sec dm renl nt st uncntr (begin 2008) |
|  | DX_ICD9 | 249.50 | sec dm ophth nt st uncn (begin 2008) |
|  | DX_ICD9 | 249.60 | sec dm neuro nt st uncn (begin 2008) |
|  | DX_ICD9 | 249.70 | sec dm circ nt st uncntr (begin 2008) |
|  | DX_ICD9 | 249.80 | sec dm oth nt st uncontr (begin 2008) |
|  | DX_ICD9 | 249.90 | sec dm unsp nt st uncon (begin 2008) |
|  | DX_ICD9 | 250.00 | diabetes uncompl type ii |
|  | DX_ICD9 | 250.01 | diabetes uncompl type i |
|  | DX_ICD9 | 250.10 | diab ketoacidosis typ ii |
|  | DX_ICD9 | 250.11 | diab ketoacidosis type i |
|  | DX_ICD9 | 250.20 | dm hyperosm coma type ii |
|  | DX_ICD9 | 250.30 | diabetes coma nec typ ii |
|  | DX_ICD9 | 250.31 | diabetes coma nec type i |
|  | DX_ICD9 | 250.40 | diab renal manif type ii |
|  | DX_ICD9 | 250.41 | diab renal manif type i |
|  | DX_ICD9 | 250.50 | diab eye manif type ii |
|  | DX_ICD9 | 250.51 | diab eye manif type i |
|  | DX_ICD9 | 250.60 | diab neuro manif type ii |
|  | DX_ICD9 | 250.61 | diab neuro manif type i |
|  | DX_ICD9 | 250.70 | diab circulat dis typ ii |
|  | DX_ICD9 | 250.71 | diab circulat dis type i |
|  | DX_ICD9 | 250.80 | diab w manif nec type ii |
|  | DX_ICD9 | 250.81 | diab w manif nec type i |
|  | DX_ICD9 | 250.90 | diab w compl nos type ii |
|  | DX_ICD9 | 250.91 | diab w compl nos type i |
|  | DX_ICD9 | 251.1 | hyperinsulinism nec |
|  | DX_ICD9 | 349.89 | CNS disorder nec |
|  | DX_ICD9 | 353.5 | neuralgic amyotrophy |
|  | DX_ICD9 | 355.9 | mononeuritis nos |
|  | DX_ICD9 | 357.2 | neuropathy in diabetes |
|  | DX_ICD9 | 362.01 | diabetic retinopathy nos |
|  | DX_ICD9 | 362.07 | diabetic macular edema (begin 2005) |
|  | DX_ICD9 | 366.41 | diabetic cataract |
|  | DX_ICD9 | 443.81 | angiopathy in other dis |
|  | DX_ICD9 | 523.8 | periodontal disease nec |
|  | DX_ICD9 | 528.9 | oral soft tissue dis nec |
|  | DX_ICD9 | 536.3 | gastroparesis (begin 1994) |
|  | DX_ICD9 | 581.81 | nephrotic syn in oth dis |
|  | DX_ICD9 | 583.81 | nephritis nos in oth dis |
|  | DX_ICD9 | 709.8 | skin disorders nec |
|  | DX_ICD9 | 713.5 | arthropathy w nerve dis |
|  | DX_ICD9 | 716.80 | arthropathy nec-unspec |
|  | DX_ICD9 | 785.4 | gangrene |
|  | DX_ICD10 | E08.00 | DM underly w hyperosmol wo nonketo hypergly-hyperosmol coma (nkhhc) |
|  | DX_ICD10 | E08.01 | DM underly w hyperosmolarity with coma |
|  | DX_ICD10 | E08.10 | DM underly w ketoacidosis without coma |
|  | DX_ICD10 | E08.11 | DM underly w ketoacidosis with coma |
|  | DX_ICD10 | E08.21 | DM underly w diabetic nephropathy |
|  | DX_ICD10 | E08.22 | DM underly w diabetic chronic kidney disease |
|  | DX_ICD10 | E08.29 | DM underly w oth diabetic kidney complic |
|  | DX_ICD10 | E08.311 | DM underly w unspec diabetic retinopathy w macular edema |
|  | DX_ICD10 | E08.319 | DM underly w unspec diabetic retinopathy wo macular edema |
|  | DX_ICD10 | E08.3211 | DM underly w mild nonprolif diabet retinopathy w macular edema, right eye |
|  | DX_ICD10 | E08.3212 | DM underly w mild nonprolif diabet retinopathy w macul edema, left eye |
|  | DX_ICD10 | E08.3213 | DM underly w mild nonprolif diabet retinopathy w macul edema, bilateral |
|  | DX_ICD10 | E08.3219 | DM underly w mild nonprolif diabet retinopath w macul edema, unspec eye |
|  | DX_ICD10 | E08.3291 | DM underly w mild nonprolif diabet retinopathy wo macul edema, right eye |
|  | DX_ICD10 | E08.3292 | DM underly w mild nonprolif diabet retinopathy wo macul edema, left eye |
|  | DX_ICD10 | E08.3293 | DM underly w mild nonprolif diabet retinopathy wo macul edema, bilateral |
|  | DX_ICD10 | E08.3299 | DM underly w mild nonprolif diabet retinopat wo macul edema, unspec eye |
|  | DX_ICD10 | E08.3311 | DM underly w moder nonprolif diabet retinopa w macul edema, right eye |
|  | DX_ICD10 | E08.3312 | DM underly w moder nonprolif diabetic retinopa w macul edema, left eye |
|  | DX_ICD10 | E08.3313 | DM underly w moder nonprolif diabetic retinopa w macul edema, bilat |
|  | DX_ICD10 | E08.3319 | DM underly w moder nonprolif diabet retinopa w macul edema, unspec eye |
|  | DX_ICD10 | E08.3391 | DM underly w moder nonprolif diabet retinopat wo macul edema, right eye |
|  | DX_ICD10 | E08.3392 | DM underly w moder nonprolif diabet retinopa wo macul edema, left eye |
|  | DX_ICD10 | E08.3393 | DM underly w moder nonprolif diabet retinopat wo macul edema, bilateral |
|  | DX_ICD10 | E08.3399 | DM underly w moder nonprolif diabe retinpat wo macul edema, unspec eye |
|  | DX_ICD10 | E08.3411 | DM underly w sev nonprolif diabet retinopat w macular edema, right eye |
|  | DX_ICD10 | E08.3412 | DM underly w sev nonprolif diabet retinopat w macular edema, left eye |
|  | DX_ICD10 | E08.3413 | DM underly w sev nonprolif diabetic retinopathy w macul edema, bilat |
|  | DX_ICD10 | E08.3419 | DM underly w sev nonprolif diabetic retinopat w macul edema, unspec eye |
|  | DX_ICD10 | E08.3491 | DM underly w sev nonprolif diabetic retinopat wo macular edema, right eye |
|  | DX_ICD10 | E08.3492 | DM underly w sev nonprolif diabetic retinopat wo macular edema, left eye |
|  | DX_ICD10 | E08.3493 | DM underly w sev nonprolif diabetic retinopat wo macular edema, bilateral |
|  | DX_ICD10 | E08.3499 | DM underly w sev nonprolif diabet retinopat wo macul edema, unspec eye |
|  | DX_ICD10 | E08.3511 | DM underly w prolif diabetic retinopathy w macular edema, right eye |
|  | DX_ICD10 | E08.3512 | DM underly w prolif diabetic retinopathy w macular edema, left eye |
|  | DX_ICD10 | E08.3513 | DM underly w prolif diabetic retinopathy w macular edema, bilateral |
|  | DX_ICD10 | E08.3519 | DM underly w prolif diabetic retinopath w macular edema, unspec eye |
|  | DX_ICD10 | E08.3521 | DM underly w prolif diabet retinopath w tract retinal detach macul, right eye |
|  | DX_ICD10 | E082 | DM underly w prolif diabet retinopath w tract retinal detach macula, left eye |
|  | DX_ICD10 | E08.3523 | DM underly w prolif diabet retinopath w tract retinal detach macul, bilat |
|  | DX_ICD10 | E08.3529 | DM underly w prolif diabet retinopath w tract retin detach macul, unspec eye |
|  | DX_ICD10 | E08.3531 | DM underly w prolif diabet retinopath w tract retina detach macul, right eye |
|  | DX_ICD10 | E08.3532 | DM underly w prolif diabet retinopathy w tract retina detach macul, left eye |
|  | DX_ICD10 | E08.3533 | DM underly w prolif diabet retinopath w tract retina detach macul, bilateral |
|  | DX_ICD10 | E08.3539 | DM underly w prolif diabet retinpath w tract retin detach macul, unspec eye |
|  | DX_ICD10 | E08.3541 | DM w prolif diab retinpath w tract retin & rhegmatog retin detach, right eye |
|  | DX_ICD10 | E08.3542 | DM w prolif diabet retinopa w tract retin & rhegmatog retin detach, left eye |
|  | DX_ICD10 | E08.3543 | DM w prolif diab retinopa w tract retal & rhegmatog retin detach, bilateral |
|  | DX_ICD10 | E08.3549 | DM w prolif diab retinpat w tract retin & rhegmatog retin detach, unspec eye |
|  | DX_ICD10 | E08.3551 | DM underly w stable prolif diabetic retinopathy, right eye |
|  | DX_ICD10 | E08.3552 | DM underly w stable prolif diabetic retinopathy, left eye |
|  | DX_ICD10 | E08.3553 | DM underly w stable prolif diabetic retinopathy, bilateral |
|  | DX_ICD10 | E08.3559 | DM underly w stable prolif diabetic retinopathy, unspecified eye |
|  | DX_ICD10 | E08.3591 | DM underly w prolif diabetic retinopathy wo macular edema, right eye |
|  | DX_ICD10 | E08.3592 | DM underly w prolif diabetic retinopathy wo macular edema, left eye |
|  | DX_ICD10 | E08.3593 | DM underly w prolif diabetic retinopathy wo macular edema, bilateral |
|  | DX_ICD10 | E08.3599 | DM underly w prolif diabetic retinopath wo macul edema, unspec eye |
|  | DX_ICD10 | E08.36 | DM underly w diabetic cataract |
|  | DX_ICD10 | E08.37X1 | DM underly w diabetic macul edema, after tx, right eye |
|  | DX_ICD10 | E08.37X2 | DM underly w diabetic macul edema, after tx, left eye |
|  | DX_ICD10 | E08.37X3 | DM underly w diabetic macul edema, after tx, bilateral |
|  | DX_ICD10 | E08.37X9 | DM underly w diabetic macul edema, after tx, unspec eye |
|  | DX_ICD10 | E08.39 | DM underly w other diabetic ophthalmic complication |
|  | DX_ICD10 | E08.40 | DM underly w diabetic neuropathy, unspecified |
|  | DX_ICD10 | E08.41 | DM underly w diabetic mononeuropathy |
|  | DX_ICD10 | E08.42 | DM underly w diabetic polyneuropathy |
|  | DX_ICD10 | E08.43 | DM underly w diabetic autonomic (poly)neuropathy |
|  | DX_ICD10 | E08.44 | DM underly w diabetic amyotrophy |
|  | DX_ICD10 | E08.49 | DM underly w other diabetic neurological complication |
|  | DX_ICD10 | E08.51 | DM underly w diabet peripheral angiopathy wo gangrene |
|  | DX_ICD10 | E08.52 | DM underly w diabetic peripheral angiopathy with gangrene |
|  | DX_ICD10 | E08.59 | DM underly w other circulatory complications |
|  | DX_ICD10 | E08.610 | DM underly w diabetic neuropathic arthropathy |
|  | DX_ICD10 | E08.618 | DM underly w other diabetic arthropathy |
|  | DX_ICD10 | E08.620 | DM underly w diabetic dermatitis |
|  | DX_ICD10 | E08.621 | DM underly w foot ulcer |
|  | DX_ICD10 | E08.622 | DM underly w other skin ulcer |
|  | DX_ICD10 | E08.628 | DM underly w other skin complications |
|  | DX_ICD10 | E08.630 | DM underly w periodontal disease |
|  | DX_ICD10 | E08.638 | DM underly w other oral complications |
|  | DX_ICD10 | E08.641 | DM underly w hypoglycemia with coma |
|  | DX_ICD10 | E08.649 | DM underly w hypoglycemia without coma |
|  | DX_ICD10 | E08.65 | DM underly w hyperglycemia |
|  | DX_ICD10 | E08.69 | DM underly w other spec complication |
|  | DX_ICD10 | E08.8 | DM underly w unspec complications |
|  | DX_ICD10 | E08.9 | DM underly wo complications |
|  | DX_ICD10 | E09.00 | drug/chem DM w hyperosm wo nonketo hypergly-hypersm coma (nkhhc) |
|  | DX_ICD10 | E09.01 | drug/chem DM w hyperosmolarity w coma |
|  | DX_ICD10 | E09.10 | drug/chem DM w ketoacidosis wo coma |
|  | DX_ICD10 | E09.11 | drug/chem induced DM w ketoacidosis with coma |
|  | DX_ICD10 | E09.21 | drug/chem induced DM w diabetic nephropathy |
|  | DX_ICD10 | E09.22 | drug/chem induced DM w diabetic chronic kidney disease |
|  | DX_ICD10 | E09.29 | Drug/chemical DM w oth diabetic kidney complication |
|  | DX_ICD10 | E09.311 | drug/chem DM w unspec diabetic retinopat w macul edema |
|  | DX_ICD10 | E09.319 | drug/chem DM w unspec diabetic retinopath wo macul edema |
|  | DX_ICD10 | E09.3211 | drug/chem DM w mild nonprolif diabet retinopat w macul edema, right eye |
|  | DX_ICD10 | E09.3212 | drug/chem DM w mild nonprolif diabet retinopathy w macul edema, left eye |
|  | DX_ICD10 | E09.3213 | drug/chem DM w mild nonprolif diabetic retinopat w macul edema, bilat |
|  | DX_ICD10 | E09.3219 | drug/chem DM w mild nonprolif diab retinopa w macul edema, unspec eye |
|  | DX_ICD10 | E09.3291 | drug/chem DM w mild nonprolif diabetic retinop wo macul edema, right eye |
|  | DX_ICD10 | E09.3292 | drug/chem DM w mild nonprolif diabet retinopat wo macul edema, left eye |
|  | DX_ICD10 | E09.3293 | drug/chem DM w mild nonprolif diabet retinopat wo macul edema, bilateral |
|  | DX_ICD10 | E09.3299 | drug/chem DM w mild nonprolif diab retinpat wo macul edema, unspec eye |
|  | DX_ICD10 | E09.3311 | drug/chem DM w moder nonprolif diabet retinpat w macul edema, right eye |
|  | DX_ICD10 | E09.3312 | drug/chem DM w moder nonprolif diabet retinopat w macul edema, left eye |
|  | DX_ICD10 | E09.3313 | drug/chem DM w moder nonprolif diabet retinopat w macul edema, bilat |
|  | DX_ICD10 | E09.3319 | drug/chem DM w moder nonprolif diab retipat w macul edema, unspec eye |
|  | DX_ICD10 | E09.3391 | drug/chem DM w moder nonprolif diab retinpat wo macur edema, right eye |
|  | DX_ICD10 | E09.3392 | drug/chem DM w moder nonprolif diab retinpat wo macul edema, left eye |
|  | DX_ICD10 | E09.3393 | drug/chem DM w moder nonprolif diabet retinopat wo macul edema, bilat |
|  | DX_ICD10 | E09.3399 | drug/chem DM w moder nonprolif diab retinpa wo macul edem, unspec eye |
|  | DX_ICD10 | E09.3411 | drug/chem DM w sev nonprolif diab retinopat w macul edema, right eye |
|  | DX_ICD10 | E09.3412 | drug/chem DM w sev nonprolif diab retinopathy w macul edema, left eye |
|  | DX_ICD10 | E09.3413 | drug/chem DM w sev nonprolif diab retinopat w macul edema, bilateral |
|  | DX_ICD10 | E09.3419 | drug/chem DM w sev nonprolif diab retinopat w macul edema, unspec eye |
|  | DX_ICD10 | E09.3491 | drug/chem DM w sev nonprolif diab retinopathy wo macul edema, right eye |
|  | DX_ICD10 | E09.3492 | drug/chem DM w sev nonprolif diab retinopat wo macular edema, left eye |
|  | DX_ICD10 | E09.3493 | drug/chem DM w sev nonprolif diabetic retinopat wo macul edema, bilat |
|  | DX_ICD10 | E09.3499 | drug/chem DM w sev nonprolif diab retinopa wo macul edema, unspec eye |
|  | DX_ICD10 | E09.3511 | drug/chem DM w prolif diab retinopat w macul edema, right eye |
|  | DX_ICD10 | E09.3512 | drug/chem DM w prolif diabetic retinopathy w macul edema, left eye |
|  | DX_ICD10 | E09.3513 | drug/chem DM w prolif diabetic retinopathy w macul edema, bilateral |
|  | DX_ICD10 | E09.3519 | drug/chem DM w prolif diabetic retinopathy w macul edema, unspec eye |
|  | DX_ICD10 | E09.3521 | drug/chem DM w prolif diab retinopa w tract retinal detach macul, right eye |
|  | DX_ICD10 | E09.3522 | drug/chem DM w prolif diab retinopat w tract retinal detach macul, left eye |
|  | DX_ICD10 | E09.3523 | drug/chem DM w prolif diab retinopat w tract retin detach macul, bilat |
|  | DX_ICD10 | E09.3529 | drug/chem DM w prolif diab retinpat w tract retin detach macul, unspec eye |
|  | DX_ICD10 | E09.3531 | drug/chem DM w prolif diab retpat w tract retin detach not macul, right eye |
|  | DX_ICD10 | E09.3532 | drug/chem DM w prolif diab retipa w tract retinal detach not macul, left eye |
|  | DX_ICD10 | E09.3533 | drug/chem DM w prolif diab retinpa w tract retinal detach not macul, bilat |
|  | DX_ICD10 | E09.3539 | drug/chem DM w prolif diab retipa w trac retin detac not macul, unspec eye |
|  | DX_ICD10 | E09.3541 | drug/chem DM w prolif diab retipat w tract reti & rhegmat detach, rt eye |
|  | DX_ICD10 | E09.3542 | drug/chem DM w prolif diab retinpa w tract reti & rhegmat detach, left eye |
|  | DX_ICD10 | E09.3543 | drug/chem DM w prolif diab retinpa w tract retin & rhegmat detach, bilat |
|  | DX_ICD10 | E09.3549 | drug/chem DM w prolif diab retinpa w tract retin & rhegmat detach, unspec |
|  | DX_ICD10 | E09.3551 | drug/chem DM w stable prolif diabetic retinopathy, right eye |
|  | DX_ICD10 | E09.3552 | drug/chem DM w stable prolif diabetic retinopathy, left eye |
|  | DX_ICD10 | E09.3553 | drug/chem DM w stable prolif diabetic retinopathy, bilateral |
|  | DX_ICD10 | E09.3559 | drug/chem DM w stable prolif diabetic retinopathy, unspec eye |
|  | DX_ICD10 | E09.3591 | drug/chem DM w prolif diabetic retinopathy wo macular edema, right eye |
|  | DX_ICD10 | E09.3592 | drug/chem DM w prolif diabetic retinopathy wo macular edema, left eye |
|  | DX_ICD10 | E09.3593 | drug/chem DM w prolif diabetic retinopathy wo macular edema, bilateral |
|  | DX_ICD10 | E09.3599 | drug/chem DM w prolif diabetic retinopathy wo macular edema, unspec eye |
|  | DX_ICD10 | E09.36 | drug/chem DM w diabetic cataract |
|  | DX_ICD10 | E09.37X1 | drug/chem DM w diab macul edema, resolved after tx, right eye |
|  | DX_ICD10 | E09.37X2 | drug/chem DM w diabetic macular edema, resolved after tx, left eye |
|  | DX_ICD10 | E09.37X3 | drug/chem DM w diabetic macular edema, resolved after tx, bilateral |
|  | DX_ICD10 | E09.37X9 | drug/chem DM w diabetic macular edema, resolved after tx, unspec eye |
|  | DX_ICD10 | E09.39 | drug/chem DM w oth diabetic ophthalmic complic |
|  | DX_ICD10 | E09.40 | drug/chem DM w neurological complic with diabetic neuropathy, unspec |
|  | DX_ICD10 | E09.41 | drug/chem DM w neurol complic w diabetic mononeuropathy |
|  | DX_ICD10 | E09.42 | drug/chem DM w neurol complic w diabetic polyneuropathy |
|  | DX_ICD10 | E09.43 | drug/chem DM w neurol complic w diabetic autonomic (poly) neuropathy |
|  | DX_ICD10 | E09.44 | drug/chem DM w neurol complic w diabetic amyotrophy |
|  | DX_ICD10 | E09.49 | drug/chem DM w neurol complic w oth diabetic neurological complic |
|  | DX_ICD10 | E09.51 | drug/chem DM w diabetic peripheral angiopathy wo gangrene |
|  | DX_ICD10 | E09.52 | drug/chem DM w diabetic peripheral angiopathy w gangrene |
|  | DX_ICD10 | E09.59 | drug/chem DM w oth circulatory complic |
|  | DX_ICD10 | E09.610 | drug/chem DM w diabetic neuropathic arthropathy |
|  | DX_ICD10 | E09.618 | drug/chem DM w other diabetic arthropathy |
|  | DX_ICD10 | E09.620 | drug/chem DM w diabetic dermatitis |
|  | DX_ICD10 | E09.621 | drug/chem DM w foot ulcer |
|  | DX_ICD10 | E09.622 | drug/chem DM w oth skin ulcer |
|  | DX_ICD10 | E09.628 | drug/chem DM w oth skin complic |
|  | DX_ICD10 | E09.630 | drug/chem DM w periodontal disease |
|  | DX_ICD10 | E09.638 | drug/chem DM w other oral complic |
|  | DX_ICD10 | E09.641 | drug/chem DM w hypoglycemia w coma |
|  | DX_ICD10 | E09.649 | drug/chem DM w hypoglycemia wo coma |
|  | DX_ICD10 | E09.65 | drug/chem DM w hyperglycemia |
|  | DX_ICD10 | E09.69 | drug/chem DM w other specified complic |
|  | DX_ICD10 | E09.8 | drug/chem DM w unspecified complic |
|  | DX_ICD10 | E09.9 | drug/chem induced DM wo complications |
|  | DX_ICD10 | E10.10 | type 1 DM w ketoacidosis wo coma |
|  | DX_ICD10 | E10.11 | type 1 DM w ketoacidosis w coma |
|  | DX_ICD10 | E10.21 | type 1 DM w diabetic nephropathy |
|  | DX_ICD10 | E10.22 | type 1 DM w diabetic chronic kidney disease |
|  | DX_ICD10 | E10.29 | type 1 DM w other diabetic kidney complic |
|  | DX_ICD10 | E10.311 | type 1 DM w unspecified diab retinopathy w macul edema |
|  | DX_ICD10 | E10.319 | type 1 DM w unspecified diabetic retinopat wo macul edema |
|  | DX_ICD10 | E10.3211 | type 1 DM w mild nonprolif diab retinopathy w macul edema, right eye |
|  | DX_ICD10 | E10.3212 | type 1 DM w mild nonprolif diab retinopathy w macul edema, left eye |
|  | DX_ICD10 | E10.3213 | type 1 DM w mild nonprolif diabetic retinopathy w macul edema, bilateral |
|  | DX_ICD10 | E10.3219 | type 1 DM w mild nonprolif diabetic retinopathy w macul edema, unspeceye |
|  | DX_ICD10 | E10.3291 | type 1 DM w mild nonprolif diab retinopathy wo macul edema, right eye |
|  | DX_ICD10 | E10.3292 | type 1 DM w mild nonprolif diab retinopathy wo macul edema, left eye |
|  | DX_ICD10 | E10.3293 | type 1 DM w mild nonprolif diab retinopathy wo macul edema, bilateral |
|  | DX_ICD10 | E10.3299 | type 1 DM w mild nonprolif diab retinopathy wo macul edema, unspec eye |
|  | DX_ICD10 | E10.3311 | type 1 DM w moder nonprolif diab retinopathy w macul edema, right eye |
|  | DX_ICD10 | E10.3312 | type 1 DM w moder nonprolif diab retinopathy w macul edema, left eye |
|  | DX_ICD10 | E10.3313 | type 1 DM w moderate nonprolif diab retinopathy w macul edema, bilat |
|  | DX_ICD10 | E10.3319 | type 1 DM w moderate nonprolif diab retinopa w macul edema, unspec eye |
|  | DX_ICD10 | E10.3391 | type 1 DM w moder nonprolif diab retinopathy wo macul edema, right eye |
|  | DX_ICD10 | E10.3392 | type 1 DM w moder nonprolif diab retinopathy wo macul edema, left eye |
|  | DX_ICD10 | E10.3393 | type 1 DM with moder nonprolif diab retinopathy wo macul edema, bilat |
|  | DX_ICD10 | E10.3399 | type 1 DM with moder nonprolif diab retinopat wo macul edema, unspec eye |
|  | DX_ICD10 | E10.3411 | type 1 DM w severe nonprolif diabetic retinopa w macular edema, right eye |
|  | DX_ICD10 | E10.3412 | type 1 DM w sev nonprolif diab retinopat w macul edema, left eye |
|  | DX_ICD10 | E10.3413 | type 1 DM w sev nonprolif diab retinopat w macul edema, bilateral |
|  | DX_ICD10 | E10.3419 | type 1 DM w sev nonprolif diab retinopat w macul edema, unspec eye |
|  | DX_ICD10 | E10.3491 | type 1 DM w sev nonprolif diab retinopat wo macul edema, right eye |
|  | DX_ICD10 | E10.3492 | type 1 DM w sev nonprolif diab retinopat wo macul edema, left eye |
|  | DX_ICD10 | E10.3493 | type 1 DM w sev nonprolif diab retinopat wo macul edema, bilateral |
|  | DX_ICD10 | E10.3499 | type 1 DM w sev nonprolif diab retinopat wo macul edema, unspec eye |
|  | DX_ICD10 | E10.3511 | type 1 DM w prolif diabetic retinopathy w macul edema, right eye |
|  | DX_ICD10 | E10.3512 | type 1 DM w prolif diabetic retinopathy w macul edema, left eye |
|  | DX_ICD10 | E10.3513 | type 1 DM w prolif diabetic retinopathy w macul edema, bilateral |
|  | DX_ICD10 | E10.3519 | type 1 DM w prolif diabetic retinopathy w macular edema, unspecified eye |
|  | DX_ICD10 | E10.36 | type 1 DM w diabetic cataract |
|  | DX_ICD10 | E10.37X1 | type 1 DM w diabetic macular edema, resolved after tx, right eye |
|  | DX_ICD10 | E10.37X2 | type 1 DM w diabetic macular edema, resolved after, left eye |
|  | DX_ICD10 | E10.37X3 | type 1 DM w diabetic macular edema, resolved after, bilateral |
|  | DX_ICD10 | E10.37X9 | type 1 DM w diabetic macular edema, resolved after, unspec eye |
|  | DX_ICD10 | E10.39 | type 1 DM w other diabetic ophthalmic complication |
|  | DX_ICD10 | E10.40 | type 1 DM w diabetic neuropathy, unspecified |
|  | DX_ICD10 | E10.41 | type 1 DM w diabetic mononeuropathy |
|  | DX_ICD10 | E10.42 | type 1 DM w diabetic polyneuropathy |
|  | DX_ICD10 | E10.43 | type 1 DM w diabetic autonomic (poly)neuropathy |
|  | DX_ICD10 | E10.44 | type 1 DM w diabetic amyotrophy |
|  | DX_ICD10 | E10.49 | type 1 DM w other diabetic neurological complication |
|  | DX_ICD10 | E10.51 | type 1 DM w diabetic peripheral angiopathy without gangrene |
|  | DX_ICD10 | E10.52 | type 1 DM w diabetic peripheral angiopathy with gangrene |
|  | DX_ICD10 | E10.59 | type 1 DM w other circulatory complications |
|  | DX_ICD10 | E10.610 | type 1 DM w diabetic neuropathic arthropathy |
|  | DX_ICD10 | E10.618 | type 1 DM w other diabetic arthropathy |
|  | DX_ICD10 | E10.620 | type 1 DM w diabetic dermatitis |
|  | DX_ICD10 | E10.621 | type 1 DM w foot ulcer |
|  | DX_ICD10 | E10.622 | type 1 DM w other skin ulcer |
|  | DX_ICD10 | E10.628 | type 1 DM w other skin complications |
|  | DX_ICD10 | E10.630 | type 1 DM w periodontal disease |
|  | DX_ICD10 | E10.638 | type 1 DM w other oral complications |
|  | DX_ICD10 | E10.641 | type 1 DM w hypoglycemia with coma |
|  | DX_ICD10 | E10.649 | type 1 DM w hypoglycemia without coma |
|  | DX_ICD10 | E10.65 | type 1 DM w hyperglycemia |
|  | DX_ICD10 | E10.69 | type 1 DM w other specified complication |
|  | DX_ICD10 | E10.8 | type 1 DM w unspecified complications |
|  | DX_ICD10 | E10.9 | type 1 DM wo complications |
|  | DX_ICD10 | E11.00 | type 2 DM w hyperosmolar wo nonketo hypergly-hyperosmol coma (nkhhc) |
|  | DX_ICD10 | E11.01 | type 2 DM w hyperosmolarity with coma |
|  | DX_ICD10 | E11.10 | type 2 DM w ketoacidosis without coma |
|  | DX_ICD10 | E11.11 | type 2 DM w ketoacidosis with coma |
|  | DX_ICD10 | E11.21 | type 2 DM w diab nephropathy |
|  | DX_ICD10 | E11.22 | type 2 DM w diab chronic kidney dis |
|  | DX_ICD10 | E11.29 | type 2 DM w other diab kidney complic |
|  | DX_ICD10 | E11.311 | type 2 DM w unspec diab retinopathy w macul edema |
|  | DX_ICD10 | E11.319 | type 2 DM w unspec diab retinopathy wo macul edema |
|  | DX_ICD10 | E11.3211 | type 2 DM w mild nonprolif diab retinopathy w macul edema, right eye |
|  | DX_ICD10 | E11.3212 | type 2 DM w mild nonprolif diab retinopathy w macul edema, left eye |
|  | DX_ICD10 | E11.3213 | type 2 DM w mild nonprolif diab retinopathy w macul edema, bilateral |
|  | DX_ICD10 | E11.3219 | type 2 DM w mild nonprolif diab retinopathy w macul edema, unspec eye |
|  | DX_ICD10 | E11.3291 | type 2 DM w mild nonprolif diab retinopathy wo macul edema, right eye |
|  | DX_ICD10 | E11.3292 | type 2 DM w mild nonprolif diab retinopathy wo macul edema, left eye |
|  | DX_ICD10 | E11.3293 | type 2 DM w mild nonprolif diab retinopathy wo macul edema, bilateral |
|  | DX_ICD10 | E11.3299 | type 2 DM w mild nonprolif diab retinopa wo mac macul edema, unspec eye |
|  | DX_ICD10 | E11.3311 | type 2 DM with mod nonprolif diab retinopat w macul edema, right eye |
|  | DX_ICD10 | E11.3312 | type 2 DM with mod nonprolif diab retinopat w macul edema, left eye |
|  | DX_ICD10 | E11.3313 | type 2 DM with mod nonprolif diab retinopat w macul edema, bilateral |
|  | DX_ICD10 | E11.3319 | type 2 DM with mod nonprolif diab retinopat w macul edema, unspec eye |
|  | DX_ICD10 | E11.3391 | type 2 DM w mod nonprolif diab retinopathy wo macular edema, right eye |
|  | DX_ICD10 | E11.3392 | type 2 DM w mod nonprolif diab retinopathy wo macul edema, left eye |
|  | DX_ICD10 | E11.3393 | type 2 DM w mod nonprolif diab retinopathy wo macul edema, bilateral |
|  | DX_ICD10 | E11.3399 | type 2 DM w mod nonprolif diab retinopathy wo macul edema, unspec eye |
|  | DX_ICD10 | E11.3411 | type 2 DM w sev nonprolif diab retinopathy w macul edema, right eye |
|  | DX_ICD10 | E11.3412 | type 2 DM w sev nonprolif diab retinopathy w macul edema, left eye |
|  | DX_ICD10 | E11.3413 | type 2 DM w sev nonprolif diab retinopathy w macul edema, bilateral |
|  | DX_ICD10 | E11.3419 | type 2 DM w sev nonprolif diab retinopathy w macul edema, unspec eye |
|  | DX_ICD10 | E11.3491 | type 2 DM w sev nonprolif diab retinopathy wo macul edema, right eye |
|  | DX_ICD10 | E11.3492 | type 2 DM w sev nonprolif diab retinopathy wo macul edema, left eye |
|  | DX_ICD10 | E11.3493 | type 2 DM w sev nonprolif diab retinopathy wo macul edema, bilateral |
|  | DX_ICD10 | E11.3499 | type 2 DM w sev nonprolif diab retinopathy wo macul edema, unspec eye |
|  | DX_ICD10 | E11.3511 | type 2 DM w prolif diab retinopathy w macul edema, right eye |
|  | DX_ICD10 | E11.3512 | type 2 DM w prolif diab retinopathy with macular edema, left eye |
|  | DX_ICD10 | E11.3513 | type 2 DM w prolif diab retinopathy with macular edema, bilat |
|  | DX_ICD10 | E11.3519 | type 2 DM w prolif diab retinopathy with macular edema, unspec eye |
|  | DX_ICD10 | E11.3521 | type 2 DM w prolif diab retinopathy w tract retinal detach macul, right eye |
|  | DX_ICD10 | E11.3522 | type 2 DM w prolif diab retinopathy w tract retinal detach macul, left eye |
|  | DX_ICD10 | E11.3523 | type 2 DM w prolif diab retinopathy w tract retinal detach macul, bilat |
|  | DX_ICD10 | E11.3529 | type 2 DM w prolif diab retinopathy w tract retinal detach macul, unspec eye |
|  | DX_ICD10 | E11.3531 | type 2 DM w prolif diab retinopa w tract retinal detach not macul, right eye |
|  | DX_ICD10 | E11.3532 | type 2 DM w prolif diab retinopathy w tract retinal detach not macul, left eye |
|  | DX_ICD10 | E11.3533 | type 2 DM w prolif diab retinopa w tract retinal detach not macul, bilateral |
|  | DX_ICD10 | E11.3539 | type 2 DM w prolif diab retinopa w tract retin detach not macul, unspec eye |
|  | DX_ICD10 | E11.3541 | type 2 DM w prolif diab retinpa w tract retin & rhegmat reti detach, right eye |
|  | DX_ICD10 | E11.3542 | type 2 DM w prolif diab retinpa w tract retin & rhegmat reti detach, left eye |
|  | DX_ICD10 | E11.3543 | type 2 DM w prolif diab retinpa w tract retin & rhegmat reti detach, bilat |
|  | DX_ICD10 | E11.3549 | type 2 DM w prolif diab retinpa w tract reti & rhegmat reti detach, unspec |
|  | DX_ICD10 | E11.3551 | type 2 DM w stable prolif diab retinopathy, right eye |
|  | DX_ICD10 | E11.3552 | type 2 DM w stable prolif diab retinopathy, left eye |
|  | DX_ICD10 | E11.3553 | type 2 DM w stable prolif diab retinopathy, bilateral |
|  | DX_ICD10 | E11.3559 | type 2 DM w stable prolif diab retinopathy, unspec eye |
|  | DX_ICD10 | E11.3591 | type 2 DM w prolif diab retinopathy wo macul edema, right eye |
|  | DX_ICD10 | E11.3592 | type 2 DM w prolif diab retinopathy wo macul edema, left eye |
|  | DX_ICD10 | E11.3593 | type 2 DM w prolif diab retinopathy wo macul edema, bilateral |
|  | DX_ICD10 | E11.3599 | type 2 DM w prolif diab retinopathy wo macul edema, unspec eye |
|  | DX_ICD10 | E11.36 | type 2 DM w diab cataract |
|  | DX_ICD10 | E11.37X1 | type 2 DM w diab macul edema, resolved after tx, right eye |
|  | DX_ICD10 | E11.37X2 | type 2 DM w diab macul edema, resolved after tx, left eye |
|  | DX_ICD10 | E11.37X3 | type 2 DM w diabetic macular edema, resolved after tx, bilateral |
|  | DX_ICD10 | E11.37X9 | type 2 DM w diabetic macular edema, resolved after tx, unspec eye |
|  | DX_ICD10 | E11.39 | type 2 DM w other diabetic ophthalmic complic |
|  | DX_ICD10 | E11.40 | type 2 DM w diabetic neuropathy, unspec |
|  | DX_ICD10 | E11.41 | type 2 DM w diabetic mononeuropathy |
|  | DX_ICD10 | E11.42 | type 2 DM w diabetic polyneuropathy |
|  | DX_ICD10 | E11.43 | type 2 DM w diabetic autonomic (poly) neuropathy |
|  | DX_ICD10 | E11.44 | type 2 DM w diabetic amyotrophy |
|  | DX_ICD10 | E11.49 | type 2 DM w other diabetic neurological complic |
|  | DX_ICD10 | E11.51 | type 2 DM w diabetic peripheral angiopathy wo gangrene |
|  | DX_ICD10 | E11.52 | type 2 DM w diabetic peripheral angiopathy w gangrene |
|  | DX_ICD10 | E11.59 | type 2 DM w other circulatory complic |
|  | DX_ICD10 | E11.610 | type 2 DM w diabetic neuropathic arthropathy |
|  | DX_ICD10 | E11.618 | type 2 DM w other diabetic arthropathy |
|  | DX_ICD10 | E11.620 | type 2 DM w diabetic dermatitis |
|  | DX_ICD10 | E11.621 | type 2 DM w foot ulcer |
|  | DX_ICD10 | E11.622 | type 2 DM w other skin ulcer |
|  | DX_ICD10 | E11.628 | type 2 DM w other skin complications |
|  | DX_ICD10 | E11.630 | type 2 DM w periodontal disease |
|  | DX_ICD10 | E11.638 | type 2 DM w other oral complications |
|  | DX_ICD10 | E11.641 | type 2 DM w hypoglycemia w coma |
|  | DX_ICD10 | E11.649 | type 2 DM w hypoglycemia wo coma |
|  | DX_ICD10 | E11.65 | type 2 DM w hyperglycemia |
|  | DX_ICD10 | E11.69 | type 2 DM w oth spec complication |
|  | DX_ICD10 | E11.8 | type 2 DM w unspecified complica |
|  | DX_ICD10 | E11.9 | type 2 DM wo complic |
|  | DX_ICD10 | E13.00 | oth spec DM w hyperosmol wo nonketo hypergly-hyperosm coma (nkhhc) |
|  | DX_ICD10 | E13.01 | oth spec DM w hyperosmolarity w coma |
|  | DX_ICD10 | E13.10 | oth spec DM w ketoacidosis wo coma |
|  | DX_ICD10 | E13.11 | oth spec DM w ketoacidosis w coma |
|  | DX_ICD10 | E13.21 | oth spec DM w diab nephropathy |
|  | DX_ICD10 | E13.22 | oth spec DM w diab chronic kidney disease |
|  | DX_ICD10 | E13.29 | oth spec DM w oth diabetic kidney complication |
|  | DX_ICD10 | E13.311 | oth spec DM w unspec diabetic retinopathy w macul edema |
|  | DX_ICD10 | E13.319 | oth spec DM w unspec diabetic retinopathy wo macul edema |
|  | DX_ICD10 | E13.3211 | oth spec DM w mild nonprolif diab retinopathy w macul edema, right eye |
|  | DX_ICD10 | E13.3212 | oth spec DM w mild nonprolif diab retinopathy w macul edema, left eye |
|  | DX_ICD10 | E13.3213 | oth spec DM w mild nonprolif diab retinopathy w macul edema, bilateral |
|  | DX_ICD10 | E13.3219 | oth spec DM w mild nonprolif diab retinopathy w macul edema, unspec eye |
|  | DX_ICD10 | E13.3291 | oth spec DM w mild nonprolif diab retinopathy wo macul edema, right eye |
|  | DX_ICD10 | E13.3292 | oth spec DM w mild nonprolif diab retinopathy wo macul edema, left eye |
|  | DX_ICD10 | E13.3293 | oth spec DM w mild nonprolif diab retinopathy wo macul edema, bilateral |
|  | DX_ICD10 | E13.3299 | oth spec DM w mild nonprolif diab retinopathy wo macul edema, unspec eye |
|  | DX_ICD10 | E13.3311 | oth spec DM w mod nonprolif diab retinopathy w macul edema, right eye |
|  | DX_ICD10 | E13.3312 | oth spec DM w mod nonprolif diab retinopathy w macul edema, left eye |
|  | DX_ICD10 | E13.3313 | oth spec DM w mod nonprolif diab retinopathy w macul edema, bilateral |
|  | DX_ICD10 | E13.3319 | oth spec DM w mod nonprolif diab retinopa w macul edema, unspec eye |
|  | DX_ICD10 | E13.3391 | oth spec DM w mod nonprolif diab retinopathy wo macul edema, right eye |
|  | DX_ICD10 | E13.3392 | Oth spec DM w mod nonprolif diab retinopathy wo macul edema, left eye |
|  | DX_ICD10 | E13.3393 | oth spec DM w mod nonprolif diab retinopathy wo macul edema, bilateral |
|  | DX_ICD10 | E13.3399 | oth spec DM w mod nonprolif diab retinopa wo macul edema, unspec eye |
|  | DX_ICD10 | E13.3411 | oth spec DM w sev nonprolif diab retinopathy w macul edema, right eye |
|  | DX_ICD10 | E13.3412 | oth spec DM w sev nonprolif diab retinopathy with macul edema, left eye |
|  | DX_ICD10 | E13.3413 | oth spec DM w sev nonprolif diab retinopathy w macul edema, bilateral |
|  | DX_ICD10 | E13.3419 | oth spec DM w sev nonprolif diab retinopathy w macular edema, unspec eye |
|  | DX_ICD10 | E13.3491 | oth spec DM w sev nonprolif diab retinopa wo macular edema, right eye |
|  | DX_ICD10 | E13.3492 | oth spec DM w sev nonprolif diabetic retinopa wo macular edema, left eye |
|  | DX_ICD10 | E13.3493 | oth spec DM w sev nonprolif diab retinopathy wo macular edema, bilateral |
|  | DX_ICD10 | E13.3499 | oth spec DM w sev nonprolif diab retinopa wo macular edema, unspec eye |
|  | DX_ICD10 | E13.3511 | oth spec DM w prolif diab retinopathy w macular edema, right eye |
|  | DX_ICD10 | E13.3512 | oth spec DM w prolif diab retinopathy w macular edema, left eye |
|  | DX_ICD10 | E13.3513 | oth spec DM w prolif diab retinopathy w macular edema, bilateral |
|  | DX_ICD10 | E13.3519 | oth spec DM w prolif diab retinopathy w macular edema, unspec eye |
|  | DX_ICD10 | E13.3521 | oth spec DM w prolif diab retinopa w tract retinal detach macula, right eye |
|  | DX_ICD10 | E13.3522 | oth spec DM w prolif diab retinopa w tract retinal detach macula, left eye |
|  | DX_ICD10 | E13.3523 | oth spec DM w prolif diab retinopa w tract retinal detach macula, bilateral |
|  | DX_ICD10 | E13.3529 | oth spec DM w prolif diab retinopa w tract retin detach macula, unspec eye |
|  | DX_ICD10 | E13.3531 | oth spec DM w prolif diab retinopa w tract retin detach not macula, right eye |
|  | DX_ICD10 | E13.3532 | oth spec DM w prolif diab retinopa w tract retinal detach not macula, left eye |
|  | DX_ICD10 | E13.3533 | oth spec DM w prolif diab retinpa w tract retin detach not macula, bilateral |
|  | DX_ICD10 | E13.3539 | oth spec DM w prolif diab retinpa w tract retin detach not mac, unspec eye |
|  | DX_ICD10 | E13.3541 | oth spec DM w prolif diab retinpa w tract retin & rhegmat detach, right eye |
|  | DX_ICD10 | E13.3542 | oth spec DM w prolif diab retinpa w tract retin & rhegmat detach, left eye |
|  | DX_ICD10 | E13.3543 | oth spec DM w prolif diab retinpa w tract retin & rhegmat detach, bilateral |
|  | DX_ICD10 | E13.3549 | oth spec DM w prolif diab retinpa w tract retin & rhegmat detach, unspec |
|  | DX_ICD10 | E13.3551 | oth spec DM w stable prolif diab retinopathy, right eye |
|  | DX_ICD10 | E13.3552 | oth spec DM with stable prolif diab retinopathy, left eye |
|  | DX_ICD10 | E13.3553 | oth spec DM with stable prolif diab retinopathy, bilateral |
|  | DX_ICD10 | E13.3559 | oth spec DM with stable prolif diab retinopathy, unspec eye |
|  | DX_ICD10 | E13.36 | oth spec DM with diab cataract |
|  | DX_ICD10 | E13.39 | oth spec DM with other diab ophthalmic complication |
|  | DX_ICD10 | E13.40 | oth spec DM with diab neuropathy, unspecified |
|  | DX_ICD10 | E13.41 | oth spec DM with diab mononeuropathy |
|  | DX_ICD10 | E13.42 | oth spec DM with diab polyneuropathy |
|  | DX_ICD10 | E13.43 | oth spec DM with diab autonomic (poly)neuropathy |
|  | DX_ICD10 | E13.44 | oth spec DM with diab amyotrophy |
|  | DX_ICD10 | E13.49 | oth spec DM with other diab neurological complication |
|  | DX_ICD10 | E13.51 | oth spec DM with diab peripheral angiopathy without gangrene |
|  | DX_ICD10 | E13.52 | oth spec DM with diab peripheral angiopathy with gangrene |
|  | DX_ICD10 | E13.59 | oth spec DM with other circulatory complications |
|  | DX_ICD10 | E13.610 | oth spec DM with diab neuropathic arthropathy |
|  | DX_ICD10 | E13.618 | oth spec DM with other diab arthropathy |
|  | DX_ICD10 | E13.620 | oth spec DM with diab dermatitis |
|  | DX_ICD10 | E13.621 | oth spec DM with foot ulcer |
|  | DX_ICD10 | E13.622 | oth spec DM with other skin ulcer |
|  | DX_ICD10 | E13.628 | oth spec DM with other skin complications |
|  | DX_ICD10 | E13.630 | oth spec DM with periodontal disease |
|  | DX_ICD10 | E13.638 | oth spec DM with other oral complications |
|  | DX_ICD10 | E13.641 | oth spec DM with hypoglycemia with coma |
|  | DX_ICD10 | E13.649 | oth spec DM with hypoglycemia without coma |
|  | DX_ICD10 | E13.65 | oth spec DM with hyperglycemia |
|  | DX_ICD10 | E13.69 | oth spec DM with oth spec complication |
|  | DX_ICD10 | E13.8 | oth spec DM with unspecified complications |
|  | DX_ICD10 | E13.9 | oth spec DM without complications |
| **Heart Failure** 17 ICD-9 DX 71 ICD-10 DX **80 Total** | DX_ICD9 | 42820 | unspecified systolic heart failure (begin 2002) |
|  | DX_ICD9 | 42821 | acute systolic heart failure (begin 2002) |
|  | DX_ICD9 | 42822 | chronic systolic heart failure (begin 2002) |
|  | DX_ICD9 | 42823 | acute on chronic systolic heart failr (begin 2002) |
|  | DX_ICD9 | 42840 | unspec cmbined syst & dias heart failr (begin 2002) |
|  | DX_ICD9 | 42841 | acute cmbined syst & dias heart failr (begin 2002) |
|  | DX_ICD9 | 42842 | chron cmbined syst & dias heart failr (begin 2002) |
|  | DX_ICD9 | 42843 | acu chro combi syst & dias hrt failr (begin 2002) |
|  | DX_ICD9 | 42830 | unspecified diastolic heart failure (begin 2002) |
|  | DX_ICD9 | 42831 | acute diastolic heart failure (begin 2002) |
|  | DX_ICD9 | 42832 | chronic diastolic heart failure (begin 2002) |
|  | DX_ICD9 | 42833 | acute on chronic diastolic heart failr (begin 2002) |
|  | DX_ICD9 | 785.51 | cardiogenic shock |
|  | DX_ICD9 | 996.83 | compl heart transplant (begin 1987) |
|  | DX_ICD9 | V15.87 | history of extracorporeal membrane oxygenatio (begin 2003) |
|  | DX_ICD9 | V42.1 | hrt transplant status |
|  | DX_ICD9 | V43.2 | hrt replacement nec (end 2003) |
|  | DX_ICD10 | A54.83 | gonococcal heart infection |
|  | DX_ICD10 | I09.81 | rheumatic heart failure |
|  | DX_ICD10 | I25.750 | atheroscler native coro art transplant hrt w unstable angina |
|  | DX_ICD10 | I25.751 | atheroscler native coron art transplant hrt w angina pect w doc spasm |
|  | DX_ICD10 | I25.758 | atheroscler native coron art transplanted hrt w oth forms angina pect |
|  | DX_ICD10 | I25.759 | atheroscler native coron art transplanted hrt w unspecified angina pectoris |
|  | DX_ICD10 | I25.760 | atherosclerosis of bypass graft coron art transplant hrt w unstable angina |
|  | DX_ICD10 | I25.761 | atheroscler bypass graft coron art transplant hrt w angina pect w doc spasm |
|  | DX_ICD10 | I25.768 | atheroscler bypass graft coron art transplant hrt w oth forms of angina pect |
|  | DX_ICD10 | I25.769 | atheroscler bypass graft coron art transplant hrt w unspecified angina pect |
|  | DX_ICD10 | I25.812 | atheroscler bypass graft coron artery of transplant hrt wo angina pectoris |
|  | DX_ICD10 | I42.7 | cardiomyopathy due to drug and external agent |
|  | DX_ICD10 | I50.1 | left ventricular failure, unspecified |
|  | DX_ICD10 | I50.20 | unspecified systolic (congestive) hrt failure |
|  | DX_ICD10 | I50.21 | acute systolic (congestive) hrt failure |
|  | DX_ICD10 | I50.22 | chronic systolic (congestive) hrt failure |
|  | DX_ICD10 | I50.23 | acute on chronic systolic (congestive) hrt failure |
|  | DX_ICD10 | I50.30 | unspecified diastolic (congestive) hrt failure |
|  | DX_ICD10 | I50.31 | acute diastolic (congestive) hrt failure |
|  | DX_ICD10 | I50.32 | chronic diastolic (congestive) hrt failure |
|  | DX_ICD10 | I50.33 | acute on chronic diastolic (congestive) hrt failure |
|  | DX_ICD10 | I50.40 | unspec systolic (congestive) & diastolic (congestive) hrt failure |
|  | DX_ICD10 | I50.41 | acute systolic (congestive) & diastolic (congestive) hrt failure |
|  | DX_ICD10 | I50.42 | chronic systolic (congestive) & diastolic (congestive) hrt failure |
|  | DX_ICD10 | I50.43 | acute on chronic systolic (congestive) & diastolic (congestive) hrt failure |
|  | DX_ICD10 | I50.810 | right hrt failure, unspec |
|  | DX_ICD10 | I50.811 | acute right hrt failure |
|  | DX_ICD10 | I50.812 | chronic right hrt failure |
|  | DX_ICD10 | I50.813 | acute on chronic right hrt failure |
|  | DX_ICD10 | I50.814 | right hrt failure due to left hrt failure |
|  | DX_ICD10 | I50.82 | biventricular hrt failure |
|  | DX_ICD10 | I50.83 | high output hrt failure |
|  | DX_ICD10 | I50.84 | end stage hrt failure |
|  | DX_ICD10 | I50.89 | other hrt failure |
|  | DX_ICD10 | I50.9 | hrt failure, unspecified |
|  | DX_ICD10 | I97.130 | postprocedural hrt failure after cardiac surgery |
|  | DX_ICD10 | I97.131 | postprocedural hrt failure after other surgery |
|  | DX_ICD10 | R57.0 | cardiogenic shock |
|  | DX_ICD10 | T82.49XA | other complication of vascular dialysis catheter, initial encounter |
|  | DX_ICD10 | T82.512S | breakdown (mechanical) of artificial heart, sequela |
|  | DX_ICD10 | T82.522S | displacement of artificial heart, sequela |
|  | DX_ICD10 | T82.532A | leakage of artificial heart, initial encounter |
|  | DX_ICD10 | T82.532S | leakage of artificial heart, sequela |
|  | DX_ICD10 | T82.592A | other mechanical complication of artificial heart, initial encounter |
|  | DX_ICD10 | T82.857D | stenosis oth cardiac prosthetic devices, implants & grafts, subseq encounter |
|  | DX_ICD10 | T86.20 | unspecified complic hrt transplant |
|  | DX_ICD10 | T86.21 | hrt transplant rejection |
|  | DX_ICD10 | T86.22 | hrt transplant failure |
|  | DX_ICD10 | T86.23 | hrt transplant infection |
|  | DX_ICD10 | T86.290 | cardiac allograft vasculopathy |
|  | DX_ICD10 | T86.298 | other complications of hrt transplant |
|  | DX_ICD10 | T86.30 | unspec complic of heart-lung transplant |
|  | DX_ICD10 | T86.31 | hrt-lung transplant rejection |
|  | DX_ICD10 | T86.32 | hrt-lung transplant failure |
|  | DX_ICD10 | T86.33 | hrt-lung transplant infection |
|  | DX_ICD10 | T86.39 | oth complic heart-lung transplant |
|  | DX_ICD10 | Z48.21 | encounter for aftercare after hrt transplant |
|  | DX_ICD10 | Z48.280 | encounter for aftercare after hrt-lung transplant |
|  | DX_ICD10 | Z92.81 | personal history of extracorporeal membrane oxygenation (ecmo) |
|  | DX_ICD10 | Z94.1 | hrt transplant status |
|  | DX_ICD10 | Z94.3 | hrt and lungs transplant status |
|  | DX_ICD10 | Z95.811 | presence of hrt assist device |
|  | DX_ICD10 | Z95.812 | presence of fully implantable artificial hrt |
| **Hypertension** 41 ICD-9 DX 54 ICD-10 DX **95 Total** | DX_ICD9 | 362.11 | hypertensive retinopathy |
|  | DX_ICD9 | 401.0 | malignant hypertension |
|  | DX_ICD9 | 401.1 | benign hypertension |
|  | DX_ICD9 | 401.9 | hypertension nos |
|  | DX_ICD9 | 402.00 | mal hyperten hrt dis nos |
|  | DX_ICD9 | 402.01 | mal hypert hrt dis w chf |
|  | DX_ICD9 | 402.10 | ben hyperten hrt dis nos |
|  | DX_ICD9 | 402.11 | benign hyp hrt dis w chf |
|  | DX_ICD9 | 402.90 | hypertensive hrt dis nos |
|  | DX_ICD9 | 402.91 | hyperten hrt dis w chf |
|  | DX_ICD9 | 403.0 | mal hypertens renal dis (begin 1980 end 1989) |
|  | DX_ICD9 | 403.00 | mal hyp ren w/o ren fail (begin 1989) |
|  | DX_ICD9 | 403.01 | mal hyp ren w renal fail (begin 1989) |
|  | DX_ICD9 | 403.1 | benign hypert renal dis (begin 1980 end 1989) |
|  | DX_ICD9 | 403.10 | ben hyp ren w/o ren fail (begin 1989) |
|  | DX_ICD9 | 403.11 | ben hyp renal w ren fail (begin 1989) |
|  | DX_ICD9 | 403.9 | hypertens renal dis nos (begin 1980 end 1989) |
|  | DX_ICD9 | 403.90 | hyp ren nos w/o ren fail (begin 1989) |
|  | DX_ICD9 | 403.91 | hyp renal nos w ren fail (begin 1989) |
|  | DX_ICD9 | 404.0 | mal hypert hrt/renal dis (begin 1980 end 1989) |
|  | DX_ICD9 | 404.00 | mal hy ht/ren w/o chf/rf (begin 1989) |
|  | DX_ICD9 | 404.01 | mal hyper hrt/ren w chf (begin 1989) |
|  | DX_ICD9 | 404.02 | mal hy ht/ren w ren fail (begin 1989) |
|  | DX_ICD9 | 404.03 | mal hyp hrt/ren w chf & rf (begin 1989) |
|  | DX_ICD9 | 404.9 | hypert hrt/renal dis nos (begin 1980 end 1989) |
|  | DX_ICD9 | 404.1 | ben hypert hrt/renal dis (begin 1980 end 1989) |
|  | DX_ICD9 | 404.10 | ben hy ht/ren w/o chf/rf (begin 1989) |
|  | DX_ICD9 | 404.11 | ben hyper hrt/ren w chf (begin 1989) |
|  | DX_ICD9 | 404.12 | ben hy ht/ren w ren fail (begin 1989) |
|  | DX_ICD9 | 404.13 | ben hyp hrt/ren w chf & rf (begin 1989) |
|  | DX_ICD9 | 404.90 | hy ht/ren nos w/o chf/rf (begin 1989) |
|  | DX_ICD9 | 404.91 | hyper hrt/ren nos w chf (begin 1989) |
|  | DX_ICD9 | 404.92 | hy ht/ren nos w ren fail (begin 1989) |
|  | DX_ICD9 | 404.93 | hyp ht/ren nos w chf & rf (begin 1989) |
|  | DX_ICD9 | 405.01 | mal renovasc hypertens |
|  | DX_ICD9 | 405.09 | mal second hyperten nec |
|  | DX_ICD9 | 405.11 | benign renovasc hyperten |
|  | DX_ICD9 | 405.19 | benign second hypert nec |
|  | DX_ICD9 | 405.91 | renovasc hypertension |
|  | DX_ICD9 | 405.99 | second hypertension nec |
|  | DX_ICD9 | 437.2 | hypertens encephalopathy |
|  | DX_ICD10 | H35.031 | hypertensive retinopathy, right eye |
|  | DX_ICD10 | H35.032 | hypertensive retinopathy, left eye |
|  | DX_ICD10 | H35.033 | hypertensive retinopathy, bilateral |
|  | DX_ICD10 | H35.039 | hypertensive retinopathy, unspecified eye |
|  | DX_ICD10 | I10 | essential (primary) hypertension |
|  | DX_ICD10 | I11.0 | hypertensive hrt disease w hrt failure |
|  | DX_ICD10 | I11.9 | hypertensive hrt disease wo hrt failure |
|  | DX_ICD10 | I12.0 | hypertensive chronic kidn dis w stg5 chron kidn dis or end stg renal dis |
|  | DX_ICD10 | I12.9 | htn chron kidn dis w stg1-stg4 chron kidn dis, or unspec chron kidn dis |
|  | DX_ICD10 | I13.0 | htn hrt & chron kid w hrt fail & stg1-stg4 chron kid, or unspec chron kid dis |
|  | DX_ICD10 | I13.10 | htn hrt & chron kid wo hrt fail, w stg1-stg4 chron kid, or unspec chron kid |
|  | DX_ICD10 | I13.11 | htn hrt & chron kidn wo hrt fail, w stg5 chron kidn or end stg renal dis |
|  | DX_ICD10 | I13.2 | htn hrt & chron kidn w hrt fail & w stg5 chron kidn, or end stg renal dis |
|  | DX_ICD10 | I15.0 | renovascular hypertension |
|  | DX_ICD10 | I15.1 | hypertension secondary to other renal disorders |
|  | DX_ICD10 | I15.2 | hypertension secondary to endocrine disorders |
|  | DX_ICD10 | I15.8 | oth secondary hypertension |
|  | DX_ICD10 | I15.9 | secondary hypertension, unspecified |
|  | DX_ICD10 | I16.0 | hypertensive urgency |
|  | DX_ICD10 | I16.1 | hypertensive emergency |
|  | DX_ICD10 | I16.9 | hypertensive crisis, unspecified |
|  | DX_ICD10 | I67.4 | hypertensive encephalopathy |
|  | DX_ICD10 | I87.9 | disorder of vein, unspecified |
|  | DX_ICD10 | I97.3 | postprocedural hypertension |
|  | DX_ICD10 | O10.111 | pre-existing hypertensive hrt disease complicating preg, first trimester |
|  | DX_ICD10 | O10.112 | pre-existing hypertensive hrt disease complicating preg, second trimester |
|  | DX_ICD10 | O10.113 | pre-existing hypertensive hrt disease complicating preg, third trimester |
|  | DX_ICD10 | O10.119 | pre-existing hypertensive hrt disease complicating preg, unspecified tri |
|  | DX_ICD10 | O10.12 | pre-existing hypertensive hrt disease complic childbirth |
|  | DX_ICD10 | O10.13 | pre-existing hypertensive hrt disease complic puerperium |
|  | DX_ICD10 | O10.211 | pre-existing hypertensive chronic kidney dis complic preg, first trimester |
|  | DX_ICD10 | O10.212 | pre-existing hypertensive chronic kidney dis complic preg, second tri |
|  | DX_ICD10 | O10.213 | pre-existing hypertensive chronic kidney dis complic preg, third trir |
|  | DX_ICD10 | O10.219 | pre-existing hypertensive chronic kidney dis complic preg, unspec tri |
|  | DX_ICD10 | O10.22 | pre-existing hypertensive chronic kidney dis complic childbirth |
|  | DX_ICD10 | O10.23 | pre-existing hypertensive chronic kidney dis complic puerperium |
|  | DX_ICD10 | O10.311 | pre-existing hypertensive hrt and chronic kidney dis complic preg, first tri |
|  | DX_ICD10 | O10.312 | pre-existing hypertensive hrt and chronic kidney dis complic preg, second tri |
|  | DX_ICD10 | O10.313 | pre-existing hypertensive hrt & chronic kidney dis complic preg, third tr |
|  | DX_ICD10 | O10.319 | pre-existing hypertensive hrt & chronic kidney dis complic preg, unspec tri |
|  | DX_ICD10 | O10.32 | pre-existing hypertensive hrt & chronic kidney dis complic childbirth |
|  | DX_ICD10 | O10.33 | pre-existing hypertensive hrt & chronic kidney dis complic puerperium |
|  | DX_ICD10 | O10.411 | pre-existing secondary hypertension complic preg, first trimester |
|  | DX_ICD10 | O10.412 | pre-existing secondary hypertension complic preg, second trimester |
|  | DX_ICD10 | O10.413 | pre-existing secondary hypertension complic preg, third trimester |
|  | DX_ICD10 | O10.419 | pre-existing secondary hypertension complic preg, unspec trimester |
|  | DX_ICD10 | O10.42 | pre-existing secondary hypertension complic childbirth |
|  | DX_ICD10 | O10.43 | pre-existing secondary hypertension complic puerperium |
|  | DX_ICD10 | O11.1 | pre-existing hypertension w pre-eclampsia, first tri |
|  | DX_ICD10 | O11.2 | pre-existing hypertension w pre-eclampsia, second tri |
|  | DX_ICD10 | O11.3 | pre-existing hypertension w pre-eclampsia, third tri |
|  | DX_ICD10 | O11.4 | pre-existing hypertension w pre-eclampsia, complic childbirth |
|  | DX_ICD10 | O11.5 | pre-existing hypertension w pre-eclampsia, complic puerperium |
|  | DX_ICD10 | O11.9 | pre-existing hypertension w pre-eclampsia, unspec tri |
